# An Atlas of Indian Genetic Diversity

**DOI:** 10.64898/2026.03.20.26348801

**Authors:** Krithika Subramanian, Chandrika Bhattacharyya, Pratheusa Machha, Ankit Mukherjee, Devashish Tripathi, Shreya Chakraborty, Sauma Suvra Majumdar, Shouvanik Sengupta, Priyanka Singh, Vinay More, Shreya Bari, MS Sreelekshmi, Eric Macwan, Debasrija Mondal, Arghya Dey, Haya Afreen, Siddhi P Jani, Payel Mukherjee, Neha Singh, Tiyasha De, Pooja Sharma, Bharathram Upilli, Arindam Maitra, Kuldeep Singh, Praveen Sharma, Nanaocha Sharma, Sunil K. Raghav, Punit Prasad, E. V. Soniya, Abdul Jaleel, M Radhakrishna Pillai, Shijulal Nelson Sathi, Madhvi Joshi, Chaitanya Joshi, Mayurika Lahiri, Santosh Dixit, L. S. Shashidhara, Nachimuthu Senthil Kumar, H. Lalhruaitluanga, Lal Nundanga, Venkataram Shivakumar, Ganesan Venkatasubramanian, Naren P. Rao, Mohd Ashraf Ganie, Imtiyaz Ahmad Wani, Ganganath Jha, Ashwin Dalal, Murali Dharan Bashyam, Pritish Kumar Varadwaj, BS Sanjeev, Yogesh Simmhan, Chirag Jain, Arun Kumar, Durai Sundar, Ishaan Gupta, Pankaj Yadav, Manikandan Narayanan, Karthik Raman, Padinjat Raghu, Radhakrishnan Sabarinathan, Sridhar Sivasubbu, Vinod Scaria, GenomeIndia Consortium, Prathima Arvind, Suman K Paine, Tulasi Nagabandi, Khader Valli Rupanagudi, Himanshu Sinha, Nidhan K. Biswas, Shweta Ramdas, Karthik Bharadwaj Tallapaka, Vijayalakshmi Ravindranath, Yadati Narahari, Kumarasamy Thangaraj, Divya Tej Sowpati, Mohammed Faruq, Bratati Kahali, Analabha Basu

## Abstract

India, the most populous country, remains significantly underrepresented in the global genomics landscape. Previous efforts to catalog Indian genetic diversity were limited in scale, scope, and representation. Here, we present the GenomeIndia dataset, comprising whole genome sequences of 9,768 healthy individuals from 83 populations spanning the ethnolinguistic and biogeographic spectrum of India. We identify 129.93 million high-confidence biallelic variants, 44.03 million of which are previously unreported in global databases. In contrast to large populations that show steady population growth and internal homogeneity, we observe low effective population sizes, significant genetic drift, and profound homozygosity in small tribal groups, likely shaped by antiquity, isolation, and endogamy. We report multiple population-specific pharmacogenomic and deleterious variants, necessitating the integration of local genetic architecture and the inclusion of underrepresented South Asian genomes in global reference resources. Finally, we highlight the limited transferability of Eurocentric polygenic scores to Indian populations, and present an imputation panel that outperforms existing resources for both rare and common variants. Together, our work fills a significant gap in the equity of global human genomics, and paves way for precision medicine strategies that will benefit a quarter of the world population.

## Main

India, with over 1.4 billion people, represents one of the most genetically complex regions in the world. Yet, it remains significantly underrepresented in global genomic databases ^1^. This disparity limits the transferability of risk prediction models and clinical tools, and constrains the development of precision medicine strategies. India’s population structure, driven by migration, complex admixture, and widespread endogamy, provides a powerful framework for understanding how demographic events and socio-cultural history influences genetic diversity. A population-scale genomic survey of India is therefore essential to improve the resolution, accuracy, and equity of global human genomics data.

Recent genomic initiatives across Africa, Latin America, and Asia have demonstrated that population-specific datasets are vital for uncovering novel variations and ensuring equitable precision health ^2–4^. In India, earlier array-based studies established how endogamy, language, and geography shape genetic structure ^5–7^. Subsequent sequencing-based studies, including GenomeAsia (n=598), IndiGen (n=1,029), and LASI-DAD (n=2,762), expanded these insights ^8–10^. However, these efforts were either modest in scale, biased toward urban populations, age-biased, or focused on specific health conditions such as dementia, thereby failing to adequately capture the population-level genetic diversity across India.

To address this gap, the GenomeIndia project systematically sampled 9,768 individuals from 83 anthropologically defined, endogamous populations spanning the ethnolinguistic and biogeographic spectrum of India. Our analyses: 1) resolve fine-scale population structure, revealing multiple, previously unresolved drifted and founder populations; 2) catalog 129.93 million high-confidence biallelic variants including many high frequency novel alleles, particularly among tribal populations; 3) reveal population-specific distributions of clinically relevant, loss-of-function, and pharmacogenetic variants; 4) identify founder populations enriched for variants influencing disease risk; 5) delineate large population groups that can power ancestry-informed design of genome-wide association studies; 6) demonstrate limited transferability of Eurocentric polygenic scores, underscoring the need for ancestry-matched prediction models; and 7) introduce a high-resolution Indian imputation panel that significantly improves South Asian genotype inference and lays the foundation for India-specific genotyping chip development.

Collectively, this work establishes the most comprehensive genomic reference for the Indian subcontinent, creating a globally relevant resource for studying human diversity and advancing equitable precision medicine.

### The GenomeIndia Dataset

The GenomeIndia initiative was designed to capture the anthropological, linguistic, and socio-cultural diversity of the Indian subcontinent. Participants were recruited from all major geographical regions of India, representing the country’s principal language families: Indo-European (IE), Dravidian (DR), Austroasiatic (AA), and Tibeto-Burman (TB). The sampling scheme encompassed both Tribal (T) and Non-tribal (NT) populations, and an additional continentally admixed outgroup (CAO), representing a wide range of social strata and different degrees of endogamy (Fig 1a). The study was implemented through the coordinated efforts of academic and medical institutions across India following uniform protocols for recruitment, sample collection, metadata annotation, and phenotypic measurements (S1; Bhattacharyya et al. 2025).

**Figure 1:**
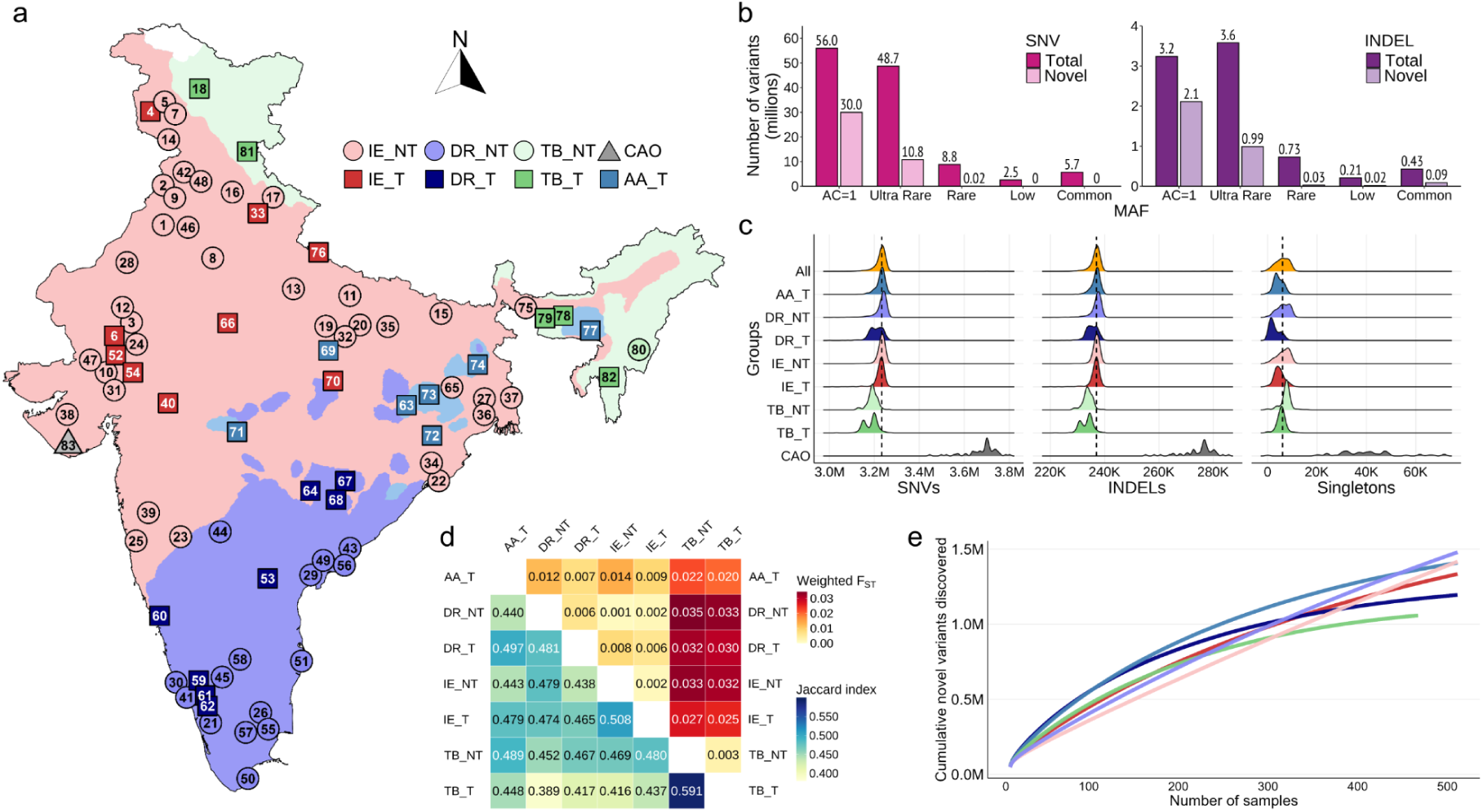
Overview of the GenomeIndia dataset. **a)** A map of India depicting the geographical distribution of the populations sampled in the GenomeIndia project. The shape of the point indicates whether the population is a recognized tribe (square) or not (circle), while the color indicates the linguistic group to which the population belongs. In addition, a continentally admixed outgroup (triangle) is also sampled. IE_T: Indo-European Tribe; IE_NT: Indo-European Non-Tribe; DR_T: Dravidian Tribe; DR_NT: Dravidian Non-Tribe; TB_T: Tibeto-Burman Tribe; TB_NT: Tibeto-Burman NonTribe; AA_T: Austro-Asiatic Tribe; CAO: Continentally Admixed Outgroup. **b)** Total and Novel (previously unreported) number of Single Nucleotide Variants (SNV; left) and small Insertions and Deletions (INDEL; right) identified in the GenomeIndia dataset, stratified by their allele frequencies. AC=1: Singletons (allele count of 1); Ultra Rare: AC > 1 and minor allele frequency (MAF) < 0.001; Rare: MAF >= 0.001 and < 0.01; Low: MAF >= 0.01 and < 0.05; Common: MAF >= 0.05. The numbers in each category, in millions, are indicated at the top of each bar. **c)** Ridge plots showing the distribution of SNVs (left), INDELs (middle), and Singletons (right) in the 8 ethnolinguistic groups as described in panel a. M - Millions; K - Thousands. The top ridge plot labelled ‘All’ indicates the distribution for the entire GI dataset. The dashed vertical line in each plot indicates the median value for the GI dataset. **d)** A matrix representing the shared allele proportion (bottom left triangle) using Jaccard index, and the Fixation index (F_ST_) of the 7 ethnolinguistic groups (excluding CAO) as described in panel a. **e)** Cumulative novel variant discovery stratified by ethnolinguistic groups as described in panel a. All groups were randomly downsampled to 500 individuals, except for TB_T, where all individuals (n = 475) were used. TB_NT are not represented in this plot due to limited sample size (n = 159).

A total of 20,195 individuals representing 83 diverse populations were enrolled in the study (Table S1.4). Whole-genome sequencing (WGS) at ∼30× coverage was performed on 10,074 individuals, including 244 trios, of whom 9,768 passed stringent quality control filters and met inclusion criteria for downstream analyses (S2). From this dataset, we identified 129,938,889 high-confidence biallelic variants, comprising ∼121 Million (M) single-nucleotide variants (SNVs) (93.7%) and ∼8M insertions/ deletions (InDels; 6.3%) (Fig 1b). Among these variants, ∼59M (45.55%) are singletons. Of the 70.74M non-singleton variants, 11.94M (9.19%) are unreported in global databases (Fig 1b, S3), demonstrating that a substantial fraction of human variation remains uncatalogued, residing within previously unsampled populations.

The allele frequency (AF) spectrum was dominated by rare variation. Among the non-singleton variants, 52.3M (73.93%) are ultra-rare (MAF < 0.1%), 9.6M are rare (0.1%-1%), 2.75M are low-frequency (1%-5%), and 6.1M are common (> 5%). This long tail of rare variation, often confined to specific endogamous populations, is critical for deciphering the genetic basis of Mendelian and complex diseases. In conjunction with genetic data, individuals are annotated with phenotypic and other metadata, including socio-demographic information, self-reported health status, anthropometric measures, complete blood counts, and biochemical assays relevant to metabolic and organ health (S1). Collectively, the GenomeIndia dataset is one of the largest and richly annotated genomic resources from South Asia, providing a foundation for genomics-informed public health and precision medicine.

### Population Structure and Genetic Differentiation

Individuals were grouped into 8 predefined ethnolinguistic categories: IE_NT, IE_T, DR_NT, DR_T, AA_T, TB_NT, TB_T, and CAO ^11^. All population analyses were done on 9,686 individuals, excluding 82 outliers who were significantly distant from their population labels in the Principal Component Analysis (S2). Variant patterns differed markedly across ethnolinguistic groups. DR_T and AA_T populations exhibited large numbers of total variants but relatively few singletons, consistent with antiquity and long-term demographic continuity (S3, Fig. 1c) ^12^. In contrast, TB populations carried fewer total variants yet a high proportion of singletons, a combination suggestive of limited founding diversity followed by population expansion (Fig. 1c, S3.1.1, S3.1.2).

### Population Differentiation and Allele Sharing Within Linguistic Families

Pairwise F_ST_ indicated significant genetic divergence in TB populations and deep differentiation of DR_T populations from the Nilgiris (Fig. 1d, Fig. S3.3.1). Jaccard similarity showed highest allele sharing within linguistic families (TB_NT–TB_T, IE_NT–IE_T, DR_NT–DR_T), supporting the utility of linguistic labels in population genetics (Fig. 1d). Reflecting widespread admixture predating endogamy ^7,13^, ∼4.9M common variants were shared across all seven groups, including ∼76,000 (1.55%) absent from 1000G, gnomAD and GenomeAsia. This core variant set may enable design of genotyping arrays with broad relevance for Indian populations and enhance multi-ancestry performance.

### Contrasting Novel Variant Discovery in Tribal and Non-Tribal Populations

Rarefaction analysis of novel variants (downsampled to ∼500 individuals per group) revealed contrasting trajectories. The non-tribal IE_NT and DR_NT populations displayed steadily rising curves, reflecting broad admixture and large rare-variant reservoirs. On the other hand, tribal groups (DR_T, AA_T, TB_T) harbored 131,257 high-frequency, population-specific novel variants, which are locally common (MAF>=1%) yet globally rare, causing their rarefaction curves to rise sharply but plateau rapidly (Fig. 1e). These findings justify our sampling design: large cohorts from multiple IE_NT and DR_NT populations to capture extensive internal diversity, and small sample sizes (n≈50–100) from isolated tribal groups.

### Geography, Language, and Biogeography Shape Genetic Structure Across India

Principal component analysis of 9,475 individuals (excluding CAO and offsprings of trios) revealed clustering by population, with IE-NT and DR-NT groups forming a substantial overlapping and an approximate north-to-south cline consistent with previous studies (Fig. 2a, S4.1) ^6–8,13^. AA_T, DR_T, and TB_T populations formed tight, isolated clusters reflecting genetic homogeneity and endogamy, while few populations showed diffuse patterns or clines suggesting admixture (Fig. 2a). Within-group PCA placed tribal populations consistently at peripheries, reflecting their genetic distinctiveness and isolation relative to nearby non-tribal populations (S4.1).

**Figure 2:**
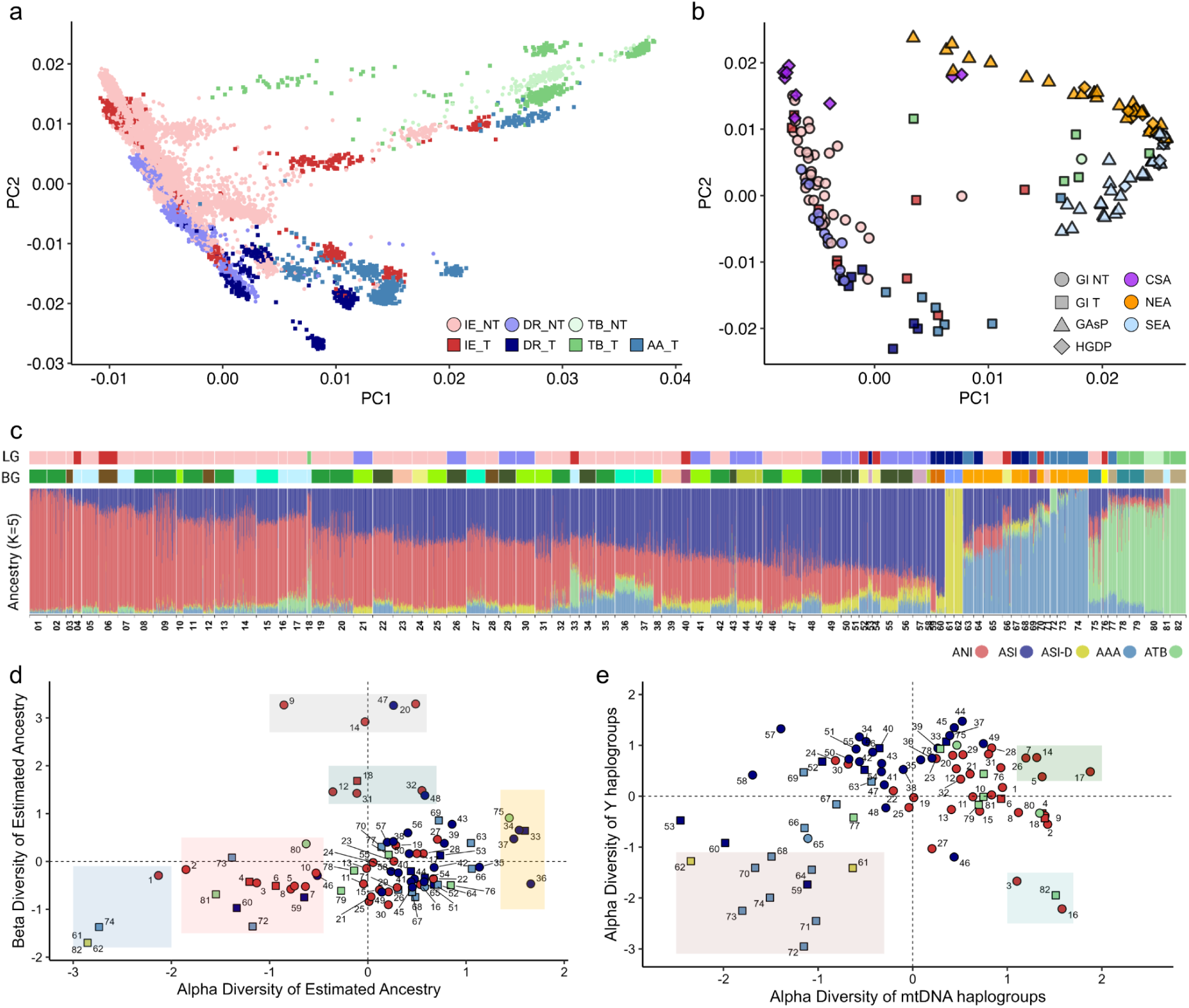
Population structure and uniparental analysis of GenomeIndia data. **a)** Principal component analysis (PCA) of 9,475 individuals from the GenomeIndia dataset, excluding population outliers and individuals from the CAO group. Each dot, representing one individual, is colored by the ethnolinguistic group they belong to, as described in Figure 1. **b)** PCA of population centroids from the GenomeIndia dataset in the context of other geographically neighboring populations sampled in the Genome Asia Project (GAsP; Triangles) and Human Genome Diversity Project (HGDP; Diamonds). The color and shape of the GI populations are the same as those of Figure 2a. **c)** ADMIXTURE analysis of unrelated (n = 9,475) individuals from 82 GI populations, excluding CAO. ANI - Ancestral North Indian; ASI - Ancestral South Indian; ASI-D - Ancestral South Indian-Dravidian; AAA - Ancestral Austro-Asiatic; ATB - Ancestral Tibeto-Burman. The top two annotation bars represent the ethnolinguistic group (LG) of each population, as shown in Figure 2a, and the biogeography (BG) of the population, respectively. The color legend for biogeography is available in Figure S4.2. **d)** Diversity analysis of admixture proportions in 82 GI populations using standardized Alpha (x-axis) and Beta (y-axis) measures. Each point corresponds to a numerically labeled population in accordance with the ADMIXTURE plot, with colors indicating the dominant ADMIXTURE cluster, and shapes indicate Tribal (squares) and Non-tribal (circles) groups. Vertical and horizontal dashed lines represent the average (z-scores of 0) Alpha and Beta diversities, respectively. Shaded areas highlight interesting groups of populations: Blue - extremely low Alpha and Beta diversities, representing founder populations; Red - low Alpha and moderate Beta diversity; Green and Grey - high and extremely high Beta diversities, respectively, but average Alpha diversity; Gold - extremely high Alpha diversity but moderate Beta diversity. **e)** Diversity analysis of uniparental markers using standardized Alpha diversity of mitochondrial (x-axis) and Y-chromosome (y-axis) haplogroups. The color and shape are the same as those of Figure 2d.

Regression modelling demonstrated a strong linkage between genetics and geography. Longitude significantly predicted PC1 (adjusted R^2^ = 0.43) and latitude predicted PC2 (adjusted R^2^ = 0.30) (S4.2, Model 1). This concordance was reinforced by incorporating 1,831 neighboring regional samples from Central South Asia (CSA), East and Southeast Asia (SEA), and North East Asia (NEA) from HGDP^14^ and GAsP ^8^. IE speakers from extreme North and Northwest India overlapped with geographically proximal CSA populations, while TB-speakers from Northeast India clustered near EA and SEA populations, reflecting regional continuity (Fig. 2b, S4.1.3). Biogeographic clustering was apparent overall and within linguistic groups (S4.1). Incorporating linguistic affiliation and biogeography dramatically improved model fit (S4.2, Models 2 and 3); a unified model incorporating all four factors explained 84% of PC1 variance and 78% of PC2 variance (S4.2: Model 4). These results reinforce that Indian genetic structure is geographically patterned and deeply intertwined with linguistic history and ecological context (S4).

### Four Major Ancestry Components and Population-Specific Drift

ADMIXTURE ^15^ analysis identified four broad ancestry components consistent with previous findings ^7^: Ancestral North Indian (ANI), Ancestral South Indian (ASI), Ancestral Austroasiatic (AAA), and Ancestral Tibeto-Burman (ATB) (S4.3). Increasing K to 5 reduced cross-validation error, revealing a fifth ancestral component representing two genetically cohesive, isolated DR_T populations from the Nilgiris hills (DR_NGH_1_02 (61) and DR_NGH_1_01(62)), shaped by strong population-specific drift consistent with F_ST_ and PCA findings (Fig. 2c, S4.3). Higher K values (>5) predominantly reflected group-specific drift effects rather than broader shared ancestries (S4.3).

To assess admixture complexity, we quantified the within-individual (Shannon’s alpha) and between-individual (Bray-Curtis beta) diversity of the admixture components for each of the 82 populations (S4.4, methods). Most populations showed low to moderate beta diversity, irrespective of alpha diversity (Fig. 2d), indicating that panmixing of populations after the initial admixture has homogenized their ancestry proportions. Ancestry “reference” groups exhibited extremely low alpha and beta diversity with minimal admixture and internal homogeneity. Conversely, highly admixed “melting pot” populations (Western Himalayas, Eastern Coastal Plains, Riverine Plains, and the Brahmaputra Valley) showed high alpha and moderate beta diversity with multiple ancestry components per individual (details in S4.4). This analysis guides population stratification for future disease gene mapping studies in India.

### Uniparental Data Analysis Reveals Sex-Biased Migrations

Uniparental markers complemented autosomal findings and revealed maternal–paternal asymmetries (Fig. 2e). mtDNA macrohaplogroup M and its basal subclades occurred at high frequencies with low diversity among AA_T and DR_T groups, consistent with antiquity, limited founding maternal lineages, and pre-Neolithic expansions (S5; Basu et al. 2003). Recent R/R0 derivatives (U, K, H, V) correlated positively with ANI ancestry (Van Oven and Kayser 2009; Brotherton et al. 2013; S5). Populations with high ANI ancestry were positioned above the regression line in the mtDNA haplogroup frequencies, indicating male-biased admixture contributing to ANI, while the ancestral haplogroup R showed no correlation, suggesting early diffusion before migrations that consolidated ANI ancestry (Larruga et al. 2017; S5).

Y-haplogroups C, H, J, L1, O, and R accounted for >90% of male lineages, with H1, R1, and R2 comprising ∼60% of all lineages (Fig. S5.3.3), consistent with previous studies ^19,20^. High frequency of R1a1 in IE-speakers correlated with high ANI ancestry (Fig. S5.3.8), while maternal lineages remained largely locally derived, confirming a male-biased Central-Asian and West-Eurasian influx ^21,22^. TB and AA populations showed distinct subclade enrichment of Y-chromosome O-lineage (O2a in TB, O1b in AA), indicating separate male-mediated dispersals, possibly from SEA ^12,23^. DR_NT populations displayed elevated H1, H3, L1, and J2 frequencies, consistent with pre-IE establishment across India ^12^. The two DR_T populations (61 and 62), classified as ASI-D by ADMIXTURE, exhibited a unique Y-haplogroup profile, characterized by the rare H3 haplogroup in two-thirds of males, which was much higher than in other Dravidian groups. The CAO group exhibited unique African-origin haplogroups (B2, E2, E1) ^24–26^. Collectively, these patterns indicate pronounced paternal heterogeneity and specific haplogroup distributions driven by historical demographic processes (S4.5, S5).

### Low Ne and High Inbreeding Burden in Tribal Communities

Like global non-African populations, Indian populations exhibited post-Out-of-Africa (OoA) recovery and the Last Glacial Maximum (LGM) expansion signatures (Li and Durbin 2011; Tallavaara et al. 2015; Gamble et al. 2005; S6). However, several tribal populations showed persistently low effective population sizes (Ne), reflecting isolation, stagnancy, and genetic drift (Figure 3a, 3b, S6). The two populations contributing to the ASI-D ancestry (61 and 62) displayed particularly low Ne trajectories, with minimal demographic expansion compared to other populations, specifically non-tribals. Clustering of the Ne trajectories identified three distinct categories, including one harboring eight tribal populations with limited growth beginning ∼2000 generations ago, followed by prolonged demographic stagnation (S6).

**Figure 3:**
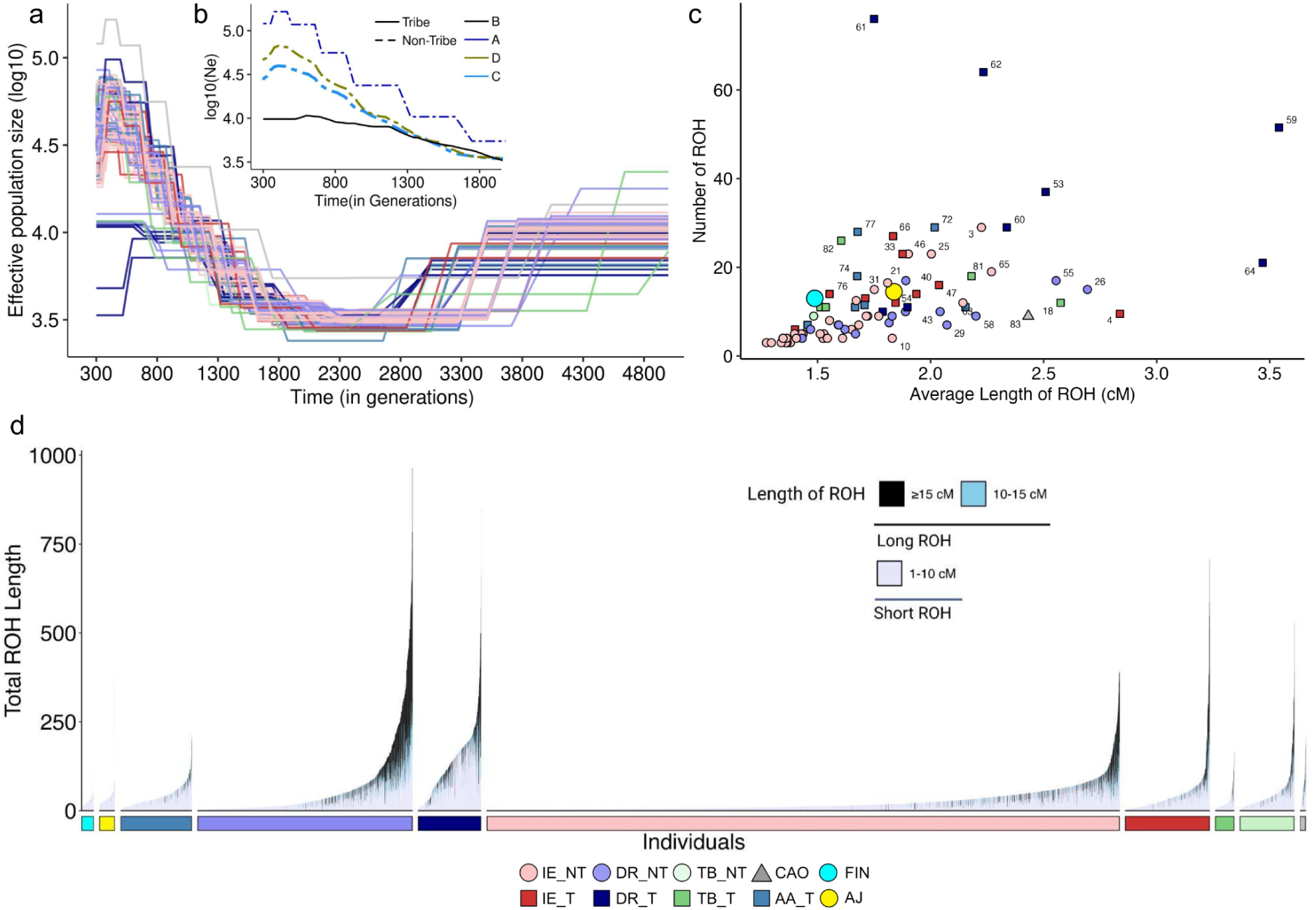
Demographic inference and Runs of Homozygosity analysis. **a)** Estimation of effective population size (Ne; y-axis)) over time (x-axis; 300-5000 generations ago) for 83 GI populations. Each line is colored by the ethnolinguistic group it belongs to, as indicated by the legend at the bottom of the figure. **b)** A summary plot depicting the change in Ne (y-axis) over time (x-axis; 300-2000 generations ago) of 4 different population clusters identified in our study. Dashed lines indicate clusters of non-tribal populations, and the solid lines represent clusters of tribal populations. For panels c) and d), Finnish (FIN) and Ashkenazi Jews (AJ) are included for global reference. **c)** A scatterplot of the average length of ROH (LROH) in centimorgans (cM) vs the number of ROH segments (NROH) of 83 GI populations. The color and shape of the GI populations are according to the ethnolinguistic group. **d)** Total ROH in cM observed in 9442 individuals from the GI cohort. The annotation bar indicates the ethnolinguistic group to which the individual belongs. The stacks in each bar are colored by the length of the ROH segments contributing to the total ROH - pale blue: short (<10 cM); sky blue: long (10-15 cM); black: very long (>15 cM).

We computed three summary measures of Runs of Homozygosity (ROH): (a) the total number of ROH segments (NROH), (b) the average ROH length (LROH), and (c) the genomic proportion in ROH (FROH). Comparison with Ashkenazi Jews (AJ) and Finnish (FIN), considered global benchmarks of inbred populations, revealed substantially higher homozygosity (Fig. 3c, Table S7.1, Table S7.2) in the GI cohort.

Median LROH >1.5cM (endogamy threshold) ^30^ was significantly exceeded in 59 of 83 GI populations of which 23 had Median LROH larger than the AJ (one-sided Kolmogorov–Smirnov test, FDR 5%, Figure 3c, Figure S7.1). Comparing NROH, which epitomize “founder effect” ^31^, revealed 22 and 21 GI populations having higher NROH than FIN (median NROH = 13) and AJ (median NROH = 14.5), respectively. As small Ne can increase cryptic-relatedness in populations, we investigated the relationship of NROH and LROH with Ne in the period just predating the Neolithic Demographic Transition (NDT), i.e. 300 generations ago (S7). Notably, with median NROH of 76 and 64 respectively, DR_T populations 61 and 62 rank among the world’s highest ^31^; >5-fold greater than AJ and FIN (Figure S7.2). Other DR_T populations (59, 53, 64) also exhibited extremely high inbreeding burden resulting from the combination of founder effects and endogamy (high NROH and LROH). We observed an inverse correlation between the median NROH and Ne, with the DR_T populations occupying the top 5 positions. However, more than 50 populations with significantly long median LROH have considerably large Ne, underscoring the impact of endogamy in the GI populations (Figure S7.2). AA_T and TB_T populations carried predominantly short LROH (<10cM), while some DR_NT showed elevated proportions of long LROH (>10cM), indicating persistent cultural endogamy (Figure 3d, Figure S7.3). Individuals from the CAO population, despite their known history of recent admixture, also showed long ROH, suggesting post-admixture isolation and ongoing endogamy (Figure S7.3).

Approximately 2,700 individuals harbor ROH in ≥1% of their genomes (FROH ≥0.01). 80% of them are from six DR_T populations (62, 61, 59, 64, 60, and 53), one AA_T (72), and two IE_T (33 and 66) populations (Fig. 3d, Fig. S7.4). Although the IE_NT as a whole had limited ROH burden yet several populations (3, 25, 46, 65) had >70% individuals with FROH ≥0.01. This suggests that fine-scale sociocultural structures influencing cryptic-relatedness may exist even within these large, non-tribal communities. These findings underscore the urgent need for population-informed genetic counseling, carrier and newborn screening (like JScreen or Dor Yeshorim programs), and rare disease diagnostics into national health initiatives for equitable precision medicine.

### Variants of Medical Importance

We identified ∼1.5 million protein-coding variants, including 763,352 non-singletons (Supplementary Figure S8.1). These comprised 25,124 putative deleterious missense variants (pDMM) (REVEL ≥ 0.644 and CADD ≥ 20) ^32,33^, across 7,465 genes (Supplementary Table S9.1) and 15,849 high-confidence loss-of-function (HC-LoF) variants across 7,076 genes, of which 9% and 17%, respectively, were absent from global variant databases, highlighting substantial uncharacterized functional variation.

Most genes are observed to carry only one HC-LoF or pDMM (Figure S10.2), as the accumulation of deleterious variants within the same gene is typically an evolutionary constraint. We used the pLI scores ^34^ to identify 134 high-pLI genes (pLI ≥ 0.9) with novel HC-LoFs and 31 with additional HC-LoFs absent from gnomAD. While globally rare, isolation and endogamy have driven these variants to higher frequencies locally (Fig 4a, Table S9.1, Table S10.1), highlighting the critical value of studying underrepresented populations for disease risk profiling and reevaluation of existing benchmarks.

**Figure 4:**
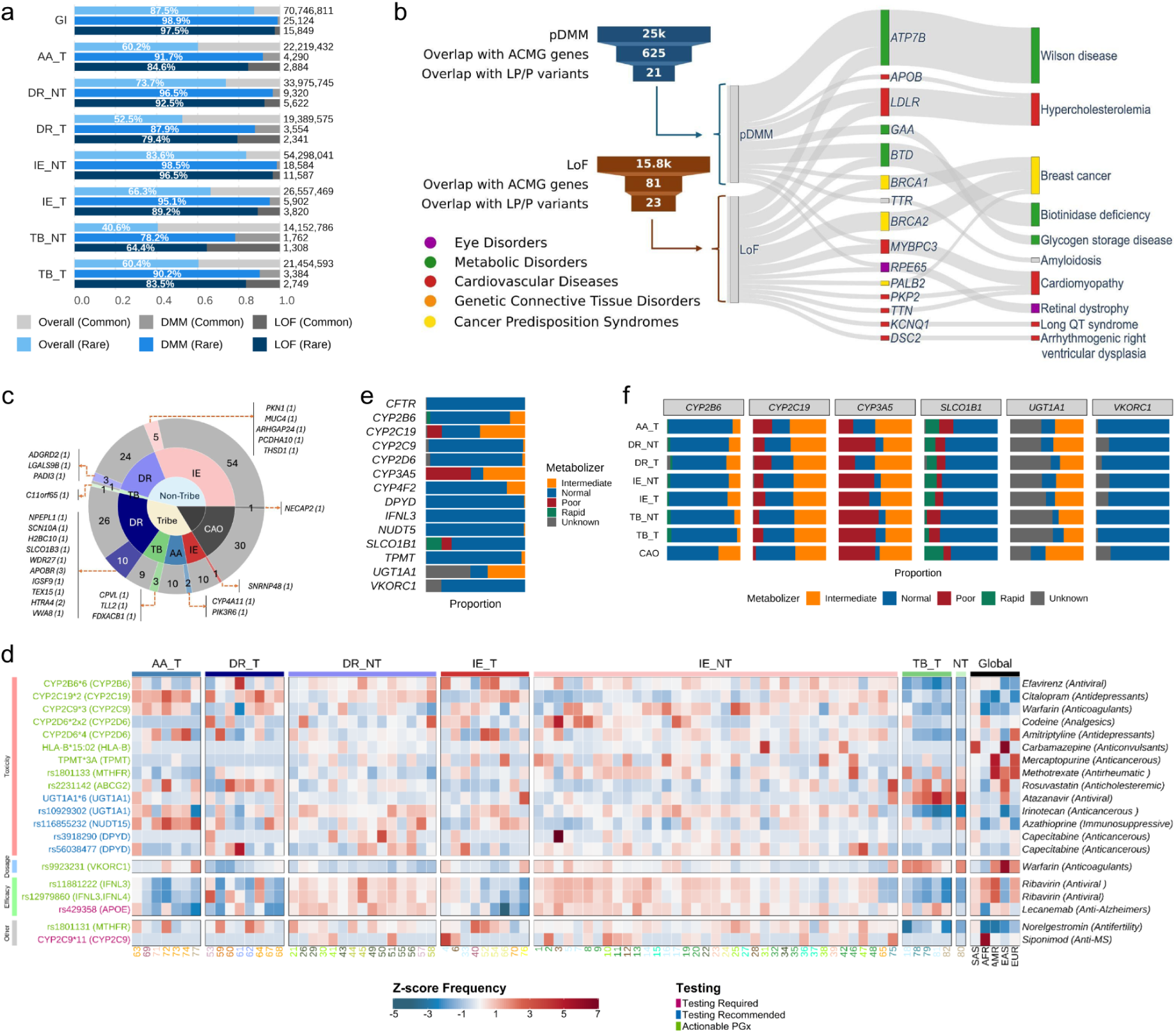
Medical relevance of the GenomeIndia dataset. **a)** Ethnolinguistic group-wise distribution of deleterious missense mutations (DMM) and loss-of-function (LoF) variants. Variants are categorized as rare (MAF < 1%; shades of blue) or common (MAF > 1%; shades of grey). The “overall” counts indicate the total number of non-singleton variants in the group. **b)** A sankey diagram representing the distribution of 44 pathogenic DMM and LoF variants overlapping medically actionable genes as per ACMG (American College of Medical Genetics and Genomics) guidelines. The overlapping gene and the implicated disease are indicated by the middle and right annotation bars. **c)** Distribution of population-specific known and novel homozygous loss-of-function variants across linguistic subgroups. The innermost ring represents major groups: Tribe, Non-Tribe, and CAO. The middle ring indicates linguistic affiliations, and the outermost ring shows counts of previously reported (grey) and novel (colored) variants. Segment sizes reflect variant counts. Six novel variants shared among populations are not shown. **d)** Z-score–standardized allele frequencies for CPIC-recommended pharmacogenomic variants (star alleles and SNPs) across 82 Indian subpopulations, grouped by linguistic and tribal affiliations, with comparisons to global 1000 Genomes populations (SAS, AFR, AMR, EAS, and EUR). Variants are annotated with associated drugs and therapeutic classes, and coloured according to CPIC recommendation categories (Testing Required, Testing Recommended, and Actionable PGx). Z-scores indicate deviation from the global mean, with white representing average frequencies, red indicating elevated frequencies of pharmacogenomic relevance, and blue indicating reduced frequencies. **e)** Proportions of metabolizer phenotypes inferred from star-allele diplotypes for key pharmacogenes across the overall GI population, with colours indicating metabolizer classes. **f)** Metabolizer phenotypes of six genes from panel (e) resolved across seven ethnolinguistic groups, highlighting group-specific implications for dose adjustment or alternative therapeutic selection.

### Spectrum and Clinical Relevance of Deleterious Variants

We identified 285 pDMMs and 483 HC-LoFs classified as pathogenic (P) or pathogenic/likely pathogenic (P/LP) (ClinVar ≥2-star). Long-term endogamy and isolation in South Asian populations can elevate globally rare deleterious variants to high local frequencies, reshaping disease risk landscapes that are invisible in global reference datasets. Illustrating this effect, we identified a splice-site loss-of-function (LoF) variant in *HGD* (rs2472804506) that is absent from gnomAD, 1000 Genomes, and GenomeAsia, rising to an allele frequency of 12.5% in a single Dravidian tribal population (DR_WGH_1_01). The most frequent pDMM, *ABCA4* rs1800553 (macular degeneration), with 225 heterozygous carriers and 2 homozygous individuals reached 5.6% in IE_WPL_2_02, while the most frequent HC-LoF, *GJB2* rs104894396 (hearing loss), occurred in 131 heterozygotes across 46 populations (∼0.6%; Table S10.2). We also noted population-specific drifting of few deleterious variants to high frequencies in genes like *ABCA4* (∼6%) and *CNGA3* (∼7% for rs141386891) in IE_WPL_2_02; *SLC22A5* rs267607052 (∼5%) in DR_NGH_1_01; *CFTR* rs397508276 (∼3%) in IE_NCH_2_01/IE_NRP_2_03.

Extending this analysis to medically actionable American College of Medical Genetics and Genomics (ACMG) “Secondary Findings” gene panel ^35^, we detected 44 P and P/LP variants (23 LoFs and 21 pDMMs) in 15 genes, largely involved in metabolic dysfunction, cardiovascular traits and lipid biology (Fig 4b). Several of these variants were specific to tribal populations, including a *TTN* LoF variant (primary dilated cardiomyopathy) in DR_T, and four *BRCA2* LoF P/LP variants in AA_T and IE_T (Tables S11.2, 11.3). We see a high burden of these variants (Supplementary figure S11.3) in tribal populations: >1% of individuals from AA_T, IE_T, and TB populations carry at least one ClinVar pathogenic allele of *ATP7B, LDLR*, or *BRCA2*.

Certain clinically important genes are underrepresented in standard deleteriousness frameworks due to unique evolutionary dynamics. *HBB*, a major contributor to haemoglobinopathies in India, exemplifies this: several pathogenic variants fall outside LoF or pDMM categories yet occur at high population-specific frequencies. The β-thalassemia splice variant c.92+5G>C exceeded 1% in 20 populations, while the sickle-cell variant rs334 reached high frequencies (>5%) in several tribal populations. These patterns likely reflect regional (malaria-driven) selection alongside endogamy and drift, rather than reduced pathogenicity. Consistent with this hypothesis, we observe multiple clinically pathogenic missense variants in non-trivial frequencies (0.01 - 1.6%; Table S5.2.1) at the *G6PD* locus on chromosome X. Several high frequency (>5%) pathogenic variations were observed in tribal populations, e.g. *SRD5A2* (disorder of sexual development) in DR_EPL_1_03, *CCDC103* (Primary Ciliary Dyskinesia) in DR_EPL_1_03, and *HPDL* (Spastic paraplegia) in TB_WHR_1_01 (full list in Table S8.4). Nearly all tribal populations (27 out of 29) harbor at least one pathogenic missense or high-confidence loss-of-function variant that reaches a frequency ≥1% within that population, compared with 36 out of 53 non-tribal populations. Despite similar numbers of pathogenic variants in tribal (120 variants across 401 individuals) and non-tribal groups (114 variants across 406 individuals), the overall proportion of individuals carrying such variants is higher in tribal populations than in non-tribal populations (17.5% vs 5.5%). The divergence of allele frequencies in populations underscores the necessity of diverse and inclusive variant catalogs for accurate assessment of disease risk.

### Repertoire of Homozygous LoF Variants and Recessive Disease Risk

Across 9,768 individuals, we detected 1,214 homozygous HC-LoF variants in 965 genes (S12, Table S12.1). Tribal populations exhibited higher burdens (one-sided Kolmogorov–Smirnov test, p < 0.0001; S12). We identified 164 population-specific and 32 novel variants (absent in dbSNP, gnomAD, 1000G, GenomeAsia), with novel variants enriched in tribal groups (DR_T; Fig 4c). Notably, a novel *APOBR* frameshift (chr16:28497670-C>CT) exclusive to DR_NGH_1_01 (∼15% frequency), was carried by three homozygous individuals exhibited to moderate-to-severe hypertriglyceridemia, indicating potential impact on lipid metabolism.

Cross-referencing mouse embryonic-lethal genes identified homozygous LoFs in >50 essential genes (Supplementary S12.1), including 23 being reported in humans for the first time ^8,36–38^. Furthermore, we identified 192 previously unreported homozygous LoF variants, expanding the catalog of human knockouts in South Asia. The most frequent, *MPZL2* rs200462584-G>A (N = 8), associated with hereditary deafness, homozygous were confined to tribal populations, reaching 7% frequency in AA_T, supporting targeted follow-up with clinical phenotyping.

Genes with multiple homozygous LoFs include *LPA* (16 homozygous, 580 heterozygous; Table S12.1), where variants rs143431368 and rs41272114 are linked to reduced Lp(a) levels and cardiovascular risk ^39–41^. We also identified LoF variants in *CD36* (5 LoFs in 6 homozygous, 226 heterozygous individuals), a glycoprotein mediating amyloid-β clearance and neurodegeneration ^42^. These results highlight the value of South Asian cohorts in characterizing gene resilience and disease mechanisms.

### Population-Specific Pharmacogenomic Profiles Enable Precision Drug Dosing

Our analysis reveals extensive pharmacogenomic (PGx) diversity, capturing 2339 of 4245 cataloged variants across 82 populations and 31 of 34 Very Important Pharmacogenes. Individuals carry a median of four actionable PGx variants (max: 9), notably affecting oncology, cardiovascular, and psychiatric therapies. Allele frequencies for critical markers like *VKORC1* (rs9923231) and *NUDT15* (rs116855232) differed significantly from global populations as well as across GI populations, with 41% frequency of VKORC1 allele in Tibeto-burman populations and *NUDT15* risk alleles highly prevalent in Austro-Asiatic groups (up to 20.8%). Additionally, *DPYD* (rs56038477, rs3918290) variants affecting fluoropyrimidine toxicity were more frequent in some of the DR and IE populations than global South Asians, offering vital insights for chemotherapy dosing. Cytochrome P450 (CYP) analysis identified population-specific metabolizer statuses; Approximately 45% of individuals, especially TB_NT, were poor *CYP3A5* metabolizers affecting tacrolimus^43^ and 15% were poor *CYP2C19* metabolizers (clopidogrel) ^44^, necessitating dose adjustment crucial for metabolizing antidepressants (Hicks et al. 2017) and opioids (Crews et al. 2014). We identified five exclusive star alleles and high enrichment of the poor metabolizer allele *CYP2D6*4* (up to 21.8%) in tribal groups, while the ultrarapid metabolizer allele *2x2 reached 5.6% in one population, posing significant risks for adverse drug reactions. Furthermore, the frequency of indeterminate metabolizer status reached 18%, significantly higher than previously reported ^47^ and highlighting a critical functional annotation gap. The unique PGx profiles we report emphasize the need for population-specific guidelines to optimize drug efficacy and safety.

Our balanced sampling strategy provided an improved resolution of population-specific clinically relevant PGx variants. For instance, while previously associated with one particular Indian ethnic community ^10,48^, the *BCHE* pathogenic variant rs104893684, was identified in 29 GI populations (Table S13.8). This variant, causally linked to anaesthesia-related complications, reached allele frequencies >1% in three populations (Table S13.8). Similarly, the deployment of GLP-1 receptor agonists in the Indian context will require PGx evaluation, given the prevalence of the missense variant rs6923761 in the *GLP1R* gene (Table S13.8).

### Addressing Eurocentric Bias

#### Transferability of Genetic Risk Prediction

To assess the transferability of Eurocentric associations, we compared the risk allele frequencies from the EBI-GWAS catalog between GI and gnomAD-European populations. Tribal groups (AA_T, DR_T) and the TB populations exhibit the largest divergence from gnomAD-EUR, whereas non-tribal Indo-European and Dravidian groups show more modest shifts (Fig. 5a). These patterns suggest that Eurocentric GWAS signals offer only partial insight into the disease risk estimates in these populations. Significant allele frequency differences limit the transferability of European-derived risk estimates, as discordant variant prevalence alters both effect contributions and statistical power.

**Figure 5:**
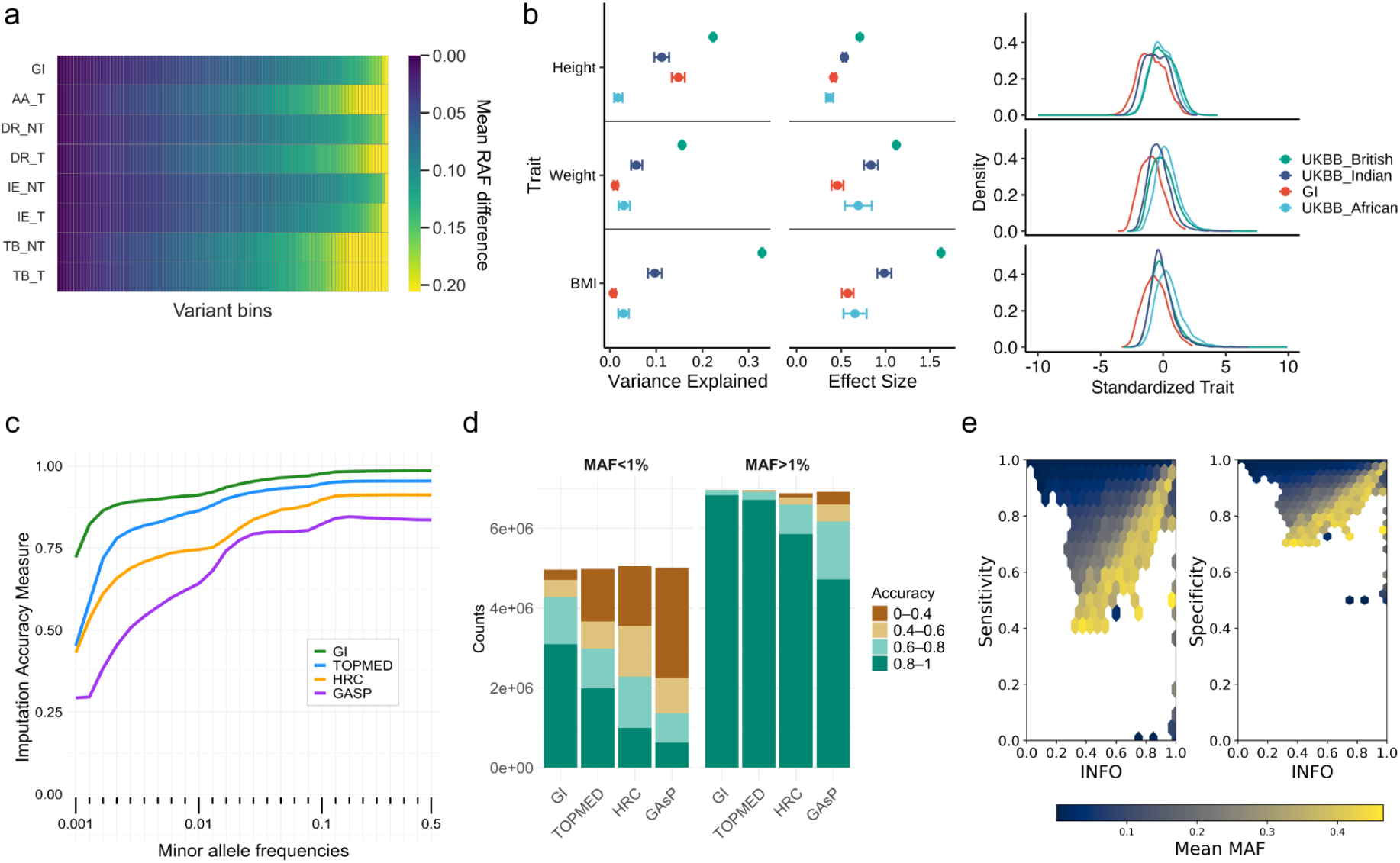
Addressing Eurocentric bias. **a)** A heatmap depicting the mean difference in risk allele frequencies (RAF) from the EBI-GWAS catalog in various GI groups. Variants are divided into percentiles based on their RAF differences, with the left-most to the right-most bins having the least difference to the most. **b)** Non-transferability of polygenic scores (PGS) to Indian and African populations, as assessed on British, Indian, and African samples from the UK Biobank (UKBB), as well as GI samples. Left and Middle: Forest plots showing the variance explained (R^2^) and effect size, respectively, for three traits - Height, Weight, and BMI. The corresponding density plots on the right show the standardized distributions for each trait. **c)** Performance of the GI imputation panel compared to three other reference panels - TOPMed, the Haplotype Reference Consortium (HRC), and the Genome Asia panel (GAsP), as measured on the array data of 7,628 South Asian ancestry individuals from the UK Biobank. Only those loci that were imputed by all the panels were included in the analysis. **d)** Allele frequency–stratified distribution of overlapping imputed variants and their corresponding imputation accuracy across different reference panels **e)** The sensitivity and specificity of imputation using the GI imputation panel, measured by comparison with whole genome sequencing data as ground truth.

We calculated European-derived polygenic scores (PGS) for height, weight, and BMI in the GenomeIndia cohort, benchmarking them against those of British, African, and Indian individuals from the UK Biobank ^49^. Consistent with the Eurocentric skew of current GWAS resources ^50^, predictive accuracy (R²) was highest in British individuals and declined with increasing genetic distance from European discovery populations, reaching its lowest values in Africans (Table S15.1, Fig. 5b). Height PGS showed comparable predictivity between UKBB Indians (R^2^ = 0.112) and GenomeIndia (R^2^ = 0.148), whereas weight and BMI PGS were substantially less predictive (GI BMI R^2^ = 0.007 vs. UKBB = 0.097). This discrepancy likely reflects the combined influence of genetic divergence and environmental heterogeneity, underscoring the need for large, independent studies in South Asia to ensure equitable polygenic risk prediction.

### The GI Imputation Panel for South Asian Ancestry

We created the GI imputation panel using phased haplotypes of 9768 Indian individuals harbouring variants with minor allele count (MAC) ≥3. The GI panel showed minimal LD divergence from 1000 Genomes South Asian SAS (mean varLD = 1.81), substantially lower than European, East Asian, or African groups (mean varLD of 6.23, 8.37, 17.29 respectively), validating its suitability for South Asian ancestry reference (Fig. S15.1a, b).

We obtained ∼400,000 array genotypes from UK Biobank ancestry-matched South Asian (UKB-SAS) individuals (N = 7,628) (Fig S15.3) and performed genotype imputation using a GI reference panel, generating approximately 50 million imputed variants with high accuracy across frequency spectra (Fig. S15.5). Comparative analyses on the shared set of variants imputed by all panels showed the GI panel outperformed widely-used reference panels GAsP ^8^, HRC ^51^, TOPMed ^52^ with 45%, 20%, and 9% average improvements in imputation quality, respectively (Fig. 5c,d). Superior performance for rare variants demonstrated that the GI panel effectively captured population-specific LD structure and rare allele frequencies, an essential characteristic of a high-quality imputation panel (Fig. 5c, S15.6). Improved performance was observed across the genome, including loci such as chr16p11.2–q11.2, a challenging centromeric region implicated in multiple diseases. These results support the panel’s robustness for large-scale population studies, with possible relevance to precision medicine applications (Fig S15.7).

Further validation using 7,628 UKB-SAS WGS-called genotypes as ground truth showed >0.99 mean pairwise Identity-By-State concordance (Fig. S15.8), 99.75% sensitivity, 99.88% specificity, and 99.75% precision. More than 99.99% of variants achieved sensitivity and specificity >0.80, with both rare and common variants imputed accurately (Fig. 5e). Only 262 variants (16 with MAF <1%) showed marginally reduced performance (sensitivity/precision <50%; specificity <60%). The performance of the GI reference panel, comparable to, or exceeding that of established, widely-used reference panels, positions it as a reliable resource for large-scale research involving diverse South Asian populations.

## Discussion

The rich and profound diversity of the Indian subcontinent offers insights into human variation that is inaccessible to studies involving more homogenous global populations. By combining large non-tribal populations alongside sampling of small tribal and isolated groups, the GenomeIndia project has captured a spectrum of human genetic diversity that is shaped by complex, sometimes sex-biased migration histories, ethnolinguistic affiliation, biogeography, and sociocultural practices. Our findings demonstrate recent endogamy as a dominant force, resulting in populations that are distinct from each other yet internally homogeneous.

Our nuanced sampling design revealed sharp demographic contrasts. Large non-tribal groups showed sustained population growth, whereas many tribal populations represent pronounced drift and strong founder effect. These groups, as well as some large populations, show extreme burdens of runs of homozygosity, often surpassing those of well-studied global isolated populations. These complementary structures in Indian populations would enable future research for a wide range of human-genetic studies, from well-powered genome-wide association analyses to rare-variant discovery, gene-function inference, and studies of human knockouts and inbreeding-related disease risk.

A key clinical consequence of the Indian genetic architecture is the enrichment of globally rare yet locally common variants, many absent from current reference datasets. A combination of extreme founder effects and genetic drift has allowed deleterious (LoF) and pathogenic variants to reach high local frequencies. When combined with careful phenotyping and longitudinal follow-up, such variants offer powerful opportunities for gene discovery, and for developing population-tailored pharmacogenetic and public-health strategies. The skew we observed in the frequency distribution of pathogenic variants suggests that the genetic disease burden in India is likely to be population specific rather than pan-Indian. While we do observe high frequencies of alleles associated with hemoglobinopathies and malarial resistance in some populations, the overall distribution of the putative pathogenic variants is functionally heterogeneous. The localized enrichment of several variants predicted to be pathogenic warrant a reassessment of their penetrance and clinical relevance.

The genetic distance of Indian populations from European-ancestry cohorts that dominate current global genomics creates challenges for clinical translation. Our analyses revealed that allele frequency differences, in conjunction with linkage disequilibrium mismatch and possible altered causal variant tagging, can lead to reduced predictive accuracy of polygenic risk estimates in Indian populations. This non-transferability emphasizes the need for population-informed genomic resources to ensure equitable medical interpretation. To partially bridge this gap, we introduced the GenomeIndia imputation panel, which substantially improved genotype inference across Indian populations, and an expanded catalogue of common and rare variants, which provides a foundation for India-specific genotyping arrays. These resources would enable cost-effective, large-scale genetic studies and facilitate clinical translation in both research and healthcare settings.

While this study represents a significant milestone, our cohort of 9,768 individuals does not fully encompass the vast, diverse population of India. To achieve a truly comprehensive map of Indian genetic variation, future studies should prioritize large-scale sampling of non-tribal populations to capture their extensive rare-variant diversity, while maintaining targeted inclusion of genetically isolated tribal groups to identify population-specific variants with high functional and clinical relevance. Nevertheless, our work provides a robust and necessary foundation for such efforts.

Finally, our analyses place India at the geographic and genetic crossroads of South, Southeast, and East Asia. The implications of GenomeIndia extend far beyond national boundaries, informing studies of ancestry, disease risk, and therapeutic response not only in India and its diaspora but across a substantial fraction of the world’s population. By providing a high-resolution reference for this underrepresented region, this project serves as a cornerstone for a more inclusive and accurate understanding of global human genetic diversity.

## Methods

### Ethics and Recruitment

The GenomeIndia consortium adhered to the principles of the Helsinki Declaration for research protocols, consent forms, sample collection, and ethical practices. These guidelines were uniformly implemented across all sampling centres and approved by their respective institutional human ethics committees (IECs). The study encompassed 83 anthropologically distinct populations comprising both tribal and non-tribal groups (S1). To ensure a balanced representation of the genetic diversity, we aimed to enroll an adequate number of volunteers for each population group, spanning 30 tribes and 53 non-tribal populations, each identified based on their ethnic, socio-cultural, biogeographic, and linguistic distinctions. Trios (6-12) were also collected from each population, with the exact number determined based on the population size, availability, and further validations. This systematic sampling strategy represents one of the most comprehensive genomic studies of Indian populations to date. Further,, these populations were grouped into seven predefined ethnolinguistic and social categories: Austroasiatic (AA), Indo-European tribal (IE-T) and non-tribal (IE-NT), Dravidian tribal (DR-T) and non-tribal (DR-NT), and Tibeto-Burman tribal (TB-T) and non-tribal (TB-NT). An additional group, continentally admixed outgroup (CAO), of African origin was excluded from this classification due to its distinct and relatively recent continental ancestry ^53^

### Sample Collection and Processing

Blood samples (n=20,195) were collected from healthy participants aged 18 and older, with their informed consent, along with biochemical assessments, sociodemographic, and anthropometric measurements (Table S1.2 and S1.3). DNA was isolated, and the samples were subjected to whole-genome genotyping. Of these, 13,242 samples were successfully genotyped. After genotyping, Hardy-Weinberg equilibrium (HWE) was assessed within each specific population. Autosomal variants in HWE and have a minor allele frequency (MAF) greater than 1% were used to estimate Identity by Descent (IBD) within populations using PLINK v1.9 (--genome command) ^54^. Individuals showing relatedness beyond the first-cousin level (PI_HAT >= 0.125) were excluded, and only one individual was retained from each related pair. In addition, trio relationships were inferred using the genotype data (Figure S2.1).

### Data Generation and Performance Benchmarking

PCR-free whole-genome sequencing (WGS) was performed for 10,074 samples. Sequencing was primarily performed at the four sequencing centres (CBR, CSIR-CCMB, CSIR-IGIB, and BRIC-NIBMG). Paired-end sequencing libraries were prepared following the standard protocols for the NovaSeq 6000 platform.

For benchmarking, cell lines corresponding to four samples from the Genome in a Bottle (GIAB) dataset were procured by CBR and sequenced at all centres. The average sequencing depths achieved were 49X for GM12878, 43X for GM24149, 48X for GM24143, and 41X for GM24385. Variant identification was performed using the DRAGEN germline variant caller pipeline. Haplotype-level comparisons, conducted with the hap.py tool ^55^, showed strong concordance with the GIAB truth set. The analysis yielded a recall of ≥0.97, a precision of 0.99, and an F1 score of 0.98 for both SNVs and INDELs. We further employed a cyclical sequencing concordance validation method, using two unrelated and one set of trio samples, randomly selected and sequenced at two centres ^11^ (Figure S2.2). Cross-centre comparisons showed an average recall of 0.99, precision of 0.97, and an F1 score of 0.98, indicating minimal batch effects. The consistent results across centres highlight the reliability and reproducibility of the project outcomes.

### Quality Filtering and Dataset Generation

The FASTQ files were processed using the germline variant caller in DRAGEN v4.0.3 software ^56^, which produced genomic Variant Call Format (gVCF) files for each sample. To ensure accurate and reliable variant detection, the dataset maintained an average sequencing coverage of at least 23X for each sample. Furthermore, each sample was evaluated for the standard quality filters of percentage callability, Het/Hom ratios, and Ti/Tv ratios. Samples that did not meet the minimum coverage and filtering criteria were excluded from the final dataset, resulting in a total of 9,768 samples (S2). The gVCF files of these 9,768 samples underwent joint-genotyping using the Illumina gVCF genotyper, resulting in a single Variant Call Format (VCF) file. Variant filtering retained only those with a ‘PASS’ in the Filter field, QUAL >30, and GQ >20. Doubletons required at least one sample with GQ >40, and singletons required GQ >40. Variants with a significant deviation from the Hardy-Weinberg Equilibrium (HWE) were also filtered out (p value ≤ 10^-11^).

### Variant Classification and Unreported Variant Discovery in GenomeIndia

The final high-confidence variant set was categorized into four groups based on minor allele frequency (MAF) within the GI dataset: ultra-rare (< 0.1%), rare (0.1 - 1%), low-frequency (1 - 5%), and common (≥ 5%) variants. Further, known and unreported genetic variants in the GenomeIndia (GI) dataset were identified through comparison with four major global databases: dbSNP (v156) ^57^, gnomAD (v4, genome and exome) ^37^, 1000 Genomes Phase 3 ^58^, and GenomeAsia (GAsP) ^8^. Variants were matched using the "CHR:POS:REF:ALT" format to determine known entries and classify variant types. The details of variant sharing analysis and identification of population-specific variants are provided in supplementary section S3.

### Analysis of Population Structure

#### F_ST_ Analysis

Pairwise genetic distances were estimated using the F_ST_ statistic ^59^, computed with EIGENSOFT (precision: six decimal places). A heatmap and dendrogram were constructed from the resulting F_ST_ matrix. To examine genetic differentiation among linguistic groups, - populations were grouped by linguistic affiliation and Tribe/Non-Tribe status, and group-wise F_ST_ values were calculated.

#### Jaccard Index

We quantified genetic similarity between populations using the Jaccard index, defined as the ratio of the number of shared variants to the total number of variants across two clusters. For each pairwise comparison, we controlled for unequal sample sizes of the clusters by randomly downsampling the larger cluster to the size of the smaller cluster. This ensured that both clusters contributed the same number of individuals to the analysis, thereby avoiding bias in variant counts. All detected variants within the downsampled sets were then included in the calculation. We randomly downsampled the larger cluster to match the sample size of the smaller cluster.

#### Novel Variant Discovery Rate

We quantified the rate of novel variant discovery using a rarefaction-based approach. For each cluster, individuals were randomly permuted, and variants contributed by the i^th^ sample were counted only if they had not been observed in the preceding (i−1) samples. To ensure comparability across clusters, rarefaction curves were generated using an equal maximum sample size of ∼500 individuals per group. Fifty bootstrap resamples of the cumulative discovery process were performed, with each bootstrap replicate consisting of 500 individuals resampled with replacement. The resulting curves represent the expected number of unique novel variants that are discovered as the sampling depth increases.

#### Principal Component Analysis

Principal Component Analysis (PCA) was performed using smartpca (version 18140) from the EIGENSOFT (v8.0.0) ^60,61^ suite to assess overall population structure in the GI dataset. To contextualize these patterns with neighboring populations from Central South, Northeast, East, and Southeast Asia globally, PCA was also conducted on a merged dataset comprising GI, Human Genome Diversity Project (HGDP), and GAsP samples.

#### ADMIXTURE Analysis

We performed unsupervised clustering using ADMIXTURE v1.3.0 ^15^ on 82 ethnic groups. One population (CAO), identified as continentally admixed, was analyzed separately. To reduce relatedness bias, children from trio samples were also removed. Linkage disequilibrium (LD) pruning was conducted using an r² threshold of 0.1 to ensure marker independence, resulting in the retention of 970,954 variants for analysis. ADMIXTURE was run with K values ranging from 2 to 14.

Based on the admixture proportion at K = 5, for each population, we estimated the within-individual diversity for each population using the Shannon index and the between-individual diversity using the Bray-Curtis distance, which measures the differences in admixture proportions for each pair of individuals within the population. Then, for each population, we took the median of within-individual and between-individual measures to estimate the average within-individual and between-individual diversity for the population.

#### Mitochondrial Analysis

The joint-called VCF from DRAGEN contained 7,912 mitochondrial variants, all of which passed DRAGEN filters (‘PASS’). We performed a series list of filtering steps on the joint-called VCF file, as detailed in Laricchia et al. 2022. We calculated the heteroplasmic fraction (HF) for each variant for each sample from the ‘Allelic Depth’ field of the VCF. HF was defined as a ratio of the number of reads supporting the variant/total number of reads at that position. We only included variants with a heteroplasmic fraction ≥ 0.10 to avoid NUMT-derived false positive variants. Variants with HF in the range of 0.90 - 1.00 were denoted as homoplasmic, and variants with VAF < 0.90 (and ≥ 0.10) were denoted as heteroplasmic.

We filtered out a predefined list of artifact-prone sites (positions 301, 302, 310, 316, 3107, 16182) since these sites have sequence contexts that make it difficult to distinguish actual variants from technical artifacts. Additionally, we applied the filter “indel_stack” to remove indels that are only present within multi-allelic calls across all samples in the set of variants. Genotypes not marked ‘PASS’ (base_quality, low_af, low_frac_info_reads, weak_evidence, mapping_quality, read_position, no_reliable_supporting_read, and too_few_supporting_reads) were marked as missing. After filtering, we annotated the variants using the Ensembl Variant Predictor tool ^63^. We annotated our variants using publicly available data files from MITOMAP ^64^ (downloaded on date 14 October 2024) to infer the clinical significance of the filtered mtDNA variants in our cohort. We used 126 MITOMAP variants with a ‘Cfrm’ (confirmed) status, being reported by at least two or more laboratories examining unrelated families. Additionally, we also report the variants which have consequences marked as ‘start lost’, ‘stop gained’ and ‘stop lost’, along with their SIFT, PolyPhen, and APOGEE2 scores, wherever applicable.

#### Mitochondrial Haplogroup Estimation

We started with all 7,912 PASS variants in the joint called VCF. We used Haplogrep 3 with Phylotree Build 17 as the reference phylogeny. To minimize the inclusion of low-confidence heteroplasmic sites, we applied a stringent heteroplasmy frequency (HF) threshold of 0.9 using Haplogrep’s *--hetLevel* parameter. We retained variants with a heteroplasmy level of ≥90% for haplogroup classification; all other heteroplasmic sites were excluded from downstream analysis. This approach ensured that only high-confidence variants, either homoplasmic or near-homoplasmic (HF ≥ 0.9), contributed to haplogroup assignment. The output included per-sample haplogroup predictions, quality scores, and lists of defining mutations based on the latest mitochondrial phylogeny.

#### ChrX Variant Filtering and Analysis

Variant-level quality filters for chrX were applied consistently with those used for autosomes, requiring a PASS status in the Filter field, a QUAL >30, and a GQ >20. However, applying the Hardy-Weinberg equilibrium (HWE) filter to chromosome X posed challenges due to male individuals possessing only a single X chromosome, rendering biallelic sites incompatible with standard HWE assumptions. To address this, HWE p-values were recalculated exclusively for female samples. Variants significantly deviating from equilibrium (p ≤ 10⁻¹¹) were then excluded from both female and male datasets, resulting in 6,059,950 biallelic variants on chrX. A Mendelian concordance rate of 99.9% was achieved, validating the accuracy of the filtering strategy prior to downstream analyses. Further, to identify unreported genetic variation on chromosome X in the GI dataset, we compared detected variants against four major public repositories - dbSNP, gnomAD, 1000 Genomes Project Phase 3, and GenomeAsia100K, following the same approach used for autosomes.

#### ChrY Analysis

Variant filtering on chromosome Y was carried out following the same strategy as for the autosomes, except that the HWE filter was not applied due to the hemizygous nature of the chromosome. This gave us a final variant set of 230,879 in chrY. Subsequent analyses were conducted exclusively on male samples from the GI dataset. To identify the unreported genetic variants across chrY, we compared the sites, along with the reference and the alt alleles, with the 1000Genome phase3, gnomAD, and dbSNP datasets, resulting in 132,097 novel variants. We assigned Y-chromosomal haplogroups to all male individuals in the GI dataset using Y-LineageTracker v.1.3.0 (Chen et al. 2021). These Y-chromosomal haplogroup assignments were integrated with autosomal PCA coordinates to investigate patterns of genetic differentiation among the various linguistic groups. Additionally, their distribution was assessed across different geographical regions to explore region-specific haplogroup patterns.

#### Demography and Effective Population Size

We used SMC++ (v1.15.4) ^66^ with default parameters and estimated the Ne for each population. The SMC++ first converts the population VCF to the smc format; here, we mask (--mask) the GRCh38 Gap tracks downloaded from the UCSC browser. We then run the estimate command to have the fitted model for each population, giving the Ne estimates over time. We used the mutation rate of 1.25e-8 per base per generation, specified the starting and ending timepoints (in generations) 100, 100000 of the model (--timepoints), and used a piecewise spline (--spline) with 12 knots (--knots), which controls the functional form used to fit the model. Using the fitted Ne trajectories obtained for each population, we generated the plots using the ggplot2 package and performed clustering using the latrend package (S6).

#### Runs of Homozygosity

Runs of homozygosity (ROH) were analyzed to assess genomic autozygosity and patterns of recent shared ancestry across populations. To ensure that only unrelated individuals were included, we excluded those with a PI_HAT ≥ 0.125, corresponding to first cousins or closer relatives. After removing the additional removal of population outliers and trio offspring, a total of 9,442 unrelated individuals were retained for downstream ROH analysis. As global references, data from the Finnish (FIN) population of the 1000 Genomes Project and the Ashkenazi Jewish (AJ) population from the TAGC dataset were included.

ROHs were identified using PLINK v1.9, excluding singleton variants and applying default parameters, except the following: --homozyg --homozyg-kb 500 --homozyg-window-snp 100 --homozyg-window-het 2 --homozyg-window-missing 20. To incorporate information on recombination, physical ROH lengths were converted into genetic distances (in centimorgans, cM) in PLINK, using a genetic map from https://github.com/odelaneau/shapeit5/tree/main/resources/maps/b38. Only ROHs with a minimum physical length of 500 Kb and a minimum genetic length of 1 cM were retained. Segments were classified into three length-based categories: 1-10 cM, 10-15 cM, and >15 cM.

#### Variant Annotation and Functional Consequences

The high-confidence variants from 9,768 samples were annotated using the ENSEMBL Variant Effect Predictor (VEP v113) ^63^. Functional consequences were assessed across minor allele count (MAC) categories - singletons, doubletons, and MAC ≥ 3. For multi-annotated variants, the principal transcript was selected using VEP’s --flag_pick option, with PICK=1. Loss-of-function (LoF) variants were further annotated using the LOFTEE plugin, pLI scores, and LOEUF plugin in VEP ^63^. To evaluate the potential impact of missense variants, we applied two predictive scoring systems: REVEL (Rare Exome Variant Ensemble Learner) ^32^ and CADD (Combined Annotation Dependent Depletion) ^33^. Missense variants with REVEL scores ≥0.644 and CADD scores ≥20 were considered deleterious (pDMM) ^32,33,67^. Clinical relevance of pDMMs and LoFs was assessed using the ClinVar database (variant_summary.txt file downloaded from the NCBI FTP repository https://ftp.ncbi.nlm.nih.gov/pub/clinvar/tab_delimited/ in February 2025). A variant was classified as clinically relevant if it was annotated as pathogenic (P) or pathogenic/likely pathogenic (P/LP) in ClinVar with a review status of ≥2 stars.

#### Homozygous Loss-of-Function Variants

For this analysis, high-quality, homozygous loss-of-function alleles present in at least one individual were considered. Variants were annotated using VEP, focusing on those with a strong predicted impact including, frameshifts, essential splice site changes, and premature stop codons (IMPACT=HIGH) and filtered using LOFTEE for high confidence (LoF=HC). For functional and clinically relevant variations, we checked for overlap with the EBI-GWAS catalog, ClinVar, and IMPC (https://www.mousephenotype.org/).

#### Pharmacogenomics

The VCF files for all 22 chromosomes were first normalized using vt ^68^ and then annotated with rsIDs from dbSNP v156 using SnpSift v5.2 ^69^ to ensure accurate retrieval of SNPs and indels. Pharmacogenomic variants were extracted from these VCFs using bcftools v1.9 ^70^, followed by conversion to PLINK format using plink v1.9^54^. Population-specific allele frequencies were computed, and Fisher’s exact tests were performed to compare allele frequency distributions across populations.

Star allele haplotypes for 58 pharmacogenomic genes were inferred using Stargazer ^71^, which also provided predicted metabolizer phenotypes based on gene activity scores. Due to their structural complexity, CYP2D6 and HLA haplotypes were called separately using Cyrius (Chen et al. 2021) and xHLA ^73^, respectively, from .bam files. Variants were additionally classified according to testing level categories, integrating evidence curated from PharmGKB and CPIC.

#### Haplotype Phasing and Genotype Imputation

Haplotype phasing was performed using SHAPEIT5 on the joint-called VCF. Phasing accuracy was evaluated by calculating switch error rates using trio data. To demonstrate imputation performance, we used array-based genotype data from 7,677 individuals of South Asian Ancestry enrolled in the UK Biobank cohort, which had been phased prior to imputation. Genotype imputation was performed using IMPUTE5, with the phased GenomeIndia haplotypes serving as the reference panel and employing a 5 Mbp overlapping window strategy.

We benchmarked the GenomeIndia reference panel against three widely used reference panels - HRC, TOPMed, and GenomeAsia - available via the Michigan Imputation Server. Individual-level imputation accuracy was assessed by comparing imputed genotypes with WGS-based genotypes from a truth set of 7628 individuals. Additionally, we examined population-level genetic differentiation between 1000 Genomes SAS sub-populations and UK Biobank (UKB) individuals of Indian ancestry from the GI reference panel to assess population-specific imputation gains. For this analysis, we downsampled the 1000 Genomes SAS WGS data to match the UKB array variant set. Both datasets were then imputed using the GI panel. Finally, we stratified variants by F_ST_ deciles and compared imputation INFO scores across these deciles.

#### Frequency Differences of Established GWAS Loci

To investigate complex trait-associated risk alleles in the GenomeIndia (GI) cohort, we retrieved European-ancestry variants from the GWAS Catalog (accessed on November 29, 2024) and intersected them with the variant set from GI data, resulting in approximately 228,000 overlapped variants. We evaluated allele frequency divergence between the Eurocentric reference and seven GI clusters using Fisher’s exact tests with multiple testing corrections, resulting in approximately 89,000 markers which are significant in at least 1 GI cluster. Statistically significant markers were categorized by their distribution across the Indian population: cluster-specific (unique to one group), partially shared, or universally shared across all seven clusters. We explicitly assessed the transferability of these Eurocentric associations by comparing risk allele frequencies between GI population clusters and gnomAD-European populations.

#### Calculation of Polygenic Scores (PGS)

We assessed the cross-population portability of European-derived polygenic scores (PGS) for height, weight, and body mass index (BMI) in the GI cohort and benchmarked their performance against British, African, and Indian ancestries in the UKBB (Bycroft et al. 2018). We utilized published PGS from the PGS Catalog ^74^ for height (PGS002804), weight (PGS004373), and BMI (PGS002313), and used the PGS_calc software for generating the PGS for the populations (S14).

## Supporting information

GenomeIndia Consortia Author List

## Data Availability

The GenomeIndia Allele Frequency data is publicly available via IBDC (https://ibdc.dbtindia.gov.in/genomeindia/downloadfile?path=9768GI_SummaryStats.tar.gz). Due to privacy laws related to participant consent for data sharing, raw sequencing data are available under controlled access via IBDC at https://ibdc.dbtindia.gov.in/data_access/. Bona fide researchers are required to submit a data access request outlining the proposed research to the Data Management Group, which will be subject to approval.

## Code Availability

Codes generated and utilized for analysis in this work are available in https://github.com/genomeindiaconsortium/Flagship-Paper-Codes.

## Acknowledgements

We thank all participants who consented to providing their samples for this project. We acknowledge the funding by the Department of Biotechnology (DBT), Ministry of Science and Technology, Government of India (BT/GenomeIndia/2018). We would like to thank Dr Rajesh S Gokhale (Secretary, DBT), Dr Suchita Ninawe (Senior Adviser, DBT), and Dr Renu Swarup (Former Secretary, DBT) for their support, and the DBT officers who have administered the GenomeIndia project : Drs Richi Mahajan, Kakali Dey Dasgupta, Onkar Tiwari, and Amit Tripathi. The authors acknowledge the guidance and technical oversight provided throughout the study by the members of the Technical Monitoring and Assessment Committee (TMAC). We acknowledge the current and past leadership of all participating institutes for their support, which was instrumental in the smooth execution of the GenomeIndia Project. The authors thank the support provided by the computational facilities of the institutes. We acknowledge the use of generative AI and large language models in refining the language of the manuscript; however, the responsibility of the final content lies with the authors. We thank the Illumina joint genotyper team for help with the early access of the joint genotyper program. Shreya Chakraborty and Sauma Suvra Majumdar are supported by the Prime Minister’s Research Fellowship. A detailed list of individuals who helped us with sample collection and other logistical operations of the projects are acknowledged in Supplementary Section S1.

