## Supplementary Material for "An Atlas of Indian Genetic Diversity"

### An Atlas of Indian Genetic Diversity
### Supplementary Information
### Table of Contents

| Title | Page |
| --- | --- |
| Section S1: Study design and description of populations | 02 |
| Section S2: Whole genome sequencing and quality control | 16 |
| Section S3: Allele Frequency Spectrum of GenomeIndia dataset | 25 |
| Section S4: Population Structure | 52 |
| Section S5: Analysis of mitochondrial DNA, X and Y chromosomes | 81 |
| Section S6: Demography and Effective Population Size (Ne) | 111 |
| Section S7: Runs of Homozygosity (ROH) analysis | 122 |
| Section S8: Annotation of variants | 129 |
| Section S9: Putative Deleterious Missense Mutations (pDMMs) | 141 |
| Section S10: High Confidence Loss-of-Function variants (HC-LoFs) | 146 |
| Section S11: Clinically actionable variants as per ACMG guidelines | 161 |
| Section S12: Homozygous Loss-of-Function variants | 169 |
| Section S13: Pharmacogenomic variants | 178 |
| Section S14: Eurocentricity of Polygenic Scores | 191 |
| Section S15: The GI Imputation Panel | 200 |

#### Supplementary section S1: Study design and description of populations

Chandrika Bhattacharyya[1], Shouvanik Sengupta[1,2], Prathima Arvind[3], Shweta Ramdas[3], Bratati Kahali[3], Kumarasamy Thangaraj[4], Analabha Basu[1,2]

[1]BRIC - National Institute of Biomedical Genomics (BRIC-NIBMG), Kolkata, India. [2]Regional Centre for Biotechnology (RCB), Faridabad, India. [3]Centre for Brain Research (CBR), IISc Campus, Bengaluru, India. [4]CSIR - Centre for Cellular and Molecular Biology (CSIR-CCMB), Hyderabad, India

#### Summary

##### Study design

The GenomeIndia project was designed systematically to capture the genetic diversity of the Indian subcontinent. The study adopted a stratified sampling approach, selecting populations to represent the major ethnolinguistic lineages, social groups, and geographic regions across India (Bhattacharyya et al., 2025). An emphasis was placed on including tribal as well as non-tribal communities from across the country, ensuring representation of historically endogamous, regionally isolated, and culturally distinct groups. The final list of 83 populations included 53 non-tribal and 30 tribal groups spread across India (see Table S1.4 for details).

Samples were collected by 13 partnering institutes (Table S1.1) through direct community engagement at field sites with active engagement of social scientists, anthropologists, local people, and community stakeholders. Biological sample collection (blood for DNA as well as basic phenotypic information) was also accompanied by the standardized collection of anthropometric and demographic information for every individual. This framework enabled a high-resolution insight into India's fine-scale population structure, demographic history, and medically relevant variation. A broad overview of the sampling and sequencing pipeline followed by the consortium is shown in Figure S1.1.

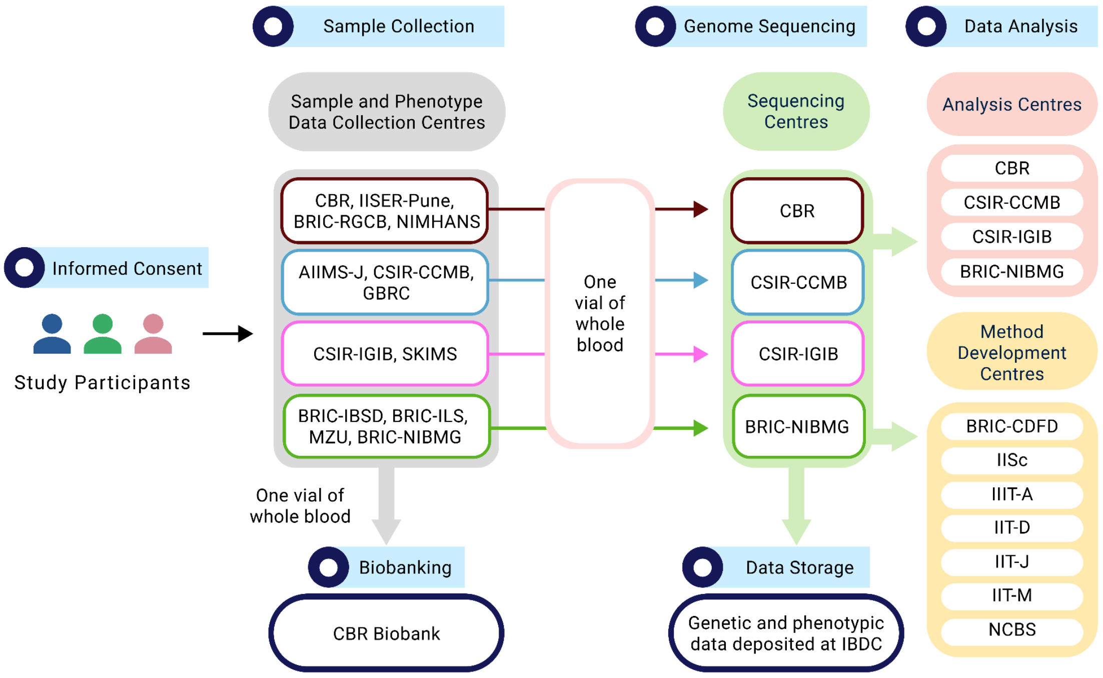

**Figure S1.1**. Overall schematic of the sample and data flow, deposition, and sharing across the GenomeIndia participating centres. See Table S1.1 for expansion of institute names.

*Sample size scheme*

Sample sizes for each population were chosen to enable robust estimates of relatively rare allele frequencies within each. Previous studies (The Indian Genome Variation Consortium, 2005) informed our definitions of large (>10 million) and small (< 1 million) population groups. To ensure that we do not miss out on the large array of rare variants in each population group, we sequenced a median of 159 unrelated individuals from each non-tribal group and 75 from each tribal group. We enrolled approximately twice the number of individuals under each population group to account for confounders such as DNA concentration and quality, to facilitate repetition of sequencing and subsequent validation in a subset of these samples, and also to ensure that there are enough unrelated individuals in each population. In total, we enrolled 20,195 self-declared healthy adults.

Our subjects included unrelated individuals of the above representative population groups across India. A small number of trios were enrolled to estimate the haplotype structure of population groups. Overall, 12 trios from large populations (>10 million) and six trios from small populations (<1 million) were collected, of which six trios from large populations and three trios from small populations were sequenced.

*Inclusion and exclusion criteria*

We included all self-declared healthy individuals aged 18 years or older in the study. Both male and female participants were recruited for the study to recruit equal numbers. We excluded individuals with self-reported history/family history of monogenic diseases, chromosomal abnormalities, bleeding or clotting disorders, and life-threatening conditions, or those who were bedridden.

**Table S1.1**. List of institutes and their responsibilities in the consortium.

| Sl. No | Name of the centres | Work Distribution |
|---|---|---|
| 1 | Centre for Brain Research, Bengaluru (CBR), CSIR-Centre for Cellular and Molecular Biology, Hyderabad (CCMB), CSIR-Institute of Genomics and Integrative Biology, New Delhi (IGIB), BRIC-National Institute for Biomedical Genomics, Kalyani (NIBMG) | Sample collection, including phenotyping, sequencing, and data processing for variant calling, and data analysis using genetic variants obtained from the project. |
| 2 | National Institute of Mental Health and Neurosciences, Bengaluru (NIMHANS), Sher-i-Kashmir Institute of Medical Sciences, Srinagar (SKIMS), All India Institute of Medical Sciences, Jodhpur (AIIMS-J), Gujarat Biotechnology Research Centre, Ahmedabad (GBRC), Indian Institute of Science Education and Research, Pune (IISER-Pune) | Sample collection, including Phenotyping |
| 3 | Mizoram University, Aizawl (MZU), Institute of Bioresources and Sustainable Development, Imphal (IBSD), Institute of Life Sciences, Bhubaneswar (ILS), Rajiv Gandhi Centre for Biotechnology, Thiruvananthapuram (RGCB) | Sample collection, including phenotyping, and data analysis using genetic variants obtained from the project |
| 4 | Indian Institute of Science, Bengaluru (IISc), Indian Institute of Information Technology, Allahabad (IIIT-A), Indian Institute of Technology Delhi, New Delhi (IIT-D) | Data analysis using ML/AI to develop novel algorithms and methods |
| 5 | National Centre for Biological Sciences, Bengaluru (NCBS), Indian Institute of Technology Madras, Chennai (IIT-M) | Data analysis using ML/AI to develop novel algorithms and methods; data analysis using genetic variants obtained from the project |

| | | |
|---|---|---|
| 6 | iBRIC - Centre for DNA Fingerprinting and Diagnostics, Hyderabad (CDFD),<br>Indian Institute of Technology, Jodhpur (IIT-J) | Data analysis using genetic variants obtained from the project |
| 7 | Centre for Brain Research, Bengaluru (CBR) | Bio-banking |
| 8 | Indian Biological Data Centre, Faridabad (IBDC) | Data storage and archival |

**Standardized procedures for participant recruitment**

*Ethics and information sheets:* All participating centres in the GenomeIndia study obtained ethical approval from their respective Institutional Ethics Committees (IECs). The 13 sample collection centres obtained informed written consent from all participants before recruitment into the study, in accordance with the ethical guidelines outlined by the Indian Council of Medical Research (ICMR). Consent forms were provided in both regional languages and English to ensure that participants clearly understood the purpose, procedures, risks, and benefits of the study in a language they were comfortable with, before agreeing to participate.

*Sample collection:* The thirteen-partnering sample collection centres across India were responsible for collecting samples from all 83 populations based on the geographical distribution of the respective populations. Individuals from small, isolated population groups were sampled from their small and respective indigenous settlements while isolated and larger populations were identified across both rural and urban settings. Outreach and community engagement programmes were conducted to highlight the importance of genetic research, explain the study's objectives, and reassure participants about data confidentiality. Wherever required, tribal populations were approached only after obtaining the necessary permissions from the respective state governments. Isolated and larger populations were engaged through local leaders, community representatives, and relevant organizations. Field visit teams from the partnering centres visited villages, organizations, and specific locations to enroll participants. Common Standard Operating Procedures (SOPs) were developed and consistently followed for the collection of biological samples and phenotypic data, including demographic and anthropometric information.

*Randomization of IDs:* All sample collection centres generated a Local ID for each participant, consisting of the centre code followed by a six-digit number. Once the sequencing centres received the blood samples, a second-level ID (random 12-character alphanumeric codes, e.g., AB12345678CD) was assigned to each sample to ensure uniqueness and anonymity. This ID was used for all downstream DNA processing and sequencing. All random IDs were generated centrally at the Centre for Brain Research (CBR), and were securely shared with the sequencing centres.

*Metadata annotation:* During the sample collection, demographic details collected for each participant included their full name, gender, place of residence, and date of birth (used to calculate age at the time of recruitment). Information on mother tongue was recorded to capture cultural and linguistic diversity. Additionally, data on whether the participant's parents belonged to the same ethnic group and whether any biological relatives were also participating in the study were recorded. Marital status, number of children, and language proficiency (both spoken and written) were also documented to provide a comprehensive socio-demographic profile.

The teams recorded educational background (highest level attained, total years of education, and medium of instruction), and socioeconomic status (current occupation and total monthly family income in rupees). Lifestyle and health-related information included the participant’s smoking and alcohol consumption habits, use of chewing tobacco, and a medical history covering major illnesses or surgeries experienced by the participant and their immediate family members (father, mother, and others). Additionally, participants were asked about current medication use, with details on the names of any medications being taken at the time of data collection.

*Phenotypic measurements and a list of phenotypes collected:* A comprehensive list of recorded parameters is listed in Tables S1.2 and S1.3; more details on the underlying phenotypic data are in an accompanying manuscript. Figure S1.2 shows the age and sex distribution of the sequenced samples.

**Table S1.2**. List of mandatory biochemical parameters assessed for all participants.

| **Biochemical Parameter** |
|---|
| Blood Glucose/HbA1c |
| HDL |
| LDL |
| Total Cholesterol |
| Triglycerides |
| Bilirubin |
| SGOT (AST) & SGPT (ALT) |
| Alkaline Phosphatase (ALP) |
| Urea, Serum |
| Creatinine, Serum |
| CBC Haemogram |

**Table S1.3**. List of sociodemographic and anthropometric measurements and lifestyle factors collected from all participants.

| **Sociodemographic measures** | **Lifestyle factors** | **Anthropometric measurements** |
|---|---|---|
| Name of the participant, City | Smoking status | Blood pressure |
| Date of Birth (age as on date) | Chewing tobacco status | Height |
| If any of your blood relations is participating in the study | Alcohol consumption status | Weight |
| Mother tongue | History of major illnesses or surgeries (Self) | Head Circumference |
| Gender | History of major illnesses or surgeries (Father) | Waist Measurement |
| Caste/ethnicity/community | History of major illnesses or surgeries (Mother) | Hip measurement |
| Father and Mother of the same caste | History of major illnesses or surgeries (Other Family Members) | |
| Marital status | Any medication being taken currently | |
| Number of children | If yes, name of medication | |
| Languages known to be spoken | | |
| Languages known to write | | |
| Highest level of education | | |
| Total years of education | | |
| Medium of education | | |
| Occupation | | |
| Income per month in rupees (Family) | | |

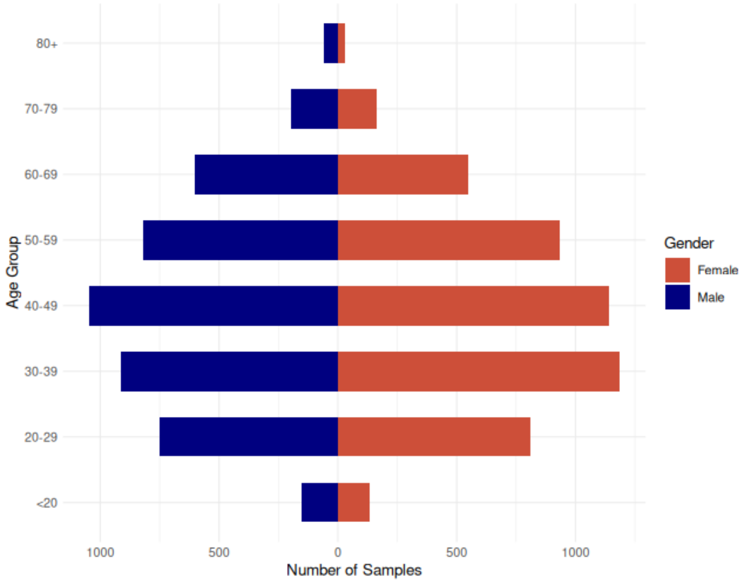

**Figure S1.2.** Age pyramid of sequenced samples in the GI project.

*Biobanking*: All 13 sampling institutions collected blood samples for biochemical tests and genetic studies. Two whole blood samples were collected in EDTA tubes for genetic studies, of which one vial was sent to the CBR Biobank for storage and future use.

The CBR Biobank serves as the primary repository to securely store and maintain DNA samples. It has been accredited in accordance with ISO 20387:2018 standard by the Quality & Accreditation Institute, Noida, India. Upon receipt, the samples are initially stored in –30°C freezers before being processed for DNA isolation. The isolated DNA, reconstituted in elution buffer, is aliquoted into smaller volumes and stored in –80°C freezers at the CBR Biobank. Currently, 20,195 samples are stored at the CBR Biobank. Of these, DNA has been successfully isolated from approximately 15,956 samples, while the remaining are in the process of DNA isolation.

**Description of the populations**

We collected 83 populations belonging to different linguistic and ethnic groups and from different biogeographic regions (FiguresS1.3, S1.4). The code structure for populations has four components, which are delimited by underscores. The first component with two letters represents the language group to which the population belongs, and the second component with three letters denotes the biogeographic zone of the primary habitat for the population. The third component has a digit which indicates whether the population is a tribe (1 for yes, 2 for no). If there are multiple populations fulfilling all these three criteria, they are listed alphabetically and numbered accordingly, as seen in the two digits of the fourth component.

The Language groups are the following:

- AA - Austroasiatic language speakers
- DR - Dravidian language speakers
- IE - Indo-European language speakers

- TB - Tibeto-Burman language speakers

As an example, if there are two non-tribal populations from the Eastern Coastal Plains, namely population XX and population YY, who speak languages belonging to the Dravidian Language Family, they will be designated as below:
XX will be coded as DR_ECP_2_01
YY will be coded as DR_ECP_2_02

The population details are provided in Table S1.4. Sample numbers reflect the number of individuals per group retained in the final dataset after joint calling and initial QC. In addition, we sampled from a Continentally Admixed Outgroup (CAO). Excluding CAO, the other 82 populations can be grouped into 7 ethnolinguistic groups (Table S1.5), as shown in Figs S1.3 and S1.4.

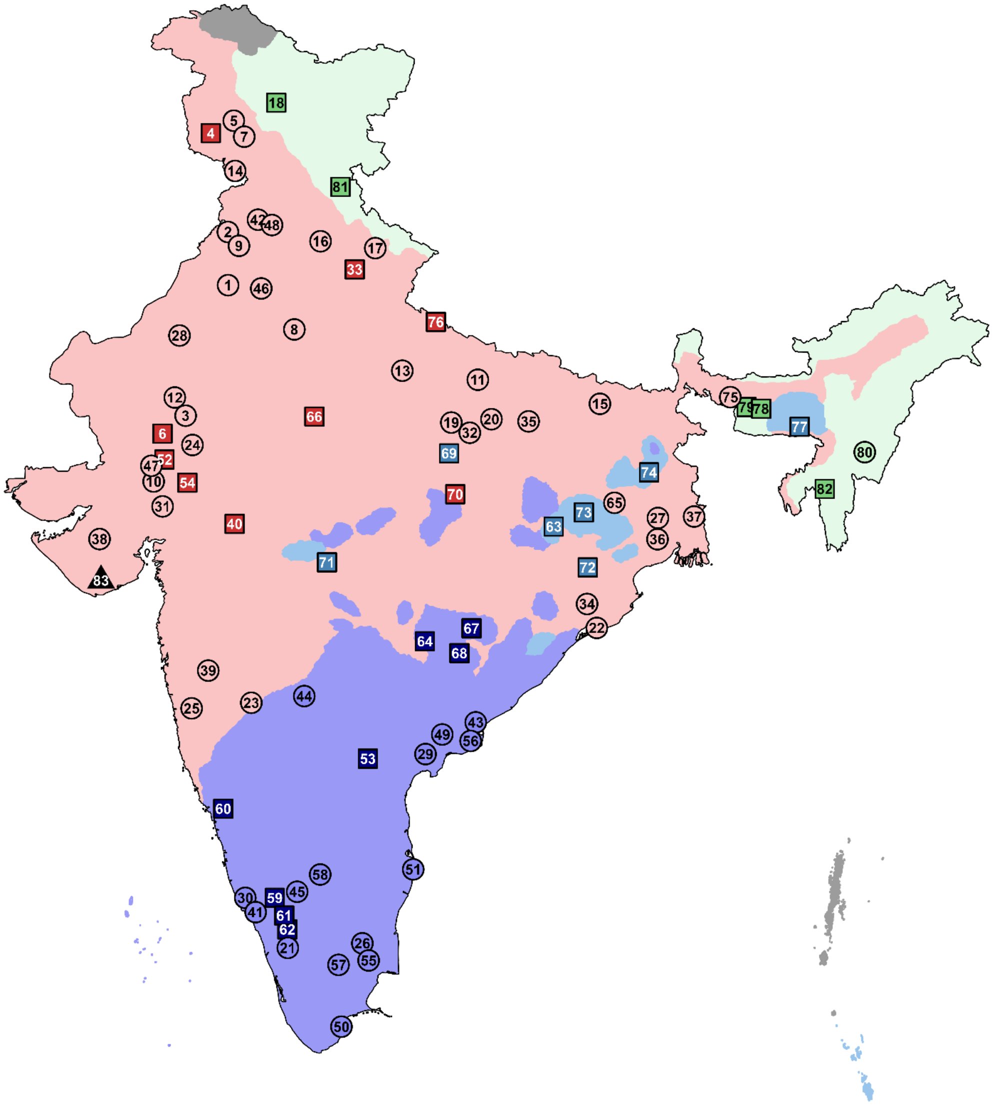

**Figure S1.3**. 83 populations coloured according to the 7 ethnolinguistic groups. Circles are used for non-tribal groups, and squares are used for tribal groups. Background color spreads are based on the spread of the linguistic groups. The points do not always reflect the sampling location, but rather an idealized location based on population history or current distribution or both.

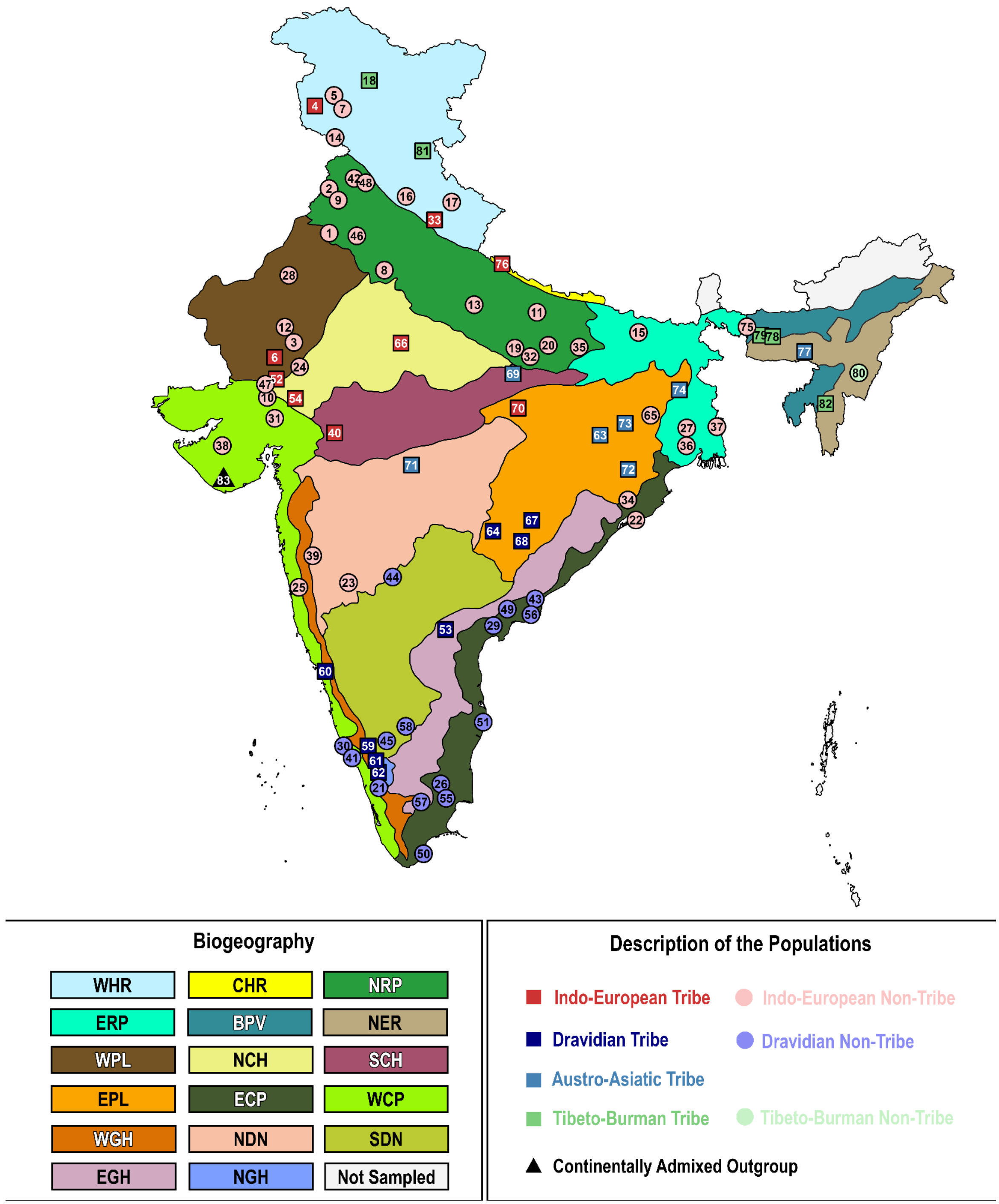

**Figure S1.4**. 83 populations coloured according to the 7 ethnolinguistic groups. Circles are used for non-tribes, and squares are for tribes. Background color spreads are based on the underlying biogeographic region. The points do not always reflect the sampling location, but rather an idealized location based on population history or current distribution or both.

**Table S1.4.** List of populations and ethnicity codes

| Numeric Code | Code | Linguistic Group | Tribe | Biogeography | Number of samples | Sample Collecting Institute |
|---|---|---|---|---|---|---|
| 01 | IE_NRP_2_05 | IE | No | Northern Riverine Plains | 149 | AIIMS-J, CBR, IGIB |
| 02 | IE_NRP_2_08 | IE | No | Northern Riverine Plains | 160 | IGIB |
| 03 | IE_WPL_2_02 | IE | No | Western Plains | 62 | AIIMS-J |
| 04 | IE_WHR_1_02 | IE | Yes | Western Himalayas | 67 | SKIMS |
| 05 | IE_WHR_2_04 | IE | No | Western Himalayas | 145 | SKIMS |
| 06 | IE_WPL_1_01 | IE | Yes | Western Plains | 161 | AIIMS-J, IGIB |
| 07 | IE_WHR_2_05 | IE | No | Western Himalayas | 140 | SKIMS |
| 08 | IE_NRP_2_02 | IE | No | Northern Riverine Plains | 161 | IGIB |
| 09 | IE_NRP_2_11 | IE | No | Northern Riverine Plains | 195 | IGIB |
| 10 | IE_WCP_2_01 | IE | No | Western Coastal Plains | 59 | GBRC |
| 11 | IE_NRP_2_10 | IE | No | Northern Riverine Plains | 166 | CBR, IGIB |
| 12 | IE_WPL_2_03 | IE | No | Western Plains | 99 | AIIMS-J, GBRC |
| 13 | IE_NRP_2_06 | IE | No | Northern Riverine Plains | 164 | CBR, IGIB |
| 14 | IE_WHR_2_01 | IE | No | Western Himalayas | 181 | SKIMS |
| 15 | IE_ERP_2_02 | IE | No | Eastern Riverine Plains | 186 | IBSD, IGIB |
| 16 | IE_WHR_2_03 | IE | No | Western Himalayas | 74 | CBR |
| 17 | IE_WHR_2_02 | IE | No | Western Himalayas | 164 | CBR, IGIB |
| 18 | TB_WHR_1_01 | TB | Yes | Western Himalayas | 38 | SKIMS |
| 19 | IE_NRP_2_12 | IE | No | Northern Riverine Plains | 154 | CBR, IGIB |
| 20 | IE_NRP_2_13 | IE | No | Northern Riverine Plains | 191 | AIIMS-J, IGIB |
| 21 | DR_WCP_2_02 | DR | No | Western Coastal Plains | 162 | RGCB |
| 22 | IE_ECP_2_02 | IE | No | Eastern Coastal Plains | 175 | ILS-B |

| Numeric Code | Code | Linguistic Group | Tribe | Biogeography | Number of samples | Sample Collecting Institute |
|---|---|---|---|---|---|---|
| 23 | IE_NDN_2_01 | IE | No | North Deccan | 163 | IISER-P |
| 24 | IE_NCH_2_01 | IE | No | North Central Highlands | 126 | AIIMS-J |
| 25 | IE_WCP_2_03 | IE | No | Western Coastal Plains | 164 | IISERP |
| 26 | DR_ECP_2_02 | DR | No | Eastern Coastal Plains | 165 | CBR, NIMHANS |
| 27 | IE_ERP_2_04 | IE | No | Eastern Riverine Plains | 162 | NIBMG |
| 28 | IE_WPL_2_01 | IE | No | Western Plains | 113 | IGIB |
| 29 | DR_ECP_2_08 | DR | No | Eastern Coastal Plains | 145 | CBR, CCMB |
| 30 | DR_WCP_2_01 | DR | No | Western Coastal Plains | 162 | RGCB |
| 31 | IE_WCP_2_04 | IE | No | Western Coastal Plains | 140 | GBRC |
| 32 | IE_NRP_2_07 | IE | No | Northern Riverine Plains | 155 | CBR, IGIB, NIBMG |
| 33 | IE_WHR_1_01 | IE | Yes | Western Himalayas | 76 | IGIB |
| 34 | IE_ECP_2_01 | IE | No | Eastern Coastal Plains | 143 | NIBMG |
| 35 | IE_NRP_2_03 | IE | No | Northern Riverine Plains | 158 | IGIB |
| 36 | IE_ERP_2_01 | IE | No | Eastern Riverine Plains | 170 | NIBMG |
| 37 | IE_ERP_2_03 | IE | No | Eastern Riverine Plains | 156 | ILSB, NIBMG |
| 38 | IE_WCP_2_05 | IE | No | Western Coastal Plains | 70 | GBRC |
| 39 | IE_NDN_2_02 | IE | No | North Deccan | 164 | IISER-P |
| 40 | IE_SCH_1_01 | IE | Yes | South Central Highlands | 80 | AIIMS-J |
| 41 | DR_WCP_2_03 | DR | No | Western Coastal Plains | 167 | RGCB |
| 42 | IE_NRP_2_04 | IE | No | Northern Riverine Plains | 161 | CBR, IGIB |
| 43 | DR_ECP_2_01 | DR | No | Eastern Coastal Plains | 56 | CCMB |
| 44 | DR_SDN_2_02 | DR | No | South Deccan | 162 | CBR, NIMHANS |
| 45 | DR_SDN_2_03 | DR | No | South Deccan | 59 | CBR |
| 46 | IE_NRP_2_01 | IE | No | Northern Riverine Plains | 167 | AIIMS-J, CBR |
| 47 | IE_WCP_2_02 | IE | No | Western Coastal Plains | 161 | GBRC, IISER-P |
| 48 | IE_NRP_2_09 | IE | No | Northern Riverine Plains | 167 | CBR, IGIB |
| 49 | DR_ECP_2_06 | DR | No | Eastern Coastal Plains | 165 | CBR |

| Numeric Code | Code | Linguistic Group | Tribe | Biogeography | Number of samples | Sample Collecting Institute |
|---|---|---|---|---|---|---|
| 50 | DR_ECP_2_05 | DR | No | Eastern Coastal Plains | 86 | CBR, CCMB, RGCB |
| 51 | DR_ECP_2_07 | DR | No | Eastern Coastal Plains | 63 | CCMB |
| 52 | IE_NCH_1_01 | IE | Yes | North Central Highlands | 77 | AIIMS-J |
| 53 | DR_EGH_1_01 | DR | Yes | Eastern Ghats | 34 | CCMB |
| 54 | IE_NCH_1_02 | IE | Yes | North Central Highlands | 67 | GBRC |
| 55 | DR_ECP_2_03 | DR | No | Eastern Coastal Plains | 148 | CCMB |
| 56 | DR_ECP_2_04 | DR | No | Eastern Coastal Plains | 125 | CCMB |
| 57 | DR_EGH_2_01 | DR | No | Eastern Ghats | 112 | CCMB, RGCB |
| 58 | DR_SDN_2_01 | DR | No | South Deccan | 32 | CBR |
| 59 | DR_WGH_1_01 | DR | Yes | Western Ghats | 45 | NIMHANS |
| 60 | DR_WGH_1_02 | DR | Yes | Western Ghats | 75 | NIMHANS |
| 61 | DR_NGH_1_02 | DR | Yes | Nilgiri Hills | 75 | RGCB |
| 62 | DR_NGH_1_01 | DR | Yes | Nilgiri Hills | 75 | RGCB |
| 63 | AA_EPL_1_01 | AA | Yes | Eastern Plateau | 91 | IBSD |
| 64 | DR_EPL_1_03 | DR | Yes | Eastern Plateau | 81 | ILS-B |
| 65 | IE_EPL_2_01 | IE | No | Eastern Plateau | 161 | NIBMG |
| 66 | IE_NCH_1_03 | IE | Yes | North Central Highlands | 75 | GBRC |
| 67 | DR_EPL_1_02 | DR | Yes | Eastern Plateau | 83 | ILS-B |
| 68 | DR_EPL_1_01 | DR | Yes | Eastern Plateau | 68 | ILS-B |
| 69 | AA_SCH_1_01 | AA | Yes | South Central Highlands | 65 | GBRC |
| 70 | IE_EPL_1_01 | IE | Yes | Eastern Plateau | 56 | AIIMS-J |
| 71 | AA_NDN_1_01 | AA | Yes | North Deccan | 50 | GBRC |
| 72 | AA_EPL_1_02 | AA | Yes | Eastern Plateau | 60 | ILS-B |
| 73 | AA_EPL_1_03 | AA | Yes | Eastern Plateau | 92 | IBSD |
| 74 | AA_EPL_1_04 | AA | Yes | Eastern Plateau | 168 | NIBMG |
| 75 | IE_BPV_2_01 | IE | No | Brahmaputra Valley | 114 | NIBMG |
| 76 | IE_CHR_1_01 | IE | Yes | Central Himalayas | 54 | CCMB |

| Numeric Code | Code | Linguistic Group | Tribe | Biogeography | Number of samples | Sample Collecting Institute |
|---|---|---|---|---|---|---|
| 77 | AA_NER_1_01 | AA | Yes | North Eastern Range | 75 | MZU |
| 78 | TB_BPV_1_02 | TB | Yes | Brahmaputra Valley | 105 | MZU |
| 79 | TB_BPV_1_01 | TB | Yes | Brahmaputra Valley | 127 | MZU |
| 80 | TB_NER_2_01 | TB | No | North Eastern Range | 159 | IBSD |
| 81 | TB_WHR_1_02 | TB | Yes | Western Himalayas | 56 | IGIB |
| 82 | TB_NER_1_01 | TB | Yes | North Eastern Range | 134 | MZU |
| 83 | CAO | NA | NA | NA | 50 | GBRC |

**Table S1.5.** Number of samples and populations per ethnolinguistic group

| Group | Number of samples | Number of populations |
|---|---|---|
| Indo-European Non-Tribe | 5,440 | 37 |
| Indo-European Tribe | 713 | 9 |
| Dravidian Non-Tribe | 1,809 | 15 |
| Dravidian Tribe | 536 | 8 |
| Tibeto-Burman Non-Tribe | 159 | 1 |
| Tibeto-Burman Tribe | 460 | 5 |
| Austro-Asiatic Tribe | 601 | 7 |
| Continentally Admixed Outgroup | 50 | 1 |

**Acknowledgements**

We acknowledge the following individuals who contributed to the sample collection, as well as the collection of anthropometric and sociodemographic data across the 13 centres.

Dr. Jagdish Goyal, Dr. Dharamveer Yadav, Dr. Jairam Yadav, Vivek Arora, Dr. Rajesh Reddy, Dr. Kamal Kant Sukla, Chandrabhan Singh, Babu Singh Shekhawat, Dr. Kanaram Meena, Ravi Nihaliya, Prashant Deora, SumanDeep Kaur, Arjun Jakhar, Bhaira Ram, Bhanwara Ram, Meenakshi Preek, Padam Singh, Sunil Kumar Poonia, Raju Ram, Ramesh Tailor, Umesh Lila Aggarwal, Surendra Kumar Bhil, Arvinda Thoudam, Teresa Tangpua, Hajarimayum Moushmi Sharma, Khuraijam Dolly Devi Mamuni Swain, Adyasha Mishra, Sourya Prakash Mishra, Sudarshan Jena, Baby Lalrintluangi, T. Vanlalhriatpuii, R. Lalengkimi, Ravi Kumar K L and Raveendra Vukkala, Society for Health and Demographic Surveillance, Birhum Population Project

(BIRPOP), Dr Abhijit Chowdhury, Dr Anamitra Barik, Dr Rajesh Kumar Rai, Mr Ashok Garai, Dr Mithun Das, Mr Sanjay Mukherjee, Ms Madhumita Pati, Mr Subhrajyoti Das, Dr Sanat Mahato, Mr Prabhat Mahato, Mr Sachin Mahato, Kalyani Cohort Studies, Mr Biltu Das, Mr Doel Sengupta, Mr Bikash Mondal, Mr Riya Roy, Ms Srilekha Halder, Mr Rabindranath Ghosh, Mr Uttam Maiti, Mr Ainur Rahaman, Ms Kakali Pal, Ms Sriparna Bose, Mr Judhajit Mishra, Mr Tithi Debnath, Mr Rabin Biswas, Late Swapan Kumar Ghosh, Mr Sukhabilash Burma, Mr Debabrata Chaki, Dr K J Sinha Roy, Mr Jogendra Nath Laskar, Dr Mrinmay Dhauria, Dr Swapan Roy, Mr Soumitra Banerjee, Ms Shreya Bhattacharyya, Dr Debdutta Bhattacharya, Dr Subrato Palo, Dr Srikant Kanungo, Dr Sanghamitra Pati, Mr Ashim Samanta, Mr Sumantra Ghosh, Mr Abhijit Santra, Mr Subhash Chandra Ghosh, Mr Bijan Bairagya, Mr Indranil Bagchi, Dr Subrata Patra, Ms Tithi Pal, Dr Kuntal Dey, Dr Disha Banerjee.

#### Supplementary Section S2: Whole Genome Sequencing and Quality Control

Chandrika Bhattacharyya[1], Krithika Subramanian[2,3], Sreelekshmi MS[4], Pratheusa Machha[4], Tiyasha De[5], Payel Mukherjee[4], Tulasi Nagabandi[4], Shreya Bari[5], Pooja Sharma[5], Shahrumi Reza[5,6], Shouvanik Sengupta[1,7], Devashish Tripathi[1,7], Vinay More[1,7], Bharathram Upilli[5], Khader Valli Rupanagudi[2], Nidhan Biswas[1,7], Analabha Basu[1,7], Bratati Kahali[2], Mohammed Faruq[5,6], Divya Tej Sowpati[4,6]

[1]BRIC - National Institute of Biomedical Genomics (BRIC-NIBMG), Kolkata, India. [2]Centre for Brain Research (CBR), IISc Campus, Bengaluru, India. [3]Manipal Academy of Higher Education (MAHE), Karnataka, India. [4]CSIR - Centre for Cellular and Molecular Biology (CSIR-CCMB), Hyderabad, India. [5]CSIR - Institute of Genomics & Integrative Biology (CSIR-IGIB), New Delhi, India. [6]Academy of Scientific and Innovative Research, Ghaziabad, India. [7]Regional Centre for Biotechnology (RCB), Faridabad, India.

#### Summary

##### Assessment of relatedness using genotyping arrays

A total of 20,195 blood samples were collected from healthy participants aged 18 and older, with their informed consent, alongside their phenotypic data (S1). Out of these, 13,242 samples were subjected to relatedness check using whole genome genotyping using either Illumina Global Screening Array (GSA) or Axiom™ Precision Medicine Research Array (PMRA) at the four sequencing centres (Fig S2.1). For this, DNA was isolated using standard protocols and dissolved in TE buffer. DNA integrity was confirmed by agarose gel electrophoresis, and its concentration and purity were assessed using NanoDrop™ spectrophotometry and Qubit fluorometer. The DNA was then used for array-based genotyping following manufacturer's protocols. Raw data was processed using Genome Studio (GSA) or Axiom™ Analysis Suite (PMRA). Samples with <95% call rate, and variants with <98% call rate or not in Hardy-Weinberg equilibrium (p-value cutoff $<10^{-5}$) were excluded. The genotypes were used to calculate the relatedness between pairs of individuals using --genome command of PLINK v1.9. Individuals who are related to each other up to first-cousin level (PI_HAT value >= 0.125) were excluded, retaining one individual per related pair. Genotyping data was also used to verify trio status. After this analysis, 10,074 samples were chosen for whole genome sequencing.

#### Sample Collection and Processing

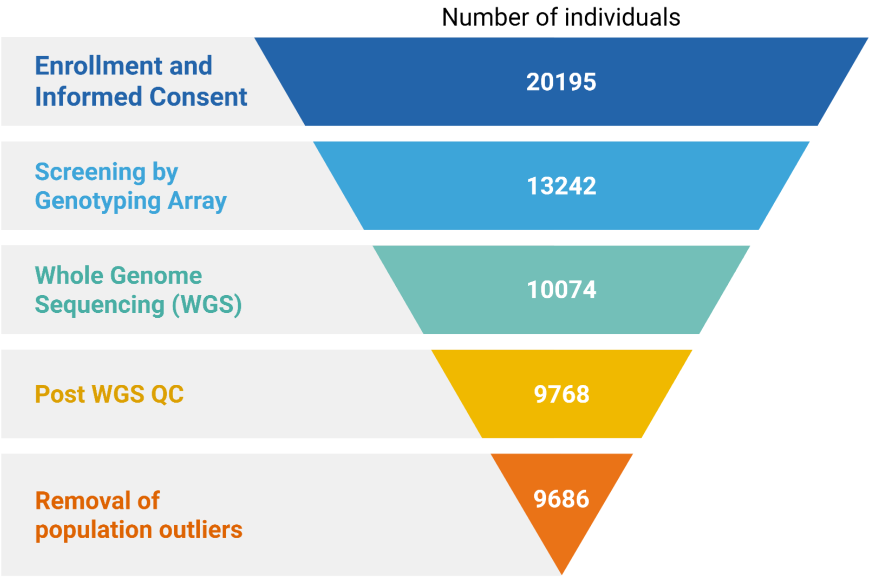

**Figure S2.1.** Funnel chart detailing the number of samples collected, processed, sequenced, and analyzed in the GenomeIndia project. The samples were collected from 83 different populations as described in Table S1.4 (Section S1).

##### Whole Genome Sequencing and Variant Calling

Whole-genome sequencing (WGS) was performed for 10,074 unrelated samples (other than the 244 trios included deliberately, by design). Sequencing was performed primarily at the four sequencing centres (BRIC-NIBMG, CBR, CSIR-CCMB, and CSIR-IGIB). Briefly, a PCR-free sequencing library was constructed for each sample using the Illumina TruSeq DNA PCR-Free HT kit. Libraries were sequenced on S4 flowcells in paired-end mode (2 x 150bp) on the Illumina NovaSeq 6000 platform, targeting a sequencing depth of 30x.

Data analysis was performed using a standardized, consistent protocol across the four centres. Raw BCL data was demultiplexed and basecalled into FASTQ files using bcl2fastq v2.20. Read QC, alignment, and variant calling was performed using DRAGEN v4.0.3. GRCh38 FASTA file containing all autosomes, sex chromosomes, mitochondrial DNA, and additional HLA decoy contigs was used as the reference genome, and was indexed using `dragen –build-hash-table`. Lane-wise FASTQ files of each sample were aligned to the reference genome, and variants were called emitting gVCF files. An example command is given below:

```
dragen -f \
-r /path/to/your/reference/<Ref> \
--fastq-list <sampleName>_FastqList.csv \
--enable-variant-caller true \
--vc-emit-ref-confidence GVCF \
--output-directory /path/to/your/output/<sampleName> \
```

```
--trim-min-quality 15 \
--trim-adapter-read1 <illumina_adapter_r1.txt> \
--trim-adapter-read2 <illumina_adapter_r2.txt> \
--output-file-prefix <sampleName> \
--enable-duplicate-marking true \
--enable-map-align-output true \
--read-trimmers adapter,quality \
--trim-min-length 50 \
--enable-bam-indexing true
```

**Performance benchmarking and cross-centre validation**

For benchmarking, cell lines corresponding to four samples from the Genome in a Bottle (GIAB) dataset (GM12878, GM24143, GM24149, GM24385), procured by the Centre for Brain Research (CBR), were sequenced at all the four centres reaching a minimum average depth of 30x. These samples were processed using the same pipeline as mentioned above, and were compared to the respective truth sets offered by the National Institute of Standards and Technology (NIST) using hap.py (Illumina/Hap.Py, 2015/2025). The analysis yielded a recall of ≥0.97, a precision of 0.99, and an F1 score of 0.98 for both SNVs and INDELs, indicating a strong concordance with the truth set across all the centres (Fig S2.2A). We further employed a cyclical sequencing concordance validation method, using one set of trio and two unrelated samples, randomly selected and sequenced at two centres (Fig S2.2B). Cross-centre comparisons showed an average recall of 0.99, precision of 0.97, and an F1 score of 0.98, indicating minimal batch effects. The consistent results across the four centres highlight the reliability and reproducibility of the project outcomes.

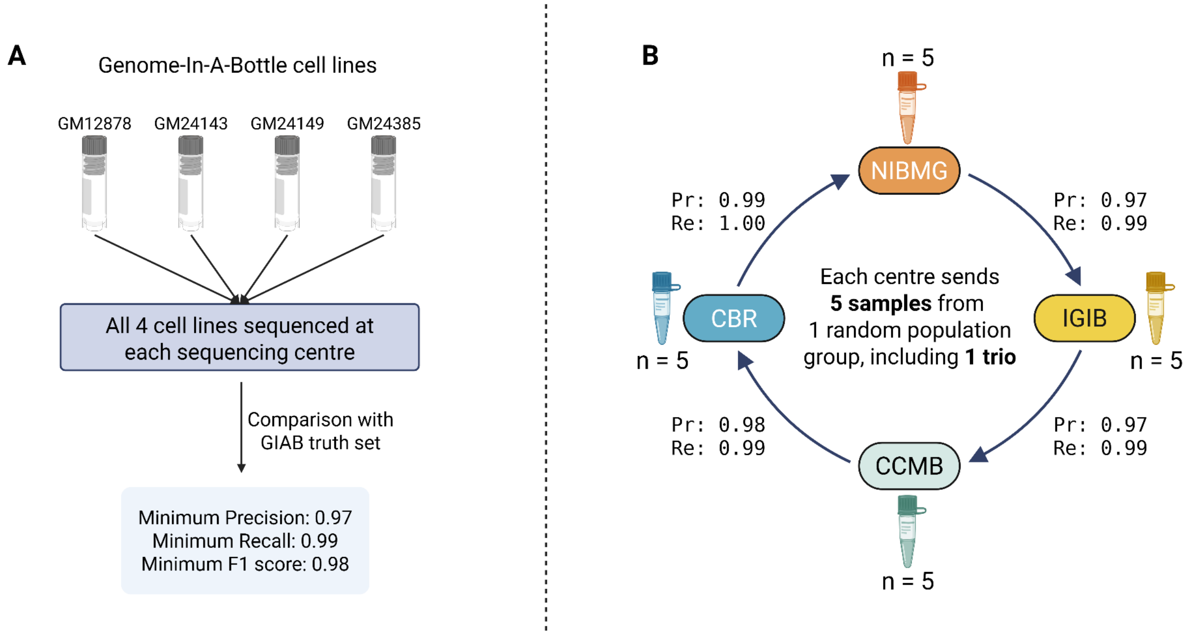

**Figure S2.2**. Performance benchmarking to ensure uniformity and reliability across the sequencing centres. A) Four GIAB cell lines were sequenced at each centre and compared with the respective truth set provided by the National Institute of Standards and Technology (NIST). B) Five DNA samples (1 trio, 2 unrelated) from a randomly chosen population group were selected and sent to another sequencing centre in a cyclical manner. The variants for each of these five samples were compared for concordance. Pr, precision; Re, recall.

#### Sample level filtering and dataset generation

From the initial set of 10,074 sequenced samples, we applied stringent quality control (QC) filters based on sequencing and alignment metrics. All retained samples achieved a minimum sequencing depth of 23x (Fig S2.3), sufficient for robust genome-wide variant detection. To ensure data quality and consistency, several per-sample metrics were evaluated: (1) Total variant count (SNVs and indels), with extreme deviations flagged as potential sequencing or processing artifacts; (2) Callability, defined as the fraction of the genome with high-confidence genotype calls, with samples retained only if callability ≥ 94%; (3) Heterozygous-to-homozygous (Het/Hom) ratio of ~1.6; and (4) Transition-to-transversion (Ti/Tv) ratio, typically between 1.98–1.99 (Fig. S2.2). This resulted in the exclusion of low-quality samples, yielding 9,871 high-quality samples for variant calling. We further performed principal component analysis (PCA) to identify and exclude 99 outlier samples, mitigating potential confounding due to population substructure. Among the remaining individuals, we detected and removed four genetically identical samples, resulting in a final dataset of 9,768 unique, high-confidence samples.

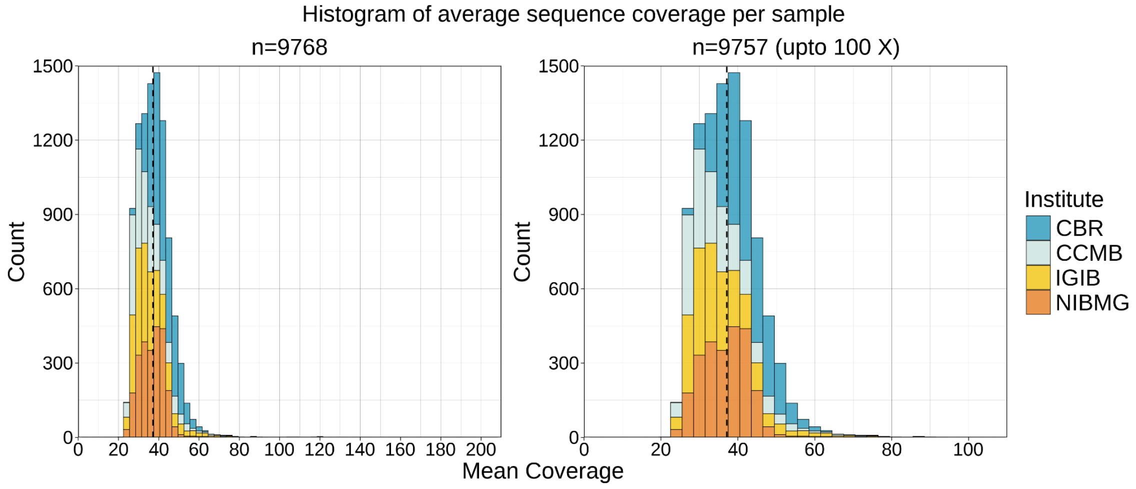

**Figure S2.3.** WGS coverage of the GenomeIndia samples. The median coverage of the analyzed samples is 37x, and the minimum is 23x. The right plot is a zoomed version where the axis is limited to 100x, removing few outliers (n = 11) with very high sequencing coverage.

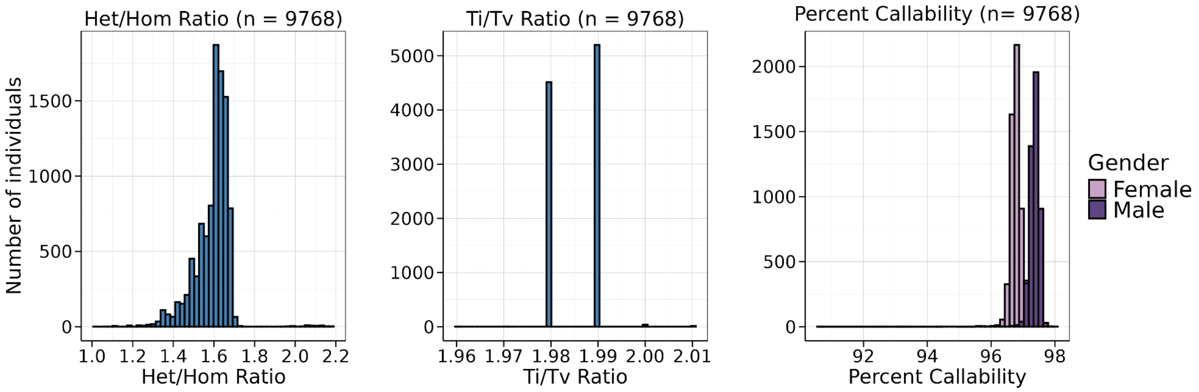

**Figure S2.4.** Quality control metrics for GenomeIndia study dataset across sequencing centres. Het/Hom ratios cluster around the expected value of ~1.6; Ti/Tv ratios remain within the expected range (1.98–1.99). Most samples show a consistent callability >96%.

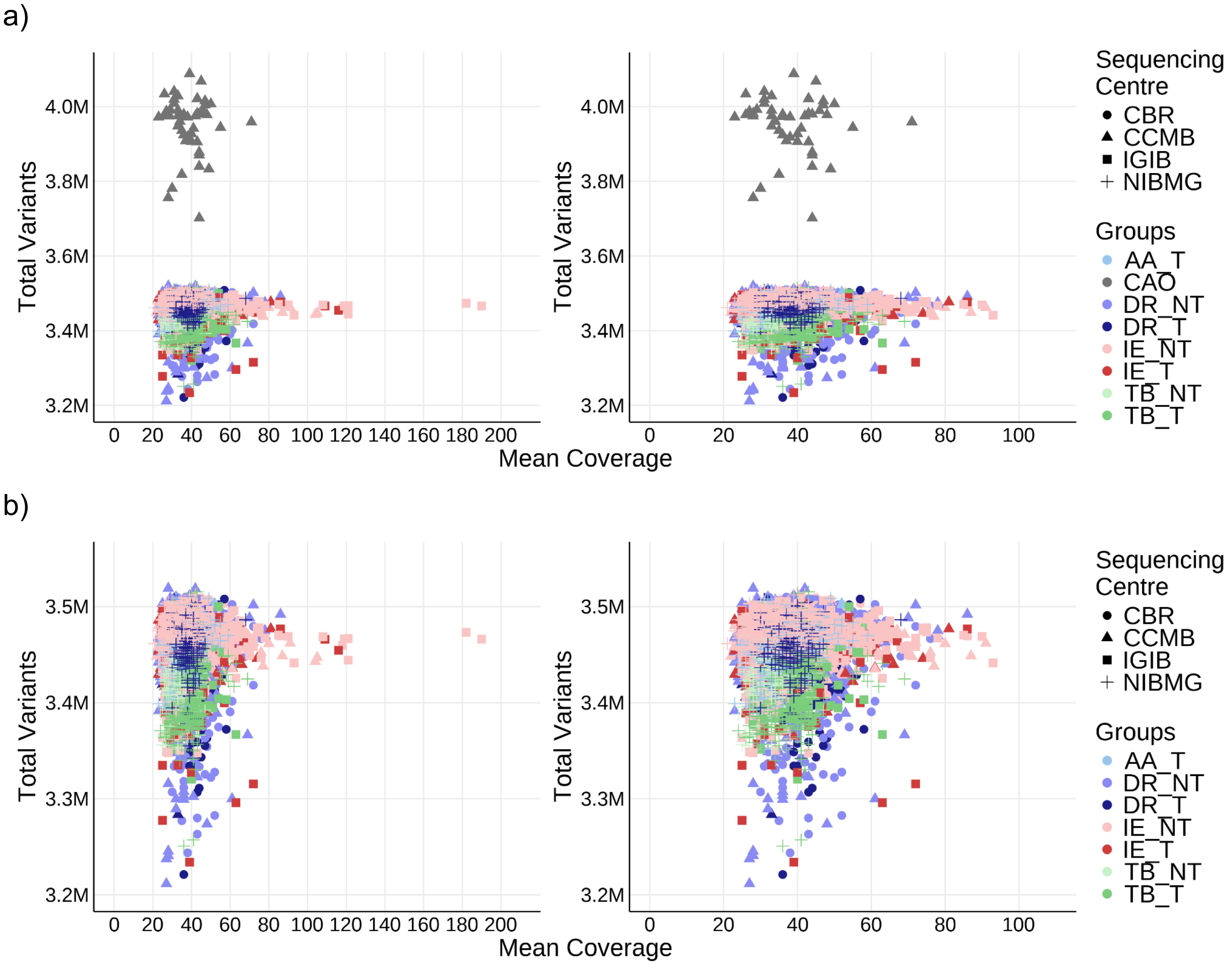

**Figure S2.5.** Showing the number of total variants plotted against their corresponding mean coverage in a) all 9768 samples and b) excluding the CAO samples. The plots highlight the fact that the detection of variants is not dependent on the sequencing coverage of the sample, as we observe no correlation between sequencing coverage and the total number of variants identified. CAO is an exception, showing higher variant counts than the other populations, and is reflective of its recent African ancestry.

**Variant identification using the DRAGEN gVCF Genotyper and variant filtering**

Joint genotyping was performed on 9,768 high-quality samples from the GenomeIndia cohort using the Illumina DRAGEN v4.2.4 Iterative gVCF Genotyper (link below). A collaborative framework between CBR and BRIC-NIBMG ensured methodological uniformity, with ~35 TB of gVCFs and quality metrics exchanged and ~0.7 million CPU hours utilized. The process involved dividing samples across ten batches of ~1,000 gVCFs each, to generate per-batch cohort and census files, which were then combined into a global census file. The global census file is then used to produce batch-wise multi-sample VCF (msVCF) files and finally to produce joint-called VCF combining all the msVCFs together. This process yielded ~188 million variants. To mitigate center-specific biases, only variants concordantly identified by both centers were retained, resulting in a robust, high-confidence dataset for downstream population and association analyses.

We implemented a stringent, allele frequency-aware QC framework. For rare variants (MAC = 1–2; singletons or doubletons in heterozygous state), we applied strict filters to reduce false positives, retaining only those that (i) passed all internal DRAGEN filters, (ii) had QUAL ≥ 30, and (iii) were supported by at least one genotype with GQ > 40. For variants with MAC ≥ 3, we extended QC to ensure robustness in more common alleles. Retained variants were required to (i) pass internal filters, (ii) have QUAL ≥ 30, (iii) exhibit call rates ≥ 98%, and (iv) be supported by at least one sample with GQ > 20. Heterozygous genotypes were additionally evaluated for allele balance (AB); calls with AB < 0.2 were masked as missing. We excluded variants deviating from Hardy–Weinberg equilibrium ($p < 1\times10^{-11}$) and those with an inbreeding coefficient of 1, which typically indicate technical artifacts or non-segregating sites. The filters used post genotyping are summarized in Table S2.1.

Link for Illumina gVCF genotyper:

https://sapac.illumina.com/science/genomics-research/articles/gVCF-Genotyper.html

**Table S2.1.** Filters used after joint-genotyping on variant calls.

| Filter | Description |
|---|---|
| **QUAL ≥ 30** | Posterior genotype probability ≥ 99.9%. (QUAL = GP(GT=0/0), Phred scale). |

| | |
|---|---|
| **NS_GT ≥ 98%** | Retained variants genotyped in ≥ 98% of samples. |
| **Inbreeding Coefficient** | 1 - O(het) / E(het). Values ≈ 0 indicate Hardy-Weinberg equilibrium; extreme negative values flagged potential issues. |
| **GQ > 20** | Required for at least one genotype call GQ > 20 at each site. Poor-quality genotype calls removed. |
| **Allelic Balance (AB)** | For heterozygous calls, AB is required to be more than 0.2, otherwise set to missing. |

**Outlier detection for population misclassification**

For each pre-defined population group, we have performed PCA and computed its centroid. The squared Mahalanobis distance (D2) is computed for each individual based on the first five PCs, with respect to its population centroid. Individuals were considered outliers if their distance is significantly large (p-value <= 1e-6) from the centroid. Examples of such outliers are shown in Figure S2.6. A total of 82 outlier individuals were detected from the final joint-called dataset of 9,768 (Table S2.2). The data from the remaining 9686 individuals was used for all population-specific analyses, whereas the full data of 9768 individuals was used for all broader analyses.

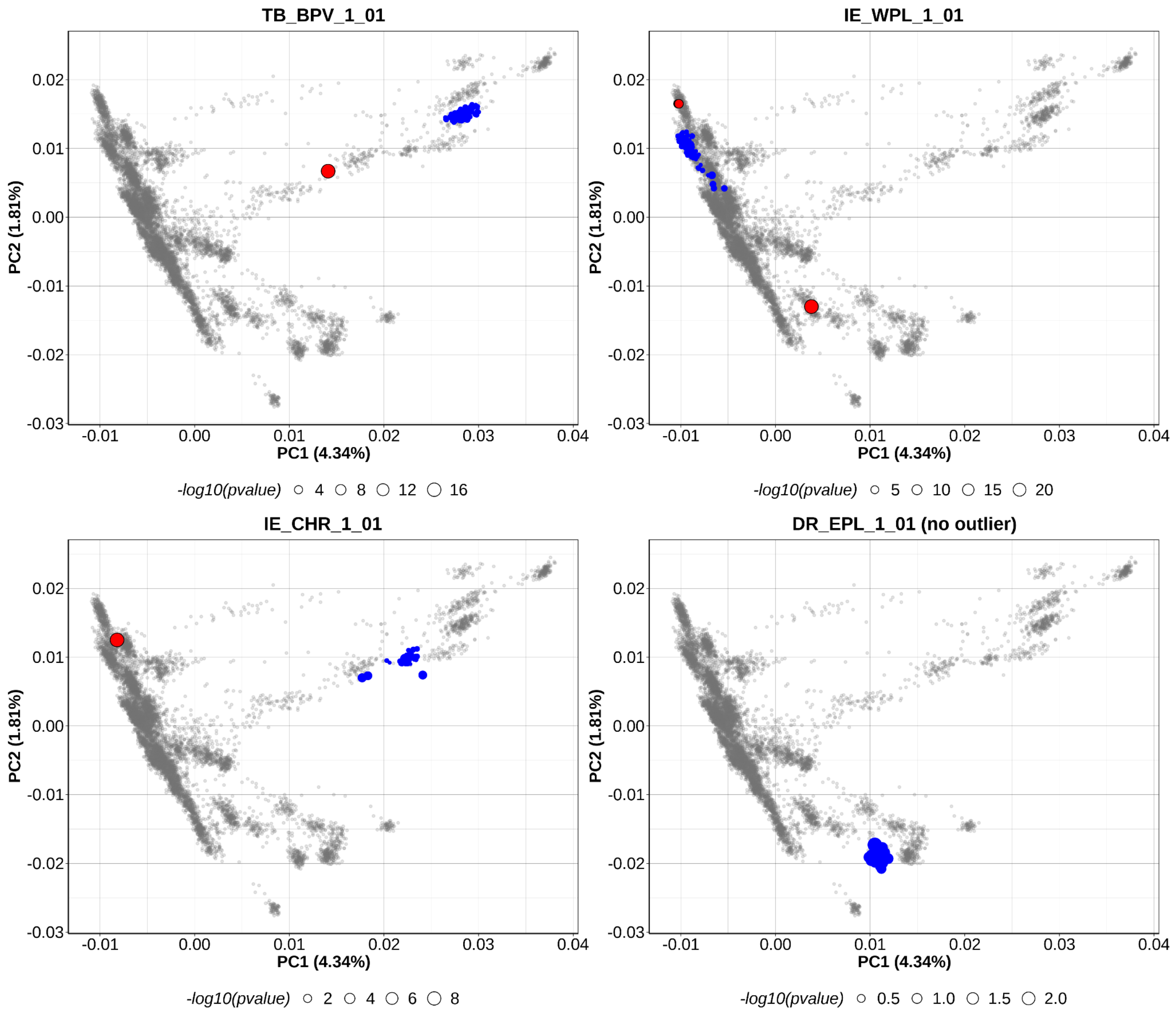

**Figure S2.6.** Outlier detection from PCA for identifying individuals with population names mislabelled. 4 representative populations are shown, where all samples of a population are colored in blue, and the detected outlier is highlighted in red. Samples from other populations are in grey. The bottom right population (DR_EPL_1_01) is an example where no outlier was detected.

**Table S2.2.** PCA-identified outlier samples (n = 82) and their group annotations

| Sample | Sex | Ethnolinguistic Group | Population | Biogeography |
|---|---|---|---|---|
| TY50898579KJ | XY | IE_non-tribe | IE_NRP_2_06 | NRP |
| XN06968311YG | XY | DR_non-tribe | DR_SDN_2_03 | SDN |

| | | | | |
|---|---|---|---|---|
| GR65199593UP | XY | IE_tribe | IE_WPL_1_01 | WPL |
| TD58196129QC | XY | IE_non-tribe | IE_ERP_2_04 | ERP |
| EM63826459TS | XY | IE_non-tribe | IE_NDN_2_02 | NDN |
| XI36126439XW | XY | IE_non-tribe | IE_WHR_2_04 | WHR |
| AG47996967OQ | XX | IE_non-tribe | IE_NRP_2_09 | NRP |
| HR28183327RS | XY | IE_non-tribe | IE_WCP_2_05 | WCP |
| IR49030528SS | XY | AA_tribe | AA_EPL_1_04 | EPL |
| QD92062170FY | XY | IE_non-tribe | IE_ERP_2_02 | ERP |
| HU56431935YE | XY | AA_tribe | AA_SCH_1_01 | SCH |
| BG72592048QY | XX | IE_tribe | IE_NCH_1_03 | NCH |
| CO31363422TQ | XY | IE_non-tribe | IE_NRP_2_02 | NRP |
| EJ52429239GX | XY | IE_tribe | IE_NCH_1_02 | NCH |
| LO76559321DT | XX | IE_tribe | IE_NCH_1_01 | NCH |
| LU12064055IV | XY | IE_tribe | IE_WPL_1_01 | WPL |
| TH31993251CH | XY | IE_non-tribe | IE_NCH_2_01 | NCH |
| WO58038890VT | XX | IE_non-tribe | IE_NRP_2_02 | NRP |
| YG54186654SI | XY | IE_non-tribe | IE_NRP_2_08 | NRP |
| BA19988738AA | XY | IE_tribe | IE_NCH_1_03 | NCH |
| EA63022348DP | XY | IE_non-tribe | IE_WHR_2_03 | WHR |
| RH88376284OT | XY | IE_non-tribe | IE_WCP_2_04 | WCP |
| TU67563736LE | XX | IE_non-tribe | IE_NRP_2_03 | NRP |
| CX93272239QM | XY | IE_non-tribe | IE_WHR_2_05 | WHR |
| PP88559301YV | XY | IE_non-tribe | IE_WCP_2_05 | WCP |
| PQ62124534DF | XX | IE_non-tribe | IE_NRP_2_11 | NRP |
| SG82435857HP | XX | IE_non-tribe | IE_NRP_2_05 | NRP |
| TB43957324RH | XY | IE_tribe | IE_WPL_1_01 | WPL |
| XA03403523EX | XY | IE_non-tribe | IE_ERP_2_03 | ERP |
| ZH24702510XR | XY | DR_non-tribe | DR_SDN_2_02 | SDN |
| BE79145216KE | XY | IE_non-tribe | IE_ERP_2_01 | ERP |
| ES85476302RN | XX | IE_non-tribe | IE_WHR_2_02 | WHR |
| JM59082891GR | XX | DR_non-tribe | DR_ECP_2_06 | ECP |
| AA13901862BS | XX | TB_non-tribe | TB_NER_2_01 | NER |
| GH56292722GY | XY | DR_non-tribe | DR_ECP_2_06 | ECP |
| HY15187359WL | XY | IE_non-tribe | IE_WPL_2_01 | WPL |
| JR46659699XB | XY | IE_tribe | IE_SCH_1_01 | SCH |

#### Supplementary Section S3: Allele Frequency Spectrum of GenomeIndia dataset

Shouvanik Sengupta[1,2], Arghya Dey[1], Krithika Subramanian[3,4], Chandrika Bhattacharyya[1], Sauma Suvra Majumdar[3,5], Mohammed Faruq[6,7], Divya Tej Sowpati[7,8], Bratati Kahali[3], Analabha Basu[1,2]

[1]BRIC - National Institute of Biomedical Genomics (BRIC-NIBMG), Kolkata, India. [2]Regional Centre for Biotechnology (RCB), Faridabad, India. [3]Centre for Brain Research (CBR), IISc Campus, Bengaluru, India. [4]Manipal Academy of Higher Education (MAHE), Karnataka, India. [5]Interdisciplinary Mathematical Sciences, Indian Institute of Science (IMI- IISc), Bengaluru, India. [6]CSIR - Institute of Genomics & Integrative Biology (CSIR-IGIB), New Delhi, India. [7]Academy of Scientific and Innovative Research, Ghaziabad, India. [8]CSIR - Centre for Cellular and Molecular Biology (CSIR-CCMB), Hyderabad, India.

#### Summary

The allele frequency spectrum (AFS) defines the basic premise of how genetic variation is distributed within a population, making it fundamental to population genetics. By characterizing the distribution of rare, intermediate, and common alleles, we make the primary inferences on key evolutionary and demographic events—such as drift, migration, bottlenecks, and selection—that have shaped the genetic diversity of different populations of the GenomeIndia Project. In addition to these insights, studying the AFS enables us to identify novel variants, including those not previously reported in global genetic studies, which is particularly important for underrepresented populations. Our identified set of common variants, along with newly discovered population-specific variants, can serve as a valuable resource for designing a new, more informative and inclusive genotyping chip tailored for India. Such a platform would strengthen the power and precision of future large-scale genetic studies by ensuring that variants relevant to the Indian population are properly captured and analyzed.

In the first subsection (S3.1), we look into the details of the distribution of total variants and singletons at:

- Individual level (9768 individuals)
- Population level (82 groups, excluding CAO) and
- Ethnolinguistic level (8 groups, including CAO)

We then look at the distribution of the novel variants across the whole population as well as across the groups (S3.2). Finally, we look into the difference in allele frequencies pertaining to populations, using $F_{ST}$ (S3.3) and the sharing of different alleles amongst the population groups (S3.4).

##### S3.1.1 The distribution of the total variants and singletons for each individual

We observe an overall trend of singleton count increasing with variant count when looking across all the 7 ethnolinguistic groups, after removing CAO individuals (as a recently and continentally admixed group they would have a lot of variants) and children of trios (as they would have a very small singleton count).

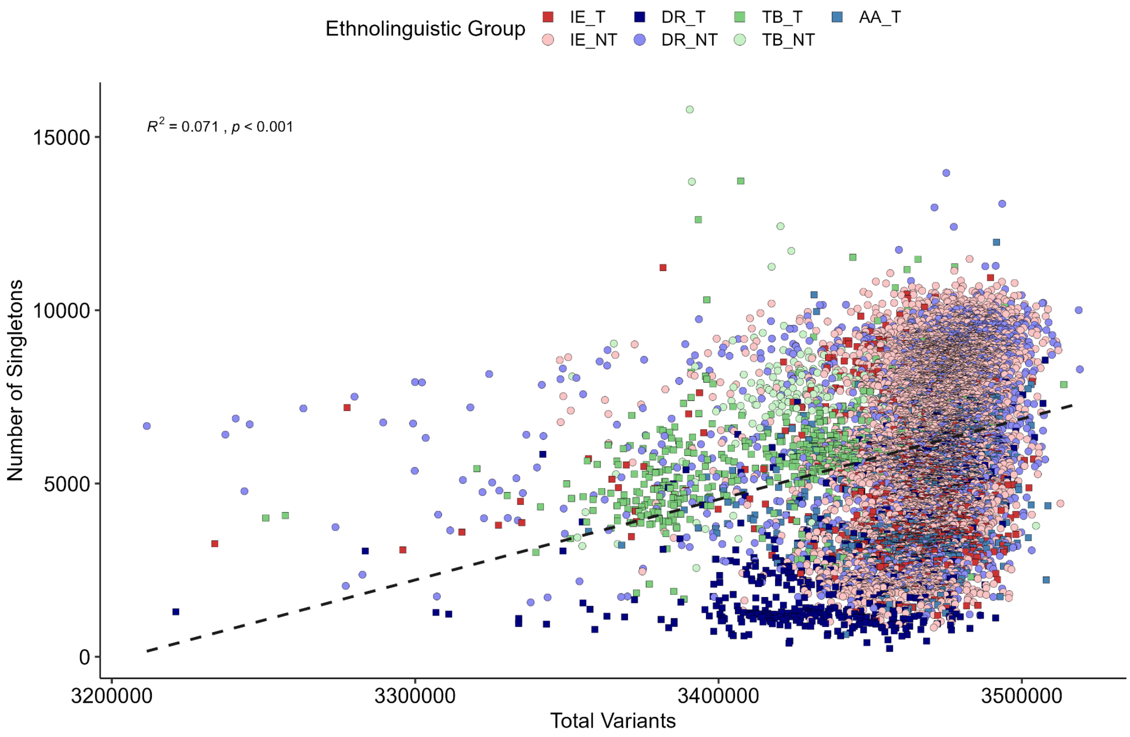

**Figure S3.1.1a.** Scatterplot of the total number of variants for each individual (X-axis) and the number of singletons (Y-axis). Individuals from the TB populations typically show fewer variants but NOT fewer singletons. Some individuals from the AA and DR tribal populations have very low singleton counts.

When looking at each of the groups separately, we observe the same positive trend in most cases, except in Indo-European tribes (Fig S3.1.1b).

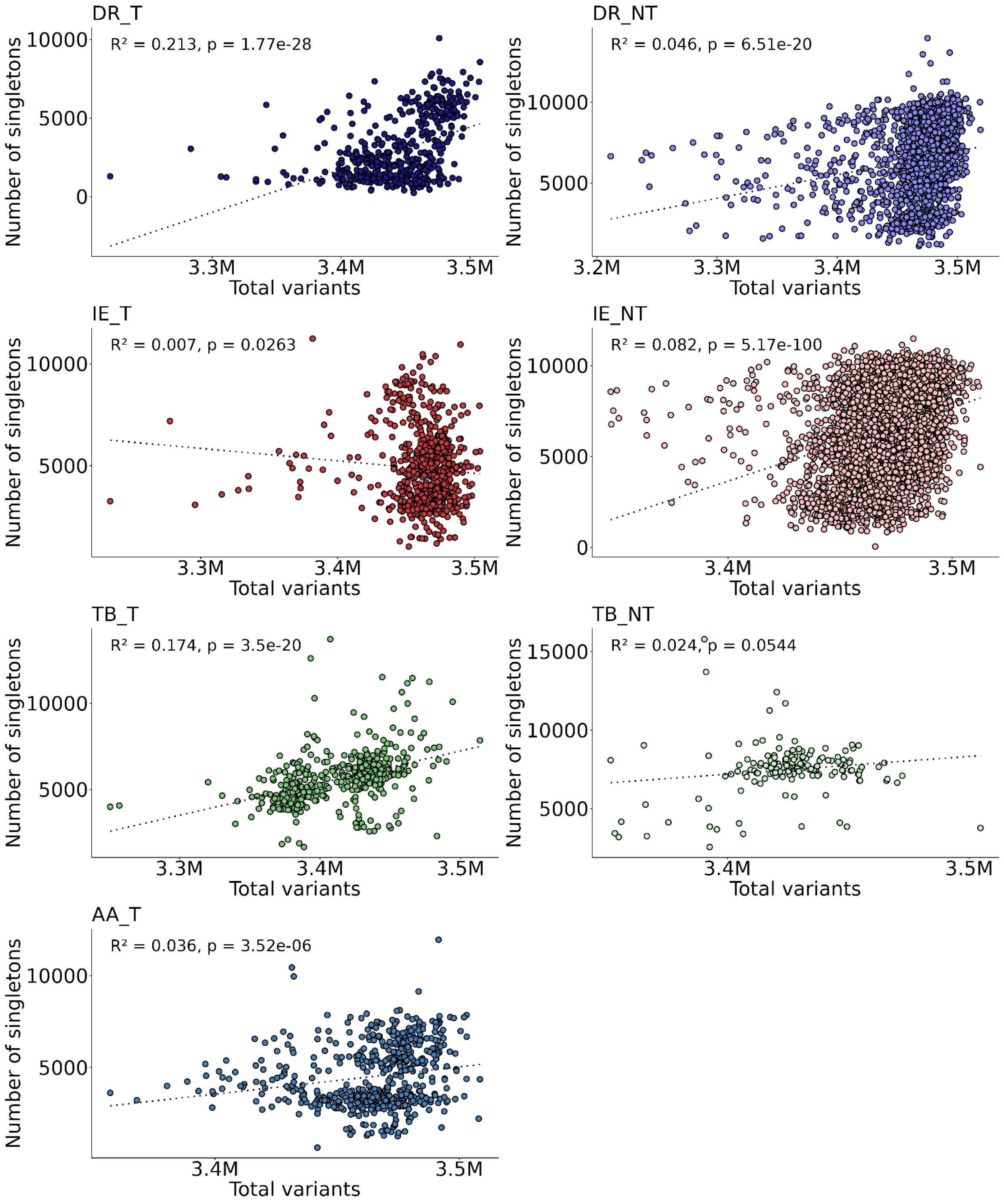

**Figure S3.1.1b.** Linguistic group-wise scatterplot of the total number of variants for each individual (X-axis) and the number of singletons (Y-axis).

##### S3.1.2 Number of variants and number of singletons for each population

The total number of variant sites per genome in the GI dataset do not vary too much except the CAO which is a continentally admixed population primarily between African and Indians. As expected the total number of variants observed in the CAO individuals are significantly higher than the other Indian population groups (Fig 1a). We also observe that the TB speaking populations from North East India generally have a lower count of the total number of variants. The Indo-European non-tribal (IE_NT) populations show lower dispersion in variant counts, as compared to other ethnolinguistic groups (Fig S3.2a).

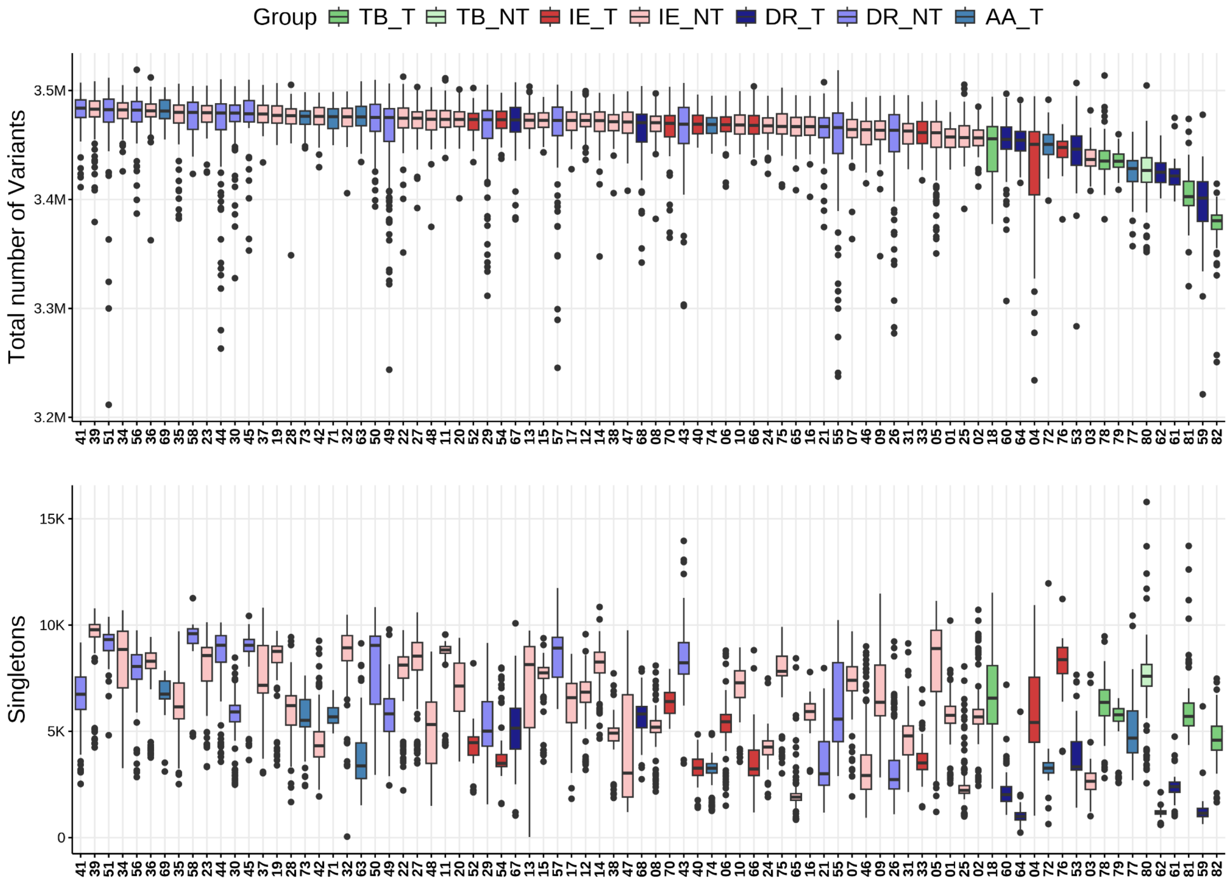

**Figure S3.1.2.** (a) Number of variant sites across individuals of 82 populations. (b) Distribution of the number of singleton variants across individuals. [CAO and PCA outliers are excluded in both (a) and (b). Children of trios were additionally excluded in (b)

##### Section S3.1.3. Distribution of variants among the 8 ethnolinguistically defined clusters

If we pool the individual populations to the 8 broad ethnolinguistic clusters, the summary statistics and the distribution of the variants within these 8 ethnolinguistic groups will be as shown in table S3.1.3 which shows the summary statistics of the variant count distributions overall as well as separately for the 7 ethnolinguistic groups. The PCA outliers are excluded from the analysis.

**Table S3.1.3.** The summary statistics of the total number of variants observed in each population and in GenomeIndia overall. As evident from Figure S3.1.3, the IE_NT have significantly lower standard deviation (sd).

| Group | min | max | mean | q1 | median | q3 | sd |
|---|---:|---:|---:|---:|---:|---:|---:|
| IE_T | 3,233,978 | 3,503,850 | 3,462,237 | 3,455,110 | 3,466,896 | 3,475,598 | 25,437.55 |
| IE_NT | 3,347,669 | 3,512,698 | 3,469,933 | 3,461,719 | 3,471,518 | 3,480,405 | 16,446.37 |
| DR_T | 3,221,138 | 3,507,684 | 3,442,284 | 3,421,214 | 3,445,420 | 3,467,715 | 33,883.65 |
| DR_NT | 3,211,536 | 3,519,088 | 3,466,443 | 3,461,158 | 3,475,708 | 3,485,586 | 34,911.14 |
| TB_T | 3,250,755 | 3,513,807 | 3,416,353 | 3,386,606 | 3,426,379 | 3,439,096 | 33,497.82 |
| TB_NT | 3,351,772 | 3,504,651 | 3,425,324 | 3,414,821 | 3,426,649 | 3,438,494 | 23,563.51 |
| AA_T | 3,357,127 | 3,508,542 | 3,465,222 | 3,456,838 | 3,469,322 | 3,478,977 | 21,752.45 |
| CAO | 3,701,477 | 4,087,906 | 3,952,880 | 3,924,773 | 3,974,901 | 3,991,519 | 76,944.89 |
| **GI Overall** | 3,211,536 | 4,08,7906 | 3,466,166 | 3,455,835 | 3,469,745 | 3,480,183 | 44,888.40 |

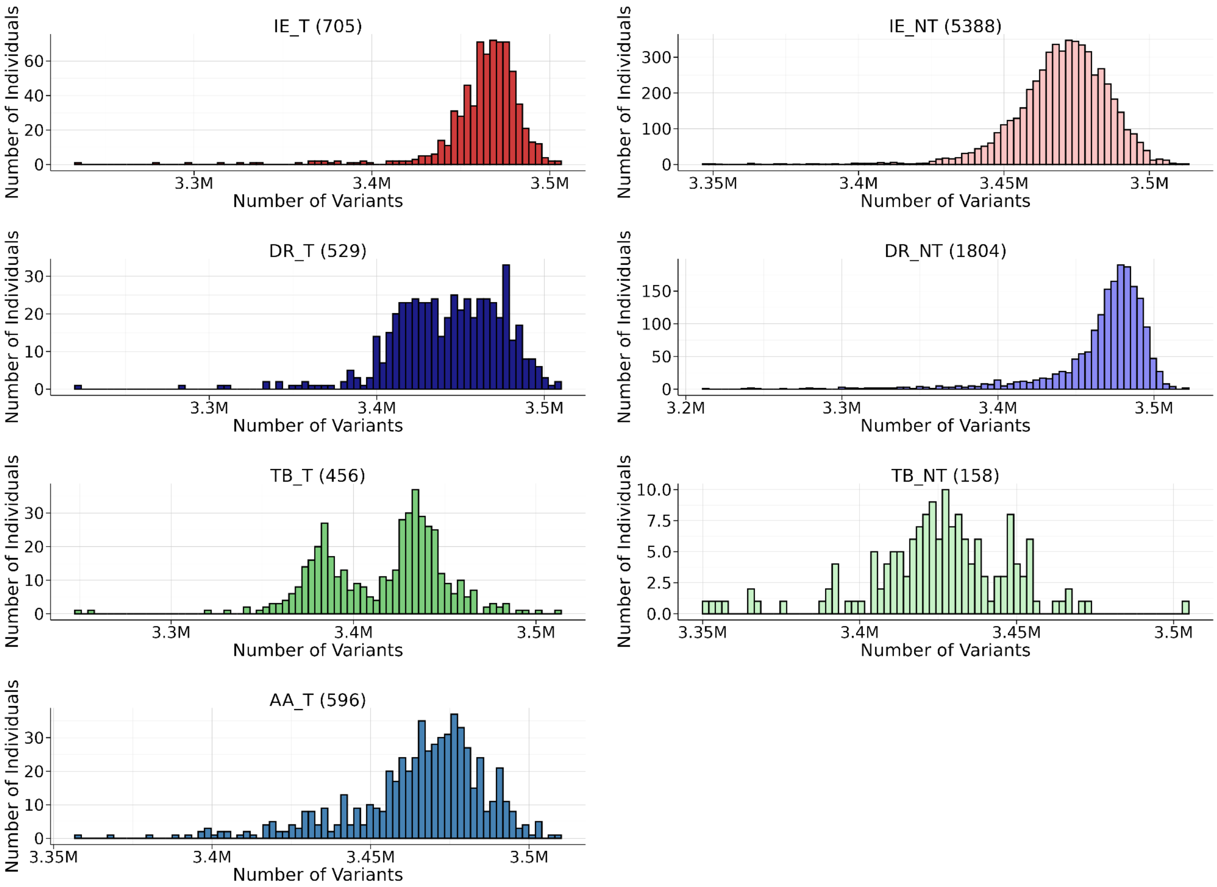

**Figure S3.1.3.** Variant histograms overall and across the different GI population groups. Numbers in parentheses indicate sample size of the group.

#### S3.2. Distribution of "novel" variants in the GI dataset

##### Identification of novel genetic variants absent/not reported from the global population

We define novel variants in the GenomeIndia cohort in two different methods as is applicable to the appropriate question. For questions which relate to population frequencies, we looked into variants which are not present in population datasets. In order to find variants which are not reported at all in any global database we define those novel variants which are not reported in any of four major global variant databases: dbSNP (build 156), gnomAD (v4.1), the 1000 Genomes Project (phase 3), or GenomeAsia. To identify such variants, we matched four key parameters "CHR:POS:REF:ALT"—chromosome, genomic position, reference allele, and alternate allele—between the GenomeIndia dataset and these databases. Variants that were novel necessarily need to have a genomic position which is not included in this list. We identified 44,034,341 novel variants, representing 33.89% of all detected variants. Among these, 24.7% of the total variants are novel singletons, and an additional 9.19% of the total variants are novel variants with a minor allele count (MAC) ≥ 2, indicating substantial representation of previously unreported genetic variation within Indian genomes.

This statistic changes significantly if we consider the novel variants which are group specific.

Table S3.2.1 documents 133,753 novel genetic variants that are commonly found within specific population clusters, yet are rare or absent in others. The majority of these variants are contributed by three tribal groups: DR_T (43.91%), AA_T (37.05%), and TB_T (11.11%). These variants occur in more than 1% of individuals within their respective clusters, underscoring their local prevalence. This pattern of distribution illustrates how antiquity of populations along with historical population structure and demographic bottlenecks have significantly shaped the genetic diversity of India's populations.

**Table S3.2.1.** Distribution of novel variants (>1%) in corresponding population clusters

| **Clusters** | **Number of novel variants (>1%)** |
|---|---|
| DR_T | 47555 |
| AA_T | 40131 |
| TB_NT | 25447 |
| TB_T | 12034 |
| IE_T | 2939 |
| DR_NT | 564 |
| IE_NT | 399 |
| >1% novel variant in multiple clusters | 4684 |

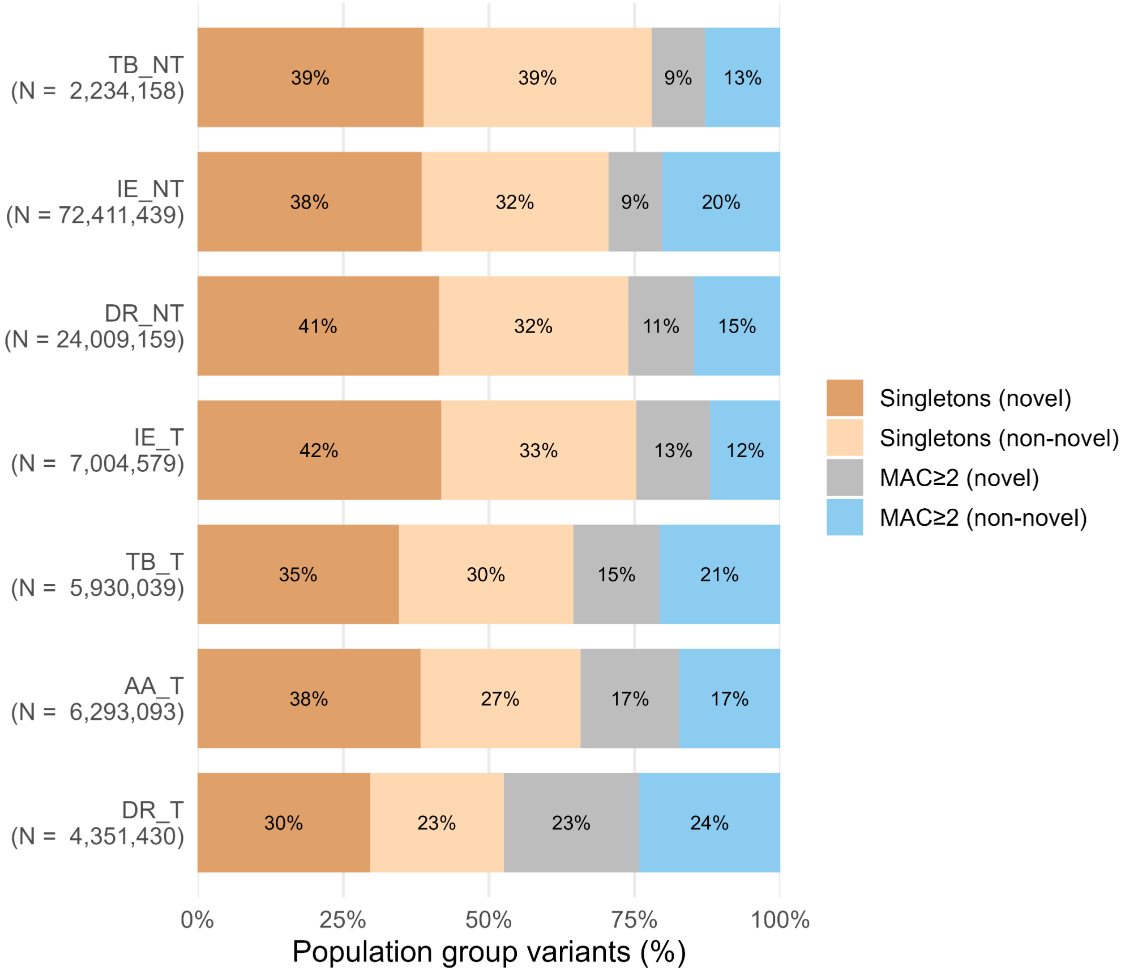

**Figure S3.2.1.** Population-group specific variant composition (%). Bars show the group-specific variant percentages of four categories: Singletons (novel), Singletons (non-novel), Non-Singletons (novel), and Non-Singletons (non-novel). Note: The total number of variants for each population group is provided below each bar plot.

**Population-specific variants identified across multiple ethnolinguistic groups**

For this analysis, we analyzed 9,636 individuals (excluding PCA outliers and the members of the CAO group) from the 82 ethnolinguistic backgrounds. Population-specific variants were defined as variants present exclusively within the individuals of a given ethnolinguistic group and not present in individuals from others. This distribution for the non-singleton population-specific variants is depicted in Figure S3.2.2.

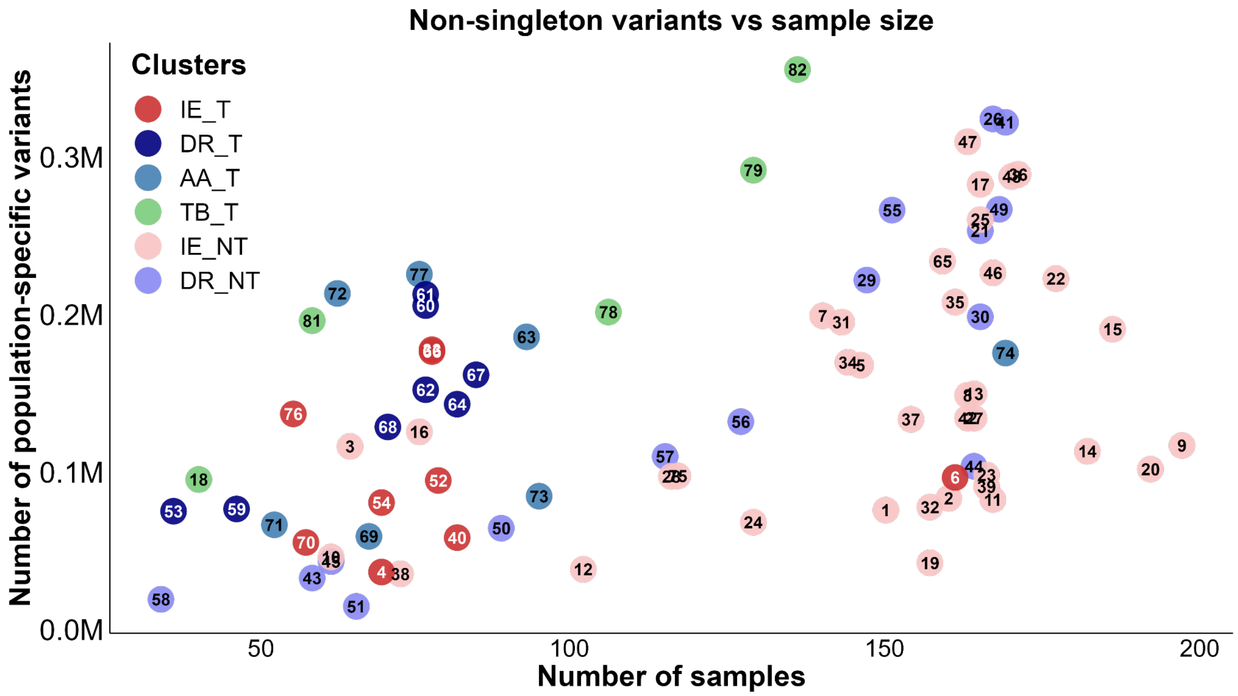

**Fig 3.2.2.** Relationship between the number of samples from a population and the number of non-singleton population-specific variants.

The above figure demonstrates that a higher sample size does not necessarily translate into a greater discovery of population-specific variants. The number of population-specific non-singleton variants increases with sample size for most IE_NT and DR_NT populations, reflecting expected discovery effects. However, at comparable sample sizes, some Austroasiatic, Dravidian, and Tibeto-Burman tribal populations harbor higher numbers of population-specific non-singleton variants than non-tribal populations. Moreover, heterogeneity for the number of population-specific variants is evident even among populations within the same linguistic cluster, indicating that extent of isolation, in addition to linguistic affiliation, could shape the accumulation of population-specific variation.

##### S3.3. $F_{ST}$ analysis

We measured the pairwise genetic distances of the study populations using the $F_{ST}$ statistic (Weir & Cockerham, 1984). We computed the pairwise $F_{ST}$ values using the EIGENSOFT software (Patterson et al., 2006; Price et al., 2006), considering a precision of up to 6 decimal points. We generated a heatmap and dendrogram based on these values. The dendrogram consisted of two apparent clusters. One of the clusters contained majorly the Tibeto-Burman (TB) speakers in one subcluster, and three Dravidian (DR) tribal populations from Nilgiri hills and the Western Ghats in another subcluster (the within cluster $F_{ST}$ values are also substantial for this cluster). The second cluster had two broad subclusters, one comprising mainly the tribal populations from Austro-Asiatic (AA), Dravidian (DR), and Indo-European (IE) linguistic families, with the other subcluster comprising mainly the Indo-European (IE) and Dravidian (DR) speaking non-tribal populations. Thus, we observe a distinction in the tribal and non-tribal populations in the $F_{ST}$ analysis.

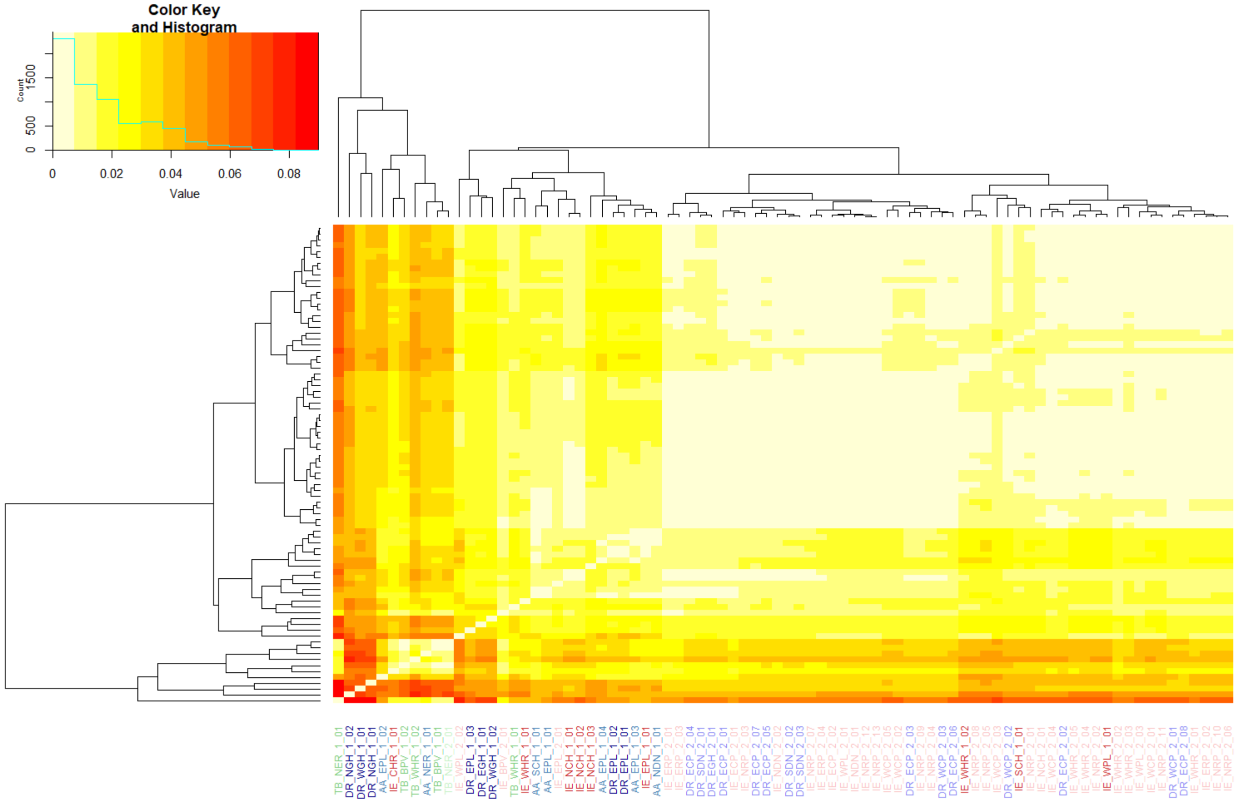

**Fig S3.3.1.** Heatmap of $F_{ST}$ statistics for 82 populations (except CAO) along with the dendrogram. The colors of the population labels represent corresponding linguistic families and Tribe/Non-Tribe affiliations.

To understand the general pattern of genetic distances among the four linguistic families (i.e. the Austro-Asiatic, Dravidian, Indo-European, and Tibeto-Burman), we grouped the populations based on their linguistic families and Tribe/Non-Tribe affiliations and computed pairwise $F_{ST}$ statistics. We observed that the Tibeto-Burman populations were more differentiated from populations belonging to other linguistic families. The Indo-European and Dravidian non-Tribe populations clustered together in the dendrogram based on the $F_{ST}$ values, whereas the Austro-Asiatic tribes shared greater genetic affinity with the Dravidian tribes, compared to other groups.

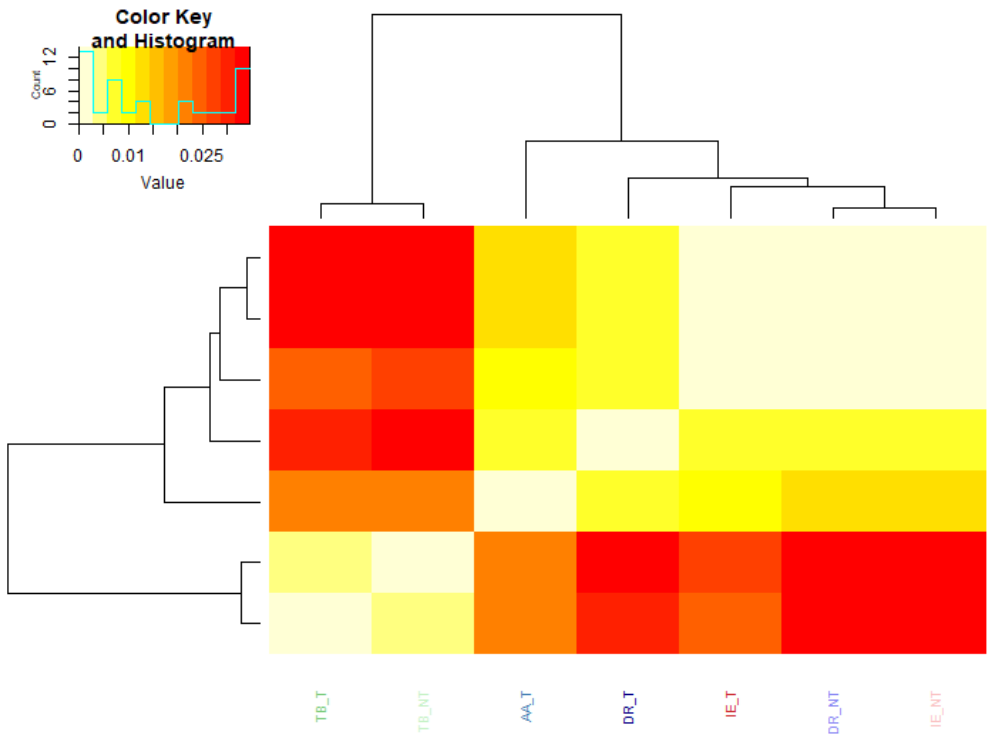

**Fig S3.3.2.** Heatmap of $F_{ST}$ statistics for populations aggregated by linguistic families and Tribe/Non-Tribe affiliations, along with the dendrogram.

**Table S3.3.1.** $F_{ST}$ matrix for populations aggregated by linguistic families and Tribe/Non-Tribe affiliations

| **Populations** | **AA_T** | **IE_T** | **IE_NT** | **DR_NT** | **DR_T** | **TB_T** | **TB_NT** |
|---|---|---|---|---|---|---|---|
| **AA_T** | 0 | 0.008787 | 0.013582 | 0.012072 | 0.007379 | 0.020141 | 0.022014 |
| **IE_T** | 0.008787 | 0 | 0.001642 | 0.001945 | 0.005848 | 0.025379 | 0.026819 |
| **IE_NT** | 0.013582 | 0.001642 | 0 | 0.001303 | 0.008236 | 0.031697 | 0.033117 |
| **DR_NT** | 0.012072 | 0.001945 | 0.001303 | 0 | 0.006081 | 0.033195 | 0.034524 |
| **DR_T** | 0.007379 | 0.005848 | 0.008236 | 0.006081 | 0 | 0.030375 | 0.031884 |
| **TB_T** | 0.020141 | 0.025379 | 0.031697 | 0.033195 | 0.030375 | 0 | 0.002951 |
| **TB_NT** | 0.022014 | 0.026819 | 0.033117 | 0.034524 | 0.031884 | 0.002951 | 0 |

**$F_{ST}$ analysis (within the four linguistic families)**

*Austro-Asiatic (AA)*
We also observed $F_{ST}$ distances within the linguistic families. Among the Austro-Asiatic speakers, the Munda-speaking populations cluster together, whereas AA_NER_1_01 which is geographically isolated (North East India) and belongs to the Mon-Khmer sub-family of AA speakers was separated and was prominently differentiated compared to other populations. (We have observed a similar pattern in the subsequent analysis on population structure, mentioned in section S4.)

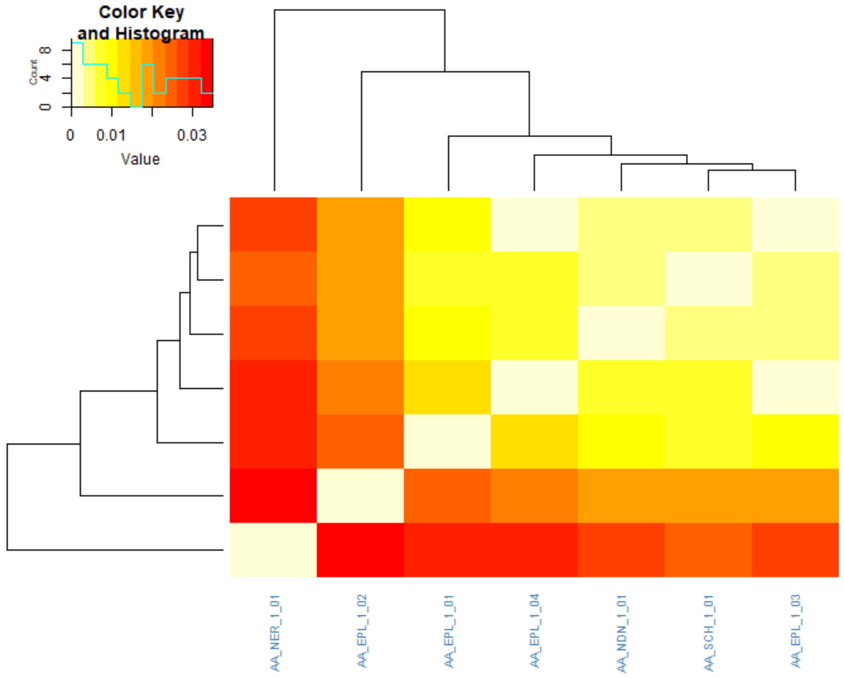

**Fig S3.3.3.** Heatmap of $F_{ST}$ statistics for **Austro-Asiatic (AA)** populations along with the dendrogram.

**Table S3.3.2.** $F_{ST}$ matrix for populations from the **Austro-Asiatic (AA)** linguistic family

| **Populations** | **AA_EPL_1_01** | **AA_EPL_1_02** | **AA_EPL_1_03** | **AA_EPL_1_04** | **AA_NDN_1_01** | **AA_NER_1_01** | **AA_SCH_1_01** |
|---|---|---|---|---|---|---|---|
| **AA_EPL_1_01** | 0 | 0.024561 | 0.009499 | 0.011939 | 0.009867 | 0.031999 | 0.007012 |
| **AA_EPL_1_02** | 0.024561 | 0 | 0.017714 | 0.021819 | 0.019343 | 0.035077 | 0.018005 |
| **AA_EPL_1_03** | 0.009499 | 0.017714 | 0 | 0.002722 | 0.005565 | 0.026729 | 0.003971 |
| **AA_EPL_1_04** | 0.011939 | 0.021819 | 0.002722 | 0 | 0.007736 | 0.030606 | 0.006002 |
| **AA_NDN_1_01** | 0.009867 | 0.019343 | 0.005565 | 0.007736 | 0 | 0.027103 | 0.004162 |

| | | | | | | | |
|---|---|---|---|---|---|---|---|
| **AA_NER_1_01** | 0.031999 | 0.035077 | 0.026729 | 0.030606 | 0.027103 | 0 | 0.025474 |
| **AA_SCH_1_01** | 0.007012 | 0.018005 | 0.003971 | 0.006002 | 0.004162 | 0.025474 | 0 |

*Dravidian (DR)*

The $F_{ST}$ analysis considering only the Dravidian speaking populations revealed separate clusters comprising the Tribe and Non-Tribe populations in the corresponding dendrogram. Three DR tribal populations, which are geographical isolates, show distinct clustering with deep branching. (We have observed a similar pattern in the subsequent analysis on population structure, mentioned in section S4.)

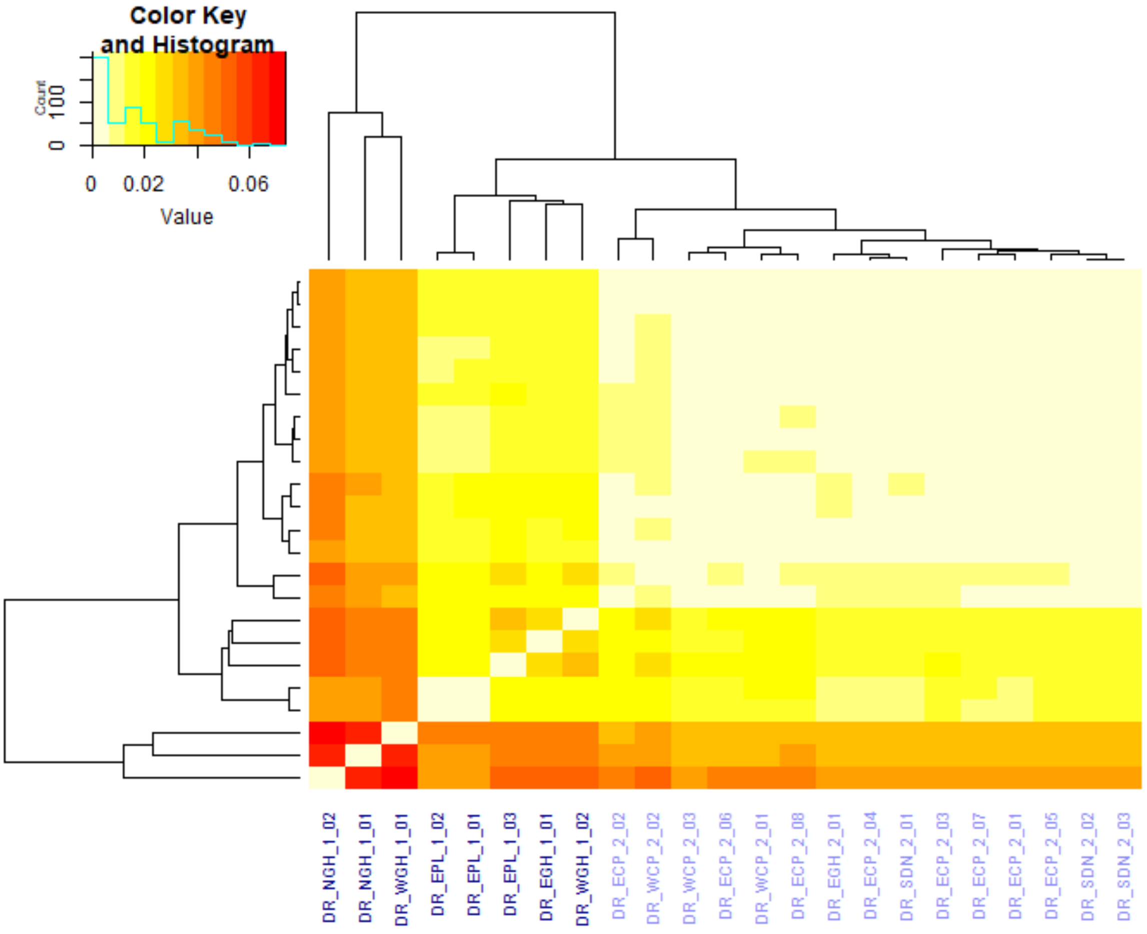

**Fig S3.3.4.** Heatmap of $F_{ST}$ statistics for **Dravidian (DR)** populations along with the dendrogram.

*Indo-European (IE)*

The FST analysis on Indo-European populations showed that IE_CHR_1_01 was highly differentiated from other populations. The dendrogram also showed multiple clusters with one prominent cluster consisting of the tribal populations: IE_EPL_1_01, IE_NCH_1_03, and IE_WHR_1_01. In general, the tribal populations tend to cluster together.

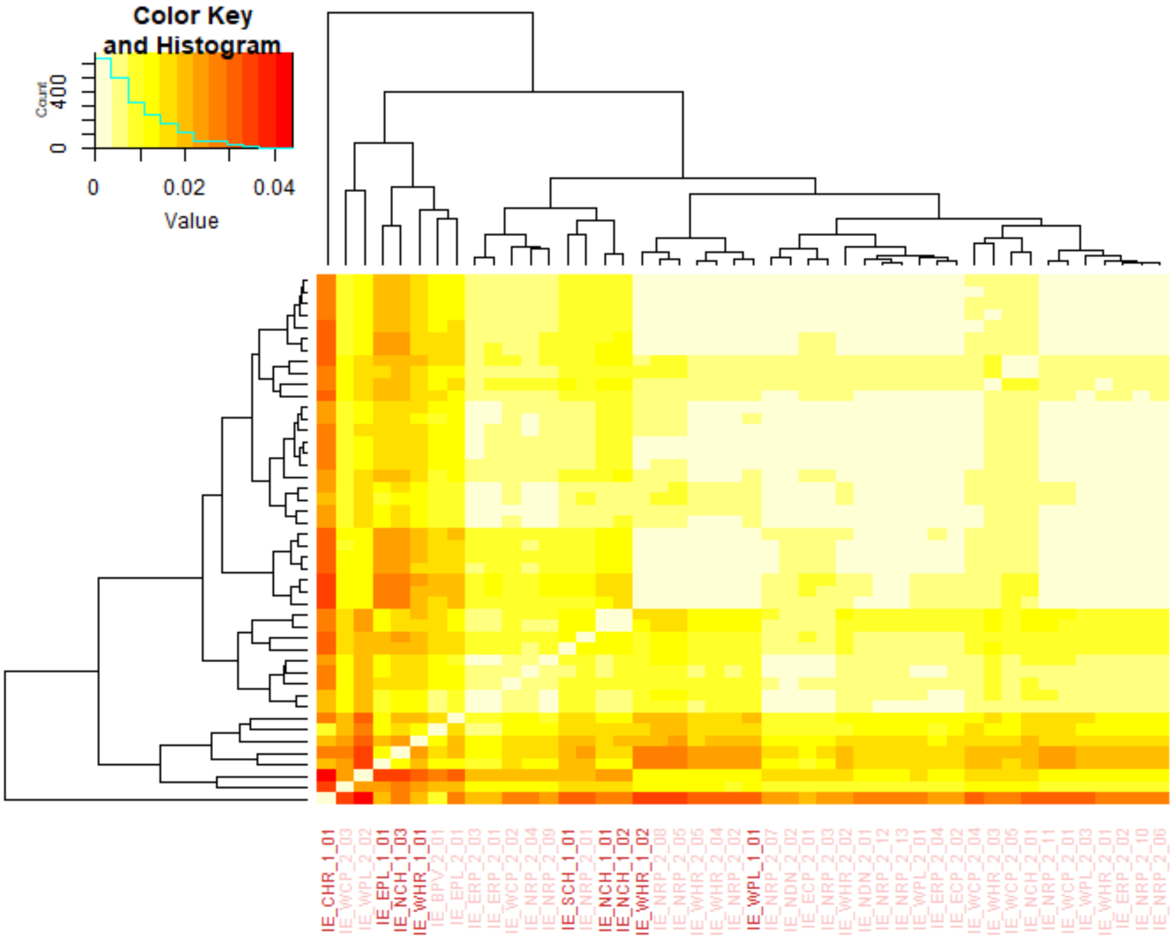

**Fig S3.3.5.** Heatmap of $F_{ST}$ statistics for **Indo-European (IE)** populations along with the dendrogram.

*Tibeto-Burman (TB)*

The $F_{ST}$ analysis on Tibeto-Burman (TB) speakers revealed that the populations from Brahmaputra Valley share more genetic affinity compared to other Tibeto-Burman populations. One TB speaking population from Western Himalayas (TB_WHR_1_01), which is more like a population isolate, is separated from the others. Whereas the other population from Western Himalayas (TB_WHR_1_02), which is significantly spread over a wide geographical space along the foothills of the Himalayas, clusters with other TB populations which are geographically from North East India.

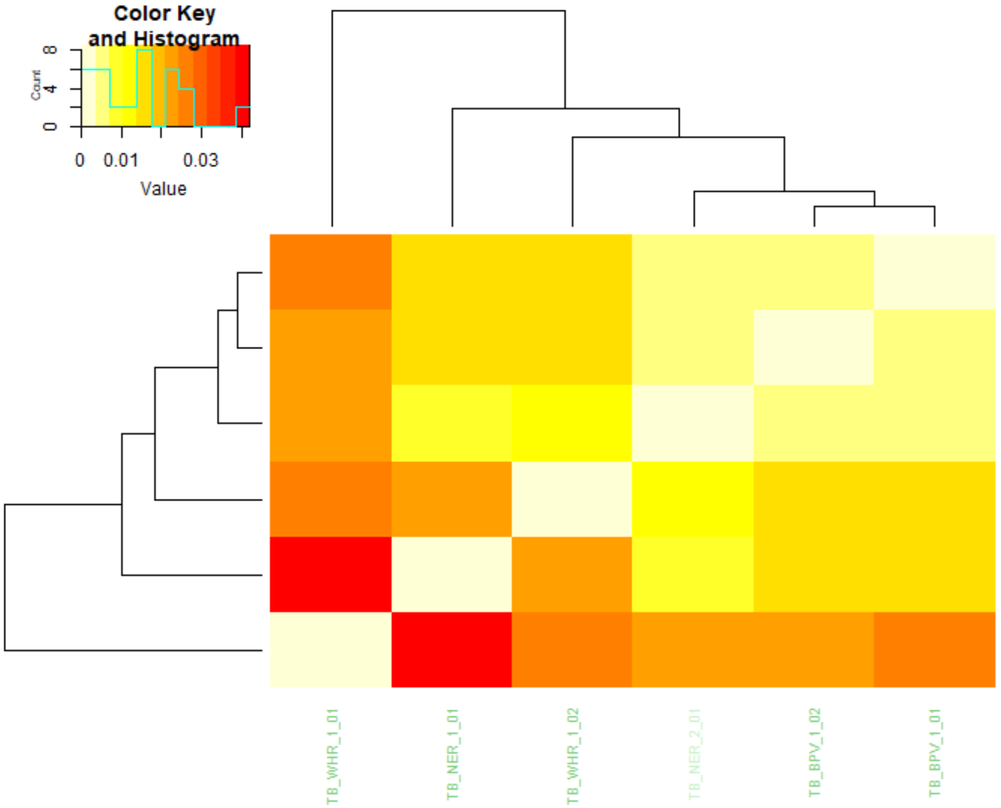

**Fig S3.3.6.** Heatmap of $F_{ST}$ statistics for **Tibeto-Burman (TB)** populations along with the dendrogram.

**Table S3.3.3.** $F_{ST}$ matrix for populations from the **Tibeto-Burman (TB)** Linguistic family

| **Populations** | **TB_BPV_1_01** | **TB_BPV_1_02** | **TB_NER_1_01** | **TB_NER_2_01** | **TB_WHR_1_01** | **TB_WHR_1_02** |
|---|---|---|---|---|---|---|
| **TB_BPV_1_01** | 0 | 0.003512 | 0.014851 | 0.005958 | 0.025107 | 0.01514 |
| **TB_BPV_1_02** | 0.003512 | 0 | 0.015204 | 0.005574 | 0.022445 | 0.014684 |
| **TB_NER_1_01** | 0.014851 | 0.015204 | 0 | 0.010178 | 0.041919 | 0.021019 |
| **TB_NER_2_01** | 0.005958 | 0.005574 | 0.010178 | 0 | 0.023869 | 0.012075 |
| **TB_WHR_1_01** | 0.025107 | 0.022445 | 0.041919 | 0.023869 | 0 | 0.025661 |
| **TB_WHR_1_02** | 0.01514 | 0.014684 | 0.021019 | 0.012075 | 0.025661 | 0 |

##### S3.4.1 Allele frequency divergence of shared variants across Indian population clusters

We conducted analysis of ~9 million genetic variants that are shared across all seven major ethnolinguistic groups in the GenomeIndia (GI) cohort: IE_Tribe, IE_NonTribe, DR_Tribe, DR_NonTribe, AA_Tribe, TB_Tribe, and TB_NonTribe. To explore population-level differences in allele frequency, we classified these shared variants based on whether their minor allele frequency (MAF) exceeded 5% within each ethnolinguistic cluster (Figure S3.4.1).

Variants classified as rare (MAF < 5%) in Indo-European (IE) and Dravidian (DR) groups often are common in Tibeto-Burman (TB) and Austroasiatic (AA) populations. Specifically, over 13% of IE/DR-specific rare variants are common in TB_T and TB_NT groups. Conversely, a substantial fraction of these TB_T rare variants, over 20%, are common in IE_NT, DR_NT, and IE_T groups. This reciprocal pattern is consistent with the existing models of population history of the Indian subcontinent where the TB and the AA populations typically were found to be relatively distant from the IE_NT and DR_NT populations (Basu et al., 2016). Similar patterns are observed even when restricting the analysis to variants with MAF ≥ 5%.

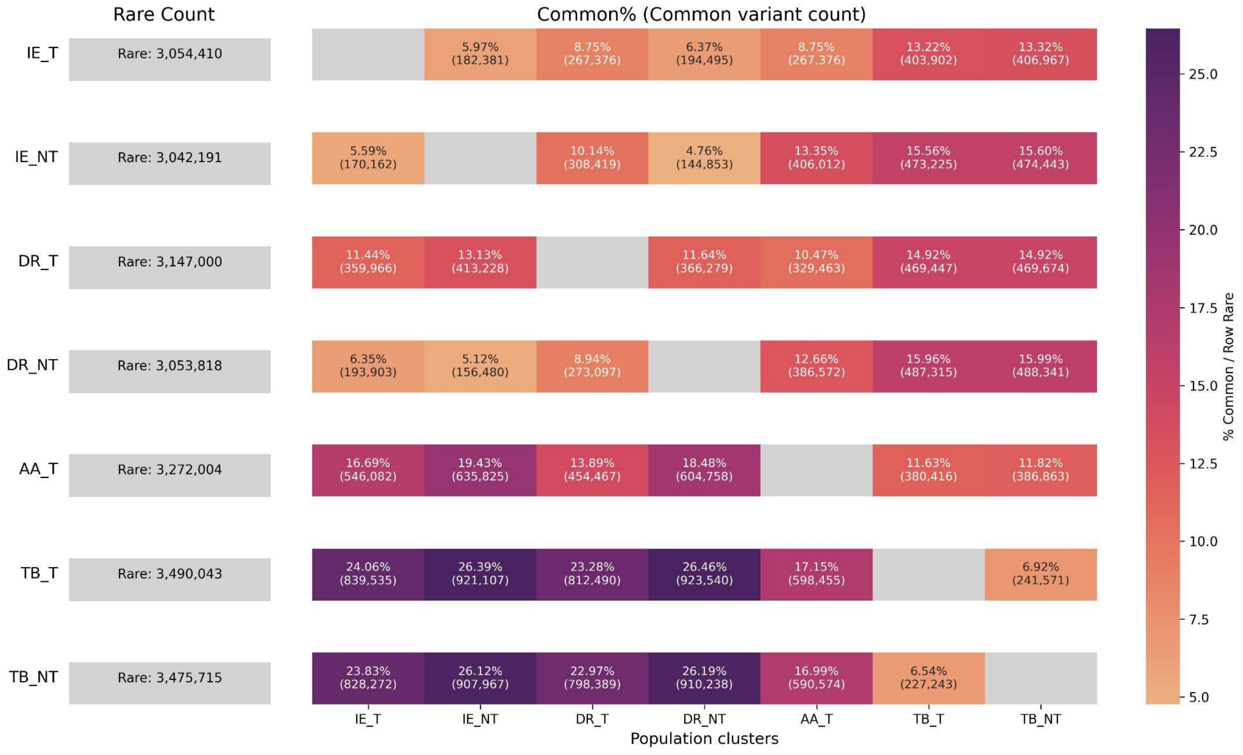

**Figure S3.4.1.** Genetic variants shared across seven major ethnolinguistic groups in the GenomeIndia cohort: IE_T, IE_NT, DR_T, DR_NT, AA_T, TB_T, and TB_NT. The left panel displays the total number of rare variants (MAF < 5%) observed within each cluster, while the gradient-colored boxes on the right visualize the proportion of those same rare variants that become common (MAF ≥ 5%) in other clusters. This framework enables the detection of allele frequency shifts across populations for shared variants.

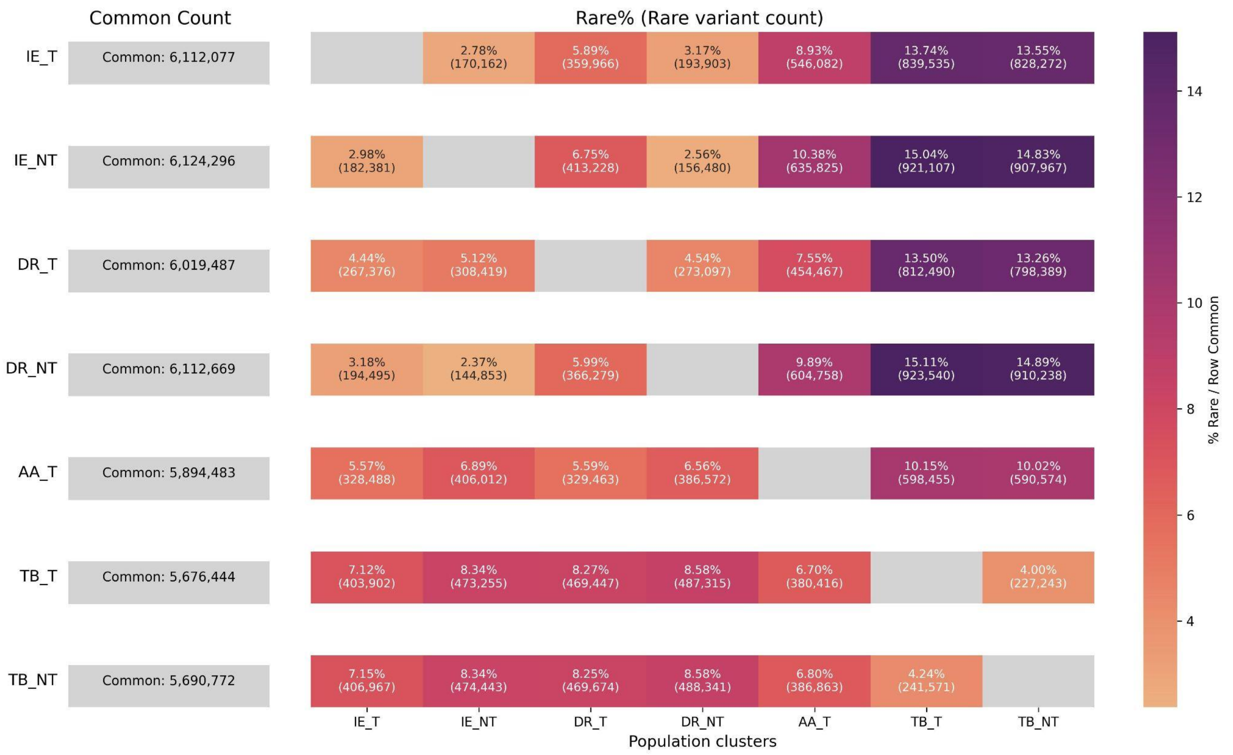

**Figure S3.4.2.** Genetic variants shared across seven major population clusters in the GenomeIndia cohort: IE_T, IE_NT, DR_T, DR_NT, AA_T, TB_T, and TB_NT. The left panel displays the total number of common variants (MAF ≥ 5%) observed within each cluster, while the gradient-colored boxes on the right visualize the proportion of those same common variants that become rare (MAF < 5%) in other clusters.

We also evaluated allele-frequency differences among the eight linguistic groups by computing pairwise Pearson correlations across common autosomal variants (Fig S3.4.3). The correlations are uniformly high, ranging from $r \approx 0.91$ up to $r \approx 0.996$ ($P < 0.001$ for all pairs). The strongest correlations are found among linguistically and geographically proximate groups, whereas comparisons involving groups with more ancient isolation or disparate linguistic affiliations (e.g., DR_NT vs TB_NT) yield slightly lower albeit still high values ($r \approx 0.913$). Taken together, this pattern demonstrates a remarkably consistent allele-frequency spectrum across diverse Indian sub-populations, reflecting a modest structure within the subcontinent populations (Kashyap et al., 2006). Nevertheless, the lower end of the range (≈0.91) indicates that some residual differentiation persists, retaining detectable fine-scale differentiation likely driven by longstanding endogamy, founder effects, and cultural or geographic isolation.

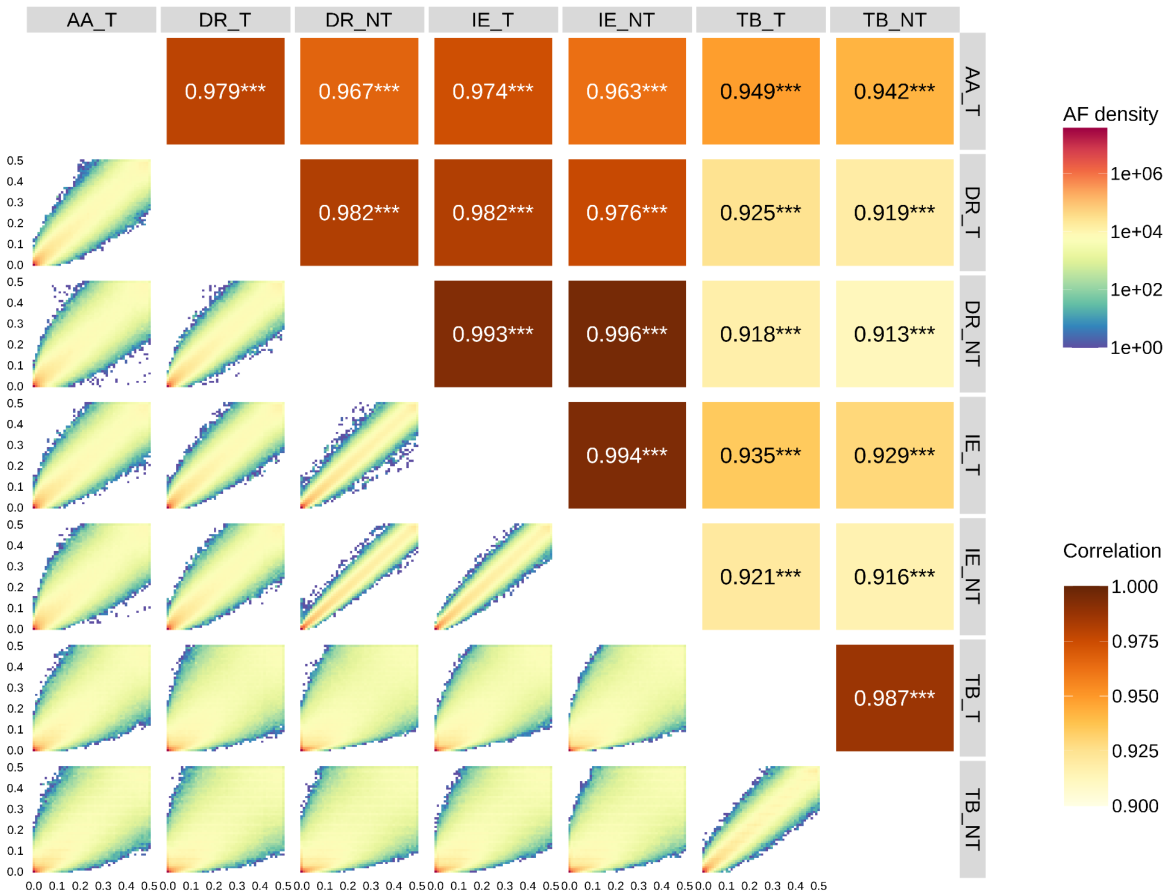

**Figure S3.4.3.** Correlation heatmaps of Minor Allele Frequencies (MAFs) across the seven ethnolinguistic groups. The x and the y-axis depict the frequencies. Each subplot highlights the correlation coefficient (Corr) between the paired groups, all marked as highly significant (***) and ranging between 0.913 to 0.996.

##### S3.4.2 Patterns of allele frequency variation across seven ethnolinguistic groups

For visualization of allele sharing between the 7 ethnolinguistic groups (CAO is not considered here), we have utilized GeoVar style plots (Biddanda et al., 2020). Variants are classified into 4 frequency-based classes - Common (>0.05), Low-Frequency (0.01 < x ≤ 0.05), Rare (0 < x ≤ 0.01) and Unobserved (0). Depending on which frequency class a variant fall into in each of the 7 groups, several categories are created as a combination of these frequency classes across the groups. Then these categories are ordered accordingly as per the variant count (Figure S3.4.4).

The plot is created on the basis of 67,315,559 variants where singleton variants, variants that were present only in the CAO are removed and the analysis was done only on the individuals who were not PCA outliers. 3114 combinations of the frequency classes, out of (47 - 1) possibilities across the 7 groups, are present in the data. The one with maximum variant count had 15,022,509 variants, which are present only in Indo-European non-tribes population with a frequency <= 1%.

As it can be observed, Indo-European non-tribes and Dravidian non-tribes have extensive sharing of alleles (~6 Million variants are rare and present in both groups) while ~4.9 Million variants in our study dataset are present across all 7 groups, with a frequency >5%. This set of ~4.9 Million variants, which are common across all the ethnolinguistic groups, are candidates which can be used to design informative genotype chips for Indian and South Asian populations. These variants can also complement as ancestry informative marker supplement for global genotype arrays. However, allele sharing counts can be biased due to unequal sample sizes (sample sizes are mentioned in parentheses beside the group labels at the bottom of the plot).

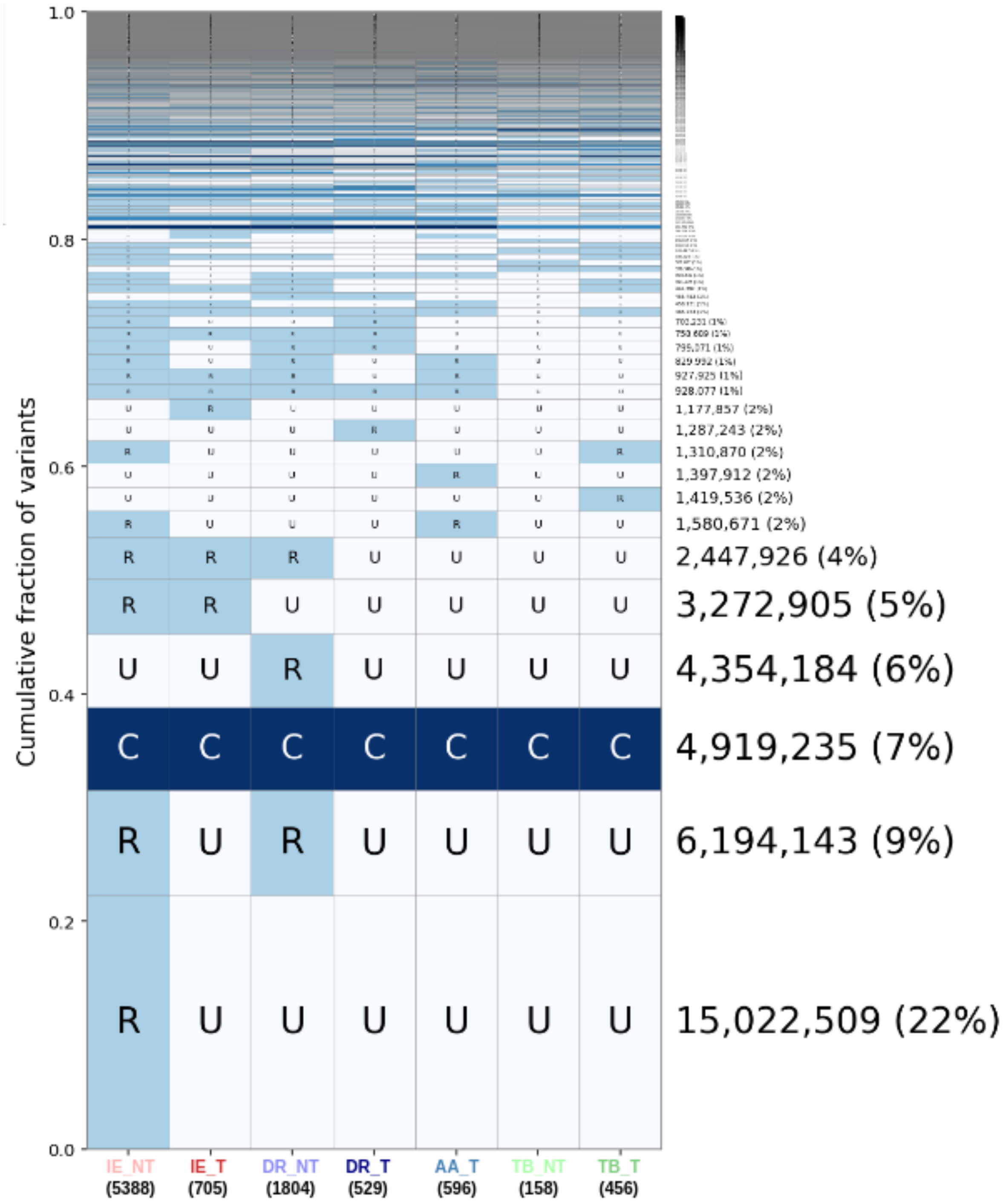

**Figure S3.4.4.** Geovar plot of 43,207,134 variants falling into the first eleven codes, explaining 64.19% of the total 67,315,558 variants considered. Numbers in parentheses indicate sample size of the group.

**Distribution of the common variants from GI in the global datasets**

We compared the frequency of ~4.9 Million common variants, which were at >5% frequency in all the 7 groups, with 1000Genomes (Auton et al., 2015) and gnomAD (Karczewski et al., 2020). Allele frequencies were obtained from the Ensembl variation database using Variant Effect Predictor (VEP) version 113.0.

**Table S3.4.1.** A comparison of the ~4.9 million variants present at >5% frequency across all ethnolinguistic groups in GenomeIndia (GI) with major global populations.

| Dataset | Common | Low-Frequency | Rare | Unobserved |
|---|---|---|---|---|
| 1000 genomes - overall | 4,668,346 (94.90%) | 47,550 (0.97%) | 537 (0.01%) | 202,802 (4.12%) |
| gnomAD - overall | 4,657,180 (94.67%) | 171,622 (3.49%) | 12,776 (0.26%) | 77,657 (1.58%) |

Compared to the 1000 genomes phase 3 original dataset, we see 84% of these variants to be common in the African superpopulation, which is the lowest. Followed by 89% in European, 91% in both American and East Asian superpopulations. Most overlap was, naturally, with South Asian superpopulation, where 96% of these variants were also common there. In all superpopulations, around 4% of these variants were unobserved (**Figure S3.4.5)**.

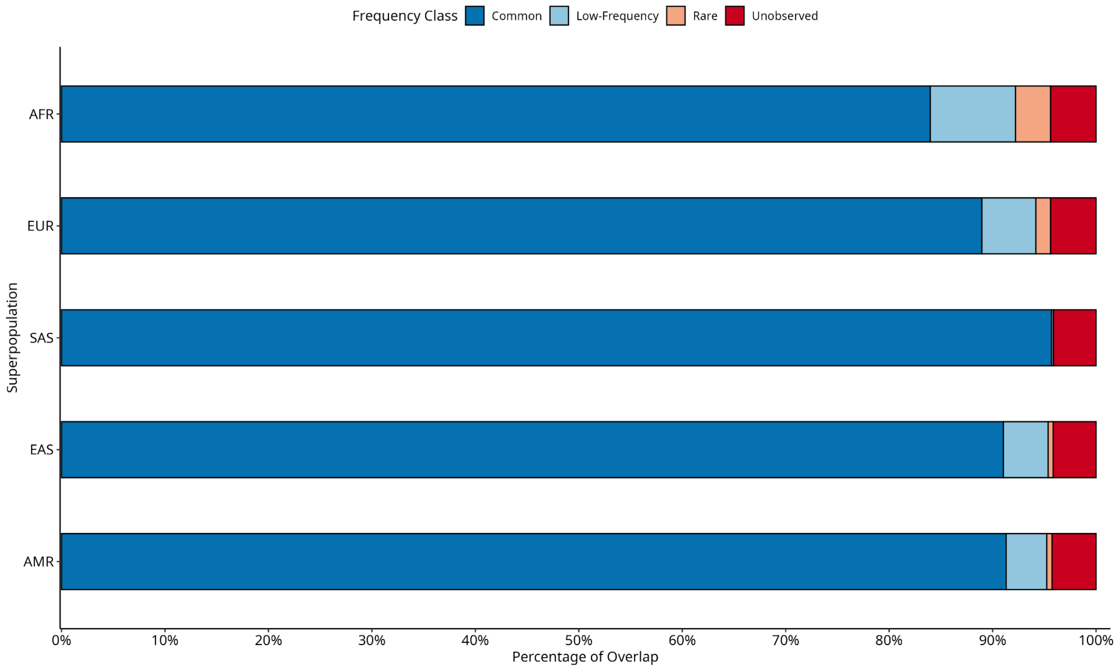

**Figure S3.4.5.** The distribution of variants common (AF ≥5%) in all the Genome India population groups, stratified by frequency bins across the 1000 genomes superpopulations

Compared to the gnomAD 4.1 dataset, there is a 7% increase in the amount of variants that fall into the common class in African superpopulation (Figure S3.4.6**)**. Other superpopulations comparable with the 1000 genomes dataset also showed 2-3% increase in this overlap. This is possibly due to higher sequencing coverage in gnomAD compared to very low sequencing coverage in the 1000 genomes phase 3 original dataset (Auton et al., 2015). For these superpopulations, around only 1.6% variants fall into unobserved class, which also supports this conclusion. Among the newly introduced superpopulations, the Middle Eastern had the highest common overlap (92%) and the Amish had the lowest (88%).

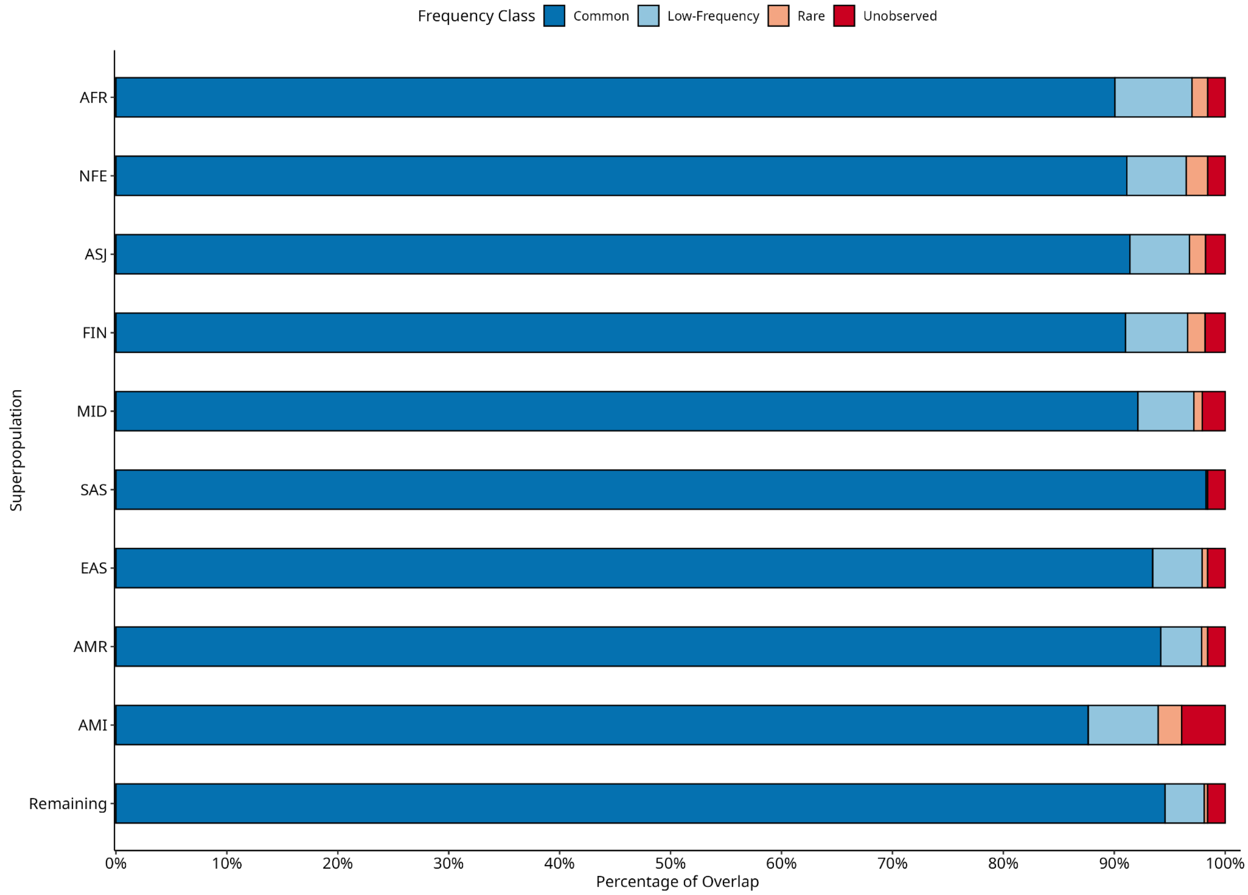

**Figure S3.4.6.** The distribution of variants common (AF ≥5%) in all the GenomeIndia population groups, stratified by frequency bins across the gnomAD superpopulations

When compared with the four GenomeAsia (Wall et al., 2019) superpopulations, the sharing was much less (Figure S3.4.7). South Asian superpopulation had the highest sharing (83.5%), followed by South-East Asian (81%), North-East Asian (80%) and Oceanian (72%). However, they have a large number of variants (12-14%) in the unobserved class, rather than in other frequency classes. This is expected as GenomeAsia had around ~67 million variants (Wall et al., 2019), while the GenomeIndia dataset has 129 million variants.

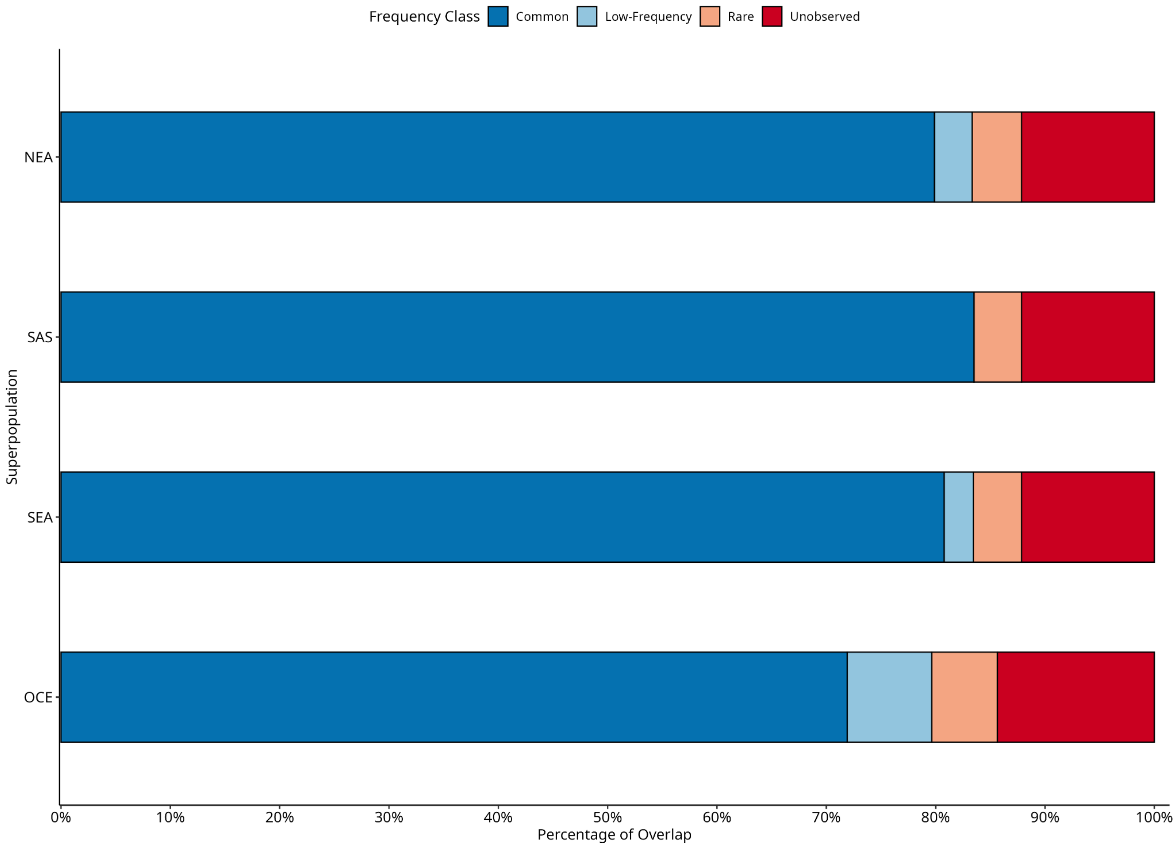

**Figure S3.4.7.** The distribution of variants common (AF ≥5%) in all the GenomeIndia population groups, stratified by frequency bins across the GenomeAsia superpopulations

**Distribution of group-specific common variants in the global datasets**

7,142,132 variants are found to be common (>0.05) in at least one of the 7 ethnolinguistic groups.

**Table S3.4.2.** A comparison of the ~7.1 million variants present at >5% frequency in at least one ethnolinguistic group in GenomeIndia (GI) with major global populations.

| Dataset | Common | Low-Frequency | Rare | Unobserved |
|---|---|---|---|---|
| 1000 genomes - overall | 5,888,779 (82.45%) | 776,199 (10.87%) | 161,430 (2.26%) | 315,724 (4.42%) |
| gnomAD - | 5,868,391 | 750,861 | 420,424 (5.89%) | 102,456 (1.43%) |

| | | |
|---|---|---|
| overall | (82.17%) | (10.51%) |

When compared at the superpopulation level, no notable deviation from the previously seen patterns were observed among the superpopulations themselves. However, in comparison with the earlier ~4.9 million variants, there is a decrease in the proportion of variants that are common, while proportions in all the other classes increased. This observation held true for all the superpopulations we earlier compared. This change can also be broadly seen at the overall frequency level when going from Table S3.4.1 to Table S3.4.2. This is expected as these variants are already showing variability in the GenomeIndia dataset itself, where it might be common in only one or just a few groups.

##### S3.4.3 Pattern of Variant Sharing between populations

**Jaccard Index:**

To quantify genetic similarity between populations, we employed the Jaccard index, defined as the ratio of number of shared genetic variants to the total number of unique variants identified across a given pair of population clusters (Jaccard P. (1908), Prokopenko et al., 2016). In order to mitigate potential biases introduced by unequal sample sizes, we implemented a random downsampling procedure, whereby the larger of each pair of clusters was subsampled to match the sample size of the smaller cluster. This standardization ensured that each cluster contributed an equal number of individuals to the analysis, thereby enabling a more equitable comparison of variant distributions.

$$Jaccard\ Index\ = |\ A \cap B\ |\ /\ |\ A \cup B\ |$$

A = variants present in cluster1
B = variants present in cluster2
$A \cap B$ = shared variants
$A \cup B$ = union of variants

We conducted three independent replicates of the random downsampling process for each pairwise comparison to ensure the robustness of this approach. Across all pairwise combinations, we observed minimal variation in the resulting Jaccard index values between replicates, suggesting that our estimates of genetic similarity were stable and not unduly influenced by stochastic effects introduced by the sampling process. All detected variants from the downsampled datasets were included in the final similarity calculations.

**Table S3.4.3.** Pairwise genetic similarity between population clusters based on Jaccard index. Minimum and maximum numbers of shared and union variants, together with the average Jaccard index, are reported for each pair of clusters across three independent random sampling replicates.

| Cluster 1 | Cluster 2 | min Shared | max Shared | min Union | max Union | Avg Jaccard Index | Sd Jaccard Index |
|---|---|---|---|---|---|---|---|
| TB_T | TB_NT | 10337492 | 10381201 | 17434457 | 17609153 | 0.59105 | 0.0014 |
| IE_NT | IE_T | 18095157 | 18108993 | 35615513 | 35630445 | 0.50809 | 0.0003 |
| AA_T | DR_T | 13042811 | 13050089 | 26240872 | 26299870 | 0.49657 | 0.0003 |
| AA_T | TB_NT | 9410562 | 9498494 | 19348973 | 19364006 | 0.48868 | 0.0004 |
| DR_NT | DR_T | 13560808 | 13592819 | 28161383 | 28241138 | 0.48127 | 0.0017 |
| IE_T | TB_NT | 9666111 | 9724952 | 20154262 | 20234305 | 0.48009 | 0.0010 |
| AA_T | IE_T | 14777741 | 14839727 | 30846996 | 30917028 | 0.47974 | 0.0013 |
| IE_NT | DR_NT | 23320920 | 23392668 | 48728841 | 48870834 | 0.47865 | 0.0003 |
| IE_T | DR_NT | 16367770 | 16375697 | 34522206 | 34575242 | 0.47379 | 0.0009 |
| IE_NT | TB_NT | 9458192 | 9566332 | 20270951 | 20313611 | 0.46918 | 0.0008 |
| DR_T | TB_NT | 8899589 | 8918030 | 19064704 | 19132662 | 0.46657 | 0.0025 |
| DR_T | IE_T | 13386928 | 13395461 | 28722953 | 28830530 | 0.46526 | 0.0007 |
| DR_NT | TB_NT | 9091739 | 9101805 | 20061788 | 20143955 | 0.45249 | 0.0002 |
| AA_T | TB_T | 11921981 | 11971631 | 26595880 | 26684783 | 0.44846 | 0.0002 |
| AA_T | IE_NT | 14672323 | 14718337 | 33064767 | 33171991 | 0.44387 | 0.0002 |
| AA_T | DR_NT | 13867280 | 13909431 | 31527768 | 31570058 | 0.44048 | 0.0006 |
| DR_T | IE_NT | 13360947 | 13380964 | 30451701 | 30584281 | 0.43825 | 0.0003 |
| IE_T | TB_T | 12501312 | 12515761 | 28620452 | 28693426 | 0.43651 | 0.0007 |

| Cluster 1 | Cluster 2 | min Shared | max Shared | min Union | max Union | Avg Jaccard Index | Sd Jaccard Index |
|---|---|---|---|---|---|---|---|
| DR_T | TB_T | 10769842 | 10798978 | 25809174 | 25878705 | 0.41733 | 0.0029 |
| IE_NT | TB_T | 12380933 | 12551463 | 29968381 | 30074994 | 0.41576 | 0.0001 |
| DR_NT | TB_T | 11372417 | 11410484 | 29233304 | 29268946 | 0.38932 | 0.0004 |

#### Supplementary Section S4: Population structure

Devashish Tripathi[1,2], Vinay More[1,2], Arghya Dey[1], Haya Afreen[1,2], Shouvanik Sengupta[1,2], Chandrika Bhattacharyya[1], Analabha Basu[1,2]

[1]BRIC - National Institute of Biomedical Genomics (BRIC-NIBMG), Kolkata, India. [2]Regional Centre for Biotechnology (RCB), Faridabad, India.

#### Summary

This section elaborates the genetic structure of India, inferred from the 82 mainland Indian populations and one continentally admixed population using autosomal genetic data. We analyze the GenomeIndia dataset at multiple scales. We highlight our findings from two major analyses: Principal Component Analysis (PCA), ADMIXTURE analysis. One prominent feature that emerges in the analysis of population structure in the extreme socio-culturally diverse Indian population is the uniformity and homogeneity within the defined populations. With long-standing endogamy, individuals from each population typically cluster together. PCA on the 82 Indian populations reveals a prominent North-South genetic cline, along with a newly reported tribal cline from Central and Eastern Plateau. We demonstrate a strong concordance between genetic structure and biogeography, with a linear model incorporating geography, language, and biogeography (Model 4) providing the best fit (Adjusted $R^2$ up to 0.844). The correlation with geography extends to neighbouring continental populations like Central South Asian, East Asians and South East Asians. Unsupervised clustering using ADMIXTURE identifies five main ancestral components—Ancestral North Indian (ANI), Ancestral South Indian (ASI), Ancestral Austro-Asiatic (AAA), Ancestral Tibeto-Burman (ATB), and a possible component from the Nilgiri Hills (ASI-D)—reinforcing the North-South cline and showing the spread of different ancestries across the subcontinent. Finally, the quantification of within and between-individual diversity identifies that individuals within a population, irrespective of their ancestry proportions, are relatively homogeneous. This homogeneity of ancestry proportions in individuals within a population serves as a support to the possible utilization of these populations in conducting large-scale genomic studies like GWAS in Indian populations. We identify five groups based on their population and individual level ancestry proportions, notably defining the 'melting pot' populations that exhibit high within-individual diversity and contributions from at least four ancestral components.

We have also used the PCA and ADMIXTURE to infer about the Continentally Admixed Outgroup (CAO) population, residing mainly along India's western and southern coasts. The CAO represents a population with relatively recent admixture between South Asian and African ancestral populations. Using supervised ADMIXTURE, PCA, and an expanded reference panel of Indian and African populations, we found that the CAO still carries substantial African ancestry; most closely related to the Luhya (LWK) of East Africa alongside notable ANI and ASI components. Their intermediate genetic position between Indian and African groups across multiple principal components further supports this admixture model. The >50% African ancestry, still present among the CAO, underlines the statement that the population has largely been an endogamous population where substantial African ancestry is still present.

##### S4.1 Principal Component Analysis

Quality control and data preparation was performed using PLINK v2.0 (Chang et al., 2015). The analysis focused on autosomal markers. From the initial set of 9768 individuals, we excluded the CAO (N = 50) population and offspring from trios (N = 243), resulting in a final dataset of 9,475 unrelated samples. As described in the GenomeAsia project (Wall et al., 2019) variants were filtered based on a Hardy-Weinberg Equilibrium (HWE) threshold of $p < 10^{-8}$, a missing genotype rate of < 0.02, and a Minor Allele Frequency (MAF) ≥ 0.01. Additionally, Linkage Disequilibrium (LD) pruning was applied using a window size of 50 SNPs, a step size of 5 SNPs, and an $r^2$ threshold of > 0.2. Following these steps, 1,253,493 variants were retained for analysis. To characterize the structure of the GI populations, we performed Principal Component Analysis (PCA) on this curated dataset using the smartpca software (version 18140) within the EIGENSOFT package (v8.0.0) (Patterson et al., 2006; Price et al., 2006).

###### S4.1.1 PCA of 82 anthropologically defined populations

The Principal component analysis of the individuals belonging to the 82 (excluding CAO and offsprings of trios) anthropologically defined populations revealed clustering by population. These results largely corroborate prior observations regarding the Indian populations (Reich et al., 2009; Basu et al., 2016; Wall et al., 2019) while offering considerably greater depth and detail.

In Figure S4.1, we observed a North-South cline that starts with the IE_NT in the upper left-hand corner and ends with the DR_T in the lower end. The other populations in this cline are admixed populations with decreasing IE-like ancestry as we move towards the lower end. This observation aligns with the earlier-reported north-south cline (Reich et al., 2009; Basu et al., 2016; Wall et al., 2019). However, we would like to highlight that population DR_NGH_1_02 is well-separated from the other DR populations in the North-South cline (E). We also observed that there are many populations whose genetic composition differs significantly from that in the North-South cline. These populations also exhibit high differentiation among themselves, with a large spread in PC space, compared to the diversity observed in the North-South cline.

We first examined a group of three populations (IE_WHR_2_03, IE_WHR_2_02, TB_WHR_1_01) from the western Himalayas, which reside among IE-speaking populations (A). In the PC space, these populations move away from the IE representative population towards TB-speaking populations. In particular, population TB_WHR_1_01 appears to have high TB-like ancestry, as indicated by a large spread along PC1, which suggests admixture with sufficient components from the two major groups, IE and TB.

We next examined a group of five populations (IE_WHR_1_01, IE_ERP_2_01, IE_ERP_2_03, IE_BPV_2_01, IE_CHR_1_01), which are IE-speaking populations, and fall in the center of PC space (C), indicating the genetic composition of these populations from the four groups: IE, DR, AA, and TB. Thus, we label these populations as the bridge populations. Populations IE_ERP_2_01 and IE_ERP_2_03 are from the Eastern Riverine Plain, population IE_WHR_1_01 is from the Western Himalayas, population IE_CHR_1_01 is from the Central Himalayas, and population IE_BPV_2_01 is from the Brahmaputra Valley. Given the geographic location of populations IE_WHR_1_01 and IE_CHR_1_01, these populations are expected to have low DR-like ancestry; however, we observed a contradiction. Populations IE_ERP_2_01, IE_ERP_2_03, and IE_BPV_2_01 are located along the geographic cline, extending from the Eastern riverine region to the northeast.

We find another major group of populations, AA_EPL_1_01, DR_EPL_1_03, IE_EPL_2_01, IE_NCH_1_03, DR_EPL_1_02, DR_EPL_1_01, AA_SCH_1_01, IE_EPL_1_01, AA_NDN_1_01, AA_EPL_1_02, AA_EPL_1_03, AA_EPL_1_04 forming a separate cluster, distant from the north-south cline (D). This group is dominated by tribal populations, except for IE_EPL_2_01, which is a non-tribal IE population. Populations IE_NCH_1_03, IE_EPL_1_01 are IE tribals, DR_EPL_1_03,DR_EPL_1_02,DR_EPL_1_01 are DR tribals, AA_EPL_1_01,AA_SCH_1_01, AA_NDN_1_01, AA_EPL_1_02, AA_EPL_1_03, AA_EPL_1_04 are AA tribals. The majority of the population resides on the Eastern Plateau and represents the DR-AA cline, which was not previously reported. Given the location of these populations in central India suggests that these populations should be an admixture of multiple groups; however, the observation from genetic data suggests these populations have primarily an AA-DR component, indicating that there could be admixture between ancestral AA and DR populations in the past, which has resulted in this cline. We observe some evidence from archaeobotany, where AA populations from the eastern part of India might have adapted some crops from DR populations (Fuller, 2007).

Another cluster we find of TB-speaking populations, except AA_NER_1_01, which is an AA-speaking population (B). All these populations are from the Northeast part of India, except TB_WHR_1_02, which resides in the western Himalayas and is representative of the TB populations. This suggests that there may be multiple routes of TB migration in India, one from the North-East and another through the North-West, along the route of the Tibetan Plateau.

Overall, we observe the geographic-genetic concordance among the Indian populations also reported in previous studies.

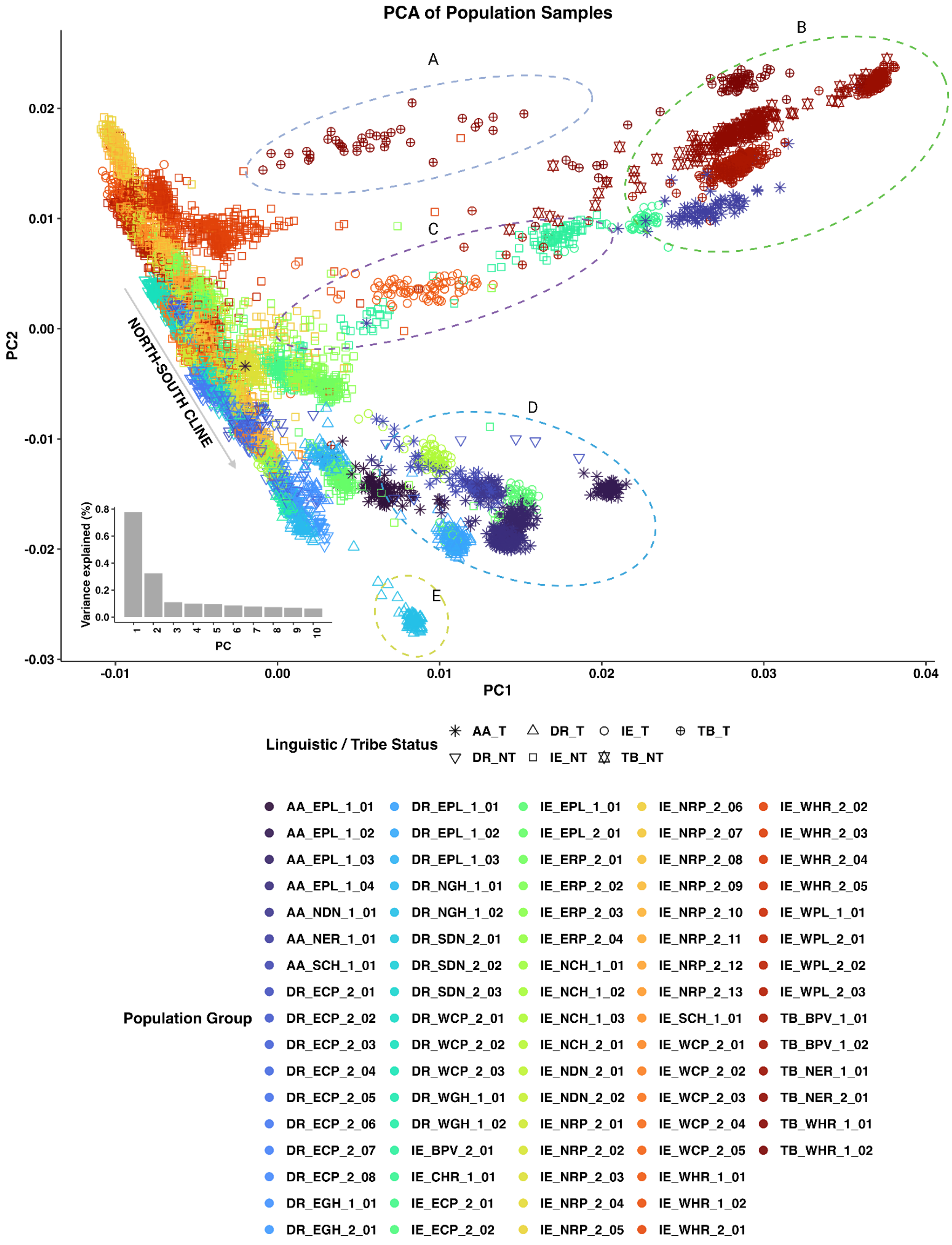
PCA of Population Samples
A
B
C
D
E
NORTH-SOUTH CLINE
PC1
PC2
0.02
0.01
0.00
-0.01
-0.02
-0.03
-0.01
0.00
0.01
0.02
0.03
0.04
Variance explained (%)
PC
Linguistic / Tribe Status
AA_T
DR_T
IE_T
TB_T
DR_NT
IE_NT
TB_NT
Population Group
AA_EPL_1_01
AA_EPL_1_02
AA_EPL_1_03
AA_EPL_1_04
AA_NDN_1_01
AA_NER_1_01
AA_SCH_1_01
DR_ECP_2_01
DR_ECP_2_02
DR_ECP_2_03
DR_ECP_2_04
DR_ECP_2_05
DR_ECP_2_06
DR_ECP_2_07
DR_ECP_2_08
DR_EGH_1_01
DR_EGH_2_01
DR_EPL_1_01
DR_EPL_1_02
DR_EPL_1_03
DR_NGH_1_01
DR_NGH_1_02
DR_SDN_2_01
DR_SDN_2_02
DR_SDN_2_03
DR_WCP_2_01
DR_WCP_2_02
DR_WCP_2_03
DR_WGH_1_01
DR_WGH_1_02
IE_BPV_2_01
IE_CHR_1_01
IE_ECP_2_01
IE_ECP_2_02
IE_EPL_1_01
IE_EPL_2_01
IE_ERP_2_01
IE_ERP_2_02
IE_ERP_2_03
IE_ERP_2_04
IE_NCH_1_01
IE_NCH_1_02
IE_NCH_1_03
IE_NCH_2_01
IE_NDN_2_01
IE_NDN_2_02
IE_NRP_2_01
IE_NRP_2_02
IE_NRP_2_03
IE_NRP_2_04
IE_NRP_2_05
IE_NRP_2_06
IE_NRP_2_07
IE_NRP_2_08
IE_NRP_2_09
IE_NRP_2_10
IE_NRP_2_11
IE_NRP_2_12
IE_NRP_2_13
IE_SCH_1_01
IE_WCP_2_01
IE_WCP_2_02
IE_WCP_2_03
IE_WCP_2_04
IE_WCP_2_05
IE_WHR_1_01
IE_WHR_1_02
IE_WHR_2_01
IE_WHR_2_02
IE_WHR_2_03
IE_WHR_2_04
IE_WHR_2_05
IE_WPL_1_01
IE_WPL_2_01
IE_WPL_2_02
IE_WPL_2_03
TB_BPV_1_01
TB_BPV_1_02
TB_NER_1_01
TB_NER_2_01
TB_WHR_1_01
TB_WHR_1_02

**Fig S4.1.** (a) The scatterplot of the first two eigenvectors of the PCA plot consisting of 9475 individuals from 82 populations from mainland India. X-axis represents PC1 and Y-axis represents PC2. Each population is colored by a unique color and has a shape based on the ethno-linguistic identity. (b) Inset - the proportion of variance explained by the first 10 principal components is shown in the bar plot. The PC components are in X-axis and the variation explained (%) in Y-axis

The PCA plots for each of the 82 populations, individually marked, on the PC1-PC2 background can be accessed in Supplementary note 1.

We assessed how known and potential sources of population structure influence genetic clustering. Language families have been one of the primary determinants of genetic clustering, particularly in Indian populations (Majumder & Basu, 2015). We have used that information and present that as Figure 2(a) in the main manuscript.

##### S4.1.2 PCA of 82 anthropologically defined populations and their relationship with biogeography

Biogeography has been a known determinant which facilitated and hindered population movement and spread (Li et al., 2008; Petkova et al., 2016). We plotted the PC coordinates of the individuals colored by their biogeographic labels to understand how well biogeography captures the genetic variation in GenomeIndia dataset. The PC1-PC2 space of the PCA plot mimicked the biogeographical distribution of these populations. We observed a North-South cline along the PC2 axis. The populations from Northern riverine plain, Western Himalayas and Western plains constituted the North end of the cline, whereas the population no. 61 residing in the Nilgiri hills constituted the South end. The eastern plateau populations, spread along the PC1 axis, form a cluster close to the North-South cline. The populations from Brahmaputra Valley and North Eastern range constitute a separate cluster from the North-South cline, corroborating with their presence in Northeast India. The Central Himalayan populations lie between the Northeast Indian populations and the North end of the North-South cline, again corroborating well with geography.

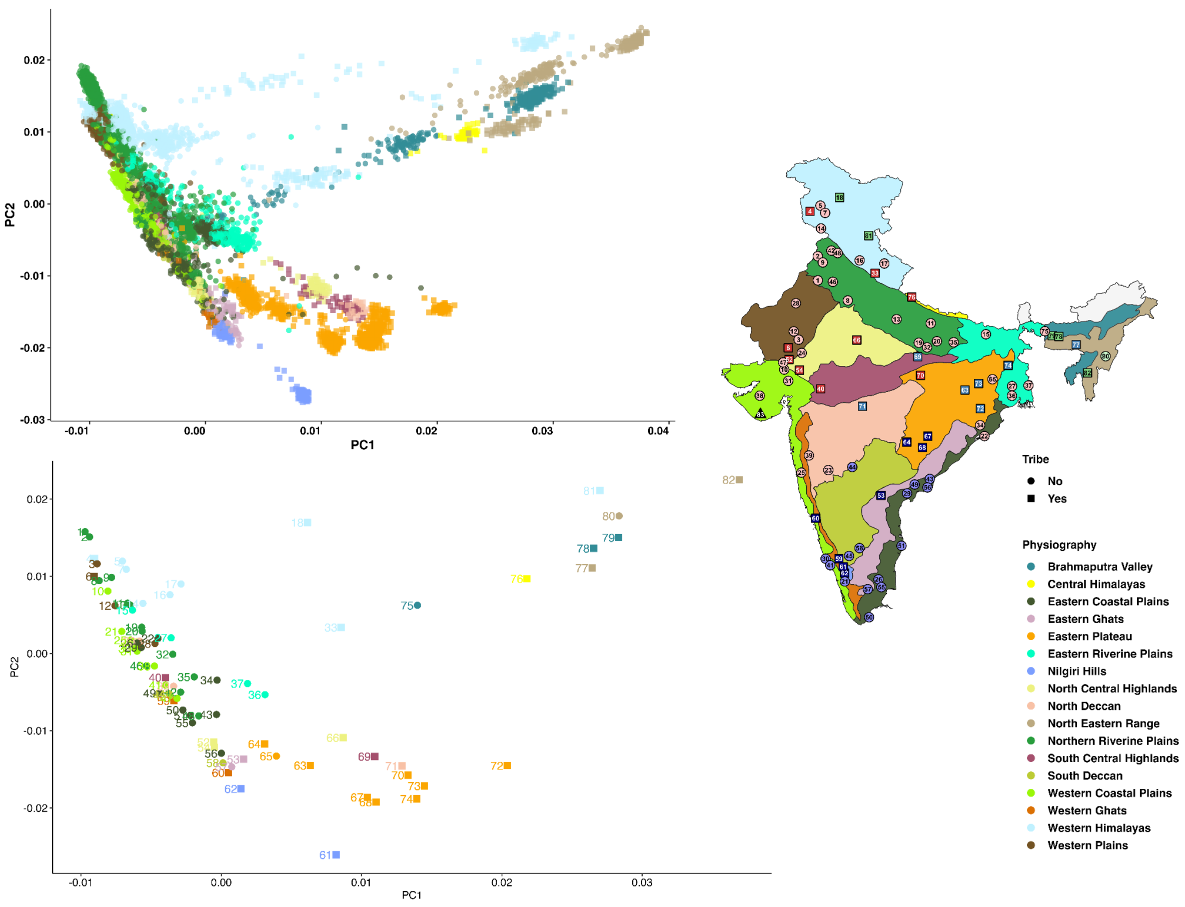

**Figure S4.2**. Top panel shows the Scatter plot of PC1-PC2 for 9475 samples from 82 population groups from mainland India, corroborating with their biogeographical presence. In the bottom panel, populations are depicted by their centroid (calculated from the first two eigenvector co-ordinates of all the individuals in that population) and are marked with their corresponding numeric code as mentioned in the Table S1.4.

#### Investigating the local population structure within each linguistic group

We have performed PCA for each of four linguistic families: Austro-Asiatic, Dravidian, Indo-European, and Tibeto-Burman to explore the population structure within each group and its concordance with the biogeography.

#### Population structure in Indo-European (IE) populations

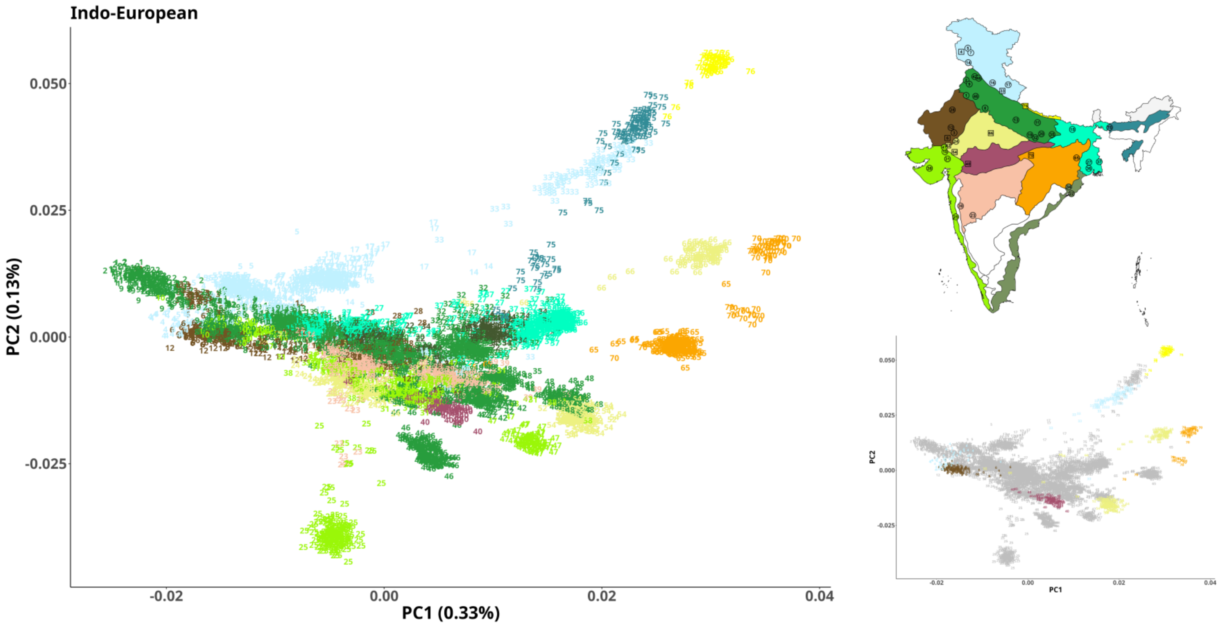

**Figure S4.3.** The scatter plot of PC1-PC2 of the Indo-European language speakers (6093), coloured according to their biogeographic location, with inset of biogeographical location of the populations along with their tribe and non-tribe status, where the non-tribal populations are all coloured in grey.

The IE language speakers are primarily non-tribal populations, both in census records as well as in our sample set. In the PCA analysis, PC1 and PC2 show a gradient pattern, indicating continuous gene flow or admixture across regions. This is overall consistent with previous findings that admixture among IE_NT groups is considerably more than tribals or DR_NT groups (Basu et al., 2016), as reflected in PC1. This reflected a North (Western Himalayas) - South (Eastern plains and North Deccan) gradient, while PC2 captured substructure along West (Western Coastal plains) - East (Brahmaputra valley). Populations from the same geographic region cluster together genetically.

The populations from the Northern riverine plain, Western Himalayas, and Western plains constituted the North end of the cline, whereas the populations residing in the North Central Highlands constituted the South end. The eastern plateau populations, spread along the PC1 axis, form a cluster close to the North-South cline. The populations from the Brahmaputra Valley, South Central Highlands, Eastern Riverine plains, and Central Himalayas do not constitute any cluster and are spread along PC2 forming east cline, which corroborates with their geography, albeit approximately. The Central Himalayan populations lie between the Northeast Indian populations and the North end of the North-South cline, again corroborating well with geography. Despite shared Indo-European language, there's strong genetic substructure—likely due to endogamy and/or geography. A non-tribal population from the western coastal plain (IE_WCP_2_03) appears distinct, possibly due to persisting endogamy, which also corroborated with their Runs of Homozygosity pattern (Section S7).

**Population structure in Dravidian (DR)**

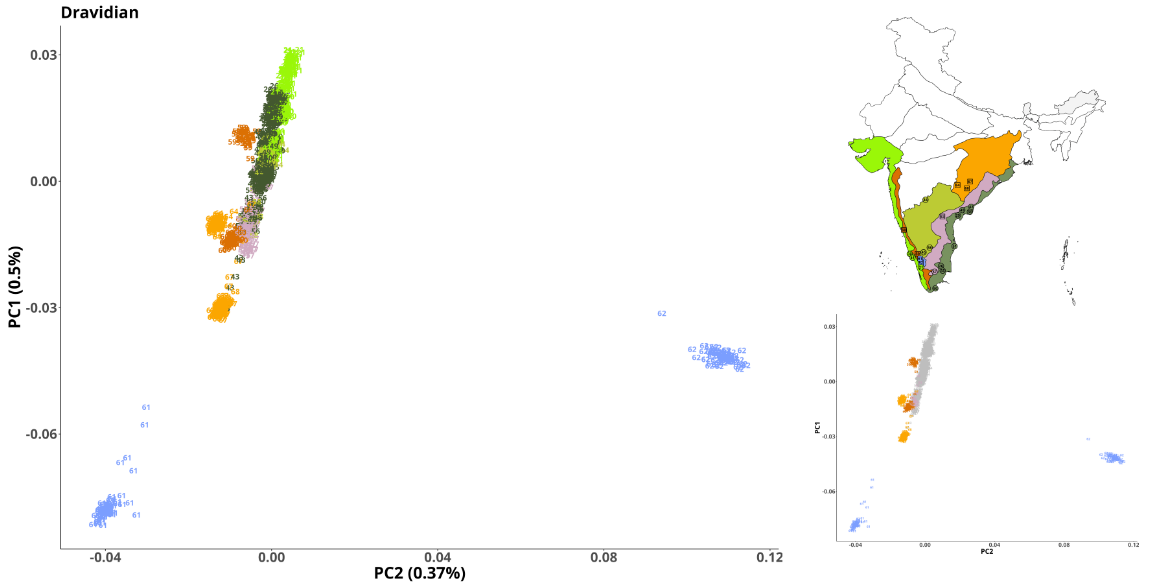

**Figure S4.4.** The scatter plot of PC1-PC2 of the Dravidian language speakers (2333), coloured according to their biogeographic location, with an inset of the biogeographical location of the populations along with their tribe vs. non-tribe status (grey).

Previous studies in genetics have indicated that some DR_NT populations are often genetically extremely similar to some IE_NT populations, whereas some DR_T populations are often genetically very close to AA populations (Basu et al., 2016). The large value of the Jaccard index, calculated in S3, on the combined pool of all DR_NT and IE_NT, as well as between DR_T and AA, also conforms to these observations. In the PCA on all DR speakers, we observe a North-South gradient along PC1. But the striking feature among the DR speakers is the isolation and separation of the tribal populations, particularly the two populations from Nilgiri hills (DR_NGH_1_01 and DR_NGH_1_02), appearing at the bottom left and right, which are genetically very distinct from the other Dravidian populations, recapitulating the observations in $F_{st}$ (S3). It is important to note here that these populations are from extreme geographical proximity and share the same biogeography. Long-term isolation from other populations, as well as each other is evident. It is also indicative that long term isolation and drift might have resulted in population-specific signatures. Overall, the DR_T populations are clustered strongly amongst themselves and show little within-group and large between-group variations. It is to be noted that these populations appear on the edges and borders of the points on the PC1-PC2 space. We also observe a North-South gradient and a subtle regional differentiation that can be seen between populations from Western Coastal plains and Eastern coastal plains.

**Population structure of Austro-Asiatic (AA)**

The AA speakers in India are exclusively tribals. Most of the AA speaking populations speak languages from the Munda subfamily whereas the AA_NER_1_01 speaks a language belonging to the Mon-Khmer sub-family. We see a clear distinction of these two language families within the

AA speakers. This is also confounded with geography as the AA_NER_1_01 are from the hills of the North-East, and the others are from the plateau region of Central and East India. The map in Figure 4.5 shows that most populations are from east-central India, and their genetic positions closely align with their geographic origin.

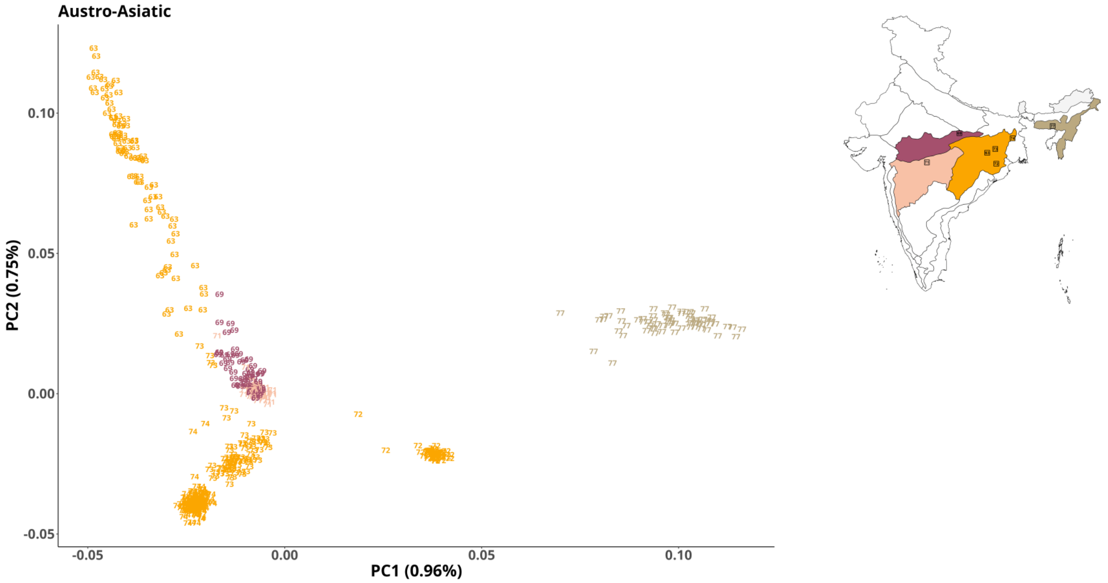

**Figure S4.5.** The scatter plot of PC1-PC2 of the Austro-Asiatic language speakers (596), coloured according to their biogeographic location, with inset of biogeographical location of the populations along with their tribe vs. non-tribe status.

#### Population structure of Tibeto-Burmans (TB)

Tibeto-Burman speakers are genetically diverse, showing widespread variation along PC1. We observed the presence of multiple genetically distinct subgroups—possibly reflecting their geographic dispersion from Northeast India to the Himalayan belt. Three genetically distinct clusters are visible, reflecting different population histories: (1) Western Himalayan highlanders, (2) North Eastern Ranges tribes, (3) Brahmaputra Valley group. Two populations from Western Himalayas, TB_WHR_1_01 and TB_WHR_1_02, showed a similar pattern as observed in the $F_{ST}$ analysis (S3). Populations from Brahmaputra Valley (TB_BPV_1_01 and TB_BPV_1_02), showing a tight cluster at the bottom, indicate their higher genetic relatedness within the same biogeographic zone. Population TB_NER_1_01, appearing in the rightmost cluster on the plot, also showed the strongest genetic affinity with the East Asian populations, compared to other TB speakers. While PC1 captures most differentiation, PC2 variation is relatively minimal within clusters.

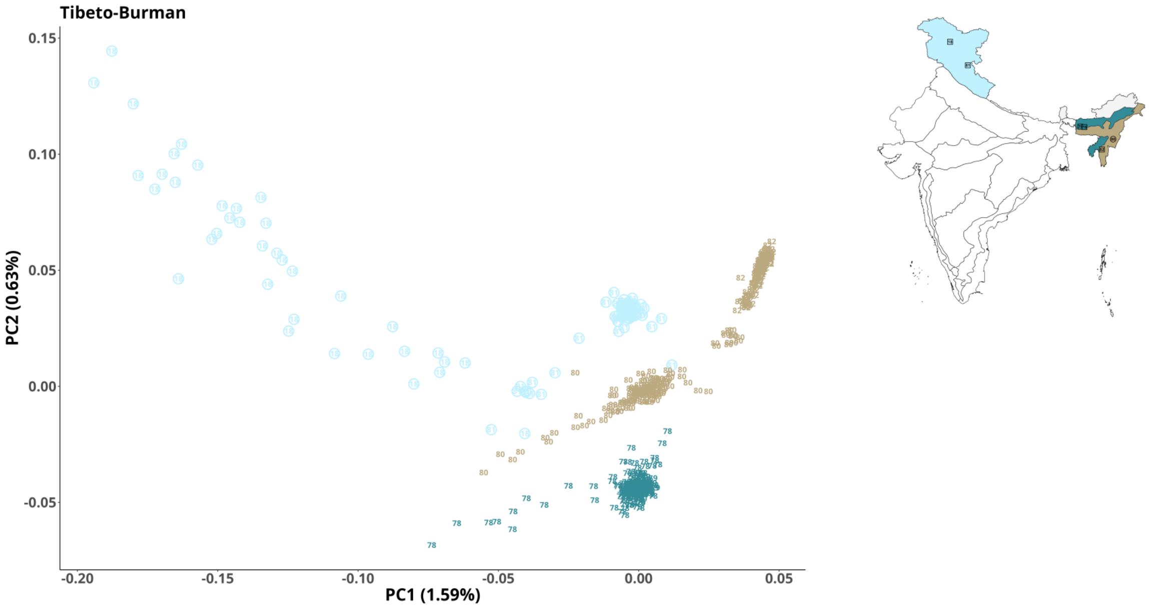

**Figure S4.6.** The scatter plot of PC1-PC2 of the Tibeto-Burman language speakers (614), coloured according to their biogeographic location, with an inset of the biogeographical location of the populations along with their tribe vs. non-tribe status. Note: Here in the above PCA plot, some points are circled, just to make them visible.

Our PCA within each language group overall reveals that Individuals speaking the same language family tend to cluster according to their biogeographic presence. There is a district pattern in the distribution of tribal populations. They typically have low within-population variation and are present along the periphery and the boundaries of the scatter-plots. In some cases, populations residing within the same biogeographical regions may form distinct clusters, as observed for AA speakers from the Eastern Plateau or DR speakers from the Nilgiri Hills. These observations reinforce that tribal populations harbour a unique spectrum of genetic variations, reflecting their antiquity and long population isolation.

##### S4.1.3 PCA of GenomeIndia populations with neighboring groups

The PCA was performed on GenomeIndia populations along with neighboring populations from Central South, Northeast, East, and Southeast Asia, using reference data from the Human Genome Diversity Project (HGDP) (Cavalli-Sforza, 2005) and Genome Asia 100k (GAsP) (Wall et al., 2019). The PC1-PC2 space of the PCA, comprising GenomeIndia (GI) populations (n = 9,475) alongside reference populations from Central South Asia (HGDP, n = 197), East Asia (HGDP, n = 223; GAsP NEA, n = 351), and South East Asia (GAsP SEA, n = 346), recapitulates the genetics-geography concordance of Indian populations with neighbouring populations in the Asian context (Basu et al., 2016).
There is documented evidence of migration and admixture along the North-West and North-Eastern corridors. We also observe a North-South cline along the PC2 where the CS Asian

populations are in close proximity with Indian populations from North and North-West. Similarly, the populations from North East India are in close proximity to some populations from East Asia and Southeast Asia.

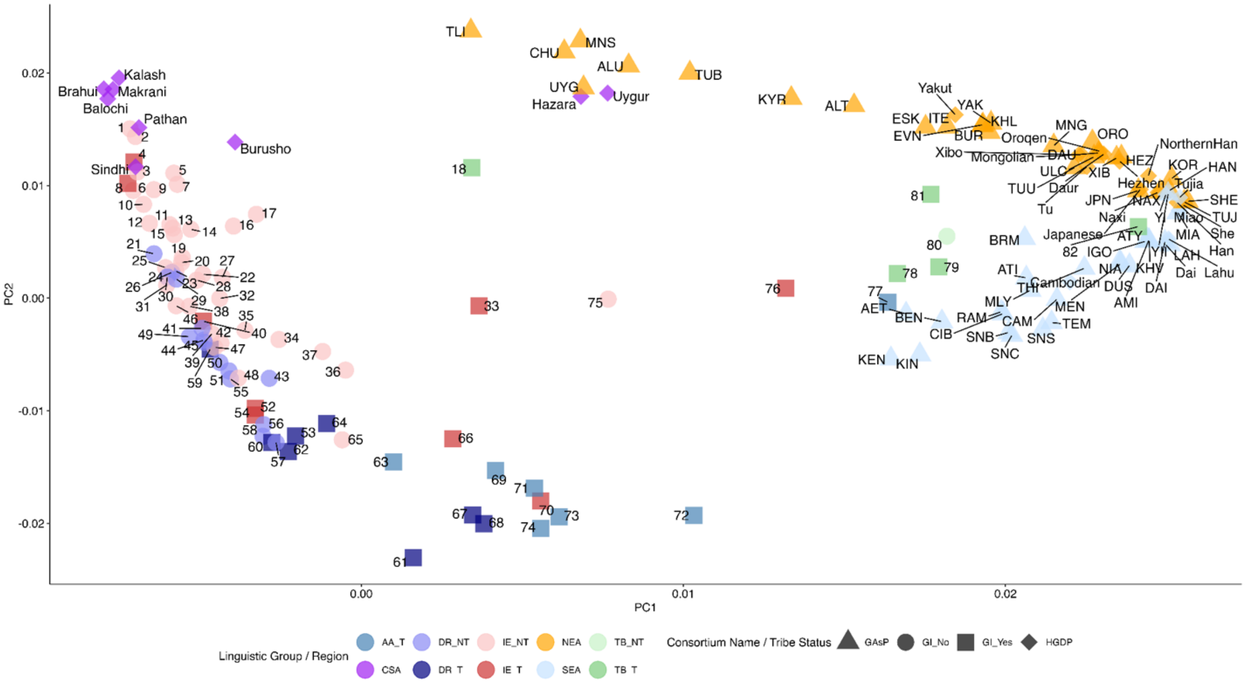

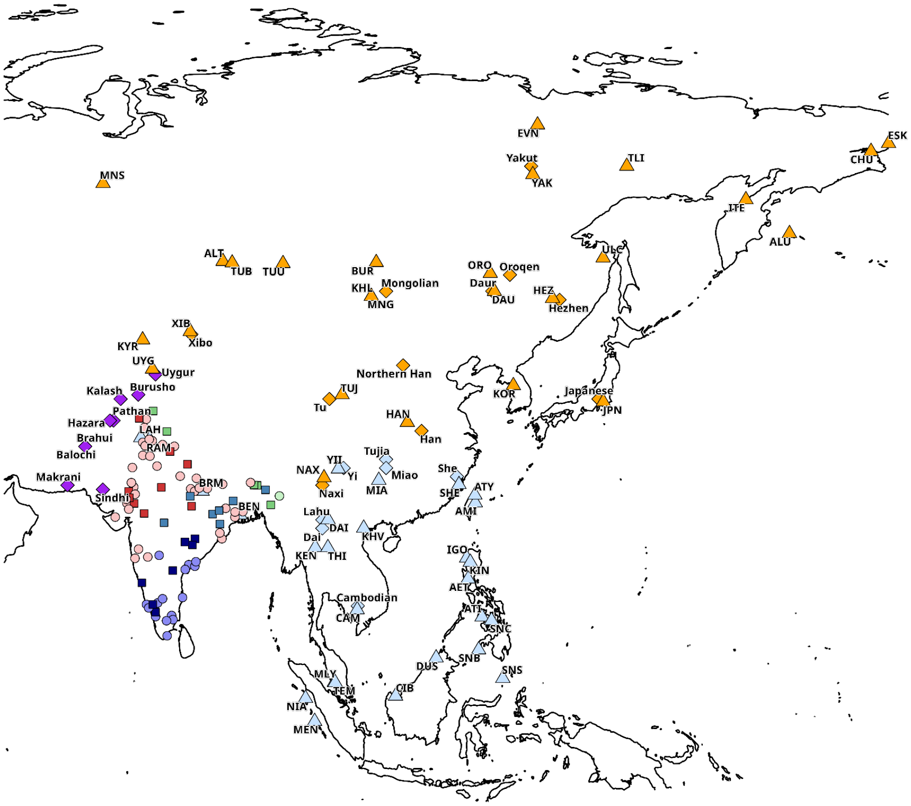

**Figure S4.7.** The top panel shows the scatter plot of PC1-PC2 with all the 82 GenomeIndia populations (the centroids using the PC1 and PC2 coordinates of all individuals from a population represent the population) with the populations from neighbouring lands. Genetic data of the populations from Central South Asia, Northeast Asia, East Asia, and Southeast Asia were taken from HGDP and GA100K. The bottom panel displays the corresponding geographical location of these populations on the map.

##### S4.2 Assessing the contribution of geographic, linguistic, and biogeographic factors to patterns of genetic variation in the Indian population

To investigate the extent to which the genetic structure of populations can be explained by geographic location, linguistic identity, and biogeographic classification, we fitted a linear model using the first two principal components (PC1, PC2) or the axes of genetic variation as the proxy of an individuals' genetic identity. We used these as outcomes and looked at how the independent variables which pertain to geography, language, and biogeography can predict the genetics of an individual. We explored four different models.

We began with **Model 1** which is the most widely used notion where we investigate how an individual's genetic identity is associated with the geographical location of the population or community.

In this initial model, we observe that latitude is significantly associated with PC2 (adjusted $R^2$ = 0.306), while longitude is significantly associated with PC1 (adjusted $R^2$ = 0.434). These results indicate that the geographic sampling locations of individuals are closely aligned with the axes of genetic variation and account for a substantial proportion of the observed structure.

**Model 1: Principal Components (PC1, PC2) regressed on latitude and longitude**

| | **PC1** | | | **PC2** | | |
|---|---|---|---|---|---|---|
| *Predictors* | *Estimates* | *std. Error* | *p* | *Estimates* | *std. Error* | *p* |
| Intercept | -0.10178 | 0.01303 | **<0.001** | 0.02971 | 0.01487 | **0.049** |
| latitude | 0.00012 | 0.00013 | 0.377 | -0.00093 | 0.00015 | **<0.001** |
| longitude | 0.00127 | 0.00016 | **<0.001** | -0.00009 | 0.00018 | 0.604 |
| Observations | 82 | | | 82 | | |
| $R^2$ / $R^2$ adjusted | 0.448 / 0.434 | | | 0.323 / 0.306 | | |

We further wanted to understand how well incorporating linguistic information along with latitude and longitude improves the adjusted $R^2$. In **Model 2,** we can see that $R^2$ adjusted improves significantly and reaches to 0.704 for PC1 and 0.523 for PC2.

**Model 2: Principal Components regressed on latitude, longitude, and linguistic identity**

| | PC1 | | | PC2 | | |
|---|---|---|---|---|---|---|
| *Predictors* | *Estimates* | *std. Error* | *p* | *Estimates* | *std. Error* | *p* |
| Intercept | -0.04248 | 0.01303 | **0.002** | 0.02032 | 0.01704 | 0.237 |
| latitude | 0.00002 | 0.00017 | 0.903 | -0.00055 | 0.00023 | **0.018** |
| longitude | 0.00067 | 0.00014 | **<0.001** | 0.00005 | 0.00018 | 0.784 |
| linguistic group (DR) | -0.01120 | 0.00316 | **0.001** | -0.00706 | 0.00413 | 0.091 |
| linguistic group (IE) | -0.01334 | 0.00253 | **<0.001** | -0.01174 | 0.00331 | **0.001** |
| linguistic group (TB) | 0.00894 | 0.00332 | **0.009** | -0.02712 | 0.00434 | **<0.001** |
| Observations | 82 | | | 82 | | |
| $R^2$ / $R^2$ adjusted | 0.722 / 0.704 | | | 0.553 / 0.523 | | |

In **Model 3**, we wanted to understand how well incorporating biogeographic classification along with latitude and longitude improves the adjusted $R^2$. We can see that $R^2$ adjusted improves significantly and reaches 0.730 for PC1 and 0.738 for PC2. We can see that biogeographic information significantly improves the model fit for PC2 compared to linguistic information, which is also expected, given that linguistic identity is not separable in the PC2 direction.

**Model 3: Principal Components regressed on latitude, longitude and biogeography**

| | PC1 | | | PC2 | | |
|---|---|---|---|---|---|---|
| *Predictors* | *Estimates* | *std. Error* | *p* | *Estimates* | *std. Error* | *p* |
| Intercept | -0.06170 | 0.02776 | **0.030** | -0.04098 | 0.02815 | 0.150 |
| latitude | 0.00015 | 0.00023 | 0.514 | -0.00008 | 0.00023 | 0.729 |
| longitude | 0.00089 | 0.00029 | **0.003** | 0.00035 | 0.00029 | 0.240 |
| Biogeography (CHR) | 0.00689 | 0.00690 | 0.322 | 0.00540 | 0.00700 | 0.443 |
| Biogeography (ECP) | -0.01604 | 0.00530 | **0.004** | 0.01883 | 0.00538 | **0.001** |
| Biogeography (EGH) | -0.00920 | 0.00696 | 0.191 | 0.02891 | 0.00706 | **<0.001** |
| Biogeography (EPL) | -0.00578 | 0.00428 | 0.182 | 0.02947 | 0.00434 | **<0.001** |
| Biogeography (ERP) | -0.02126 | 0.00433 | **<0.001** | 0.01280 | 0.00439 | **0.005** |
| Biogeography (NCH) | -0.00796 | 0.00631 | 0.212 | 0.02537 | 0.00640 | **<0.001** |
| Biogeography (NDN) | -0.00761 | 0.00650 | 0.247 | 0.02191 | 0.00659 | **0.001** |
| Biogeography (NER) | 0.00557 | 0.00455 | 0.226 | -0.00652 | 0.00462 | 0.163 |
| Biogeography (NGH) | -0.00349 | 0.00749 | 0.643 | 0.03709 | 0.00760 | **<0.001** |
| Biogeography (NRP) | -0.01845 | 0.00491 | **<0.001** | 0.01198 | 0.00498 | **0.019** |
| Biogeography (SCH) | -0.00817 | 0.00621 | 0.193 | 0.02392 | 0.00630 | **<0.001** |
| Biogeography (SDN) | -0.01171 | 0.00670 | 0.085 | 0.02358 | 0.00679 | **0.001** |
| Biogeography (WCP) | -0.01251 | 0.00654 | 0.060 | 0.01656 | 0.00664 | **0.015** |
| Biogeography (WGH) | -0.00901 | 0.00747 | 0.232 | 0.02662 | 0.00758 | **0.001** |
| Biogeography (WHR) | -0.01076 | 0.00551 | 0.055 | 0.00590 | 0.00558 | 0.295 |

| | | | | | | |
|---|---|---|---|---|---|---|
| Biogeography (WPL) | -0.01510 | 0.00653 | **0.024** | 0.01039 | 0.00663 | 0.122 |
| Observations | 82 | | | 82 | | |
| $R^2$ / $R^2$ adjusted | 0.790 / 0.730 | | | 0.796 / 0.738 | | |

In **Model 4**, we wanted to understand how well incorporating linguistic information and biogeographic classification, along with latitude and longitude, improves the adjusted $R^2$. We can see that $R^2$ adjusted improves significantly and reaches 0.844 for PC1 and 0.780 for PC2.

**Model 4: Principal Components regressed on latitude, longitude, linguistic identity and Biogeography**

| | **PC1** | | | **PC2** | | |
|---|---|---|---|---|---|---|
| *Predictors* | *Estimates* | *std. Error* | *p* | *Estimates* | *std. Error* | *p* |
| Intercept | -0.03414 | 0.02674 | 0.207 | -0.05484 | 0.03270 | 0.099 |
| latitude | 0.00012 | 0.00025 | 0.625 | 0.00027 | 0.00031 | 0.387 |
| longitude | 0.00056 | 0.00025 | **0.030** | 0.00051 | 0.00031 | 0.106 |
| linguistic group (DR) | -0.00604 | 0.00333 | 0.074 | 0.00214 | 0.00407 | 0.601 |
| linguistic group (IE) | -0.00834 | 0.00231 | **0.001** | -0.00359 | 0.00283 | 0.209 |
| linguistic group (TB) | 0.00852 | 0.00300 | **0.006** | -0.01277 | 0.00367 | **0.001** |
| Biogeography (CHR) | 0.01510 | 0.00547 | **0.008** | -0.00003 | 0.00669 | 0.997 |
| Biogeography (ECP) | -0.00991 | 0.00439 | **0.028** | 0.01345 | 0.00537 | **0.015** |
| Biogeography (EGH) | -0.00453 | 0.00561 | 0.423 | 0.02348 | 0.00686 | **0.001** |
| Biogeography (EPL) | -0.00125 | 0.00383 | 0.746 | 0.02237 | 0.00469 | **<0.001** |
| Biogeography (ERP) | -0.01102 | 0.00371 | **0.004** | 0.00802 | 0.00454 | 0.082 |
| Biogeography (NCH) | -0.00206 | 0.00534 | 0.702 | 0.02229 | 0.00653 | **0.001** |
| Biogeography (NDN) | -0.00416 | 0.00596 | 0.488 | 0.01931 | 0.00729 | **0.010** |
| Biogeography (NER) | 0.00359 | 0.00352 | 0.312 | -0.00761 | 0.00430 | 0.082 |
| Biogeography (NGH) | 0.00046 | 0.00611 | 0.940 | 0.03258 | 0.00747 | **<0.001** |
| Biogeography (NRP) | -0.01107 | 0.00408 | **0.009** | 0.00701 | 0.00499 | 0.165 |
| Biogeography (SCH) | -0.00520 | 0.00544 | 0.343 | 0.01879 | 0.00665 | **0.006** |
| Biogeography (SDN) | -0.00746 | 0.00543 | 0.174 | 0.01797 | 0.00664 | **0.009** |
| Biogeography (WCP) | -0.00786 | 0.00563 | 0.167 | 0.01371 | 0.00688 | 0.051 |
| Biogeography (WGH) | -0.00537 | 0.00606 | 0.379 | 0.02161 | 0.00741 | **0.005** |
| Biogeography (WHR) | -0.00766 | 0.00432 | 0.081 | 0.00182 | 0.00529 | 0.733 |
| Biogeography (WPL) | -0.00954 | 0.00545 | 0.085 | 0.00695 | 0.00666 | 0.301 |
| Observations | 82 | | | 82 | | |
| $R^2$ / $R^2$ adjusted | 0.884 / 0.844 | | | 0.837 / 0.780 | | |

These findings expand our understanding of confounding factors in genetics and its relationship with various other variables like culture, physical geography and biogeography. These results also indicate that the sampling in GenomeIndia captures a large proportion of the genetic diversity structured along the contours of geography, language, and biogeography. From the perspective of planning future research studies, this analysis provides us with an instrument to define ‘homogeneous’ populations in genetic studies like large scale studies of association.

**S4.3 Unsupervised Clustering - ADMIXTURE analysis**

To resolve population structure at a finer scale and identify ancestral components, we performed unsupervised clustering analysis on 9,475 samples from 82 populations. This dataset was established after excluding the CAO group and cryptic related individuals. Quality control of autosomal markers was performed using PLINK v2.0 (Chang et al., 2015). Variants were filtered based on a Hardy-Weinberg Equilibrium (HWE) p-value $< 10^{-8}$, a missing genotype rate < 0.02, and a Minor Allele Frequency (MAF) ≥ 0.01. To mitigate the effects of linkage disequilibrium, pruning was applied using a window size of 50 SNPs, a step size of 5 SNPs, and an $r^2$ threshold of 0.1. This resulted in a final dataset of 970,954 variants. Maximum likelihood clustering was then conducted using ADMIXTURE v1.3.0 (Alexander et al., 2009) with the default parameters. The --cv flag was employed to perform cross-validation procedures to estimate the optimal number of clusters (K).

We observed four major clusters which we can map to four ancestral populations of mainland India, namely ANI (Ancestral North Indian), ASI (Ancestral South Indian), AAA (Ancestral Austro-Asiatic) and ATB (Ancestral Tibeto-Burman), based on the geographic and linguistic affiliations of the populations, and by comparison with the previous findings (Moorjani et al., 2013; Basu et al., 2016). In addition to these, we identified a distinct cluster corresponding to two Dravidian speaking tribal populations (populations 61 and 62) from the Nilgiri Hills, which will henceforth be referred to as ASI-D. Notably, these populations also formed a separate cluster in the PCA analysis shown in Figure S4.1, reinforcing the ADMIXTURE results.

We observed a genetic cline from North to South India, indicating a gradual shift in ancestry components among the mainland Indian populations. This clinal pattern represents a gradual decrease of ANI ancestry proportions from north to south, accompanied by a corresponding increase of ASI ancestry proportions, a pattern that was observed predominantly among Indo-European and Dravidian speaking non-tribal populations. Notably, DR_NT populations exhibited higher proportions of ANI ancestry, compared to the DR_T populations, which aligns with PCA inference, as both IE_NT and DR_NT showed a diffuse pattern along the north-south cline. This trend was also reflected in variant-sharing analyses S3.

The ATB (Ancestral Tibeto-Burman) component is primarily present among the populations from north-east India. However, these groups also displayed varying proportions of other ancestry components, suggesting a possible admixture with other Indian populations. IE language speaking populations from the Western Himalayas, like 16, 17,18, and 33, exhibited a notable proportion of ATB ancestry while residing in the neighbourhood of IE populations. This admixture pattern aligns with their biogeographical habitat of living in a mountainous region. The AA_T (77) from the North Eastern region showed majorly ATB component, along with the AAA component.

The AAA component is majorly identified among the Austroasiatic-speaking populations, from the central and eastern parts of mainland India, with varying levels of ASI admixture. Notably, the AAA ancestry component was not confined solely to these populations. It was also detectable, to a lesser extent, across several other population groups, noticeably among some IE_NT populations 34, 35, 36, and 37 from the eastern part of India. This suggests there was likely broader historical spread and genetic influence of AA groups across the subcontinent.

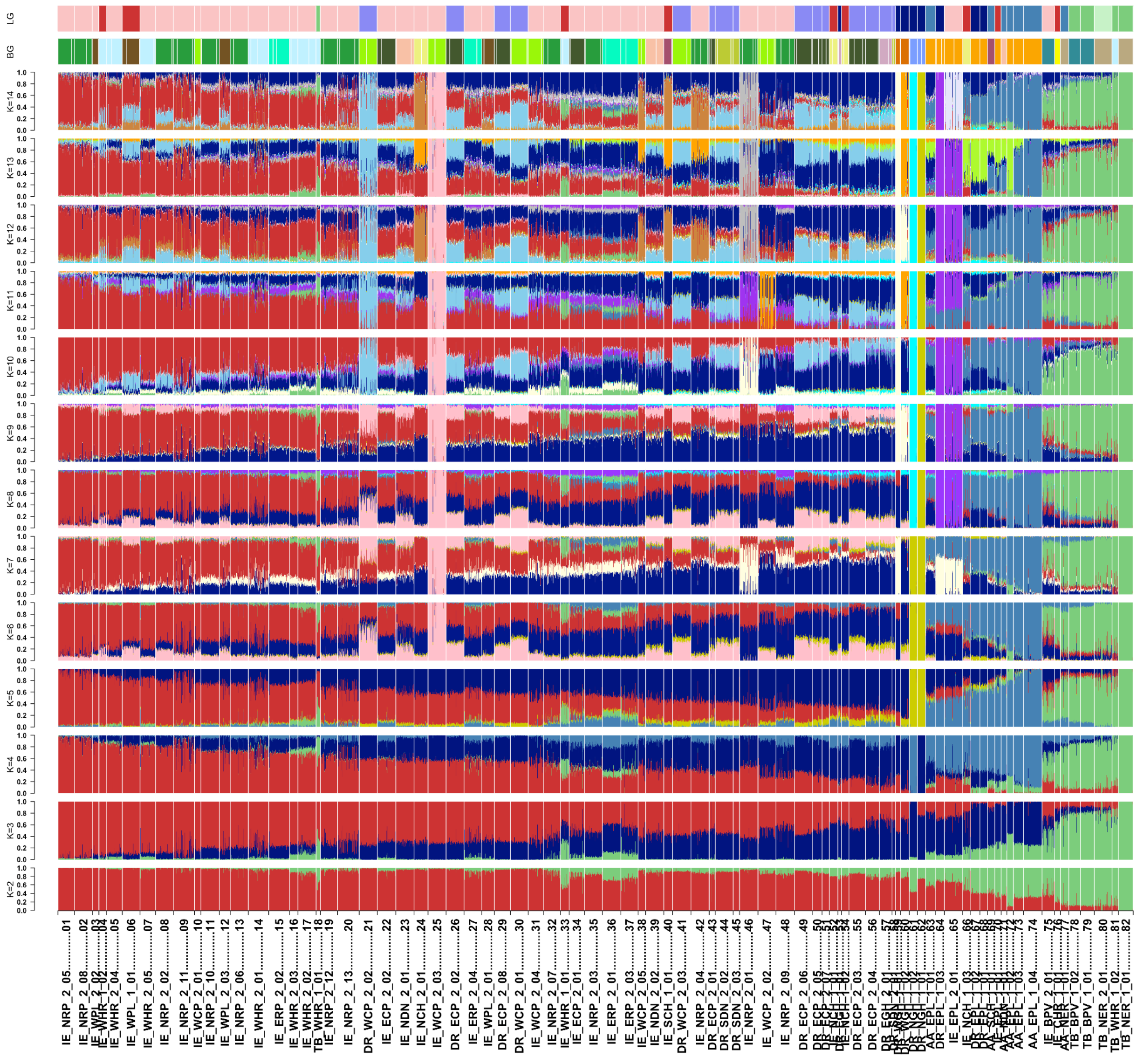

**Figure S4.8.** Admixture Plots: Ancestry estimates for 9,475 individuals from 82 mainland Indian populations at K = 2 to K=14 . Each individual is represented as a vertical bar partitioned into colored segments, with segment lengths proportional to ancestry contributions from Various components.

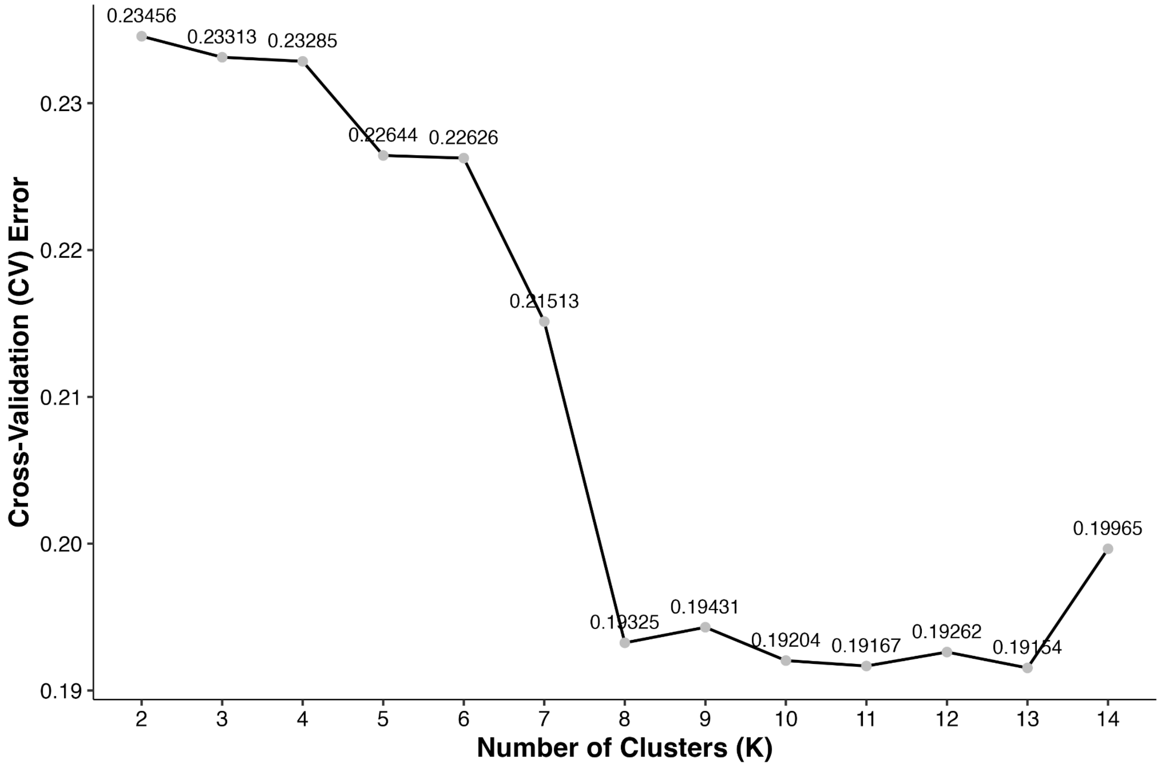

**Figure S4.9.** Cross-validation errors of Admixture run (Unsupervised clustering) from K=2 to K=14

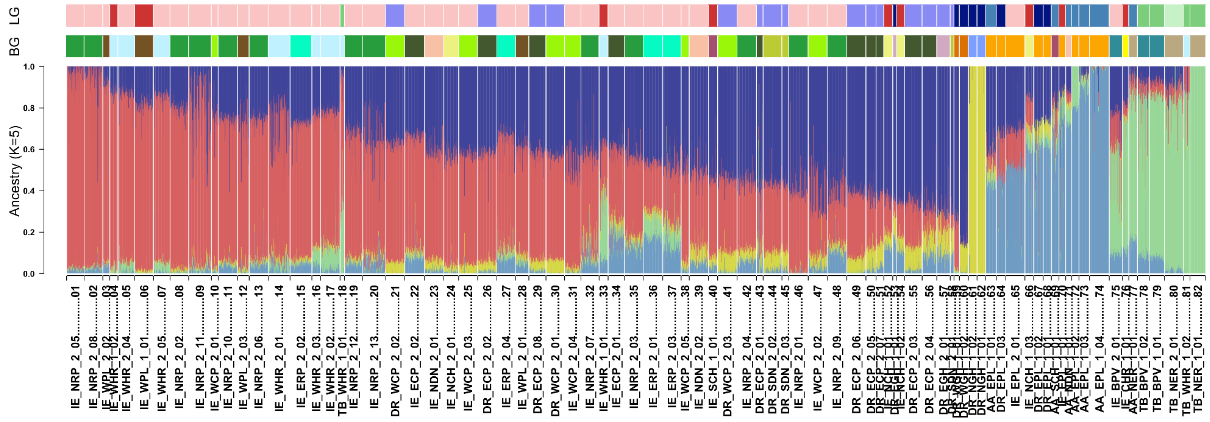

**Figure S4.10.** Admixture plot of the Mainland India Populations. Ancestry estimates for 9,475 individuals from 82 mainland Indian populations at K = 5. Each individual is represented as a vertical bar partitioned into colored segments, with segment lengths proportional to ancestry contributions from five components. Red and dark blue represent ANI (Ancestral North Indian) and ASI (Ancestral South Indian), respectively. Sky blue and pale green denote the inferred AAA (Ancestral Austroasiatic) and ATB (Ancestral Tibeto-Burman) components. Dark yellow indicates a distinct ancestry component, referred to here as the (ASI-D), predominantly observed in populations from the Nilgiri Hills. The upper bar (LG) denotes linguistic group, and the lower bar (BG) indicates the biogeographic region of each population.

We observe a North-South cline along the PC2 axis, comprising mainly the IE and DR speaking populations. The IE populations residing mostly in North India constitute one end of this cline, whereas the DR populations residing mostly in South India constitute the other end. The DR populations (61 and 67) do not cluster with other DR populations, indicating their genetic isolation. The AA populations residing in Central and East India cluster close to this cline. However, the AA populations no. 72 and 77 cluster away from other AA populations, suggesting genetic isolation in case of population no. 72 and genetic closeness with TB speakers for population 77. Some of the DR and IE tribal populations cluster with the AA tribes. The TB speakers constitute a separate cluster spread along the PC1 axis, corroborating their geographical presence in Northeast India. The TB population no. 18 is spread along PC1 between the IE populations and other TB populations, suggesting an admixture between these two linguistic families.

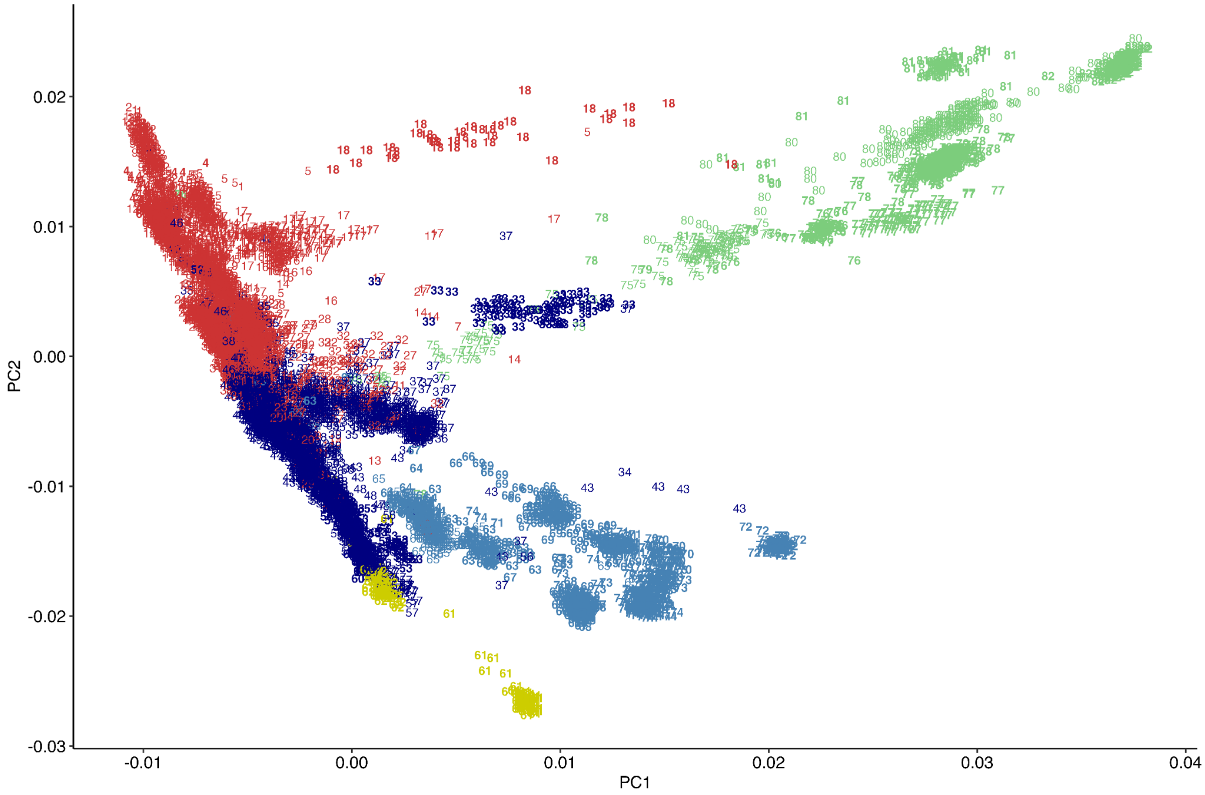

**Figure S4.11.** The scatterplot of the first two eigenvectors of the PCA plot consists of 9475 individuals from 82 populations from mainland India. The X-axis represents PC1, and the Y-axis represents PC2.  Here, each individual is coloured according to the major ancestry component out of ANI (Ancestral North Indian), ASI (Ancestral South Indian), AAA (Ancestral Austroasiatic), ATB (Ancestral Tibeto-Burman), and ASI-D.

##### S4.3.1 Refining Population Structure Inference through Iterative ADMIXTURE Analysis

From the ADMIXTURE results (Fig. 4.13), we observed that increasing the number of inferred ancestral components (K) beyond 5 led to the appearance of distinct ancestry signals in several individual populations. The cross-validation (CV) error continued to decline steadily between K = 6 and K = 8, indicating incremental improvements in model fit. However, this steady decline was accompanied by the emergence of population-specific clusters, particularly at higher K values, suggesting that some of the additional components may be capturing fine-scale structure or population-specific drift rather than broadly shared ancestry. These patterns likely reflect the influence of highly differentiated groups with unique demographic histories, such as strong founder effects and long-term isolation.

To test this hypothesis, we conducted the ADMIXTURE analysis in which we removed populations that consistently formed unique clusters at higher K values. These included populations 20, 25, 46, 59, 61, 62, 64, and 65. In the initial analysis, these groups repeatedly appeared as standalone components after K = 4 or K = 5, a pattern we interpret as the genetic imprint of founder effects, long-term isolation, and pronounced genetic drift. Such forces can drive allele frequencies away from regional patterns, resulting in artificial inflation of inferred ancestry components.

By excluding these small endogamous isolated populations, we aimed to achieve a more stable and interpretable model of population structure across the remaining dataset. This suggests that due to the long-term isolation and population-specific drift, they were identified as a distinct ancestral component by ADMIXTURE.

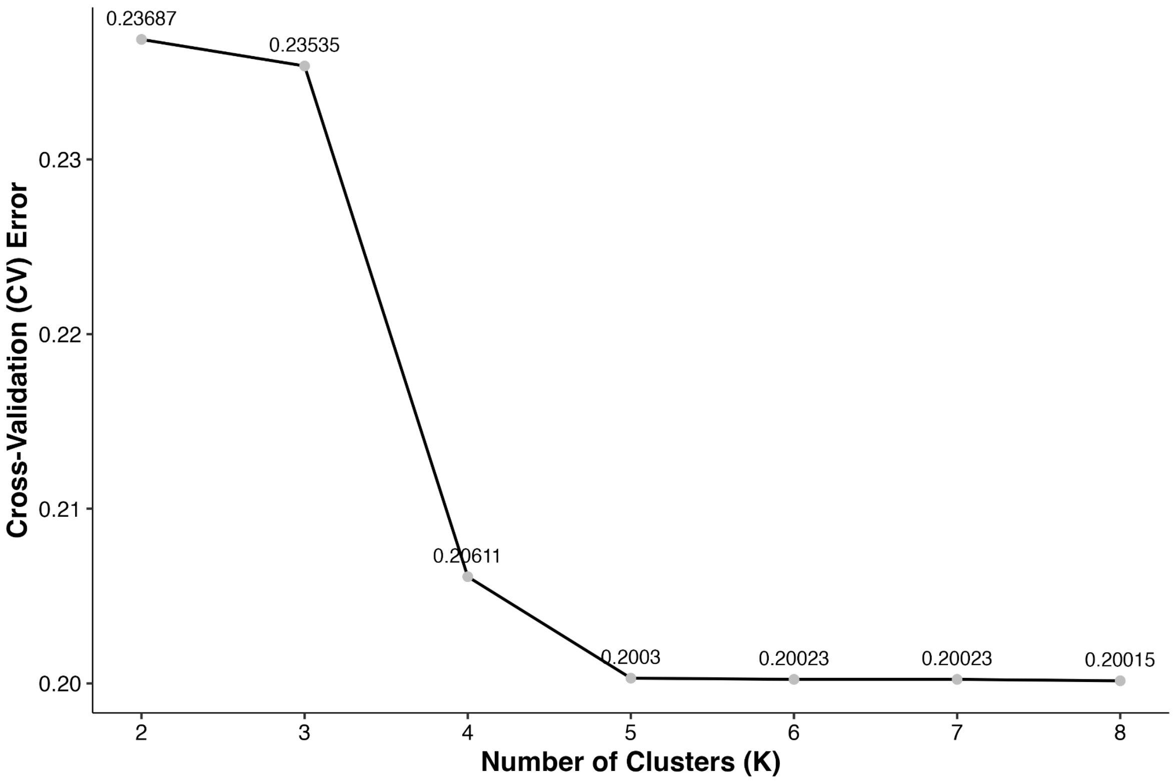

**Figure S4.12.** CV errors of the Admixture run after removing 8 populations from all the whole dataset

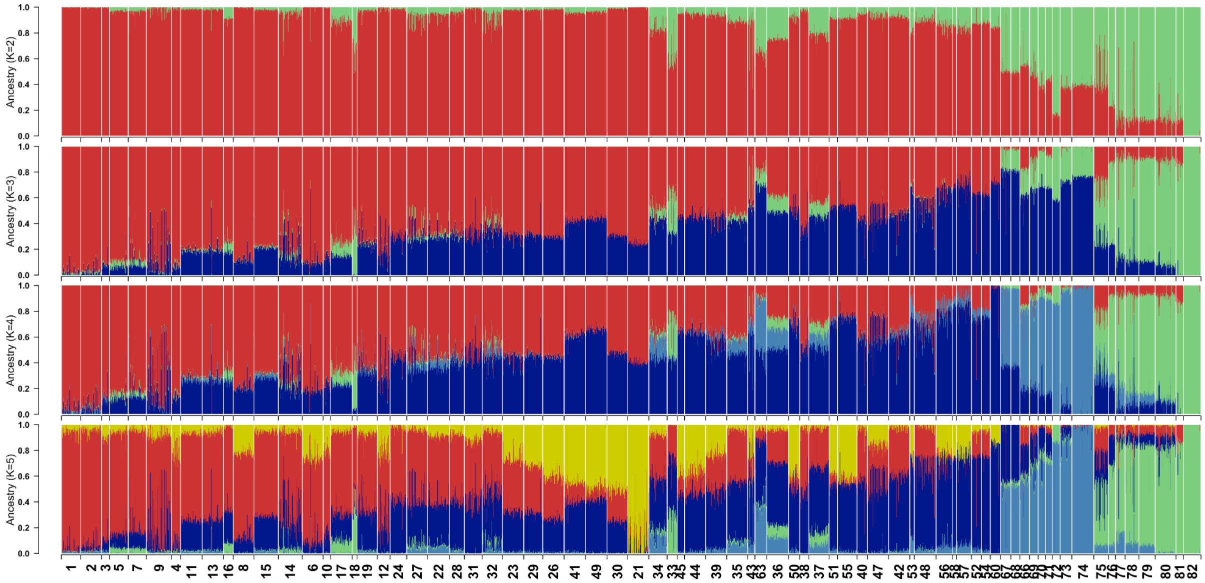

**Figure S4.13.** Admixture plot of the Mainland India Populations. Ancestry estimates for 8541 individuals from 74 mainland Indian populations at different K. Each individual is represented as a vertical bar partitioned into colored segments, with segment lengths proportional to ancestry contributions from five components. Red and dark blue represent ANI (Ancestral North Indian) and ASI (Ancestral South Indian), respectively. Sky blue and pale green denote the inferred AAA (Ancestral Austroasiatic) and ATB (Ancestral Tibeto-Burman) components. Dark yellow indicates a distinct ancestry component, referred to here as the (ASI-D) based on initial K=5 Admixture results, predominantly observed in populations from the Nilgiri Hills.The upper bar (LG) denotes linguistic group, and the lower bar (BG) indicates the biogeographic region of each population.

We observed a substantial decrease in the cross-validation (CV) error, particularly at K = 4 and K = 5, up to the point where one remaining population again formed a separate cluster. And from that point, the CV errors remain stable.

This pattern was similarly observed in the full dataset with all ethnic groups, where increasing K after 4 groups or 5 groups led to the separation of specific groups. Our analysis suggests that ADMIXTURE, when applied to datasets with numerous populations under varying demographic histories, can identify some as distinct clusters, which in turn influences the CV scores. Therefore, understanding the demographic context of each population is crucial when interpreting ADMIXTURE results. Based on this refined approach, we selected K = 5 for the admixture analysis as the most informative value, as it provides a balance between model complexity and explanatory power.

Our ADMIXTURE results reinforce the complexity of interpreting population structure in this context. In deeply structured regions like South Asia, unsupervised clustering methods may capture recent demographic isolation rather than ancient divergence, leading to potential misinterpretations. These results emphasize the importance of integrating demographic modeling with genetic clustering analyses when reconstructing deep population histories.

**S4.4 Quantifying the diversity in Indian populations based on admixture proportions**

**Method**

Based on the admixture proportion at K = 5, for each population, we estimated the within-individual diversity (or abundance of the admixture proportions) using the Shannon index. Similarly we calculated the between-individual diversity (or the difference in the admixture proportions between two individuals) using Bray-Curtis distance.

Let,

$$P_{ij} = (p_1, p_2, \dots, p_k);\ \sum_{S=1}^{K} p_S = 1$$

be the ancestry proportions of the $i^{th}$ individual in the $j^{th}$ population, which consists of K ancestral components.

For each of the 82 populations, we have used K=5 ADMIXTURE results to quantify the within-individual (alpha) diversity using the Shannon index and between-individual (Beta) diversity using the Bray-Curtis distance from the vegan R package.

The Shannon index is defined as

$$H = -\sum_{S=1}^{K} p_S \log(p_S)\ ; K = 5$$

The Bray-Curtis distance is defined as

$$d_{jk} = \frac{\sum_i |x_{ij} - x_{ik}|}{\sum_i (x_{ij} + x_{ik})}$$

Where $x_{ij}$ and $x_{ik}$ refers to the ancestry proportions on ancestral components (columns) i and individuals (rows) j and k.

After generating the diversity index for each population, we took the median of within-individual and between-individual measures to estimate the average within-individual and between-individual diversity for the population.

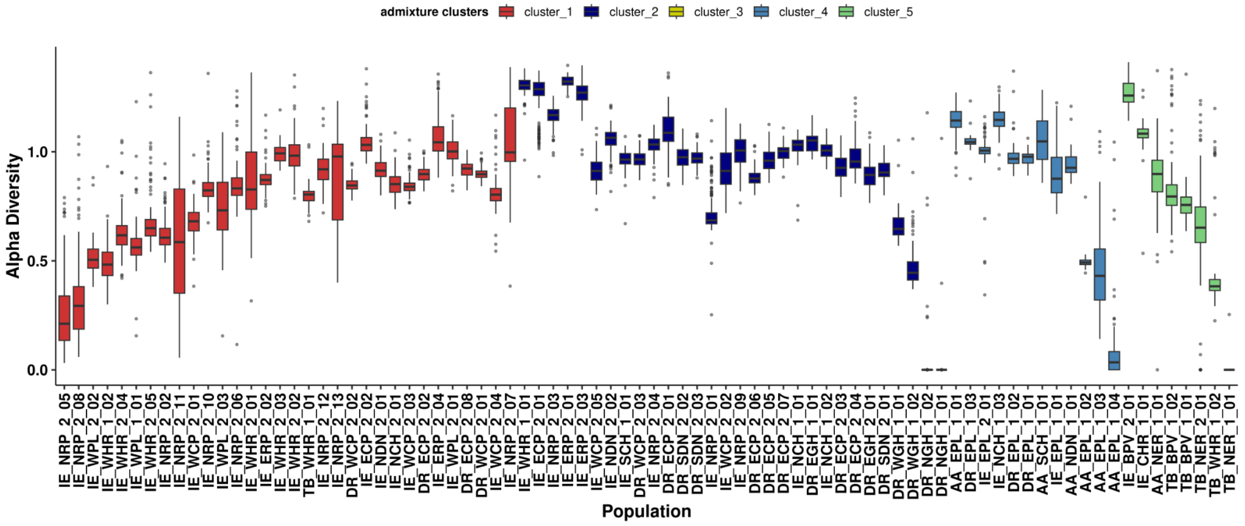

**Figure S4.14.** Distribution of the within-individual diversity across 82 populations based on admixture proportions(K=5) colored by the admixture cluster labels. cluster 1 (ANI), cluster 2 (ASI), cluster 3 (ASI-D), cluster 4 (AAA), and cluster 5 (ATB).

In general, we observe that within each admixture cluster, the alpha diversity decreases with increase in the ancestry of the representative ancestral component.

Then, for each population, we take the median of within-individual and between-individual measures to have an estimate of the average within-individual and between-individual diversity for the population.

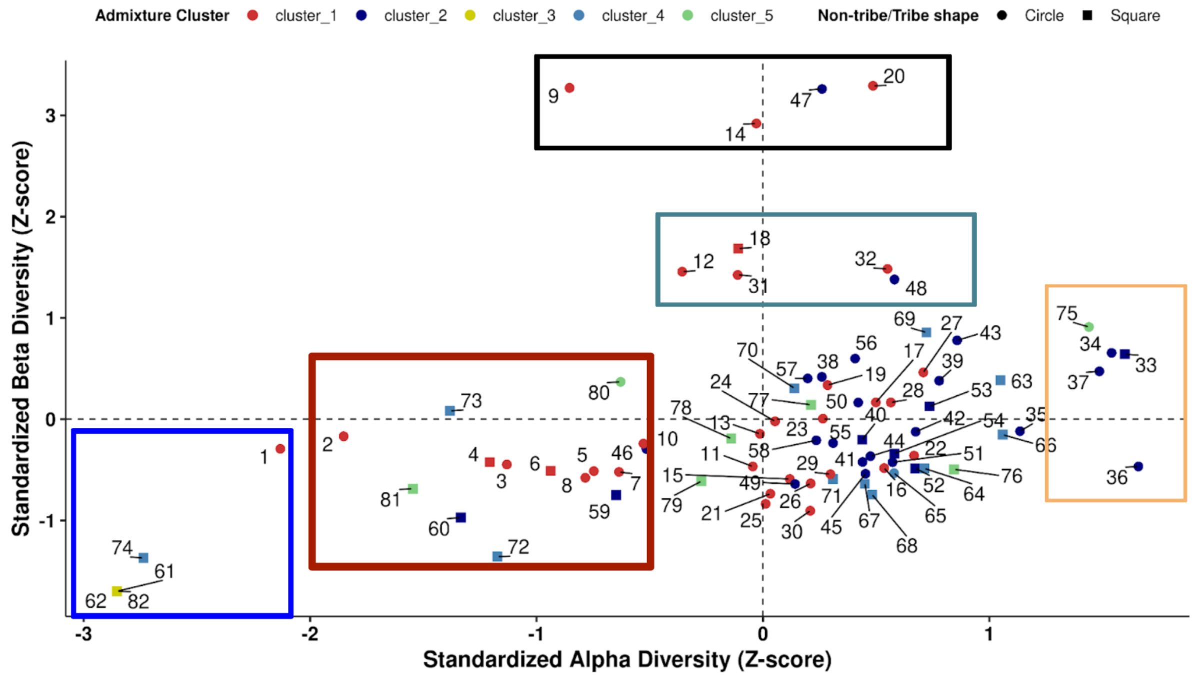

**Figure S4.15.** Populations are plotted based on the standardized median within-individual diversity on the x-axis and standardized median between-individual diversity on the y-axis. Each point represents a population (numerically labeled), colored by its corresponding ADMIXTURE cluster and shaped by social classification, tribal populations are denoted by squares and non-tribal populations by circles. This figure is a part of Figure 2.

We have summarized our observations into the following five major points, also highlighted in the above figure.

**Blue Box**: This cluster consists of representative populations corresponding to each major ancestral group: ANI, AAA, ATB, and ASI-D. As these are representative of specific ancestries, they exhibit the lowest within-individual and between-individual diversity. Notably, population 1, representative of ANI ancestry, shows higher within-individual diversity relative to the other reference groups and moderate between-individual diversity, suggesting slight variability in ancestral proportions among individuals within the population.

**Black Box**: Populations in this region show extremely high between-individual diversity and moderate within-individual diversity. ADMIXTURE analysis indicates that these populations have ancestry components from at least two distinct ancestral groups. Principal component (PC) analysis reveals broad population structure with multiple subclusters, consistent with elevated inter-individual variation in ancestry components.

**Dark Cyan Box**: Similar to the black box, these populations exhibit high between-individual diversity and moderate within-individual diversity. They also possess ancestry from two to three ancestral components. However, in contrast to the black box populations, these groups show lower between-individual variation in ancestral components.

**Light Orange Box**: Populations in this region display high within-individual diversity and moderate between-individual diversity. ADMIXTURE analysis shows contributions from at least four ancestral populations. Principal component analysis identifies these groups as "**melting pot**" populations, those that connect all representative ancestries and likely received gene flow from multiple ancestral groups in successive admixture events. The biogeographic location of these populations is Western Himalayas(33), Eastern Coastal Plains (34), Eastern Riverine Plains (36,37), and Brahmaputra Valley(75). Population 33 has significantly high ATB and ASI ancestry compared to ANI ancestry, given that the population geographically resides with high ANI ancestry populations.

**Dark Red Box**: This group contains populations with both low within-individual and low between-individual diversity. Populations with high ANI ancestry **(populations 2–10)** display a pattern of increasing within-individual diversity as ANI ancestry decreases. Interestingly, between-individual diversity remains largely unchanged with this increase in within-individual diversity, suggesting a strong role for social structure and endogamy in maintaining genetic homogeneity, which can be achieved through panmixing within the group after initial admixture. Among Tibeto-Burman groups, population 80 shows relatively moderate within-individual and between-individual diversity, likely due to the presence of three ancestry components, whereas population 81, with only two ancestral components, has lower diversity.

Other populations fall outside these designated boxes and exhibit moderate within-individual and between-individual diversity. These populations lie along genetic clines and derive ancestry from multiple groups. The moderate between-individual diversity observed may reflect endogamy, resulting in relatively homogeneous ancestry proportions across individuals within populations, even amidst admixture.

##### S4.5 Analysis of Continentally Admixed Outgroup (CAO)

Among the diverse populations sampled from mainland India, the CAO group is of particular interest. Previous studies have elucidated this population's admixture with African lineages, specifically Bantu-speaking groups (Narang et al., 2011; Shah et al., 2011). The availability of whole-genome sequencing (WGS) data for 49 CAO individuals, integrated with the comprehensive Genome India dataset, provides a unique opportunity to analyze this group in greater detail. We sought to clarify the fine-scale population structure and demographic history of the CAO group using a combination of genomic tools.

To characterize the ancestry of the CAO population, we conducted a supervised ADMIXTURE analysis. Our initial framework included all mainland Indian populations alongside African groups from the HGDP dataset. We ran ADMIXTURE at K = 6, based on prior findings that the genetic structure of mainland India can be described by five major ancestral components, to which we added a sixth: African ancestry, represented by the Yoruba (YRI) population.

The CAO group displayed a distinct admixture signal, with components deriving from Ancestral North Indian (ANI), Ancestral South Indian (ASI), and African Yoruba lineages (Figure 4.16). A minor signal corresponding to Ancient Ancestral Austroasiatic (AAA) ancestry was also detected. These findings reinforce earlier evidence of admixture involving both South Asian and African source populations.

To refine this signal and more precisely identify the African source population, we expanded our supervised analysis. We constructed a dataset of 1,784 samples, comprising 849 individuals from mainland India—selected to represent ANI, ASI, and geographically proximal groups (populations 6, 31, 38, 47, and 60)—and 14 African populations, of which five were designated as ancestry donors. These included Yoruba (YRI), Mbuti, Biaka, Gambian (GWD), and Luhya (LWK)—representing a transect from West to South-Central Africa. CAO group showed the highest mean African ancestry proportion (~56.5%) from the Luhya (LWK) population, a Bantu-speaking group from western Kenya included in the 1000 Genomes Project (Figure 4.17). The Luhya are known to occupy a transitional genetic position between East and Central African populations, which may reflect an ancient gene flow corridor linking regions along the Indian Ocean.

In addition, the CAO individuals carried ~28% ANI ancestry and ~10% ASI ancestry, consistent with a South Asian-African admixture model. The samples also show variation in ancestry Figure 4.19 . Taken together, these results provide compelling evidence that the CAO group is an admixed population, with the African ancestral contribution deriving not from West Africa (e.g., Yoruba), but more specifically from a group genetically closest to the Luhya of East Africa.

We also used PCA to look at samples defined earlier in Figure 4.17 The CAO samples—labeled as population 83 in our dataset—consistently projected closer to African populations along the first two principal components (PC1 and PC2) Figure 4.20. which revealed a clear intermediate position for the CAO between mainland South Asian and African populations. These results strongly support a model of admixture, in which the CAO group derives ancestry from both African and South Asian sources, shaping the unique genetic structure observed today.

In the following sections on mitochondrial and Y-chromosome haplogroup diversity (Supplementary section S5), we observed that in both cases CAO harbours more than 70% of their haplogroups which are unique to African populations (mitochondrial haplogroup L and Y-chromosome haplogroups E1, E2 and B2). Demographic analysis of the CAO population (S5) also substantiates their complex population history.

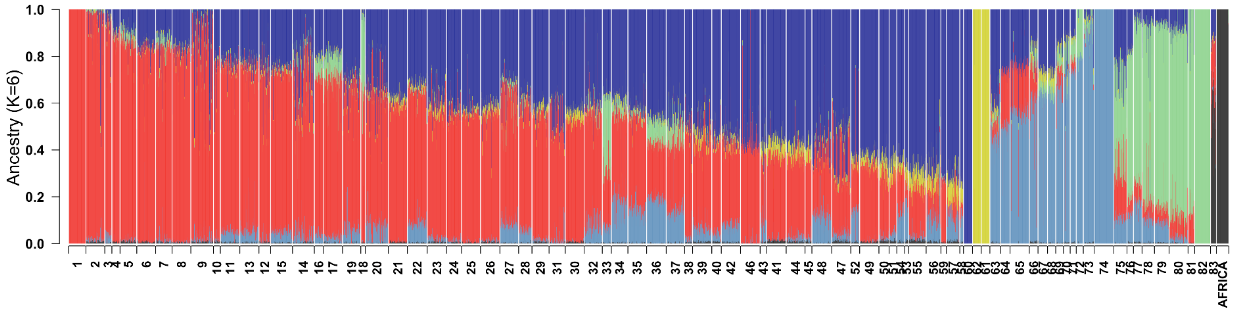

**Figure S4.16.** Supervised admixture plot including all mainland Indian populations and their ancestry components (ANI, ASI, AAA, ATB, ASI-D), along with an African reference population (YRI). The CAO population shows evidence of admixture between Indian (ANI and ASI) and African (YRI) ancestries

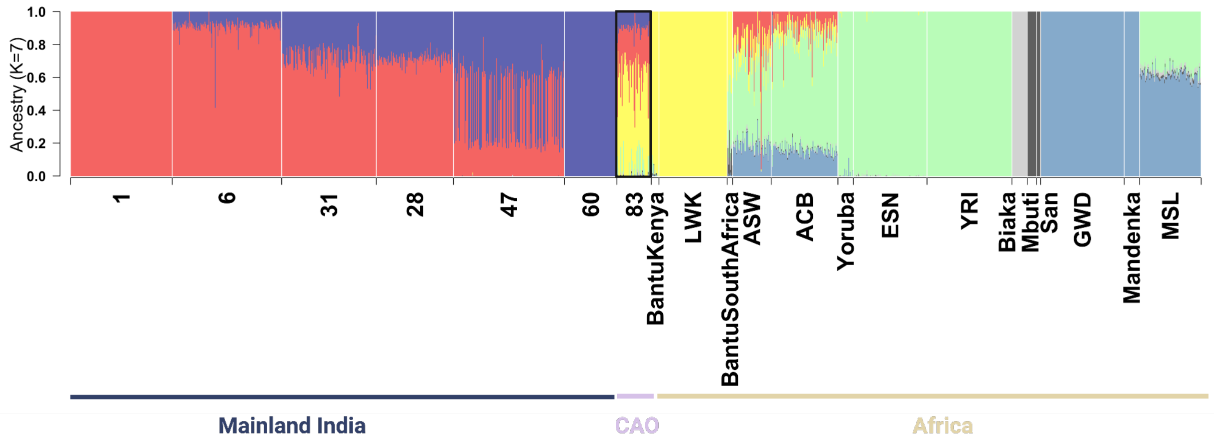

**Figure S4.17.** Supervised admixture plot showing the proportions of ANI and ASI ancestry across populations, alongside five African reference groups (Yoruba [YRI], Mbuti, Biaka, Gambian [GWD], and Luhya [LWK]). The CAO (83) sample displays evidence of admixture between mainland Indian and Luhya-like African ancestries.

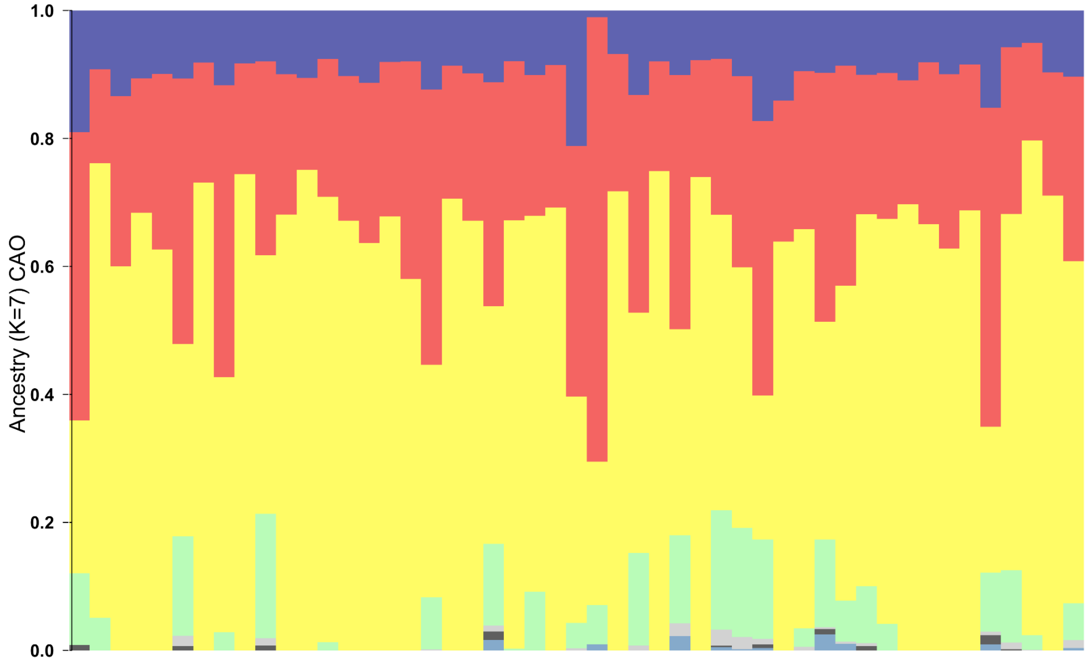

**Figure S4.18.** Admixture plot of CAO samples showing varying proportions ASI, ANI and African Ancestry.

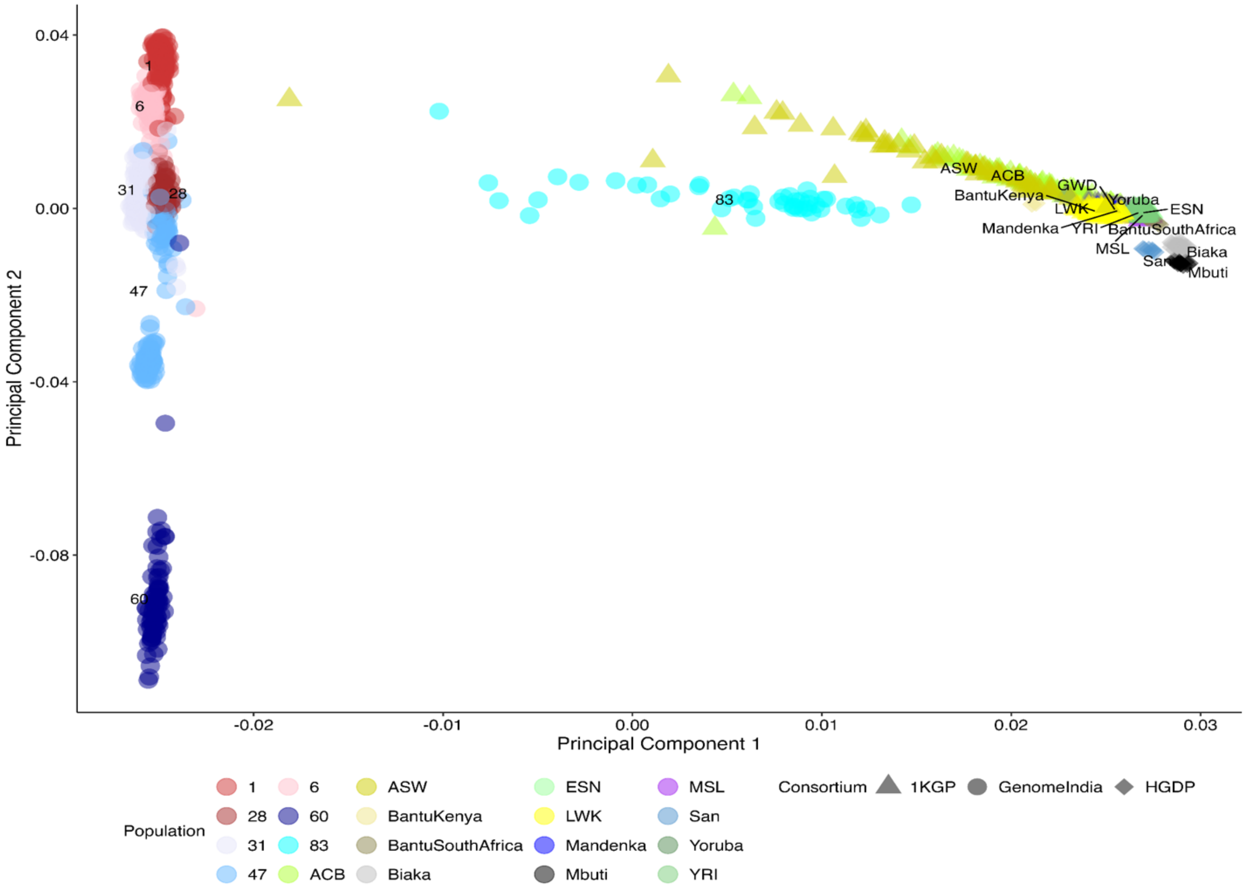

**Figure S4.19.** The plot illustrates the first two principal components (PC1 vs. PC2), utilizing whole-genome data to visualize genetic structure. The African and Indian populations form distinct clusters at opposite ends of the PC1 axis. Population 83 (the continentally admixed group) occupies an intermediate position in the PCA space, forming a cline that connects the African and Indian clusters. Notably, along the second principal component (PC2), Population 83 clusters more closely with the LWK (Luhya in Webuye, Kenya) population compared to other African groups.

#### Supplementary Section S5: Mitochondrial DNA, X, and Y Chromosome Analysis

Eric Macwan[1], Debasrija Mondal[1], Pratheusa Machha[2], Shouvanik Sengupta[3,4], Pooja Tayade[2], Analabha Basu[3,4], Shweta Ramdas[1], Divya Tej Sowpati[2,5]

[1]Centre for Brain Research (CBR), IISc Campus, Bengaluru, India. [2]CSIR - Centre for Cellular and Molecular Biology (CSIR-CCMB), Hyderabad, India. [3]BRIC - National Institute of Biomedical Genomics (BRIC-NIBMG), Kolkata, India. [4]Regional Centre for Biotechnology (RCB), Faridabad, India. [5]Academy of Scientific and Innovative Research, Ghaziabad, India.

##### Summary

Section 5.1 - Mitochondrial DNA analysis
Section 5.2 - Chromosome X analysis
Section 5.3 - Chromosome Y analysis

##### Section 5.1 - Mitochondrial DNA

###### Mitochondrial haplogroup distribution

Haplogrep3 was used to estimate haplogroups for all sequenced samples. The distribution of haplogroup quality scores from Haplogrep3 is shown in figure S5.1.1.

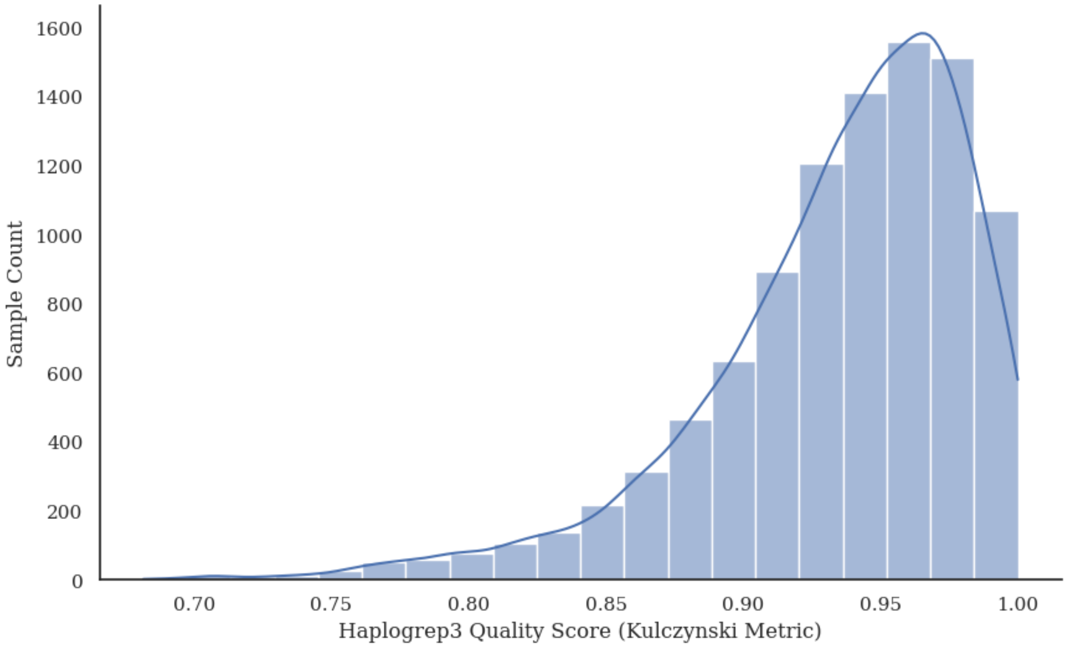

**Figure S5.1.1.** Distribution of Haplogrep quality scores for 9,768 samples

The distribution of mitochondrial haplogroups across the Indian subcontinent reveals distinct patterns in haplogroup frequency, population structure, and regional association. Consistent with previous studies, the M, U, and R haplogroups have the highest frequencies in GI; 54% of GI samples have the M haplogroup, followed by 15% with the U haplogroup and 13% with the R haplogroup. (Figure S5.1.2A). Large and isolated populations both contribute to haplogroup diversity; however, isolated populations form a substantial proportion of the genetic composition for many haplogroups, with ~50% of samples forming sublineages of the M branch, D, G, C, and Z coming from isolated groups (Figure S5.1.2B).

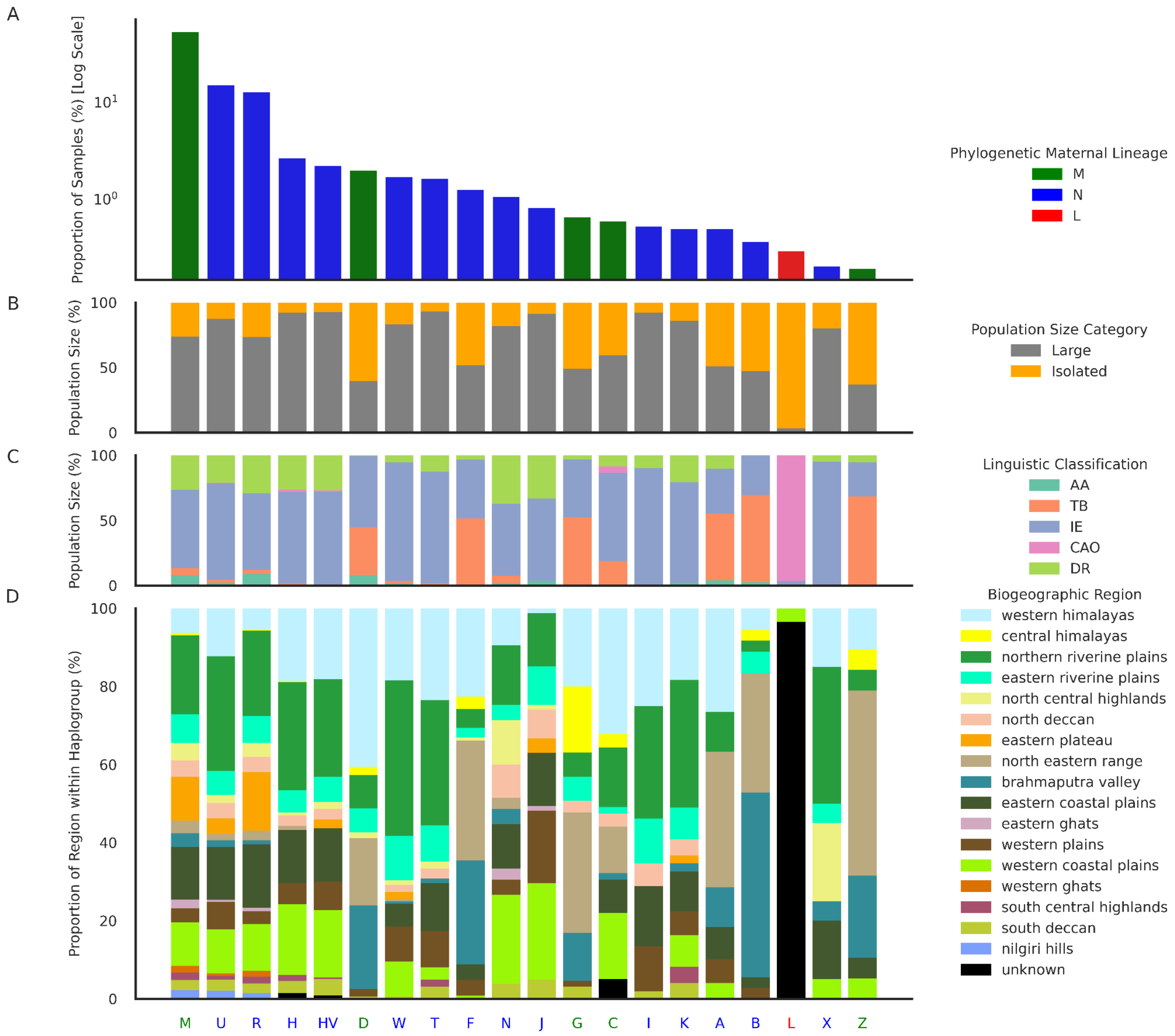

**Figure S5.1.2**. Distribution of mitochondrial haplogroups across India by frequency, population size, linguistic class and biogeographic region. (A) Proportion of individuals (log-scaled) assigned to each mitochondrial haplogroup, colour-coded by macro-haplogroup lineage: M (green), N (blue), and L (red). (B) Proportion of samples in each haplogroup derived from isolated (gray) vs. large (orange) populations. (C) Linguistic affiliation of populations contributing to each haplogroup: Austroasiatic (AA), Tibeto-Burman (TB), Indo-European (IE), Continental African

Origin (CAO), and Dravidian (DR). (D) Biogeographic distribution of each haplogroup, with stacked bars representing the proportion of individuals from various regions of India. Each bar corresponds to a distinct mitochondrial haplogroup, ordered on the x-axis by descending frequency.

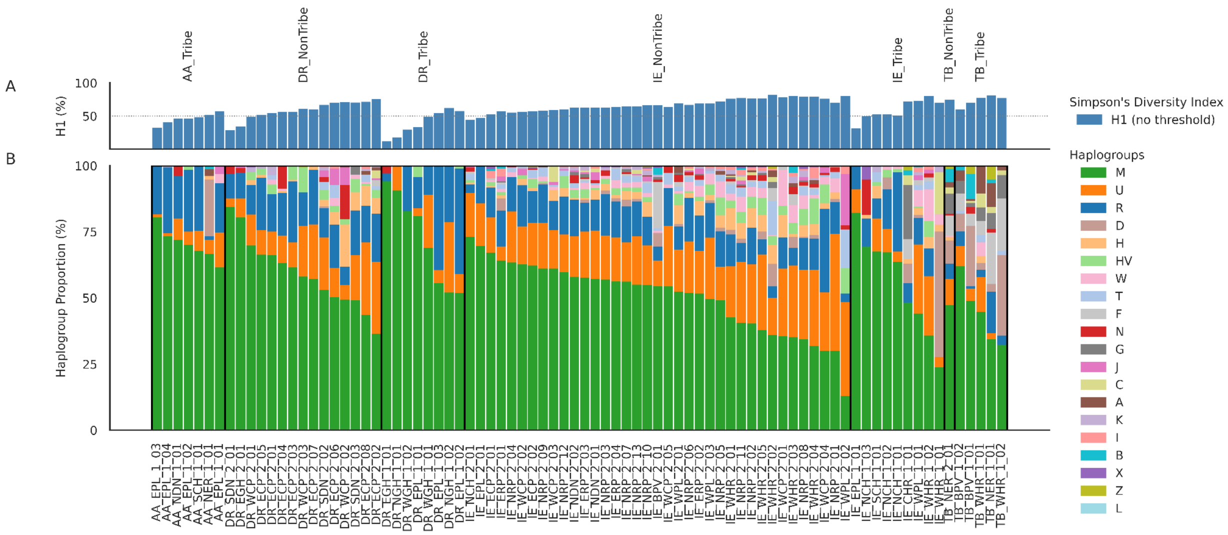

**Figure S5.1.3**. Mitochondrial haplogroup frequencies and diversity across ethnic groups. (A) The Simpson's Diversity Index (H1) is expressed as a percentage for each ethnic group, representing mitochondrial haplogroup diversity without any frequency threshold. The horizontal dotted line at 50% serves as a reference. (B) Mitochondrial haplogroup proportions (%) for each ethnic group, color-coded according to haplogroup identity. Haplogroups were filtered and ordered based on their overall frequency and assigned distinct colours. Ethnic groups are grouped and sorted by their linguistic classification and haplogroup M frequency. The diversity index highlights variation in haplogroup heterogeneity, while the stacked bars illustrate the relative haplogroup composition within each population.

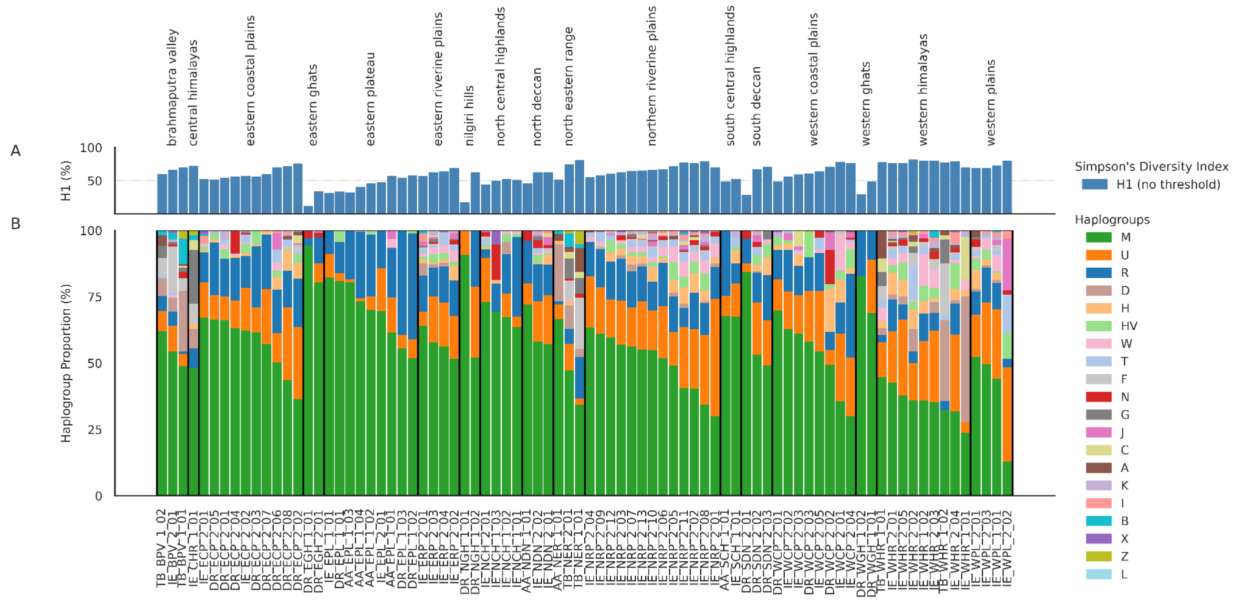

**Figure S5.1.4** Distribution and diversity of mitochondrial haplogroups across Indian ethnic groups, organized by biogeographic regions. (A) Simpson's diversity index (*H1*) is shown for each group, expressed as a percentage. The dotted line at 50% provides a reference for intermediate diversity. (B) Proportional composition of mitochondrial haplogroups within the same groups. Haplogroups are represented in distinct colors according to the haplogroup color scheme. Ethnic groups are arranged by biogeographic region, with bounding boxes marking regional clusters along the x-axis. Together, the panels illustrate variation in haplogroup diversity and composition across both ethnic and biogeographic classifications, highlighting regional patterns of maternal lineage structure.

#### Haplogroup proportions and admixture

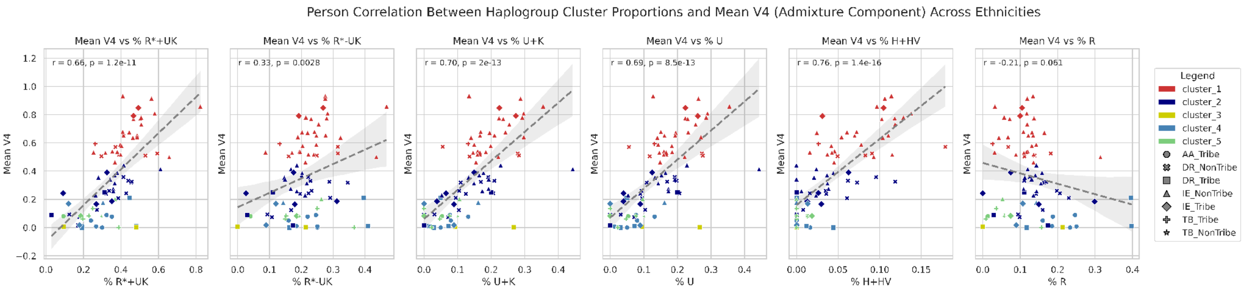

**Figure S5.1.5**. Pearson correlation between the proportion of individuals with specific mitochondrial haplogroup clusters and the mean V4 ancestry component—interpreted as a proxy for Ancestral North Indian (ANI) ancestry—across Indian ethnic groups. Each subplot corresponds to one of six defined haplogroup clusters: R+UK*, R-UK*, U+K, U, H+HV, and R. The x-axis shows the proportion of individuals within an ethnicity belonging to the respective haplogroup cluster, and the y-axis shows the mean V4 (ANI) ancestry score for that ethnicity. Ethnic groups are colored according to their assigned admixture cluster (Cluster 1–5), which were defined based on genome-wide V4 ancestry proportions. Marker shapes reflect linguistic classification. A dashed grey regression line and shaded 95% confidence interval are plotted in each panel. Pearson correlation coefficients (r) and corresponding p-values are noted in the top-left of each subplot.

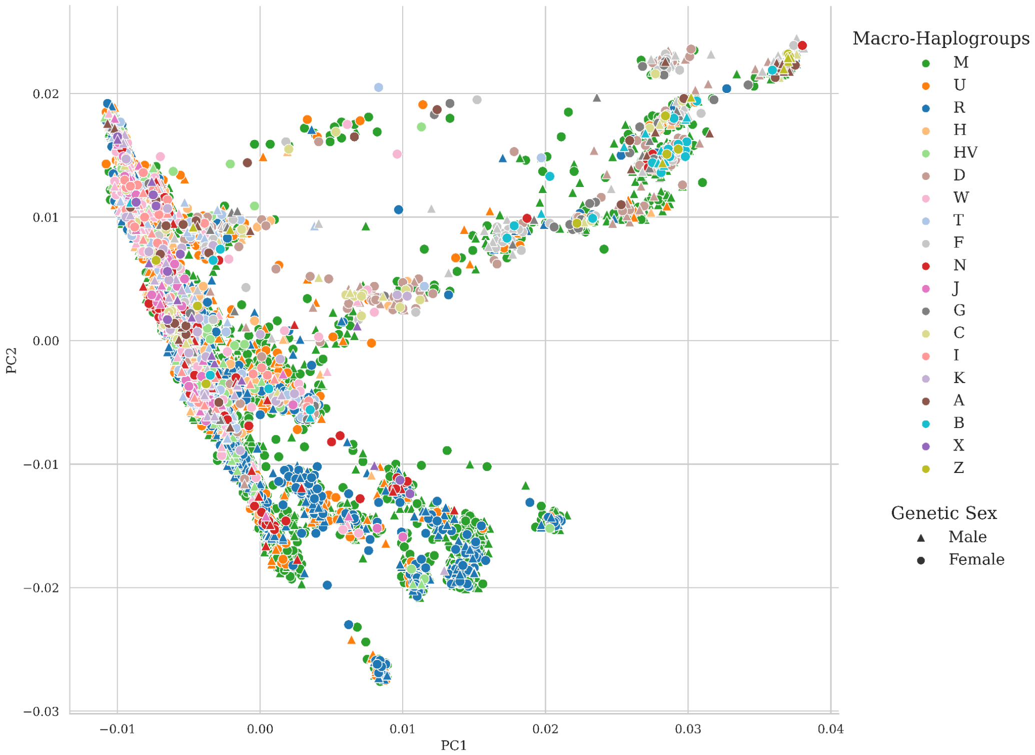

**Figure S5.1.6.** Principal component analysis (PCA) of 9,768 GenomeIndia samples based on autosomal genetic variation. Each point represents an individual, colored according to their mitochondrial haplogroup and ordered by frequency across the dataset. Haplogroups belonging to macro-lineages M, N, R, U, and other subclades are distinguished by distinct colors, as defined in the study's haplogroup color scheme. The shape of the points represents genetic sex, with upward triangles denoting males and circles denoting females. Haplogroups from African macro-lineage L were excluded for clarity.

#### Pathogenic variants in the mitochondrial genome

To obtain the list of high-confidence mtDNA variants, we performed a list of filtering steps on the joint-called VCF file as detailed in Laricchia et al. 2022.

*Variant filtering:* We filtered out a predefined list of artifact-prone sites (positions 301, 302, 310, 316, 3107, 16182) since these sites have sequence contexts that make it difficult to distinguish actual variants from technical artifacts. Additionally, we applied the filter "indel_stack" to remove indels that are only present within multi-allelic calls across all samples in the set of variants. DRAGEN itself imposes 8 other filters (base_quality, low_af, low_frac_info_reads, weak_evidence, mapping_quality, no_reliable_supporting_read, read_position and too_few_supporting_reads). We filtered out all the variants that were flagged by these filters, and retained only the variants marked PASS, over and above the artifact prone sites and indel stack filters. After filtering, we annotated variants with the Ensembl Variant Predictor tool (McLaren et al. 2016).

*Heteroplasmy estimation*: We calculated the heteroplasmic fraction (HF) for each variant for each sample from the information in the 'Allelic Depth' field of the VCF. HF was defined as a ratio of the number of reads supporting the variant/total number of reads at that position. We only included variants with heteroplasmic fraction ≥ 0.10 to avoid NUMT-derived false positive variants. Variants with HF in the range of 0.90-1.00 were denoted as homoplasmic and variants with HF < 0.90 (and ≥ 0.10) were denoted as heteroplasmic.

*Pathogenicity annotation*: We annotated all PASS marked variants with publicly available data files from MITOMAP (Lott et al. 2013) (disease mutations, download date 14 October 2024) to infer the clinical significance of the filtered mtDNA variants in our cohort. We used 126 MITOMAP variants having **Cfrm (confirmed)** status, being reported by at least two or more laboratories looking at unrelated families. Additionally, we also report the variants which have consequences marked as start-lost, stop-gained and stop-lost, along with their SIFT, PolyPhen and APOGEE2 scores wherever applicable (for the variants in the protein-coding region).

*Results*

After filtering the variants, there were 5,877 unique variants from 5,258 unique positions. These represent 31.73% of the mitochondrial genome. These 5,877 variants include 5,667 SNVs, 141 insertions and 69 deletions. Of these variants, 4,317 (73.46%) are homoplasmic only, while 417 are heteroplasmic only (7.1%), and 1,143 both heteroplasmic and homoplasmic (19.45%). 5,228 (88.96%) of the variants are rare (MAF ≤ 0.5%). 587 (9.99%) of the variants are low-frequency (0.5% < MAF ≤ 5%), and 62 (1.05%) of the variants are common. (Figure S5.1.7A). The allele frequency spectrum of all high-confidence variants can be seen in Figure S5.1.7B.

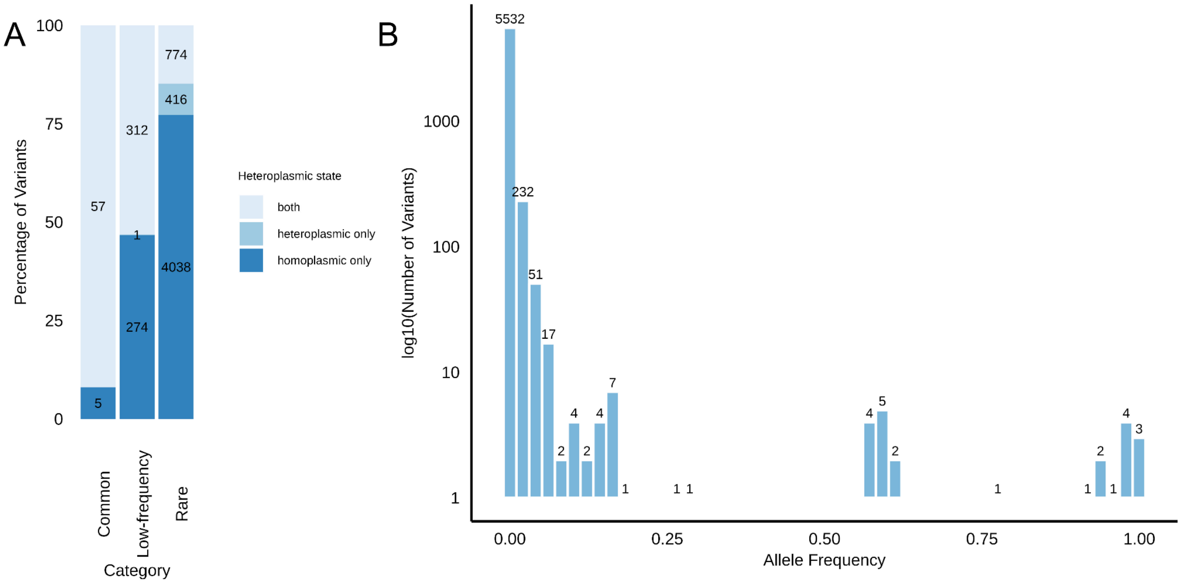

**Figure S5.1.7.** (A) Distribution of heteroplasmies for common, low-frequency, and rare mtDNA variants. The numbers inside the bars indicate the numbers of variants in each heteroplasmic state for each allele frequency category, and the colours denote variants that occur as homoplasmic only (dark blue), heteroplasmic only (light blue) and both (grey) (B) Distribution of allele frequencies of high-confidence mitochondrial variants in GenomeIndia. Bars show the number of variants in each allele frequency bin; values above bars indicate counts. The y axis is in the log(10) scale.

Of the 5,877 variants, 4,163 (70.84%) are in the protein coding region, followed by 815 (13.87%) in the control region, 450 (7.66%) in the rRNA coding, 369 (6.28%) in the tRNA coding and 80 (1.36%) in the intergenic region. 36 individuals carry 11 known pathogenic variants (Table S5.1.1), representing a burden in 1 in 271 individuals. Of these variants, two variants (chrM:3460G>A and chrM:7445A>G) have a higher heteroplasmy in GI than has been observed in gnomAD, suggesting their potential pathogenicity (Figure S5.1.8A). Given the late age-of-onset of many of the conditions associated with these variants, a phenotypic confirmation of variant effects is not possible in this cohort.

Besides known pathogenic variants, we also identified 10 start-lost, stop-gained, or stop-lost variants in the GenomeIndia data (Table S5.1.2). These include two variants (chrM:3307A>G and chrM:7444G>T) which are absent in gnomAD. The comparison of heteroplasmic fractions of these variants in GenomeIndia vs gnomAD is shown in Figure S5.1.8B.

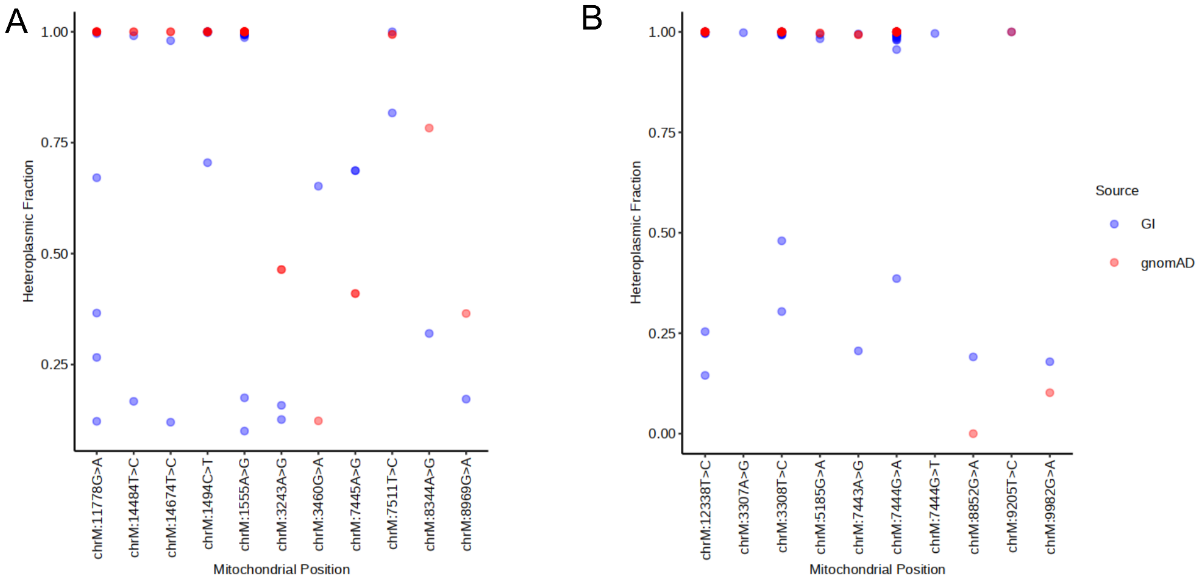

**Figure S5.1.8.** (A) Comparison of heteroplasmic fractions for known pathogenic variants between GenomeIndia (GI) and gnomAD (B) Comparison of heteroplasmic fractions for start-lost, stop-gained, and stop-lost mitochondrial variants between GenomeIndia (GI) and gnomAD

#### Section S5.2 - ChromosomeX

##### QC and filtering of variants

Variant-level quality filters for chrX were applied similar to those used for autosomes, i.e., required to pass the internal DRAGEN filters, QUAL ≥ 30, exhibit call rates ≥ 98%, and at least one sample with GQ > 20. However, applying the Hardy-Weinberg equilibrium (HWE) filter to chromosome X posed challenges due to males possessing only a single X chromosome, rendering biallelic sites incompatible with standard HWE assumptions. To address this, HWE p-values were recalculated exclusively for female samples. Variants significantly deviating from equilibrium ($p \leq 10^{-11}$) were excluded from both female and male datasets, resulting in 6,059,950 biallelic variants on chrX.

We further performed a Mendelian concordance check on chrX for the trios. Variants located in the centromeric and pseudoautosomal (PAR) regions were excluded for this analysis, and we specifically examined variants present in the child but absent in both parents. Given the hemizygosity of chrX in male offspring, any heterozygous calls would indicate potential errors or inconsistencies. After applying quality control filters, concordance rates improved, ranging from 99.7% to over 99.9% (Fig. S5.2.1). These results indicate high accuracy of the variant calls, minimal Mendelian inconsistencies, and confirm the reliability of the chrX genotypes for downstream analyses.

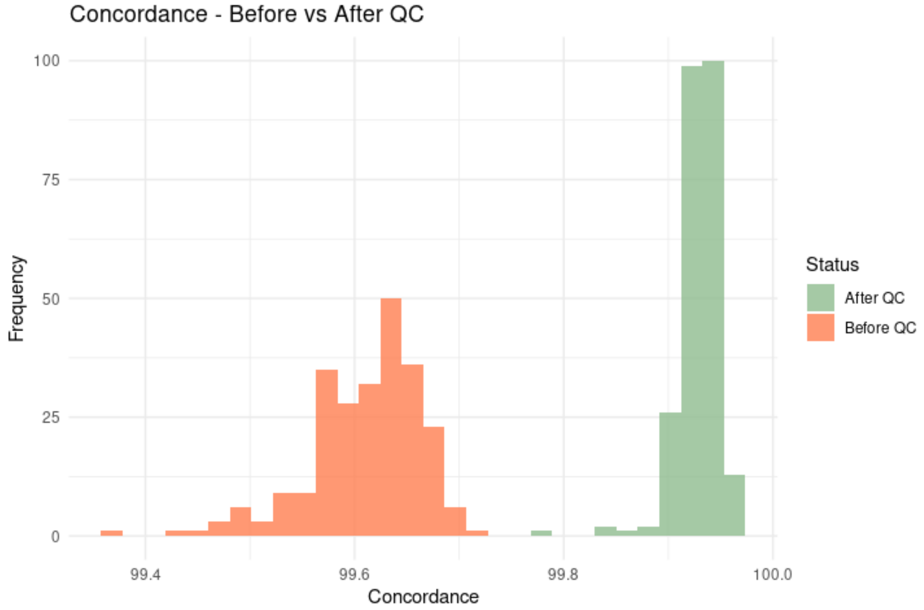

**Figure S5.2.1.** Mendelian concordance rates on chrX for 244 trios before and after QC. Concordance improved from 99.4–99.7% to 99.7–99.9% following QC, reflecting the efficacy of the applied filtering steps.

#### Annotation and identification of pathogenic variants

Chromosome X variant annotation using the VEP, in alignment with the strategy employed for autosomes, enabled the assessment of potential functional impacts. VEP categorized 1,110 variants as HIGH impact, 22,700 as MODERATE, 19,216 as LOW, and the remainder as MODIFIER (Fig S5.2.2). For the HIGH impact variants, we further looked for loss of function (LoF) variants annotated to be of high confidence (HC). These were then filtered for variants that have less than 1% occurrence in any of the 1000Genome or gnomAD populations as well as in the Genome India dataset itself. These are referred to as loss-of-function (LoF) variants and in chrX we identified 587 such variants distributed across 243 unique genes, with 123 genes harboring 2 or more variants.

ClinVar annotation further identified 17 variants as pathogenic or likely pathogenic (Table S5.2.1), of which 16 carried a clinical significance assertion of two-stars and one a three-star, reflecting strong to moderate confidence based on concordance among multiple submitters in clinVar. Notably, 11 of these 17 variants were located within the *G6PD* gene, consistent with prior reports of a higher burden of pathogenic *G6PD* variants among South Asian populations. Of the 17 clinVar pathogenic variants, only two - both within the G6PD gene had a MAF greater than 0.01 in the GI dataset, with the remaining being rare. Furthermore, all but four of these variants were observed only in the heterozygous state.

#### G6PD gene variants in Indian populations

Glucose-6-phosphate dehydrogenase (G6PD) protects erythrocytes from oxidative damage, and deficiency caused by G6PD-impairing variants predisposes individuals to hemolytic anemia under oxidative stress. These variants occur at high frequencies in malaria-endemic regions, where partial G6PD deficiency confers protection against severe *Plasmodium falciparum* infection, reflecting long-term balancing selection between malaria resistance and hemolytic risk (Luzzatto et al., 1969). This relationship also has important pharmacogenomic implications, as low G6PD activity increases susceptibility to drug-induced hemolysis during treatment with 8-aminoquinoline antimalarials such as primaquine (Luzzatto et al., 2020; Rajasekhar et al., 2024; Recht et al., 2014), whereas 4-aminoquinolines, including chloroquine and hydroxychloroquine, are considered relatively safer and less likely to cause hemolysis in G6PD-deficient individuals (Kane, 2012; Schilling et al., 2020). Several G6PD variants are well characterized, including rs78478128 (Orissa variant) prevalent among Indian tribal populations, rs137852314 (Mahidol variant) reported as the most frequent in Myanmar, and rs137852339 (Kerala variant) together with the Mediterranean variant (rs5030868), which represent the most common G6PD-deficient alleles across South and Southeast Asia (Devendra et al., 2020).

**Table S5.2.1.** Pathogenic and likely pathogenic chrX variants (ClinVar assertions ≥2 stars), their predicted consequence types, associated disorders, and allele frequencies in the dataset. All variants occur at low frequencies (AF < 0.01). Several variants map to the G6PD gene, which is associated with hemolytic anemia.

| #Variation | Consequence | Existing_ID | Gene Symbol | AF (GI) | ClinVar_condition |
|---|---|---|---|---|---|
| chrX_130136086_G/A | missense_variant | rs724160020 | AIFM1 | 0.0001 | Charcot-Marie-Tooth, neuropathy X, Combined oxidative phosphorylation deficiency |
| chrX_8699693_C/T | splice_donor_5th_base_variant,intron_variant | rs773138384 | ANOS1 | 0.0001 | Hypogonadotropic hypogonadism 1 with or without anosmia |
| chrX_154533596_C/G | missense_variant | rs137852318 | G6PD | 0.0004 | Anemia, nonspherocytic hemolytic, due to G6PD deficiency |
| chrX_154534390_G/A | missense_variant | rs137852330 | G6PD | 0.0024 | |
| chrX_154534419_G/A | missense_variant | rs5030868 | G6PD | 0.0036 | |
| chrX_154534495_C/T | missense_variant, splice_region_variant | rs137852314 | G6PD | 0.0077 | |
| chrX_154535247_G/A | missense_variant | rs979416826 | G6PD | 0.0001 | |
| chrX_154535277_T/C | missense_variant | rs1050829 | G6PD | 0.0008 | |
| chrX_154532389_C/T | missense_variant | rs137852324 | G6PD | 0.0007 | |
| chrX_154533044_C/T | missense_variant | rs137852339 | G6PD | 0.0127 | |
| chrX_154535963_G/A | missense_variant | rs138687036 | G6PD | 0.0001 | |
| chrX_154535996_A/G | missense_variant | rs137852349 | G6PD | 0.0009 | |
| chrX_154536168_G/C | missense_variant | rs78478128 | G6PD | 0.0157 | |
| chrX_101398011_C/T | missense_variant | rs111422676 | GLA | 0.0003 | Fabry disease |
| chrX_101400692_G/C | missense_variant | rs397515870 | GLA | 0.0001 | |
| chrX_38408988_G/A | missense_variant | rs66724222 | OTC | 0.0004 | Ornithine carbamoyltransferase deficiency |
| chrX_154953961_G/A | missense_variant | rs137852428 | F8 | 0.0001 | Hereditary factor VIII deficiency disease |

##### Novel genetic variants on chrX

To identify unreported genetic variation on chromosome X within the GI dataset, we systematically compared identified variants against four major public variant repositories: dbSNP, gnomAD, the 1000 Genomes Project Phase 3, and GenomeAsia100K. Variants were considered novel if their genomic coordinates along with reference/alternate alleles were absent from all four databases. Among the variants identified in the analysis as unique to the GI dataset, VEP categorised 2,526,947 as novel variants, including 2,350,337 SNVs and 176,610 indels. Functional annotation of these variants was performed using VEP to infer potential biological impact. The distribution of predicted variant consequences is summarized in Figure S5.2.2.

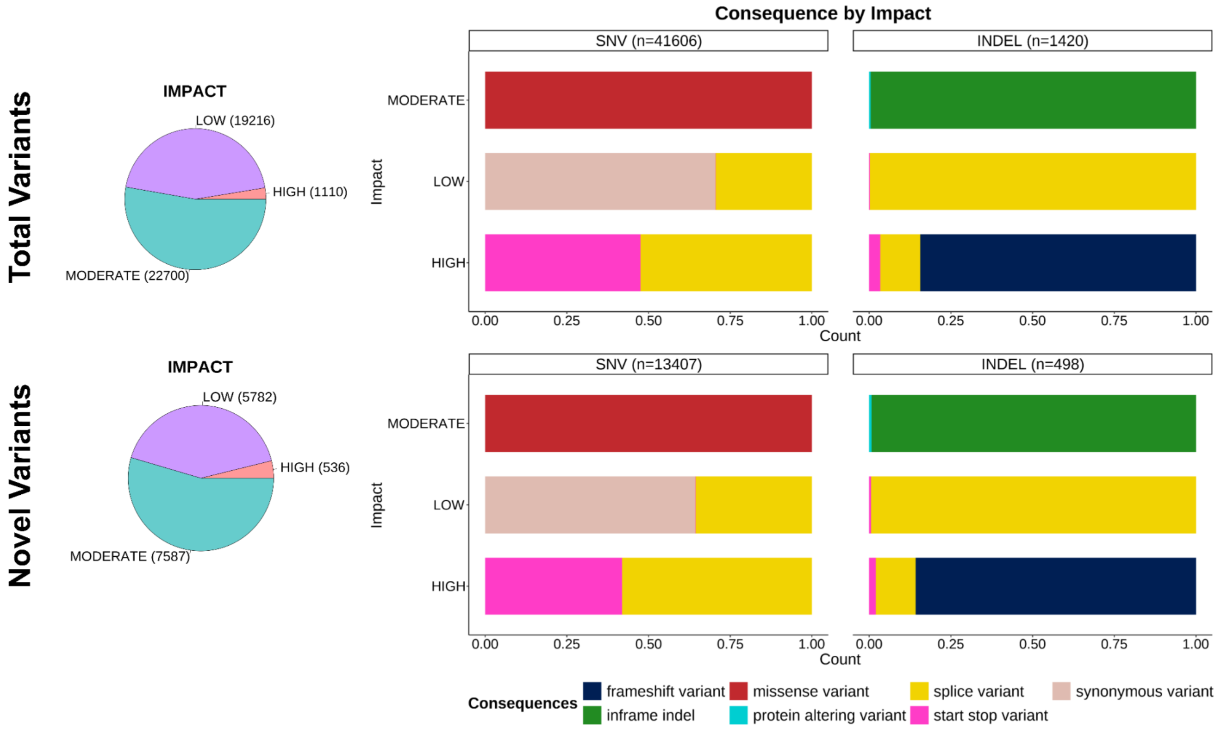

**Figure S5.2.2.** Pie charts depict the distribution of impact categories (LOW, MODERATE, HIGH) for total and novel variants in chrX. Bar charts illustrate the proportion of consequence types (frameshift, missense, start, inframe indel, splice, synonymous) within each impact category for SNVs and INDELs. MODIFIER variants (total=6,000,816 and novel=2,513,042) are not represented here.

#### Population specific variants of chrX

We assessed the distribution of chrX variants across the 82 populations by categorizing them as private (unique to the population), shared across some populations, or shared across all populations (Fig.S5.2.3). We identified that the majority of the variants were shared across all populations, with counts ranging from ~60,000 to 80,000 per population. A small fraction of the variants were population-specific, typically fewer than 600 variants per population. Variants shared among populations displayed an intermediate range (~5,000 to 10,000), possibly suggestive of the geographical or linguistic-level sharing of genetic variation.

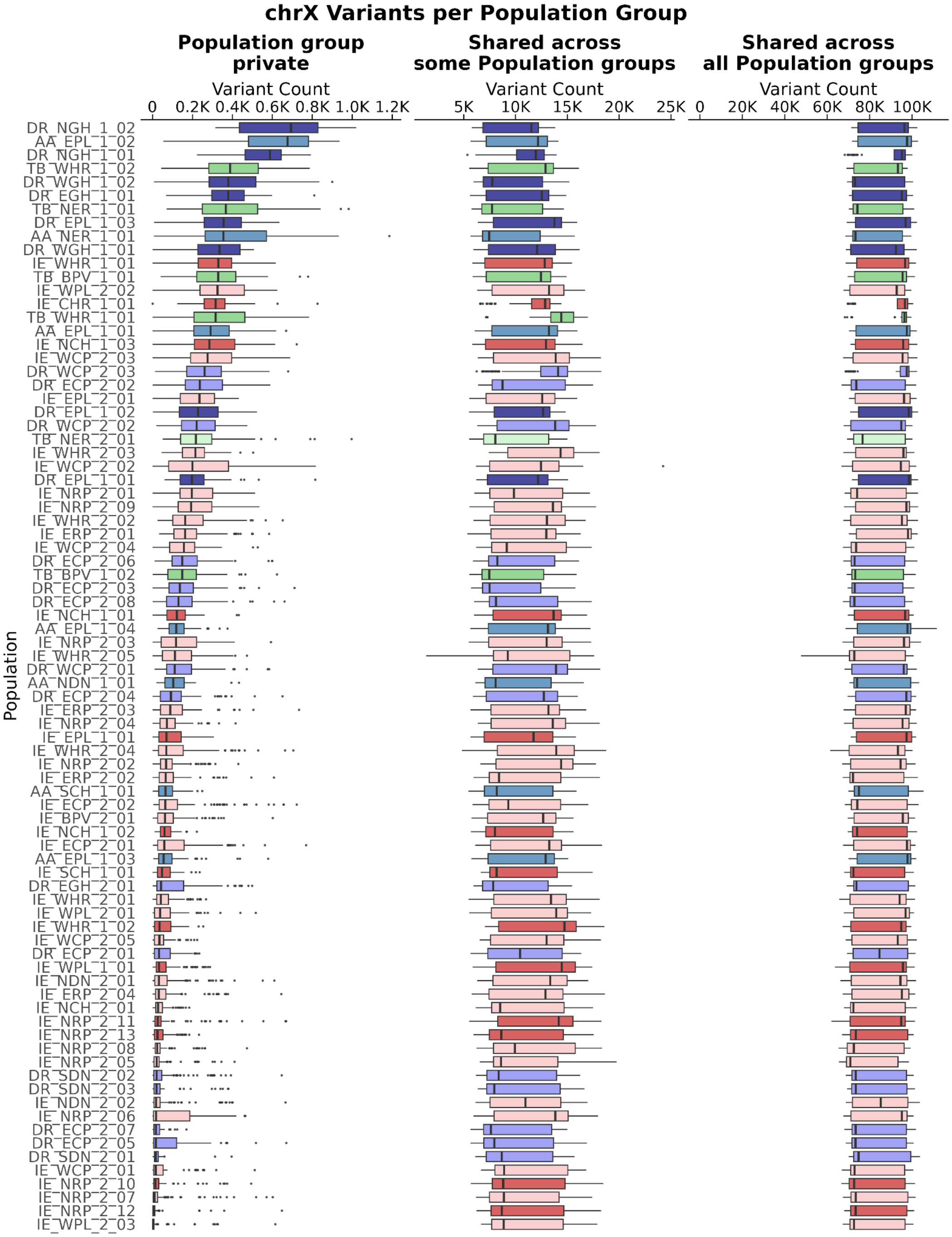

**Figure S5.2.3.** ChrX variants were categorized as private (restricted to a single population), shared across multiple populations (>1), or present in all populations. Tribal populations - primarily those speaking Dravidian and Tibeto-Burman languages - harbour the largest number of population-specific variants, consistent with long-term genetic isolation and reduced gene flow with neighboring groups. This pattern highlights the distinct evolutionary histories and demographic trajectories shaping ChrX diversity within these groups.

#### Section S5.3 - ChromosomeY

##### QC filtering of variants and annotation

A separate vcf file was generated for the chrY with only XY samples (n = 4,694). Variants were retained if they passed DRAGEN filters, had a QUAL score ≥ 30, call rates ≥ 98%, and at least one sample with GQ > 20. The resulting chrY call set comprised 230,879 genetic variants, of which 212,113 were biallelic. Variant annotation was performed using Ensembl VEP, which identified 43 HIGH impact variants - predominantly start-stop codon, frameshift, and some splice site changes, 158 MODERATE impact variants, and 289 LOW impact variants.

##### Novel genetic variants

To identify the unreported genetic variants across chrY, we compared the sites with 1000Genome phase3, gnomAD and dbSNP dataset, resulting in 131,097 as novel variants. These were further annotated by VEP to classify them into different impact classes (Fig. S5.3.1).

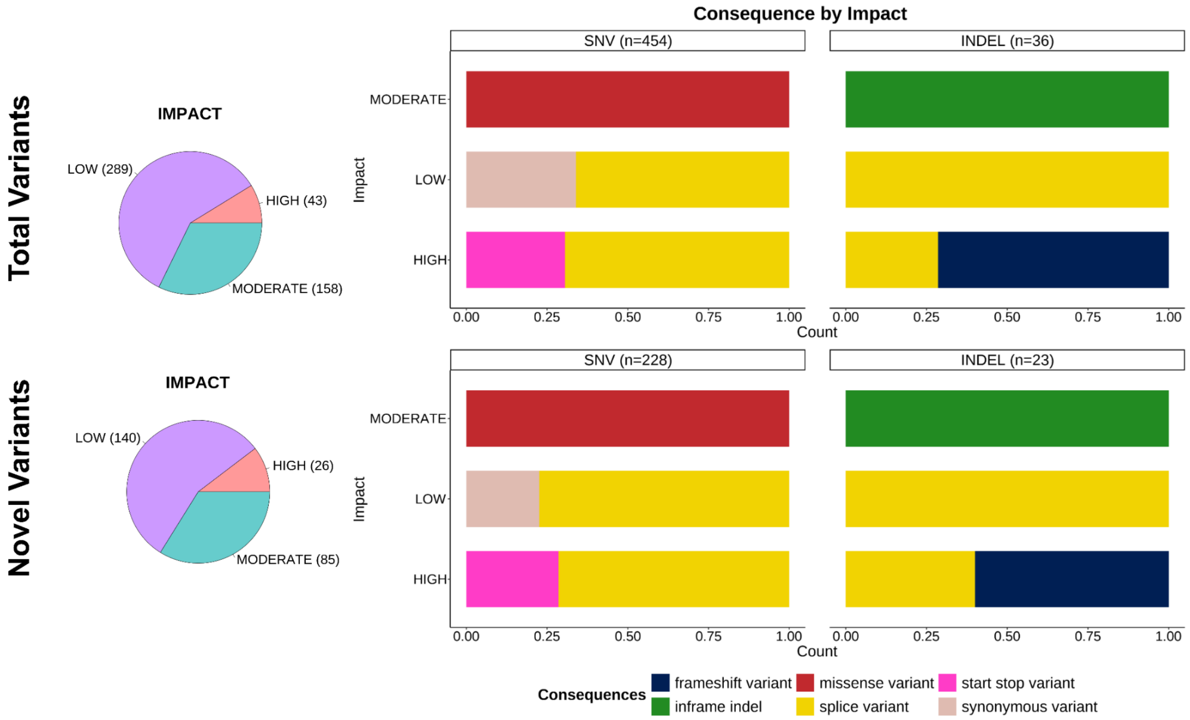

**Figure S5.3.1.** Pie charts depict the distribution of impact categories (LOW, MODERATE, HIGH) for total and novel variants in chrY. Bar charts illustrate the proportion of consequence types (frameshift, missense, start, inframe indel, splice, synonymous) within each impact category for SNVs and INDELs. MODIFIER variants (total=211,623 and novel=131,656) are not represented here.

**chrY haplogroups**

Given the strictly paternal mode of inheritance and the largely non-recombining nature of the Y chromosome (NRY), Y-chromosomal markers serve as powerful tools for tracing patrilineal ancestry and exploring population-level genetic diversity. We assigned Y-chromosomal haplogroups to all male individuals in the GI dataset using Y-LineageTracker v.1.3.0 (Chen et al., 2021). These Y-chromosomal haplogroup assignments were integrated with autosomal PCA coordinates to investigate patterns of genetic differentiation among the various linguistic groups (Fig S5.3.2). Additionally, their distribution was assessed across different linguistic groups (Fig S5.3.3) as well as geographical regions (Fig S5.3.4) to explore linguistic or region-specific haplogroup patterns.

ChrY haplogroups C, H, J, L1, O, R1, and R2 together comprise over >90% of male lineages in the GI dataset, with H1, R1, and R2 accounting for the majority. Distinct ethnolinguistic differences are evident (Fig S5.3.5 and Fig S5.3.6). The R haplogroups, particularly R1 and R2 are strongly associated with Ancestral North Indian (ANI) ancestry, linked to Central Asian and West Eurasian populations. Indo-European Non-Tribal populations are characterized by a massive dominance of R1 and R2. Haplogroup R1a1 shows its highest prevalence in the Indo-European populations. Haplogroup H1 (Yellow) is an ancient lineage, often called the Indian haplogroup, which originated in western India about 30,000 years ago (Mahal & Matsoukas, 2018). There is a pervasive presence of H1 across the entire subcontinent, particularly dominant in many Dravidian populations, supporting the hypothesis that Dravidian-speaking groups were possibly widespread throughout India before the arrival of Indo-European speakers. Additionally, the R component generally appears to decrease in proportion as one moves away from the North and West toward the South Deccan and Nilgiri Hills, reflecting the documented gradient of ANI ancestry running through the majority of IE and Dravidian populations.

Austroasiatic and Tibeto-Burman groups show extreme dominance of haplogroups O1 and O2 compared to the others. The near-total lack of R-lineages in TB_T and AA_T, separates them as a distinct cluster, supporting their entry from the northeastern corridor. The CAO group displays a unique Y-haplogroup profile marked by African-origin haplogroups B2, E1, and E2 (Underhill et al., 2001; Karafet et al., 2008; Gomes et al., 2010). Within the biogeographic groups, populations from the north-eastern region exhibit higher frequencies of haplogroups D1, O1, and O2, while those from the Eastern Plateau display relatively lower haplogroup diversity, with most individuals belonging to O1 and H1, aligning with the TB group. Interestingly, haplogroup R1 is absent in four of the nine Eastern Plateau populations, despite its presence across all other regional groups. These patterns underscore the complex interplay between geography, language, and patrilineal genetic structure in shaping population differentiation.

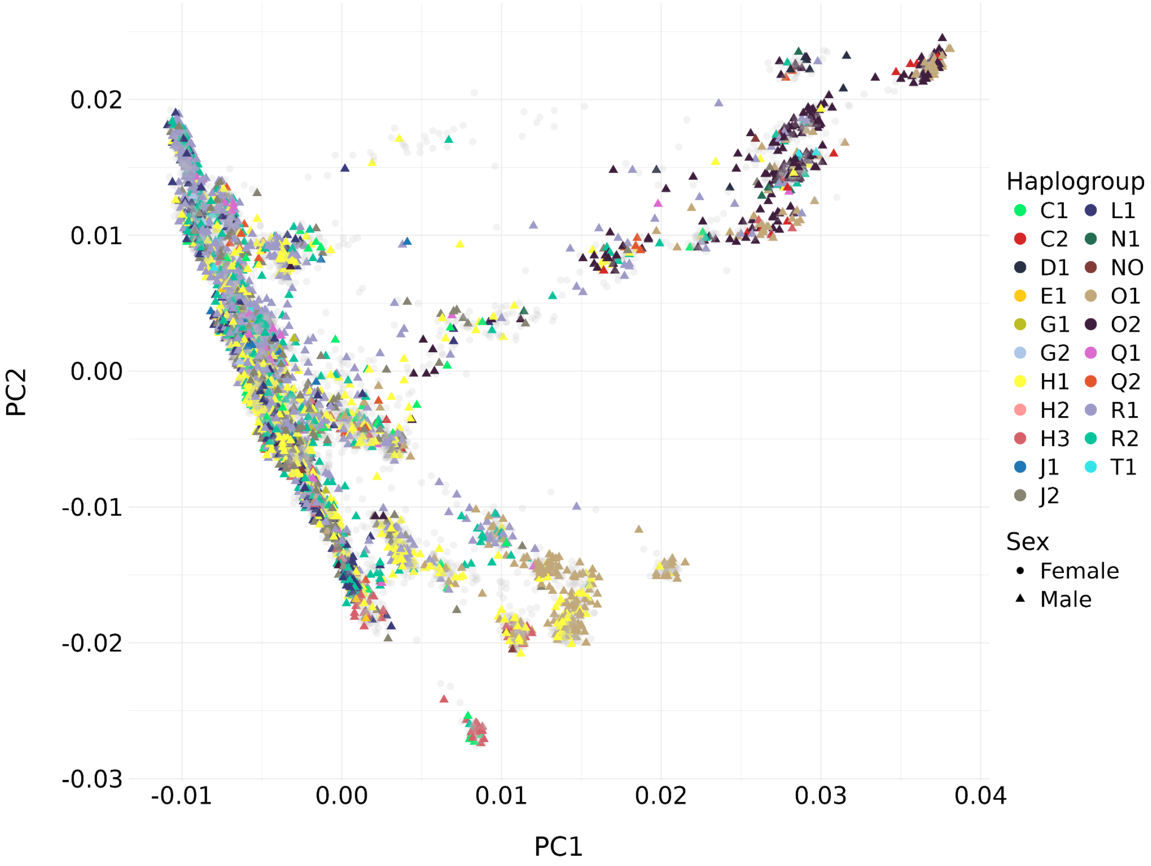

**Figure S5.3.2.** Principal Component Analysis (PCA) of the GenomeIndia samples, revealing clustering patterns based on chrY haplogroups. The plot displays the distribution of individuals across the first two principal components (PC1 and PC2). Each point represents an individual, with colors indicating different haplogroups (e.g., C1, L1, N1, etc.) and shapes distinguishing genetic sex. R1 constitutes the predominant haplogroup in the dataset. Individuals carrying O1 and O2 - haplogroups characteristic of Tibeto-Burman and Austroasiatic speakers form distinct clusters in the lower regions and upper-right of the plot, respectively. In contrast, the majority of samples belonging to the R1 and H1 haplogroups show substantial overlap, mirroring the well-documented continuum between ANI- and ASI-related ancestries across the Indian subcontinent.

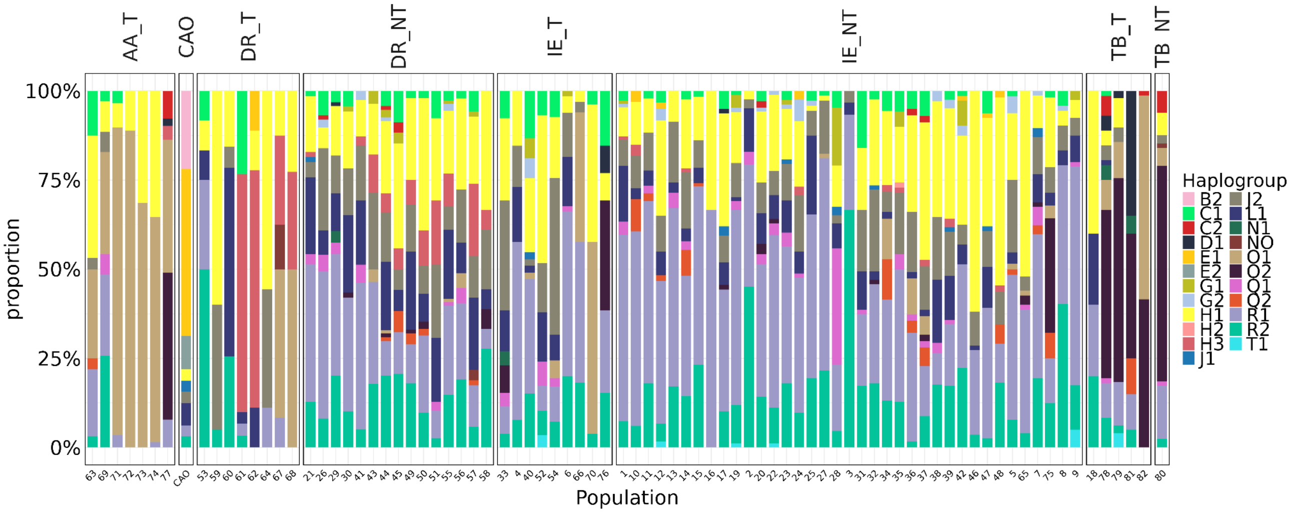

**Figure S5.3.3.** Frequency distribution of chrY haplogroups across major ethnolinguistic groups in the GenomeIndia dataset. H1 and R1 represent the two most prevalent haplogroups, though their relative frequencies vary across populations. Indo-European populations show a clear predominance of R1, with H1 constituting the next major lineage. Dravidian populations show higher frequencies of H1, H3, L1, J2 and R1; G1 and G2 are restricted to Dravidian non-tribals, indicating localized paternal lineages.Austroasiatic and Tibeto-Burman populations exhibit strong enrichment of O1 and O2, reflecting their distinct demographic histories and ancestral origins. In contrast, CAO populations carry a highly distinctive haplogroup composition dominated by B2 and E2 - lineages typically associated with African ancestry, highlighting their unique paternal genetic background within the Indian subcontinent.

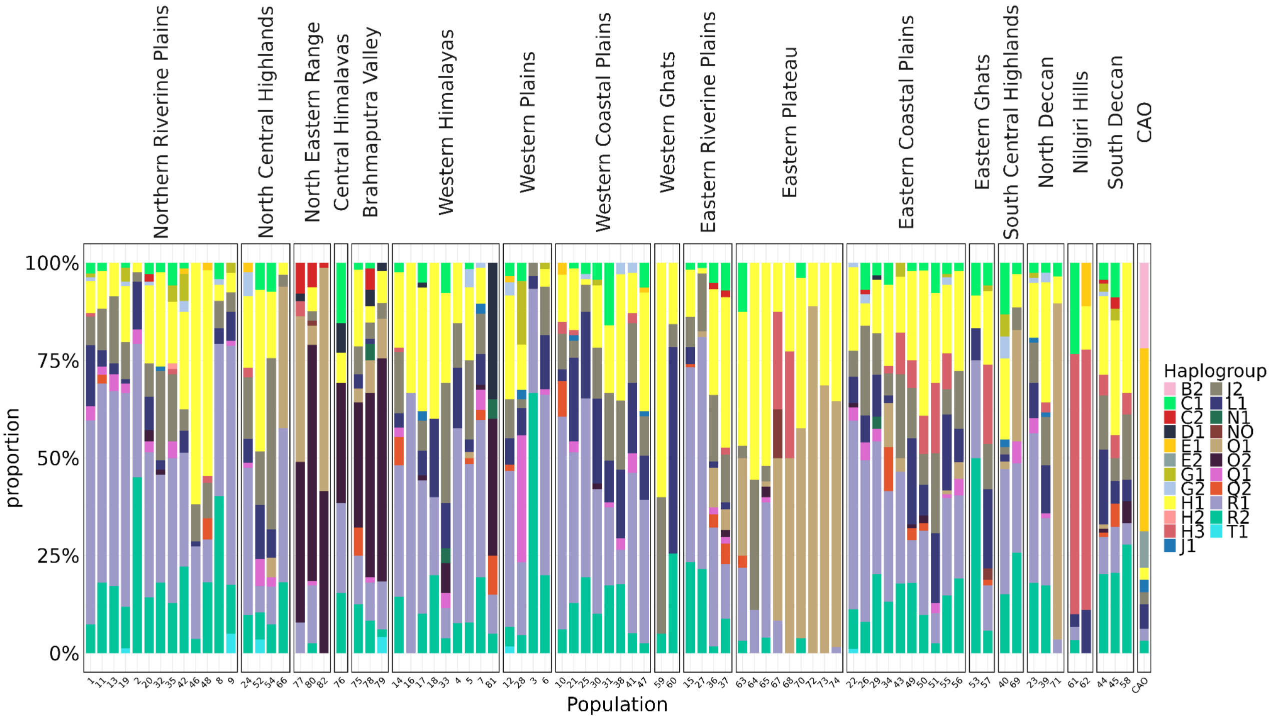

**Figure S5.3.4.** Frequency distribution of chrY haplogroups in GenomeIndia samples placed in their respective biogeographical groups. Geographic distribution reveals a north-south cline with R1 and R2 haplogroups being dominant in populations located in the Northern Riverine Plains and Western Plains. Haplogroup H1 is visible as a significant component in populations across nearly every geographic region, including the Northern Riverine Plains, Western Ghats, Eastern Plateau. The dominance of the O lineages in the North Eastern Range, Central Himalayas, and Brahmaputra Valley. This visually supports the finding that a major wave of humans entered India through the northeast corresponds to populations speaking Tibeto-Burman (TB) and Austro-Asiatic (AA) languages.

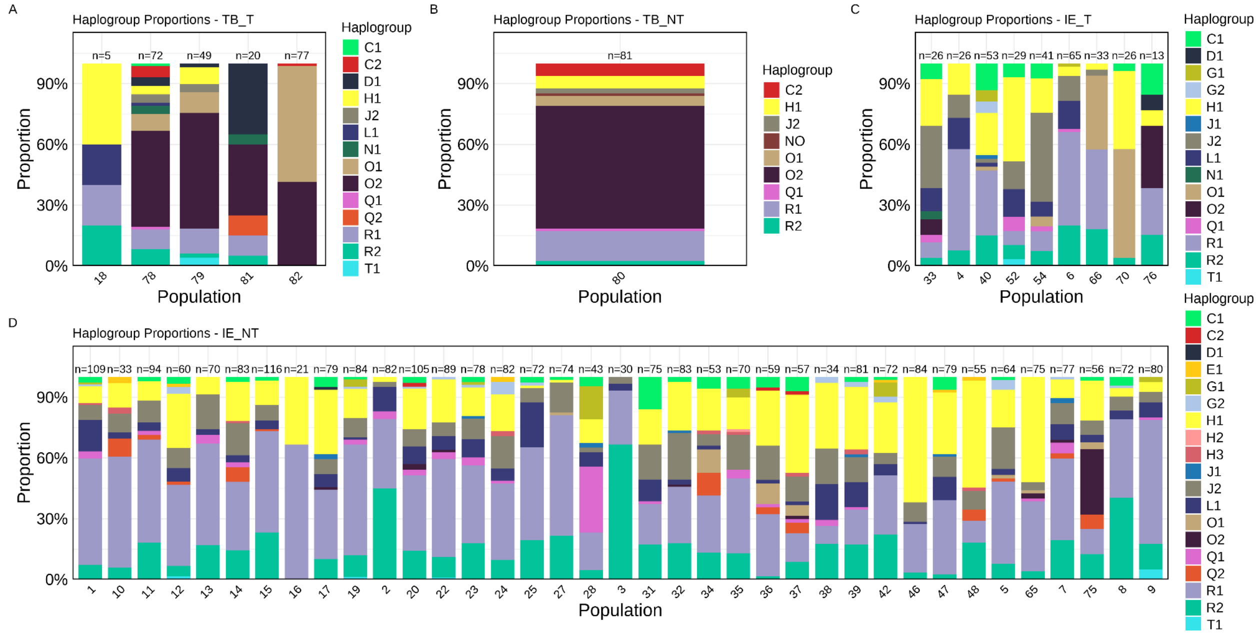

**Figure S5.3.5.** (A) Haplogroup distribution across the populations of AA tribe - characterized by a consistent dominance of O1 haplogroup across all.
(B) Y-chromosomal haplogroup composition in the CAO population. This group exhibits a highly distinctive paternal profile, dominated by haplogroups E1 and B2, both of which are typically of African origin. Additional presence of E2, along with minor contributions from J2, L1, R1, and R2, reflects limited admixture but preserves a predominantly African paternal signature.
(C) Y-chromosomal haplogroup composition in the Indo-European populations (IE_T). The haplogroup profiles reveal haplogroups R1 and H1 to form the predominant components across most populations, mirroring the typical paternal ancestry of South Asian Indo-European speakers.
(D) Distribution of Y-chromosomal haplogroups among Indo-European non-tribal (IE_NT) populations. The IE_NT group shows a clear dominance of the R1 haplogroup, which constitutes the major paternal lineage across nearly all populations. H1 is the next most frequent haplogroup and appears consistently across populations, followed by lower but notable contributions from haplogroups such as L1, J2, O1, O2 and R2, reflecting the complex demographic history of northern and central India.

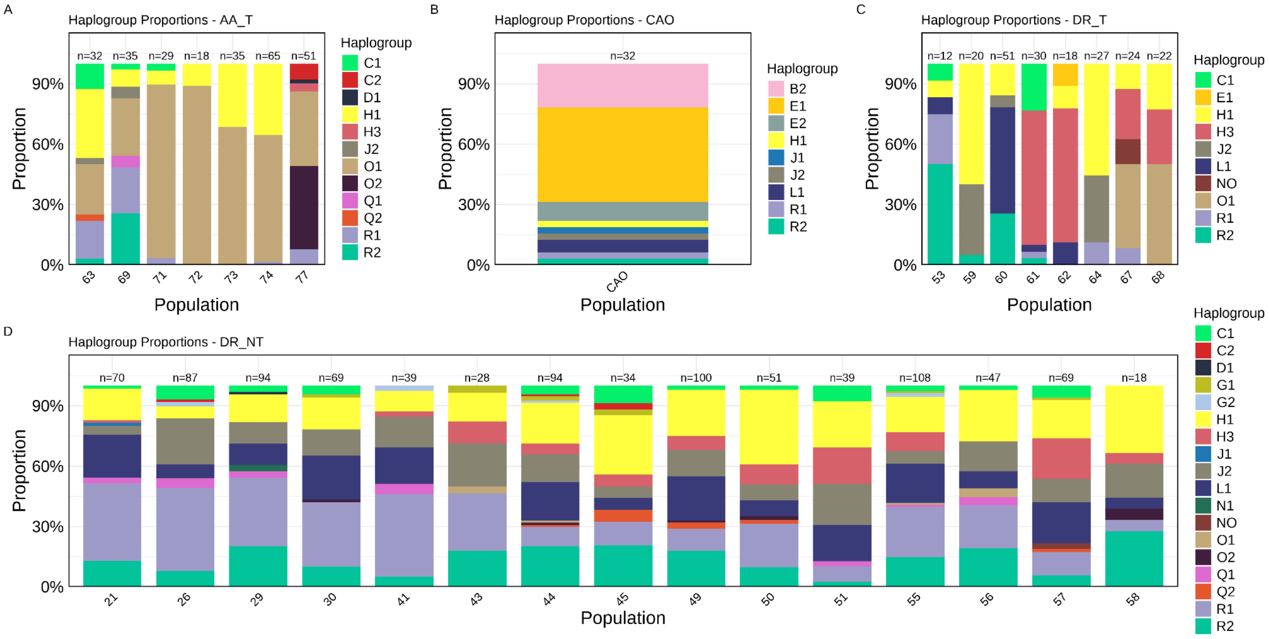

**Figure S5.3.6.** (A) The proportional distribution of Y-chromosomal haplogroups across the Tibeto-Burman tribal (TB_T) group. The TB_T group exhibits a consistent dominance of haplogroup O2, which constitutes the majority ancestry component across all populations, reflecting the characteristic paternal lineage associated with East Asian and Tibeto-Burman expansions. Few populations further show minor but notable contributions from haplogroups O1, J2, H1, and L1.
(B) Y-chromosomal haplogroup composition in Tibeto-Burman non-tribal (TB_NT) group. The plot shows a striking skew toward the O2 haplogroup, which dominates the paternal ancestry of this group. Smaller but consistent contributions from haplogroups O1, R1, H1, and C2 reflect secondary components of the paternal gene pool, while J2 is present at modest levels.
(C) Y-chromosomal haplogroup composition in the DR_T populations. They display haplogroup distributions dominated by H1 and R2, consistent with the deep antiquity of these lineages in peninsular India. Several populations exhibit strikingly high frequencies of haplogroup H3.
(D) Y-chromosomal haplogroup distribution across Dravidian non-tribal (DR_NT) populations. Dravidian groups show a broad and diverse haplogroup spectrum, with substantial contributions from H1, H3, L1, R1, and J2. Several populations also carry moderate levels of haplogroups O1, O2, and C1, demonstrating varying degrees of shared ancestry. Notably, G1 and G2 appear in some Dravidian non-tribal populations but are largely absent elsewhere, suggesting them to be population-specific.

#### Associations between autosomal ancestral components and paternal lineage frequencies

We further examined the relationship between autosomal ancestral components and the population-level proportion of the dominant Y-chromosomal haplogroup, stratified by ethnolinguistic affiliation. Indo-European and Dravidian groups show positive correlations with

ANI ancestry (Fig. S3.5.7), largely driven by the Y-haplogroups R1, R2 and H1. We then focused specifically on the R1a1 sub-haplogroup, given its well-documented widespread distribution and high frequency across Eurasia, Central Asia and the Indian subcontinent (Sharma et al., 2009). Consistent with previous reports, we observe that the distribution of R1a1 is strongly associated with Indo-European–speaking populations and with higher mean levels of ANI ancestry (Fig. S3.5.8).

Further analysis of correlations with other ancestral components revealed a statistically significant negative association in the DR non-tribal group, with ASI ancestry explaining approximately 63% of the variance in the proportion of the dominant Y-chromosomal haplogroup in the populations (Fig. S3.5.9). This pattern is consistent with previous studies showing that non-tribal Dravidian populations experienced disproportionate paternal gene flow associated with ANI-related ancestry, resulting in the expansion of Y-haplogroups such as R1a, R2 and H1 (Basu et al., 2016; Moorjani et al., 2013). A similar, though weaker, significant negative correlation is also observed in the Indo-European non-tribal group.

Extending this analysis to ASI-D ancestry, a distinct ancestry component from the Nilgiri hills, Indo-European and Dravidian populations, particularly the non-tribal populations show significant negative correlations with dominant Y-haplogroup frequencies (Fig. S3.5.10), whereas Tibeto-Burman populations show no meaningful association ($r = 0.092$). In contrast, correlations between AAA ancestry and dominant Y-haplogroup frequencies are weak and non-significant across Indo-European and Dravidian groups, indicating limited association (Fig. S3.5.11). Tibeto-Burman and Austroasiatic populations instead show modest positive trends, largely reflecting high frequencies of O-lineages. Finally, the ATB ancestry component shows its strongest and most consistent association within Tibeto-Burman populations themselves, with generally weak or negligible correlations in other groups (Fig. S3.5.12). This pattern suggests distinct demographic histories for Tibeto-Burman and Austroasiatic populations, consistent with their contrasting dominant Y-haplogroups - O2 in Tibeto-Burman populations and O1 in Austroasiatic, despite both lineages being broadly associated with East and Southeast Asian origins.

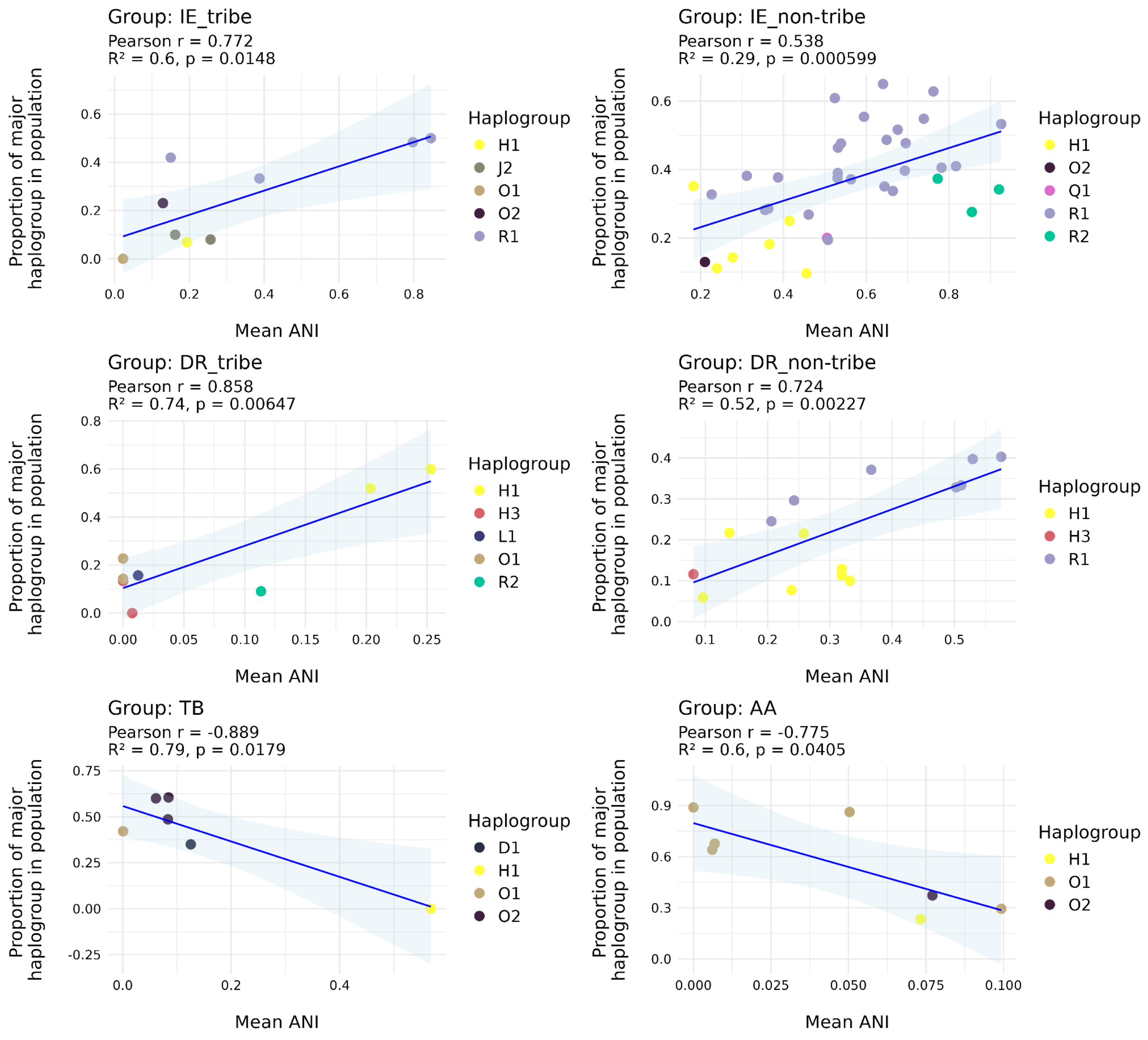

**Figure S5.3.7.** Relationship between autosomal ANI (Ancestral North Indian) ancestry and dominant Y-chromosomal haplogroups across the GI populations.

Each panel corresponds to an ethnolinguistic group and illustrates the relationship between the mean autosomal ANI (Ancestral North Indian) ancestry (x-axis) and the population-level frequency of the predominant Y-chromosomal haplogroup (y-axis). Each point represents a population and is coloured according to its major Y-haplogroup. Solid lines denote linear regression fits, with shaded areas indicating 95% confidence intervals. Pearson's correlation coefficient (r), $R^2$, and two-sided p-values are shown within each panel.

Positive correlations in Indo-European and Dravidian groups are driven primarily by haplogroups R1, R2 and H1, consistent with previous evidence that ANI ancestry reflects West Eurasian related gene flow into South Asia. In contrast, Tibeto-Burman and Austroasiatic populations show negative correlations, largely reflecting high frequencies of O1 and O2 lineages, which are characteristic of East and Southeast Asian paternal ancestry in these groups.

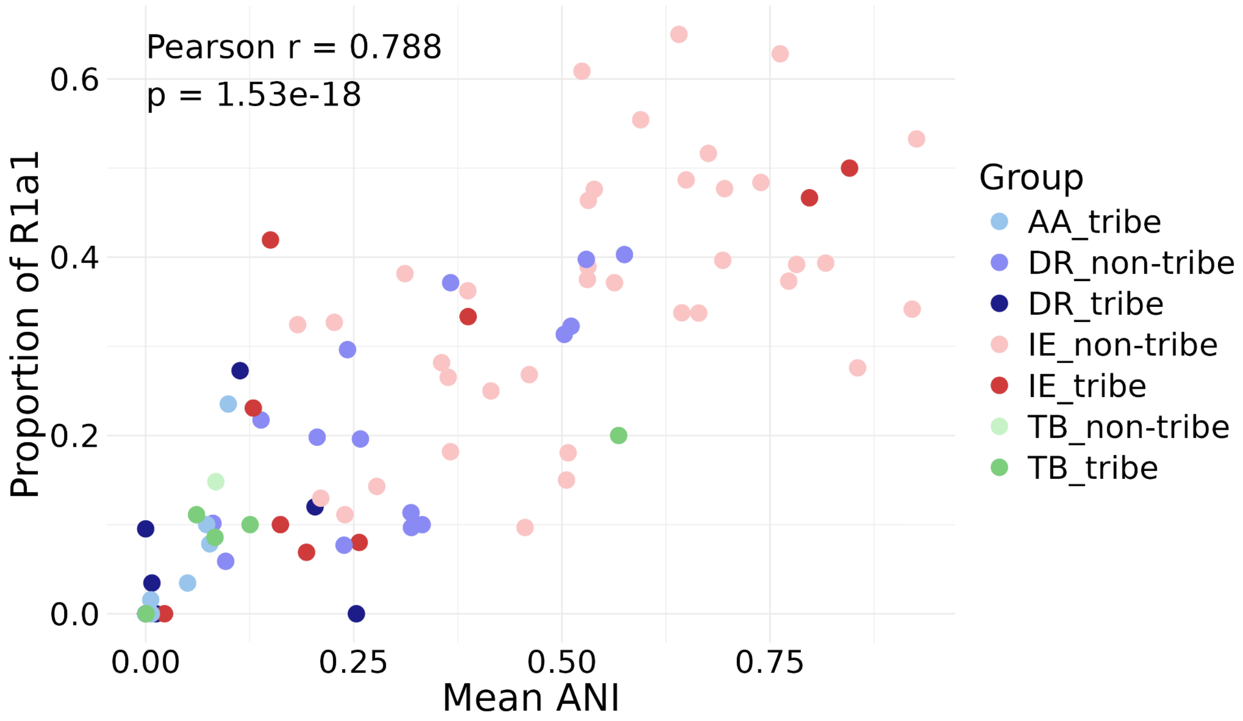

**Figure S5.3.8.** Relationship between genome-wide ANI ancestry (mean) and Y-chromosome haplogroup R1a1 frequencies (proportion of individuals with R1a1 haplogroup) across the populations, colored by their ethnolinguistic category A strong positive correlation is observed between mean ANI and R1a1 frequency (Pearson r = 0.788, $p = 1.53 \times 10^{-18}$), indicating that populations with greater contributions from ANI-related ancestry tend to exhibit higher frequencies of R1a1. Indo-European non-tribal groups occupy the upper range of both ANI and R1a1, followed by the Dravidian populations. The Austroasiatic, and Tibeto-Burman tribal populations cluster at low ANI and low R1a1 frequencies.

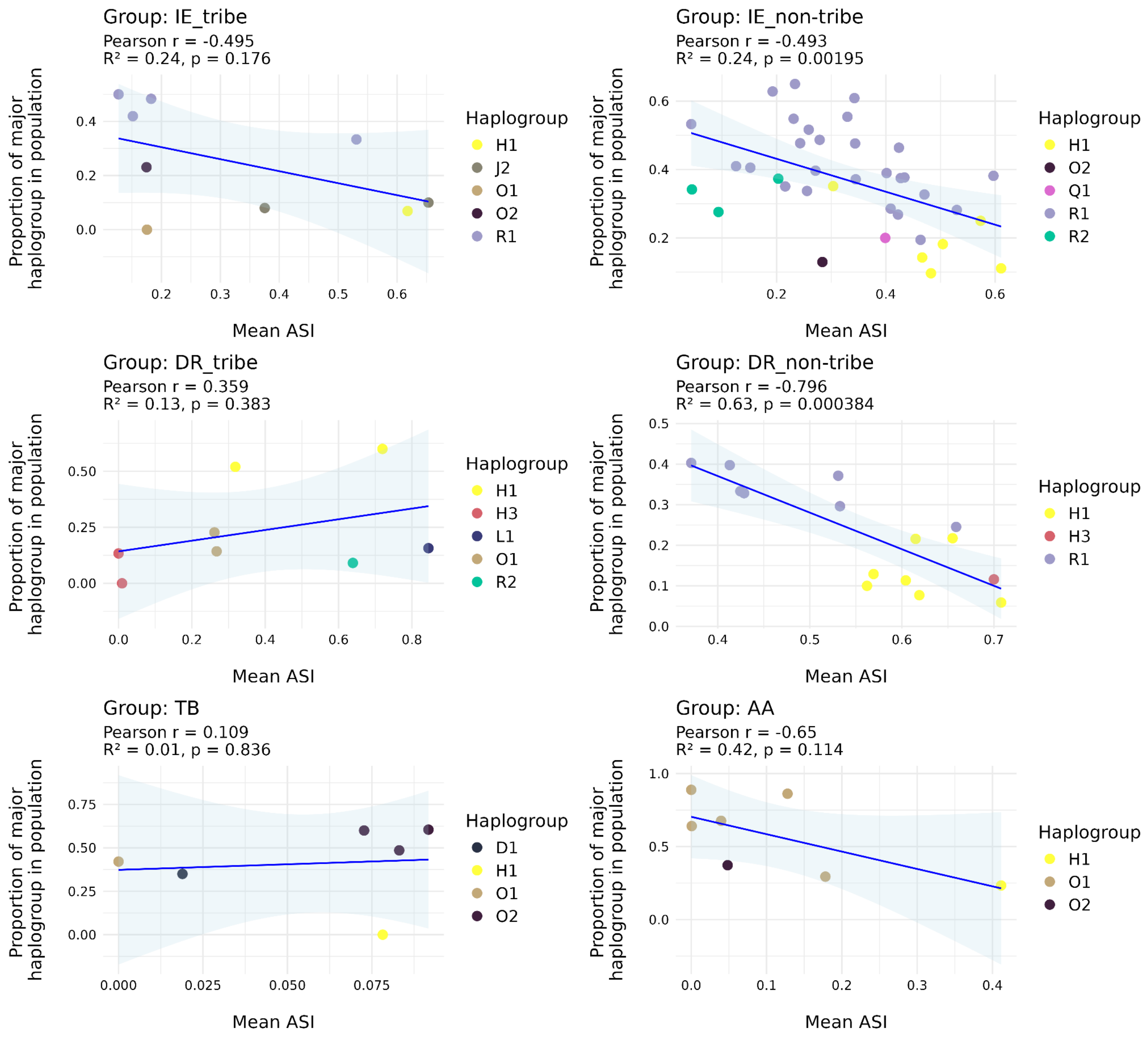

**Figure S5.3.9.** Relationship between autosomal ASI (Ancestral South Indian) ancestry and dominant Y-chromosomal haplogroups across the GI populations. The Indo-European and Dravidian non-tribal groups show negative correlations between ASI ancestry and the frequency of their dominant Y-haplogroups, particularly those enriched in ANI-associated lineages such as R1, R2 and H1, consistent with the reciprocal relationship between ANI and ASI components. Dravidian tribal populations, however, show a non-significant ($p>0.05$) weak positive correlation. In contrast, Tibeto-Burman populations show little to no association, whereas the Austroasiatic group tends toward negative correlation, reflecting the persistence of O-lineages despite variation in ASI ancestry.

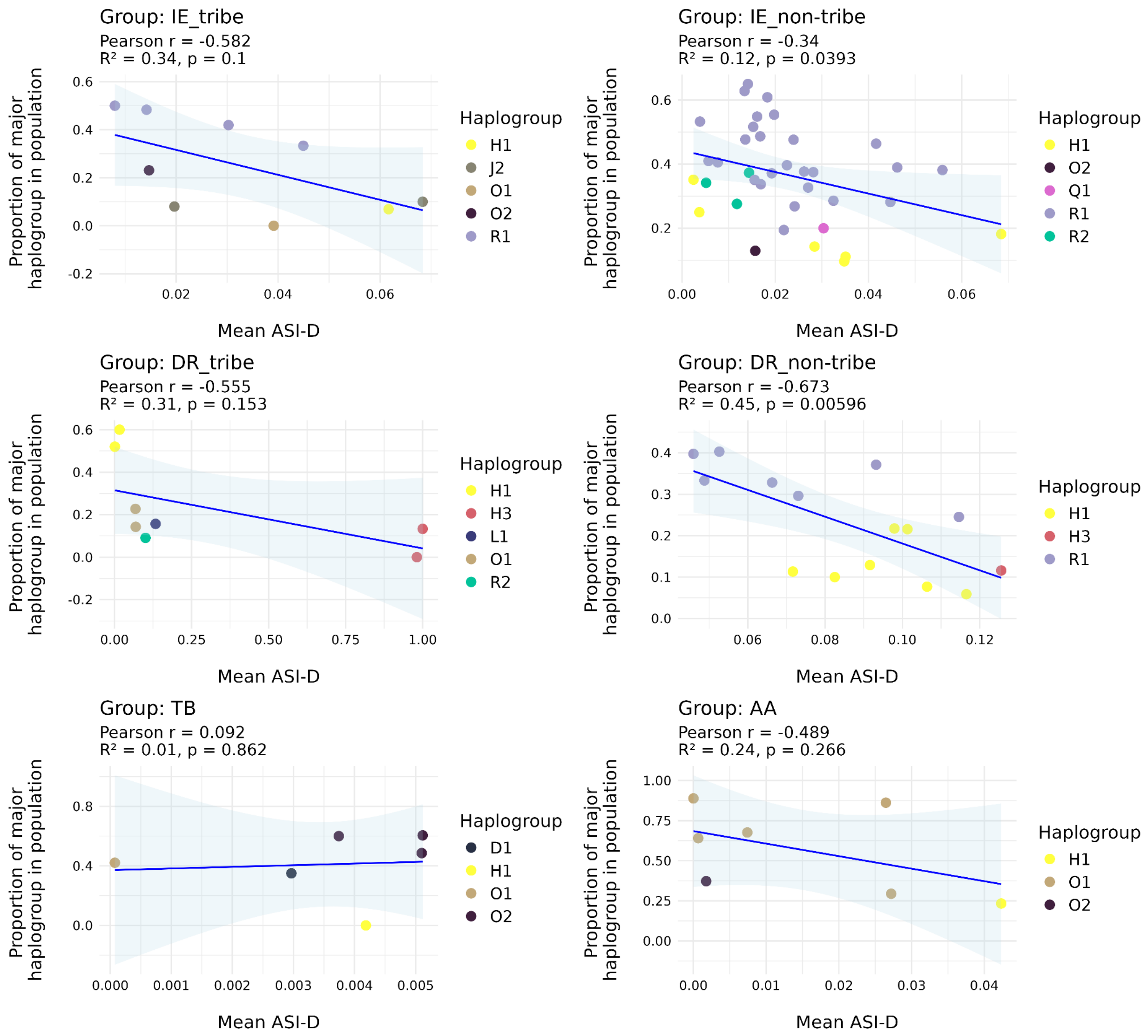

**Figure S5.3.10.** Relationship between autosomal ASI-D (a distinct ancestral component, from the Nilgiri hills) ancestry and dominant Y-chromosomal haplogroups across the GI populations. The Mean ASI-D component is associated with negative correlations in all groups, except the Tibeto-Burman populations, achieving significance primarily in non-tribal populations. A statistically significant negative correlation was found in the DR_non-tribe population, where the ASI-D component accounts for 45% of the variance (R2=0.45). A significant, weaker negative correlation was also present in the IE_non-tribe group. The TB group showed a negligible positive correlation.

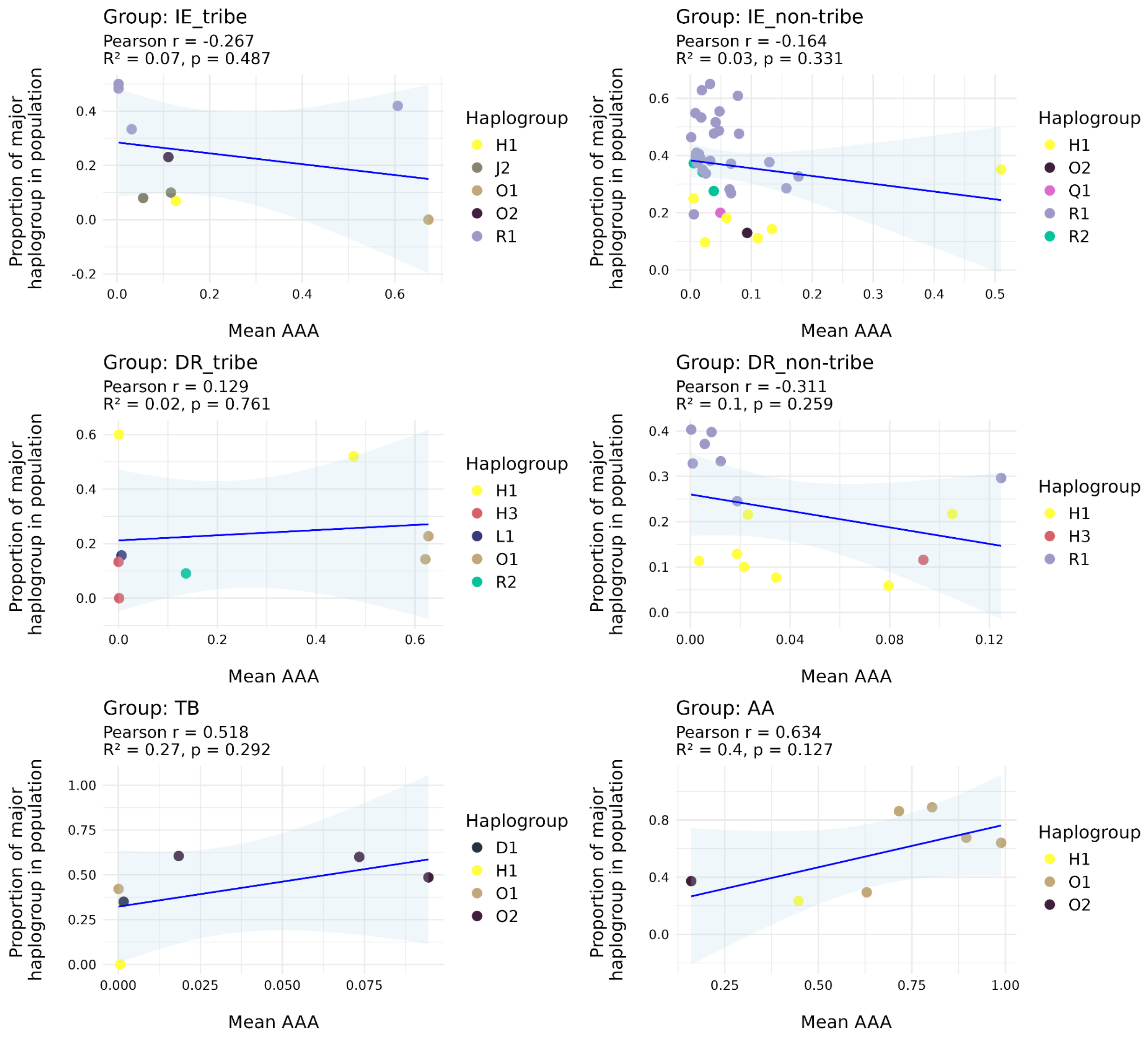

**Figure S5.3.11.** Relationship between autosomal AAA (Ancestral Austro-Asiatic) ancestry and dominant Y-chromosomal haplogroups across the GI populations. Across Indo-European and Dravidian groups, correlations between AAA ancestry and dominant Y-haplogroup frequencies are weak and non-significant, indicating limited coupling between this autosomal component and specific paternal lineages. Tibeto-Burman and Austroasiatic groups show positive trends, largely reflecting high frequencies of O-lineages, suggesting that AAA ancestry covaries modestly with East and Southeast Asian–associated paternal ancestry in these groups.

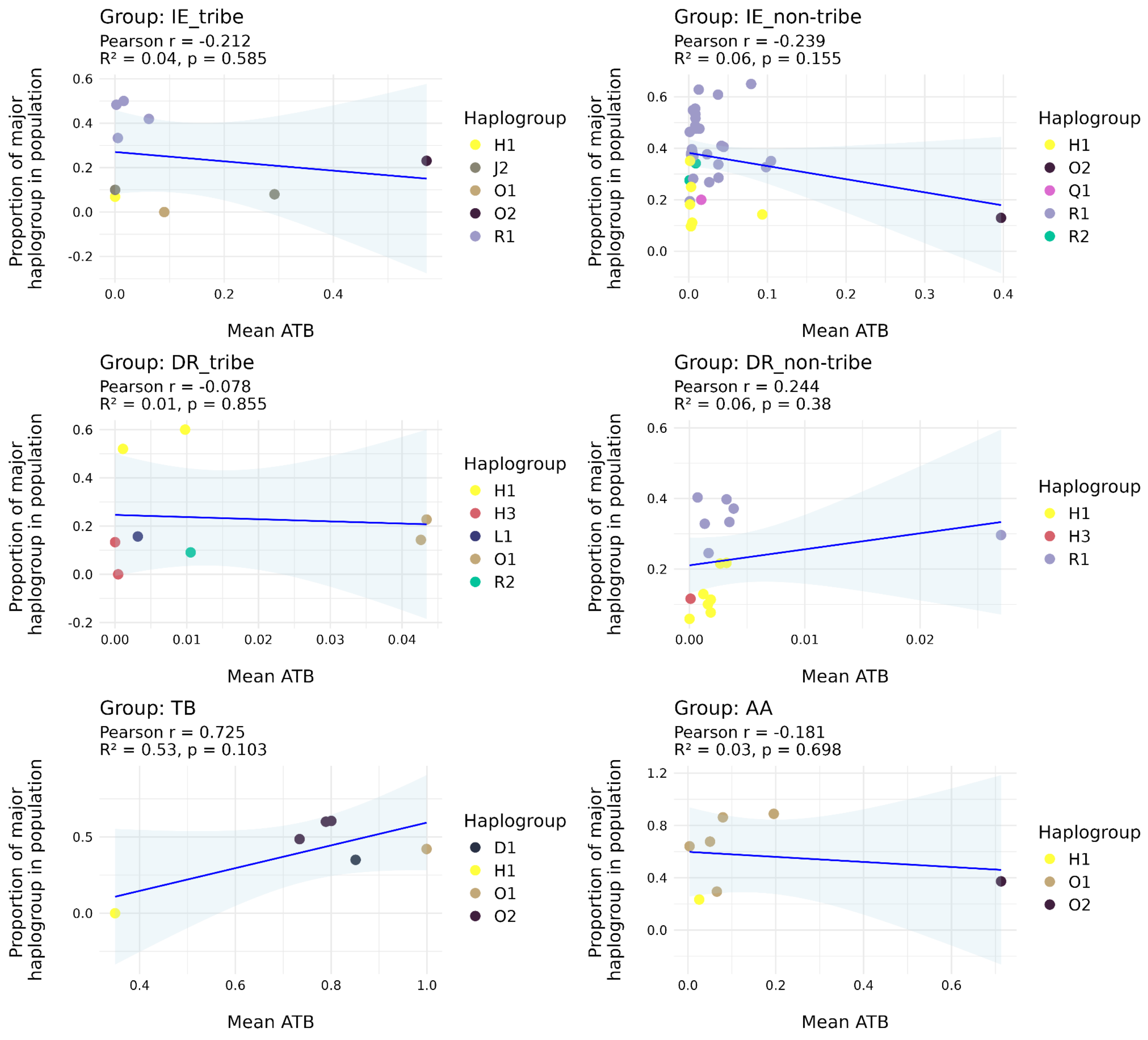

**Figure S5.3.12.** Relationship between autosomal ATB (Ancestral Tibeto-Burman) ancestry and dominant Y-chromosomal haplogroups across the GI populations.

Indo-European and Dravidian groups show weak or absent correlations between ATB ancestry and dominant Y-haplogroup frequencies, indicating minimal association between this autosomal component and paternal lineages in these populations. In contrast, Tibeto-Burman populations show a positive trend, consistent with increasing frequencies of O-lineages (particularly O2 haplogroup) with higher ATB ancestry. Austroasiatic groups display no consistent association. These results suggest that ATB ancestry is primarily informative for Tibeto-Burman demographic history and shows limited correspondence with Y-chromosomal variation outside this linguistic group.

#### Section S5.4 - Uniparental diversity analysis and evidence for sex-biased Migrations

Similar to our analysis of alpha diversity using ADMIXTURE proportions, we performed alpha diversity analysis on uniparental chromosomes (chrM - maternal, chrY - paternal). For this, we calculated population-wise alpha-diversity values of both mitochondrial and Y-chromosome haplogroups and converted them into z-scores. The relationship between z-scores of mitochondrial and y-chromosome diversities is shown in Figure S5.4.1.

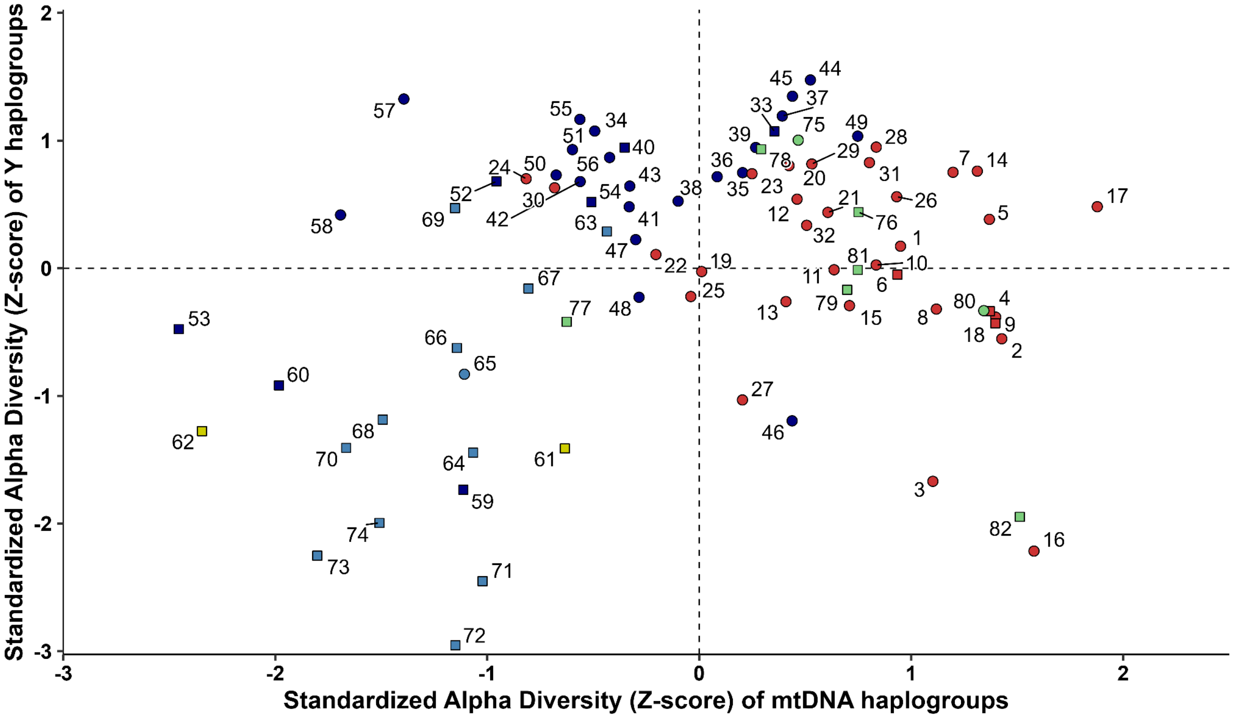

**Figure S5.4.1.** Population-wise Alpha-diversity analysis of mitochondrial (x-axis) and Y-chromosome (y-axis) haplogroups. Markers are colored by the ADMIXTURE cluster. Frequency of mtDNA haplogroups, Y haplogroups in each population and calculated diversity values with Z-scores are given in Supplementary Table S5.4.1.

The genomic architecture of South Asian populations reflects a long history of migration, social stratification, and sex-biased admixture. Consistent with asymmetric gene flow, analysis of Y-chromosomal haplogroup distributions across linguistic and social groups reveals a strong positive association between ANI ancestry and the prevalence of dominant paternal lineages in both Indo-European and Dravidian populations. In IE populations the frequent Y-haplogroups are R1 and R2, whereas in the DR populations, haplogroups H1 and H3 are the most frequent. The earliest documented samples of R1a were found in hunter-gatherers from Russia, with its subsequent expansion across Europe linked to Bronze Age steppe migrations (Haak et al., 2015), coinciding with the dispersal of Indo-European languages. The divergence of haplogroup R1 into R1a and R1b is estimated at approximately 25,000 years before present, with subsequent diversification of R1a likely occurring in the Middle East, near present-day Iran (Underhill et al., 2015).

In contrast, H1 haplogroup is often categorised as an indigenous Indian lineage (Cordaux et al., 2004; Sahoo et al., 2006; Sengupta et al., 2006). It likely originated with the initial modern human settlement of the subcontinent, constituted a major paternal component of the Indus Valley Civilization, and persists as the predominant indigenous paternal signature in South India, albeit with reduced frequency in upper-caste groups due to later migrations (Narasimhan et al., 2019). However, we observe a significant inverse relationship between ASI ancestry and the frequency of these major Y-chromosome haplogroups in non-tribal populations. ASI ancestry explains approximately 63% of the variance in the Dravidian non-tribal group ($r = -0.796$, $p = 0.000384$) and exhibits a similarly negative correlation in Indo-European non-tribal populations ($r = -0.493$, $p = 0.00195$). This pattern suggests that the expansion of haplogroups such as R1a, R2, and H1 within these populations was driven by disproportionate paternal gene flow from ANI-related sources, resulting in a partial replacement of indigenous ASI paternal lineages while retaining substantial autosomal admixture. These dynamics align with the timing and structure of South Asian admixture modeled by Moorjani et al., (2013), and suggest that ANI-related gene flow was frequently mediated by male-biased demographic processes rather than symmetric population mixing. The observed excess of Y-chromosomal haplogroup R frequencies relative to autosomal ANI ancestry proportions, yet relatively rare European-associated mtDNA lineages, is indicative of strongly male-biased gene flow, consistent with models of paternal lineage amplification and limited female migration from ANI-source (Basu et al., 2016).

We further observe a distinction in the prevalence of O haplogroups (O1 and O2), which serve as the dominant paternal lineages for eastern groups of TB and AA populations. In TB populations, the O2 haplogroup correlates positively with the ATB genetic component, while in AA groups, both O1 and O2 show a moderate positive correlation with the Ancestral AAA component. O is the major haplogroup in East Asia (Karafet et al., 2008). In our data, TB and AA groups show significant negative correlations with ANI ancestry ($r = -0.889$, $p = 0.0179$ and $r = -0.775$, $p = 0.0405$, respectively), revealing that their paternal ancestries were distinct from those in IE and DR groups. This suggests a paternal genetic structure that remained comparatively resilient to ANI-driven lineage expansion. Collectively, these results support a model wherein ANI-associated genomic input into TB and AA populations occurred primarily via autosomal admixture, while their dominant paternal lineages maintained greater phylogenetic continuity. This is consistent with previously proposed scenarios of asymmetric admixture (Basu et al., 2016).

#### Supplementary Section S6: Demography and Effective Population Size (Ne)

Devashish Tripathi[1,2], Vinay More[1,2], Analabha Basu[1,2]

[1]BRIC - National Institute of Biomedical Genomics (BRIC-NIBMG), Kolkata, India. [2]Regional Centre for Biotechnology (RCB), Faridabad, India.

#### Summary

The global human population has seen a rapid growth over the last 10000 years, having an estimated present-day size of ~8 billion. Mainland India has an estimated population size of ~1.4 billion, which contributes ~18% to the global population. The whole genome sequence datasets from the present-day populations in mainland India provide insights into the demographic history of the populations and the relationship among them. We observe most Indian populations to show a steady increase post the Last Glacial Maxima (LGM), a phenomenon consistent with global population growth following LGM. However, this phenomenon is far less generalizable if we delve into the details. Applying a clustering algorithm on the 'longitudinal' dataset (here the population size change trajectory acts as the longitudinal data), identifies three distinct groups of population. Two of the three groups show an overall increase in population size in recent times, whereas the third group, consisting mostly of AA and DR_T populations, shows negligible population growth and as well as population decline.

The pattern of effective population size (Ne) changes over time can be linked to the patterns of genetic variants and the disease susceptibility of the populations. Theoretical population genetics predicts that the population explosion post-bottleneck would result in an excess of novel rare variants in the populations, whereas populations that have experienced a decline or only a small increase in effective population size (Ne) tend to lose rare variants through genetic drift, leading to a shift toward intermediate-frequency alleles. However, some deleterious variants can drift to higher frequencies, increasing the burden of certain rare diseases (Keinan & Clark, 2012).

We used SMC++ (v1.15.4) (Terhorst et al., 2017) with default parameters and estimated the Ne for each population. The SMC++ first converts the population VCF to the smc format; here, we mask (--mask) the GRch38 Gap tracks downloaded from the UCSC browser. We then run the estimate command to have the fitted model for each population, giving the Ne estimates over time. We used the mutation rate of 1.25e-8 per base per generation, specified the starting and ending timepoints (in generations) 100, 100000 of the model (--timepoints), and used a piecewise spline (--spline) with 12 knots (--knots), which controls the functional form used to fit the model. Using the fitted Ne trajectories obtained for each population, we generated the plots using the ggplot2 package and performed clustering using the latrend package (4).

We color the estimated population trajectories (Ne) with respect to their major ancestral component (as inferred by ADMIXTURE), and we observed that all the populations (except CAO) have gone through a bottleneck ~2700-3700 generations before present, coinciding with the Out of Africa (OoA) human migration event. Assuming a generation time of 26.9 years (Wang et al., 2023), the time of the bottleneck translates to ~72,630-99,530 years before present (YBP), which aligns with the OoA human migration event. We start to see an increase in the Ne sizes post 2000 generations before present, which translates to 53,800 YBP (Terhorst et al., 2017). In the last ~1000 generations, we observe that the populations in mainland India have differential patterns. ASI-D consists of isolated populations with the smallest Ne in recent times, and a subset of ASI populations have moderate Ne in recent times. Predominantly, ANI populations consistently had higher estimated Ne in recent times compared to other populations.

The CAO population, which is recently admixed between two continental ancestral populations, has an inflated Ne estimate as parts of the genome come from the Africans that have diverged from Indian populations during the OoA event. The genomic regions coming from Africans resulted in a lower probability of coalescence in recent times, which indirectly translated into a higher effective population size in recent times.

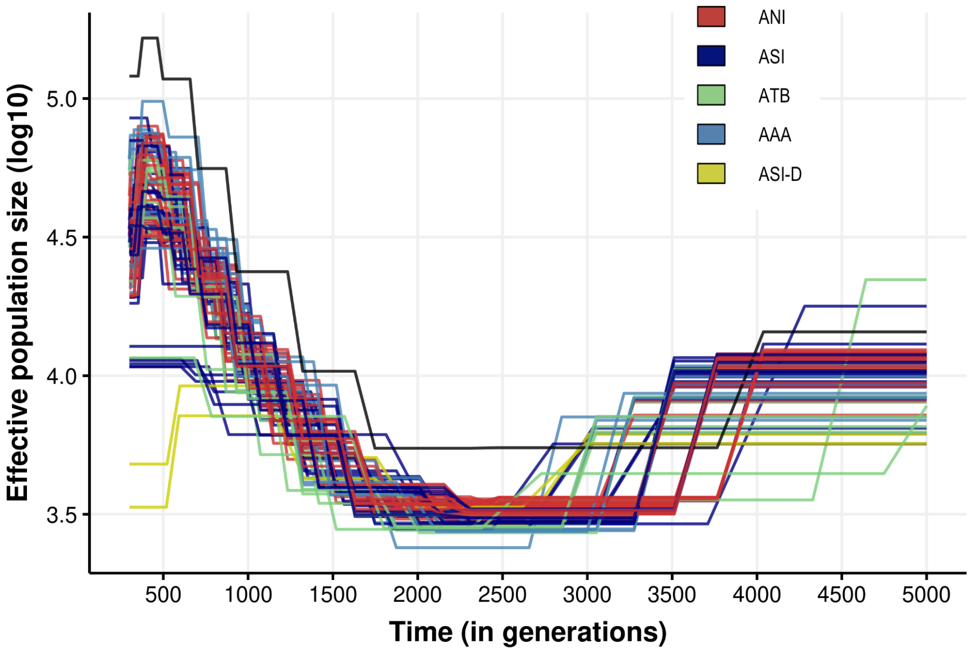

**Figure S6.1.** Plot consists of 83 populations' estimated Ne trajectories. Populations are colored based on the assigned admixture cluster. The X-axis represents the Time (in generations), ranging from 300 to 5000 generations, and the Y-axis represents Ne. **"Black line"** represents the continentally admixed population between the African and Indian populations.

We colored the populations based on 7 ethno-linguistic groups and also plotted the median Ne estimates for each group. We observed that the DR_T group has the lowest Ne expansion compared to other ethnolinguistic groups.

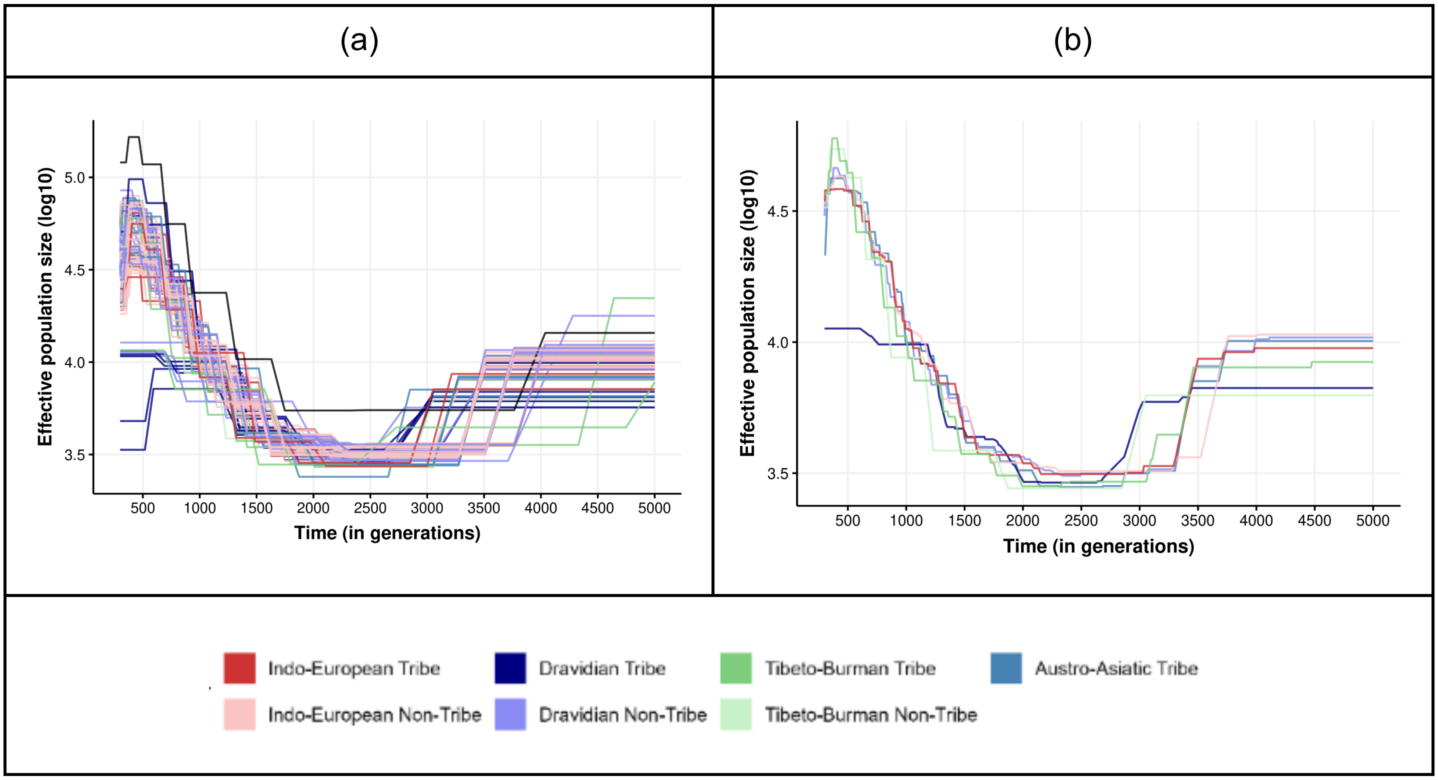

**Figure S6.2.** A) Plot consists of 83 populations' estimated Ne trajectories. Populations are colored based on the assigned Linguistic-tribe cluster. The X-axis represents the Time (in generations), ranging from 300 to 5000 generations, and the Y-axis represents Ne. "Black line" represents the continentally admixed population between the African and Indian populations. B) Plot contains the 7 linguistic-tribes groups' median estimated Ne trajectories. Populations are colored based on the assigned Linguistic-tribe cluster. The X-axis represents the Time (in generations), ranging from 300 to 5000 generations, and the Y-axis represents Ne. From the above figure, we can see that the Dravidian tribal populations have a lower Ne compared to other population groups.

From the above figures S6.1 and S6.2, we can see that the Dravidian tribal and a subset of Dravidian non-tribal populations have lower to moderate Ne compared to other population groups.

##### Cluster analysis on the estimated Ne trajectories

We then performed the clustering on the longitudinal data to cluster the populations with similar Ne trajectories to investigate and identify the populations with differential patterns of Ne trajectories. We performed the cluster analysis using two-step clustering through latent growth curve modeling and k-means (Teuling et al., 2024). Cluster analysis has been performed in two scenarios, one using the Ne estimates up to 10000 generations before present, and also up to 2000 generations before present. The idea is to look into the clusters when populations Ne have differential patterns in recent times with more resolution, and also look into the Ne trajectories, incorporating older population history to understand how population history changed in the past.

**Within-cluster mean absolute error (WMAE) distribution**

**Ne estimate up to 2000 generations**

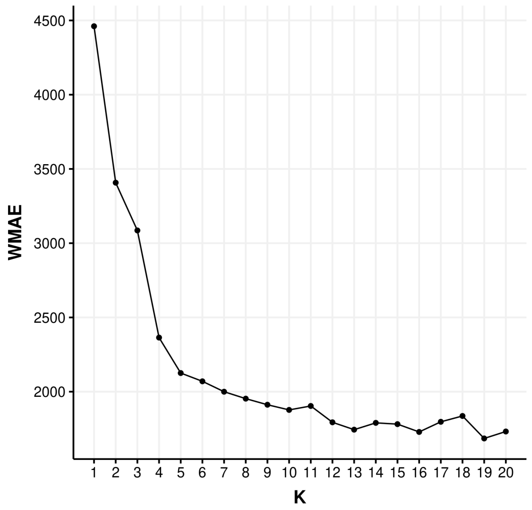

**Ne estimate up to 10000 generations**

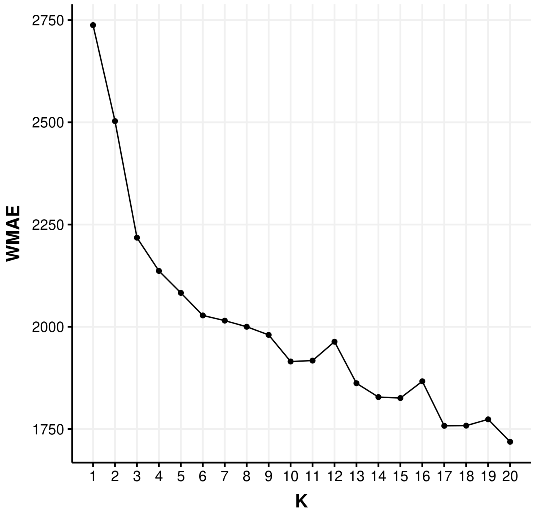

**Figure S6.3.** Within-cluster mean absolute error (WMAE) distribution

Based on the WMAE metric distribution, we chose the number of clusters (K) to be 4 to cluster the Ne trajectories in both scenarios.

In Figure S6.4, **Cluster A** contains the CAO population, which is recently admixed between two continental ancestral populations, and has an inflated Ne estimate as the ancestors are coming from populations that have diverged long ago, i.e, during the OoA event.

**Cluster B** consists of 9 non-IE tribal populations that have consistently lower Ne compared to other groups. In clusters C and D, populations have expanded, and the clusters are dominated by non-tribal IE populations.

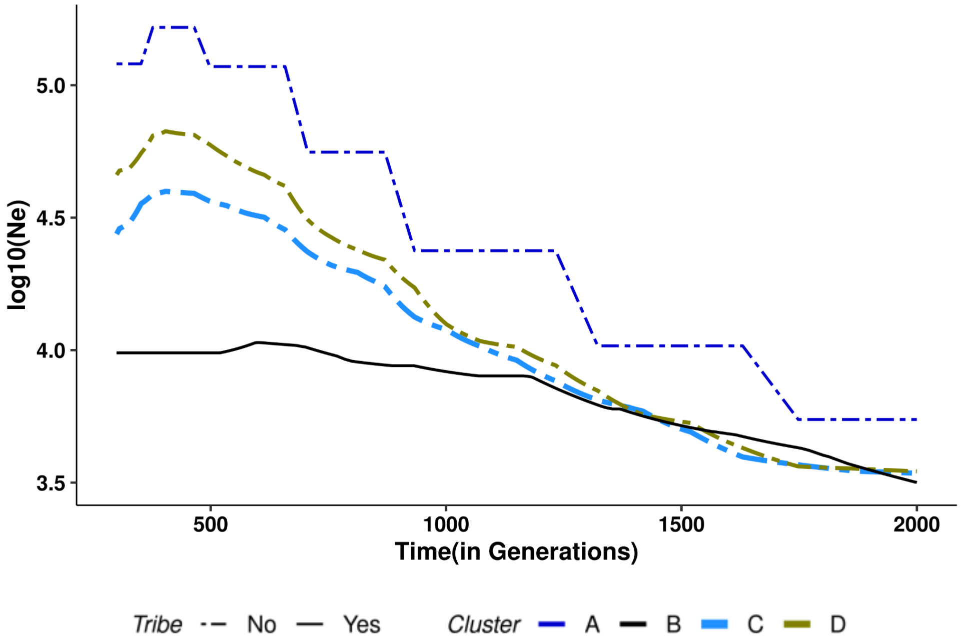

**Figure S6.4.** This consists of K = 4 fitted trajectories. The X-axis represents the Time (in generations), ranging from 300 to 2000 generations, and the Y-axis represents Ne. Each fitted trajectory is classified into tribe/non-tribe.

In figure **S6.5**, In all non-African populations, we observe a bottleneck post-out-of-Africa migration event ~2700-3700 generations ago. **Cluster A** contains the CAO population, which is admixed between African and Indian populations. **Cluster B** consists of 8 non-IE tribal populations that have consistently lower Ne compared to other groups. In **clusters C and D,** populations have expanded, and the clusters are dominated by non-tribal IE populations.

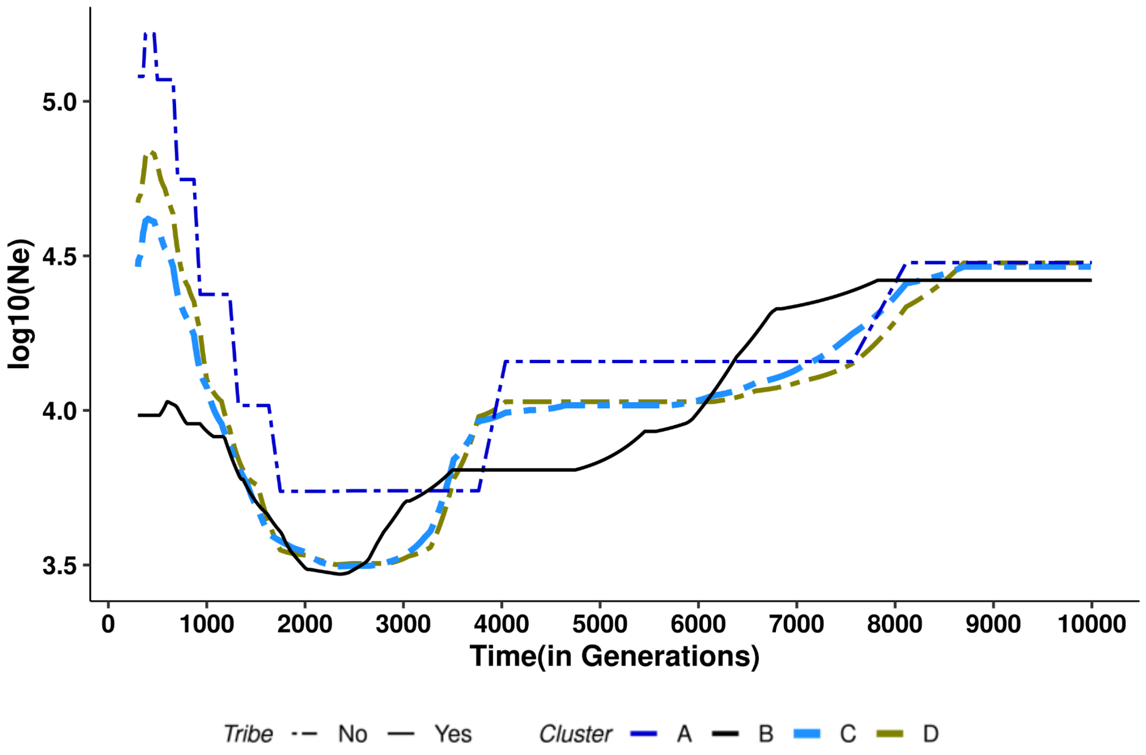

**Figure S6.5.** consists of K = 4 fitted trajectories. The X-axis represents the Time (in generations), ranging from 300 to 10000 generations, and the Y-axis represents Ne.

From **figures S6.4 and S6.5**, it is evident that there exist at least two broad population histories within mainland India. A group that contains only tribal populations from non-IE populations and other groups that are dominated by non-tribal populations. Non-tribal populations have been primarily from the agriculturist societies, whereas other tribal groups are isolated and historically practice mixed subsistence: from hunting-gathering, fishing, pastoralism, and occasional farming, a transition in cultural history might have shaped their demographic history. This altered demographic history would have a profound impact on their disease burden and is discussed in detail in the next section.

##### Divergence time estimation between NGH (ASI-D) and nonNGH populations in Cluster D

We estimated the divergence time between NGH populations (DR_NGH_1_01, DR_NGH_1_02) and the non-NGH population (DR_EGH_1_01) within Cluster D using the SMC++ split command. First, we inferred the marginal demographic histories for the NGH and non-NGH groups independently. Second, using these marginal histories alongside the joint site frequency spectrum (SFS), we modeled the joint demographic history to estimate the time of population split. Our results indicate a divergence time of approximately 600 generations.

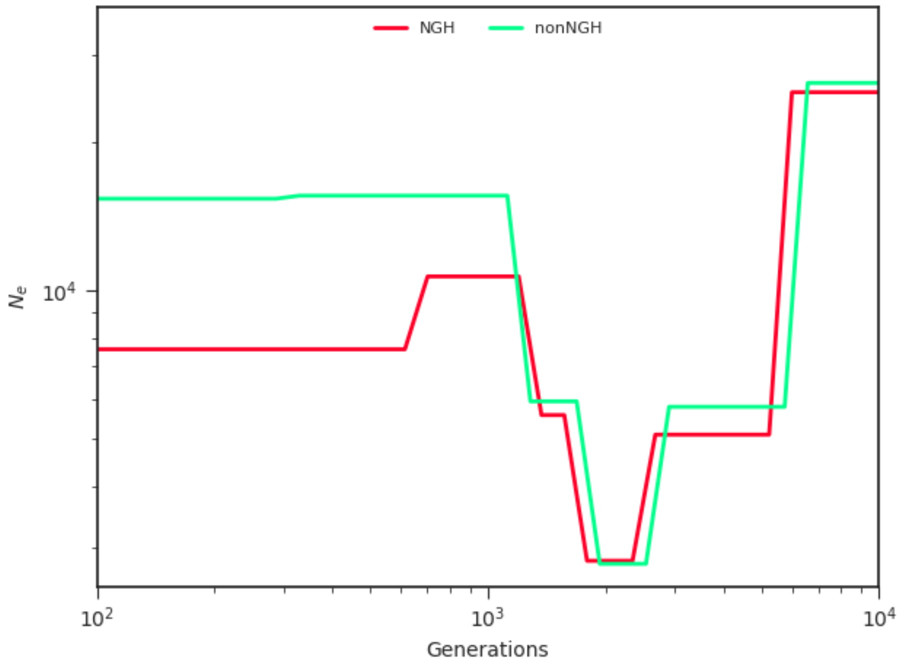

**Figure S6.6.** Joint demographic history of the NGH and nonNGH population. The X-axis represents the Time (in generations), ranging from 100 to 10000 generations, and the Y-axis represents Ne.

**Conclusion**

In summary, our analysis provides one of the first comprehensive and fine-scale delineations of demographic history across such a large number of mainland Indian populations. By leveraging whole-genome sequence data from 83 groups and modeling their effective population size (Ne) trajectories using SMC++, we capture major shared demographic events as well as striking population-specific deviations that illuminate the complexity of India's demographic landscape.

Across virtually all populations, we observe a pronounced bottleneck around ~2700–3700 generations before present, aligning well with the timing of the Out-of-Africa migration when translated using established estimates of generation time. Beyond this shared ancient history, however, the trajectories diverge substantially in more recent periods. Our clustering results demonstrate that non-agriculturist tribal populations—particularly those belonging to non-IE linguistic groups—consistently exhibit much smaller Ne values in the recent past compared to agriculturist, predominantly IE non-tribal populations. This reduction in Ne among isolated groups, many of whom historically practiced mixed subsistence strategies rather than agriculture, suggests a fundamentally different demographic trajectory following the Neolithic demographic transition (NDT). In contrast, IE non-tribal populations exhibit sustained and pronounced expansions in Ne, consistent with demographic processes associated with agricultural societies.

These findings are consistent with the patterns observed in the population structure analysis (S4). In the PCA, tribal populations form isolated, well-separated clusters, whereas non-tribal IE and DR populations fall along a cline. This strong structuring in PCA space matches the demographic signals recovered from the Ne trajectories: groups that cluster separately in PCA—most notably the isolated tribal populations—also show reduced recent Ne and distinct demographic histories. Thus, two independent analyses converge on the same conclusion: tribal populations in mainland India have experienced long-term isolation and markedly different population size histories compared to non-tribal agriculturist groups.

Finally, the contrasting demographic histories we observe—particularly the reduced recent Ne and long-term isolation among many tribal groups—have direct consequences for disease burden. Such demographic conditions are known to increase homozygosity, elevate the frequency of deleterious variants, and reshape the landscape of rare disease risk. These effects, and their manifestation in Indian populations, are examined in detail in the following sections through ROH-based analyses and related approaches.

**Table S6.1. The table describing the cluster labels of different populations**

| Code | Population_Order | cluster_2K | cluster_10K |
|---|---|---|---|
| CAO | 83 | A | A |
| DR_EGH_1_01 | 53 | B | B |
| DR_WGH_1_01 | 59 | B | B |
| DR_WGH_1_02 | 60 | B | B |
| DR_NGH_1_01 | 62 | B | B |
| DR_ECP_2_05 | 50 | B | B |
| DR_ECP_2_06 | 49 | B | B |
| DR_NGH_1_02 | 61 | B | B |
| TB_WHR_1_02 | 81 | B | B |
| IE_NRP_2_01 | 46 | C | C |
| IE_NRP_2_03 | 35 | D | C |
| IE_WCP_2_01 | 10 | C | C |
| IE_NRP_2_04 | 42 | C | C |
| IE_SCH_1_01 | 40 | C | C |
| IE_NCH_1_01 | 52 | C | C |
| IE_WHR_1_01 | 33 | D | C |

| | | | |
|---|---|---|---|
| DR_EPL_1_01 | 68 | D | C |
| IE_NCH_2_01 | 24 | C | C |
| IE_NDN_2_01 | 23 | C | C |
| DR_ECP_2_01 | 43 | B | C |
| IE_NCH_1_02 | 54 | C | C |
| IE_WPL_1_01 | 6 | C | C |
| TB_BPV_1_01 | 79 | D | C |
| DR_ECP_2_02 | 26 | C | C |
| IE_NRP_2_05 | 1 | C | C |
| AA_EPL_1_02 | 72 | C | C |
| DR_ECP_2_03 | 55 | C | C |
| IE_WHR_2_03 | 16 | C | C |
| IE_NRP_2_06 | 13 | C | C |
| IE_ECP_2_01 | 34 | C | C |
| IE_NRP_2_07 | 32 | C | C |
| AA_NER_1_01 | 77 | C | C |
| IE_NRP_2_08 | 2 | C | C |
| AA_SCH_1_01 | 69 | C | C |
| IE_WCP_2_02 | 47 | C | C |
| IE_ERP_2_01 | 36 | C | C |
| IE_ERP_2_02 | 15 | C | C |
| DR_ECP_2_04 | 56 | C | C |
| IE_NDN_2_02 | 39 | C | C |
| TB_NER_2_01 | 80 | C | C |
| TB_NER_1_01 | 82 | C | C |
| DR_WCP_2_01 | 30 | C | C |
| DR_WCP_2_02 | 21 | D | C |
| IE_WPL_2_02 | 3 | C | C |
| DR_EGH_2_01 | 57 | C | C |
| IE_WCP_2_04 | 31 | D | C |
| TB_BPV_1_02 | 78 | D | C |

| | | | |
|---|---|---|---|
| IE_ERP_2_04 | 27 | C | C |
| IE_BPV_2_01 | 75 | C | C |
| IE_WPL_2_03 | 12 | C | C |
| IE_NRP_2_09 | 48 | C | C |
| IE_NCH_1_03 | 66 | C | C |
| AA_EPL_1_04 | 74 | C | C |
| IE_NRP_2_10 | 11 | C | C |
| IE_NRP_2_11 | 9 | C | C |
| IE_NRP_2_12 | 19 | C | C |
| IE_CHR_1_01 | 76 | C | C |
| DR_WCP_2_03 | 41 | C | C |
| DR_SDN_2_03 | 45 | C | C |
| IE_WCP_2_05 | 38 | C | C |
| DR_SDN_2_01 | 58 | D | D |
| IE_NRP_2_02 | 8 | D | D |
| IE_EPL_1_01 | 70 | D | D |
| TB_WHR_1_01 | 18 | D | D |
| IE_WPL_2_01 | 28 | D | D |
| AA_EPL_1_01 | 63 | D | D |
| IE_WHR_2_01 | 14 | D | D |
| IE_WHR_2_02 | 17 | D | D |
| DR_EPL_1_02 | 67 | D | D |
| IE_WHR_1_02 | 4 | D | D |
| IE_WHR_2_04 | 5 | D | D |
| IE_WHR_2_05 | 7 | D | D |
| IE_WCP_2_03 | 25 | D | D |
| AA_NDN_1_01 | 71 | D | D |
| IE_EPL_2_01 | 65 | D | D |
| DR_SDN_2_02 | 44 | D | D |
| DR_EPL_1_03 | 64 | D | D |
| AA_EPL_1_03 | 73 | D | D |

| | | | |
|---|---|---|---|
| IE_ERP_2_03 | 37 | D | D |
| IE_ECP_2_02 | 22 | D | D |
| DR_ECP_2_07 | 51 | D | D |
| DR_ECP_2_08 | 29 | D | D |
| IE_NRP_2_13 | 20 | D | D |

#### Supplementary Section S7: Runs of Homozygosity (ROH) analysis

Chandrika Bhattacharyya[1], Vinay More[1,2], Pratheusa Machha[3], Divya Tej Sowpati[3,4], Analabha Basu[1,2]

[1]BRIC - National Institute of Biomedical Genomics (BRIC-NIBMG), Kolkata, India. [2]Regional Centre for Biotechnology (RCB), Faridabad, India. [3]CSIR - Centre for Cellular and Molecular Biology (CSIR-CCMB), Hyderabad, India. [4]Academy of Scientific and Innovative Research, Ghaziabad, India.

#### Summary

Runs of Homozygosity (ROH) occur when identical haplotypes are inherited from each parent, resulting in extended regions of homozygous genotypes. Analyzing the number and size of these ROH provides valuable insights into a population's demographic history and cultural practices. Population bottleneck or founder effect increases genome-wide homozygosity, resulting in shorter ROH lengths over successive generations due to recombination. In contrast, recent or enduring endogamy, a cultural practice of marrying within communities, generates an excess of longer ROH.

In the GI dataset, we noted that almost all populations have a burden of ROH to varying degrees. A combination of founder effect, endogamy, and isolation has given rise to unique patterns of ROH distribution among the populations of mainland India. In multiple cases, the burden of ROH exceeds what is observed in Finnish and Ashkenazi Jewish populations. These observations have important implications in understanding disease burden and the design of population-specific genetic screens in India.

**Methods:** We have removed 82 population misclassified individuals (as mentioned in S2) and 244 children from the trio sets. A total of 9,442 unrelated individuals were retained for downstream ROH analysis. To provide a broader context for our findings, we have included whole genome sequence data of individuals from two other populations, widely studied for their ROH burden, the Finnish (FIN) (Jakkula et al., 2008) and the Ashkenazi Jewish (AJ) (Kang et al., 2017).

ROHs were identified using PLINK v1.9, excluding singleton variants. We modified the following parameters: --homozyg --homozyg-kb 500 --homozyg-window-snp 100 --homozyg-window-het 2 --homozyg-window-missing 20; to ensure a minimum physical length of detected ROH as 500 Kb. To incorporate information on recombination, we converted the physical lengths of ROHs into genetic distances (in centimorgans, cM) using a genetic map from https://github.com/odelaneau/shapeit5/tree/main/resources/maps/b38, in PLINK. Only ROHs with a minimum genetic length of 1 cM were included in the final analysis.

**Results**

To quantify the burden of ROH, for each individual, we have estimated the following three measures: a) total number of ROH (NROH), b) average length of ROH (LROH), and c) genomic proportion in ROH ($F_{ROH}$). For $F_{ROH}$, we have estimated the proportion of the total physical length of ROH with respect to the total genome length (3.1 gigabases). Accordingly, for each population, we calculated the median values of NROH, LROH, and FROH, and compared these with the Ashkenazi Jewish (AJ) and Finnish (FIN) populations (Table S7.1). We compared the ROH measures for each population, separately with FIN and AJ, using one-sided two-sample Kolmogorov–Smirnov (KS) tests. A population was considered significantly different if the FDR-adjusted p-value < 0.05, and the median value is higher than the reference populations (Table S7.2).

**Table S7.2.** Number of populations showing significant difference in the distribution of ROH measures (one-sided Kolmogorov–Smirnov tests, FDR 5%), relative to Finns (FIN) and Ashkenazi Jews (AJ)

| Reference | LROH | NROH | FROH |
|---|---|---|---|
| FIN | 59 | 22 | 39 |
| AJ | 23 | 21 | 28 |

We observed that within the individuals from the majority of the populations (63 out of 83), the median length of ROH is more than 1.5 cM. ROH longer than 1.5 cM, is generally considered a threshold for identifying the influence of endogamy, because only a very small proportion of individuals from outbred populations harbour ROH of this length (McQuillan et al., 2008).

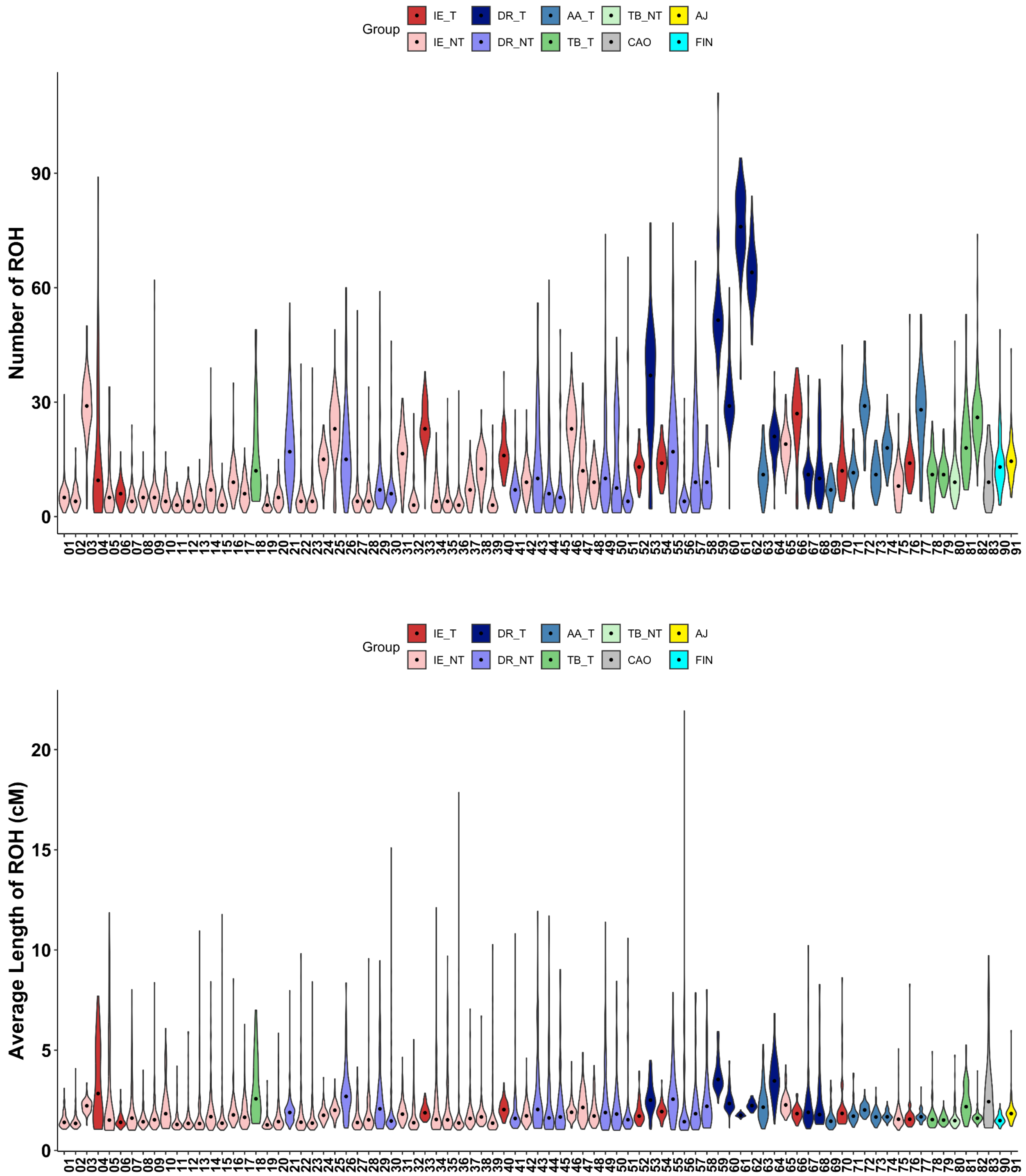

**Figure S7.1.** Distribution of the total number of ROH (NROH) (top panel) and average length of ROH (LROH) (bottom panel) across all 83 populations, along with FIN and AJ. The median is shown as a black circle.

We have also noted that these populations have considerably larger effective population sizes as estimated in recent times (Figure S7.2), highlighting that cultural endogamy is widespread among the populations of mainland India, which is getting reflected in the distribution of longer tracts of ROH even in large populations.

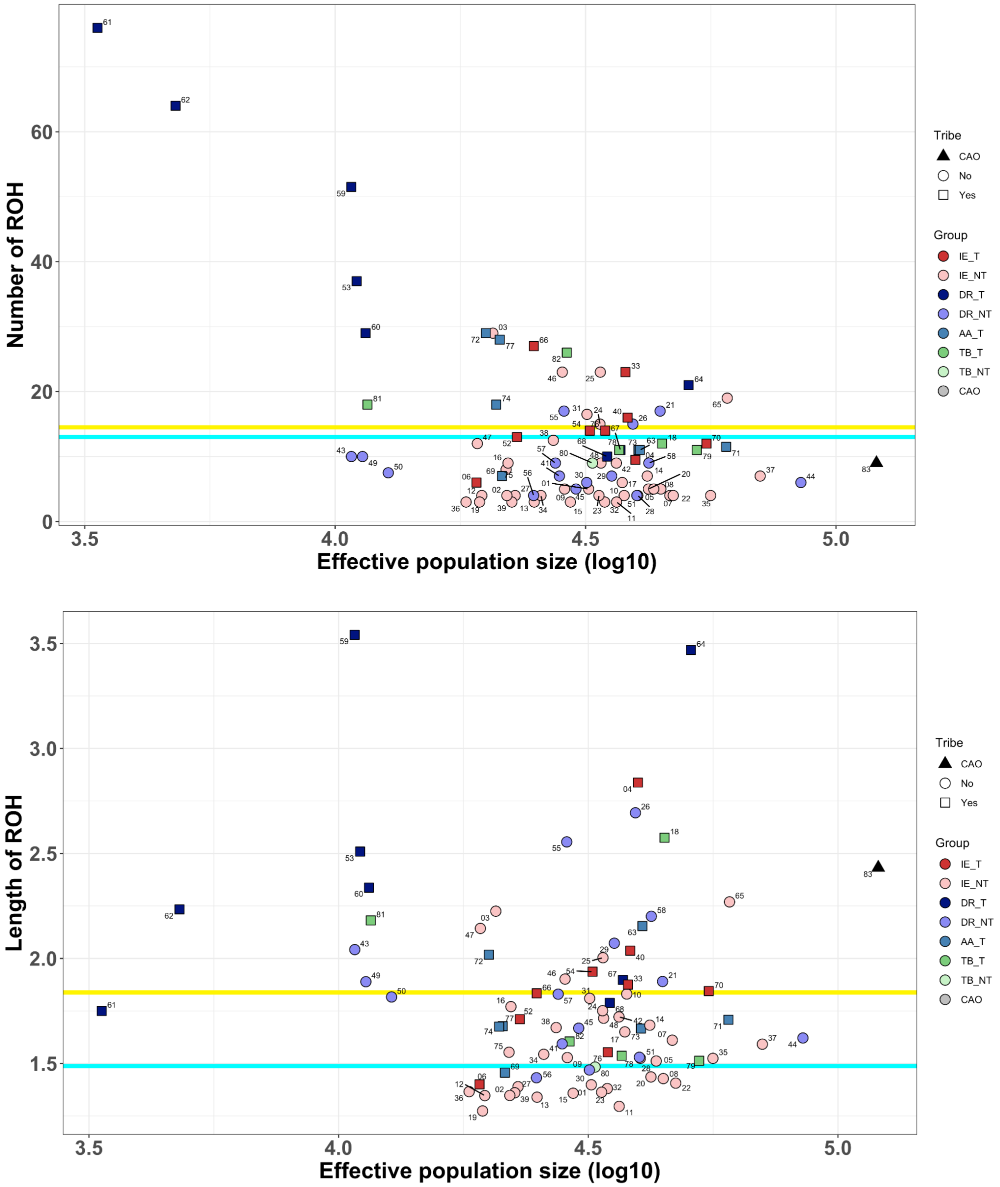

**Figure S7.2.** The median value of the total number of ROH (NROH) (top) and length of ROH (bottom) is plotted for each population along with their estimated effective population size, at 300 generations ago. Assuming a generation time of 26.9 years, the time reflects the post neolithic period. Populations with small population sizes harbour large numbers of short ROH. In contrast, several populations with relatively large population size, carrying ROH with median length > 1.5 cM, arise due to endogamy. Horizontal lines of cyan and yellow mark the median values of NROH and LROH observed in Finnish (FIN) and Ashkenazi Jewish (AJ) populations.

Generally, long ROH (>10 cM) indicate more recent parental relatedness due to endogamy (Sahoo et al., 2021), whereas the shorter ones (<10 cM) reflect a small effective population size. AJ individuals have a median of 1 long ROH segment and 14 short ROH segments, while FIN individuals have a median of 1 long ROH segment and 12 short ROH segments.

We have observed that DR_T populations harbor a large number of ROH, specifically short ROH (1-10 cM length) (Figure S7.3). Two DR_T populations from Nilgiri hills (61 and 62), who have the smallest Ne in recent times, have the largest burden of short ROH. Other tribal populations, such as AA_T and TB_T, also possess a higher number of short ROH than both Ashkenazi Jewish (AJ) and Finnish. In contrast, populations from DR_NT harbour a large number of long ROH (>10 cM), suggesting endogamy, a cultural practice persistent within the community.

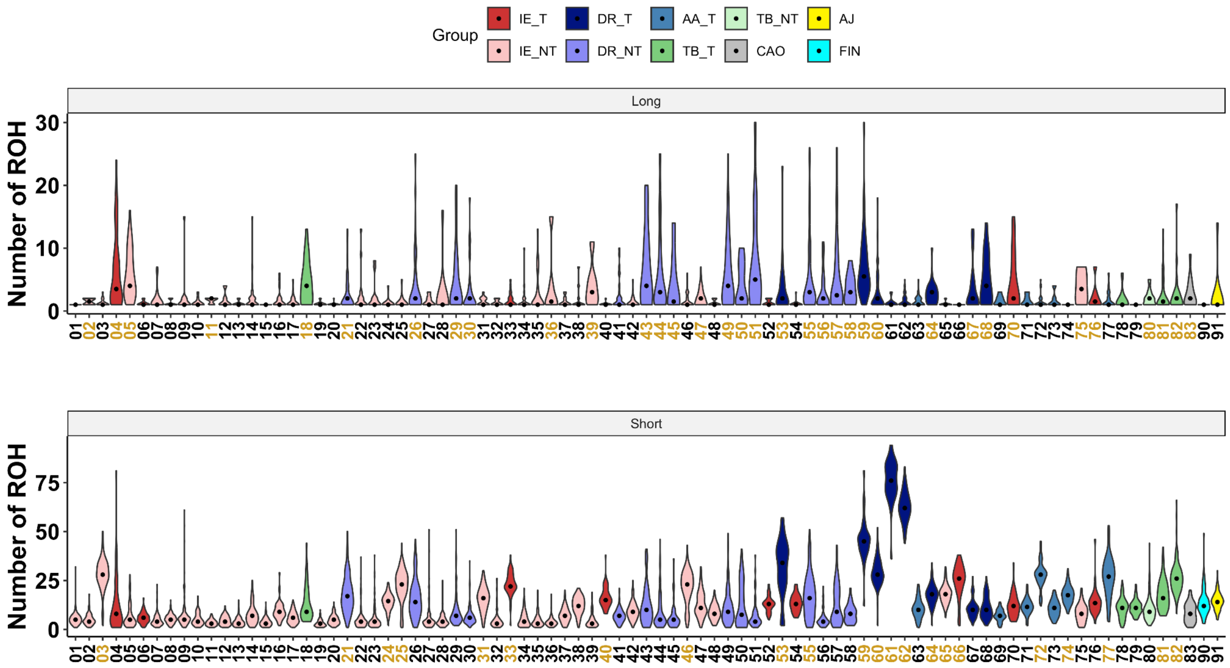

**Figure S7.3.** Distribution of the total number of ROH (NROH) across all 83 populations, along with FIN and AJ. The median is shown as a black circle. For each population, long ROH (>10 cM) are plotted above and shorter ROH (< 10 cM) are plotted below. Populations with higher median NROH than Ashkenazi Jewish (AJ) are highlighted in gold. AJ individuals have a median of 1 long ROH segment and 14 short ROH segments; while FIN individuals have a median of 1 long ROH segment and 12 short ROH segments.

Overall, we identified 35 populations with median number of long ROH greater than AJ, while 22 populations have short ROH more than AJ.

In total, we have identified 2727 individuals, across all the 83 populations, having more than 1% of their autosomal genome with ROH measuring 1.5 Mb and longer, here termed as $F_{ROH}$; while among the individuals from AJ, the median $F_{ROH}$ is 0.0075, and only 21% individuals harbour 1% ROH in their genome. For 6 out of 8 DR_T populations (Except for two DR_T populations from the Eastern Plateau), we observed that more than 80% of the individuals have $F_{ROH}$ more than 0.01.

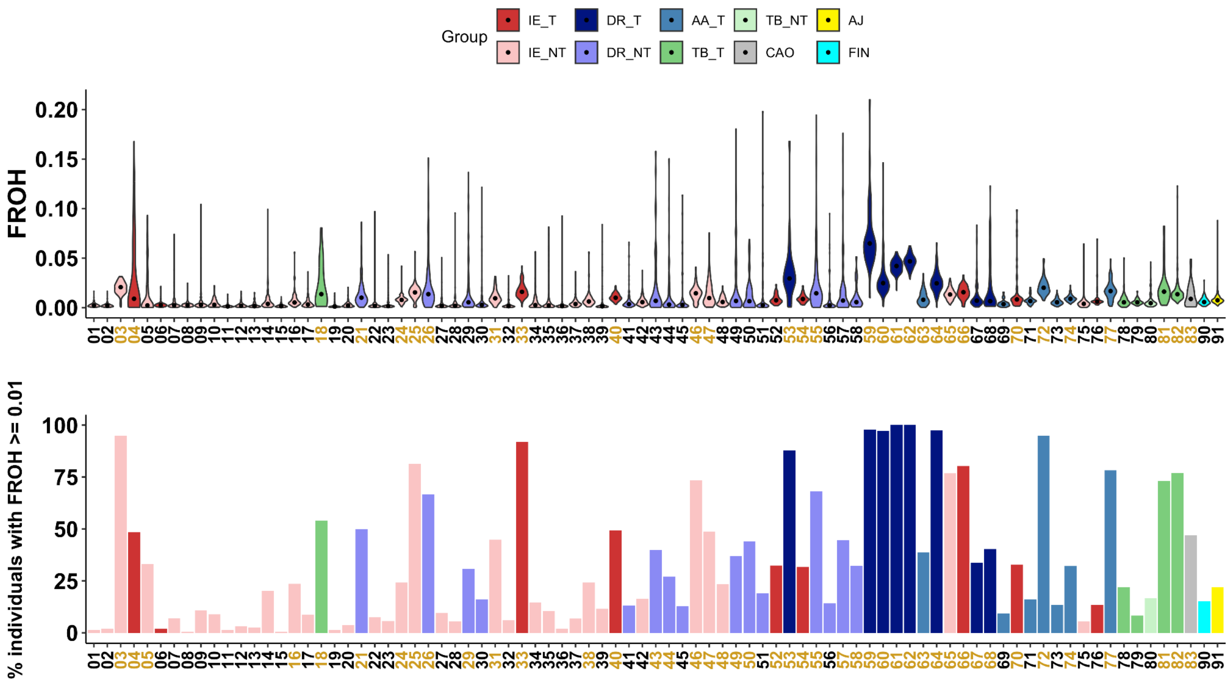

**Figure S7.4.** In the Top panel, the distribution of FROH is shown across all 83 populations, along with FIN and AJ. The median is shown as a black circle. Populations with higher median FROH than Ashkenazi Jewish (AJ) are highlighted in gold.
In the bottom panel, the percentage of individuals who have FROH ≥ 0.01 is plotted for each population. Here, populations that have more individuals having FROH ≥ 0.01 than Ashkenazi Jewish (AJ) are highlighted in gold.

Interestingly, although the IE_NT group did not show a notable ROH burden based on either number or length, some IE_NT populations (Population 3, 25, 46, 65) showed markedly high homozygosity, with more than 70% of individuals having FROH ≥ 0.01. This indicates that fine-scale sociocultural structure influencing parental relatedness may exist even within these large, non-tribal communities.

#### Supplementary section S8: Annotation of GI variants

Sauma Suvra Majumdar[1,2], Krithika Subramanian[1,3], Rupanwita Majumder[1], Priyanka Singh[1], Mohammed Faruq[4,5], Divya Tej Sowpati[5,6], Analabha Basu[7,8], Bratati Kahali[1]

[1]Centre for Brain Research (CBR), IISc Campus, Bengaluru, India. [2]Interdisciplinary Mathematical Sciences, Indian Institute of Science (IMI- IISc), Bengaluru, India. [3]Manipal Academy of Higher Education (MAHE), Karnataka, India. [4]CSIR - Institute of Genomics & Integrative Biology (CSIR-IGIB), New Delhi, India. [5]Academy of Scientific and Innovative Research, Ghaziabad, India. [6]CSIR - Centre for Cellular and Molecular Biology (CSIR-CCMB), Hyderabad, India. [7]BRIC - National Institute of Biomedical Genomics (BRIC-NIBMG), Kolkata, India. [8]Regional Centre for Biotechnology (RCB), Faridabad, India.

#### Summary

##### Overall workflow of sections S8 - S12:

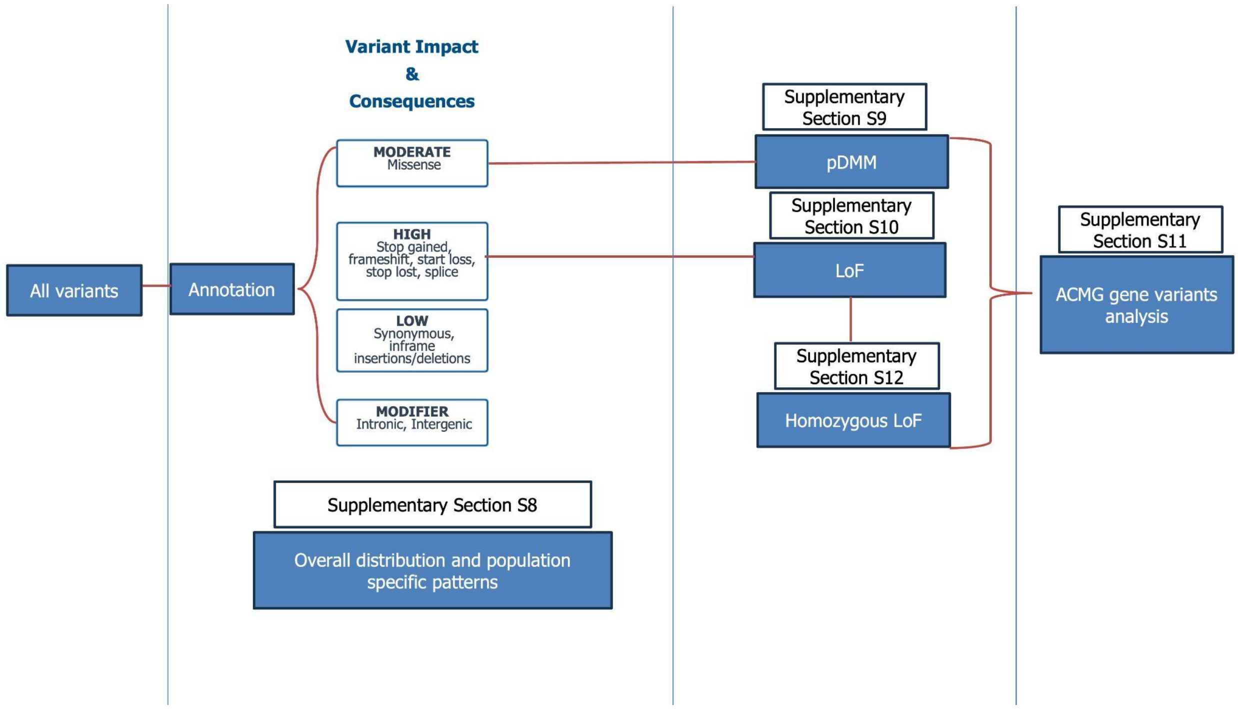

##### Introduction:

Sections S8-S12 presents analysis regarding comprehensive functional annotation and medical relevance of genetic variants identified in the GenomeIndia study.

We present the comprehensive functional annotation of all small variants in the GI dataset and their distribution in different populations. We prioritized these variants by utilizing the Variant Effect Predictor (VEP) and LOFTEE to define high-confidence loss-of-function (HC-LoF) and putative deleterious missense variants (pDMMs, based on REVEL and CADD). We integrated data from ClinVar and the GWAS Catalog to investigate pathogenicities and disease relationships.

#### Methods

##### Genomic Annotation

We annotated the small variants (SNPs and short InDels) using VEP v113 (McLaren, William et al. 2016). For each variant with multiple annotations, the annotation corresponding to the principal transcript was chosen using the "flag_pick" command. To prioritize canonical transcript effects, we filtered for variants with PICK = 1. We further filtered transcripts by choosing "protein_coding" in the BIOTYPE field column. The variants that were present on exonic regions of these protein-coding transcripts were designated as protein-coding variants.

We identified loss-of-function (LoF) by annotating using the Loss-Of-Function Transcript Effect Estimator (LOFTEE) plugin (Karczewski et al., 2020) within the Variant Effect Predictor (VEP) annotation framework. We retained only high-confidence (HC) LoF variants, defined as protein-truncating changes including frameshift, splice donor, splice acceptor, and stop-gained variants. Further, we evaluated the potential impact of missense variants; for that, we used two predictive scoring systems: REVEL (Rare Exome Variant Ensemble Learner) https://sites.google.com/site/revelgenomics/ and CADD (Combined Annotation Dependent Depletion) https://cadd.gs.washington.edu/ . REVEL integrates outputs from 13 individual tools (e.g., MutPred, FATHMM, PolyPhen-2, SIFT, PROVEAN, and others) to generate a combined pathogenicity score. Missense variants with REVEL scores ≥0.644 and CADD scores ≥20 were considered deleterious. We assessed the pathogenicity of the variants by cross-referencing with ClinVar and identified known pathogenic mutations in the whole GI dataset. ClinVar file was downloaded from the NCBI FTP repository https://ftp.ncbi.nlm.nih.gov/pub/clinvar/tab_delimited/ in February 2025. Variants annotated as pathogenic (P) or pathogenic/likely pathogenic (P/LP) (review status ≥2 stars) were classified as clinically relevant. Gene Ontology analysis was performed using PANTHER (Mi et al., 2013).

#### Results

##### Annotation of variants and their distribution across different populations reveal distinct patterns

Of the total 129.9 million variants we uncovered from 9768 individuals, 70.74 million (12.3 million novel) were non-singletons, and 59.2 million (32 million novel) variants were singletons in the dataset. Expectedly, the majority of the variants fall in the MODIFIER category (including intronic, intergenic, 3'/5'UTR variants, etc.), occurring in non-coding regions of the genome. In the non-singleton and singleton categories, 801,710 variants and 815,082 variants belonged to the coding regions of the genome, respectively. Of them, only 0.043% (24,190) of singleton SNVs, and 0.02% (15,813) of non-singleton SNVs are classified as HIGH IMPACT variants. HIGH-impact INDELs appear to be more frequent than HIGH-impact SNVs, particularly among singletons, where they account for 0.44% (14,320) of variants (Supplementary Table S8.1). Further granular distribution of variant consequences is provided in Supplementary Figure S8.1.

**Table S8.1.** Distribution of annotated small variants across different categories

| *VARIANT TYPE* | *CATEGORY* | *NOVELTY* | *MODIFIER* | *MODERATE* | *HIGH* | *LOW* | *TOTAL* |
|---|---|---|---|---|---|---|---|
| *SNVS* | **Non-singletons** | **Known** | 54,041,274 | 367,286 | 13,494 | 334,262 | 54,756,316 |
| | | **Novel** | 10,970,662 | 32,727 | 2,319 | 25,536 | 11,031,244 |
| | **Singletons** | **Known** | 25,385,831 | 325,863 | 15,292 | 238,778 | 25,965,764 |
| | | **Novel** | 29,778,722 | 118,842 | 8,898 | 79,583 | 29,986,045 |
| *INDELS* | **Non-singletons** | **Known** | 3,638,323 | 5,552 | 6,858 | 8,152 | 3,658,885 |
| | | **Novel** | 1,294,842 | 1,138 | 2,293 | 2,093 | 1,300,366 |
| | **Singletons** | **Known** | 1,119,388 | 3,678 | 5,880 | 4,416 | 1,133,362 |
| | | **Novel** | 2,093,155 | 2,164 | 8,440 | 3,248 | 2,107,007 |
| | | | | | | | **129,938,989** |

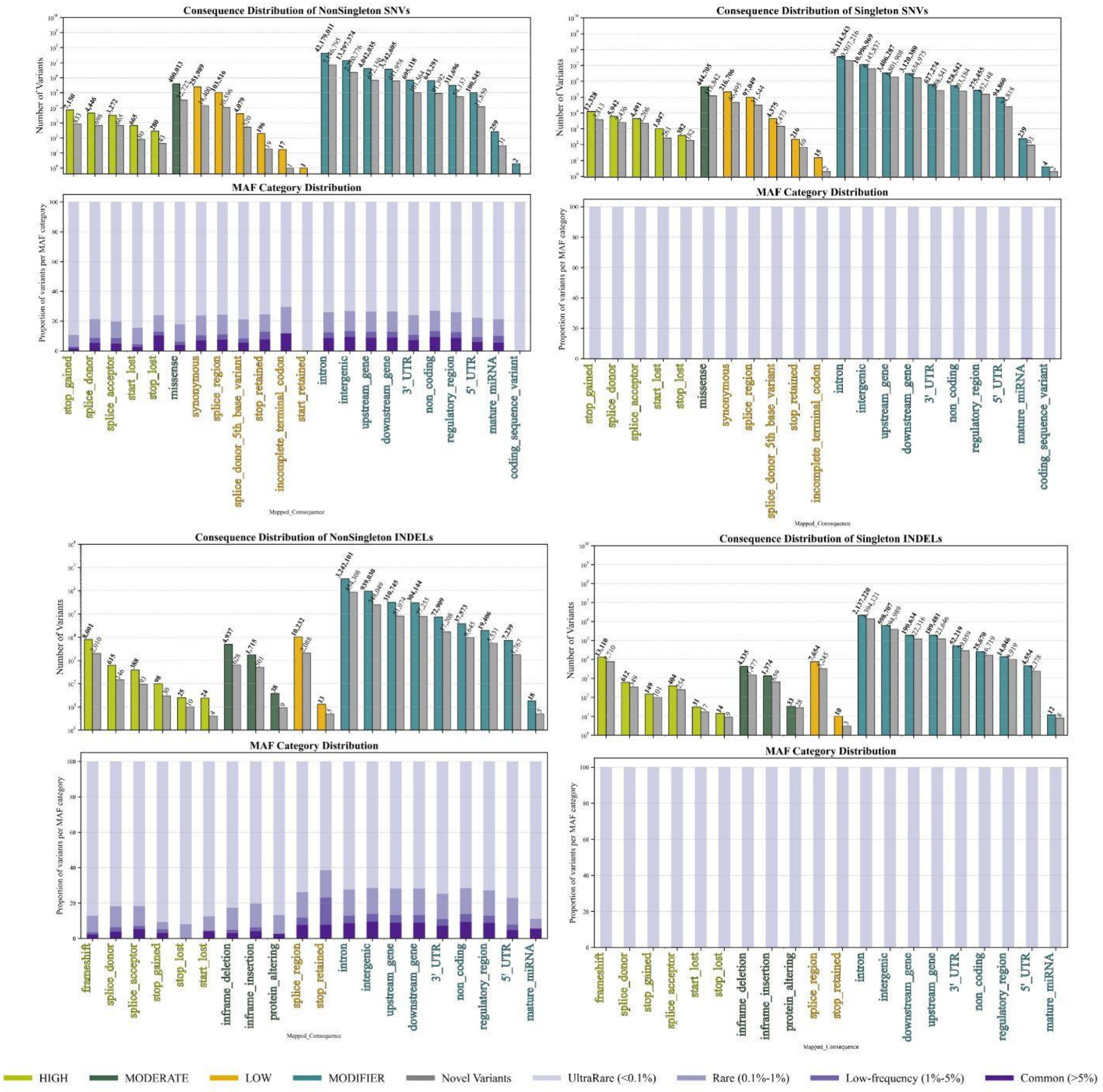

**Figure S8.1**. Distribution of functional consequences of non-singleton and singleton SNVs and InDels in the GenomeIndia dataset. The color of their impact highlights the individual consequences, and the novel variants in the same category are plotted as grouped grey bars. The proportion of the respective variants in each frequency category is depicted in the stacked bar below every consequence.

We identified a significant burden of potentially protein-disrupting variations, comprising 54,903 high-impact variants (18,143, ~33% of these being novel)—namely, frameshift, stop-gain, start-loss, stop-loss, and splice-site mutations. An individual carried, on average, a substantial load of these variants, including 83 frameshift indels, 54 stop-gain mutations, and 36 splice-site alterations, collectively affecting 8,537 genes across the dataset. Further heterogeneity was observed among different population groups (Supplementary Table S8.2).

**Table S8.2.** Average individual carrier rates of protein-coding variants in different populations

| | CAO | IE_NT | DR_NT | IE_T | DR_T | AA_T | TB_T | TB_NT |
|---|---|---|---|---|---|---|---|---|
| **Frameshift** | 91.16 | 81.206 | 82.435 | 82.669 | 85.166 | 81.426 | 78.009 | 76.83 |
| **Frameshift_novel** | 13.32 | 14.119 | 14.263 | 14.375 | 15.272 | 13.948 | 12.861 | 12.40 |
| **Missense** | 8305.34 | 7521.93 | 7506.59 | 7521.42 | 7464.07 | 7399.86 | 7175.21 | 7134.7 |
| **Missense_novel** | 6.68 | 4.676 | 6.161 | 7.258 | 17.294 | 11.846 | 12.486 | 9.264 |
| **Stop_gained** | 55.62 | 53.037 | 52.959 | 53.911 | 54.460 | 55.033 | 52.353 | 52.754 |
| **Stop_gained_novel** | 0.06 | 0.0456 | 0.0635 | 0.109 | 0.315 | 0.1564 | 0.1518 | 0.0754 |
| **Stop_lost** | 15.56 | 15.228 | 15.547 | 15.270 | 15.410 | 15.121 | 13.365 | 13.264 |
| **Stop_lost_novel** | 0 | 0.002 | 0.001 | 0.0014 | 0.072 | 0.006 | 0.016 | 0 |
| **Start_lost** | 9 | 7.986 | 7.966 | 8.1528 | 7.833 | 7.868 | 7.253 | 6.974 |
| **Start_lost_novel** | 0 | 0.006 | 0.006 | 0.0252 | 0.016 | 0.0183 | 0.016 | 0.0188 |
| **Splice_acceptor** | 20.58 | 16.778 | 16.677 | 17.251 | 17.272 | 17.445 | 19.071 | 19.094 |
| **Splice_acceptor_novel** | 0 | 0.018 | 0.0320 | 0.0575 | 0.093 | 0.103 | 0.079 | 0.088 |
| **Splice_donor** | 24.08 | 19.337 | 19.704 | 18.8317 | 18.291 | 18.183 | 17.86 | 17.441 |
| **Splice_donor_novel** | 0.36 | 0.456 | 0.508 | 0.4950 | 0.667 | 0.542 | 0.678 | 0.622 |
| **Synonymous** | 10256.8 | 8861.48 | 8832.39 | 8857.28 | 8771.88 | 8701.69 | 8407.53 | 8335.21 |
| **Synonymous_novel** | 3.2 | 1.972 | 2.577 | 3.265 | 7.447 | 5.262 | 5.445 | 3.943 |

Figure S8.2 illustrates the differential burden of homozygous and heterozygous genotypes in tribal and non-tribal populations. The total number of protein-coding variations (PCVs) was lower in tribal populations, consistent with overall variant trends (Supplementary Figure S3.1.2; Table S3.1.3); however, the heterozygous-to-homozygous ratios remained largely stable across groups, being only marginally lower in the tribal cohort. Additionally, distinct carrier rates for these protein-coding variants were observed (Supplementary Table S8.2).

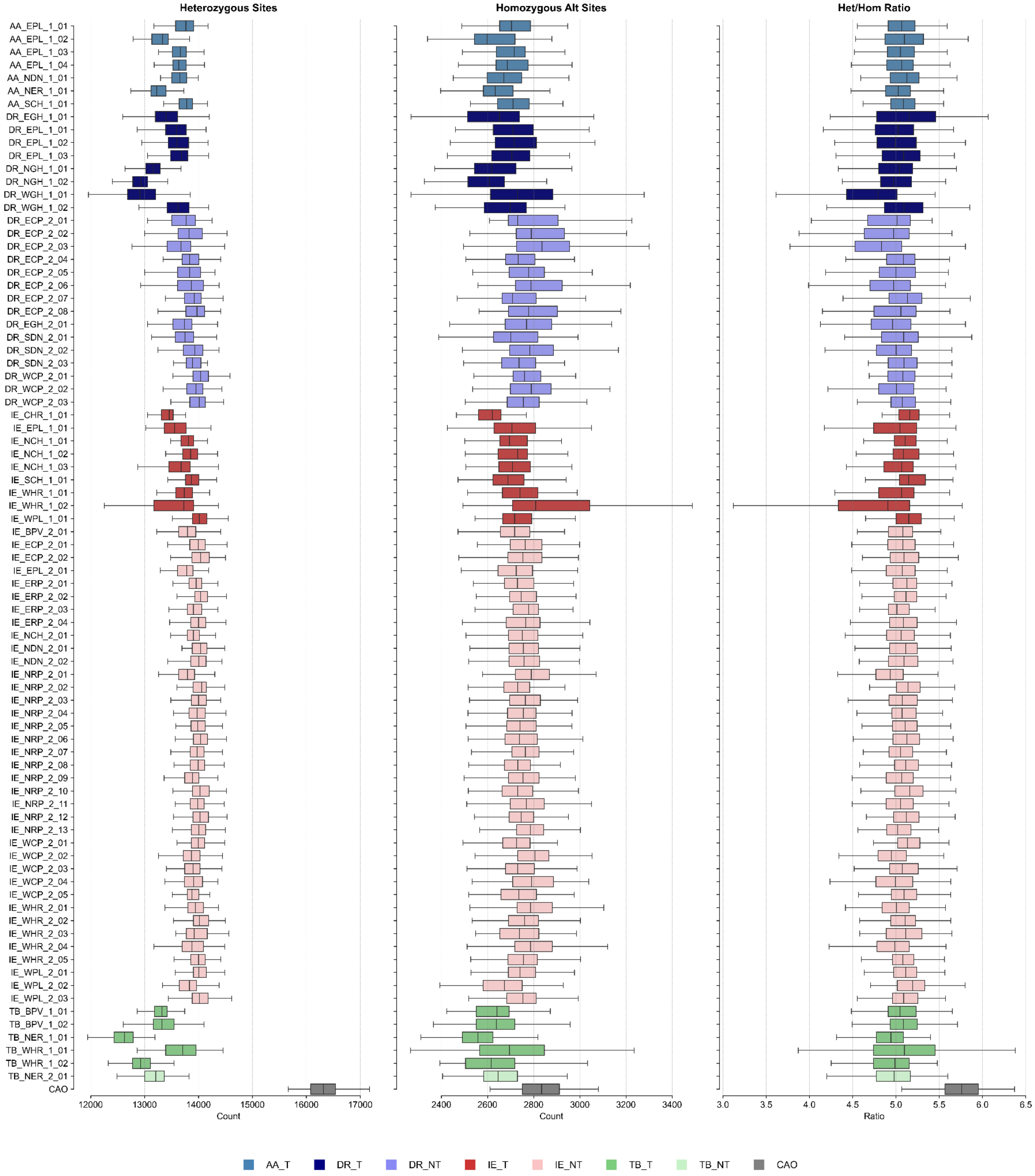

**Figure S8.2.** Box plots showing distribution of heterozygous(left), homozygous(middle) genotypes of protein coding variants across 83 populations and their heterozygous to homozygous ratios (right).

Looking into the landscape of protein-coding variations (PCVs), including missense and synonymous variants, we uncovered that the tribal populations (AA_Tribe, IE_Tribe, DR_Tribe, TB_Tribe), when compared to their non-tribal counterparts, generally exhibit higher burdens (ranging from 152 to 192 PCVs/Sample) (Table S8.3)

**Table S8.3.** PCV distribution across clusters:

| Population | AA_T | IE_NT | CAO | IE_T | DR_T | DR_NT | TB_NT | TB_T |
|---|---|---|---|---|---|---|---|---|
| **PCVs/Sample** | 157.33 | 44.52 | 849.08 | 167.35 | 152.18 | 84.5 | 332.09 | 191.64 |
| **PopSpecific/Total** | 0.052 | 0.192 | 0.18 | 0.027 | 0.073 | 0.077 | 0.0157 | 0.051 |
| **Novel/Total** | 0.009 | 0.01 | 0.001 | 0.005 | 0.017 | 0.008 | 0.002 | 0.009 |
| **Prop_UltraRare** | 0.253 | 0.689 | 0 | 0.298 | 0.188 | 0.482 | 0 | 0.245 |
| **Prop_Rare** | 0.462 | 0.201 | 0.2 | 0.478 | 0.458 | 0.342 | 0.512 | 0.463 |
| **Prop_LowFreq** | 0.120 | 0.043 | 0.33 | 0.09 | 0.16 | 0.07 | 0.196 | 0.119 |
| **Prop_Common** | 0.163 | 0.066 | 0.47 | 0.133 | 0.193 | 0.104 | 0.29 | 0.172 |
| **(UR+R)/C Ratio** | 4.38 | 13.49 | 0.43 | 5.84 | 3.35 | 7.87 | 1.76 | 4.12 |
| **PopSpecific/Sample** | 8.26 | 8.57 | 155.72 | 4.59 | 11.22 | 6.58 | 5.22 | 9.83 |
| **Novel/Sample** | 1.56 | 0.48 | 1.12 | 0.86 | 2.6 | 0.74 | 0.92 | 1.88 |
| **Sample_size** | 601 | 5440 | 50 | 713 | 536 | 1809 | 159 | 460 |

Prop_ = Proportion

***pDMM:***

Using the pathogenicity criteria defined by REVEL scores ≥0.644 and CADD scores ≥20, we identified 25,124 non-singleton putative deleterious missense mutations (pDMM) variants across 7,465 genes. (Detailed analysis in S9)

***Loss of Function Variants:***

We further used the LOFTEE plugin to classify a subset of HIGH IMPACT variants as high confidence (HC) loss of function (LoF) variants. For variants having at least 2 alleles in the population, we observe that 15,849 are high-confidence (HC) LoF spanning 7075 genes. Analysis of these LoF variants reveals distinct distribution patterns across different consequence and allele frequency categories, providing insights into the genetic landscape of LoF variation.

(a)

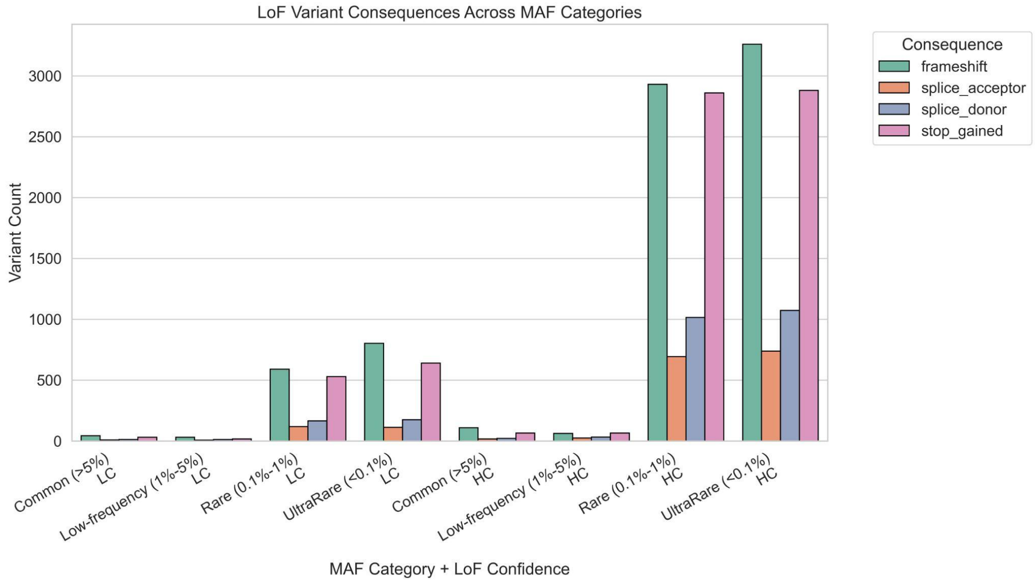
LoF Variant Consequences Across MAF Categories
Consequence
frameshift
splice_acceptor
splice_donor
stop_gained
Variant Count
0
500
1000
1500
2000
2500
3000
Common (>5%) LC
Low-frequency (1%-5%) LC
Rare (0.1%-1%) LC
UltraRare (<0.1%) LC
Common (>5%) HC
Low-frequency (1%-5%) HC
Rare (0.1%-1%) HC
UltraRare (<0.1%) HC
MAF Category + LoF Confidence

(b)

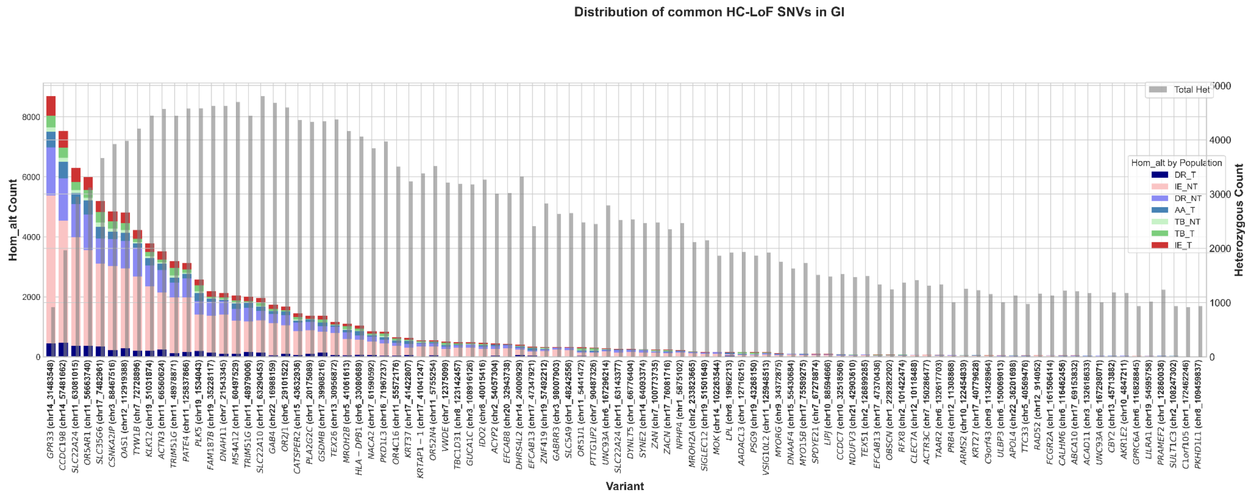
Distribution of common HC-LoF SNVs in GI
Hom_alt Count
Heterozygous Count
Total Het
Hom_alt by Population
DR_T
IE_NT
DR_NT
AA_T
TB_NT
TB_T
IE_T
Variant

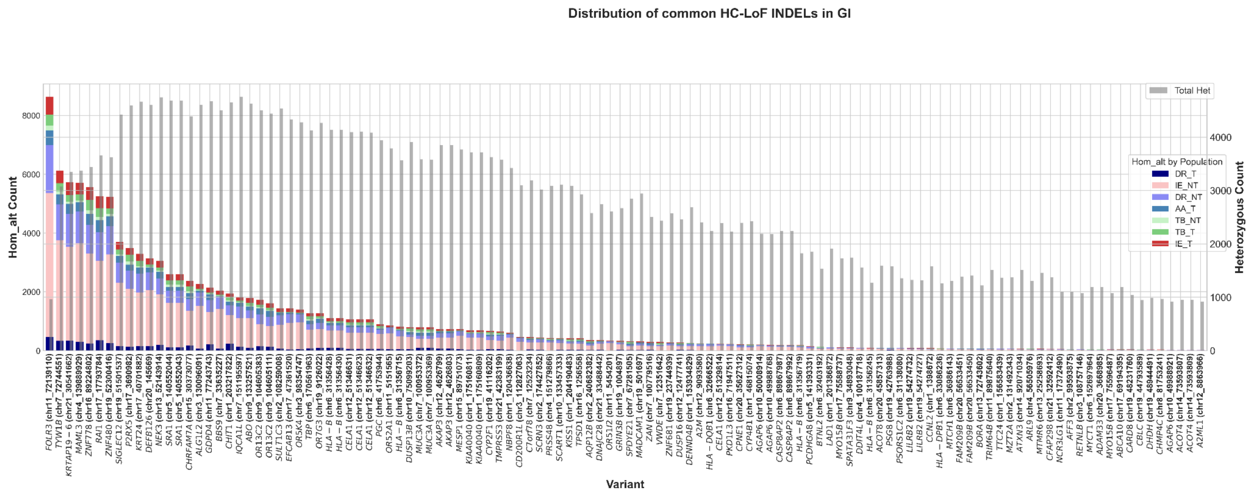

**Figure S8.3**. ***(a)*** *Counts of different HC-LoFs and LC-LoFs (Low confidence) in the dataset having at least 2 minor alleles in the population. The majority of the HC-LoFs are in the ultrarare category.* ***(b)*** *Distribution of HC-LoF variants [Top-SNV, Bottom-Indel] which are common in the population (>5%). Each stacked bar represents the distribution of homozygous alternate (1/1) genotyped variants in different populations, and the grey bars indicate the number of heterozygous variants for that variant in the whole GI population. For INDELs, rs137962621 and rs111599831 are excluded from the plot as they are exclusively present in outlier samples and the CAO population.*

High-confidence loss-of-function (HC LoF) variants are predominantly observed in the rare (0.1%–1%) and ultra-rare (<0.1%) categories, quite expectedly because of their deleterious nature. Among the consequences, frameshift and stop_gained variants are the most frequent, followed by splice donor and splice acceptor disruptions, all of which can severely impair gene function (Supplementary figure S8.3). This highlights that impactful LoF events are generally kept at low frequencies in the population due to their potentially damaging effects (Supplementary S10 has the LoF details in populations). We also studied the LoFs that were common in the population (>5% MAF) and Gene Ontology analysis showed overrepresentation of the less-constrained immune and olfactory genes (Supplementary figure S8.3,), a well-known phenomenon in population genomics (Ignatieva et al., 2014; Karczewski et al., 2020).

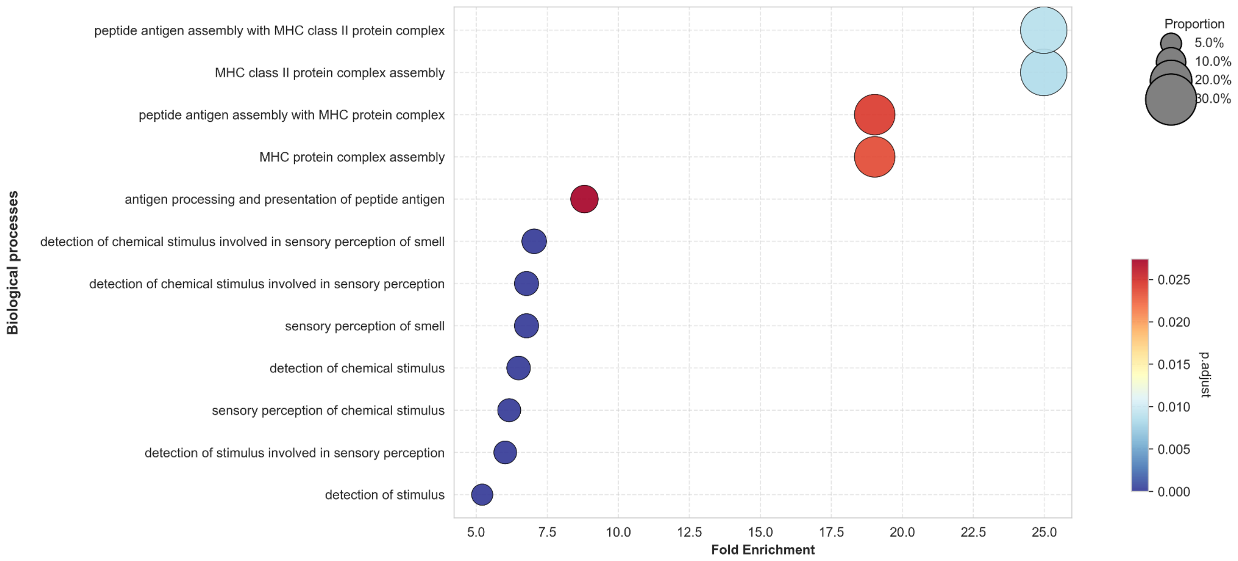

**Figure S8.4:** Functional enrichment analysis of common LoF variants. Dot plot showing significantly enriched Biological Process terms for genes harboring LoF variants with a MAF >= 5%. The x-axis indicates fold enrichment, representing the ratio of observed to expected genes in each category. The bubble size represents the proportion of overlap (the ratio of LoF-impacted genes to the total number of genes in the GO term), while the color gradient represents the FDR-adjusted p-value, with blue indicating the most significant enrichment (p.adjust < 0.01).

#### Pathogenic variants

We characterized the landscape of clinically relevant variation within the dataset, identifying 986 variants annotated as Pathogenic(P) or Pathogenic/Likely Pathogenic(P/LP) in ClinVar. Of these, 858 were classified as missense or high confidence loss-of-function (HC- LoF) variants. Notably, 244 of these variants exhibited a minor allele frequency (MAF) of ≥ 1% across the 83 analyzed populations. Most of the variants that have MAF ≥ 10% are originating from AA_T and DR_T populations. The highest prevalence was observed in an HFE missense variant, which reached a MAF of 26.6% in the AA_EPL_1_02 population (Figure S8.5). A comprehensive inventory of these P and P/LP variants is provided in Supplementary Table S8.4.

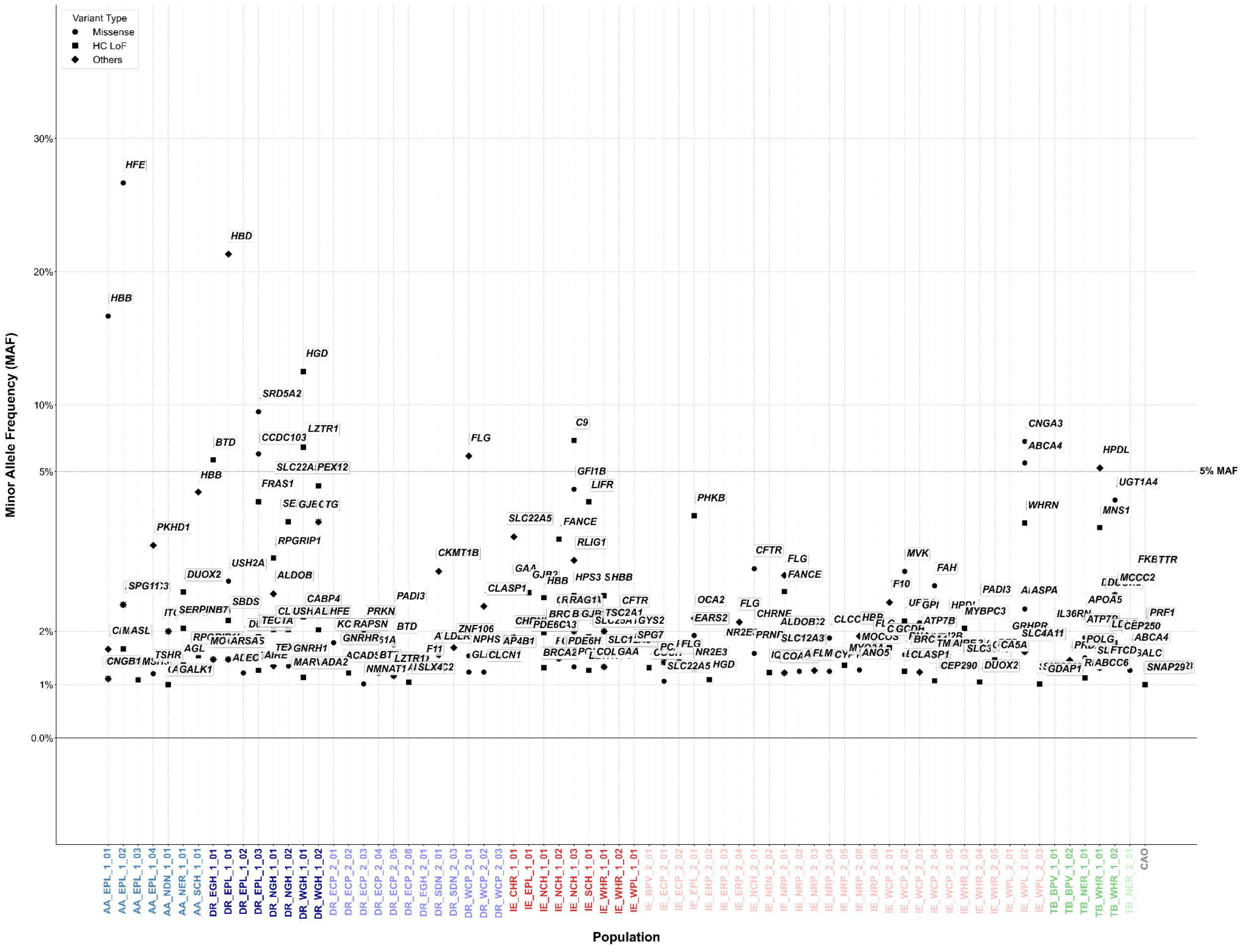

**Figure S8.5:** Minor allele frequency (MAF) distribution of top pathogenic (P) and likely pathogenic (P/LP) variants across populations. "Top" variants represent those reaching their highest frequency in the population where they are most prevalent; only variants with MAF ≥ 1% are shown. Underlying data also available to peruse in Table S8.4.

This plot displays the frequency of specific genomic variants within the population where they are most prevalent. Only variants with a MAF >=1% are shown. The y-axis utilizes a non-linear scale to expand the 0–5% frequency range for higher resolution, with a horizontal grid line marking the 5% threshold. Data points are distinguished by shape based on variant consequence: circles represent missense variants, squares represent high-confidence loss-of-function (LoF) variants, and diamonds represent other variant types. Each point is annotated with its corresponding gene symbol.

#### Supplementary Section S9: Landscape of putative deleterious missense mutations (pDMM)

Priyanka Singh[1], Sauma Suvra Majumdar[1,2], Ankit Mukherjee[3], Mohammed Faruq[3,4], Bratati Kahali[1]

[1]Centre for Brain Research (CBR), IISc Campus, Bengaluru, India. [2]Interdisciplinary Mathematical Sciences, Indian Institute of Science (IMI- IISc), Bengaluru, India. [3]CSIR - Institute of Genomics & Integrative Biology (CSIR-IGIB), New Delhi, India. [4]Academy of Scientific and Innovative Research, Ghaziabad, India.

#### Summary

In this section, we identified the putative deleterious missense variants (pDMMs) by applying pathogenicity thresholds of REVEL and CADD scores. We then characterized these pDMMs by assessing their novelty, population-specific patterns across linguistic groups and ethnicities, and clinical relevance.

##### Methods

To evaluate the potential impact of missense variants, we applied two predictive scoring systems: REVEL (Rare Exome Variant Ensemble Learner) (Ioannidis et al., 2016) and CADD (Combined Annotation Dependent Depletion) (Rentzsch et al., 2019). Missense variants with REVEL scores ≥0.644 and CADD scores ≥20 were considered deleterious (Bergquist et al., 2024; Ioannidis et al., 2016; Rentzsch et al., 2019). We assessed the pathogenicity of the variants by cross-referencing with ClinVar and identified known pathogenic mutations in the set of pDMMs. ClinVar file was downloaded from the NCBI FTP repository https://ftp.ncbi.nlm.nih.gov/pub/clinvar/tab_delimited/ in February 2025. Variants annotated as pathogenic (P) or pathogenic/likely pathogenic (P/LP) (review status ≥2 stars) were classified as clinically relevant.

Additionally, novel missense variants absent from public variant repositories i.e. dbSNP, gnomAD, 1000G and GenomeAsia were identified and population-specific patterns across the major Indian linguistic groups and ethnicities were investigated.

##### Results

###### *Identification and Frequency Spectrum of pDMMs*

In total, we identified 25,124 pDMM (non-singletons) across 7,465 genes (Supplementary Table S9.1). As expected, these variants were predominantly rare, with only ~1% (N = 272) showing an MAF ≥1%. Among 25,124 variants, 2,223 pDMMs across 1,809 genes were absent from global variant databases (Supplementary Table S9.1, column J). The distribution of allele frequencies across linguistic groups revealed majority of these pDMMs were rare but a fraction of variants in tribal populations, showed a shift toward higher allele frequencies, consistent with the effects of long-term endogamy and genetic drift that could elevate deleterious allele frequencies in isolated populations.

##### *Cluster-wise Enrichment and Founder Signatures*

Clusterwise frequency distribution also revealed 612 pDMMs displaying elevated allele frequencies in at least one of the population clusters (Supplementary Table S9.2). A detailed list of 612 variants and corresponding frequencies are provided in Supplementary Table S9.1 (column V). Notably, tribal groups such as DR_T (168 variants) and AA_T (140 variants) harbored a larger number of pDMMs occurring at relatively higher allele frequencies compared to other clusters (except TB_NT), likely shaped by founder effects and population isolation.

**Table S9.2.** Enriched pDMMs with higher allele frequency (MAF ≥ 0.01) in specific linguistic clusters

| Population Cluster | No. of enriched pDMMs (MAF ≥0.01 in cluster & MAF <0.01 in all others) |
|---|---|
| DR_T | 168 |
| AA_T | 140 |
| IE_T | 22 |
| TB_T | 86 |
| DR_NT | 39 |
| IE_NT | 15 |
| TB_NT | 142 |

In addition, we identified 24 variants that were common to both DR_T and AA_T tribes but remained rare in other populations (Supplementary Figure S9.1). Among these, a notable signal in *SPINT1*, with allele frequencies of ~6% in DR_T and ~3% in AA_T, while remaining rare across other clusters. Of the total 251 individuals carrying this *SPINT1* variant, 61 belonged to DR_T (40 from DR_NGH_1_02/DR_NGH_1_01), and 38 belonged to AA_T (28 from AA_EPL_1_03/AA_EPL_1_04), indicating that this variant is concentrated within specific subgroups. Reduced SPINT1 levels are implicated in placental insufficiency and elevated stillbirth risk, and Spint1 deficiency leads to lethality in mice (Kaitu'u-Lino et al., 2020). Taken together, this deleterious missense variant warrants future investigations in these populations to assess potential reproductive and clinical consequences. Similarly, rs560961886 (G>T) variant in *CYSLTR2* shows elevated frequencies in DR_T (MAF = 2.45%) and AA_T (MAF = 1.09%) clusters. Among the 93 carriers, 25 were from DR_T (23 from DR_NGH_1_01) and 12 were from AA_T (10 from AA_EPL_1_04), indicating marked population-specific enrichment. *CYSLTR2* encodes a leukotriene receptor targeted by gemilukast, a dual $CysLT_1R$/$CysLT_2R$ antagonist used in asthma management (Itadani et al., 2015), highlighting potential pharmacogenomic relevance in these tribes.

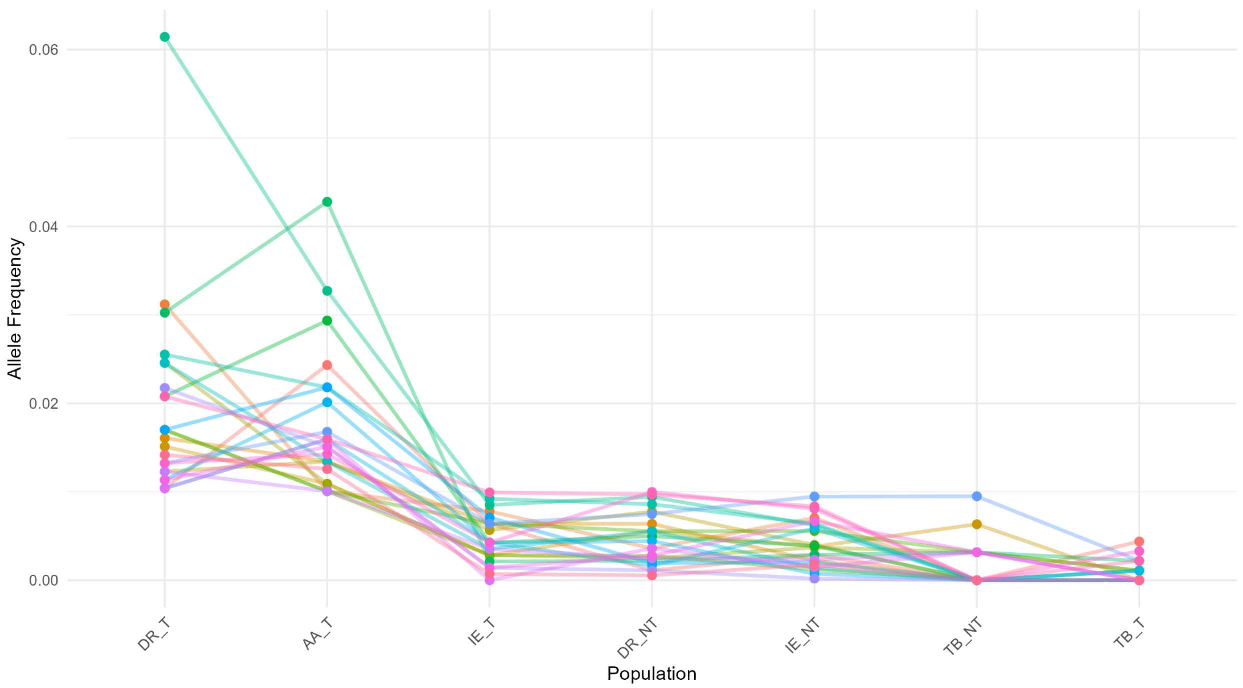

**Figure S9.1.** Allele frequency distribution of 24 variants with higher allele frequency in DR_T and AA_T (MAF ≥ 0.01) compared to other clusters (MAF < 0.01).

We also examined allele frequency distributions at finer resolution across individual populations. We investigated population-specific allele frequency enrichment across 82 populations (excluding CAO) and 9,651 variants that exhibit a pattern of being globally rare (MAF <0.01) yet locally enriched (MAF≥ 0.01) in specific populations. These variants are summarized in Supplementary Table S9.3.

***Allele frequency comparison with gnomAD***

We assessed minor allele frequency (MAF) differences across all linguistic population clusters by comparing them with the corresponding frequencies reported in gnomAD. A comparative analysis of these pDMM allele frequencies between the GenomeIndia (GI) cohort and the south-Asian specific gnomAD dataset (gnomAD-SAS) revealed substantial population-specific differences. These disparities were particularly pronounced in the tribal groups, notably the DR_T and AA_T populations. For example rs201493353:C>T in *SPAG1* (Sperm-associated antigen 1) showed an MAF of 3% in DR_T compared to 0.03% in gnomAD-SAS. While no homozygotes are reported in gnomAD, 49 heterozygotes and 2 homozygotes were observed in GI, primarily from DR_T (DR_NGH_1_01 and DR_NGH_1_02), known to exhibit founder effects (Supplementary section S4). The distinct allele frequency spectrum observed in these groups underscores the strong demographic and evolutionary imprint shaping the distribution of deleterious variants within Indian populations (Supplementary Figure S9.2).

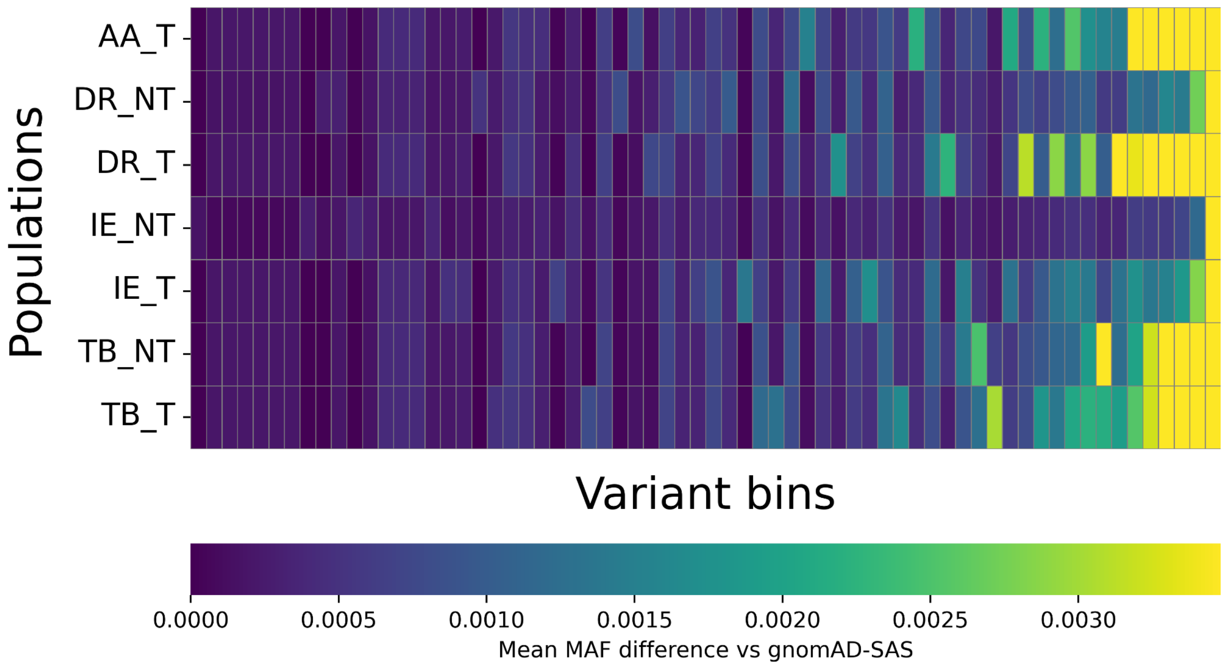

**Figure S9.2.** Heatmap showing MAF differences of all pDMM between GI and gnomAD-SAS. The plot illustrates MAF differences for pDMM across Indian tribal and non-tribal linguistic groups relative to gnomAD. The differences in variants are binned into groups of ~400 and plotted for better visualization. Yellow represents greater divergence in allele frequencies, while purple represents minimal divergence.

***ClinVar annotated pathogenic variants***

ClinVar annotated 285 pDMMs as P or P/LP (Supplementary Table S9.1, column K-M). Among these, the most frequent variant, rs1800553 in *ABCA4*, is a risk factor of Stargardt disease, retinitis pigmentosa, early-onset retinal dystrophy, and age-related macular degeneration. It was identified in 225 individuals in the heterozygous state and in 2 individuals as homozygotes, appearing in multiple populations but reaching their highest frequencies in IE_WPL_2_02 (~6%), indicative of endogamy. Another notable pathogenic *BCHE* variant (rs104893684) associated with butyrylcholinesterase deficiency, which increases sensitivity to muscle relaxants, is found in 29 populations and exceeds MAF ≥1% in three groups, with frequencies approaching ~2% in DR_ECP_2_01 and IE_NRP_2_01.

***Variants Reported in the GWAS Catalog***

We performed a look-up of the 25,124 pDMMs on the EBI-GWAS catalog (https://www.ebi.ac.uk/gwas/) to check for known disease associations of these variants. We found that 155 variants in 147 genes were previously reported in the GWAS Catalog (Supplementary Table S9.1, column N). Importantly, 67 of these variants, although globally rare, are enriched in at least one of the 82 populations. The overlap of these pDMMs were observed for loci associated with complex disease traits, example, Alzheimer's disease, early-onset schizophrenia, type 2 diabetes, coronary artery disease, chronic inflammatory disorders, nonalcoholic fatty liver disease, chronic obstructive pulmonary disease, stroke, and multiple types of cancers. This phenomenon is consistent with a polygenic disease architecture, in which modest functional perturbations across many genes contribute incrementally to risk rather than acting as highly penetrant causal mutations. In Indian population-specific cohorts, where such coding variants can differ substantially in frequency, enable the identification of risk-modifying alleles and pathways that may be not so prominent in predominantly European studies. The knowledge of these variants can help increase power in rare-variant burden and gene-based collapsing tests in population-specific manner, while also offering a principled framework for enhancing polygenic risk scores through functional annotation–informed weighting.

#### Supplementary Section S10: Spectrum and Clinical Relevance of Loss-of-Function Variants

Shouvanik Sengupta[1,2], Analabha Basu[1,2], Chandrika Bhattacharyya[1]

[1]BRIC - National Institute of Biomedical Genomics (BRIC-NIBMG), Kolkata, India. [2]Regional Centre for Biotechnology (RCB), Faridabad, India.

##### Summary

In this section, we characterize the landscape of high-confidence Loss-of-Function (HC-LoF) variants identified in the GenomeIndia dataset, along with their potential functional and clinical significance. We have compared with major global genomic datasets to evaluate the distribution and novelty of these HC-LoF variants. Our findings highlight a significant number of LoF variants that are both population-specific and clinically relevant, underscoring the importance of studying populations of diverse ancestries in genomics-driven public health strategies.

###### Methods

Using the LoFTEE (version 1.0.4_GRCh38) (Karczewski et al., 2020) plugin of VEP 113.0, we identified 45,031 high-confidence loss-of-function (HC-LoF) variants on the canonical transcripts of 12,067 genes. These HC-LoF belong to the following classes of variants: frameshift, splice donor, splice acceptor, and stop-gained. For comparison with global datasets, we have considered 1000 Genomes (1000G), gnomAD v4, and GA100K. To assess the clinical relevance of these HC-LoFs, we have considered ClinVar (Landrum et al., 2014) annotations.

###### Results

Out of 45,031, 29,182 are singletons. 15,849 non-singleton HC-LoFs were mapped to 7,076 genes. Among the non-singletons, 3,898 variants were identified as unreported in 1000 Genomes (1000G), gnomAD, and GA100K. These unreported HC-LoFs correspond to 2,976 genes. Notably, we discovered 1,346 unreported HC-LoFs mapped to 1,171 genes that have no previously reported HC-LoFs. Additionally, we identified 2,552 unreported HC-LoFs occurring alongside 4,830 previously reported HC-LoFs in 1,805 genes.

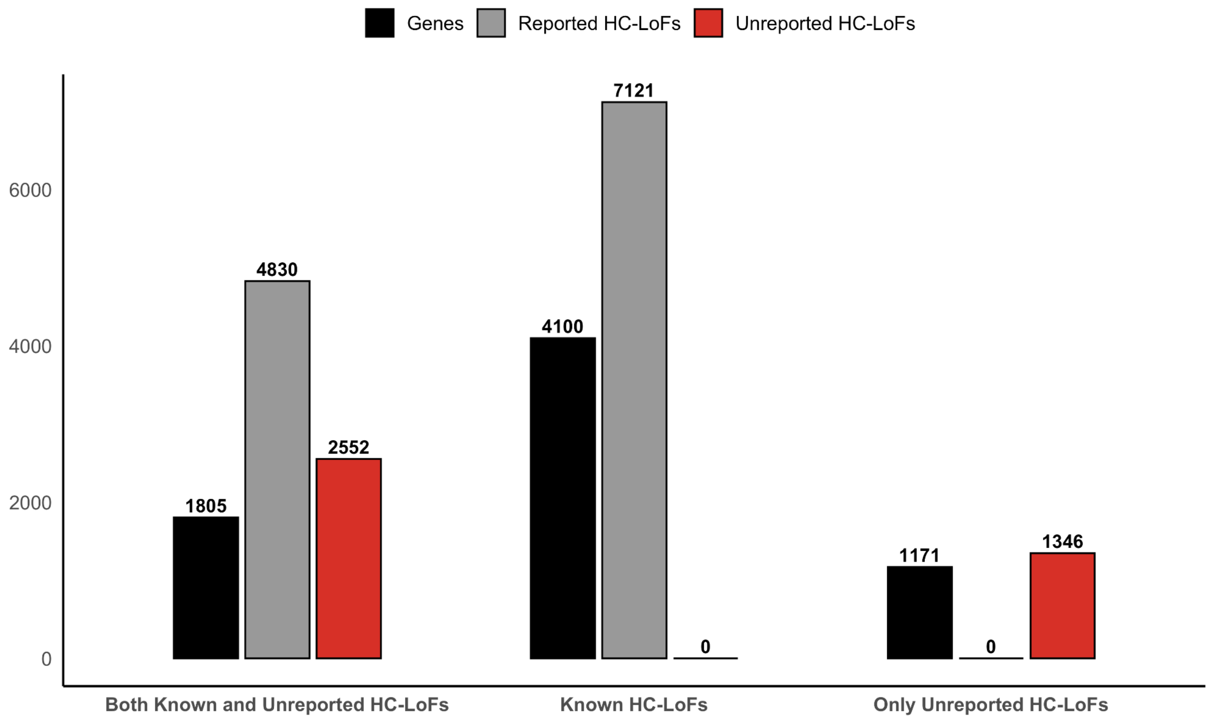

**Figure S10.1.** Total number HC-LoFs and the mapped Genes. Here, the singleton LoFs are not considered.

Most genes are observed to contain 1 HC-LoF, as accumulation of multiple HC-LoFs in a single gene is typically prevented by evolutionary constraints. We identified at least 590 genes where we discovered one additional, previously unreported HC-LoF alongside one already known, which could be informative about how the evolutionary constraint is acting on them.

Genes with Probability of Loss-of-function Intolerant (pLI) more than 0.9, are typically considered as intolerant to loss-of-function. Here, we discovered 165 genes, with pLI >= 0.9, harboring both newly discovered and previously reported HC-LoFs. It highlights the importance of studying isolated/underrepresented populations in refining the evolutionary constraint metrics and improving population-based predictions in medical genetics.

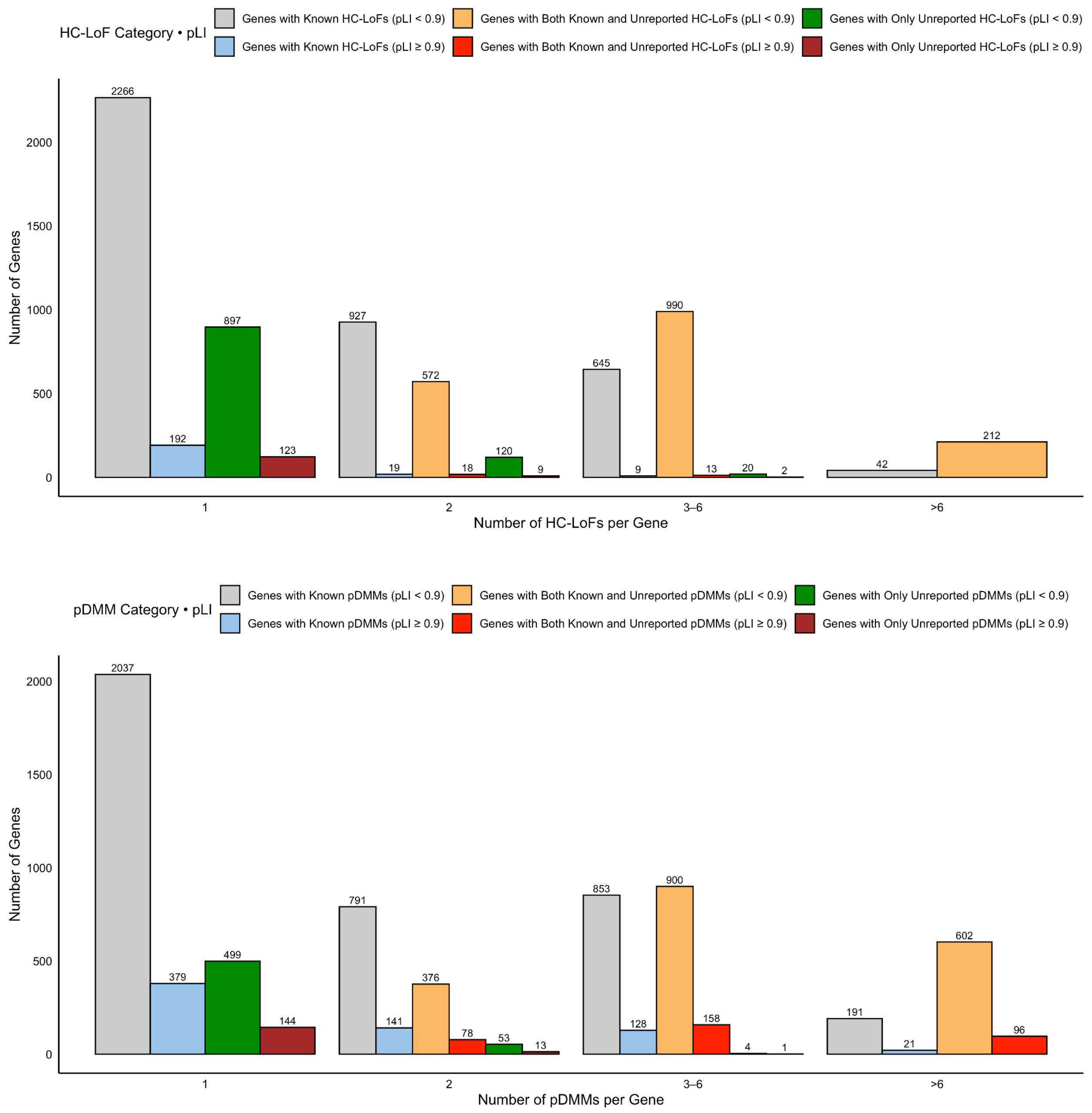

**Figure S10.2.** Number of HC-LoFs (top) and pDMMs (bottom) observed in a gene**.**

On average, each individual carries 27.6 HC-LoFs as homozygous and 78.97 HC-LoFs as heterozygous, with a population-specific pattern as shown in the figure S10.3. There are 43 populations that have an average number of HC-LoFs at the individual level of more than 27.6, of which 25 are of tribal ancestry.

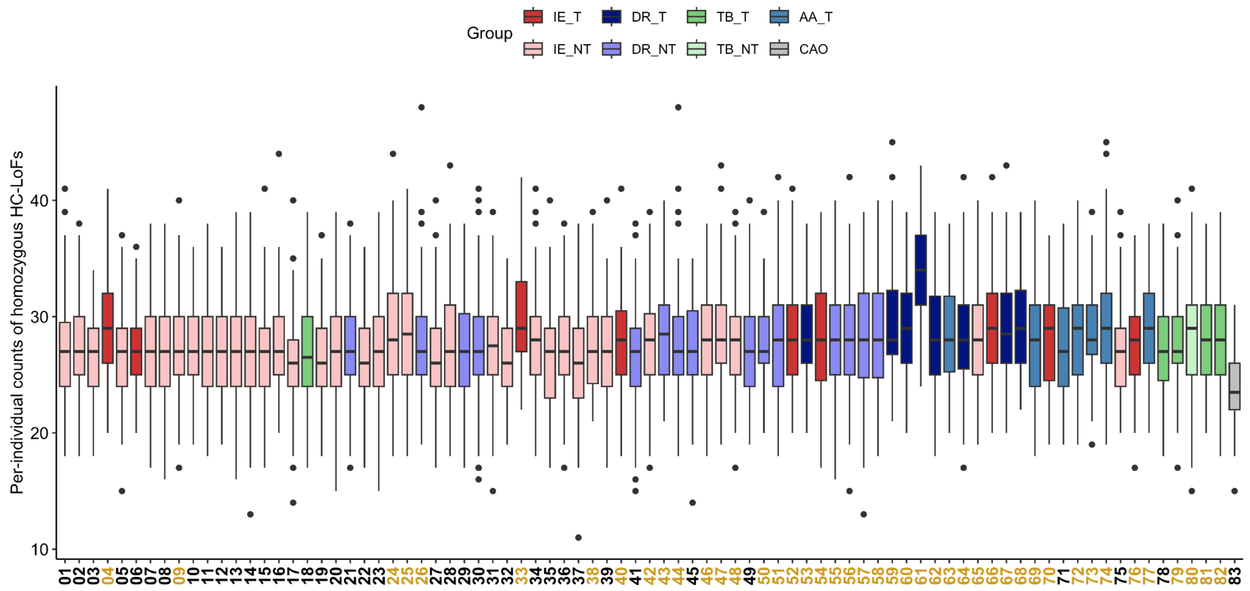

**Figure S10.3.** Distribution of homozygous loss-of-function variants (HC-LoFs) per individual across populations. Populations with a higher mean burden of homozygous LoFs than the GenomeIndia dataset average (27.6 LoFs per individual) are highlighted in gold.

Among these 3,898 previously unreported HC-LoFs, to prioritize variants for potential pathogenic roles, we compared their allele frequencies across populations. Since truly pathogenic HC-LoFs are unlikely to occur at high frequencies in multiple populations, we analyzed their distribution by assessing both the number of populations in which they are observed and the maximum allele frequency in any single population.

We observed 19 HC-LoFs which are unlikely to be pathogenic, as these are present across multiple populations in relatively high frequency (Figure S10.4).

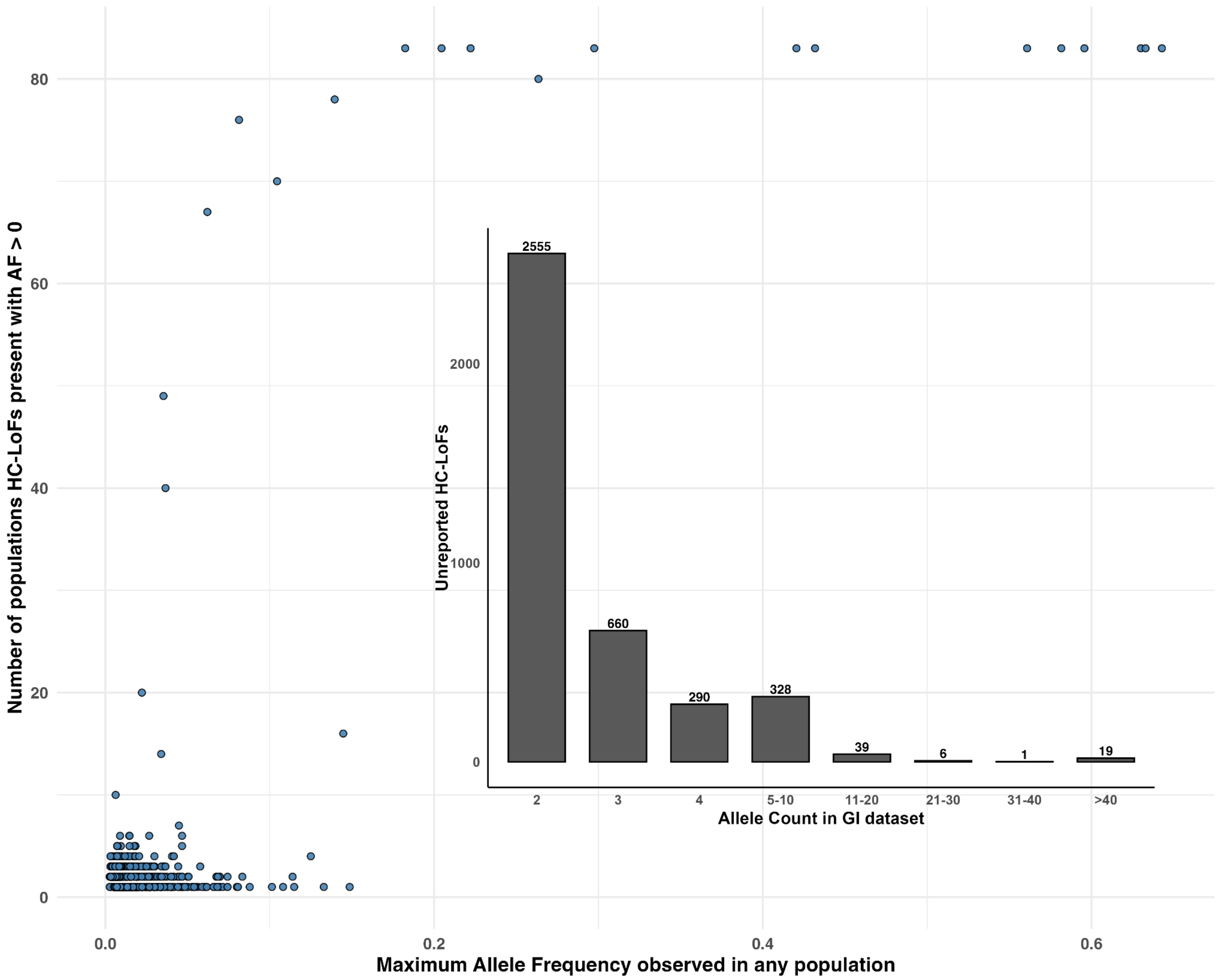

**Figure S10.4.** Distribution of previously unreported loss-of-function (LoF) variants across populations, stratified by the number of populations in which each variant was observed and by the maximum allele frequency recorded. The inset depicts the allele count distribution of these LoF variants across the entire dataset.

In contrast, we observed 1,402 LoFs exhibiting population-specific patterns, each with an allele frequency greater than 1% in at least one population (Table S10.1). We considered these LoFs as candidates for potential pathogenic roles, possibly increased in frequency due to population-specific demographic processes. Taking into consideration that the minimum sample size is ~50 in GI populations. So, we have further selected the variants that are non-singletons within a population

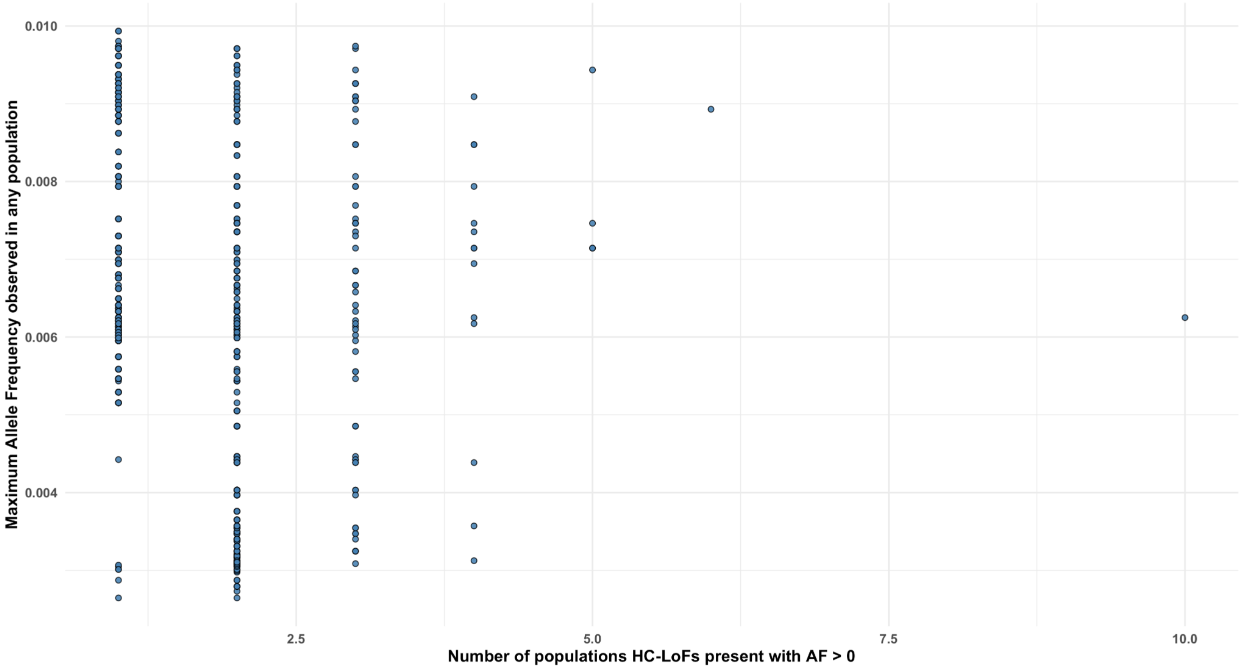

**Figure S10.5.** Distribution of 2477 unreported LoFs across all the populations, which are NOT considered for further analysis.

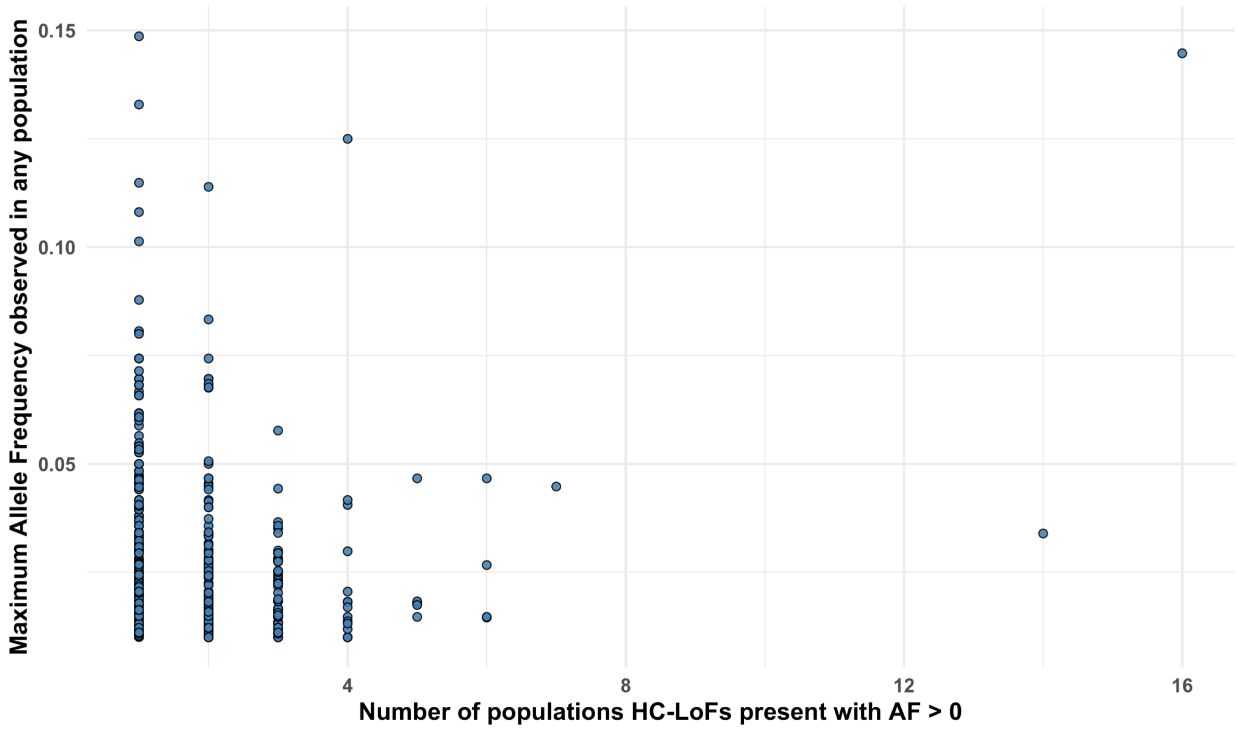

**Figure S10.6.** Distribution of 1402 unreported LoFs across all the populations, which are considered for further analysis. These variants exhibit population-specific patterns, each with an allele frequency greater than 1% in at least one population

These 1402 LoFs mapped to 1250 genes, with 57 genes with pLI >= 0.9. Although no homozygous individuals were observed for these high pLI genes, the corresponding LoFs were present at moderate frequencies in several isolated populations. Given the small effective population sizes and endogamous practices of these groups, there remains a risk of homozygous occurrence for these variants in future generations.

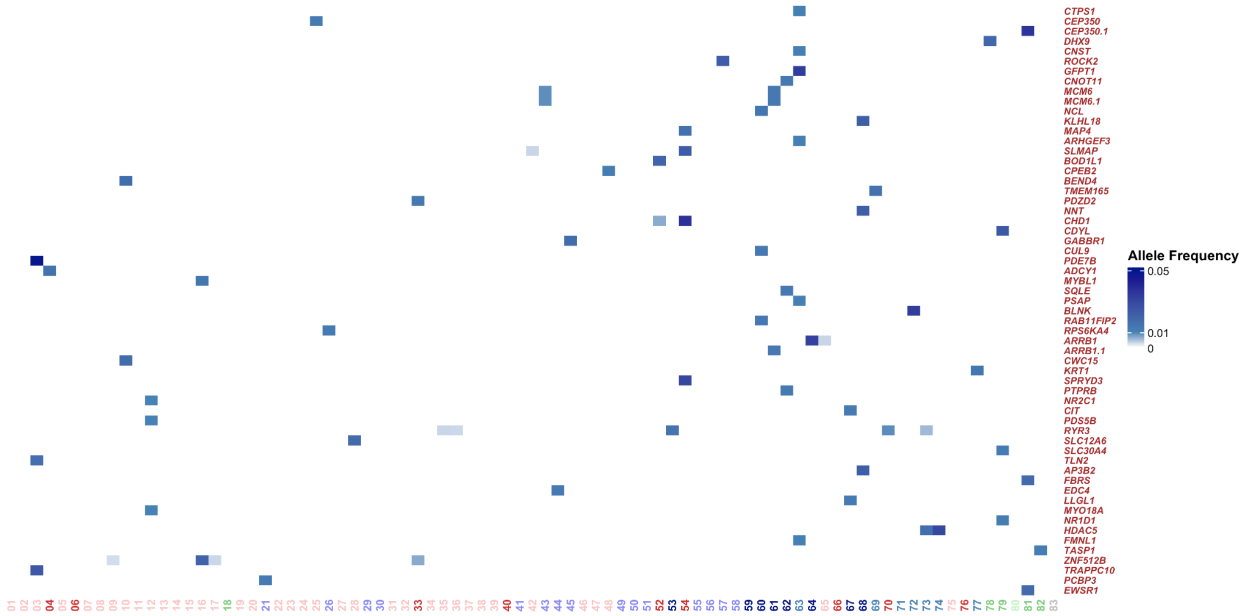

**Figure S10.7.** HC-LoFs which are present in <= 20 populations and in >= 1% AF in the populations in the genes with high pLI (>= 0.9).

We identified a total of 2,868 individuals carrying these 1401 HC-LoFs, including 60 individuals from 32 populations, who were homozygous for 54 distinct HC-LoFs. Out of the 51 genes to which these HC-LoFs are mapped, one notable example is *LAIR1*, where we observed the variant in 31 individuals across 16 populations. Importantly, 3 individuals were found to carry the variant in the homozygous state. *LAIR1* (Leukocyte-associated immunoglobulin-like receptor 1), plays a role in the human immune response to malaria, particularly in severe malarial anemia (SMA) (Achieng et al., 2019). We also noted that this variant is present among the individuals in the populations from malaria-endemic regions.

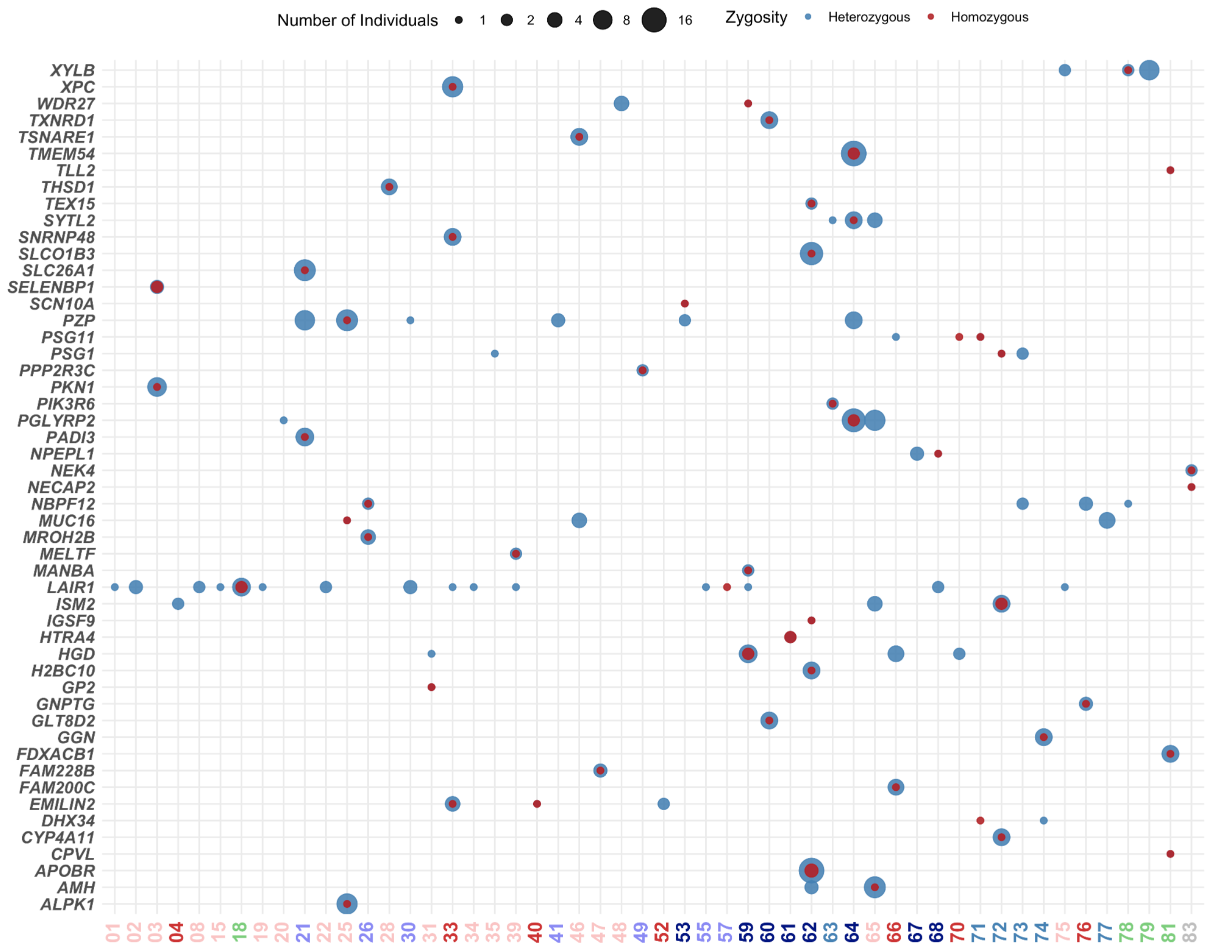

**Figure S10.8.** Dot plot visualization of the homozygous burden of previously unreported loss-of-function (LoF) variants across populations. The x-axis represents the 51 genes harboring these LoF variants.

To get a deeper insight into the clinical relevance of non-singleton HC-LoFs, we considered ClinVar annotations (≥2-star review status). A variant was considered of clinical relevance if it was annotated as pathogenic or pathogenic/likely pathogenic by ClinVar. Thus, taking the intersection of the two tools, we identified 483 variants (301 SNVs and 182 InDels) (Figure S10.9), of which 94.2% of variants were rare (0 < x < 0.01) and 19 were not reported in previous population genomic studies (1000 genomes, gnomAD and GA100K).

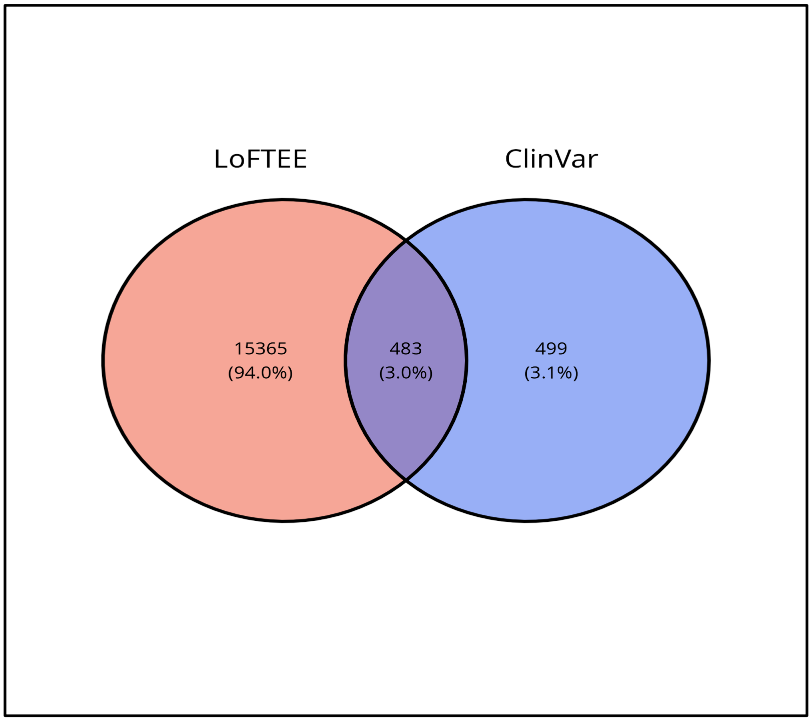

**Figure S10.9.** Venn diagram showing distribution of pathogenic variants identified by the two tools

Among the 483 variants, 219 are doubletons and 223 have allele count of 3-10 (Fig S10.10). Only three variants contribute to the >50 allele count bin - chr15:45101227 TGAAC>T (allele count 55), chr7:66994210 A>G (allele count 87) and Chr13:20189511 C>T (allele count 131).

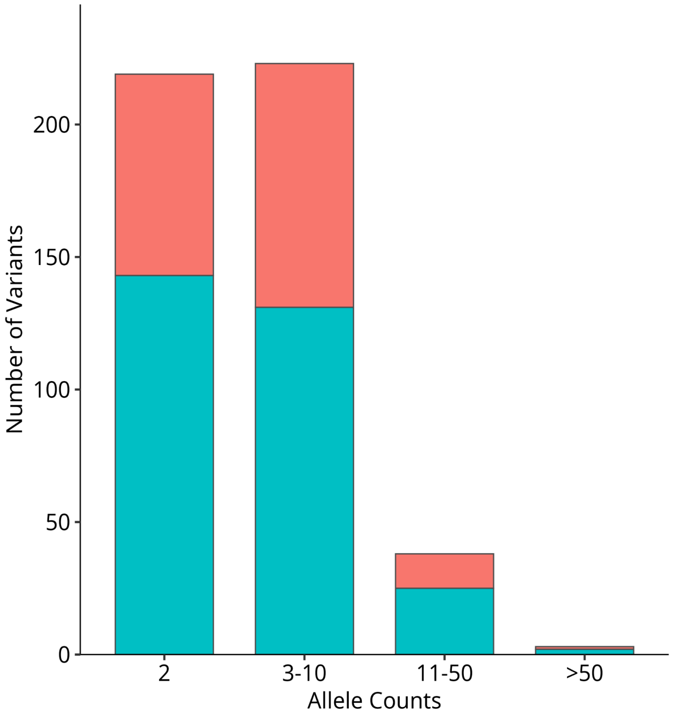

**Figure S10.10.** Allele count distribution of the 483 variants, divided into 5 bins and categorized by variant type.

2,139 individuals (22.08% of GI) carry at least one of these 483 variants (Fig S10.11). 10 individuals carry one of these variants in a homozygous condition, and they are only from Indo-European and Dravidian groups. Each of these individuals belongs to a different population, except for two individuals from population DR_WGH_1_01 (59) - a Dravidian tribal population (Fig S10.11).

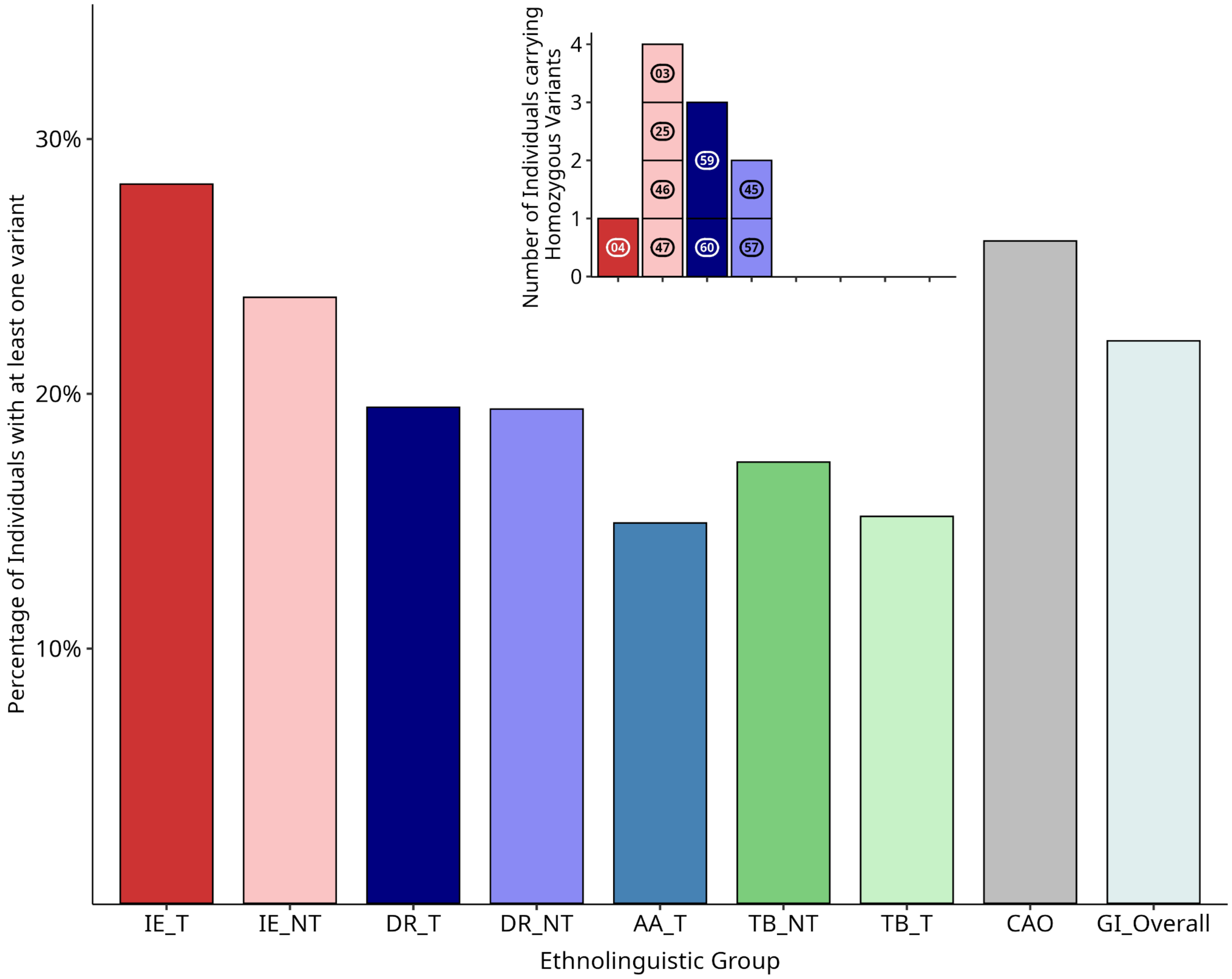

**Figure S10.11.** Percentage of individuals in the ethnolinguistic groups carrying at least 1 variant among the 483, in heterozygous or homozygous state. The inset plot shows the number of individuals, belonging to the population and the groups, carrying one of these variants in a homozygous state.

We observed that some populations carry many clinically relevant variants at relatively high frequencies, likely due to their demographic history and higher levels of homozygosity (Figure S10.12). As in the figure, the populations that are in the left pane harbor these pathogenic HC-LoF variants in high frequency. These populations, mostly tribal, also showed a distinctive pattern in their demographic history. Populations, influenced by both founder effects and consanguinity, may therefore be important targets for carrier screening and rare disease gene discovery.

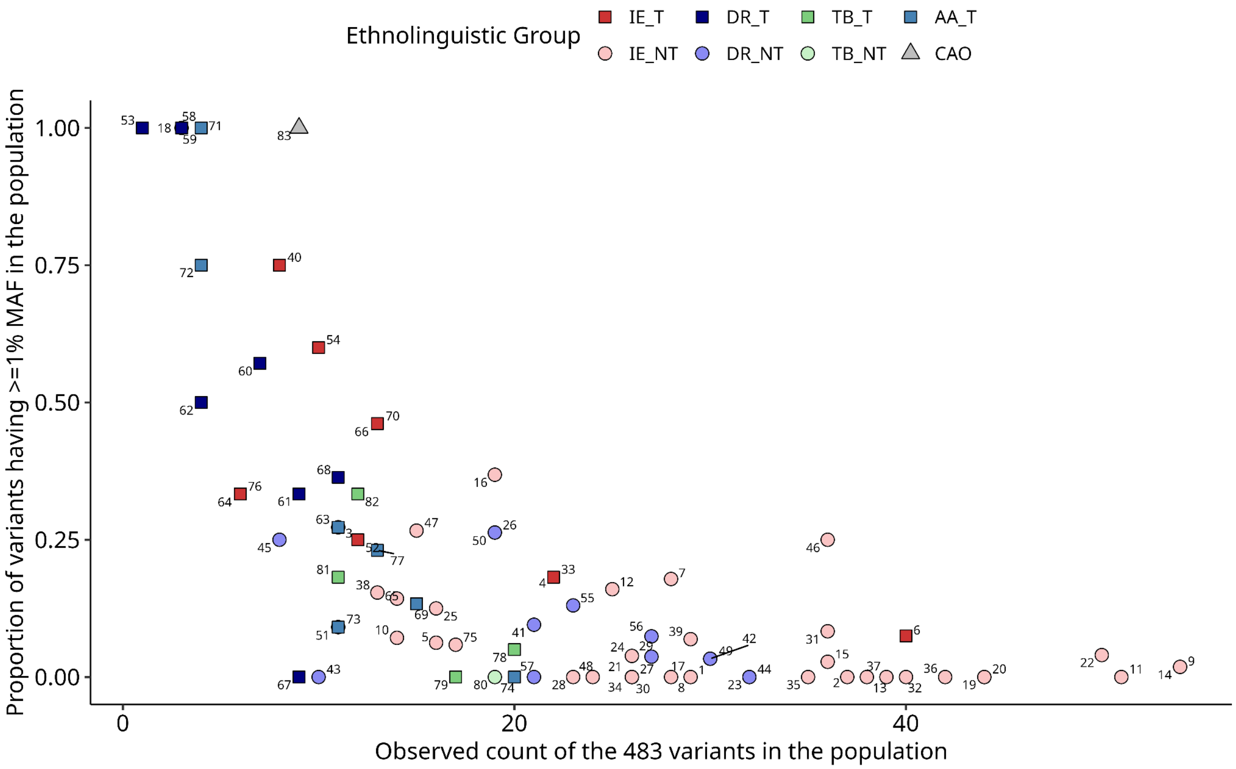

**Figure S10.12.** Population-wise distribution of the 483 variants, showing the observed counts in the populations on the X-axis and the proportions of the variants that have ≥1% allele frequency among the observed ones in the populations, on the Y-axis.

Among the 483 variants, the highest observed allele frequency in the overall GenomeIndia superpopulation was just 0.6%. However, when we looked at the maximum observed allele frequency in any of the 83 populations, we saw 133 variants having frequency ≥1% and 4 having ≥5%, in at least one of the populations (Table S10.2). This indicates the high degree of heterogeneity in the frequency distribution across the different populations. Compared to global data from 1000 genomes, gnomAD and GA100K, 127 variants are rare (<1%) or unobserved there but have ≥1% frequency in at least one GenomeIndia population (Fig 10.13).

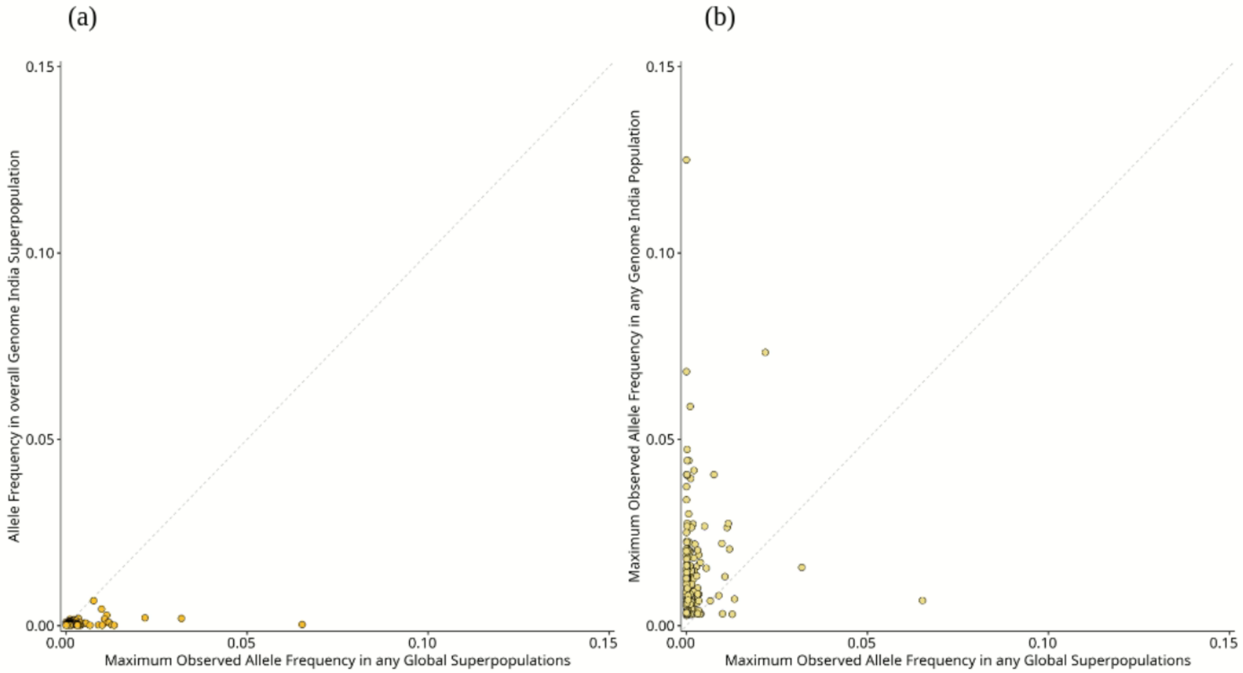

**Figure S10.13.** Allele frequency comparisons between maximum observed frequency in any of the global (1000 genomes, gnomAD and GA100K) superpopulations and - (a) frequency in overall GI dataset and (b) maximum frequency in any of the 83 constituent populations.

The variant, which is present in the highest frequency in pan-GI data (0.6%), is a stop-gained in the *GJB2* gene, associated with hearing loss (13:20189511, p.Trp24Ter). This variant is present in 131 individuals, in a heterozygous condition, across 46 populations, while in 17 populations, as more than 1%. Out of all the variants not reported in previous genomic studies such as GA100K, 1000G, and gnomAD, one variant is in the *HGD* gene (3:120675791, c.87+1G>A), associated with Alkaptonuria. This variant is present in more than 1% across three isolated populations, while in one population, the frequency is 12.5% (Population 59); there are two individuals with a homozygous condition.

Overall, these 133 variants are annotated to 115 genes, associated with predominantly metabolic and intellectual disorders (Fridman et al., 2021). Notably, we identified variants in *SBDS* (Shwachman-Diamond syndrome: an exocrine pancreatic dysfunction), *GYS2* (Glycogen storage disease) and *XDH* (Xanthinuria) genes, present across multiple (> 20) populations, even for some in more than 1% frequencies.

Proceeding with these 133 variants, we observe the highest number of variants (3) belonging to the *BRCA2, DUOX2, GJB2* and *LZTR1* genes. These are followed by several genes with 2 variants and the rest with just a single variant in each case. (Supplementary Table S10.3) .

**Table S10.3.** Genes associated with the 133 variants that have ≥1% allele frequency in any of the 83 populations and the number of such variants corresponding to the gene. Genes with >1 variants are mentioned.

| Gene | Number of Variants |
|---|---|
| *BRCA2* | 3 |
| *DUOX2* | 3 |
| *GJB2* | 3 |
| *LZTR1* | 3 |
| *BTD* | 2 |
| *HBB* | 2 |
| *HGD* | 2 |
| *FANCE* | 2 |
| *MOCOS* | 2 |
| *MSH3* | 2 |
| *MYBPC3* | 2 |
| *NPHP4* | 2 |
| *RPGRIP1* | 2 |
| *SBDS* | 2 |

#### Supplementary Section S11: Variants in medically relevant ACMG genes reveal population-level risks

Sauma Suvra Majumdar[1,2], Krithika Subramanian[1,3], Priyanka Singh[1], Bratati Kahali[1]

[1]Centre for Brain Research (CBR), IISc Campus, Bengaluru, India. [2]Interdisciplinary Mathematical Sciences, Indian Institute of Science (IMI- IISc), Bengaluru, India. [3]Manipal Academy of Higher Education (MAHE), Karnataka, India.

#### Summary

In this section, we analyzed the variants present in ACMG genes to understand the potential disease risk owing to these gene variants at the population level.

##### Methods

##### Carrier frequencies for variants in ACMG genes

We analyzed the variants in the ACMG v3.2 gene list of 94 clinically relevant genes. We used the exonic variants of these genes to calculate variant and gene carrier rates as follows:

$$VCR = \frac{(AC - Hom)}{(0.5 \times AN)}$$

Where AC is the alternate allele count for the variants, AN is the allele number. The gene carrier rates are calculated as follows:

$$GCR_g = 1 - \prod_{i=1}^{v}(1 - VCR_i)$$

Here, $VCR_i$ is the variant carrier rate for variant i, and v is the number of variants of interest in gene g. These calculations were performed separately for each population group. The GCR (gene carrier rate) values range from 0 to 1. A GCR close to 1 implies a high probability that an individual from the population carries at least one exonic variant for the gene, indicating high prevalence in the population, while a GCR close to 0 implies a low probability of being a carrier.

##### Results

For the whole GI dataset, we calculated variant densities for HIGH and MODERATE variants in ACMG genes. Variant density was calculated by normalizing the number of variants("missense", "frameshift", "stop_gained", "splice_donor", "splice_acceptor", "start_lost","stop_lost") per gene by the length of the gene (variants per kilobase), in which 5 of the top 10 genes were related to cardiac and lipid disorders (TableS11.). For further analysis of the variants in ACMG genes, we focused on the non-singleton loss-of-function (LoF) variants and putative deleterious missense mutations (pDMMs). We uncovered 81 (46 SNV and 25 INDELs) loss-of-function (LoF) variants distributed across 28 ACMG genes (Table S11.2), and 625 pDMMs that overlapped across 65 ACMG genes (Table S11.3). In these 81 LoFs, 13 were novel; and in the 625 pDMMs, 42 were novel (Table S11.4).

Several (17) of these genes that presented with LoF (*APOB, CASQ2, DSC2, DSG2, DSP, KCNQ1, LDLR, MYBPC3, MYH7, MYL2, PCSK9, PKP2, RYR1, RYR2, TNNT2, TRDN, TTN*) are known to be implicated in cardiac and lipid related disorders as per OMIM (www.omim.org ). The LoF variants in these genes also showed differences in allele frequencies with other South Asian populations like gnomAD-SAS, with the highest difference showed for a frameshift variant in *MSH6*- a gene known for Lynch syndrome . Other ACMG genes like *APOB,BTD, RPE65, BRCA2, LDLR* etc have LoF variants in the GI dataset that are absent in gnomAD SAS (figure S11.1). This makes them interesting candidates for future clinical studies, for example for ascertaining the true medical relevance in the populations, assessing the penetrance and pathogenicity from sufficient number of studies, in conjunction with appropriate clinical phenotyping. The *MSH6* variant, along with others showed population-level differences in frequencies in the GI dataset itself. *MSH6* and *TRDN* are the only ACMG genes that exhibited homozygous LoFs in the dataset (figure S11.2) .

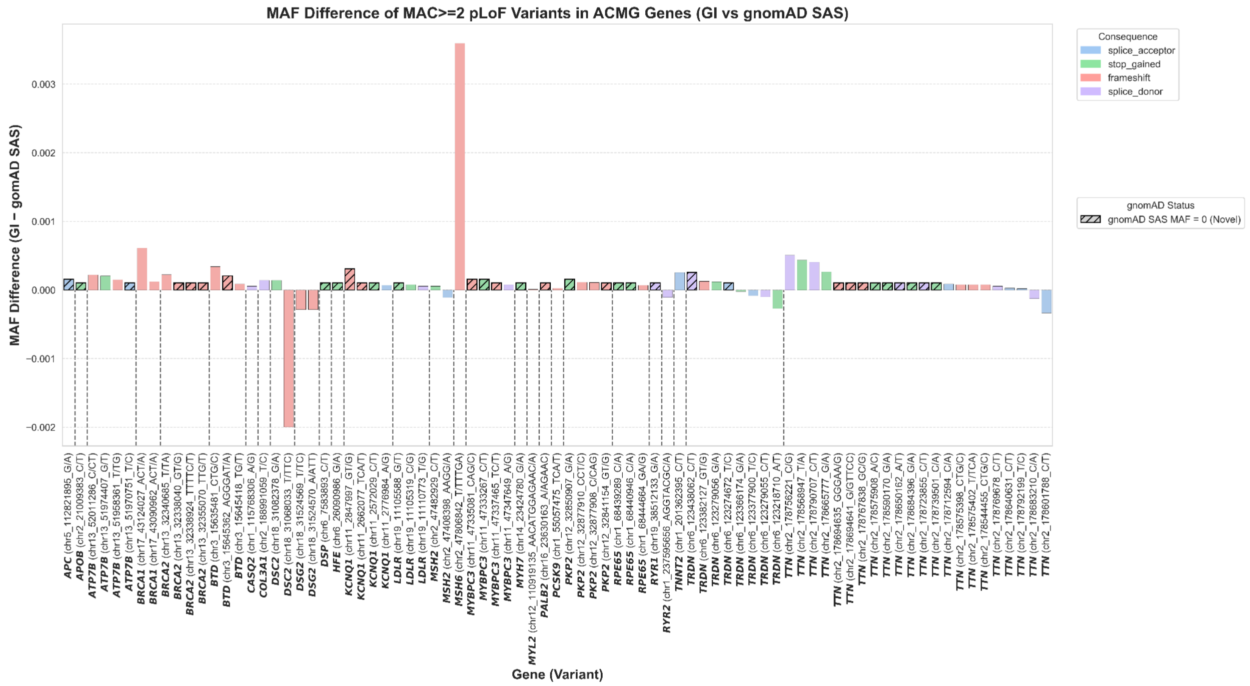

**Figure S11.1**. Plot showing frequency differences in LoF mutations in ACMG genes between gnomAD-SAS and the GenomeIndia dataset, grouped by different genes. A few of these variations are not reported in gnomAD SAS (marked with red borders on the bar) but are found in the GenomeIndia dataset.

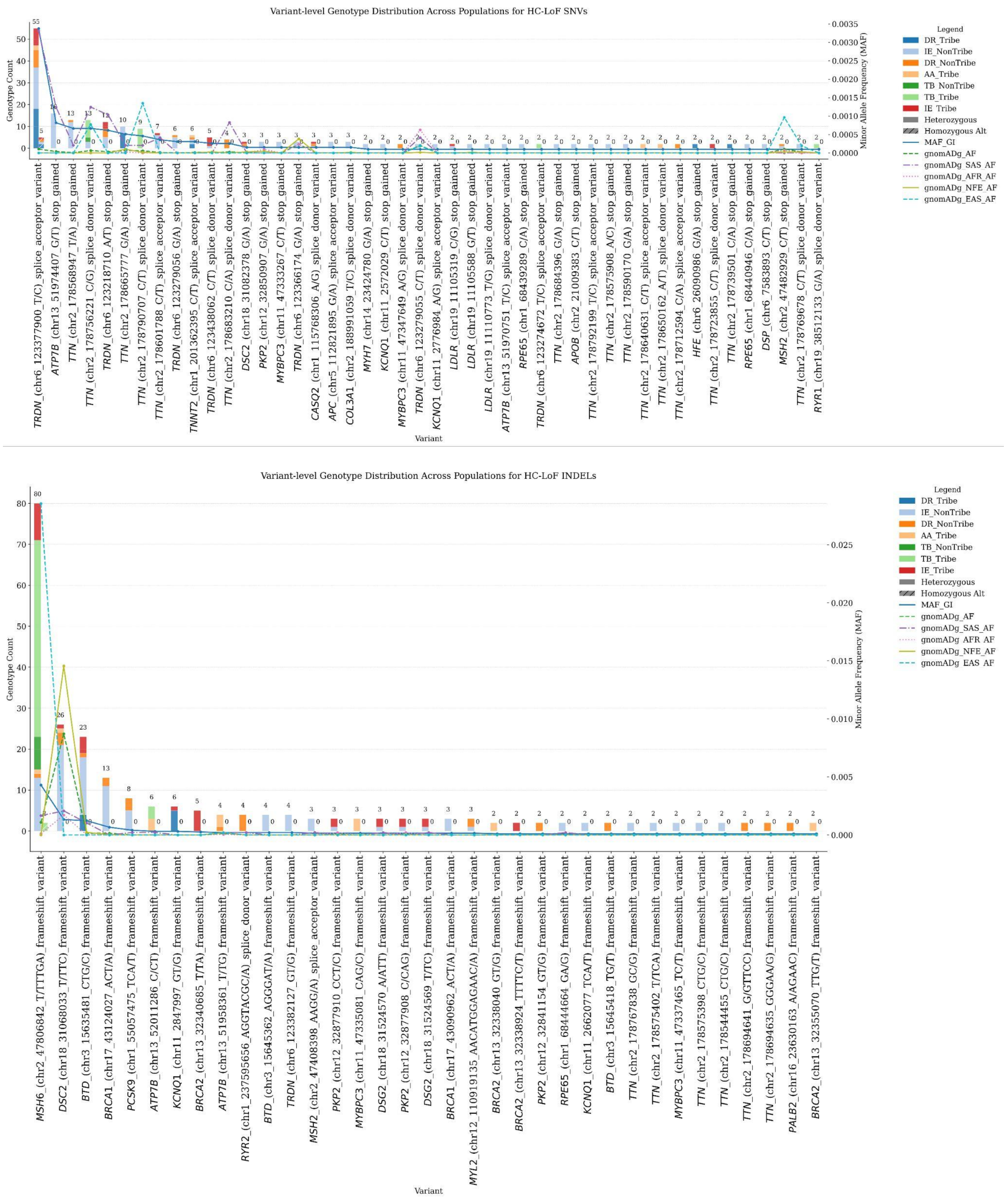

**Figure S11.2.** Distribution of heterozygous and homozygous counts of LoF variants in ACMG genes across different populations. The light-colored bars depict heterozygous counts, and the shaded grouped bars depict homozygous counts for that variant across the populations.

Overlapping with Clinvar, we found 21 LP/P variants from the pDMM category, with *LDLR* and *ATP7B* showing the largest share at 8 and 6 variants, respectively. For LoFs we found 23 variants overlapped with Clinvar with *ATP7B* and *BRCA2* being the highest at 4 each, followed by *TTN, LDLR* and *BTD,* each with 3 variants (Fig 4B, Table S11.2, Table S11.3).
Notable uncovered variant examples of ACMG genes: (Table S11.2, Table S11.3)

- The splice acceptor variant in the *TRDN* gene (rs578024729), associated with cardiomyopathy and previously noted as South Asian-specific in gnomAD, was found in 66 individuals, including five homozygotes. This variant demonstrates a wide distribution across most Indian linguistic groups (excluding TB tribe and nontribe), confirming a broad population risk.

- Furthermore, specific high-risk alleles show population stratification: Two of three P/LP/P variants in *LDLR* (rs121908029, rs774069731) were restricted to the IE_NT population. Four P/LP/P variants in the *ATP7B* gene (associated with Wilson's disease) (rs1954023351, rs137853287, rs2139975409, rs572147914) were also exclusively found in the IE_NT population.

- A *TTN* LoF likely pathogenic variant rs2530607571 associated with primary dilated cardiomyopathy and absent in gnomAD was found exclusively in the DR_Tribe population (DR_NGH_1_01).

- An *ATP7B* P/LP LoF variant (rs1954023351) was found exclusively in the tribal populations (AA_T and TB_T).

- A likely pathogenic *RPE65* missense variant rs61751277 associated with retinopathy, previously undiscovered in SAS and EAS populations in gnomAD, was found exclusively in the TB_Tribe populations.

- A SAS specific variant in gnomAD, rs181396238, which is LP/P in *BTD* gene associated with biotinidase deficiency, was exclusively found in 5 individuals in the DR_NonTribe population.

- Four P/LP/P *BRCA2* LoF variants were specific to the Tribal populations. Rs878853573 (AA_Tribe), rs1057517565 (IE_Tribe), rs876659617 (IE_Tribe), rs2137556981 (AA_Tribe)

The comprehensive sampling strategy employed by the GenomeIndia Project has been instrumental in elucidating these insights, underscoring its significant contribution to advancing genomic medicine within the South Asian population. We further calculated the gene carrier rate of ClinVar pathogenic or likely pathogenic variants of the above-mentioned ACMG genes. Genes like *ATP7B, BTD, LDLR, GAA and BRCA2* showed the highest burden, with at least one population exhibiting a carrier rate of 0.5% in the dataset, with *BRCA2* pathogenic or likely pathogenic variants limited to tribal populations (figure S11.3).

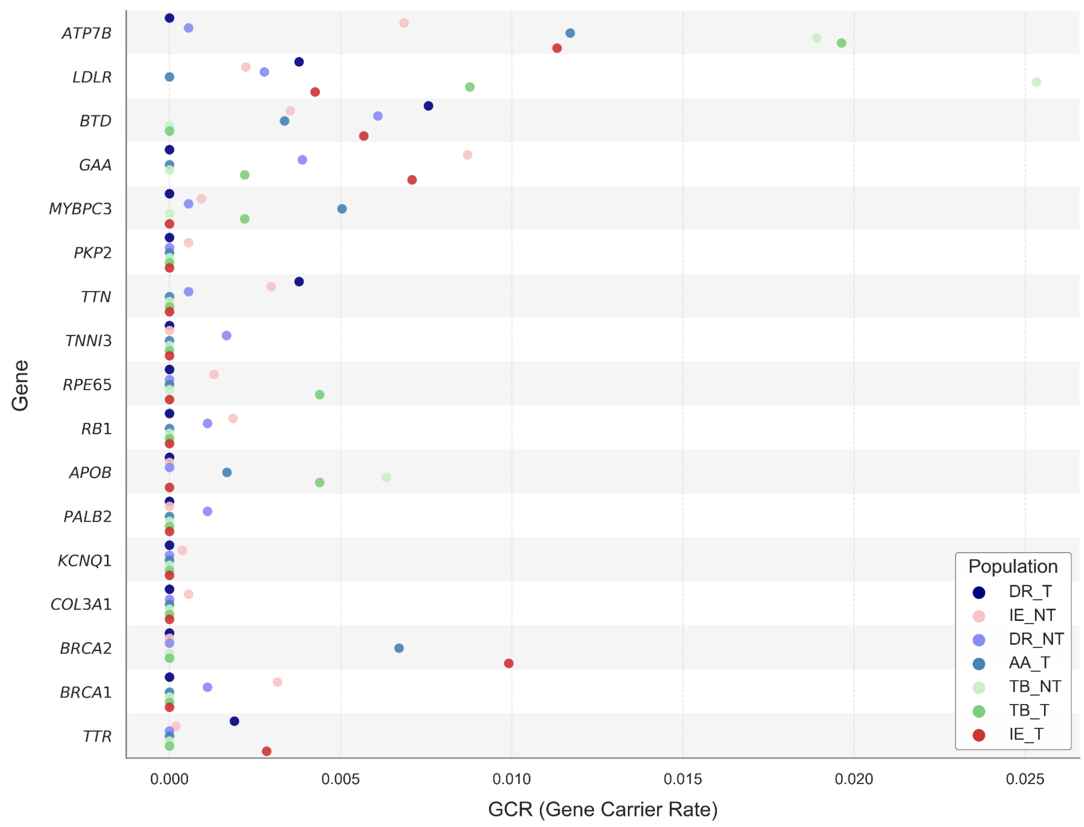

**Figure S11.3.** Figure showing the carrier rates of pathogenic alleles in ACMG genes per population.

**Table S11.4.** List of novel deleterious missense and LoF variants in ACMG genes:

| Chrom | Pos | Gene | Ref | Alt | Consequence | Populations |
|---|---|---|---|---|---|---|
| chr1 | 68444828 | *RPE65* | T | C | missense_variant | AA_EPL_1_04=0.90% \| IE_NCH_1_01=0.66% |
| chr1 | 68446851 | *RPE65* | G | C | missense_variant | IE_WHR_2_04=0.70% |
| chr1 | 201075534 | *CACNA1S* | C | A | missense_variant | DR_NGH_1_02=2.03% |
| chr1 | 237500864 | *RYR2* | G | T | missense_variant | IE_NRP_2_06=0.62% |
| chr1 | 237674817 | *RYR2* | A | C | missense_variant | DR_ECP_2_07=0.79% \| DR_SDN_2_03=0.85% |
| chr1 | 237707053 | *RYR2* | A | C | missense_variant | IE_BPV_2_01=0.44% \| IE_ERP_2_04=0.31% |
| chr10 | 86923456 | *BMPR1A* | T | C | missense_variant | DR_ECP_2_02=0.61% |

| | | | | | | |
|---|---|---|---|---|---|---|
| chr10 | 119677009 | *BAG3* | G | C | missense_variant | DR_WGH_1_02=1.35% |
| chr13 | 48381388 | *RB1* | T | G | missense_variant | IE_WCP_2_05=1.43% |
| chr13 | 48465262 | *RB1* | T | C | missense_variant | IE_NRP_2_08=0.32% \| IE_WPL_2_03=0.51% |
| chr13 | 51946314 | *ATP7B* | C | G | missense_variant | IE_WHR_2_01=0.56% |
| chr15 | 48430703 | *FBN1* | T | C | missense_variant | IE_NDN_2_02=0.31% \| IE_WHR_2_02=0.31% |
| chr15 | 48495565 | *FBN1* | G | A | missense_variant | DR_ECP_2_03=1.01% |
| chr15 | 48520736 | *FBN1* | T | G | missense_variant | IE_NCH_2_01=0.79% |
| chr15 | 48534204 | *FBN1* | A | C | missense_variant,splice_region_variant | DR_WCP_2_01=0.62% \| DR_WCP_2_03=0.30% |
| chr16 | 15747624 | *MYH11* | A | G | missense_variant | IE_WPL_2_02=1.61% |
| chr18 | 31074778 | *DSC2* | T | A | missense_variant | IE_WHR_1_01=1.33% |
| chr18 | 31086724 | *DSC2* | A | C | missense_variant | TB_BPV_1_01=1.19% |
| chr18 | 31521166 | *DSG2* | T | C | missense_variant | AA_EPL_1_02=1.67% |
| chr19 | 38451824 | *RYR1* | G | A | missense_variant | TB_WHR_1_02=1.79% |
| chr19 | 38466203 | *RYR1* | T | G | missense_variant | DR_ECP_2_06=0.61% |
| chr19 | 38504776 | *RYR1* | C | T | missense_variant | IE_NRP_2_03=0.63% |
| chr19 | 38537916 | *RYR1* | A | G | missense_variant | DR_ECP_2_03=0.34% \| IE_ERP_2_02=0.27% |
| chr2 | 47476398 | *MSH2* | T | G | missense_variant | IE_BPV_2_01=0.44% \| IE_WPL_2_03=0.51% |
| chr2 | 178539451 | *TTN* | C | T | missense_variant | DR_WCP_2_01=1.85% |
| chr2 | 178539573 | *TTN* | T | C | missense_variant | DR_NGH_1_02=2.70% |
| chr2 | 178552017 | *TTN* | A | G | missense_variant | IE_NCH_1_03=4.67% |
| chr2 | 178553204 | *TTN* | A | G | missense_variant | IE_NRP_2_10=0.61% |
| chr2 | 178564131 | *TTN* | A | G | missense_variant | AA_EPL_1_04=0.60% |
| chr2 | 178618634 | *TTN* | A | G | missense_variant | IE_NCH_1_01=0.66% \| IE_NCH_1_02=0.75% |
| chr2 | 178722727 | *TTN* | C | T | missense_variant | IE_NRP_2_07=0.65% |
| chr2 | 178734937 | *TTN* | C | G | missense_variant | IE_NCH_1_03=2.00% |
| chr2 | 178770622 | *TTN* | C | A | missense_variant | AA_EPL_1_03=2.17% \| AA_EPL_1_04=0.90% \| |

| | | | | | | |
|---|---|---|---|---|---|---|
| | | | | | | AA_SCH_1_01=0.77% \| IE_NRP_2_06=0.31% |
| chr2 | 188999885 | *COL3A1* | A | T | missense_variant | DR_WCP_2_01=0.62% |
| chr2 | 189010198 | *COL3A1* | A | G | missense_variant | IE_NRP_2_02=0.31% \| IE_NRP_2_13=0.26% |
| chr3 | 38597777 | *SCN5A* | G | C | missense_variant | IE_NDN_2_02=0.61% |
| chr3 | 52452202 | *TNNC1* | T | A | missense_variant | IE_NRP_2_10=0.61% |
| chr6 | 7584059 | *DSP* | A | G | missense_variant | AA_EPL_1_01=1.11% |
| chr7 | 128837497 | *FLNC* | C | T | missense_variant | TB_NER_1_01=0.75% |
| chr7 | 128844851 | *FLNC* | A | T | missense_variant | AA_EPL_1_03=1.09% |
| chr7 | 128847711 | *FLNC* | G | C | missense_variant | DR_SDN_2_03=1.70% |
| chr7 | 151565783 | *PRKAG2* | T | A | missense_variant | AA_EPL_1_01=1.11% |
| chr1 | 68439289 | *RPE65* | C | A | LoF | IE_ECP_2_01=0.71% |
| chr1 | 68440946 | *RPE65* | C | A | LoF | IE_NRP_2_10=0.61% |
| chr14 | 23424780 | *MYH7* | G | A | LoF | IE_WCP_2_04=0.71% |
| chr19 | 38512133 | *RYR1* | G | A | LoF | TB_BPV_1_02=0.97% |
| chr2 | 21009383 | *APOB* | C | T | LoF | IE_ECP_2_01=0.71% |
| chr2 | 178575908 | *TTN* | A | C | LoF | IE_NRP_2_05=0.68% |
| chr2 | 178684396 | *TTN* | G | A | LoF | IE_NRP_2_06=0.31% \| IE_NRP_2_07=0.32% |
| chr2 | 178694635 | *TTN* | GGGAA | G | LoF | DR_ECP_2_01=1.79% |
| chr2 | 178694641 | *TTN* | G | GTTCC | LoF | DR_ECP_2_01=1.79% |
| chr2 | 178767838 | *TTN* | GC | G | LoF | IE_WCP_2_02=0.62% |
| chr6 | 7583893 | *DSP* | C | T | LoF | IE_WCP_2_02=0.62% |
| chr6 | 26090986 | *HFE* | G | A | LoF | DR_EPL_1_03=1.27% |
| chr6 | 123274672 | *TRDN* | T | C | LoF | TB_NER_1_01=0.75% |

#### Supplementary Section S12: Homozygous Loss-of-Function variants

Priyanka Singh[1], Sauma Suvra Majumdar[1,2], Bratati Kahali[1]

[1]Centre for Brain Research (CBR), IISc Campus, Bengaluru, India. [2]Interdisciplinary Mathematical Sciences, Indian Institute of Science (IMI- IISc), Bengaluru, India.

#### Summary

In this section, we present a comprehensive catalog of homozygous loss-of-function (LoF) variants observed in the GenomeIndia cohort and identify the individuals carrying them. We describe the distribution of these homozygous variants across genes and individuals carrying them across populations; highlight variants that are novel or enriched in specific endogamous populations; and assess their clinical and therapeutic relevance using established knowledgebases. Together, these analyses reveal population-specific homozygous LoF landscapes and form the basis for future studies into gene function and disease biology in diverse Indian ancestries.

##### Methods

We identified high-confidence loss-of-function (LoF) variants occurring in the homozygous state in at least one individual. These homozygous LoF variants were further classified into (i) novel variants and (ii) population-specific variants. Clinically relevant variants were identified using ClinVar (https://www.ncbi.nlm.nih.gov/clinvar/), retaining those labeled as pathogenic or pathogenic/likely pathogenic with a ≥2-star review status (downloaded February, 2025). Additionally, we examined overlap with known trait-associated variants from the GWAS Catalog (https://www.ebi.ac.uk/gwas/). Importance of the homozygous variants was also assessed by checking if they are present in essential developmental genes based on lethality in homozygous knockout mouse models, as reported by the International Mouse Phenotyping Consortium (IMPC) https://www.mousephenotype.org/. To assess the pharmacological relevance of genes harboring homozygous LoF variants, we retrieved all drug target genes listed in the Drug Repurposing Hub (https://repo-hub.broadinstitute.org/repurposing#home), encompassing preclinical, clinical, and launched stages, and identified overlaps with the documented homozygous LoF genes.

##### Results

A central goal of biomedical research is to characterize the function of every human gene. Traditionally, geneticists studied gene function by inactivating, or “knocking out,” genes in model organisms such as mice and inferring human gene roles from observed physiological or behavioral changes. However, results from animal models do not always reflect human biology. Interestingly, naturally occurring LoF variants, which disrupt gene function, when occurring in a homozygous state, often referred to as human knockouts (Karczewski et al., 2020; Saleheen et al., 2017; Sun et al., 2024; Wall et al., 2019), serve as powerful natural models to directly investigate gene function, essentiality, and their impact on human health and disease. Accordingly, here we present a comprehensive catalogue of homozygous HC-LoF variants, along with their corresponding genes and the distribution across population clusters of individuals carrying these variants within the GI dataset.

Focusing on homozygous HC-LoF variants, we identified 1,214 such variants in 965 genes, out of a total of 15,849 non-singleton HC-LoF variants in our dataset (Figure S12.1a). The alternate allele frequency (AAF) distribution of homozygous HC-LoF variants revealed a wide spectrum, from ultra rare to common alleles. Notably, for 33 variants, the allele was a major allele (AAF above 50%), and unlikely to be deleterious at such high frequency in our dataset; accordingly, they were not considered for further analysis. Among the remaining 1,181 variants spanning 940 protein-coding genes, the majority were frameshift (n = 502), followed by stop-gained (n = 399), splice donor (n = 163), and splice acceptor variants (n = 117) (Figure S12.1b and Supplementary Table S12.1).

**a.** **b.**

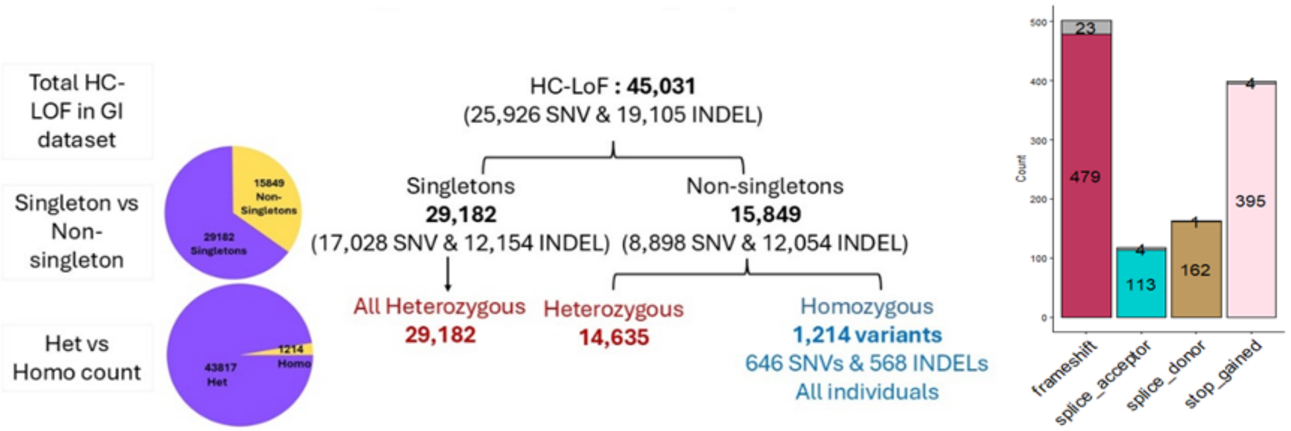

**Figure S12.1.** a) Flowchart summarizing the distribution of high-confidence loss-of-function (HC-LoF) variants identified in GI dataset. b) Distribution of 1181 homozygous HC-LoF based on consequence. Stacked bar representing counts of known (colored) and novel (grey) variants.

***Higher number of novel homozygous LOFs in tribal populations***

Out of the 1,181 homozygous HC-LoF variants, we identified several LoFs with potential functional or clinical significance that warrant further investigation (Figure S12.2). Among these, 32 novel variants were found in the GI dataset, predominantly enriched in tribal groups, particularly the DR_T, reflecting genetic distinctiveness shaped by founder effects, endogamy and long-term isolation (Table S12.1, column R).

Notably, a novel homozygous frameshift variant (chr16:28497670-C>CT) in *APOBR*, a receptor critical for dietary lipid uptake and the transport of triglyceride-rich lipoproteins, was identified in three individuals from the DR_T (DR_NGH_1_01). *APOBR* functions independently of *APOE*, and mutations in this gene have been implicated in lipid metabolism disorders. Phenotypic data revealed that while two female homozygotes (aged 35 and 36) had moderate triglyceride levels (131 and 140 mg/dL), a male homozygote (aged 25) exhibited marked hypertriglyceridemia (387 mg/dL), suggesting that studying this variant further in these populations may help understand the contribution to dysregulated lipid profiles. These findings underscore how isolated, highly endogamous populations can serve as natural genetic laboratories for studying genes harboring homozygous LoF variants, and facilitate future study targets for understanding the functional consequences of homozygous occurrences, thereby uncovering variants with direct clinical relevance that are largely absent from broader global datasets.

***Homozygous LoFs absent in gnomAD***

Additionally, comparison with gnomAD, one of the largest and widely used population-scale genomic resources, uncovers 192 variants in our dataset observed in the homozygous state for the first time, representing homozygous individuals not previously reported in any population within gnomAD (Supplementary Table S12.1, column S).

Notably, a variant rs941571543-C>T in the *GUF1* gene was detected in 31 heterozygous and 2 homozygous individuals, all from the DR_T (DR_NGH_1_02), whereas no heterozygotes or homozygotes for this variant are reported in gnomAD, highlighting the value of underrepresented endogamous populations in uncovering novel homozygous occurrences.

***Population Specific LOFs***

Furthermore, we identified 164 variants that occur exclusively in one of the eight population clusters, with both heterozygous and homozygous individuals confined to that group (Main Fig.4c and Supplementary Table S12.1, column T). Among these 164 cluster-specific variants, 123 are further restricted to a single population when assessed across the 82 populations (Supplementary Table S12.2), reflecting the impact of endogamous practices leading to larger instances of homozygous variants in individuals in certain populations. Given their restricted distribution, systematic screening of homozygous individuals from these populations can enable investigations of variant impact and penetrance, offering insights into human gene tolerance and population-level disease risk.

***Tribe vs Non-Tribe burden***

The distribution of total homozygous LoF variants per individual was compared between tribe and non-tribe populations using the one-sided Kolmogorov–Smirnov test, revealing a significant difference ($p < 0.0001$, performed in Rv4.4.2), with tribal individuals exhibiting a higher burden per individual than their non-tribal counterparts.

***Homozygous individuals in genes lethal in mice***

To further assess functional relevance, we explored the International Mouse Phenotyping Consortium (IMPC) database, a global initiative aimed at systematically generating and phenotyping knockout mice for every protein-coding gene in the mouse genome, providing a comprehensive resource for understanding gene function and their essentiality. Leveraging this resource, we identified over 50 genes including *LIFR, HTT, FOCAD* etc, whose homozygous knockout is embryonically lethal in mice yet humans carrying homozygous LoFs variants in these same genes are viable, highlighting a unique set of likely tolerated homozygous individuals that may inform functional and clinical studies (Supplementary Table S12.1, column V), with appropriate clinical and diagnostic phenotyping. For example, we identified a splice-donor variant in *FOCAD* (rs754077384-G>GT) in 8 heterozygotes and 1 homozygote, all exclusively present in IE_NT group (IE_NCH_2_01 (~2.4%) & IE_WCP_2_05 (~3%)). This South Asian–specific variant has no previously reported homozygotes in gnomAD. *FOCAD* is involved in focal adhesions, microtubule dynamics, and cell cycle regulation, and is associated with severe congenital liver disease (www.omim.org/ ) and pediatric liver cirrhosis (Shao, 2022).

Notably, homozygous LoF variants in 23 of these genes are reported in humans for the first time in this study when compared with existing large-scale human knockout datasets (Karczewski et al., 2020; Saleheen et al., 2017; Wall et al., 2019; Sun et al., 2024). These include *LIFR* (rs773896661), which is associated with Stüve–Wiedemann syndrome (OMIM: 601559); *DHX38* (rs753918448), associated with retinitis pigmentosa (OMIM: 618220) and specifically observed in the DR_NGH_1_01 population (~13%), again highlighting the utility of studying underrepresented endogamous communities where seemingly harmful variants could be found in homozygous states. The viability of these homozygous individuals opens new avenues for studying such naturally occurring putative human knockouts to investigate the extent of penetrance, mechanisms of tolerance and functional impact of these homozygous LoFs on gene function.

##### *LoFs in GI linked to GWAS findings*

We identified 49 homozygous LoF variants previously reported in the GWAS Catalog (Supplementary Table S12.1, column Y). These include rs114285050-G>A in *GPR151*, which has been associated with reduced BMI and lower risk of type 2 diabetes. Another noteworthy gene, *LPA*, encodes lipoprotein(a) [Lp(a)], elevated levels of which are a recognized marker of cardiovascular risk (Small et al., 2024). In our dataset, we identified six homozygous LoF variants in *LPA* (rs143431368-T>C, rs41272114-C>T, rs200154828-G>A, rs199536939-C>G, rs753812356-C>CA, and rs559580002-C>T). Among these, the splice-acceptor variant rs143431368-T>C has been reported in the GWAS Catalog to lower Lp(a) levels (Lim et al., 2014) and reduce coronary heart disease (CHD) risk. Notably, no homozygotes for this variant have been reported in the South Asian population in gnomAD; however, we observed 12 carriers, including two homozygous individuals from the DR_T population. Another variant, rs41272114-C>T, though not listed in the GWAS Catalog, has been previously linked to decreased Lp(a) and coronary artery disease (CAD) risk (Kyriakou et al., 2014). This variant shows a MAF of 0.026 in our dataset, with 489 heterozygotes and 10 homozygous individuals, predominantly from the IE and AA clusters. Overall, these findings highlight the presence of functionally relevant *LPA* LoF variants that may exert a protective effect against cardiovascular disease risk, that can only be confirmed with further functional, clinical and structural studies. Next, a stop-gained variant (rs17602729-G>A) in *AMPD1*, was observed in 477 heterozygotes and 13 homozygotes. While the alternate allele has been associated with increased glomerular filtration rate and poorer kidney function in humans, complete knockout of *AMPD1* is lethal in mice (Figure S12.2). Furthermore, a variant reported in the African population, *A2ML1* (rs143864957-AAG>A), which has been significantly associated with lower expression of the *A2ML1* gene in patients with chronic kidney disease and hypertension, was found exclusively in the CAO group in the GI dataset, which is an African admixed population in India (Figure S12.2).

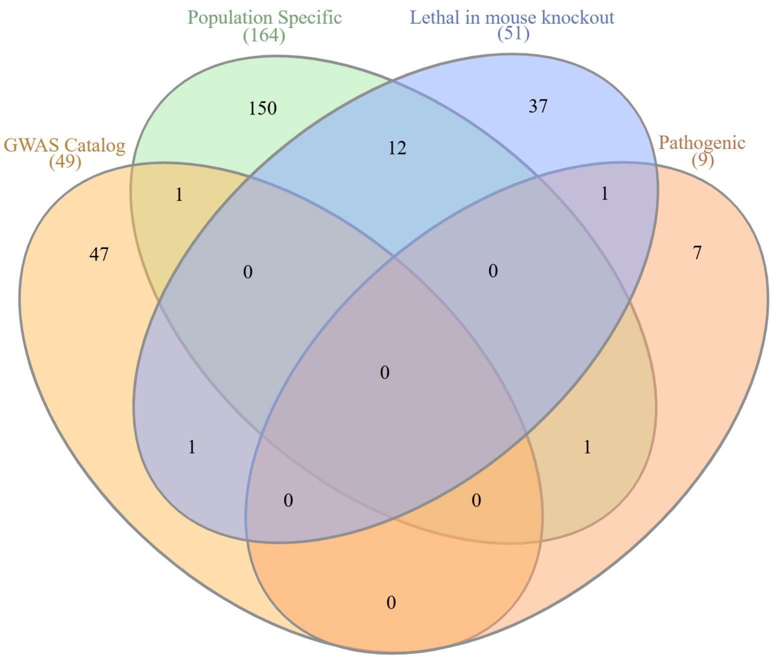

**Figure S12.2**. Overlap of LoF variants across four categories: population-specific, lethal in mouse knockout, pathogenic, and GWAS Catalog variants. The numbers indicate the count of variants shared or unique to each category.

##### *Pharmacological relevance*

To investigate their pharmacological relevance, we compared our gene list (N=940) with known drug targets from the Drug Repurposing Hub. This analysis identified 77 genes (~8.2%) that overlapped with known drug targets across preclinical, clinical, and launched stages, suggesting that a subset of homozygous LoF variants carried by individuals may represent pharmacologically relevant candidates.

##### *Frequency distribution of Homozygous LoFs*

Of the 1,181 homozygous HC-LoF variants, the majority (N = 817) were rare (AF <1%), spanning 686 protein-coding genes in 993 individuals (Supplementary Table S12.1, column Q). The distribution of these individuals across population clusters is provided in Table S12.3. Besides, we performed an observed-to-expected analysis of homozygous genotypes across 15,437 annotated rare HC-LoFs. This analysis provided insight into the representation and distribution of homozygous genotypes in the dataset. We identified 817 rare LoF variants with at least one homozygous genotype, compared to an expected average of 928.2 allele frequency–matched synonymous variants with at least one homozygote, representing a 12% deficit of homozygous LoF genotypes. For the neutral expectation, each rare LoF variant was matched to a synonymous variant of similar allele frequency, which is unlikely to be under strong selective pressure. This depletion indicates that a fraction of homozygous LoF genotypes are absent from our cohort, could be due to early lethality or severe disease, and are therefore not observed in healthy adults.

**Table S12.3.** Distribution of 993 individuals carrying rare LoF variants across populations

| Linguistic group | Tribal | Non-Tribal | Total number of individuals in each population |
|---|---|---|---|
| IE | 75 | 338 | 413 |
| DR | 133 | 202 | 335 |
| TB | 97 | 34 | 131 |
| AA | 84 | - | 84 |
| CAO | - | - | 30 |
| Total | 389 | 574 | 993 |

***Frequency-based patterns across populations***

***i) rare across populations***

Examining the frequency spectra of these rare variants across all 82 populations (excluding CAO), revealed that only 78 variants, when present, remained consistently rare (Supplementary Table S12.4). These rare homozygous LoFs likely arose from independent mutational or founder events and are expected to be maintained at low frequencies, given their deleterious consequences. Such variants illustrate the value of studying diverse populations, as each of them contribute distinct homozygous LoF occurrences that may not be captured in larger but more homogeneous datasets.

***ii) impact of fine-scale population structure***

The majority of the remaining variants (N = 709) display a more gradual frequency spectrum, being common in some populations yet rare in others, or occurring as homozygous in one population but only as heterozygous in others. A subset of population-specific variants that are rare in the overall dataset reached higher frequencies (AF ≥ 1%) in particular populations. These variants therefore represent globally rare but locally enriched LoF variants shaped by endogamy, founder effects, local selection, and population-specific demographic histories (Supplementary Table S12.5).

For instance, two LoF variants in *RETSAT* (rs774169688-G>GGT and rs759186033-CAG>C) (Supplementary Table S12.5) are present in hom-LoF condition. *RETSAT* encodes retinol saturase, an enzyme involved in lipid metabolism, with previous studies linking its expression to hepatic fat, triglyceride levels, and glycemic control (Pang et al., 2017). These variants were observed in 174 heterozygous individuals and 8 homozygous individuals. Notably, all homozygous carriers were exclusively found in tribal populations (AA_EPL_1_02 (2), TB_WHR_1_02 (1), AA_NER_1_01 (1), DR_EPL_1_02 (1), IE_EPL_1_01 (1), TB_WHR_1_01 (1), TB_NER_1_01 (1)). The allele frequency of these variants reached up to 10% in tribal populations, likely reflecting strong endogamy and founder effects, resulting in locally higher frequencies of these variants, which may contribute to differential susceptibilities to diseases.

Similarly, a splice-site variant in *IFIH1* (rs35732034-C>T; c.2807+1C>T), was observed in 159 heterozygotes and 2 homozygotes. The two homozygous carriers belonged to DR_ECP_2_03 (1 individual; AF ≈ 2%) and AA_NER_1_01 (1 individual; AF ≈ 10%). Heterozygotes are previously found to be associated with early-onset inflammatory bowel disease, while homozygotes show extreme susceptibility to respiratory RNA viruses (Asgari et al., 2017). Similarly, rs754285553 (G>GCT), a frameshift variant in *ZFHX3*, gene in which LoF variants have been reported to cause syndromic intellectual disability (Pérez Baca et al., 2024), was observed in 106 individuals, including 2 homozygotes. The two homozygotes belonged to tribal population i.e. IE_CHR_1_01 (1 individual) and TB_BPV_1_02 (1 individual). Also, there can be LoF variants that are homozygous in one founder group, and may persist elsewhere only as heterozygous carriers, reflecting differences in genetic drift, bottlenecks, or historical mating patterns. For example, rs4986893 (G>A) in the *CYP2C19* gene was identified in 134 carriers, the majority of whom were heterozygous, with only one homozygous individual observed (TB_BPV_1_01; AF ≈ 11%). In gnomAD, homozygotes are reported exclusively in East Asians, and this represents the first report of homozygosity in the South Asian population. The alternate allele “A” is pharmacogenomically important, as it has been associated with an increased risk of adverse outcomes when treated with clopidogrel in acute coronary syndrome. It has also been linked to risk for major depressive disorder and bipolar disorder in the Han Chinese population (Zhang et al., 2022). Discovering such variants from the Indian population has been made feasible by the sampling of diverse ethnolinguistic groups in the GenomeIndia study.

**Concluding Insights**

The unique population characteristics and demographic histories of the populations sampled in GenomeIndia provide an unprecedented opportunity to identify individuals with homozygous LoF and heterozygous carriers for the same variants at appreciable frequencies. Nevertheless, interpretation of their deleteriousness must be made with caution because the enrichment of homozygous LoFs in isolated (primarily tribal) populations may reflect demographic history rather than neutral tolerance. Our findings reveal that both group-specific and shared homozygous LoF variants contribute uniquely to the landscape of homozygous occurrences in the population. It further illustrates how the definition of “rare” is highly context-dependent, variants that are rare in the overall GI dataset may not remain rare when examined within individual population clusters reflecting the strong influence of endogamy, drift and population structure in the Indian subcontinent. Such cross-population contrasts are particularly informative for understanding gene essentiality and resilience. The differences in homozygous LoF tolerance across populations might be influenced by functional constraints or other biological factors that need to be investigated further.

By integrating data from multiple ancestrally diverse populations, we thus capture a more comprehensive view of human gene loss, revealing a continuum from private/rare to widely distributed LoF alleles. This catalog of homozygous LoF variants provides a foundation for future research that can be integrated with functional and clinical perspectives, enabling linkage of gene disruption with phenotype, resilience, and disease risk, and for informing precision medicine approaches in underrepresented ancestries.

#### Supplementary Section S13: Landscape of pharmacogenomic variations in GI populations

Ankit Mukherjee[1], Chandrika Bhattacharyya[2], Mohammed Faruq[1,3]

[1]CSIR - Institute of Genomics & Integrative Biology (CSIR-IGIB), New Delhi, India. [2]BRIC - National Institute of Biomedical Genomics (BRIC-NIBMG), Kolkata, India. [3]Academy of Scientific and Innovative Research, Ghaziabad, India.

#### Summary

We curated the risk allele frequencies of known pharmacogenomic variations in the GenomeIndia (GI) dataset across 82 distinct Indian populations, broadly categorized into seven ethnolinguistic identities, as described earlier. To investigate how GenomeIndia has captured the complex genetic diversity of the Indian population, we examined the variants against the South Asian (SAS) superpopulation from the 1000 Genomes Project (Auton et al., 2015), which includes multiple South Asian subgroups and serves as a reference.

##### Methods

The GenomeIndia initiative analyzed whole genome sequencing (WGS) data from 9768 individuals across 82 Indian populations, representing four major ethno-linguistic groups: Austro-Asiatic (AA), Dravidian (DR), Indo-European (IE), and Tibeto-Burman (TB). To characterize the pharmacogenomic landscape of Indian populations, we integrated curated variant and annotation data from three major PGx resources: PharmGKB (Gong et al., 2021), PharmVar (Gaedigk et al., 2020), and the Clinical Pharmacogenomics Implementation Consortium (CPIC) (Relling et al., 2020).

Variant processing involved normalization using vt (Tan et al., 2015), annotating with dbSNP v156 using SnpSift v5.2 (Cingolani et al., 2012), and extracting PGx-relevant SNPs and indels using bcftools v1.9 (Danecek et al., 2021). Variants were converted into PLINK v1.9 (Purcell et al., 2007) format to enable population-specific allele frequency estimation and genotype analysis. Genotype counts and risk-allele distributions were analyzed to identify populations within India which are potentially at increased pharmacogenetic risk relative to other groups. Risk-allele frequencies were also compared with global populations from the 1000 Genomes Project, especially the South Asian (SAS) population, and Z-score normalization was applied to highlight populations exhibiting higher risk-allele frequencies for specific drugs relative to other GenomeIndia populations and global references.

Haplotype (star allele) analysis was performed on 58 pharmacogenes using Stargazer (S.-B. Lee et al., 2019), while CYP2D6 diplotypes were evaluated using Cyrius (Chen et al., 2021) due to the gene's complex structural variability. For the highly polymorphic HLA region, four-digit typing was carried out using xHLA (Xie et al., 2017), which allows accurate allele resolution from WGS data. Allele frequencies of star alleles were computed, and metabolizer phenotypes (poor, intermediate, normal, and ultrarapid) were assigned based on activity scores derived from diplotypes. GenomeIndia haplotyped star alleles were stratified by linguistic and tribal affiliations to assess interpopulation variability in drug metabolism and pharmacokinetic response.

#### Results and Discussion

##### Variant Curation

We integrated pharmacogenomic variant information from PharmGKB and PharmVar v6.2, along with drug-specific clinical guidelines and testing levels from CPIC and PharmGKB. Of the 4245 pharmacogenomic variants cataloged by PharmGKB and PharmVar v6.2 , 2831 were found in the GI dataset. Among these, 2339 have associated drug annotations. These 2339 variants comprise 2099 SNPs and 17 indels from 991 genes, as well as 223 SNPs located outside of any known gene. The occurrence of PharmGKB variants in GI is as follows: 29% (33/114) of level 1, 89% (32/36) of level 2, 84.3% (2240/2658) of level 3, and 92.6% (150/162) of level 4 variants. Notably, 287 pharmacogenomic variants were identified in 31 of the 34 Very Important Pharmacogenes (VIP) reported by PharmGKB across different levels of evidence. Furthermore, among these 31 VIP genes, variants for 29 are present in more than two individuals **(Table S13.1)**. In addition, we identified 317 star alleles across 55 pharmacogenes out of the 58 genes genotyped using Stargazer, and 86 CYP2D6 haplotypes inferred using Cyrius (**Table S13.2)**.

##### Prioritizing actionable pharmacogenomic variants and star allele haplotypes

To identify pharmacogenomic variants crucial to GI populations, we prioritized variants based on PharmGKB evidence levels and allele frequency differences relative to the South Asian (SAS) population. For PharmGKB Level 1 and Level 2 variants, those with an allele frequency in at least one GenomeIndia (GI) population exceeding that observed in SAS were selected. For PharmGKB Level 3 variants, variants exhibiting an overall allele frequency difference of at least 5% compared to SAS were included. In addition, variants annotated with a significant testing level according to PharmGKB drug label annotations and CPIC guidelines were also added. Applying these criteria to the 2,339 drug-associated variants, we prioritized a subset of 49 pharmacogenomic variants **(Table S13.3, Figure S13.1)**.

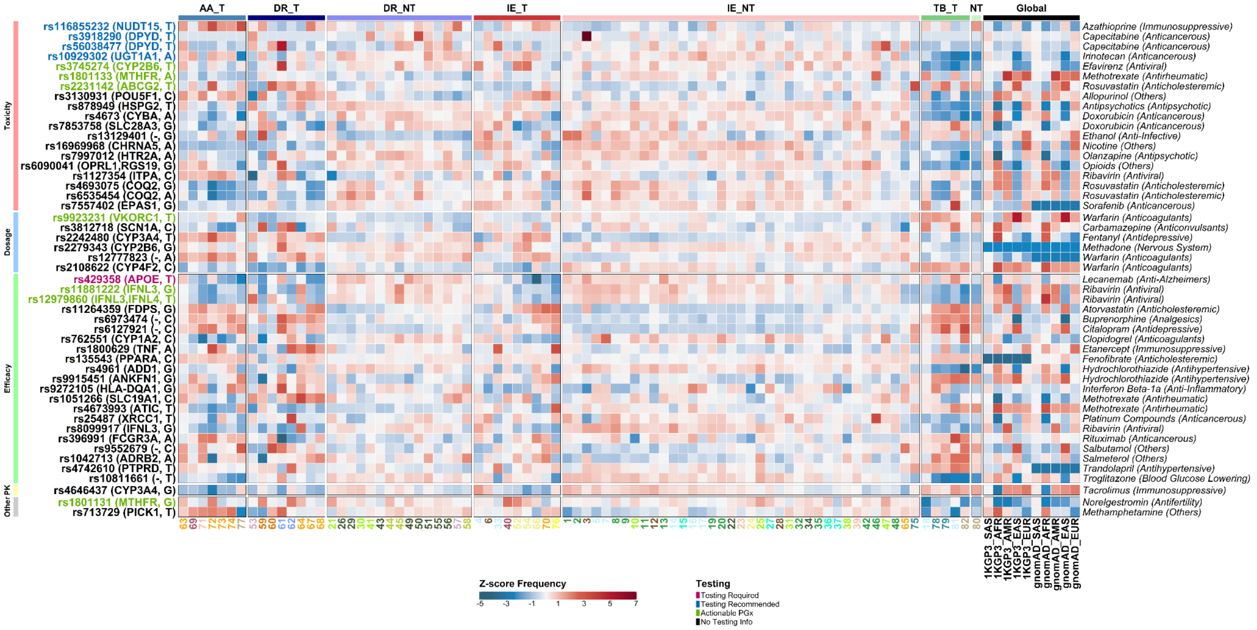

**Fig. S13.1.** Allele frequency comparison between pharmacogenomic variants across 82 Indian subpopulations and the global populations (1000 Genome and gnomAD). The Indian populations are clustered on the basis of tribal status and linguistics, while the variants have been annotated by CPIC recommendations associated with drug kinetics.

Using these prioritized variants, risk genotype frequencies were calculated and subsequently aggregated at the drug-population level. For each variant, genotype frequencies were computed based on the risk allele. Variant-level risk genotype frequencies were then summed across all variants associated with a given drug, stratified by population and drug response category (e.g., toxicity, dosage, efficacy), and the aggregated risk burden was normalized by the number of unique variants associated with each drug. This normalized metric represents the average contribution of risk-associated genotypes per variant for a given drug within a population. Overall, this approach yielded 4,018 drug-drug response-population-variant interactions **(Table S13.4, Figure S13.2)**.

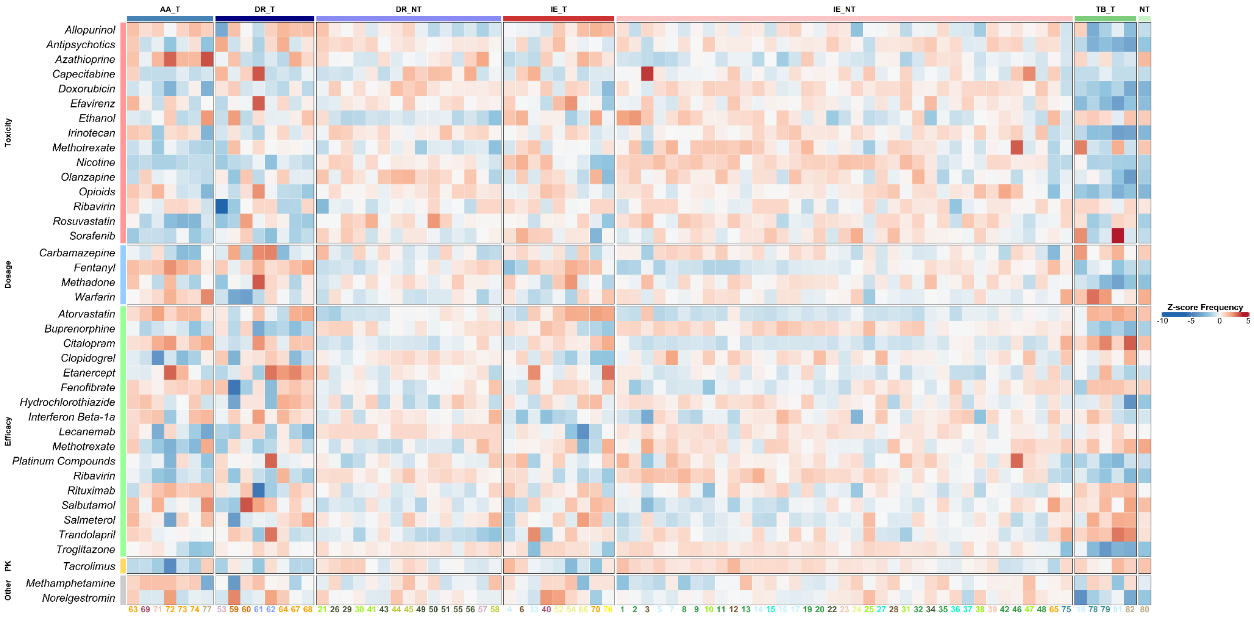

**Fig. S13.2.** Drug-genotype assessment showing the normalized risk genotype frequencies (homozygous and heterozygous) of SNPs grouped on the basis of drugs.

Star-allele haplotypes relevant to GI populations were prioritized based on a haplotype frequency at least 10% higher than that observed in South Asian (SAS) populations in at least one GI population or supported by PharmGKB and CPIC testing levels. From the star alleles identified using Stargazer, Cyrius, and xHLA, a total of 32 star alleles were selected **(Table S13.5, Figure S13.3)**.

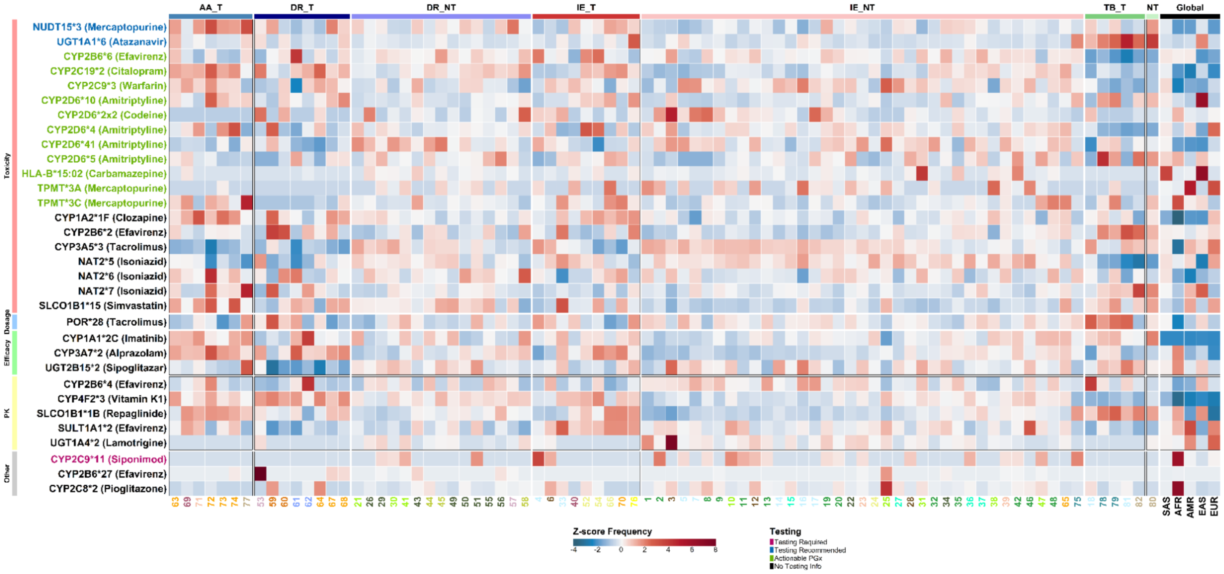

**Fig. S13.3.** Allele frequency comparison between star alleles across 82 Indian subpopulations and the global 1000 Genome populations. The Indian populations are clustered on the basis of tribal status and linguistics, while the star alleles have been annotated by CPIC recommendations associated with drug kinetics.

Ultimately, from these 49 key SNPs and 32 selected star alleles, a final set of 20 important pharmacogenomic variants was compiled **(Table S13.6 and, S13.7, Figure S13.4)**. Each of these 20 variants is associated with an actionable testing level as designated by PharmGKB (i.e., 'actionable PGx,' 'testing required,' or 'testing recommended'). A variant-drug pair is a genetic variant (SNP or star allele) for which a pharmacogenetic association with a particular drug is established, indicating a potential impact on drug response. Building upon our curated set of 20 important pharmacogenomic variations, our analysis revealed that the majority of these unique variant-drug pairs mapped to medicines crucial in several key therapeutic areas: antineoplastics (4 pairs, 20%), antivirals (4 pairs, 20%), anticoagulants (2 pairs, 10%), and antidepressants (2 pairs, 10%). This distribution underscores the significant clinical utility of pharmacogenetic testing to optimize therapeutic strategies, particularly within these critical drug classes.

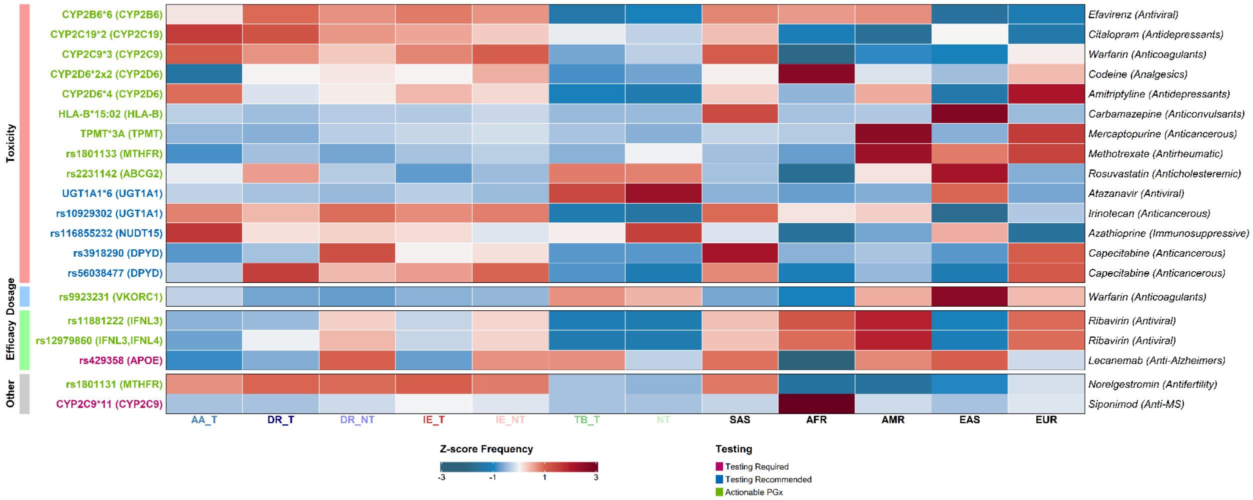

Fig. S13.4. Actionable pharmacogenetic variants (SNPs and star alleles) distilled from Fig. S13.1 and Fig. S13.3, showing normalized allele frequencies across linguistic tribal groups and global reference populations. Variants are colour-coded according to CPIC testing levels (Testing Required, Testing Recommended, and Actionable PGx). Rows are clustered by drug-response category (Toxicity, Dosage, Efficacy, and Other) and annotated on the left, while the corresponding drug names and their therapeutic classes are indicated on the right.

#### Individual pharmacogenetic burden

At an individual level, we assessed the pharmacogenetic burden using the 20 finalized pharmacogenomic variations **(Figure S13.5)**. For each individual, this burden was quantified as the sum of risk genotypes among the actionable SNPs and the count of non-normal metabolizer phenotypes derived from the actionable star allele diplotypes. Across the cohort, each Indian individual carries a median of 4 such actionable pharmacogenetic variants, with a range from 0 to 9. Notably, individuals exhibiting the highest pharmacogenetic burden are predominantly concentrated within the Indo-European (IE) non-tribal and Dravidian (DR) non-tribal speaking populations.

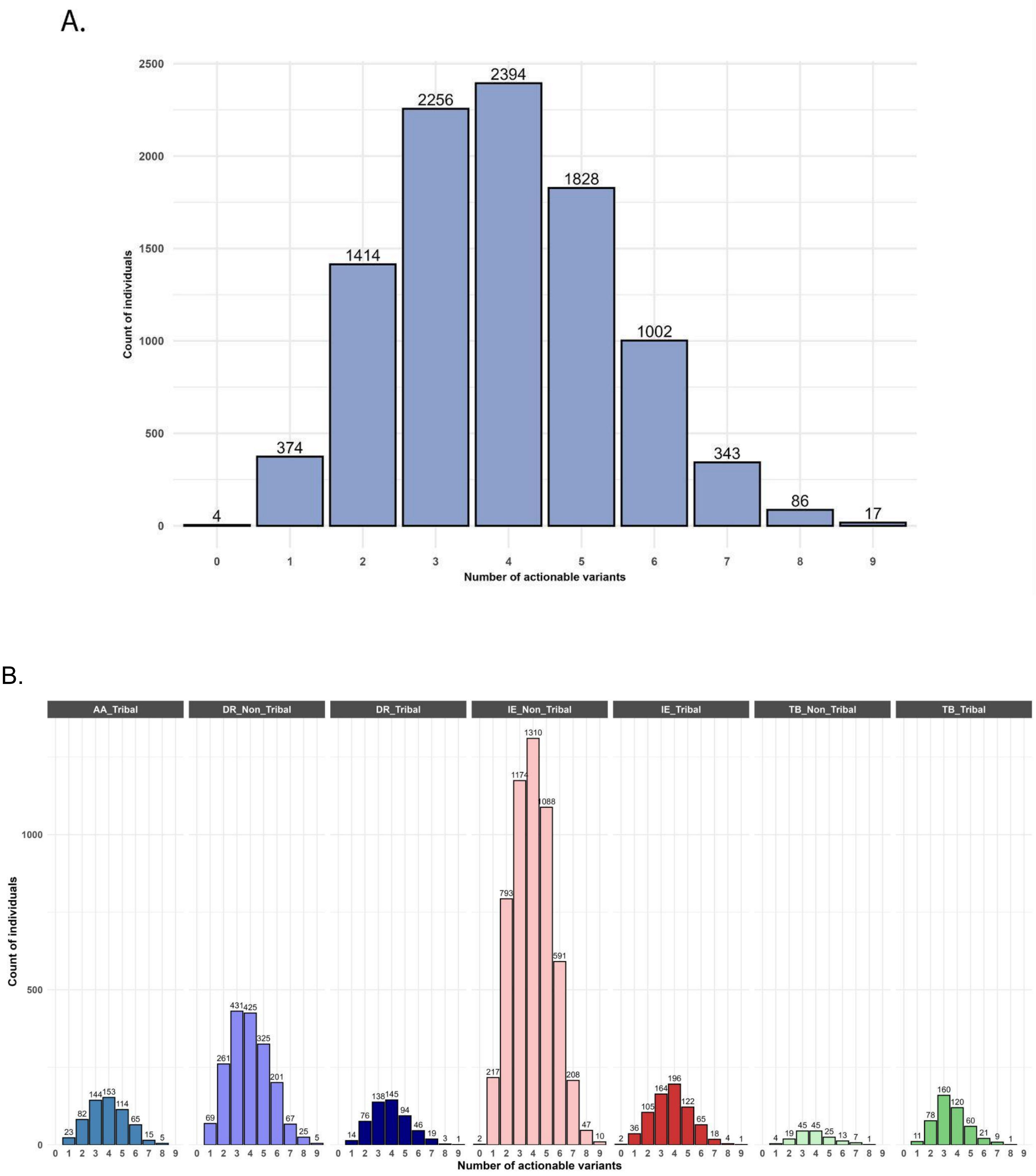

**Fig. S13.5.** Distribution of actionable variants in the GI cohort

(A) Histogram of actionable variants per individual. It displays the frequency distribution of the number of actionable variants carried by each individual in the GI dataset.
(B) This panel illustrates the distribution of actionable variant counts across different linguistic and tribal populations within the GI cohort.

**Population-specific frequencies of key SNPs:**

Significant differences in allele frequencies were identified for SNPs using Fisher's exact test **(Table S13.3 and, S13.6)**. These differences were observed across a broad range of linguistic and tribal/non-tribal groups and further elucidated at a granular level among the 82 distinct populations within the cohort.

**VKORC1 variant rs9923231**

We observed that the variant rs9923231, located 2 kb upstream of the VKORC1 gene and known to influence warfarin dosing (Johnson et al., 2017), exhibited an overall frequency of 0.1896 across the entire GI cohort. This rs9923231 variant further displayed a unique geographical distribution among Indian populations; particularly higher allele frequencies (relative to the global SAS population (0.143)) were observed in populations residing in high-altitude regions such as the Central Himalayas, the Eastern Himalayas, and the Northeastern range, for example, in populations 78 (TB_BPV_1_02, 0.4143), 80 (TB_NER_2_01, 0.3962), and 18 (TB_WHR_1_01, 0.3947). Comparatively, ethnic groups residing in low-altitude plains, from the Eastern and Western Coastal Plains and the Northern Riverine Plains, demonstrated lower risk allele (T) frequencies, with populations 48 (IE_NRP_2_09, 0.0928) and 51 (DR_ECP_2_07, 0.0556) showing frequencies considerably below the global SAS (0.143). Given that the T allele is associated with increased warfarin sensitivity, individuals with the TT genotype (513/9768) may require a decreased dose of warfarin to avoid over-anticoagulation and bleeding risks. This spatial stratification suggests a potential role of environmental adaptation, particularly in response to chronic hypoxia experienced by people residing in high-altitude areas.

**NUDT15 variant rs116855232**

Among other interesting variants, the missense NUDT15 variant rs116855232 (risk allele: T), associated with azathioprine and mercaptopurine dosage and toxicity (Pratt et al., 2022), has been reported with significant frequency among Asian populations, i.e., GI: 0.0789, Indigen: 0.08, 1KGP3-SAS: 0.0674, and 1KGP3-EAS: 0.0957. Further, the GenomeIndia dataset offers more granularity, enabling more precise identification of specific populations that may benefit from preemptive pharmacogenomic testing. For instance, the highest allele frequencies were observed in tribal populations of Austro-Asiatic (AA) origin, with GI population 72 (AA_EPL_1_02) exhibiting the peak at 0.2083. Most other AA tribal populations also showed notably high frequencies, including populations 77 (0.2067), 73 (0.1413), 71 (0.14), 63 (0.1374), and 74 (0.1369), with only population 69 having a frequency lower than in global SAS. In contrast, non-tribal Indo-European-speaking individuals in population 16 exhibited a comparatively lower frequency (0.0203) of the risk allele. These inter-population differences in allele frequency were statistically significant (Fisher's exact test, p-value < 0.05) when comparing the 82 populations. Stratifying by genotype, we could identify a high number of individuals with the heterozygous CT genotype (n = 1390/9768), suggesting substantial carrier burden for the NUDT15 risk allele. Endogamy prevalent in many Indian populations, particularly within tribal and closed caste systems, can elevate the frequency of the homozygous TT genotype (observed: 76/9768; expected: 61/9768) over generations through increased rates of consanguinity and within-group mating. This warrants targeted intervention for specific pharmacotherapy in a population-specific manner.

**DPYD variants rs56038477 and rs3918290**

Notably, a missense and a splice donor variant in the DPYD gene (rs56038477 [GI: 0.0159] and rs3918290 [GI: 0.0025], respectively) associated with capecitabine and fluorouracil toxicity (Pratt et al., 2024) occur at a considerably high frequency in certain Indian populations as compared to South Asians (rs56038477 [SAS: 0.0166] and rs3918290 [SAS: 0.0075]). As an example, consider population 61 (DR_NGH_1_02, 0.0867) belonging to the Dravidian-speaking tribal group and population 3 (IE_WPL_2_02, 0.0645) of non-tribal Indo-European origin. The GI dataset revealed 6 individuals with the high-risk homozygous genotype and 349 individuals with the heterozygous genotype for the DPYD variants. This provides knowledge about the risk of adverse drug reactions (ADRs) related to 5-FU-DPYD interaction.

**Analysis of clinically relevant star alleles:**

Among several star alleles, CYP3A5, CYP2C19, and CYP2D6 show population-specific differences in frequencies within GI populations and with global datasets **(Table S13.6, Figure S13.6)**.

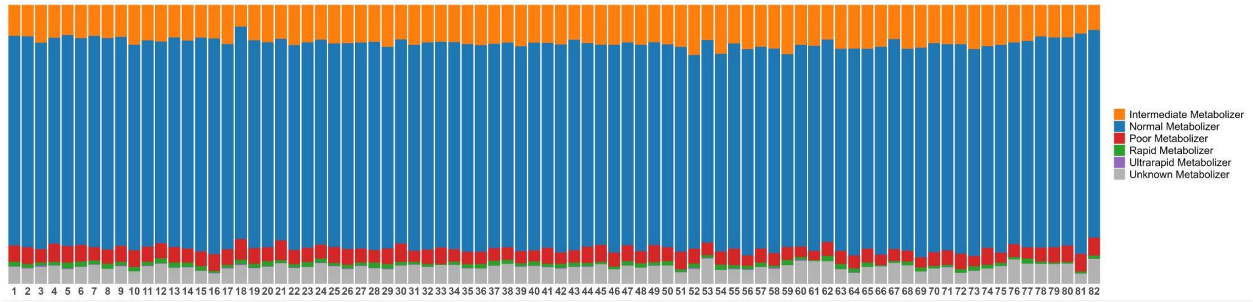

**Fig. S13.6.** Stacked bar plot representing the metabolic profiles across 82 Indian subpopulations.

The CYP3A5 diplotype *3/*3, which results in a loss of CYP3A5 enzyme activity, is associated with poor metabolism of tacrolimus, an immunosuppressant used in organ transplantation (Birdwell et al., 2015). Individuals with poor metabolizer diplotypes require lower doses of tacrolimus or alternate medications to avoid drug accumulation and toxicity. In the GI cohort, this diplotype was observed in approximately 45% of the population, the highest being in the Tibeto-Burman non-tribal groups, indicating that genetic testing is recommended before organ transplantation using immunosuppressives. CYP2C19 haplotypes that affect the metabolism of clopidogrel, a blood thinner, are used to prevent cardiac problems like strokes and heart attacks (C. R. Lee et al., 2022). It gets converted into an active form in the liver by an enzyme encoded by a gene, CYP2C19. Haplotypes *2 and *3 in the gene CYP2C19 significantly reduce the clopidogrel active metabolite formation, increasing on-treatment platelet reactivity and thus causing increased risk for adverse cardiac and cerebrovascular events by ineffective thrombosis prevention. There are approximately 15% poor metabolizers and 45% intermediate metabolizers of clopidogrel in the GI populations who should avoid use of clopidogrel if possible and use alternative medications.

**Population specific rareIndian-exclusive CYP2D6 alleles**

We observed five non-singleton CYP2D6 alleles that occur exclusively in the Indian population **(Table S13.5, S13.6)**. This was also reported in the Indigen CYP2D6 study (Sivadas et al., 2024), but the GI study offers more granularity in highlighting the specific populations, their linguistics, tribal status, and their primary residence.

**Table S13.9:** Rare CYP2D6 haplotypes identified in GenomeIndia (GI) populations.

The table summarizes rare CYP2D6 haplotypes detected in the GenomeIndia cohort, highlighting the GI populations in which each haplotype attains its highest observed frequency. For each haplotype, the corresponding population, maximum allele frequency, major ethno-linguistic group, and biogeographic region of India are reported.

| CYP2D6 allele | Indigen | SAS \| EAS \| EUR | GI All | Population (Frequency) | Linguistic Group | Biogeography |
|---|---|---|---|---|---|---|
| *86 | 0.02 | 0.02 \| 0.0002 \| 0 | 0.02 | 60 (0.09) | DR_T, DR_NT | Plain regions |
| *111 | 0.008 | 0.008 \| 0 \| 0.004 | 0.0039 | 31 (0.03) | IE_T, IE_NT | Plain regions |
| *112 | 0.003 | 0.002 \| 0 \| 0 | 0.0038 | 61 (0.03) | IE_NT, DR_NT | Plain regions |
| *113 | - | 0.008 \| 0 \| 0 | 0.0051 | 25 (0.04) | IE_NT, DR_NT | Plain regions |
| *99 | 0.0005 | 0.002 \| 0 \| 0 | 0.00081 | 54 (0.02) | IE | - |

Notably, the GI dataset showed the highest prevalence of alleles of unknown/indeterminate metabolizer status, higher than reported by Indigen and SAS (GI: 18%; Indigen: 4.7%; SAS: 5%). This emphasizes the importance of characterizing the unknown diplotypes that can elucidate the knowledge of CYP2D6 metabolizer phenotypes. The CYP2D6 gene showing haplotype variation across Indian subpopulations affects the metabolism of psychotropic and analgesic drugs. The *4 haplotype, encoding a non-functional enzyme, is associated with poor metabolism of tricyclic antidepressants such as amitriptyline (Hicks et al., 2017). This haplotype was found at a higher frequency in tribal groups within the GI dataset: —population 74 (Austro-Asiatic, 0.2178) and population 52 (Indo-European, 0.2083), —compared to global South Asians (0.0896). Genotype analysis revealed 66 individuals with the *4/*4 diplotype having a poor metabolizer status and a significantly increased risk of adverse drug reactions when treated with standard doses of amitriptyline. Contrarily, the *2x2 haplotype, characterized by multiple copies of functional CYP2D6 alleles, is associated with ultrarapid metabolism of codeine, leading to increased risk of opioid toxicity. This haplotype was observed at a high frequency in population 3 (non-tribal Indo-European, 0.0564), which is markedly elevated compared to South Asians (0.0069). These findings emphasize the need for pharmacogenomic testing to guide antidepressant and analgesic prescribing in Indian populations with extreme metabolizer phenotypes.

**Star alleles affecting antiviral drug response**

Similarly, variants CYP2B6*6(Desta et al., 2019), rs12979860, rs11881222 (Lange & Zeuzem, 2011), and UGT1A1*6 (Gammal et al., 2016) affecting the activity of antiviral drugs such as efavirenz, ribavirin atazanavir, and atazanavirribavirin show unequal distribution of allele frequency across Indian populations **(refer to Table S13.6)**, the highest being in Dravidian-speaking tribal groups (61: DR_NGH_1_02, 0.653 [SAS: 0.375], 53: DR_EGH_1_01, 0.4265 [SAS: 0.2404]), non-tribal groups (45: DR_SDN_2_03, 0.3305 [SAS: 0.2288]), and Tibeto-Burman tribal populations (81: TB_WHR_1_02, 0.2946 [SAS: 0.014]).

Some other clinically relevant variants include:

**BCHE variant rs104893684 (Table S13.8).**

The missense variant rs104893684 (A>G) in the BCHE gene is associated with reduced butyrylcholinesterase enzyme activity and poses a clinical risk during anaesthesia (David et al., 2015). This variant is classified as pathogenic/likely pathogenic in ClinVar. In our dataset, we identified this allele in 45 individuals across 29 populations, with all carriers being heterozygous except for a single individual from population 13 (IE_NRP_2_06), who carried the homozygous minor allele (allele frequency: 0.01524).

Previous reports highlighted elevated frequencies of this variant in the Vysya community of Tamil Nadu, India. Our results extend this observation by demonstrating notable frequencies in several additional Indian populations, all of which belong to Indo-European or Dravidian linguistic groups. Three populations, population 13 (IE_NRP_2_06; 0.01524), population 43 (DR_ECP_2_01;

0.01786), and population 46 (IE_NRP_2_01; 0.01796) exhibit allele frequencies exceeding 1%, all of which are higher than the reported global South Asian (SAS) frequency of 0.0075.

**GLP1R variant rs6923761 (Table S13.8)**

We identified the PharmGKB level 3 missense variant rs6923761 (G>A) in the GLP1R gene which affects the efficacy of GLP-1 receptor agonists (like semaglutide and liraglutide) in reducing HbA1c levels (Dawed et al., 2023). This variant is present in 164 individuals, with 82 being homozygous for the minor 'AA' genotype. It is particularly prevalent in Indo-European and Dravidian linguistic groups, with 36 populations showing a higher frequency than the global SAS (0.1256), the highest being in population 4 (IE_WHR_1_02, 0.2463).

These findings highlight the need for appropriate pharmacogenomics testing before drug treatment.

#### Supplementary section S14: Eurocentric bias and polygenic scores (PGS)

Devashish Tripathi[1,2], Debasrija Mondal[3], Chandrika Bhattacharyya[1], Divya Tej Sowpati[4,5], Shweta Ramdas[3], Analabha Basu[1,2]

[1]BRIC - National Institute of Biomedical Genomics (BRIC-NIBMG), Kolkata, India. [2]Regional Centre for Biotechnology (RCB), Faridabad, India. [3]Centre for Brain Research (CBR), IISc Campus, Bengaluru, India. [4]CSIR - Centre for Cellular and Molecular Biology (CSIR-CCMB), Hyderabad, India. [5]Academy of Scientific and Innovative Research, Ghaziabad, India.

#### Summary

##### Eurocentric Bias in Public Genomic Datasets and the Underrepresentation of South Asians

Public genomic resources have greatly advanced our understanding of human genetic variation and disease risk, yet they remain strongly Eurocentric (Dokuru et al., 2024), with individuals of European ancestry vastly overrepresented relative to their share of the global population. Major datasets - including the 1000 Genomes Project, UK Biobank, gnomAD, and the GWAS Catalog - are disproportionately European, leading to limited transferability of predictive models and variant interpretations to non-European groups. This imbalance is particularly pronounced for India, a region of exceptional genetic diversity shaped by deep population structure, endogamy, and founder events. Underrepresentation of Indian genomes hampers accurate disease-risk prediction and variant classification, underscoring the need for improved inclusion in global reference resources.

To place these findings in a global context, we then compared our dataset with publicly available reference datasets (Fig S14.1). Correlations with the South Asian (SAS) subset of the 1000 Genomes Project were higher than those obtained with the South Asian subset of gnomAD, despite the fact that 1000 Genomes' SAS samples are mostly diaspora rather than native Indian. This suggests that the allele-frequency spectrum in 1000 Genomes SAS better approximates the core Indian-subcontinental gene-pool than gnomAD's SAS subset, which is more affected by Eurocentric sampling and heterogeneous recruitment. Together, these observations underscore that while allele-frequency patterns across Indian populations are remarkably consistent, large global reference resources continue to incompletely reflect South Asian genomic diversity - highlighting the urgent need for more inclusive sampling and unbiased representation in future global datasets.

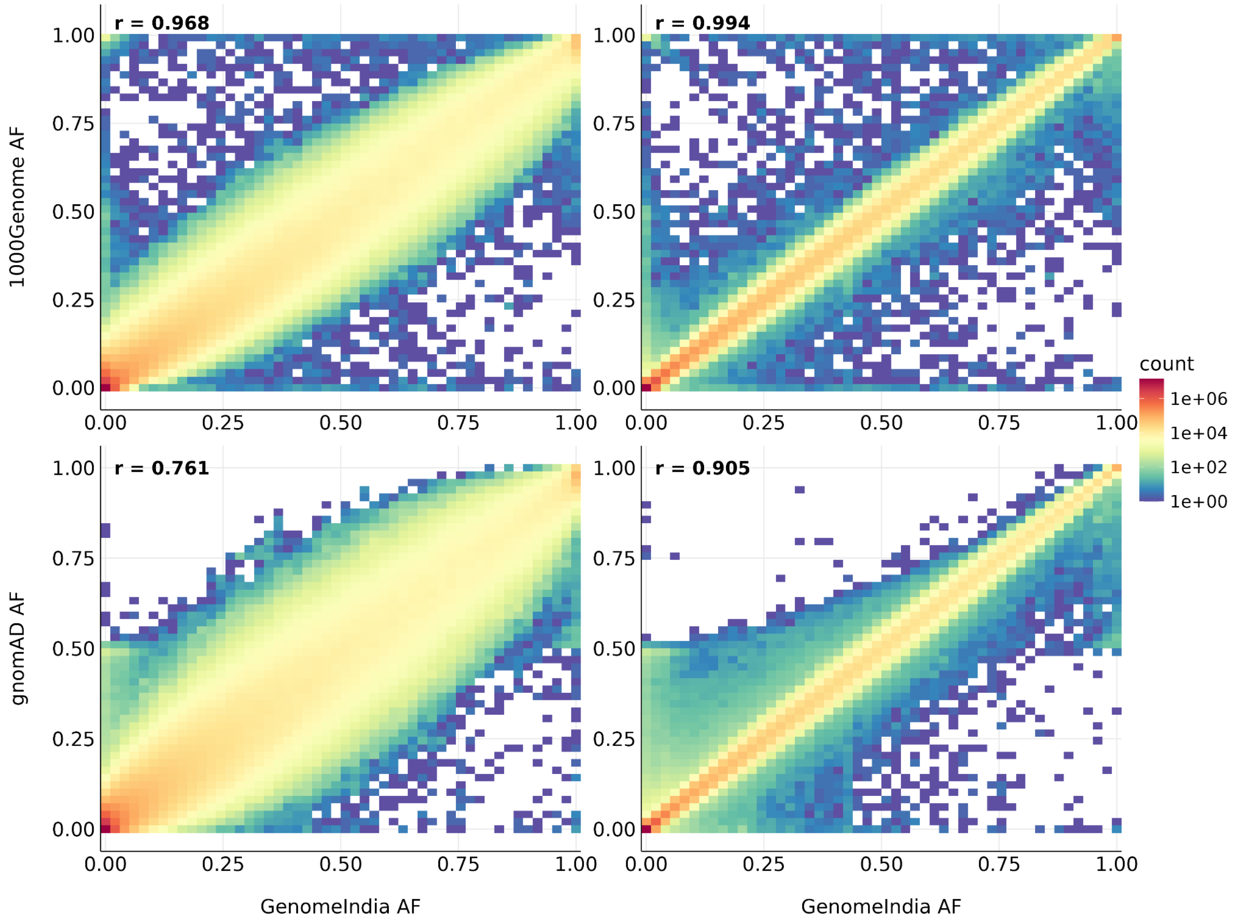

**Figure S14.1.** Allele frequency concordance between GenomeIndia and public reference datasets. Scatter-density plots show correlations in allele frequencies between GenomeIndia and the 1000 Genomes Project (top left), with the South Asian (SAS) subset from 1000 Genomes project shown on the top right, and between GenomeIndia and gnomAD (bottom left), with the SAS subset from gnomAD shown on the bottom right. Pearson's r values indicate consistently higher concordance with 1000 Genomes (r = 0.968 - 0.994) compared to gnomAD (r = 0.761 - 0.905). The stronger correlation with 1000 Genomes likely reflects its more balanced representation of South Asian ancestry, whereas lower concordance with gnomAD highlights persistent Eurocentric sampling bias and limited South Asian coverage in global genomic resources.

##### Phenotype data

In this section, we presented the distribution of height, weight, and BMI across all the GI populations stratified by Sex. We further assessed the portability of PGS developed using individuals of European ancestry on Europeans, Africans, and Indians from the UK Biobank (UKBB) cohort and GI populations.

All phenotypic data (blood biochemistry, anthropometry, socio-demographic data, and lifestyle) were collated and cleaned at the Centre for Brain Research. This process included standardizing all columns to the same scale, removing any identifying information, and removing extreme outlier values (*more details of phenotype data are provided in a separate manuscript*). Of the 9,768 sequenced samples, 9,330 samples have phenotype data available.

Figures S14.2, S14.3, and S14.4 show the distribution of height, weight, and body mass index (BMI) across ethnicities, separated by sex. Detailed distributions and values for other phenotypic data are included in an accompanying manuscript, and can be visualized on the GenomeIndia phenotype data browser.

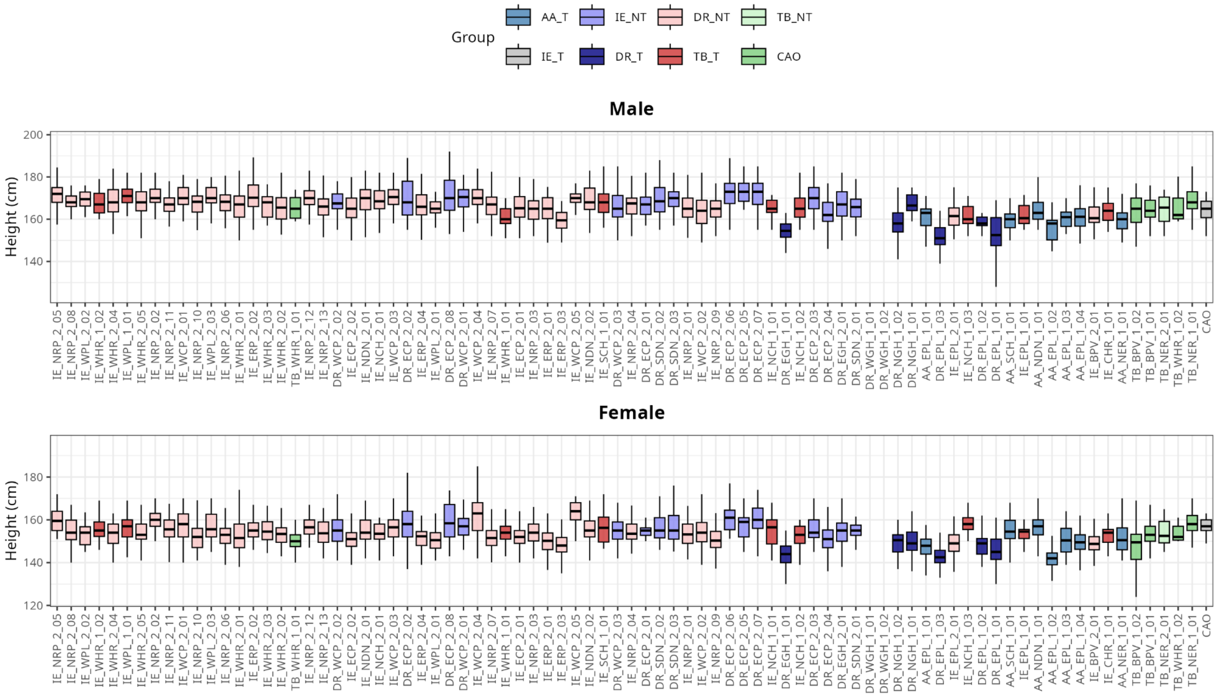

**Figure S14.2**. Distribution of height across 83 ethnicities in males (top) and females (bottom)

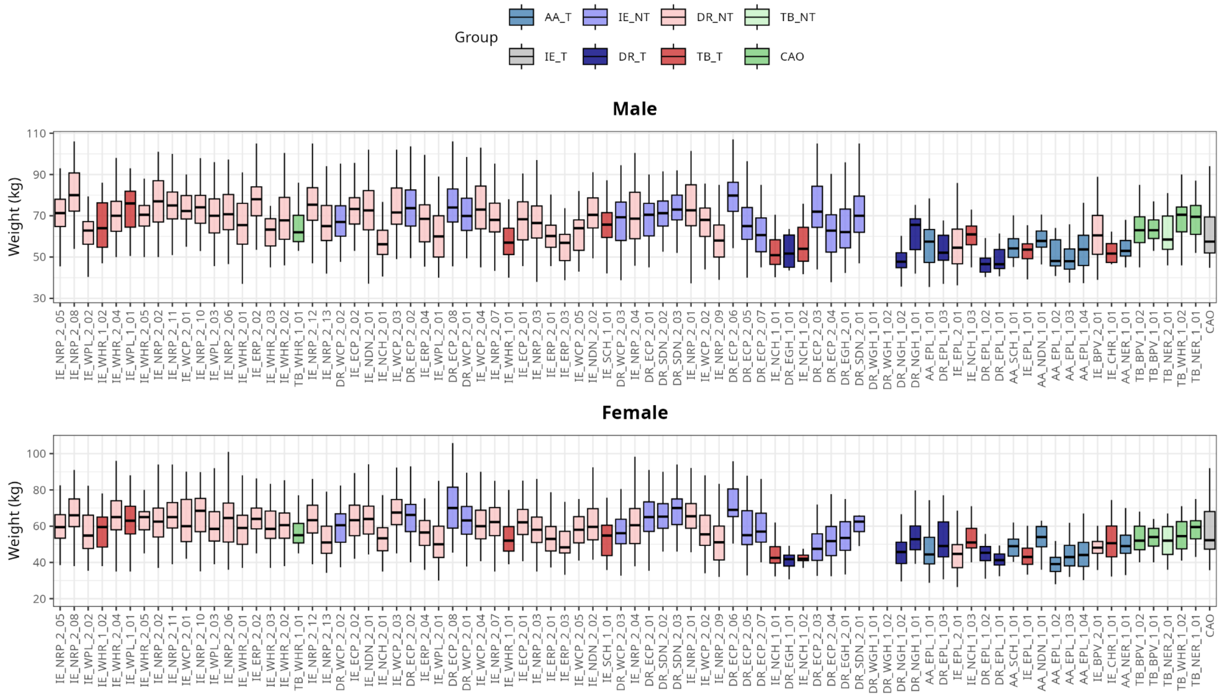

**Figure S14.3**. Distribution of weight across 83 ethnicities in males (top) and females (bottom)

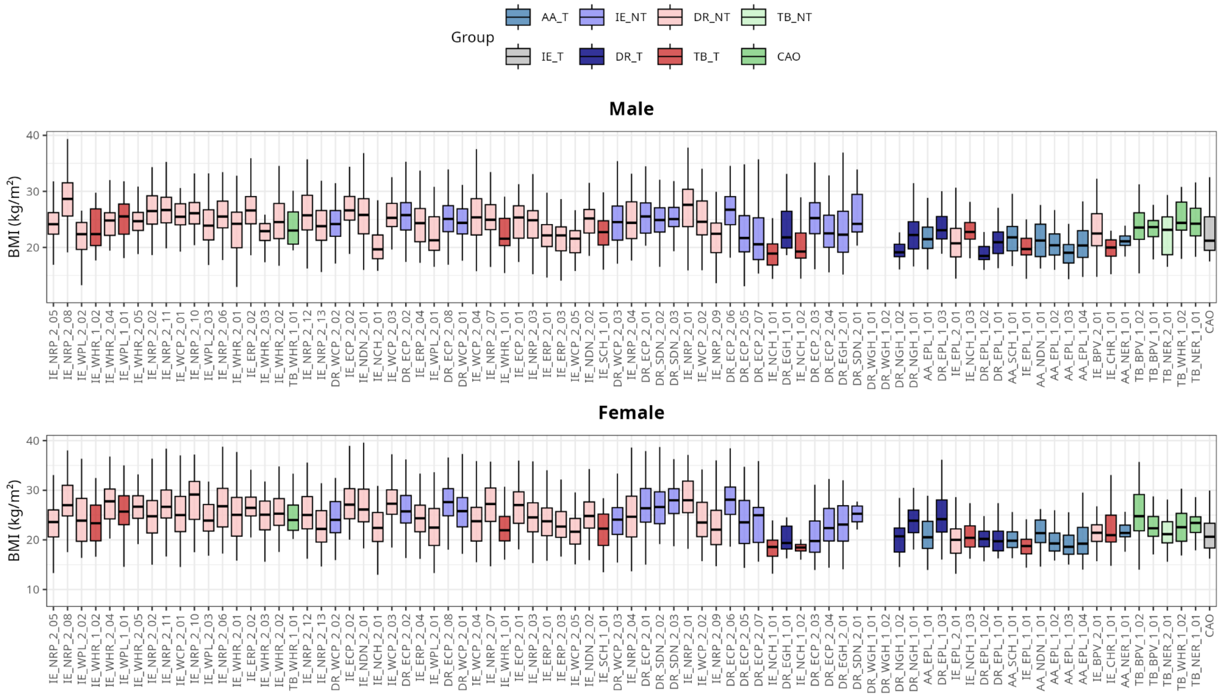

**Figure S14.4**. Distribution of BMI across 83 ethnicities in males (top) and females (bottom)

#### Investigating the portability of PGS to GenomeIndia populations

We generated the sex-stratified phenotype distributions of height, weight, and BMI for British, African, and Indian individuals from UKBB and GI. The height distribution for females in GI is lower compared to females in UKBB. However, the differences were more pronounced in both males and females, with significantly lower weights and BMIs compared to the British, Africans, and Indians from the UKBB.

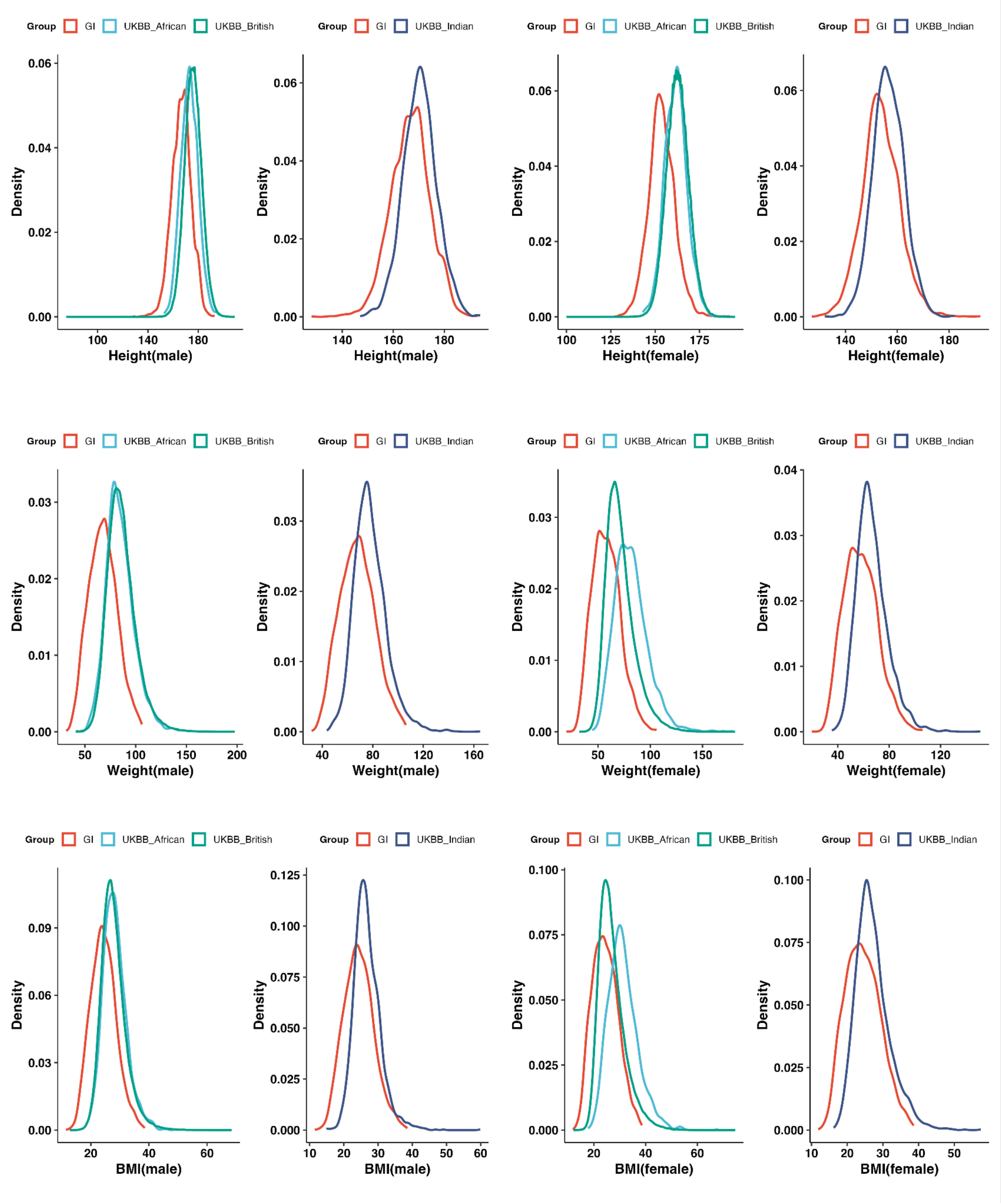

**Figure S14.5.** Phenotype distribution of height, weight, and BMI stratified by Sex (Male/Female) in British, Africans, and Indians from the UKBB and GI. GI populations show large differences in phenotypic distribution compared to other populations in UKBB, especially weight and BMI

We assessed the cross-population portability of European-derived polygenic scores (PGS) for height, weight, and body mass index (BMI) in the GI cohort and benchmarked their performance against British, African, and Indian ancestries in the UKBB (Bycroft et al., 2018). We utilized published PGS from the PGS Catalog for height (PGS002804), weight (PGS004373), and BMI (PGS002313), and used the PGS_calc software for generating the PGS for the populations (Lambert et al., 2024). As expected from the strong Eurocentric bias in existing GWAS resources (Martin et al., 2019), predictive accuracy ($R^2$) was highest in the British population and showed a large reduction in non-European groups, with the steepest decline in individuals of African ancestry (Table S14.1). This observation is due to pronounced differences in allele frequencies and LD structure between African and European populations. For example, the $R^2$ for height decreased from 0.223 in British individuals to 0.017 in Africans, and for BMI from 0.329 to 0.029.

A comparison of PGS performance between Indians from UKBB and the GI cohort revealed more nuanced, trait-specific patterns. For height, a trait with high heritability and relatively low environmental modulation, PGS performance was broadly consistent across Indian subpopulations ($R^2$ = 0.148 in GI vs. 0.112 in UKBB Indians). In contrast, weight and BMI showed substantial divergence: while the BMI PGS retained moderate predictive ability in UKBB Indians ($R^2$ = 0.097), it explained almost no variance in the GI cohort ($R^2$ = 0.007). These results suggest that reduced portability is shaped not only by genetic distance from Europeans but also by cohort-specific environmental exposures and potential Gene-by-Environment interactions. The superior performance of European-derived weight-related PGS in UKBB Indians may partly reflect environmental similarity with the UK population, whereas the distinct lifestyle and nutritional context of the GI cohort reduces translatability.

Overall, our findings indicate that the portability gap arises from a combination of population genetic divergence and environmental mismatch, reinforcing the need for more ancestrally diverse GWAS for improving the portability of PGS.

**Table S14.1.** Performance of European-derived PGS for height, weight, and BMI on UKBB and GI Populations.

| Trait | Cohort | Population | PGS | | N |
|---|---|---|---|---|---|
| | | | $R^2$ (95% CI) | Coefficient (95% CI) | |
| Height | UKBB | African | 0.017(0.009,0.027) | 0.369(0.329,0.410) | 3123 |
| Height | UKBB | British | 0.223(0.221,0.225) | 0.710(0.708,0.712) | 425708 |

| | | | | | |
|---|---|---|---|---|---|
| Height | UKBB | Indian | 0.112(0.096,0.128) | 0.536(0.515,0.558) | 5467 |
| Height | GI | Indian | 0.148(0.134,0.162) | 0.414 (0.391,0.437) | 8277 |
| Weight | UKBB | African | 0.030(0.019,0.043) | 0.692 (0.542, 0.843) | 3075 |
| Weight | UKBB | British | 0.156(0.154,0.158) | 1.119 (1.112, 1.125) | 419255 |
| Weight | UKBB | Indian | 0.057(0.046,0.070) | 0.835 ( 0.757,0.913) | 5508 |
| Weight | GI | Indian | 0.011(0.007,0.016) | 0.457 (0.524, 0.391) | 8254 |
| BMI | UKBB | African | 0.029(0.018,0.041) | 0.655 (0.525, 0.785) | 3119 |
| BMI | UKBB | British | 0.329(0.327,0.331) | 1.622 (1.615, 1.629) | 425270 |
| BMI | UKBB | Indian | 0.097(0.082,0.112) | 0.985 (0.906,1.064) | 5461 |
| BMI | GI | Indian | 0.007(0.004,0.012) | 0.573 (0.507,0.640) | 8179 |

In the original GWAS summary statistics used to construct the weight PGS, the majority of variant effect sizes were negative due to the coding of the effect allele. This resulted in a PGS where higher scores corresponded to lower weight. To standardize interpretation across traits and avoid confusion, we re-oriented the weight PGS by multiplying the score by −1, ensuring that a higher PGS reflects a higher genetically predicted weight. This transformation does not affect model fit, variance explained ($R^2$), or statistical significance; it only changes the direction of the regression coefficient for interpretability.

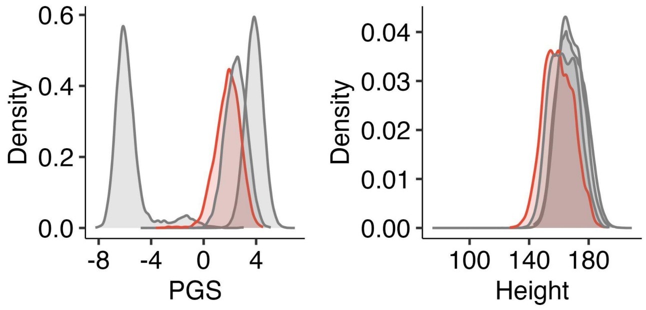

**Figure S14.6**. Estimated PGS of height on African, British, Indian (from UK BioBank) and GI populations (left panel). Observed distributions of height in the same populations (right)

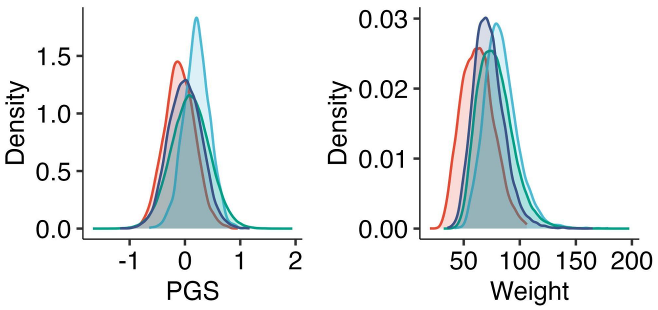

**Figure S14.7**. Estimated PGS of weight on African, British, Indian (from UK BioBank) and GI populations (left panel). Observed distributions of weight in the same populations (right)

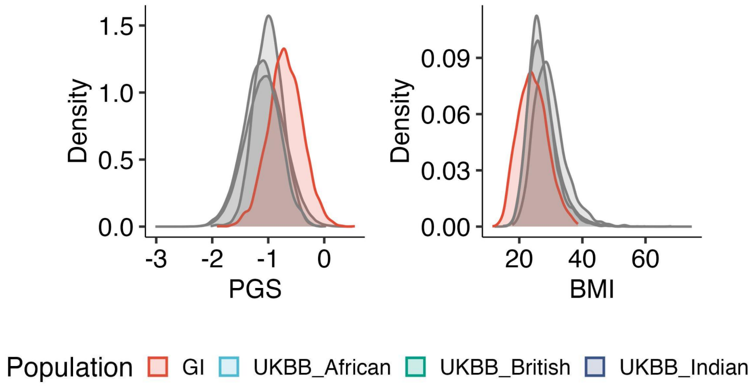

**Figure S14.8**. Estimated PGS of BMI on African, British, Indian (from UK BioBank) and GI populations (left panel). Observed distributions of BMI in the same populations (right)

#### Supplementary section S15: Design and testing of GI Imputation Panel

Shreya Chakraborty[1,2], Bratati Kahali[1]

[1]Centre for Brain Research (CBR), IISc Campus, Bengaluru, India. [2]Interdisciplinary Mathematical Sciences, Indian Institute of Science (IMI- IISc), Bengaluru, India.

##### Summary

Central to the success of genome-wide association studies is genotype imputation, a statistical technique that leverages reference panels to infer untyped variants, thereby increasing variant density and power for discovery. However, the accuracy of imputation and thereby the validity of downstream analyses, could critically depend on the ancestral match between study samples and the reference panel. One of the key contributions of this study is the development of an Indian population-specific haplotype reference panel that outperforms existing panels in genotype imputation performance. The panel designed using GI data is able to achieve superior imputation accuracy at specific genomic loci compared to its closest competitors, the GenomeAsia (GAsP) and the TOPMed panels, underscoring its practical utility in fine-scale genetic analyses. This panel is enriched for ultra-rare variants enabling high-resolution imputation at both the individual and variant level. Importantly, the panel supports accurate imputation within specific subpopulations as well as also across datasets representing the broader genetic diversity of the Indian population. On the whole, this imputation panel clearly becomes the most comprehensive and reliable resource available for researchers studying Indian genetics. In particular, it holds much promise for downstream applications in population genetics, disease mapping, and genome-wide association studies within the understudied, genetically diverse Indian population.

##### Phasing

###### Methods

We utilized SHAPEIT5 (Hofmeister et al., 2023) to infer haplotype structure from 9768 WGS-derived genotypes. Prior to phasing, we utilized the data on 244 trios to filter out variants which show a Mendelian Inheritance Error rate >5%. We utilized this trio data to phase the genotypes in two stages. At first, we phased the variants with MAF >0.1% and subsequently, formed a scaffold with these phased variants upon which we phased the rarer variants. We assessed the accuracy of phasing by computing the switch error rate.

##### Haplotype Reference panel construction and LD characterization

**Methods:** We characterized linkage disequilibrium (LD) patterns across the genome using the complete variant call set. We calculated the raw VarLD (Ong & Teo, 2010) scores to assess regional differences in LD between our population and four 1000 genomes superpopulations (EUR, AFR, SAS, EAS). This analysis was based on the overlapping set of variants between our population and these superpopulations. The genome was divided into sliding windows spanning 50 SNPs. In each genomic window, we calculated the LD matrix of each of the two populations being compared (say GI and SAS) and constructed two SNP-by SNP correlation matrices respectively. Each element of the population-specific LD matrix would represent the signed $r^2$ between two variants obtained as:

$$\frac{(p_{AB} - p_A p_B)^2}{p_A p_a p_B p_b}(-1)^{I(p_{AB} < p_A p_B)}$$

where $p_{AB}$ denotes the frequency of haplotype AB, $p_A, p_a, p_B, p_b$ denote the respective allele frequencies and the indicator function $I(p_{AB} < p_A p_B)$ taking value one when the condition is satisfied and zero otherwise.

This LD matrix summarizes how variants are correlated (magnitude and direction) within the said window for a given population. We then performed an Eigen decomposition of each of the LD matrices and matched the ranked eigenvalues of each of the LD matrices in order. Next, we obtained the sum of the absolute differences between the ranked eigenvalues as the VarLD scores for the genomic window under consideration in the following manner:

$$\sum_i |\lambda_i^{GI} - \lambda'^{POP}_i|$$

where $\lambda_i$ and $\lambda'_i$ denote the rank-matched eigen values of the LD matrices for the GI population and the 'POP' under consideration respectively, where POP$\in$ {AFR, EUR, EAS, SAS}.

We slide the window along the genome to obtain the VarLD score profiles for each of the population comparisons.

We first constructed the haplotype reference panel using variants with a minor allele count (MAC) ≥ 3. To characterize LD patterns within this panel, we calculated localized pairwise linkage disequilibrium ($r^2$) across the genome in sliding windows of 50 kb. The resulting LD matrices were combined, and variants exhibiting pairwise LD $r^2 > 0.2$ were identified and are reported (see Data Availability). To assess population-specific variation in LD structure, we analyzed patterns of allelic correlation within each of the seven population clusters. Mean $r^2$ values were summarized across 50-kb windows and plotted against physical distance to evaluate the extent and decay of LD across the genome. Additionally, LD score (LDSC) estimates for the panel were computed and are provided to facilitate downstream analyses in genetically similar populations (see Data Availability).

**Results**

We observe regional divergences in LD patterns of our panel compared to 1000 genomes superpopulation (Figure S15.1 a,b).

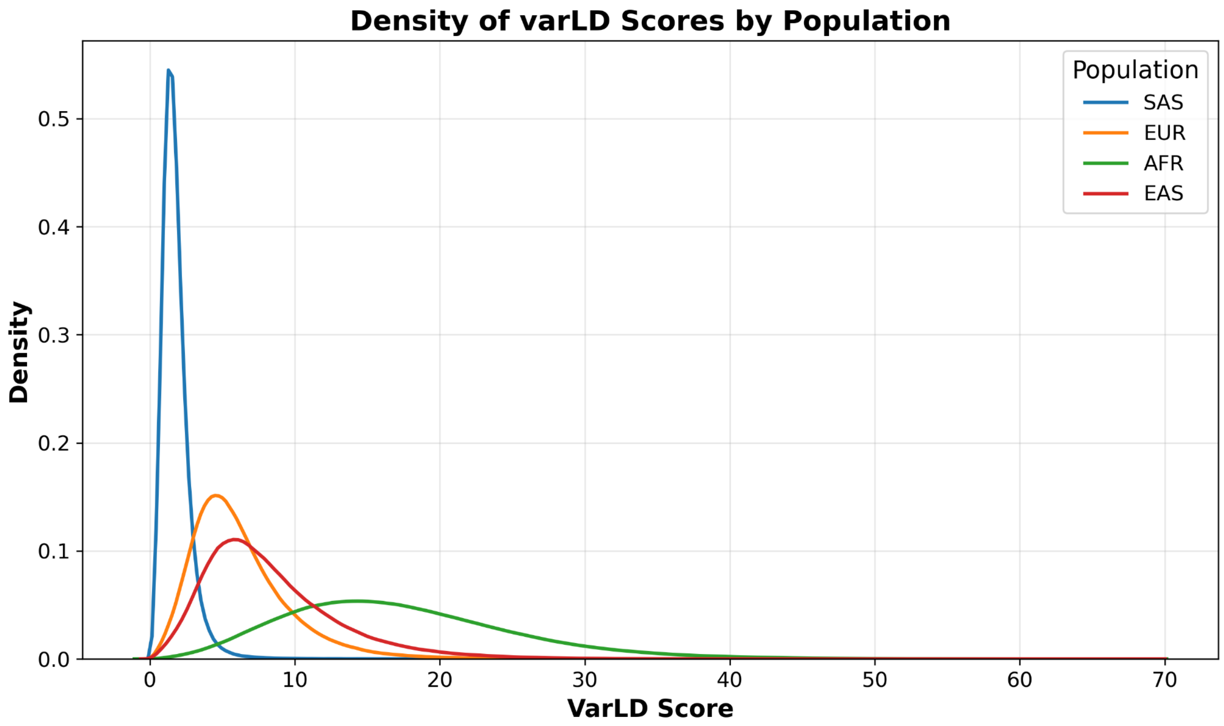

**Figure S15.1a.** Density distributions of varLD scores comparing the LD divergence of the GI panel 1000G subpopulations. The GI panel shows minimal LD divergence from the 1000 (SAS) population (mean varLD = 1.81), reflecting concordance with regional LD structure. In contrast, European, East Asian, and African populations exhibit progressively higher divergence (mean varLD = 6.23, 8.37, and 17.29, respectively), underscoring the panel's power to capture fine-scale regional variation often missed by global reference datasets (LD divergence)

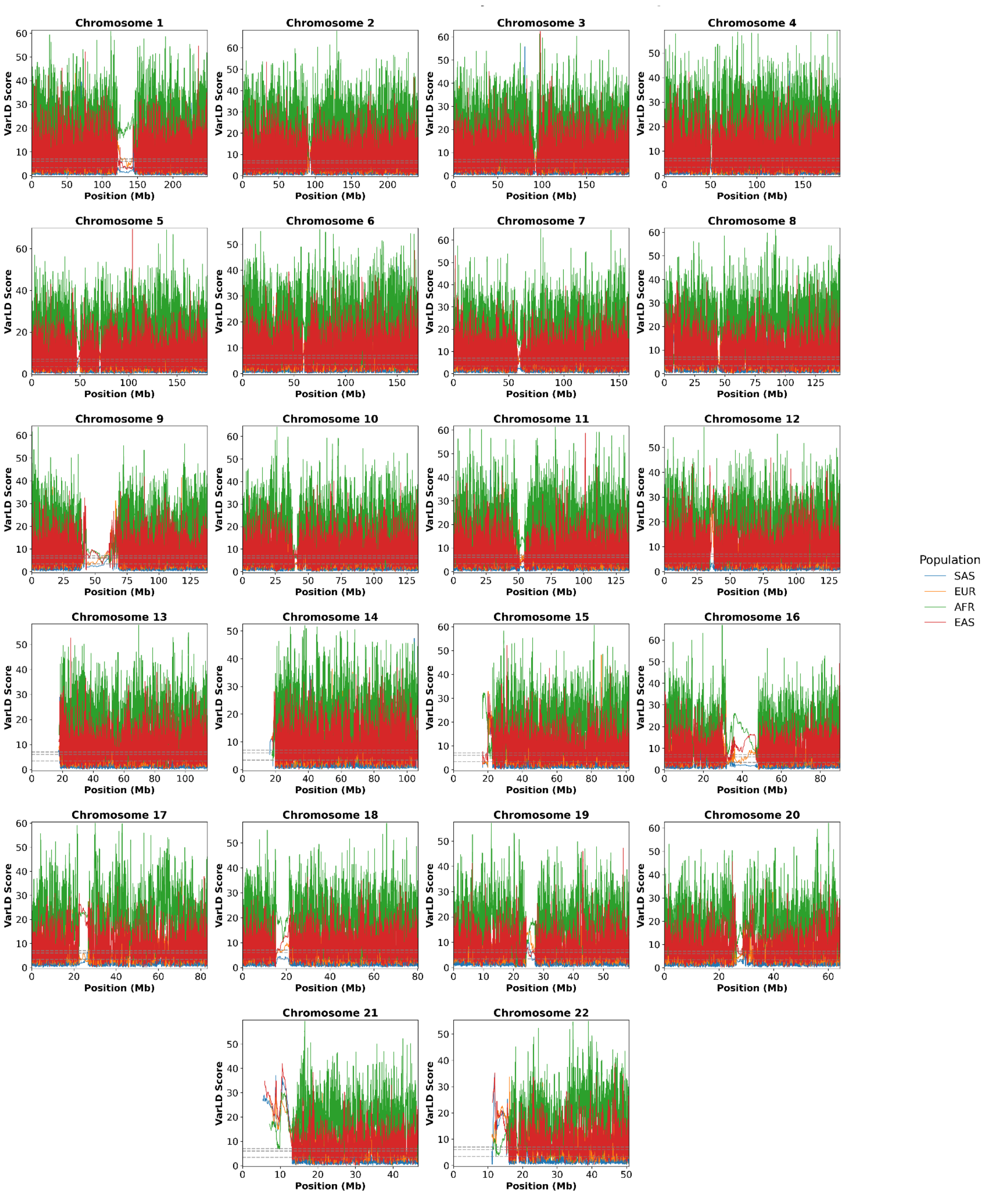

**Figure S15.1b.** This figure illustrates regional differences in local linkage disequilibrium (LD) patterns between each 1000 Genomes superpopulation and the GI haplotype reference panel. The y-axis represents varLD scores, where higher values indicate greater divergence in LD structure. In each panel, grey dashed lines mark the 95th, 99th, and 99.99th percentiles of the genome-wide varLD score distribution, highlighting regions with significantly elevated LD divergence.

Regions with raw VarLD scores near or below zero indicate little to no LD divergence between populations, while regions exceeding the dashed percentile lines represent those with the highest LD differentiation. As expected, the curve for the SAS population lies closest to zero, followed by EUR, EAS, and AFR, reflecting increasing divergence in LD structure relative to the reference population.

Across all seven GI sub-population specific clusters, LD decreased with increasing inter-marker distance, consistent with the expected effect of recombination (Figure S15.2). Overall, mean $r^2$ values ranged from approximately 0.80 at close marker spacing (<1 kb) and declined to around 0.55 at 50 kb. The LD decay trajectories were largely overlapping among clusters, indicating comparable haplotype structures and recombination histories. We formally tested whether LD decay trajectories differed across clusters using a linear model with an interaction term between physical distance and cluster (Mean_$r^2$ ~ Distance bin * Cluster). ANOVA on this model indicated that LD decays significantly with distance ($F = 2569.49$, $P < 1e\text{-}170$), as expected. Neither the cluster main effect ($F = 0.87$, $P = 0.53$) nor the Distance bin × Cluster interaction ($F = 0.16$, $P = 0.99$) was significant, indicating from our current observations that baseline LD levels and the rate of decay are effectively the same across all population clusters.

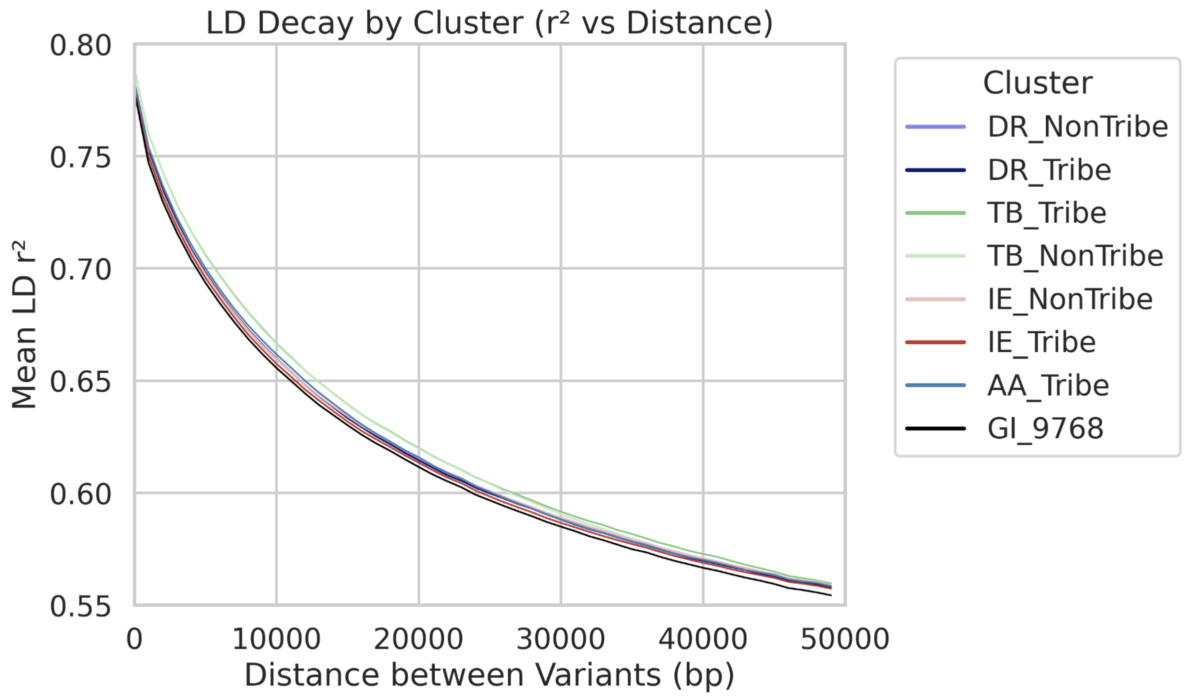

**Figure S15.2.** LD decay across seven population clusters. Mean pair-wise LD ($r^2$) is plotted against inter-marker physical distance (bins up to 50 kb) for each cluster.

#### Imputation

##### Methods

We evaluated the performance of our haplotype reference panel by performing imputation on array-based genotypes of 7677 individuals with self-reported South Asian ancestry enrolled in the UKBiobank cohort (UKB-SAS). We computed principal components from our GI populations and overlaid these 7677 target samples to assess genetic similarity between UKB-SAS and the GI population using EIGENSOFT (Price et al., 2006) (Fig S15.3).

We applied quality control filters on the array data based on sample and genotype missingness, heterozygosity, deviation from hardy weinberg equilibrium, and minor allele frequency. This target genotyping data was also phased using SHAPEIT5 in a similar fashion prior to imputation. Genotype imputation was performed using the phased GenomeIndia haplotypes as a reference panel. We performed imputation considering overlapping chunks of size 5Mbp using the IMPUTE5 (Rubinacci et al., 2020) software. To ensure that the imputed variants reflect the correct population-level frequencies, we compared the reference allele frequencies between the target population and the reference population and assess the concordance allele frequencies (Figure S15.4).

To visualize the completeness and accuracy of the imputed genotypes, we also generated a coverage plot that displays the proportion of imputed genotypes across low and common variants against the INFO scores (Figure S15.5).

To perform a comparative analysis on the performance of the newly created GI reference panel, we also carried out imputation with three different widely known haplotype reference panels- HRC (Haplotype Reference Consortium), TOPMed, and Genome Asia. The composition of these panels is described in Table S15.1.

**Table S15.1.** *Composition of the haplotype reference panels*

| Panel | Samples | Sites | Ancestry | Panel content |
|---|---|---|---|---|
| GI (GRCh38) | 9,772 | 129,929,266 | Indians | Chr1-22 |
| TOPMED (GRCh38) | 133,597 | 445,600,184 | Multi-ethnic | Chr1-22 and X SNPs and INDELs (No singletons) |
| HRC (GRCh37) | 32,470 | 39,635,008 | Mostly European | Chr 1-22 and X; SNPs only; MAC>5 |
| Genome Asia (GRCh37) | 1,654 | 21,494,814 | Asians | Chr 1-22 Biallelic SNPs |

We compared the distribution of the number of imputed variants and INFO scores across different minor allele frequency bins - namely rare, low frequency, and common. The INFO metric is a commonly used metric that is provided as an output by IMPUTE programs. This estimates how much statistical information about the population allele frequency is retained in the imputed genotypes compared to the true genotypes (Das et al., 2018). Under Hardy-Weinberg equilibrium, it is equal to Minimac's $r^2$ measure which estimates the squared correlation coefficient between imputed and true, unobserved genotypes (https://genome.sph.umich.edu/wiki/Minimac3_Info_File#Rsq).

To investigate how allele frequency (MAF) influences the reliability of imputation, we assessed the imputation accuracy across variants in different minor allele frequency bins. Comparative analyses showed that the GI panel substantially outperformed other widely used reference panels such as GAsP, HRC, and TOPMed with an average increase in imputation quality of 55%, 27%, and 21%, across the allele-frequency spectrum. The improvement was especially pronounced for rare variants, with improvements of 93%, 36%, and 9%, and for ultra-rare variants (MAF as low as 0.00015), where the GI panel achieved striking gains of 281%, 208%, and 433%, respectively. This superior performance was maintained even when the analysis was restricted to variants imputed by all panels, with the GI panel still showing 45%, 20%, and 9% higher imputation quality across all frequency bins represented in the shared variant set, demonstrating the robustness of its performance across the allele-frequency spectrum.

To assess the overlap between imputed variants from different reference panels, we visualized the intersection sets of imputed variants. We then computed all possible pairwise and multi-panel intersections of the imputed variant sets to identify variants exclusively imputed by GI panel and variants imputed by all panels. To evaluate the consistency of imputed genotypes across different reference panels, we also plotted the $R^2$ values against MAF bins for each variant shared across the panels (refer to Fig 5C,D-main text).

In addition to the imputation quality metrics reported by the software, we assessed imputation accuracy by calculating aggregate $R^2$ defined as the squared Pearson correlation between imputed genotype dosages and hard-called WGS genotypes which we have considered as the ground-truth dataset. Aggregate $R^2$ values were then plotted against minor allele frequencies. (Fig S15.6).

Next, we visualized how imputation accuracy varied across the genome by exploring the relationship between $R^2$ (imputation accuracy) and genomic coordinates. We carried out this analysis for all four panels and plotted accuracy vs genomic coordinates (Fig. S15.7).

Next, to assess the accuracy of imputed genotypes at the individual level, we considered imputed genotypes of a subset of 7628 individuals for whom WGS-based genotypes were present. We compared imputed genotypes and WGS-based genotypes, using identity-by-state (IBS) DST metric using plink1.9 (Purcell et al., 2007) (Fig. S15.8).

To evaluate variant-level imputation performance, we considered the WGS-based genotypes as "true" genotypes and the imputed genotypes as "observed" ones. We then performed a multi-class classification and generated per variant sensitivity specificity and precision in the manner specified in Fig S15.M0 given below.

```
#============================================
#Actual(b)            Imputed(a)
#--------------------------
#mat=               0|0      0|1 or 1|0       1|1
#                   ------------------------------
#        0/0        0,0      0,1              0,2
#        0/1        1,0      1,1              1,2
#        1/1        2,0      2,1              2,2
#
#============================================
```

```
TP=(0,0),       FN=(0,1)+(0,2), FP=(1,0)+(2,0), TN=(1,1)+(1,2)+(2,1)+(2,2),
TP=(1,1),       FN=(1,0)+(1,2), FP=(0,1)+(2,1), TN=(0,0)+(0,2)+(2,0)+(2,2)
TP=(2,2),       FN=(2,0)+(2,1), FP=(0,2)+(1,2), TN=(0,0)+(0,1)+(1,0)+(1,1)
```

**Figure S15.M0**. This schema represents a confusion matrix, where the leftmost column outlined in pink represents the actual/true WGS-derived genotypes (0/0,0/1,1/1- these represent homozygous REF, heterozygous of homozygous ALT genotypes). The header outlined in orange represents the phased genotypes as obtained after genotype imputation. The green box represents a 3x3 matrix with each cell representing the index of that cell.

For each variant, we thus calculated $TP_C$ (true positives), $FN_C$ (false negative), $FP_C$ (false positive), and $TN_C$ (true negative) for each class C (=0/0,0/1, and 1/1) (represented in blue box, in respective order). We then computed micro average using the following formula

Sensitivity=$\sum_C \frac{TP_C}{TP_c+FN_C}$

Specificity= $\sum_C \frac{TN_C}{TN_c+FP_C}$

Precision=$\sum_C \frac{TP_C}{TP_c+FP_C}$

We collapsed all variants into a single confusion matrix and then calculated the overall metrics. We obtained plots illustrating sensitivity, specificity, and precision (y-axis) across imputation accuracy (x-axis) shaded according to the average minor allele frequency (MAF) to assess the performance of our reference panel at variant level (Refer to Fig 5E in main text).

To evaluate the reliability of our GI reference panel in imputing genotypes across genetically close yet distinct South Asian subgroups, we analyzed imputation accuracy stratified by population differentiation (FST quartiles) relative to the GI panel (Fig S15.9). Variants from the 1000 Genomes (Auton et al., 2015) SAS subpopulations were downsampled to match those present on our Affy array. The subpopulations were grouped into four categories: BEB, STU-ITU, PJL-GIH, and the broader SAS population. Additionally, array-based genotypes from the UKB-SAS cohort were included. FST was calculated using Hudson's method as implemented in PLINK2, based on the set of overlapping variants between the GI reference panel and each subpopulation. To ensure independence among variants, linkage disequilibrium pruning was performed with a window size of 50, step size of 5, and an $r^2$ threshold of 0.2.

**Results**

We inferred the haplotype structure using 9,768 whole-genome sequenced Indian samples with 70,746,809 variants having a minor allele count (MAC) ≥ 2. We utilized the data on 244 trios to filter out 9,895 variants with a Mendelian Inheritance Error rate >0.5%. We then phase the haplotypes consisting of the remaining 70,736,914 variants, while accounting for these 244 related samples. At first, we phased the variants with MAF >0.1% and then used the resulting phased haplotypes as a scaffold to phase the rarer variants. We observed an approximate switch error rate of ~0.5% which is indicative of accurate phasing of the haplotypes. We then selected variants with a MAC ≥ 3 to construct our haplotype reference panel for imputation.

We performed genotype imputation on a target sample of 7767 UKB-SAS individuals. Before proceeding to genotype imputation, we observed that these target samples overlapped majorly with the IE nontribe and DR_NonTribe populations, and were broadly reflective of the diversity of the GI population (Fig S15.3).

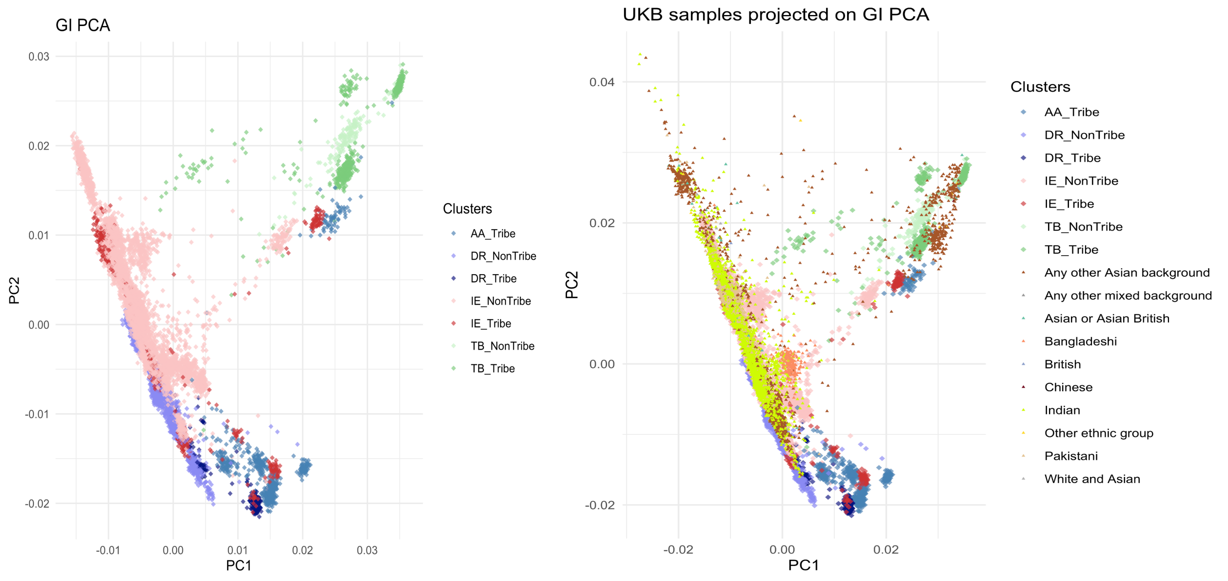

**Figure S15.3**. Principal Component Analysis of genetic variation in the GI panel. 7677 individuals have been projected onto the PCA space defined by the GI reference panel to visualize population structure and genetic similarity without influencing the principal components.

After applying quality control filters, we retained high-quality genotype data for approximately 400,000 typed genetic variants which will serve as the anchor for obtaining imputed haplotypes and consequently to obtain the untyped variants. Pre-imputation diagnostics reveal 99.4% concordance between the non-REF allele frequency between the GI panel and the target 7767 array-based genotypes (Fig S15.4).

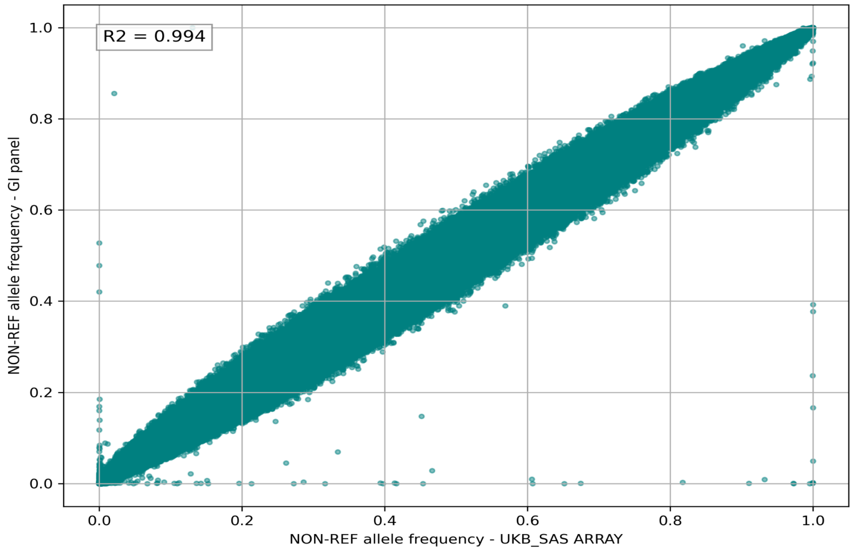

**Figure S15.4**. Concordance in non-REF allele frequency between the array-derived genotypes of UKB-SAS and the GI reference panel

As expected, imputation completeness was positively correlated with INFO score across minor allele frequency bins (Fig S15.5). For common variants, over 95% of sites achieved INFO scores ≥ 0.8, indicating high-quality imputation and near-complete genotype recovery. Low-frequency variants showed greater variability in imputation quality, with some proportion falling below the INFO ≥ 0.8 threshold. Nonetheless, approximately >80% of low-frequency variants still achieved INFO scores ≥ 0.8, suggesting moderate-to-high reliability in these regions. The coverage plot shows a modest declining pattern in the proportion of well-imputed genotypes (INFO ≥ 0.8) in lower MAF bins, reflecting the expected difficulty of imputing rarer variants due to limited haplotype diversity. Nonetheless, our panel maintains relatively high imputation quality for these variants, demonstrating robust performance even in low-frequency regions.

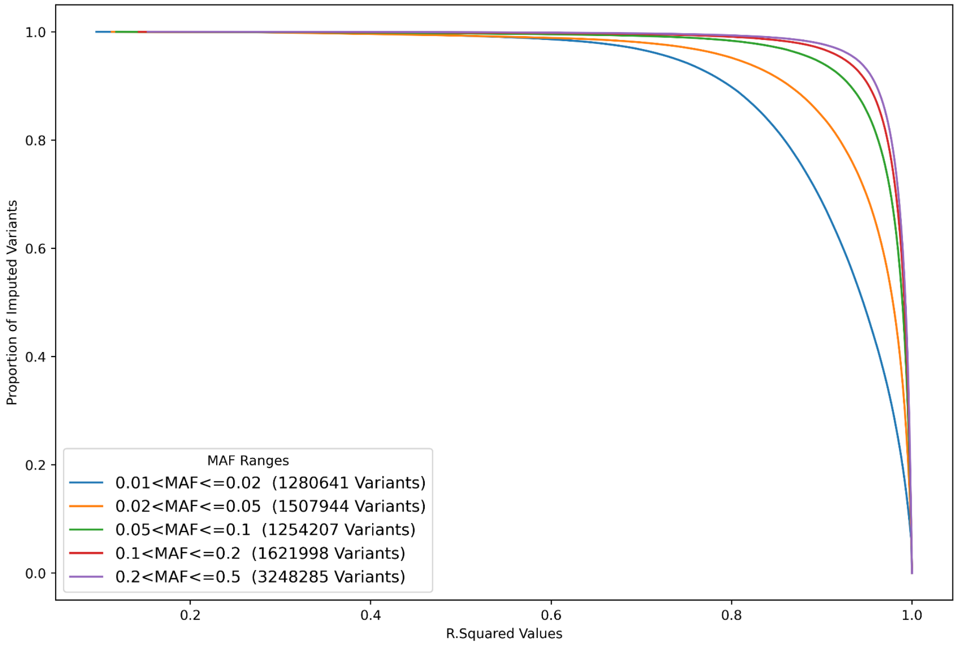

**Figure S15.5.** Coverage plots illustrating the proportion of high-quality variants in relation to their mean imputation accuracy, across gradients of low-frequency to common variants (represented in different colors)

Aggregate $R^2$ analysis showed that GI consistently outperformed HRC and GASP across the allele-frequency spectrum. TOPMed is well known to demonstrate superior performance due to its larger number of haplotypes and broader representation of rare alleles (up to minor allele count 2), which allows more accurate haplotype copying (O'Connell et al., 2021; Sengupta et al., 2023). However, we observe that GI achieves comparable performance to TOPMed for variants with MAF ≥ 1%, while maintaining superior accuracy for rarer variants down to a minor allele frequency of 0.1% (Fig S15.6).

Differences between aggregate $R^2$ and software-derived imputation quality metrics are expected and reflect fundamental differences in their definitions: software-derived metrics provide model-based estimates of imputation uncertainty derived from posterior genotype variances, whereas aggregate $R^2$ is an empirical measure of concordance between imputed genotype dosages and sequencing (usually regarded as true) hard-called WGS genotypes. However, in the absence of ground-truth sequencing data, software-derived imputation quality metrics remain the standard for assessing variant reliability for downstream association analyses. Accordingly, the relative performance that we observed using the software-derived metrics in the main Figure 5C is also reflected in Fig. S15.6 given below.

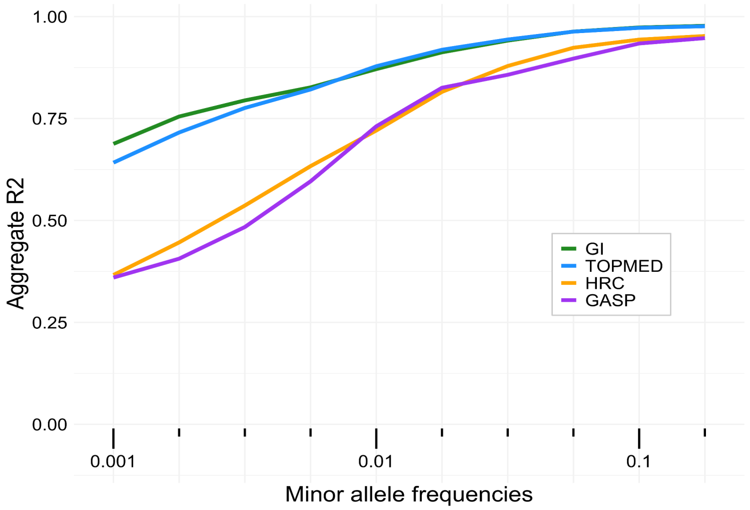

**Figure S15.6.** Aggregate $R^2$ stratified by minor allele frequency for the overlapping variant set imputed by all panels, the corresponding WGS-derived genotypes for which were available.

The genome-wide plots illustrating the relationship between imputation quality and genomic coordinates revealed consistently high imputation accuracy across most regions for the GI panel with median accuracy values exceeding 0.85 (Fig S15.7). Only a few genomic intervals exhibited localized reductions in accuracy which may warrant cautious interpretation in downstream analyses. Notably, this panel consistently outperformed its closest competitor, the GASP panel, across the genome, demonstrating higher accuracy values (at least 30% higher) even in small pockets regions where accuracy from all other panels tend to decline.

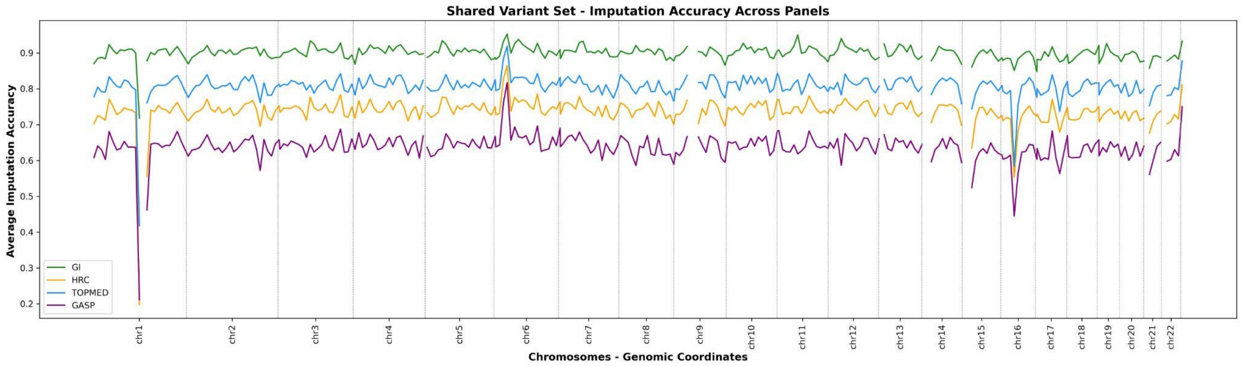

**Figure S15.7.** Distribution of imputation accuracy across genomic coordinates, when imputed to GI, TOPMed, HRC and GASP panels. GI consistently outperforms all panels across all coordinates, even with a few local drops in performance at specific intervals

At the individual level, we observed that mean pairwise identity-by-state (IBS) measure, DST > 0.99, between WGS and imputed genotypes (Figure S15.7). Using WGS-called genotypes as the ground truth, the imputed genotypes showed an overall sensitivity of 99.75%, specificity of 99.88%, and precision of 99.75%. At the variant level, most variants showed high INFO scores and high sensitivity and specificity (Figure 5E). Both rare and common variants are imputed accurately, with common variants (lighter colors) generally displaying high sensitivity and specificity. Only a small subset—mainly rare variants (darker colors at the bottom right part of each panel)—exhibit lower metrics (sensitivity,precision <20%; precision <60%).

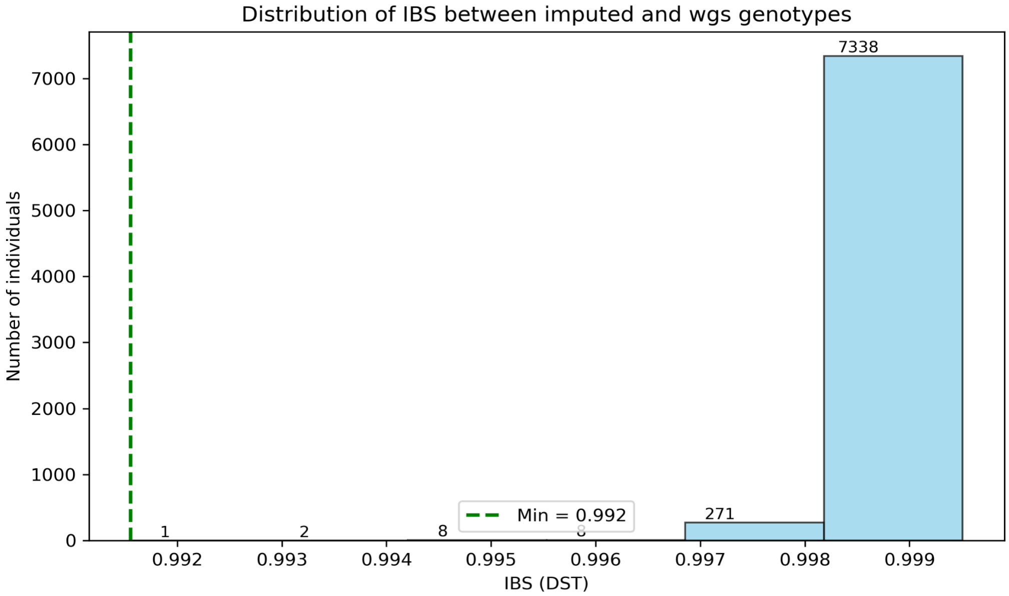

**Figure S15.8**. Accuracy of imputed genotypes at the individual level as assessed by Identity by state

Our analysis of imputation accuracy (INFO scores) across population differentiation (FST quartiles) in South Asian populations reveals important nuances (Fig S15.9). While UKB-SAS shows improved imputation accuracy with increased differentiation despite a lower proportion of rare variants, the 1000 Genomes SAS populations exhibit decreasing accuracy as differentiation rises, even with variable rare variant proportions. This indicates that MAF acts as a confounding factor: rare variants generally have lower imputation accuracy, but their distribution varies across FST quartiles. Therefore, imputation performance can depend on the complex interplay of both the allele frequency spectrum and the genetic similarity between the reference panel and target populations. Overall, the observed consistency of imputation performance across groups with modest genetic differentiation suggests that the GI reference panel effectively captures shared variation across South Asian subpopulations. However, given the relatively narrow FST range observed, it is difficult to draw definitive conclusions about this trend. Future studies with larger, well-characterized cohorts from individual subpopulations are essential to validate and refine imputation performance and genetic differentiation measures at finer scales.

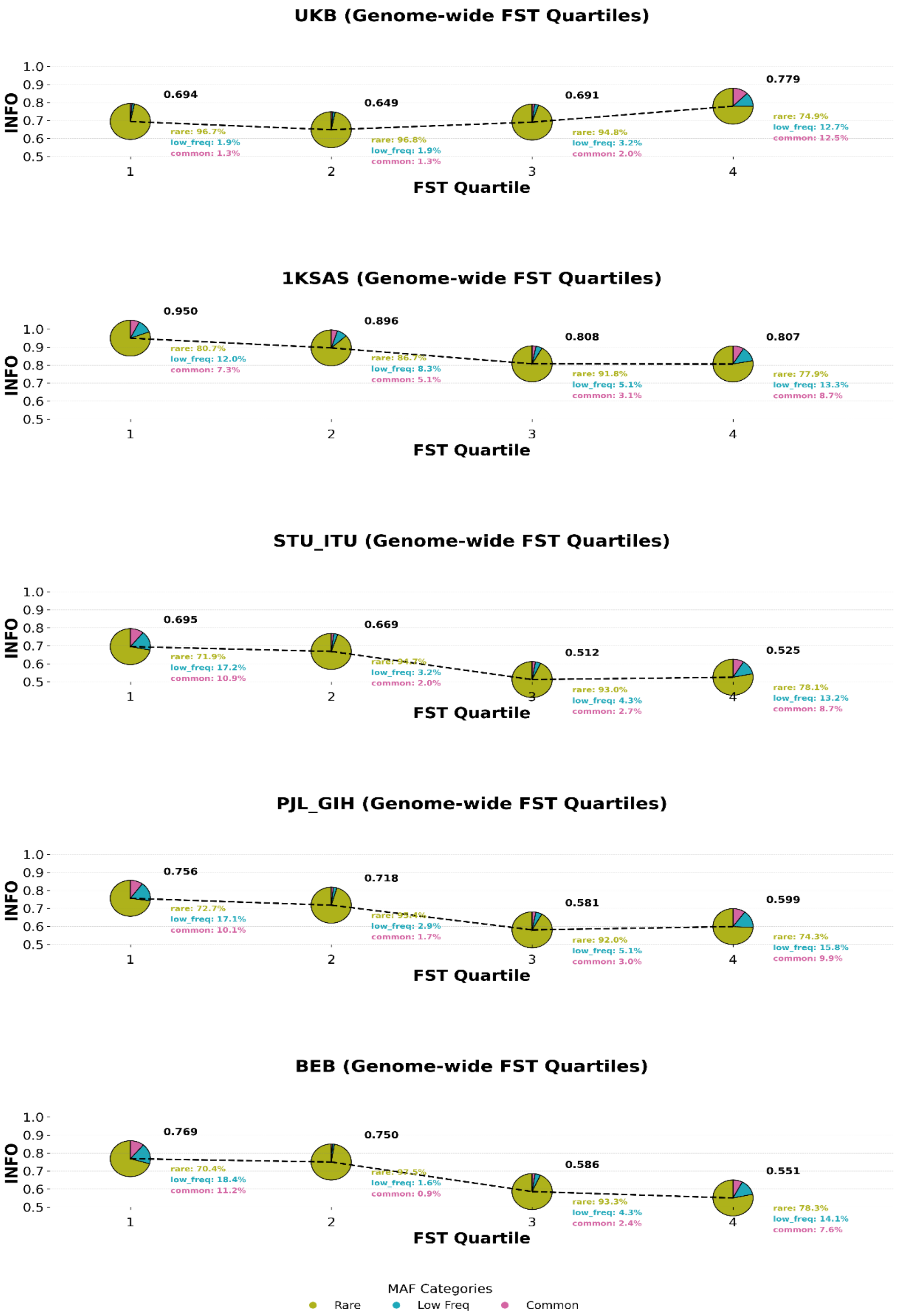

**Figure S15.9**. Imputation accuracy (INFO scores) across population differentiation (FST quartiles) for South Asian populations. UKB-SAS array data, and 1000 Genomes SAS along with its subpopulations (STU-ITU, PJL-GIH, BEB), all downsampled to array data, were imputed using the GI reference panel. Abbreviations: STU - Sri Lankan Tamil in the UK; ITU - Indian Telugu in the UK; PJL - Punjabi in Lahore, Pakistan; GIH - Gujarati Indians in Houston, Texas, USA; BEB: Bengali in Bangladesh. The pie charts overlaid on the figure depict the percentage composition of rare, low-frequency, and common variants within each FST quartile bin. The centres of the pie charts represent the mean imputation accuracy per bin. The dotted line connecting the centers indicates the overall trend.

Together, these results support the broader applicability of the GI reference panel across South Asian subpopulations, providing a practical and efficient solution for imputation in regions where population-specific reference datasets remain limited.
