## Supplementary Note 1 for "An Atlas of Indian Genetic Diversity"

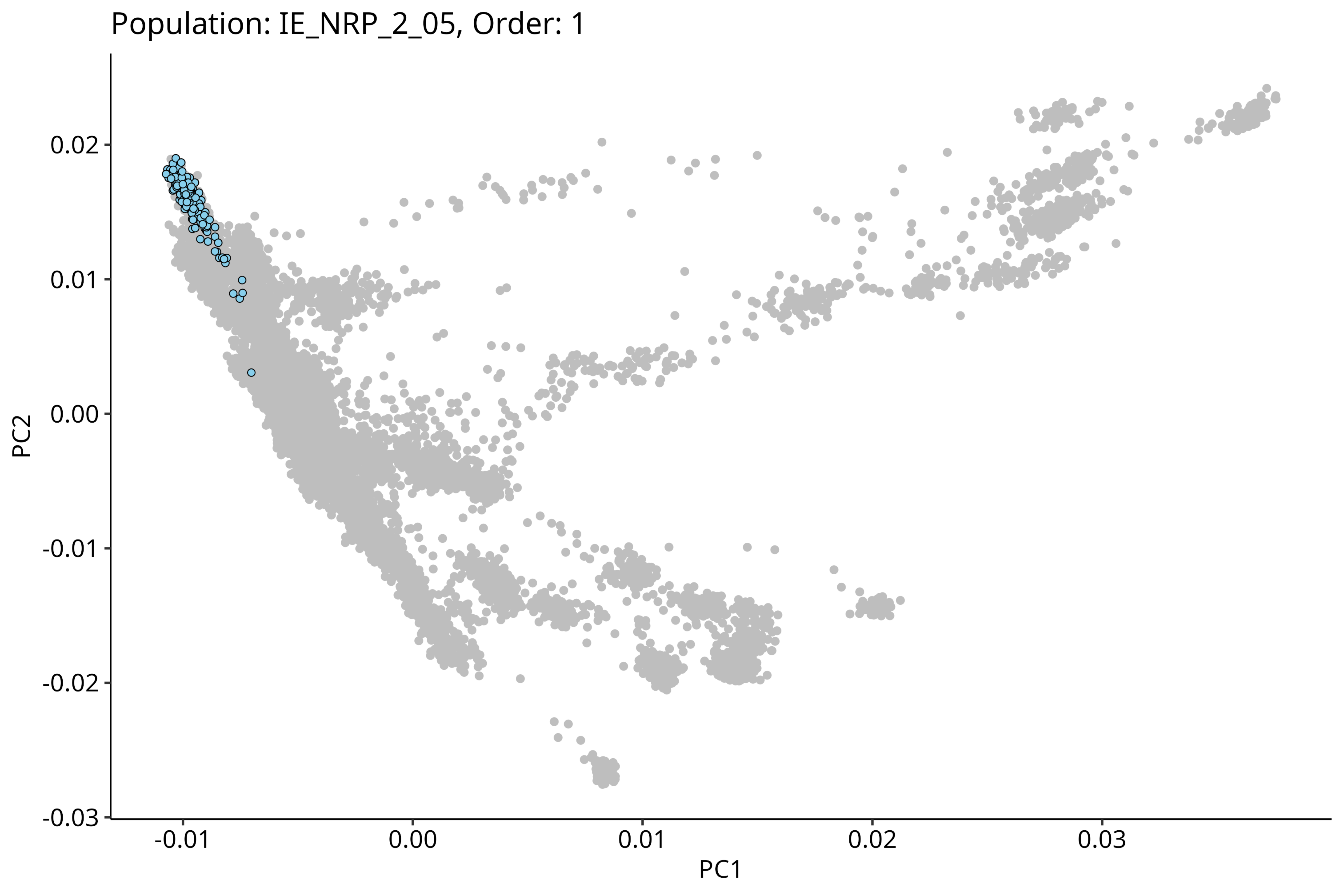

Population: IE_NRP_2_05, Order: 1
PC2
0.02
0.01
0.00
-0.01
-0.02
-0.03
-0.01
0.00
0.01
0.02
0.03
PC1

Population: IE_NRP_2_08, Order: 2

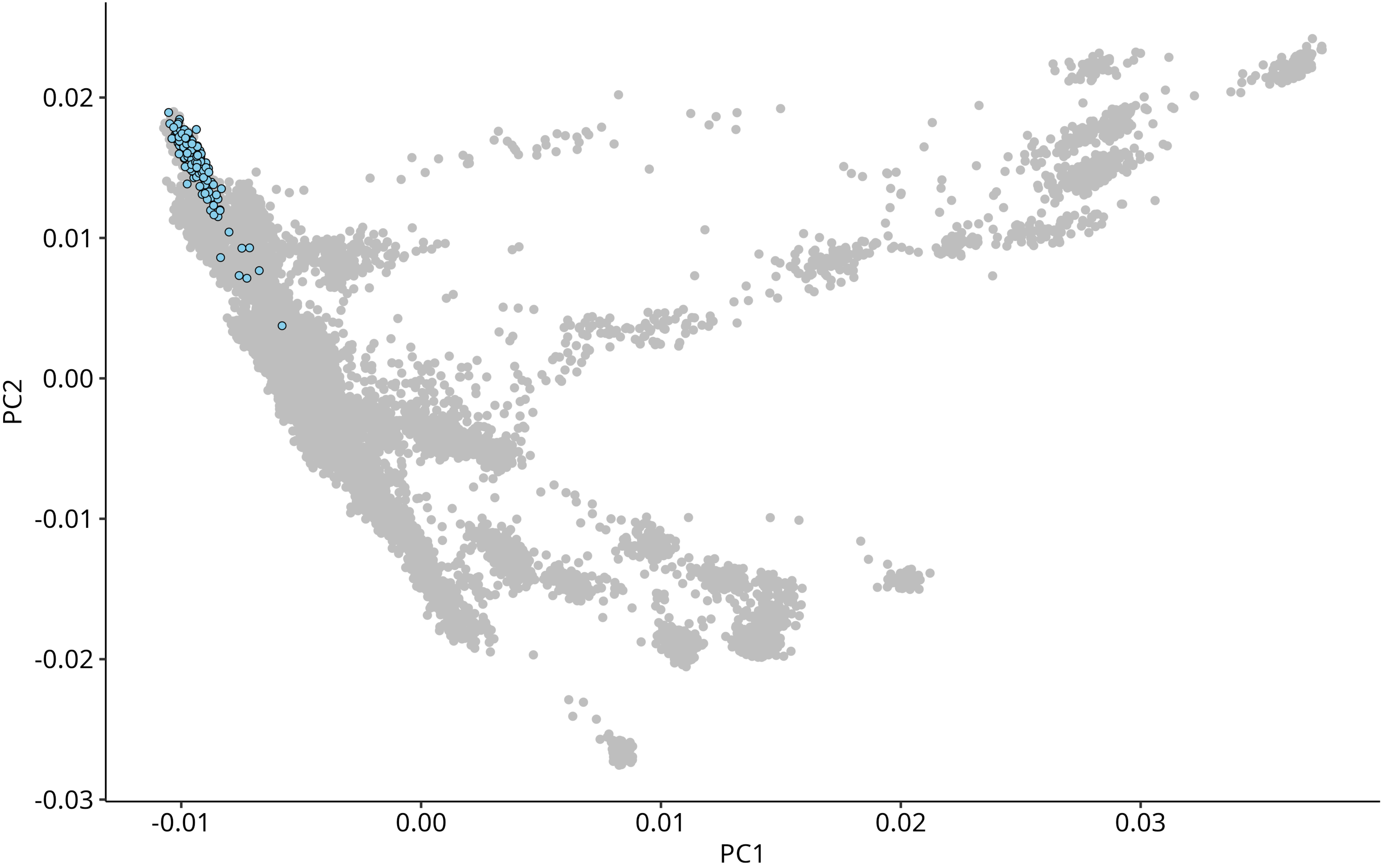

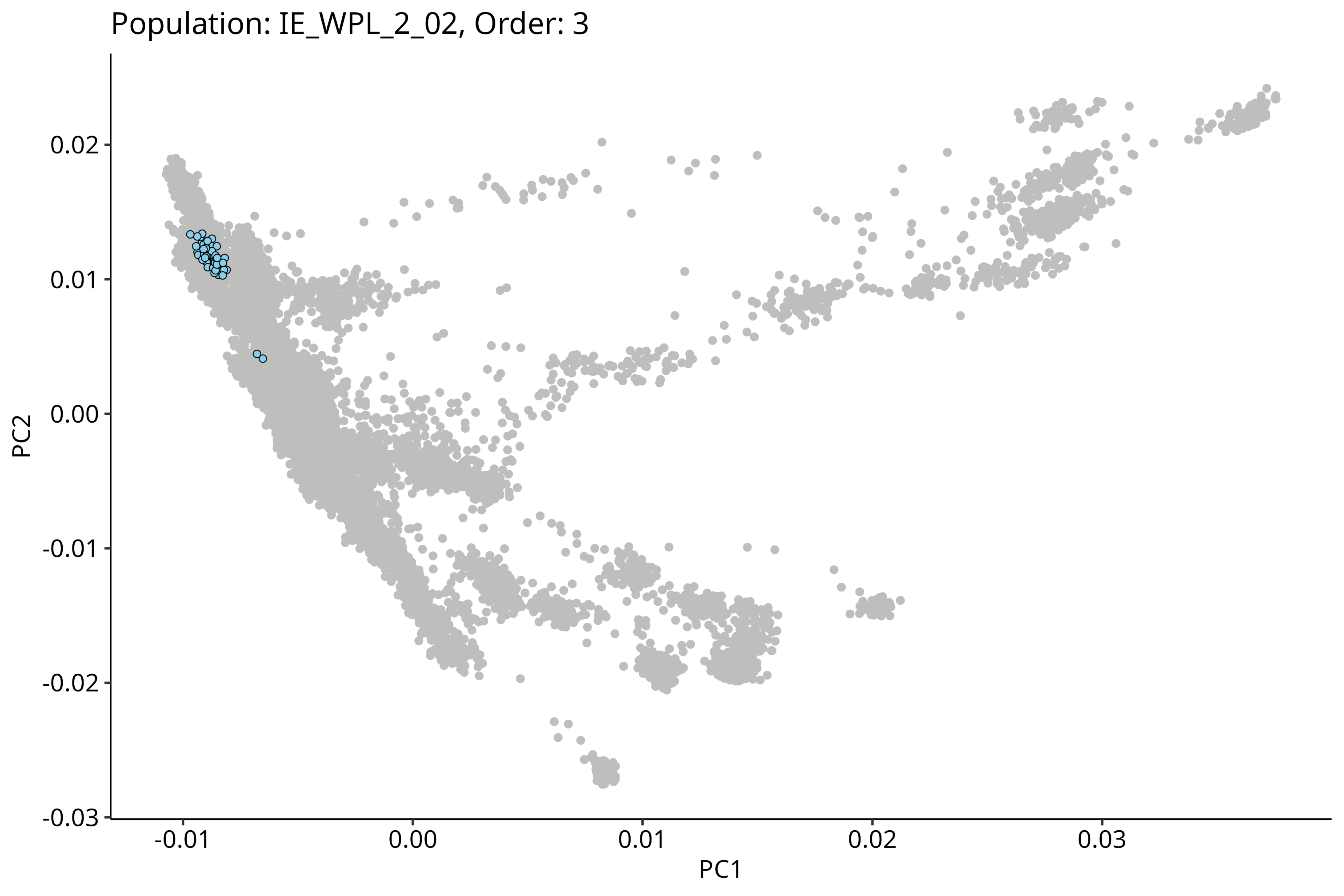
Population: IE_WPL_2_02, Order: 3
0.02
0.01
0.00
-0.01
-0.02
-0.03
PC2
-0.01
0.00
0.01
0.02
0.03
PC1

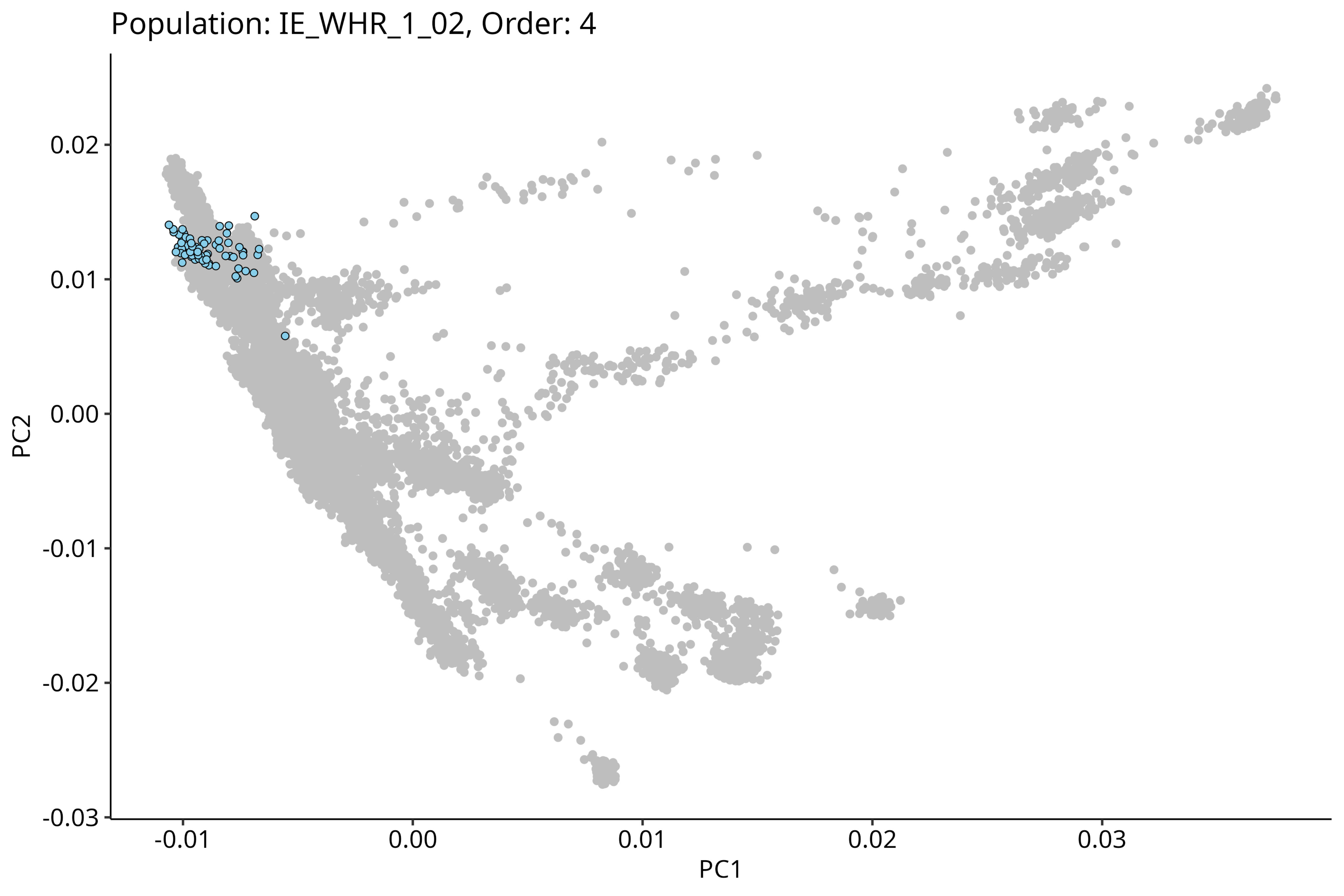
Population: IE_WHR_1_02, Order: 4
PC2
PC1
0.02
0.01
0.00
-0.01
-0.02
-0.03
-0.01
0.00
0.01
0.02
0.03

Population: IE_WHR_2_04, Order: 5

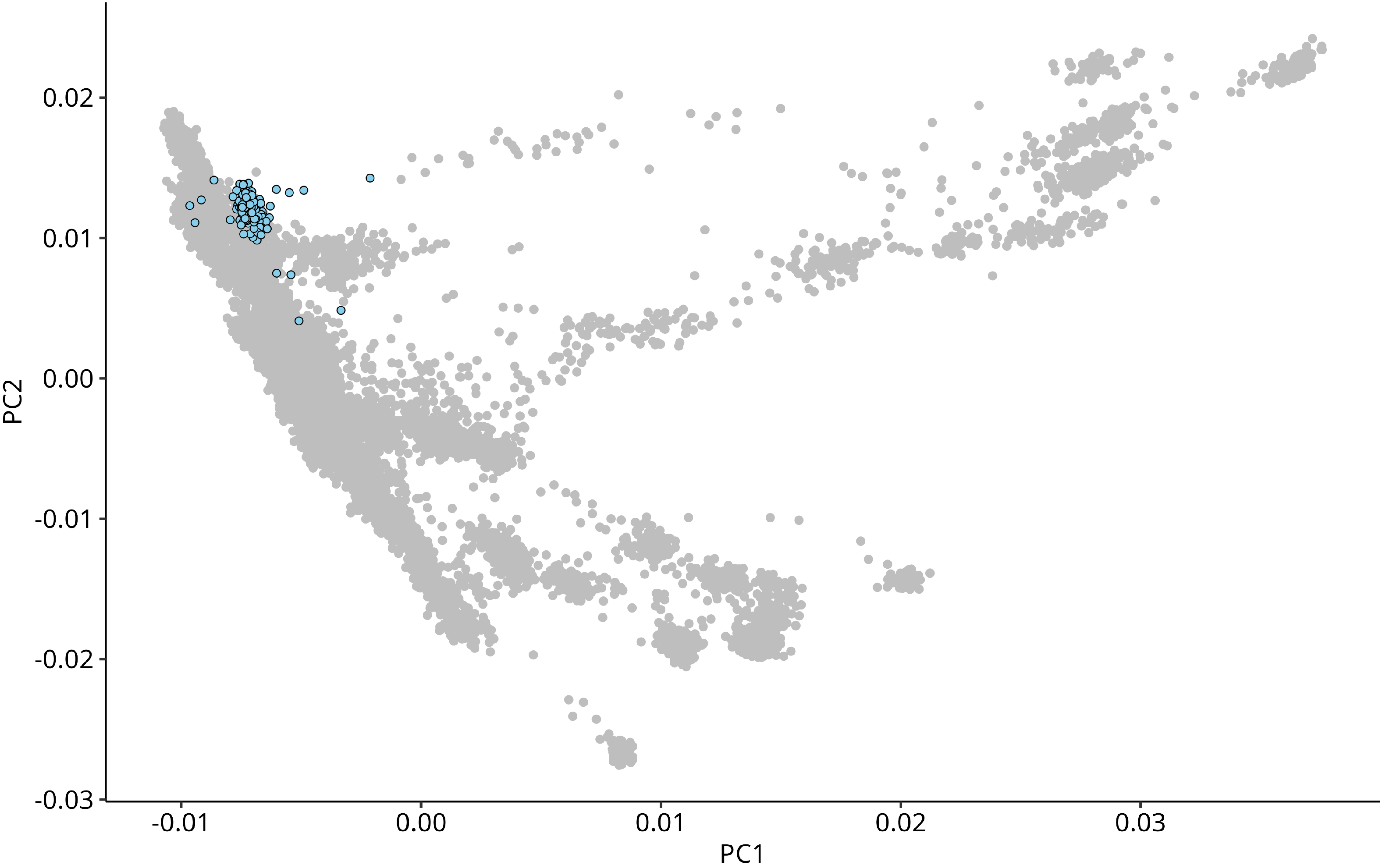

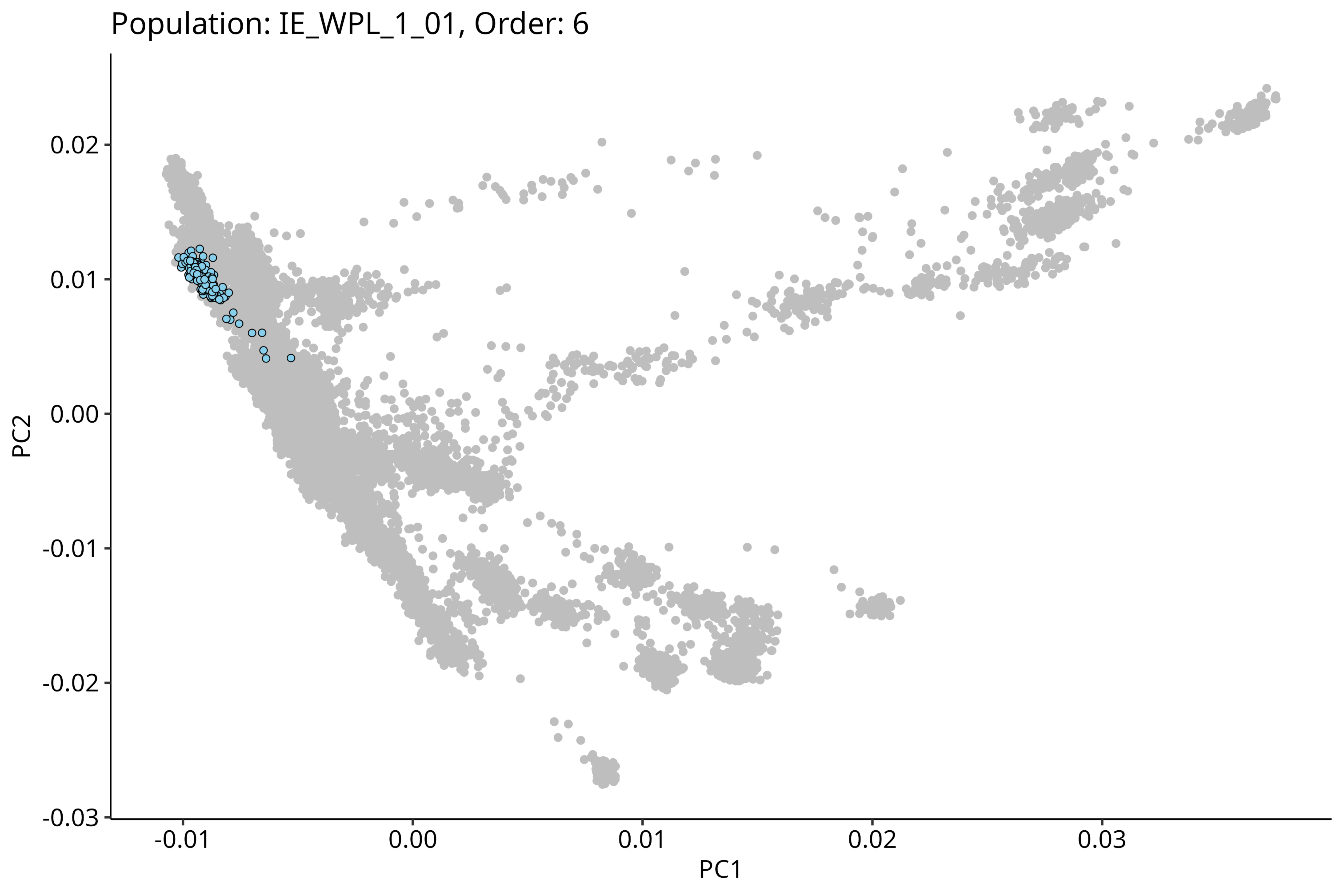
Population: IE_WPL_1_01, Order: 6
0.02
0.01
0.00
-0.01
-0.02
-0.03
PC2
-0.01
0.00
0.01
0.02
0.03
PC1

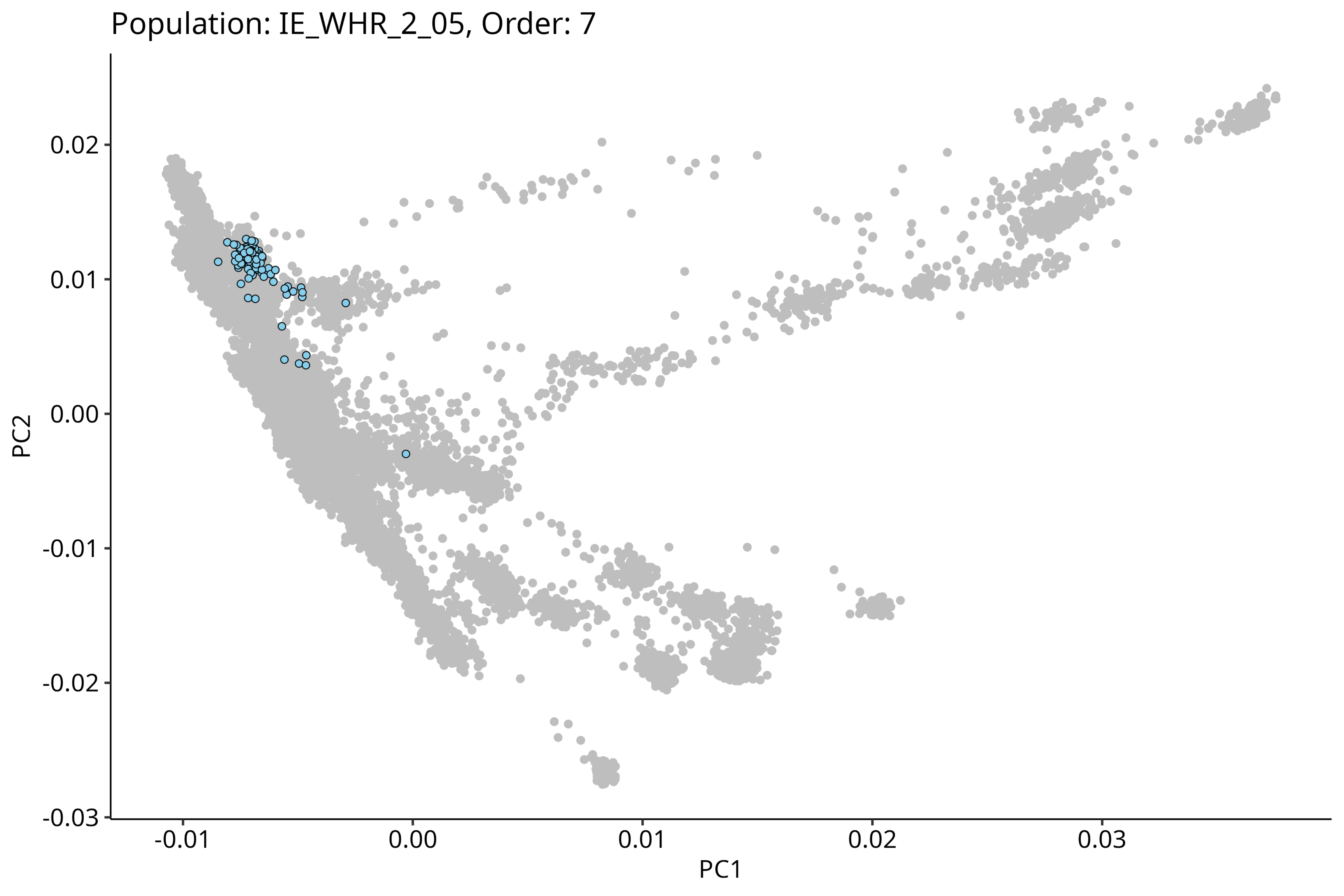

Population: IE_WHR_2_05, Order: 7
PC2
0.02
0.01
0.00
-0.01
-0.02
-0.03
-0.01
0.00
0.01
0.02
0.03
PC1

Population: IE_NRP_2_02, Order: 8

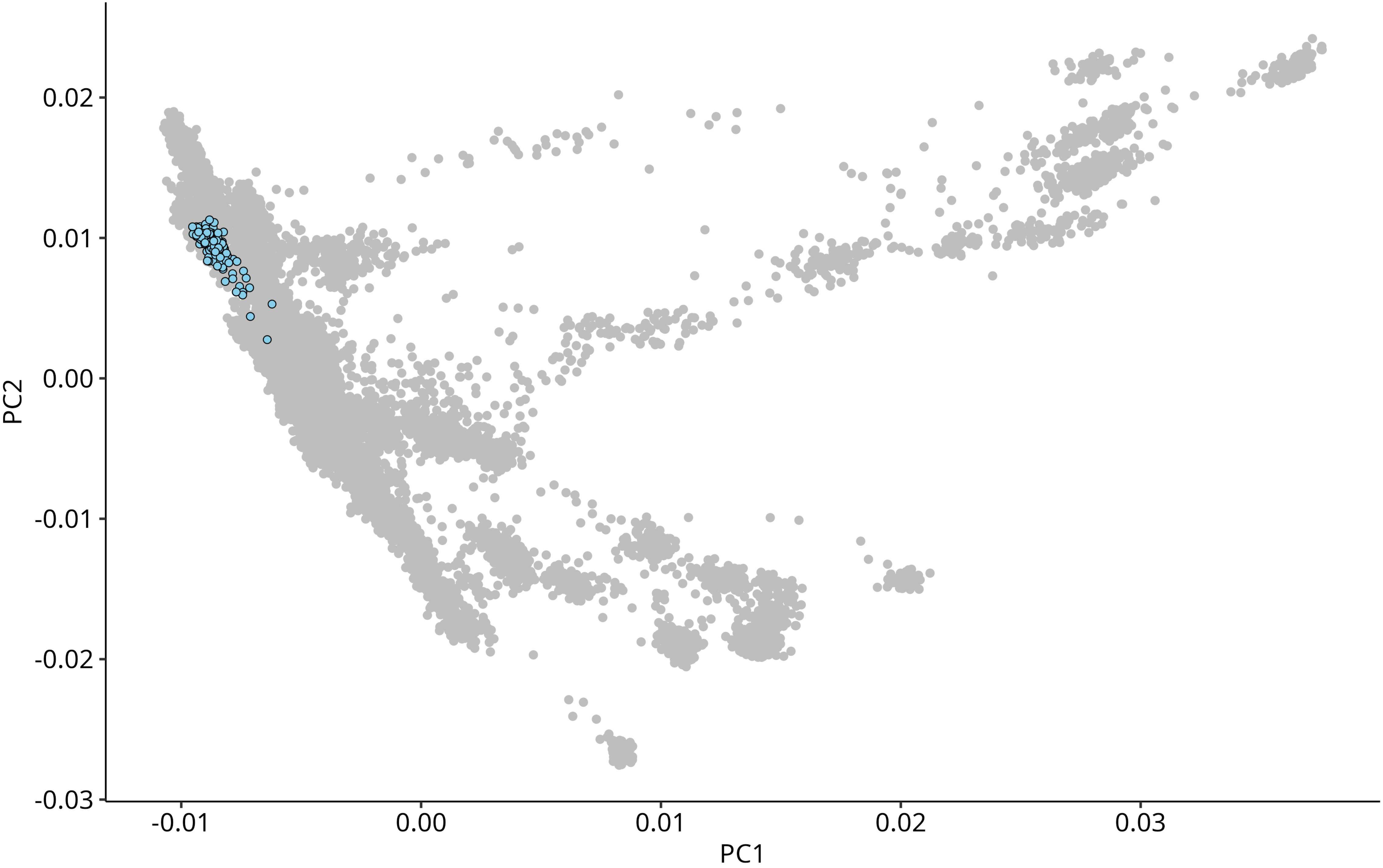

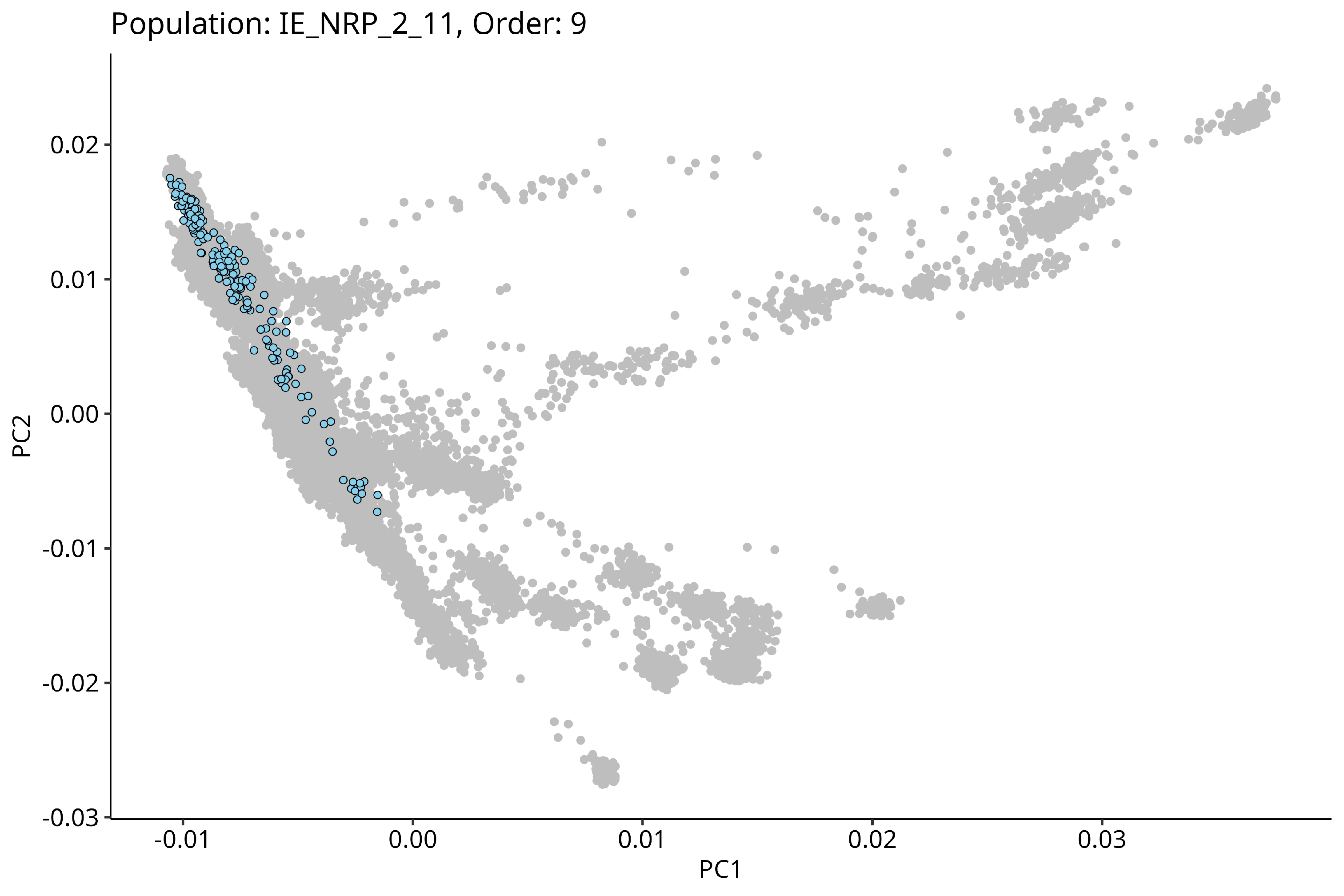
Population: IE_NRP_2_11, Order: 9
PC2
0.02
0.01
0.00
-0.01
-0.02
-0.03
PC1
-0.01
0.00
0.01
0.02
0.03

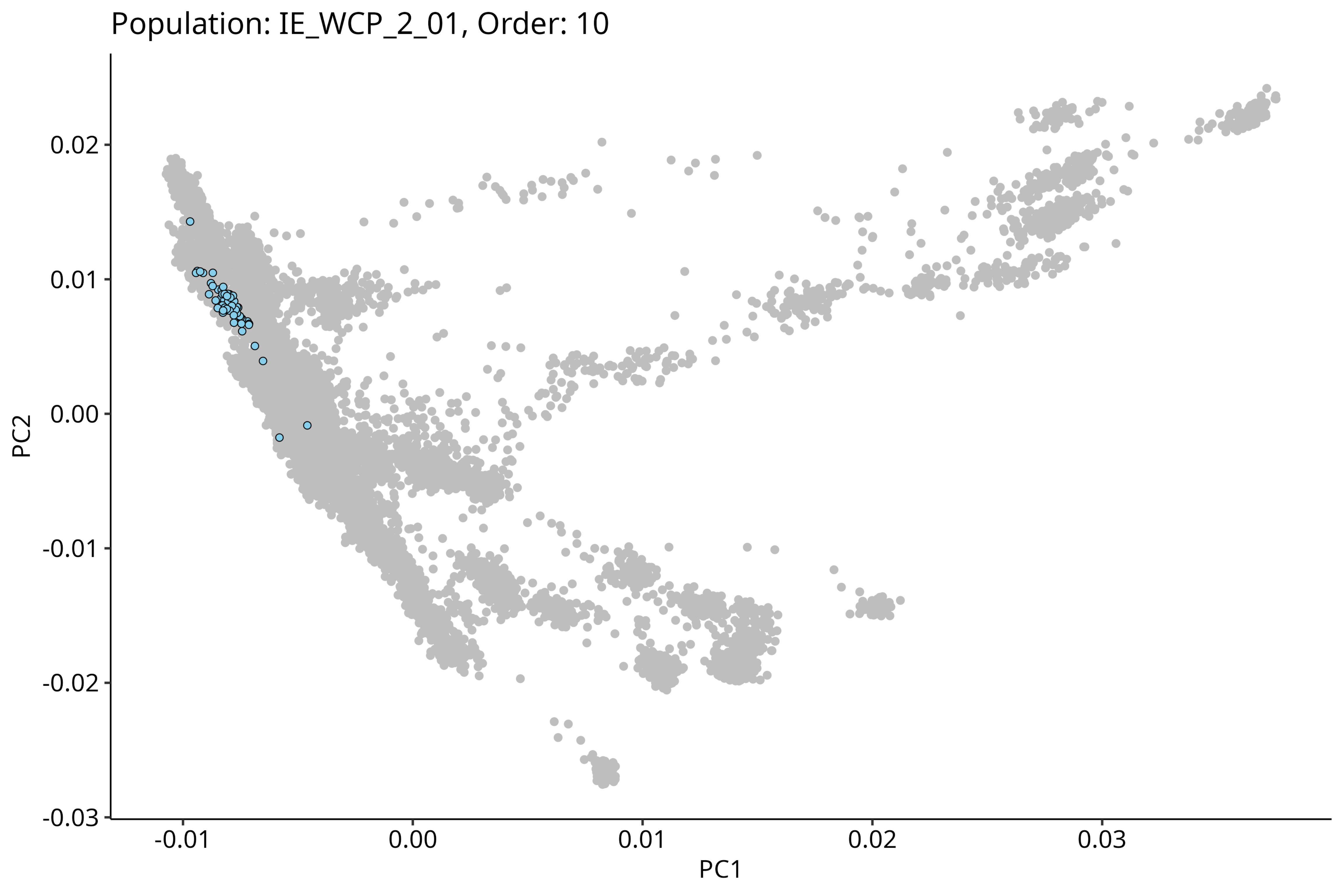
Population: IE_WCP_2_01, Order: 10
0.02
0.01
0.00
-0.01
-0.02
-0.03
PC2
-0.01
0.00
0.01
0.02
0.03
PC1

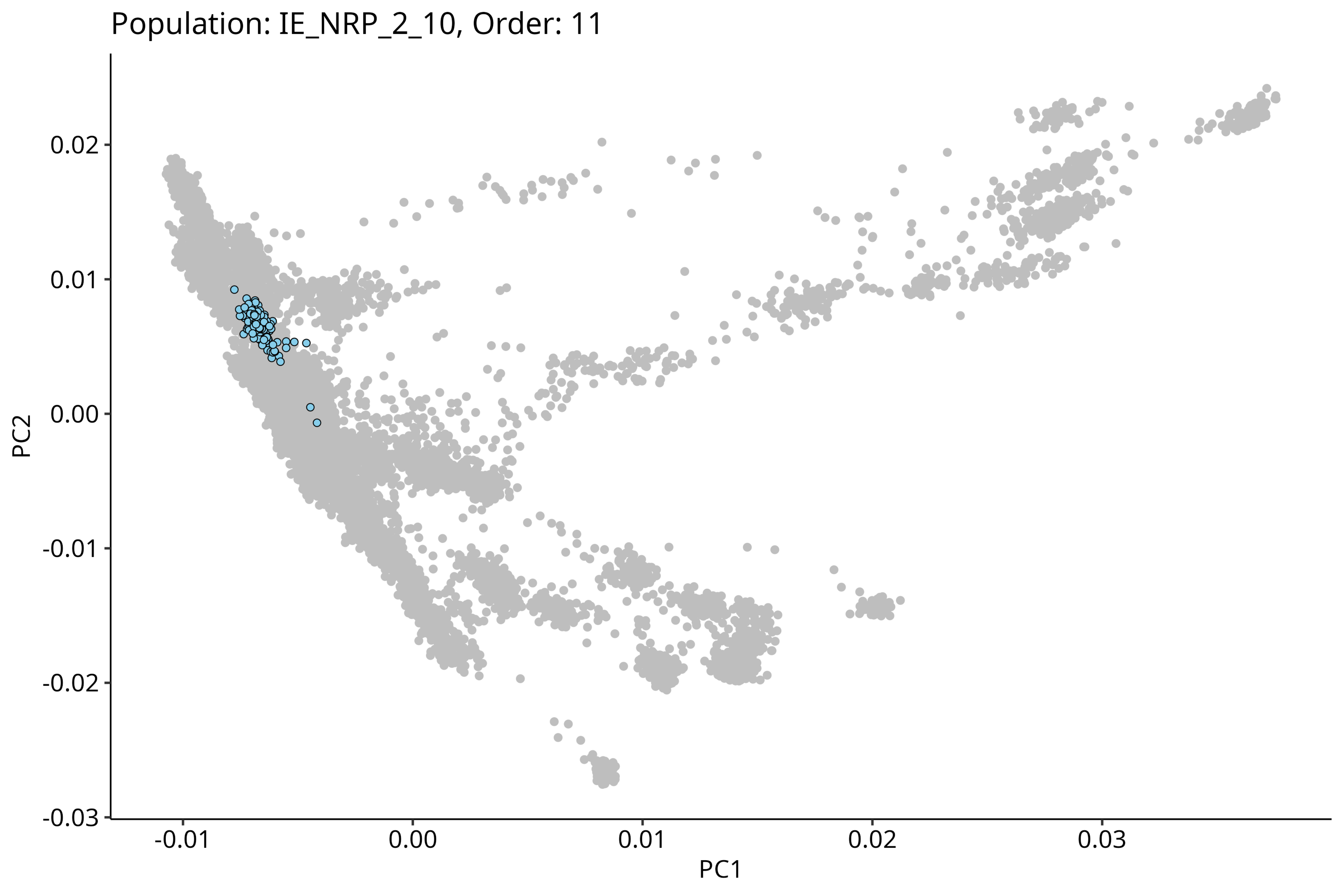
Population: IE_NRP_2_10, Order: 11
PC2
0.02
0.01
0.00
-0.01
-0.02
-0.03
PC1
-0.01
0.00
0.01
0.02
0.03

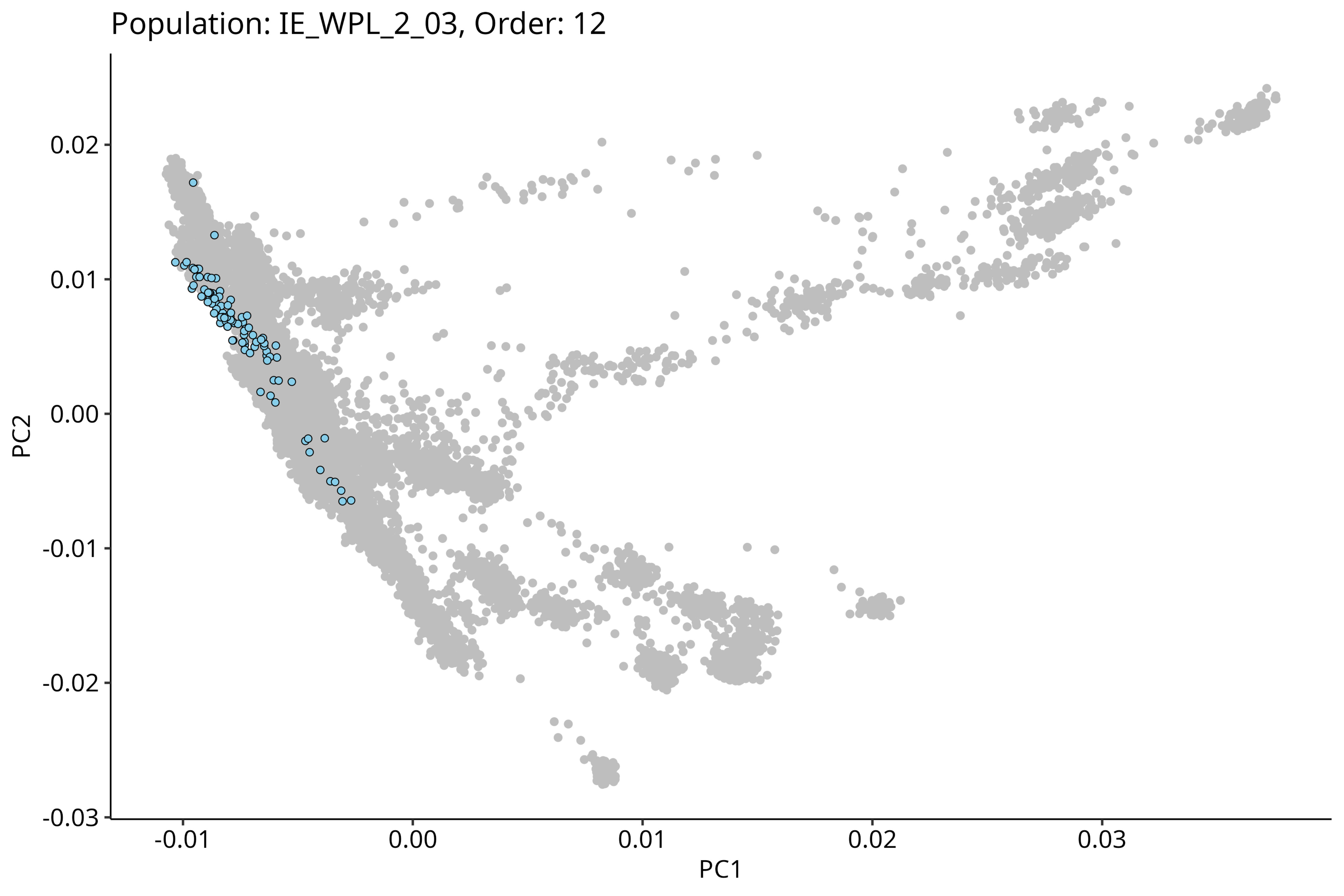
Population: IE_WPL_2_03, Order: 12
0.02
0.01
0.00
-0.01
-0.02
-0.03
PC2
-0.01
0.00
0.01
0.02
0.03
PC1

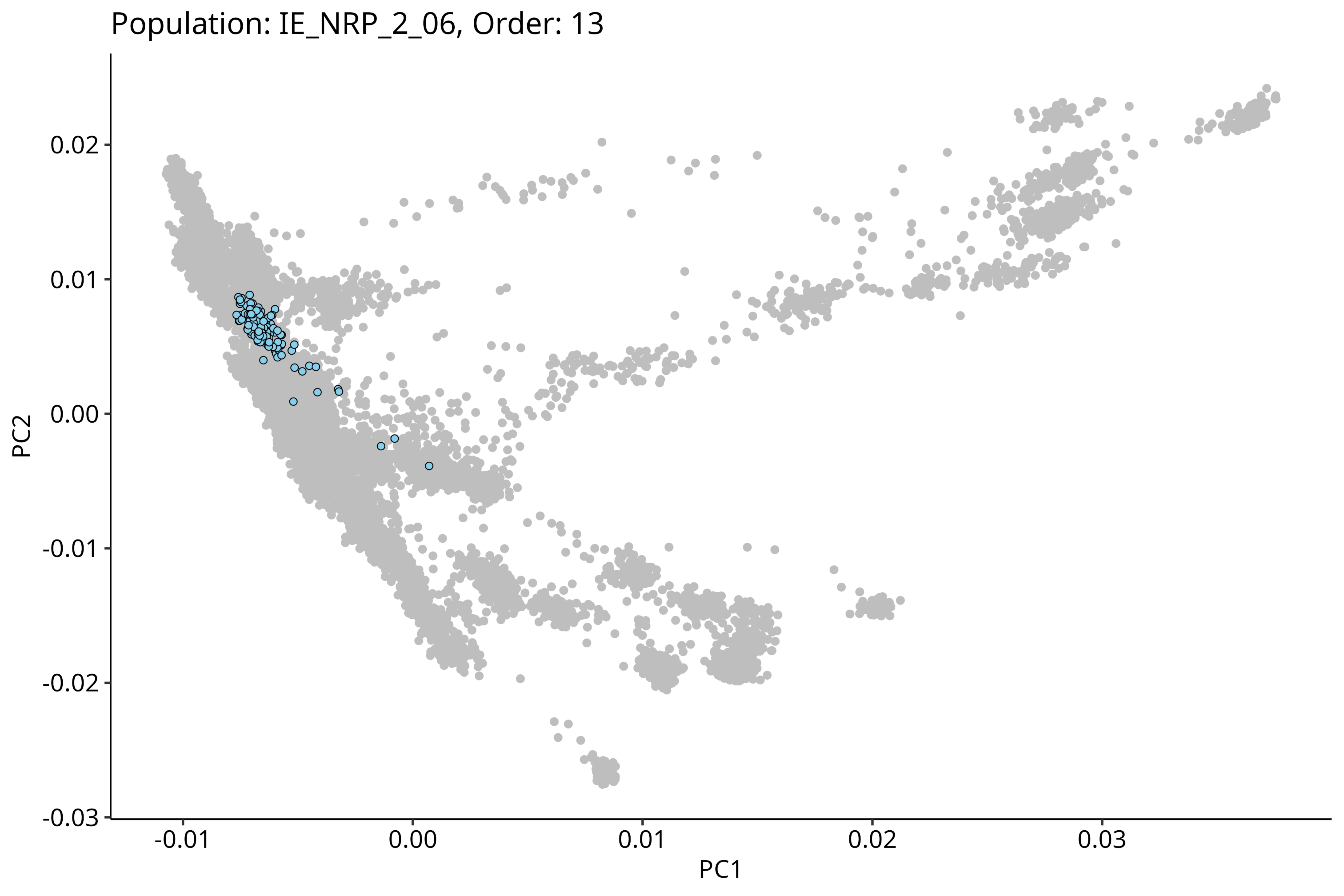

Population: IE_NRP_2_06, Order: 13
0.02
0.01
0.00
-0.01
-0.02
-0.03
PC2
-0.01
0.00
0.01
0.02
0.03
PC1

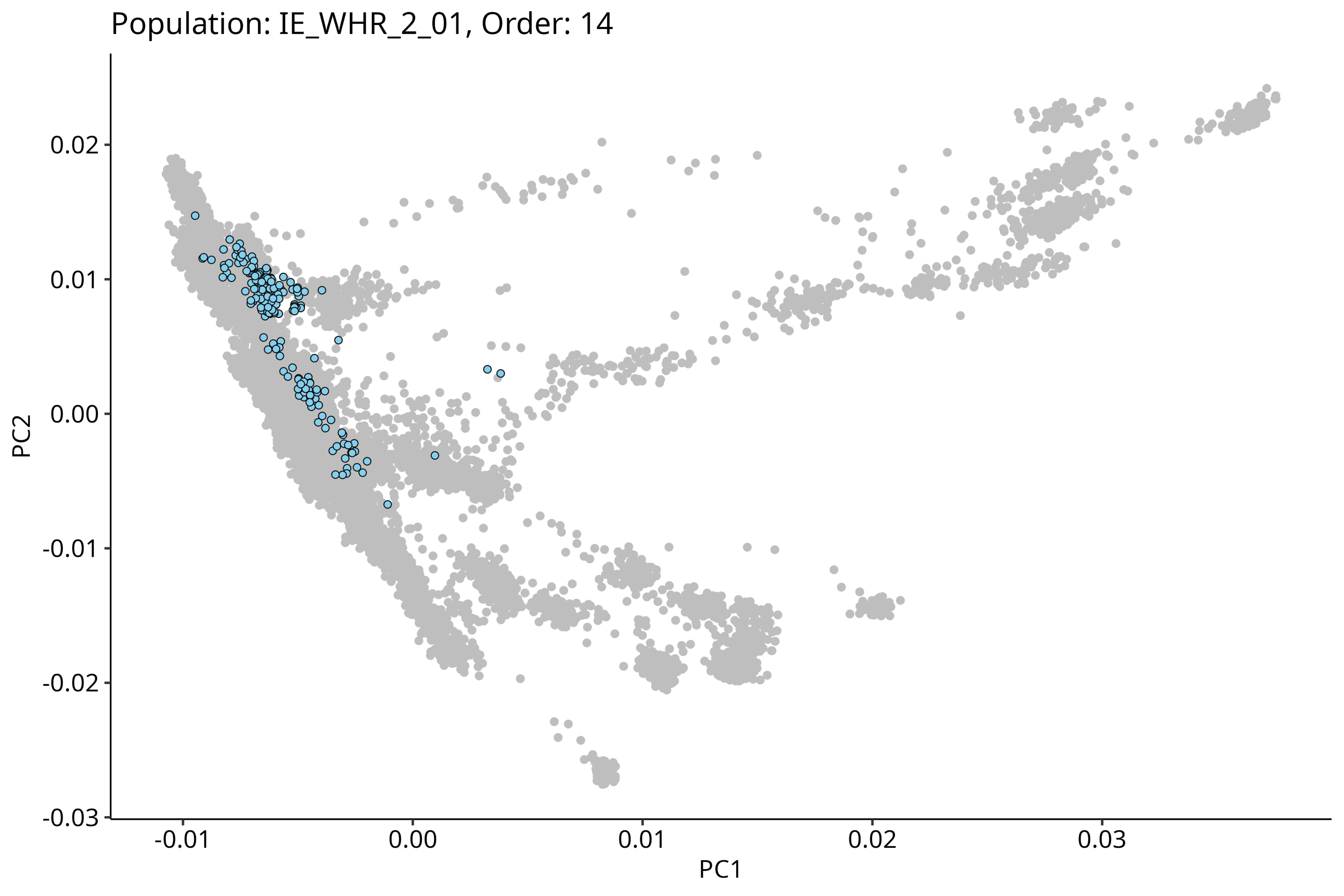

Population: IE_WHR_2_01, Order: 14
0.02
0.01
0.00
-0.01
-0.02
-0.03
PC2
-0.01
0.00
0.01
0.02
0.03
PC1

Population: IE_ERP_2_02, Order: 15

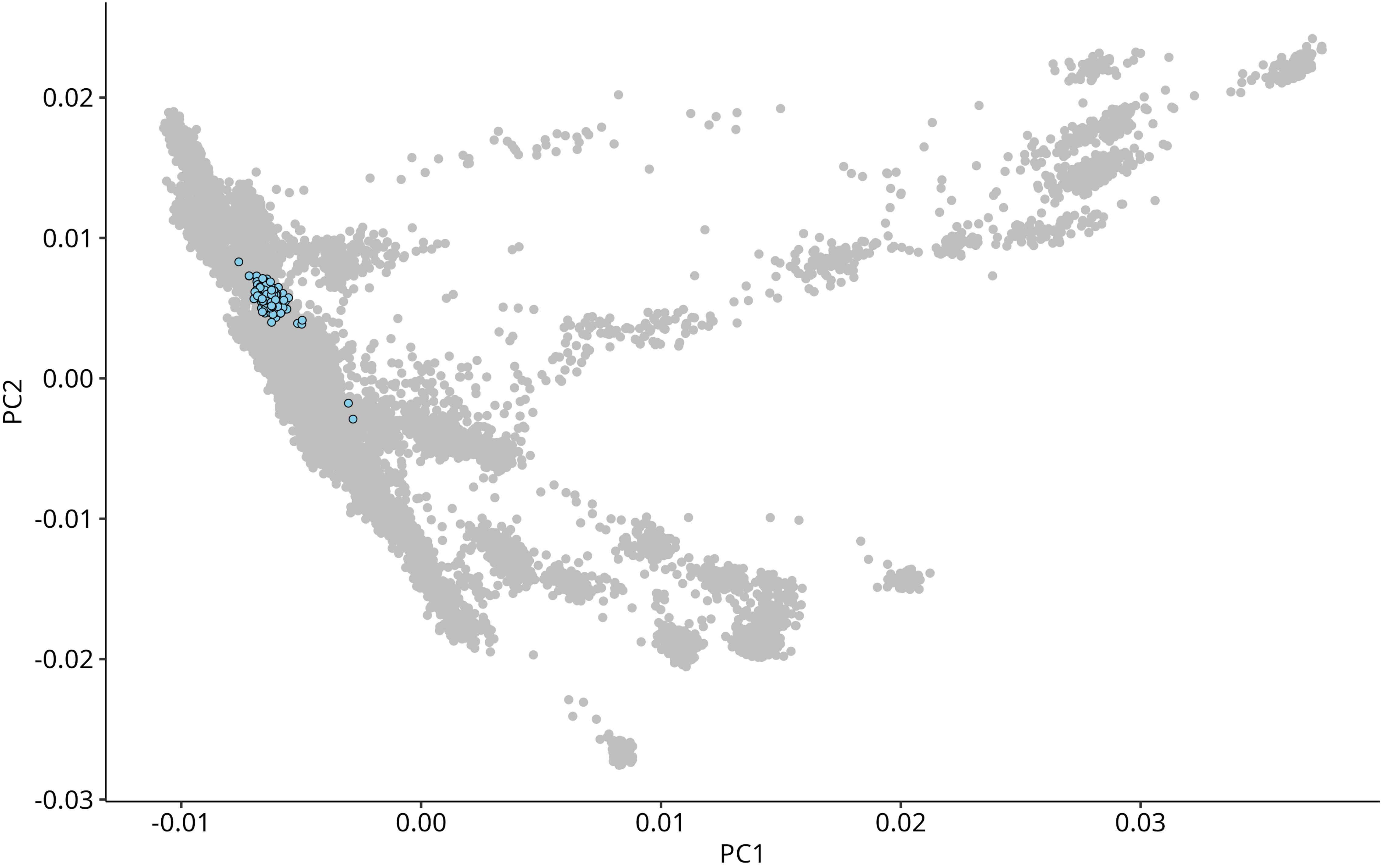

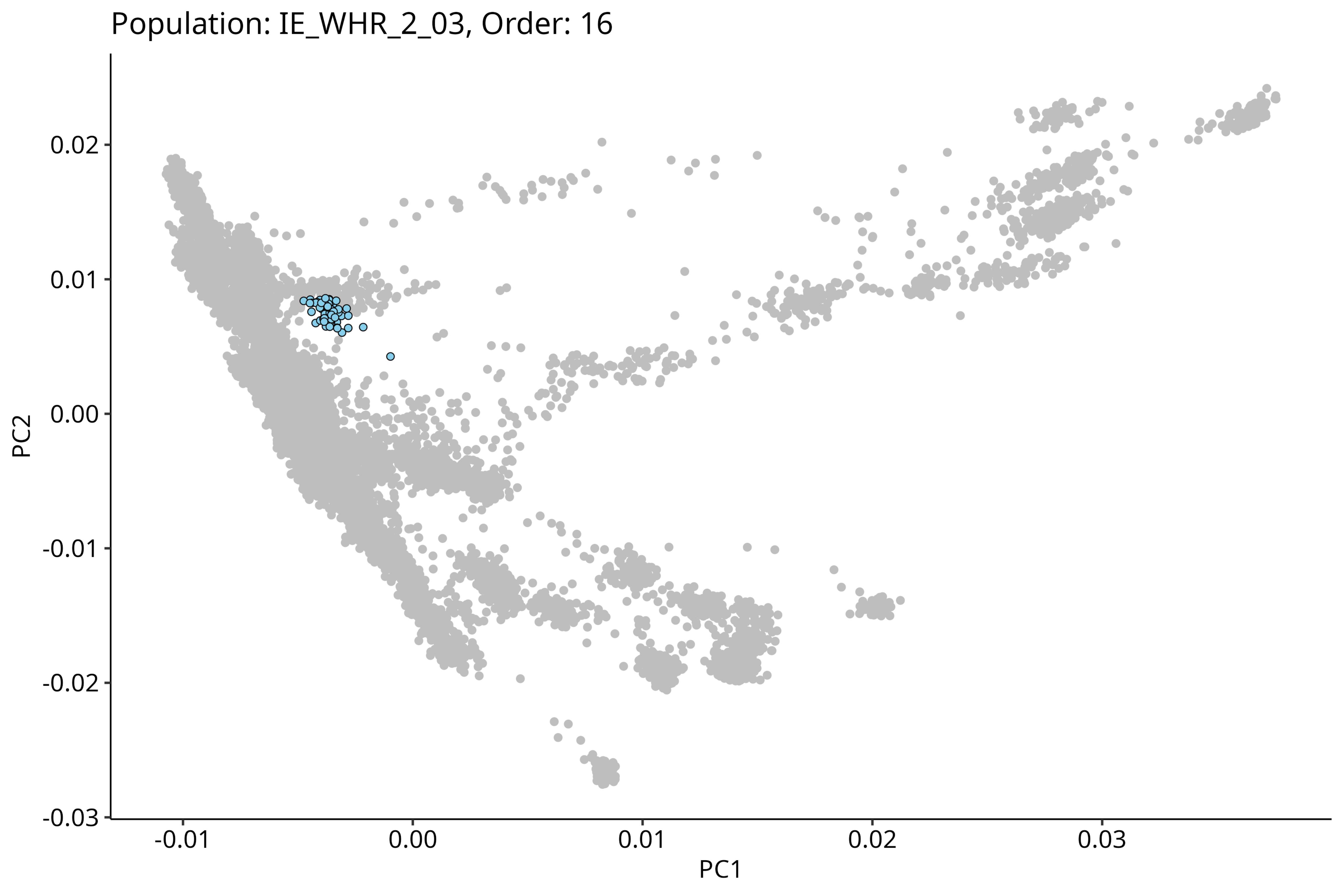
Population: IE_WHR_2_03, Order: 16
PC2
0.02
0.01
0.00
-0.01
-0.02
-0.03
PC1
-0.01
0.00
0.01
0.02
0.03

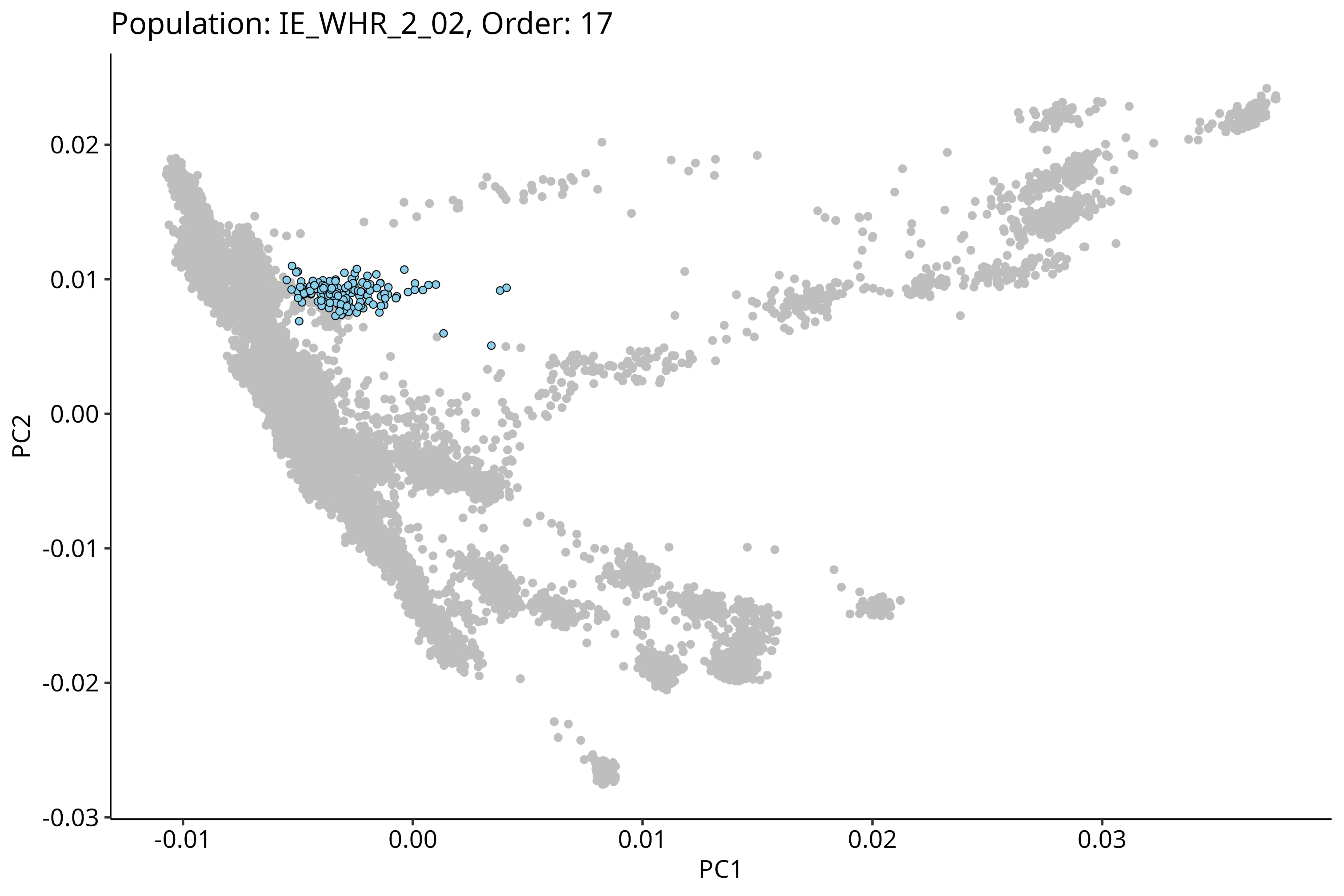
Population: IE_WHR_2_02, Order: 17
PC2
0.02
0.01
0.00
-0.01
-0.02
-0.03
-0.01
0.00
0.01
0.02
0.03
PC1

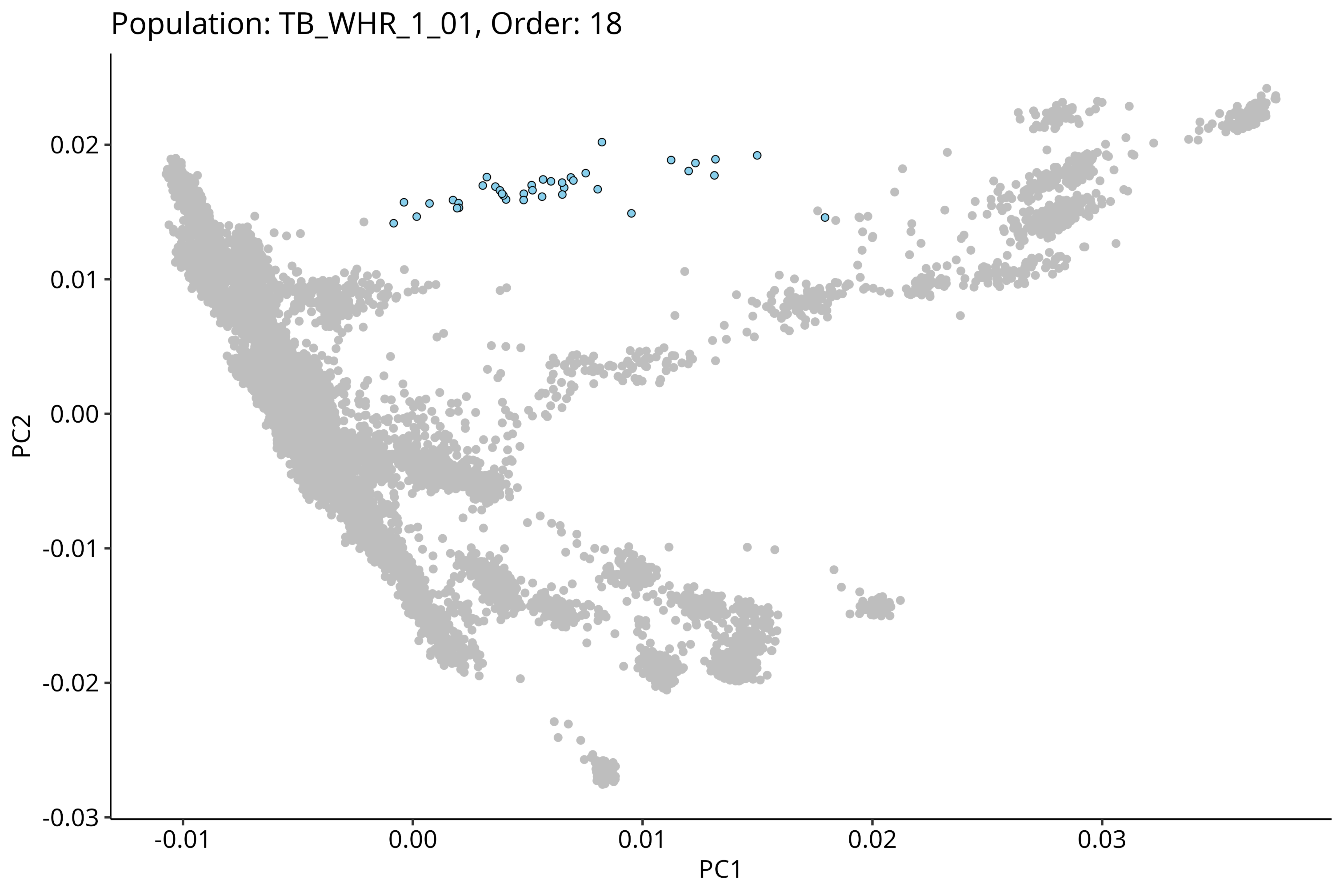
Population: TB_WHR_1_01, Order: 18
PC2
0.02
0.01
0.00
-0.01
-0.02
-0.03
PC1
-0.01
0.00
0.01
0.02
0.03

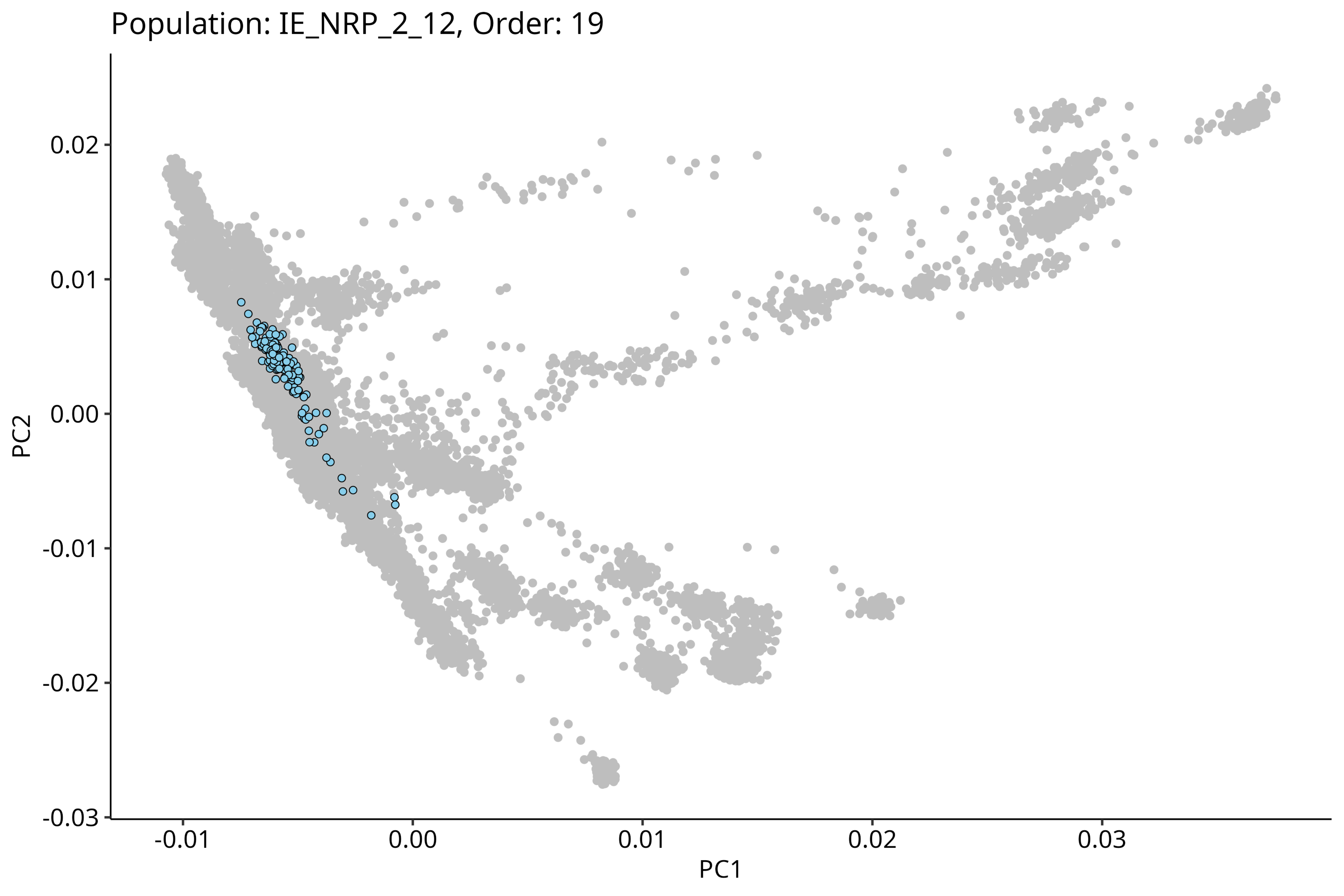

Population: IE_NRP_2_12, Order: 19
0.02
0.01
0.00
-0.01
-0.02
-0.03
PC2
-0.01
0.00
0.01
0.02
0.03
PC1

Population: IE_NRP_2_13, Order: 20

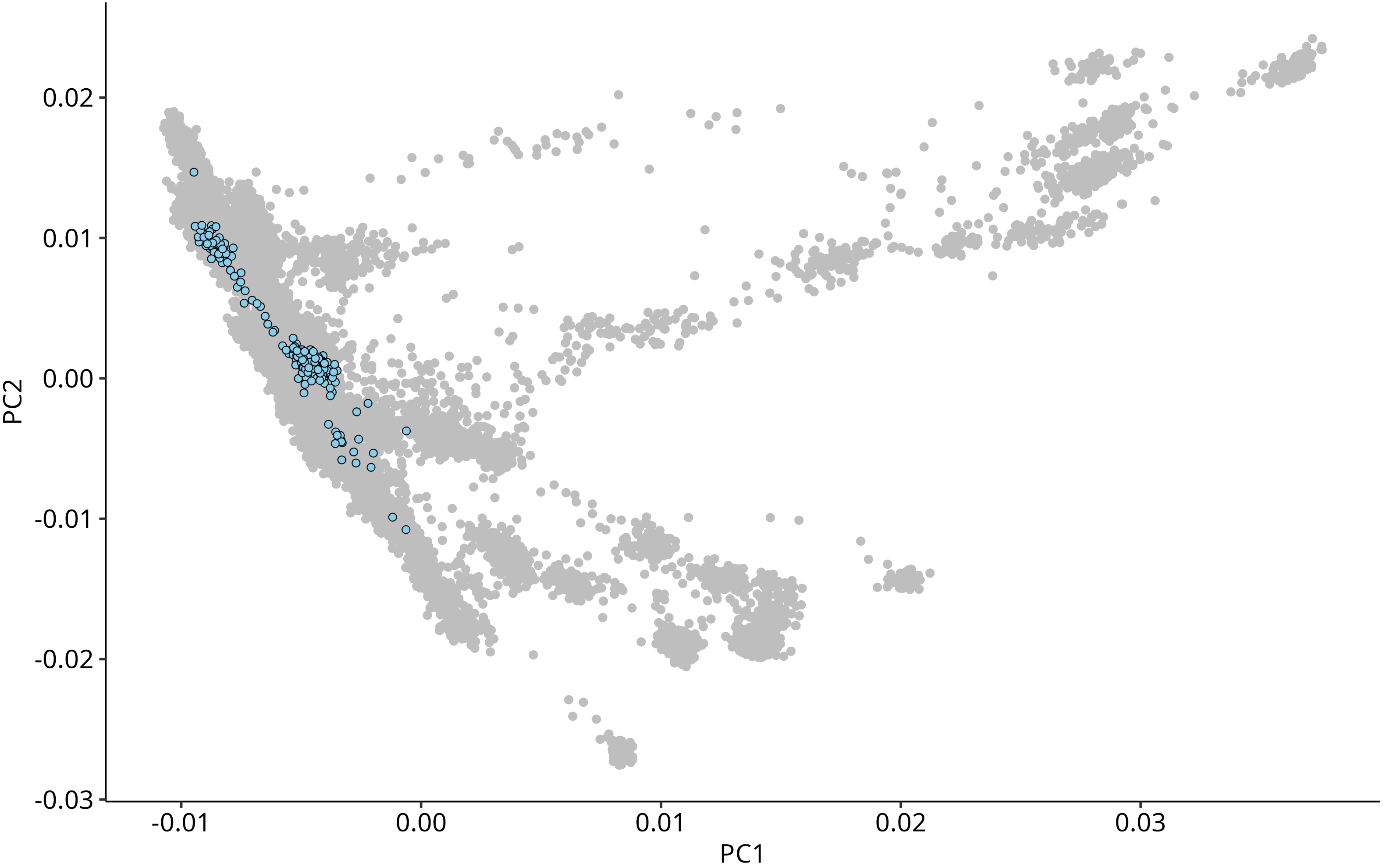

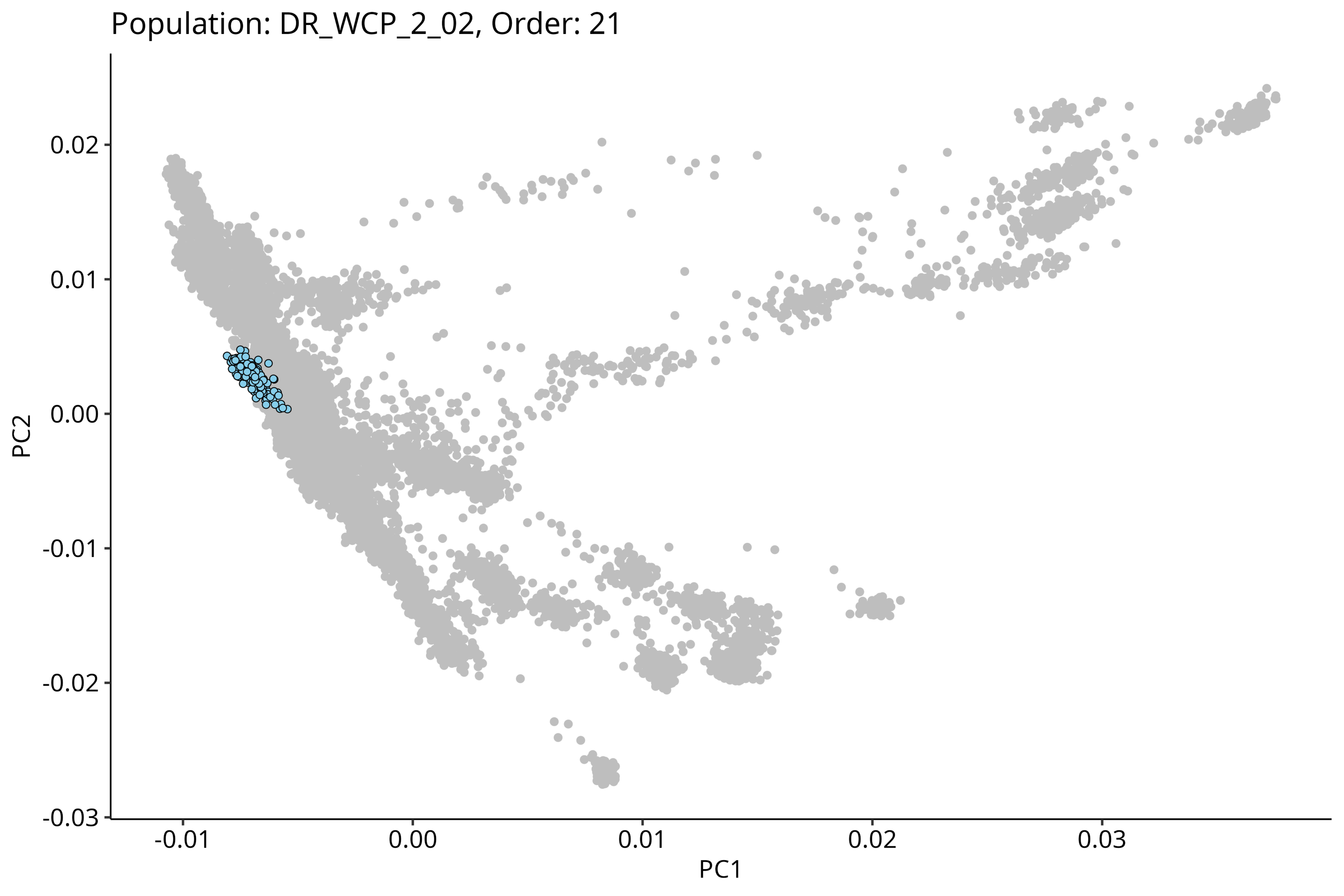

Population: DR_WCP_2_02, Order: 21
PC2
PC1
0.02
0.01
0.00
-0.01
-0.02
-0.03
-0.01
0.00
0.01
0.02
0.03

Population: IE_ECP_2_02, Order: 22

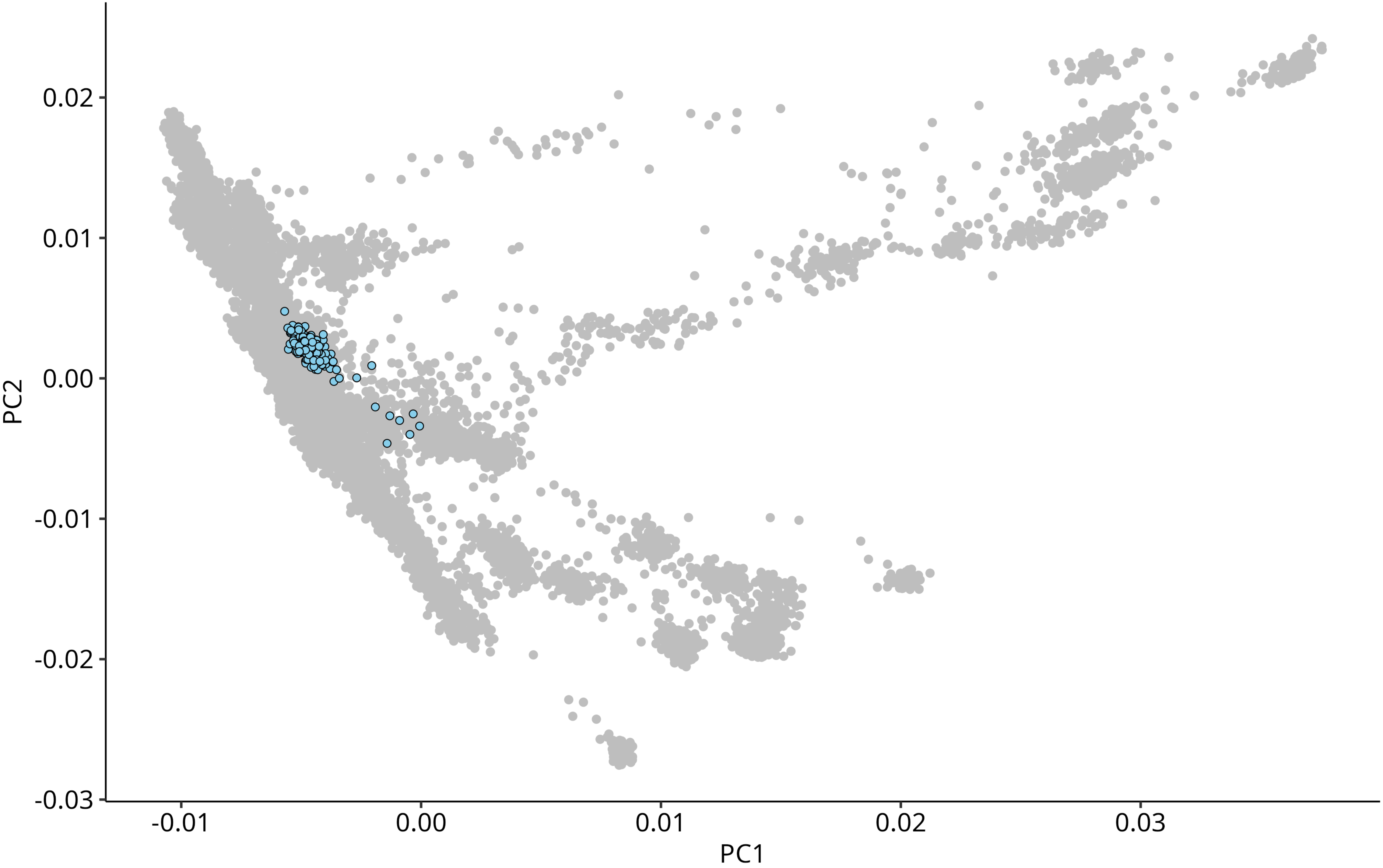

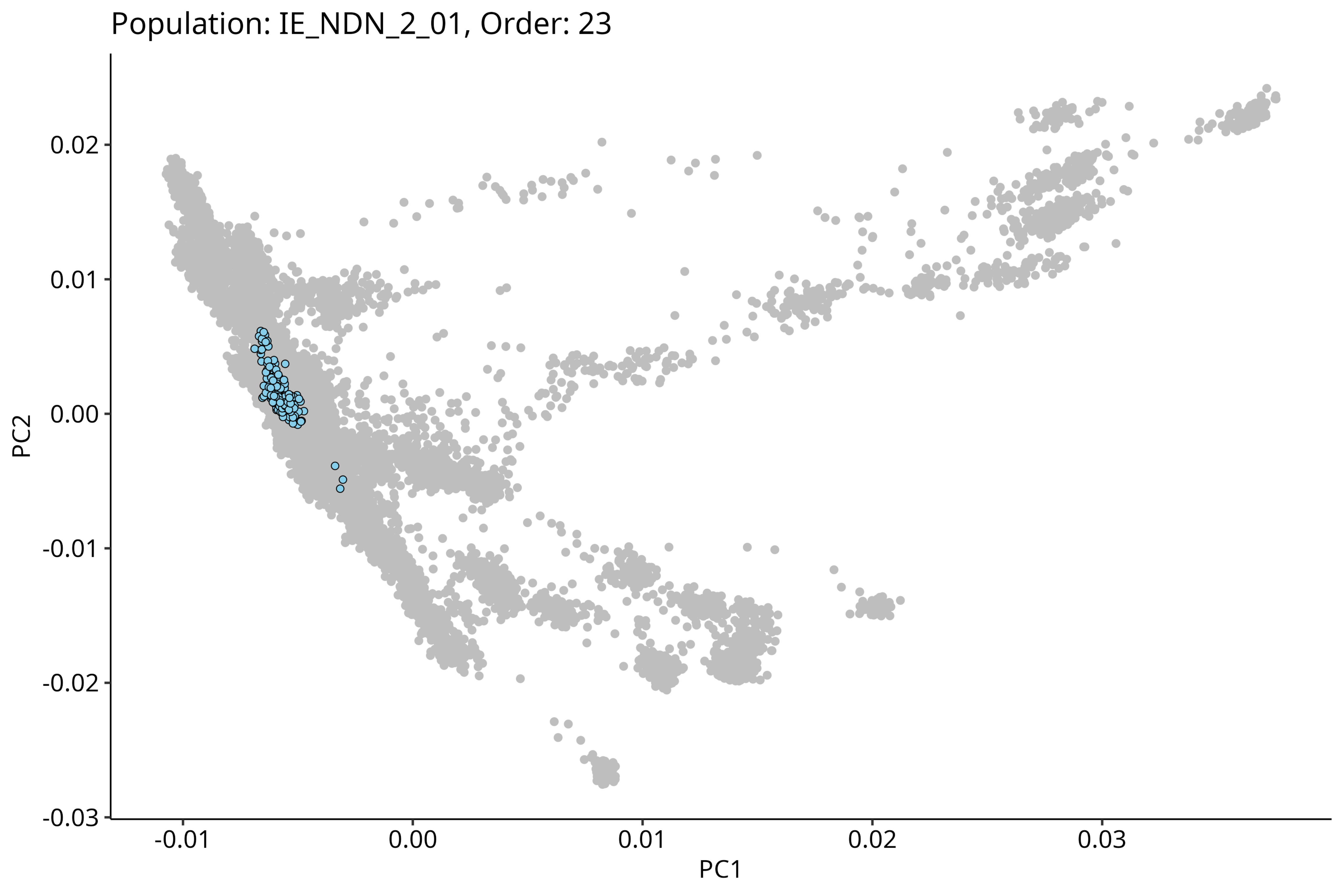
Population: IE_NDN_2_01, Order: 23
PC2
0.02
0.01
0.00
-0.01
-0.02
-0.03
PC1
-0.01
0.00
0.01
0.02
0.03

#### Population: IE_NCH_2_01, Order: 24

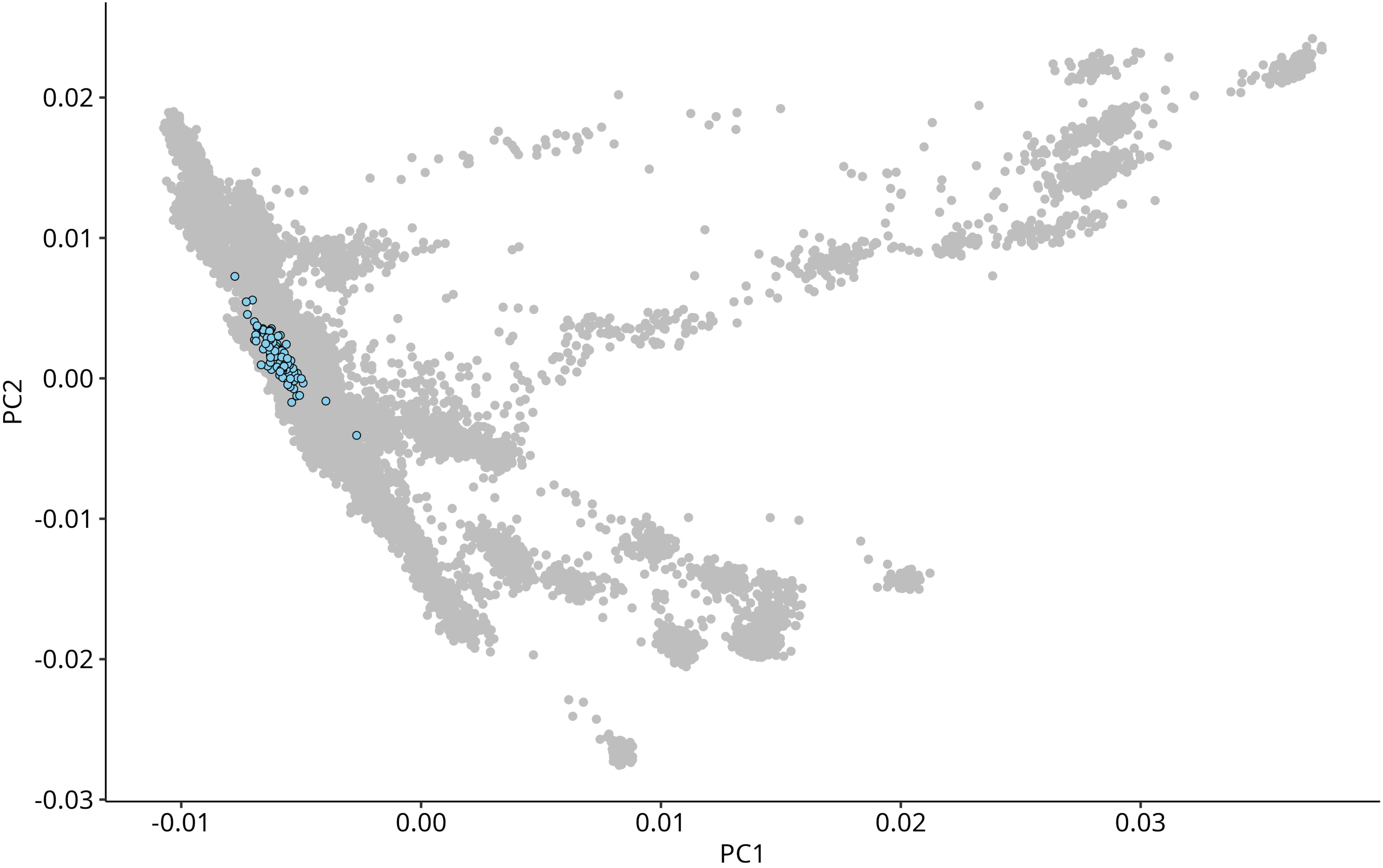

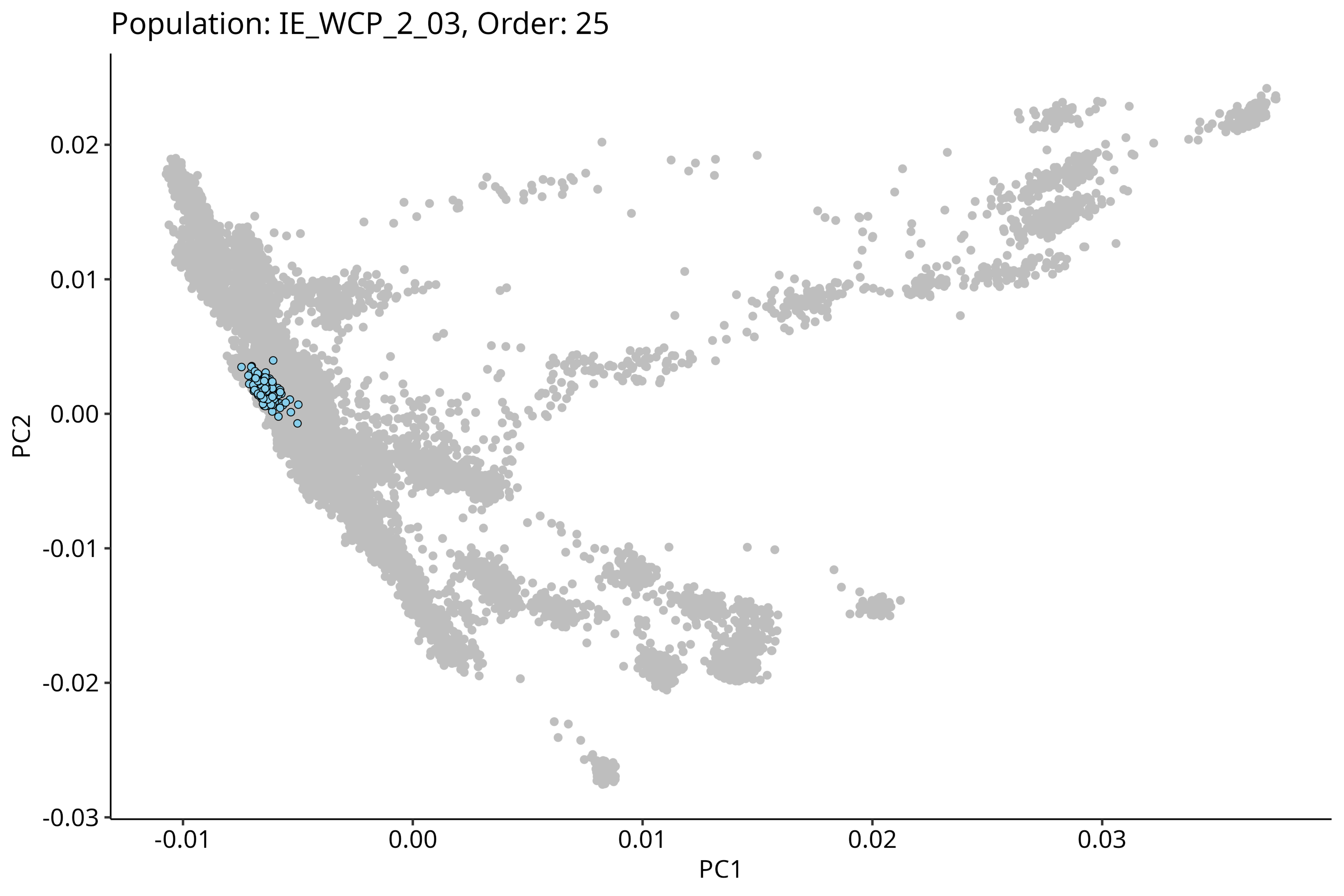

Population: IE_WCP_2_03, Order: 25
0.02
0.01
0.00
-0.01
-0.02
-0.03
PC2
-0.01
0.00
0.01
0.02
0.03
PC1

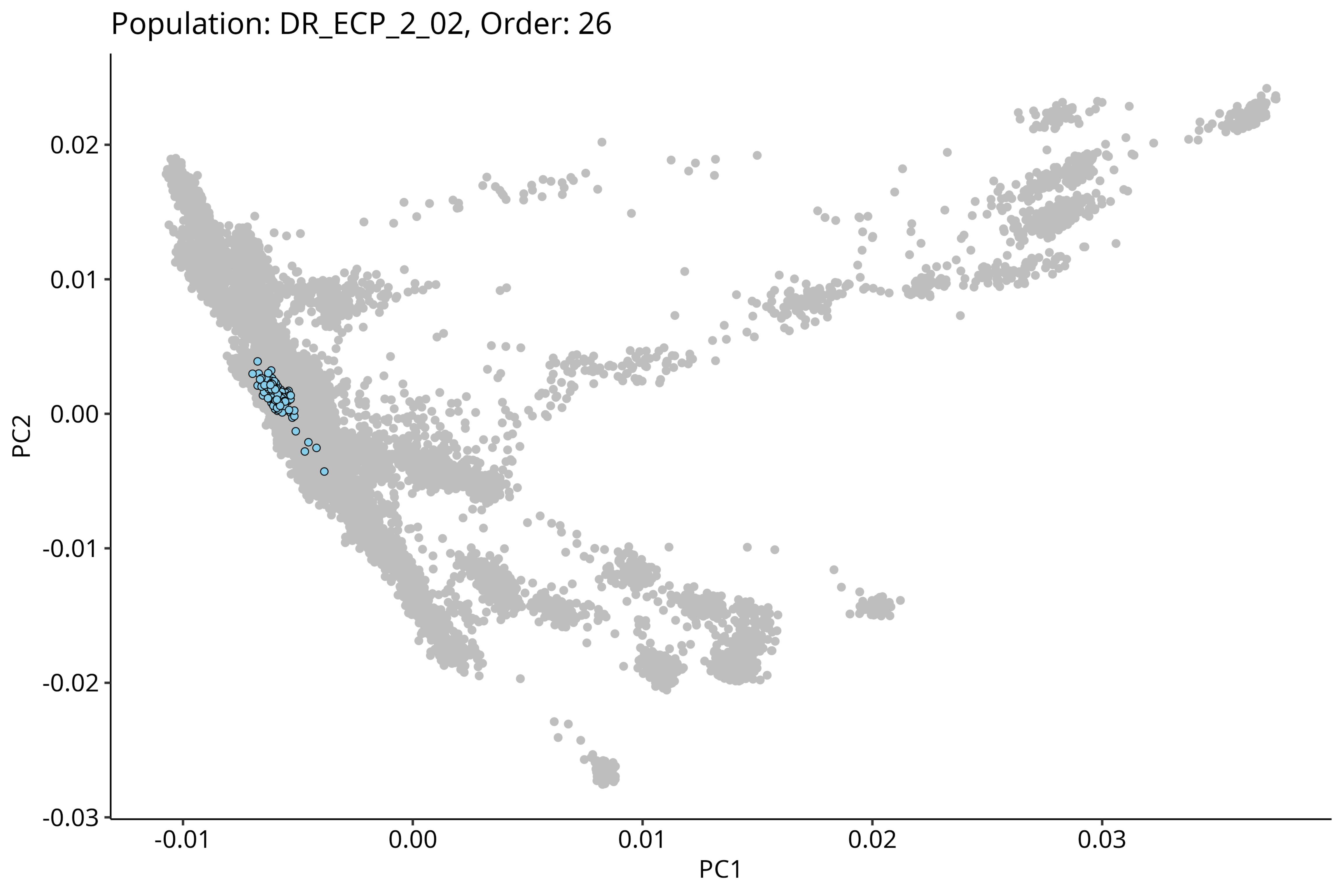
Population: DR_ECP_2_02, Order: 26
0.02
0.01
0.00
-0.01
-0.02
-0.03
PC2
-0.01
0.00
0.01
0.02
0.03
PC1

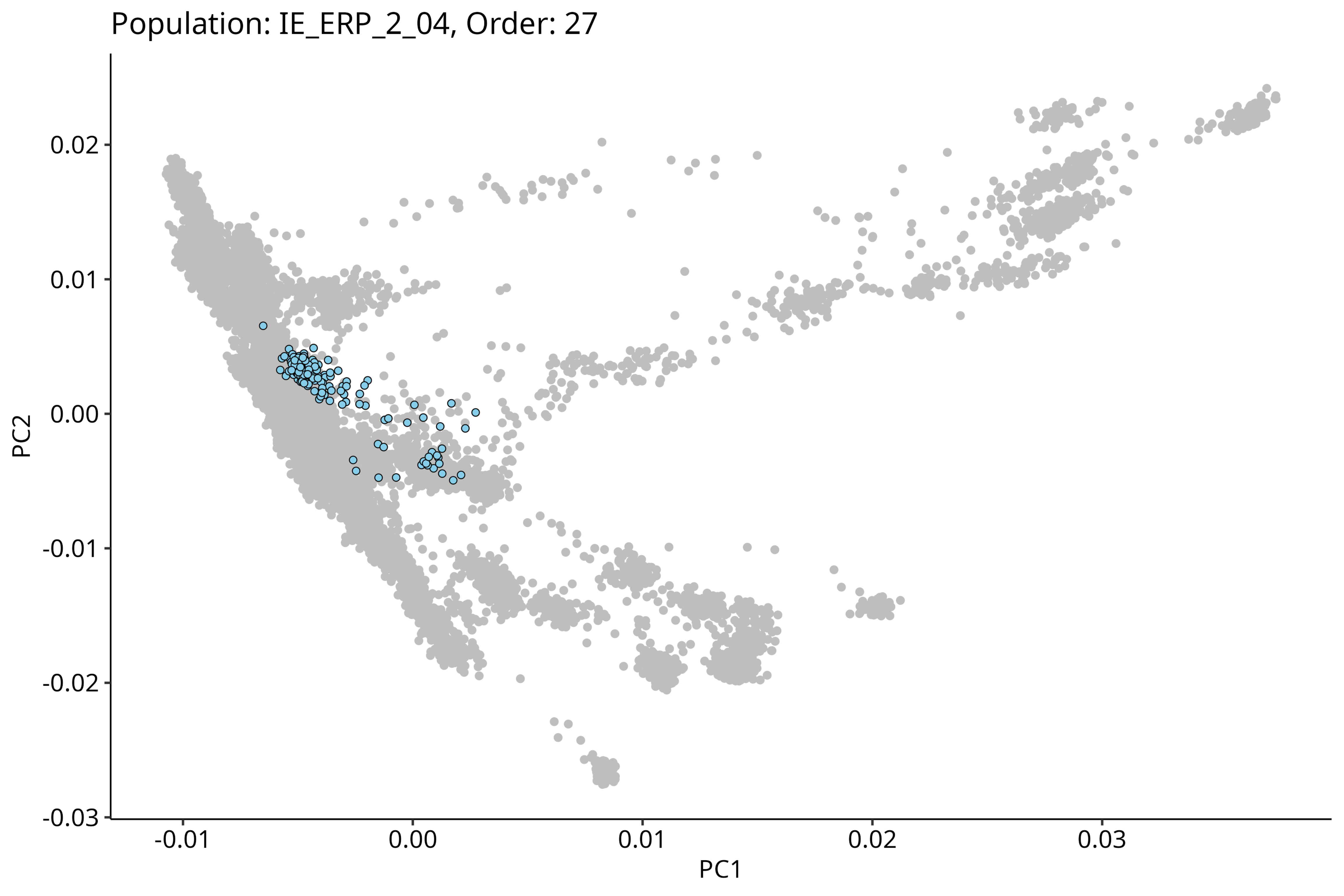

Population: IE_ERP_2_04, Order: 27
PC2
PC1
0.02
0.01
0.00
-0.01
-0.02
-0.03
-0.01
0.00
0.01
0.02
0.03

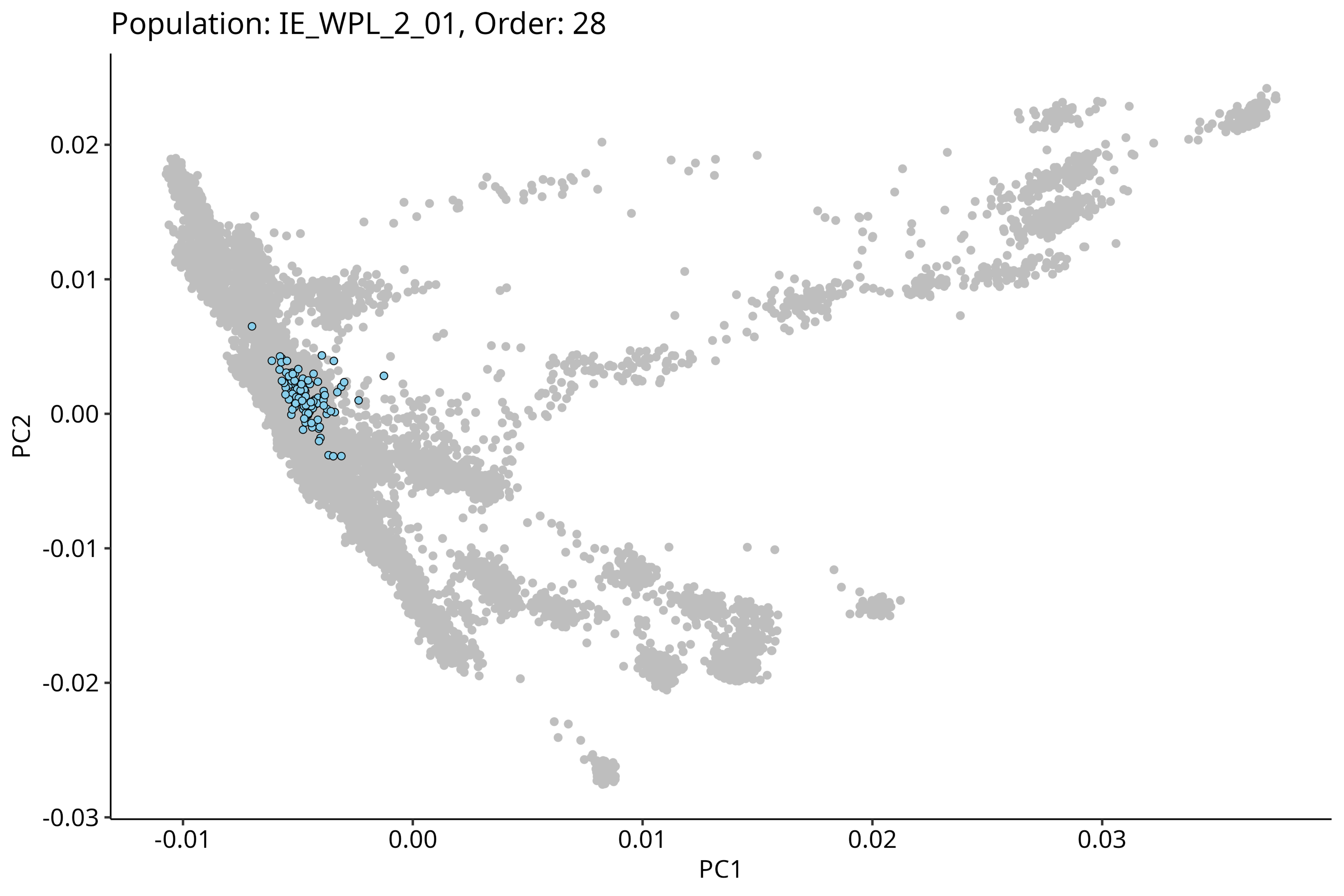

Population: IE_WPL_2_01, Order: 28
0.02
0.01
0.00
-0.01
-0.02
-0.03
PC2
-0.01
0.00
0.01
0.02
0.03
PC1

Population: DR_ECP_2_08, Order: 29

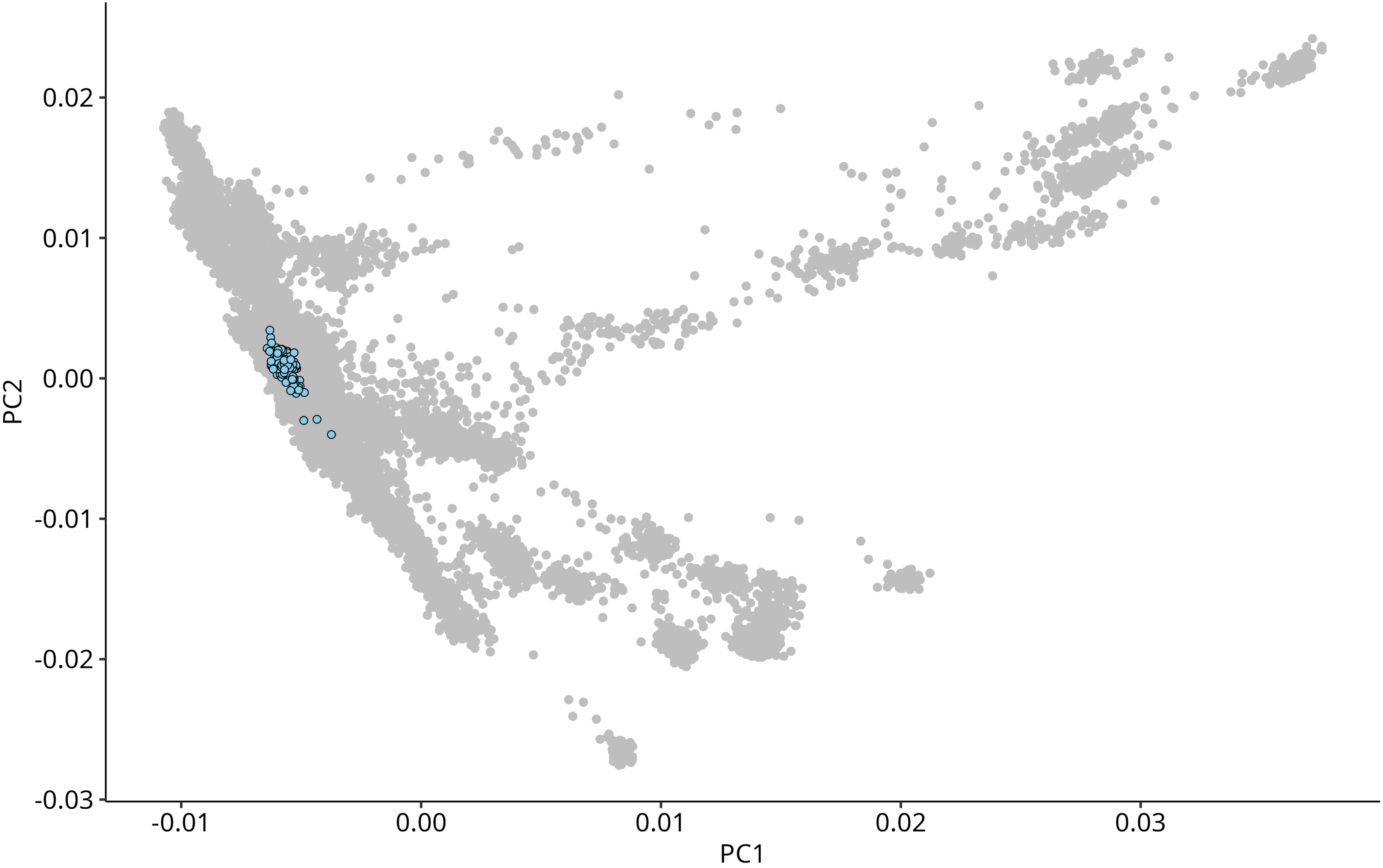

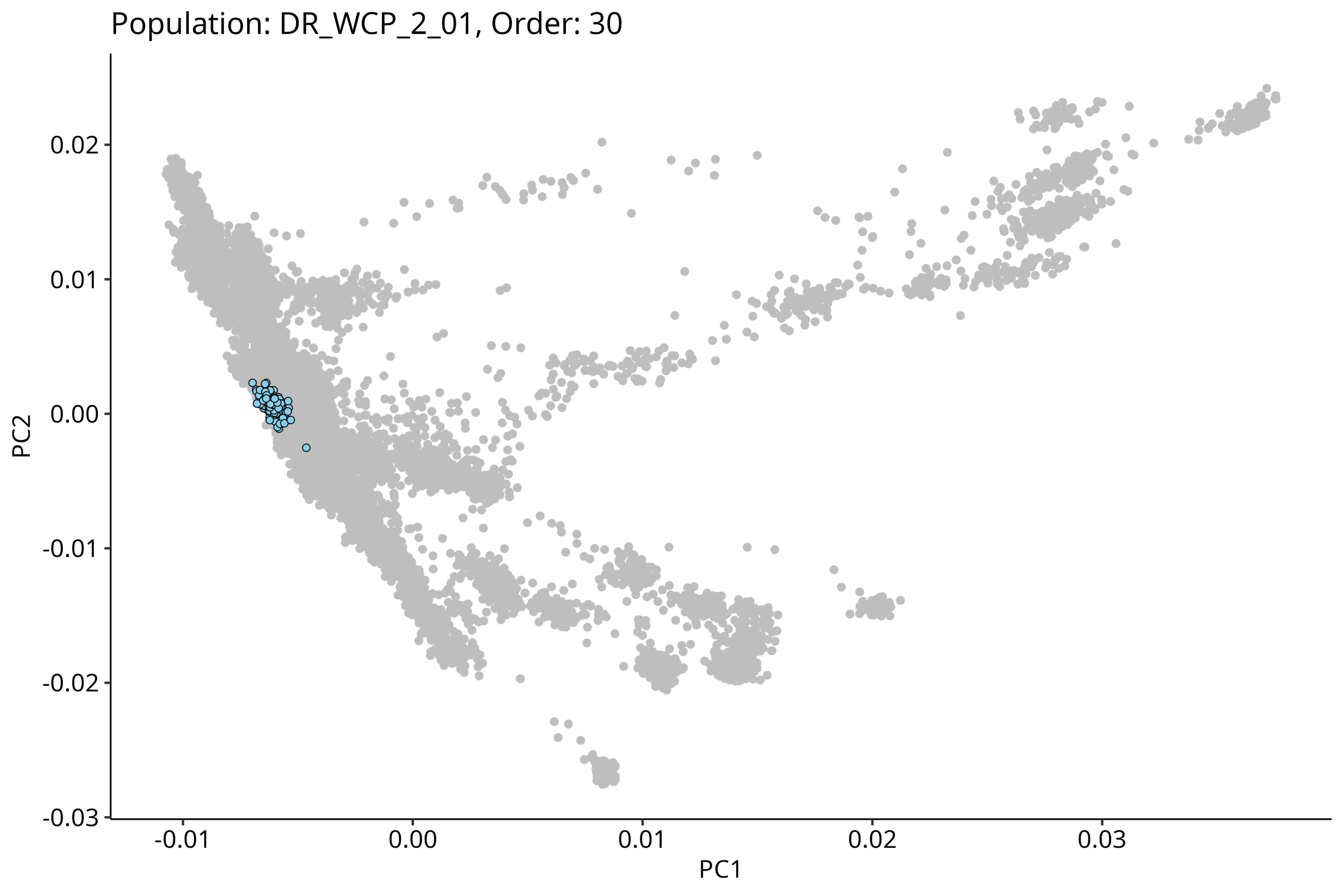

Population: DR_WCP_2_01, Order: 30
0.02
0.01
0.00
-0.01
-0.02
-0.03
PC2
-0.01
0.00
0.01
0.02
0.03
PC1

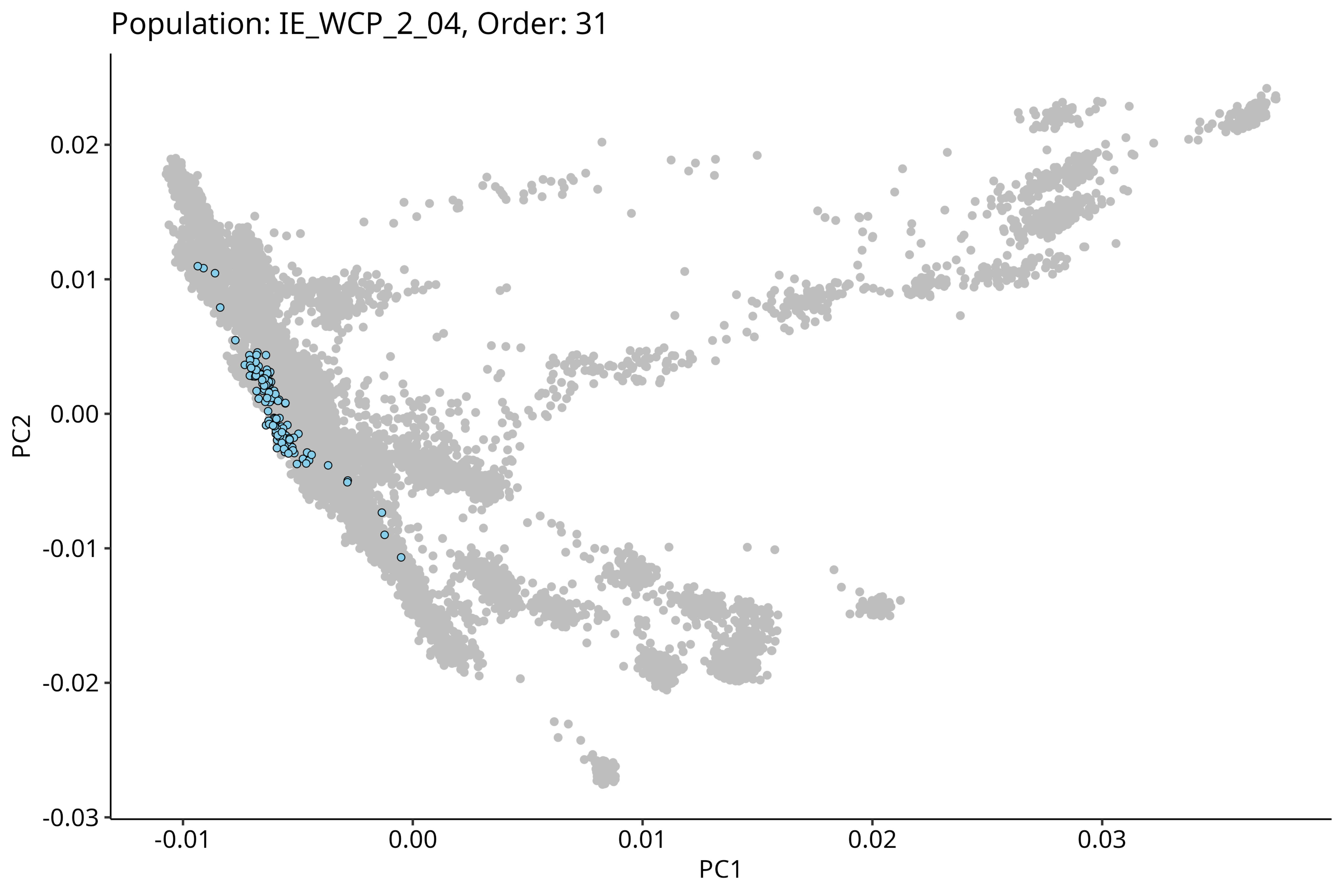
Population: IE_WCP_2_04, Order: 31
PC2
0.02
0.01
0.00
-0.01
-0.02
-0.03
-0.01
0.00
0.01
0.02
0.03
PC1

### Population: IE_NRP_2_07, Order: 32

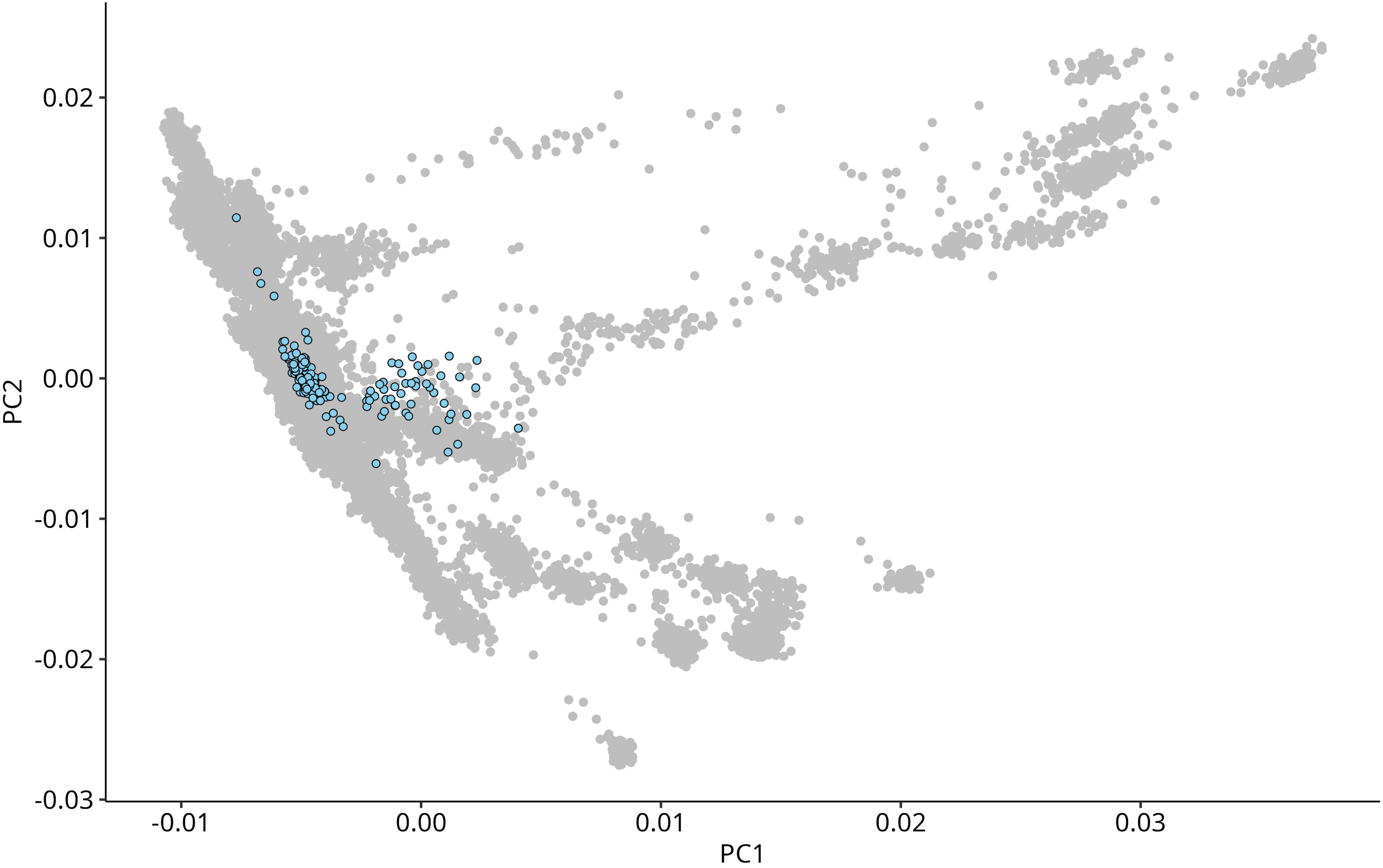

Population: IE_WHR_1_01, Order: 33

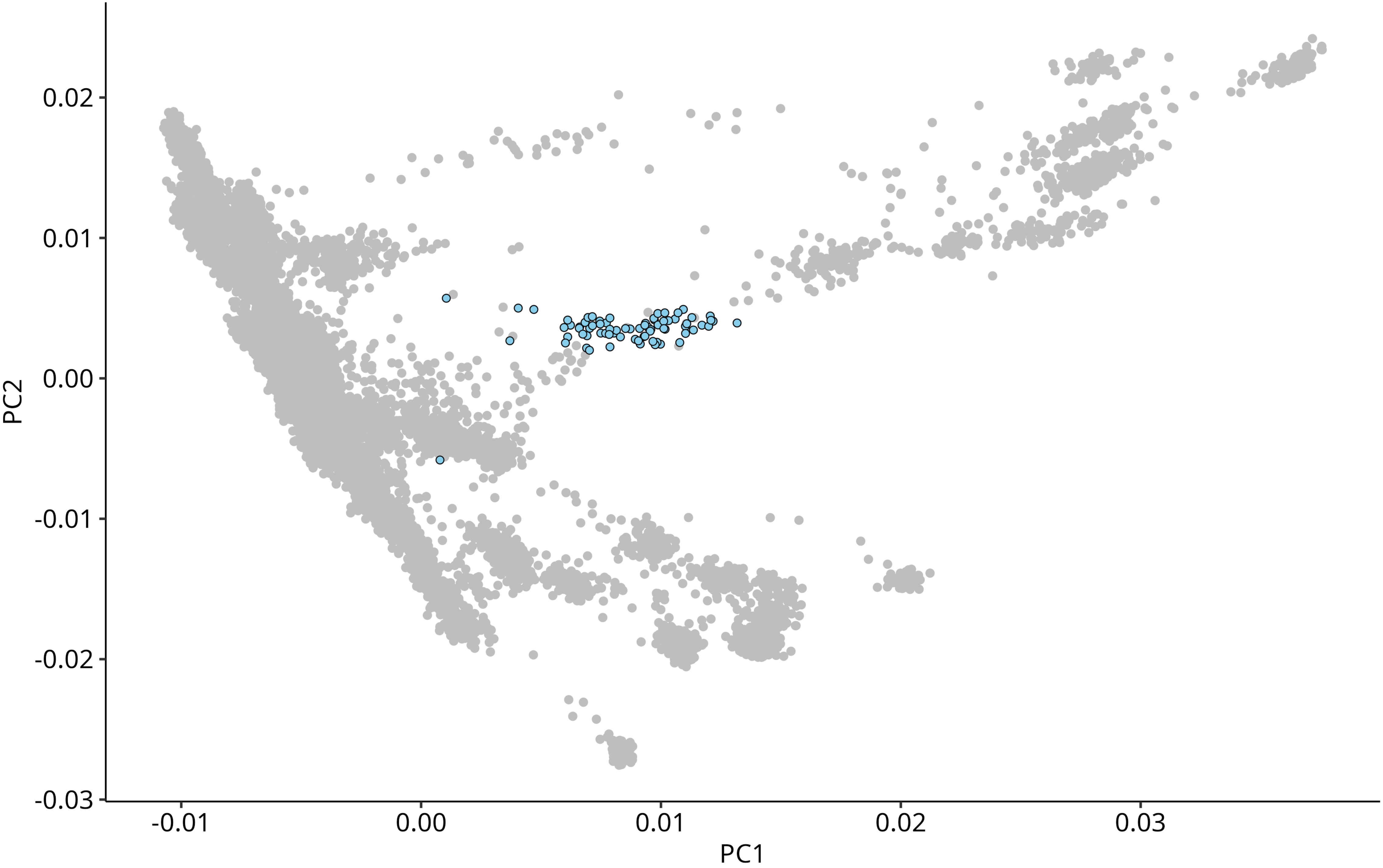

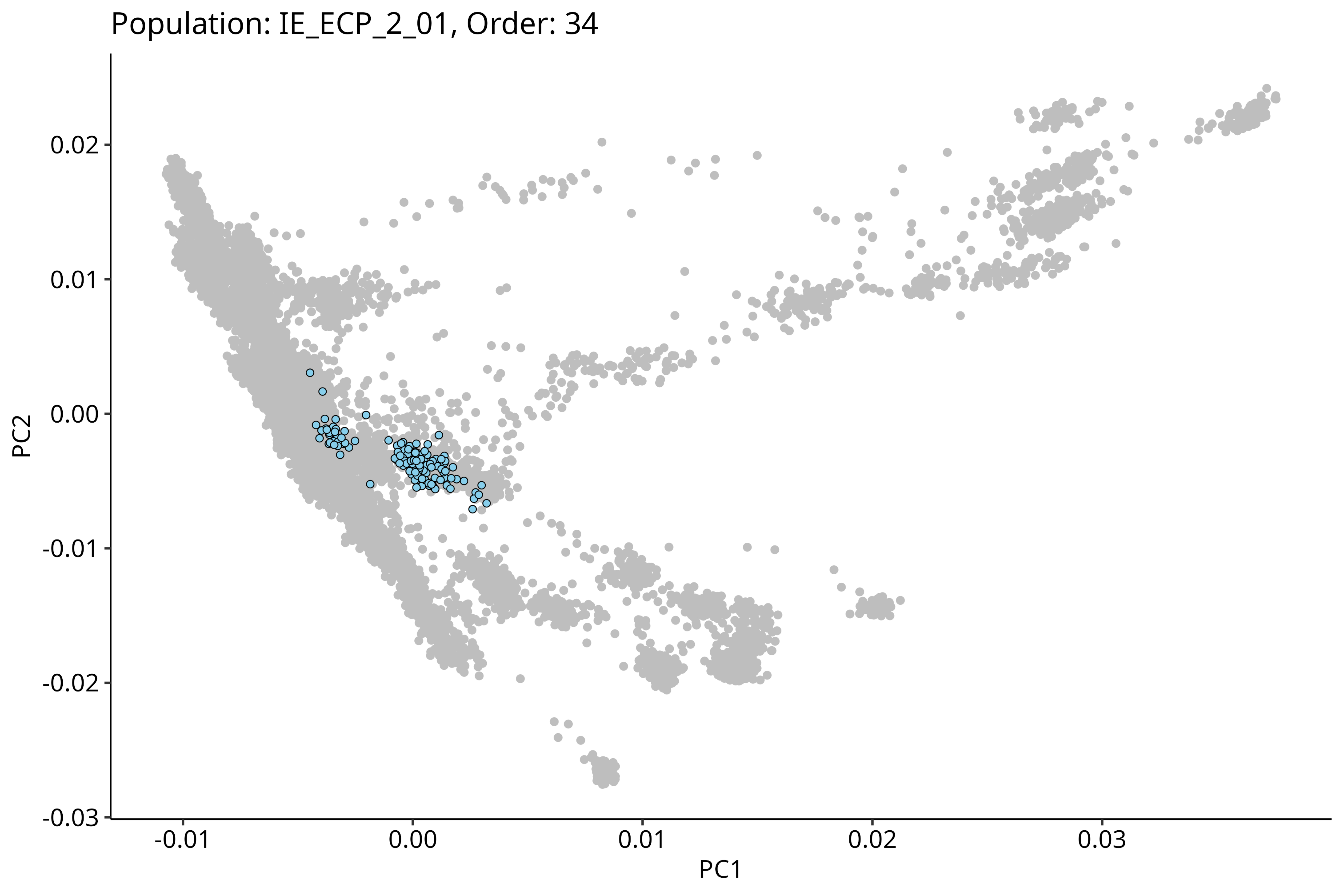

Population: IE_ECP_2_01, Order: 34
0.02
0.01
0.00
-0.01
-0.02
-0.03
PC2
-0.01
0.00
0.01
0.02
0.03
PC1

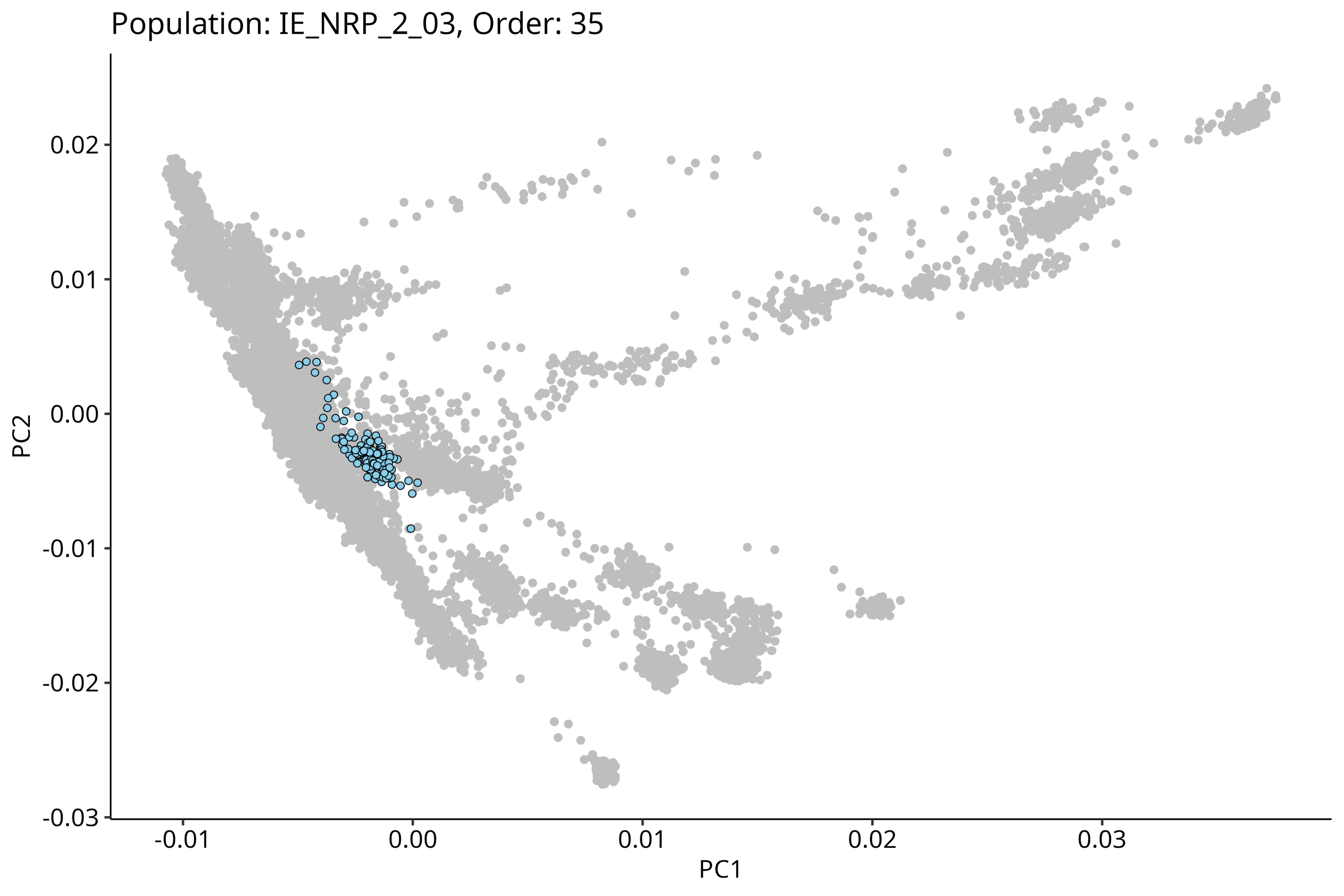
Population: IE_NRP_2_03, Order: 35
0.02
0.01
0.00
-0.01
-0.02
-0.03
PC2
-0.01
0.00
0.01
0.02
0.03
PC1

Population: IE_ERP_2_01, Order: 36

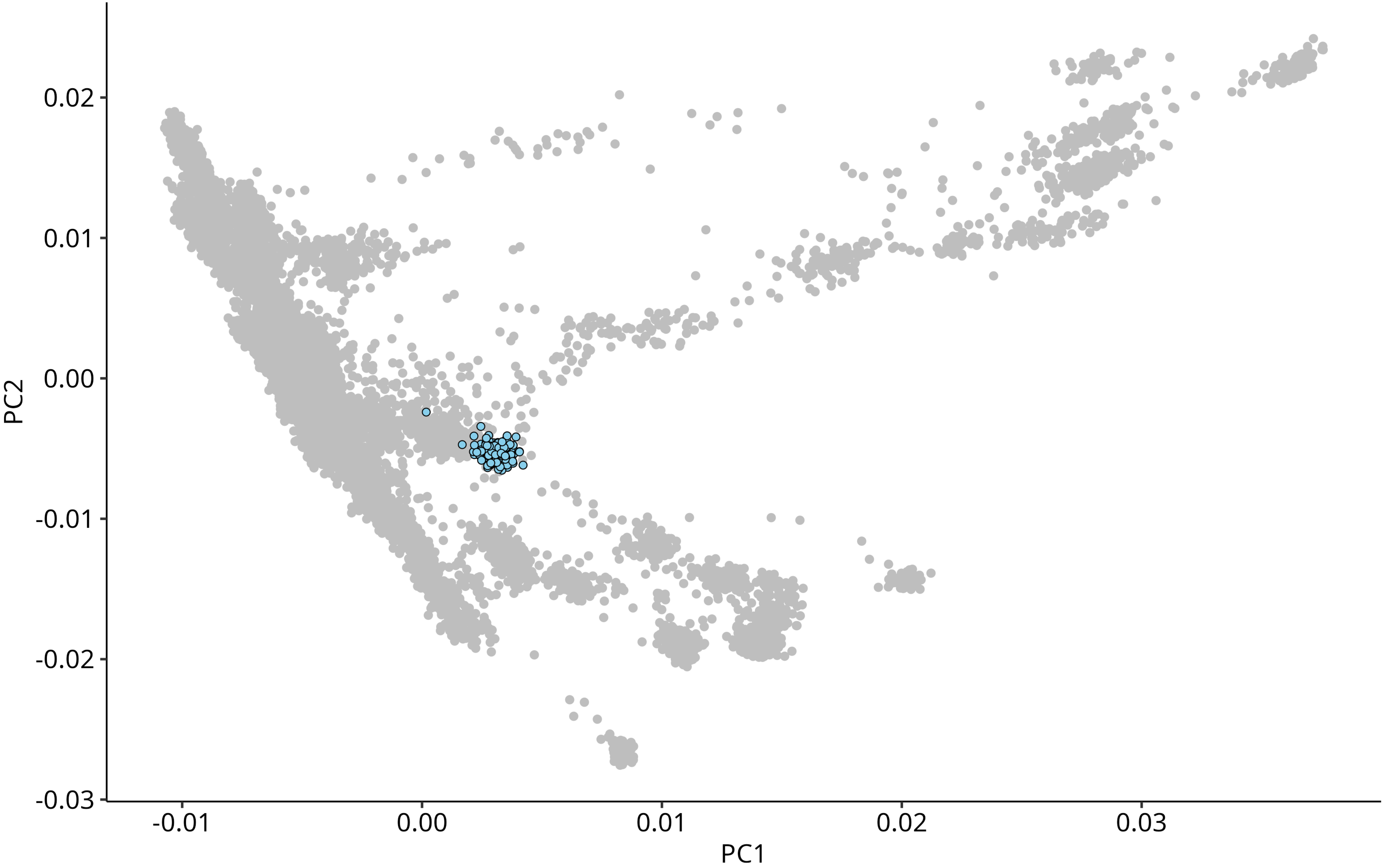

### Population: IE_ERP_2_03, Order: 37

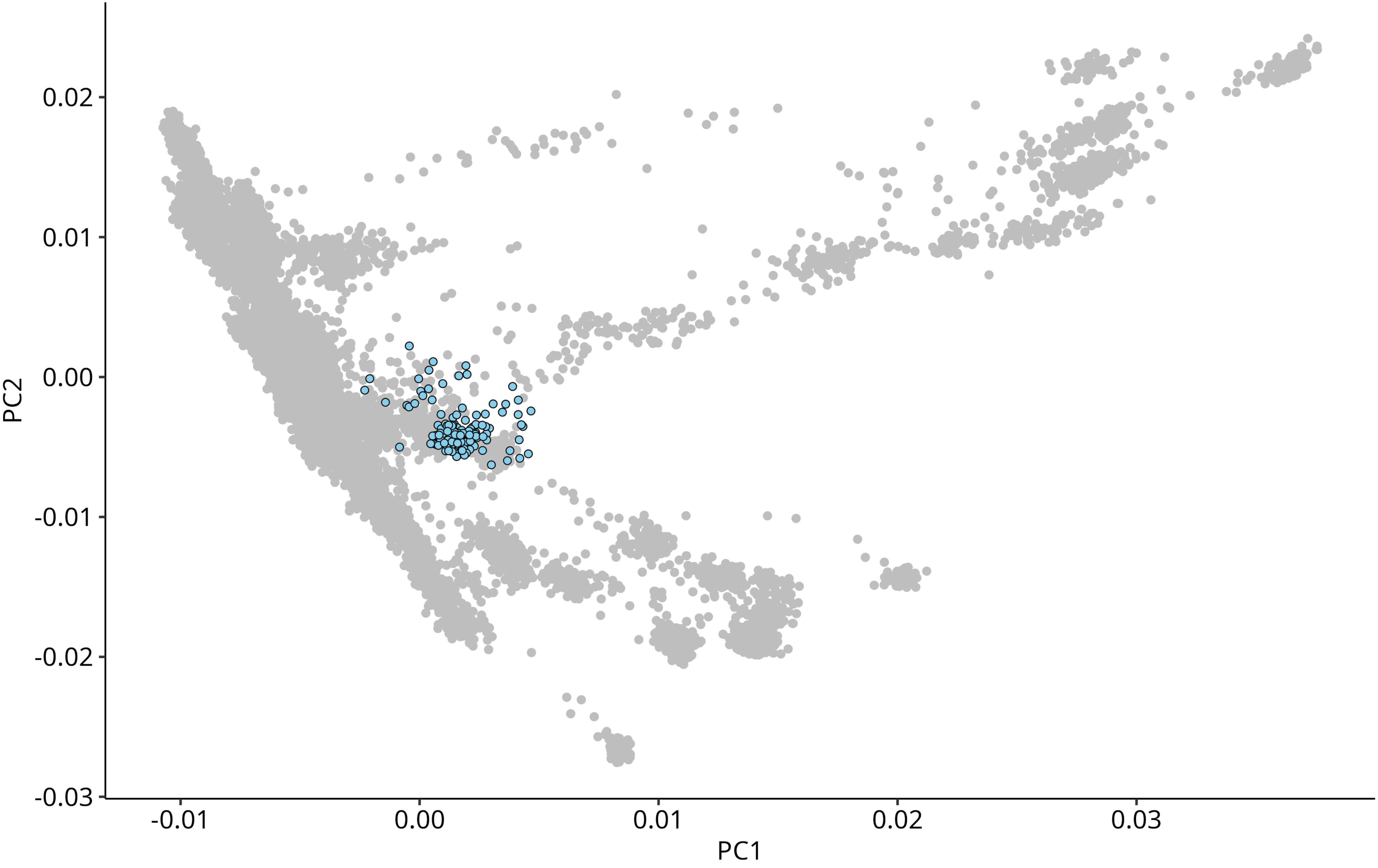

Population: IE_WCP_2_05, Order: 38

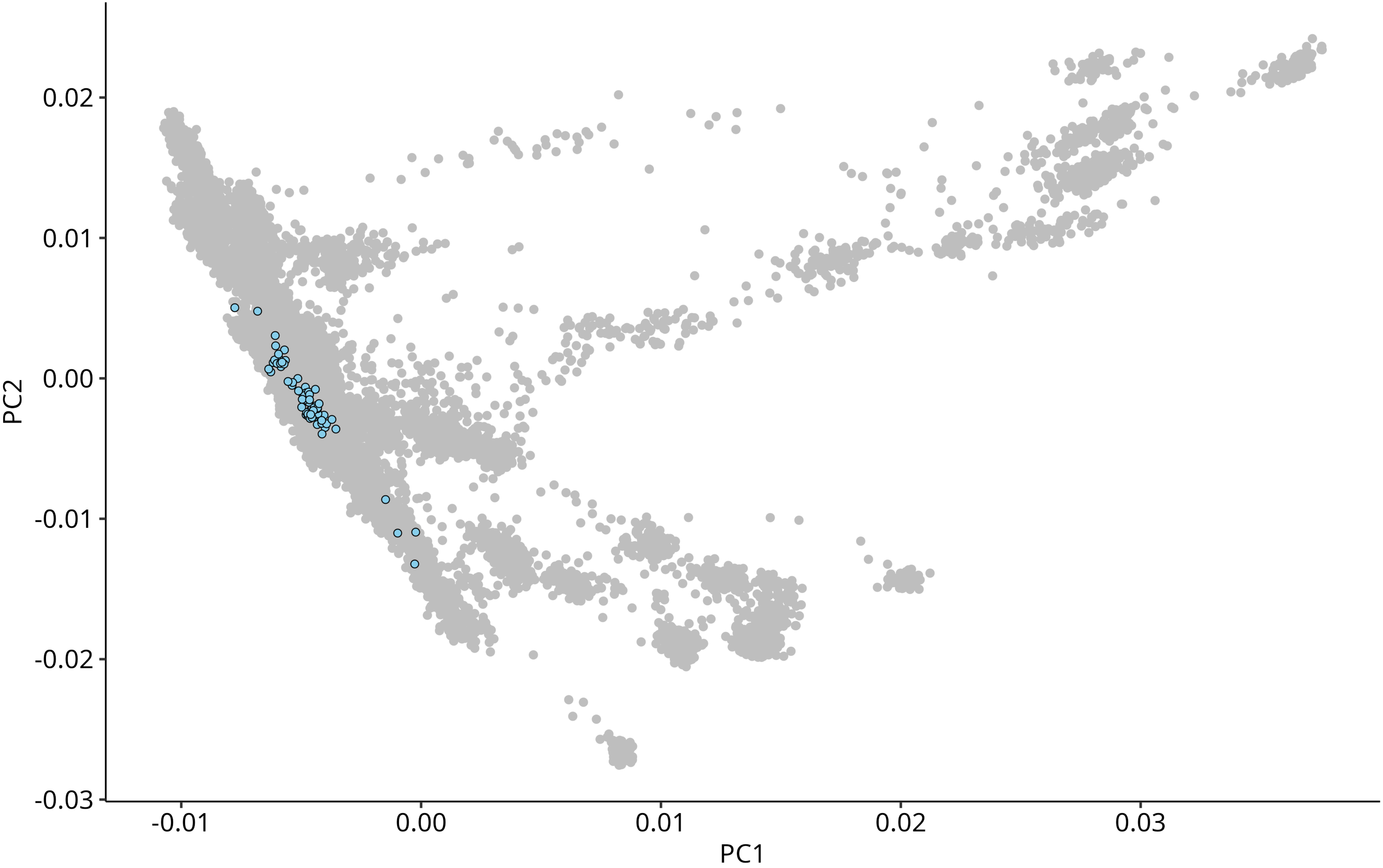

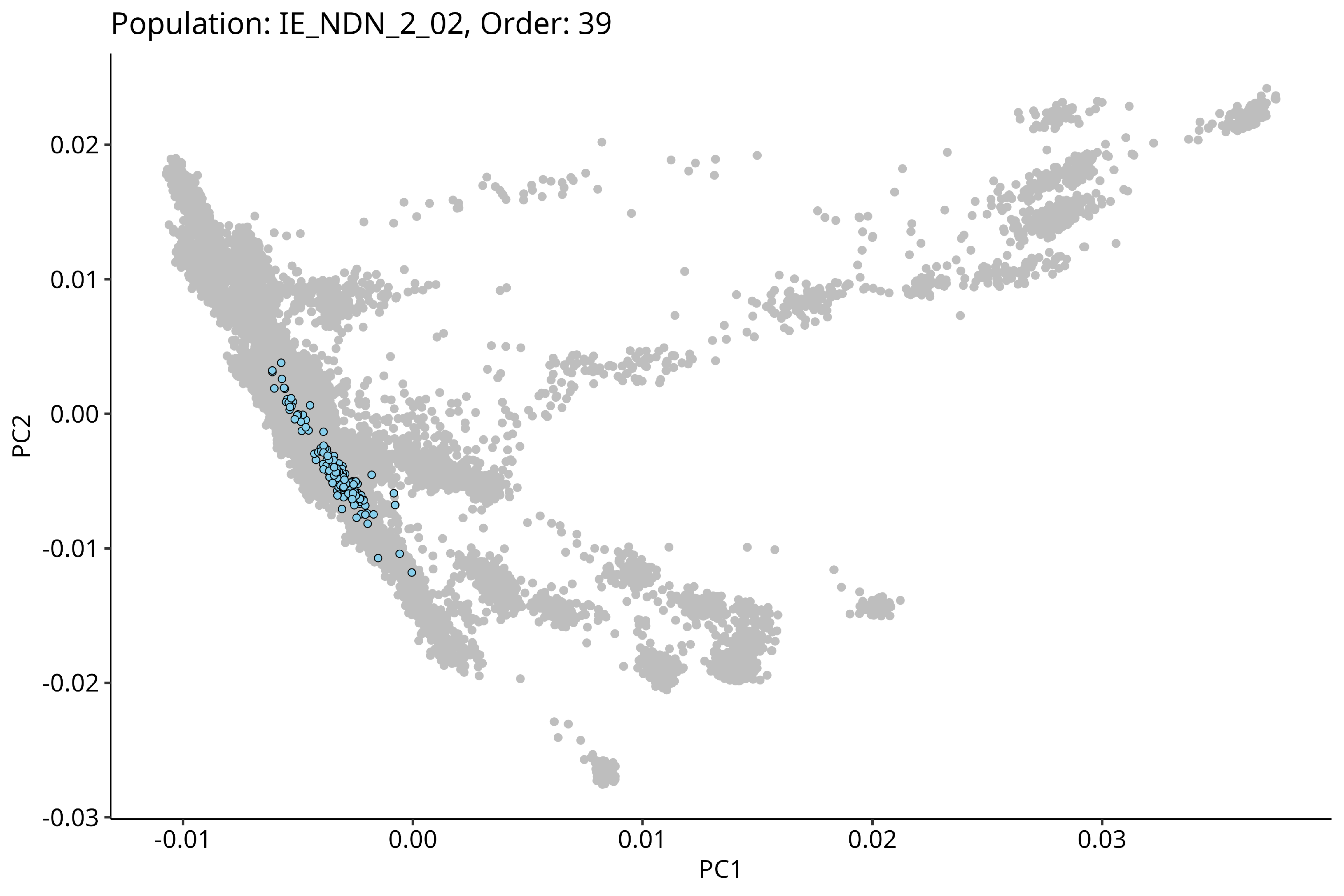
Population: IE_NDN_2_02, Order: 39
PC2
PC1
0.02
0.01
0.00
-0.01
-0.02
-0.03
-0.01
0.00
0.01
0.02
0.03

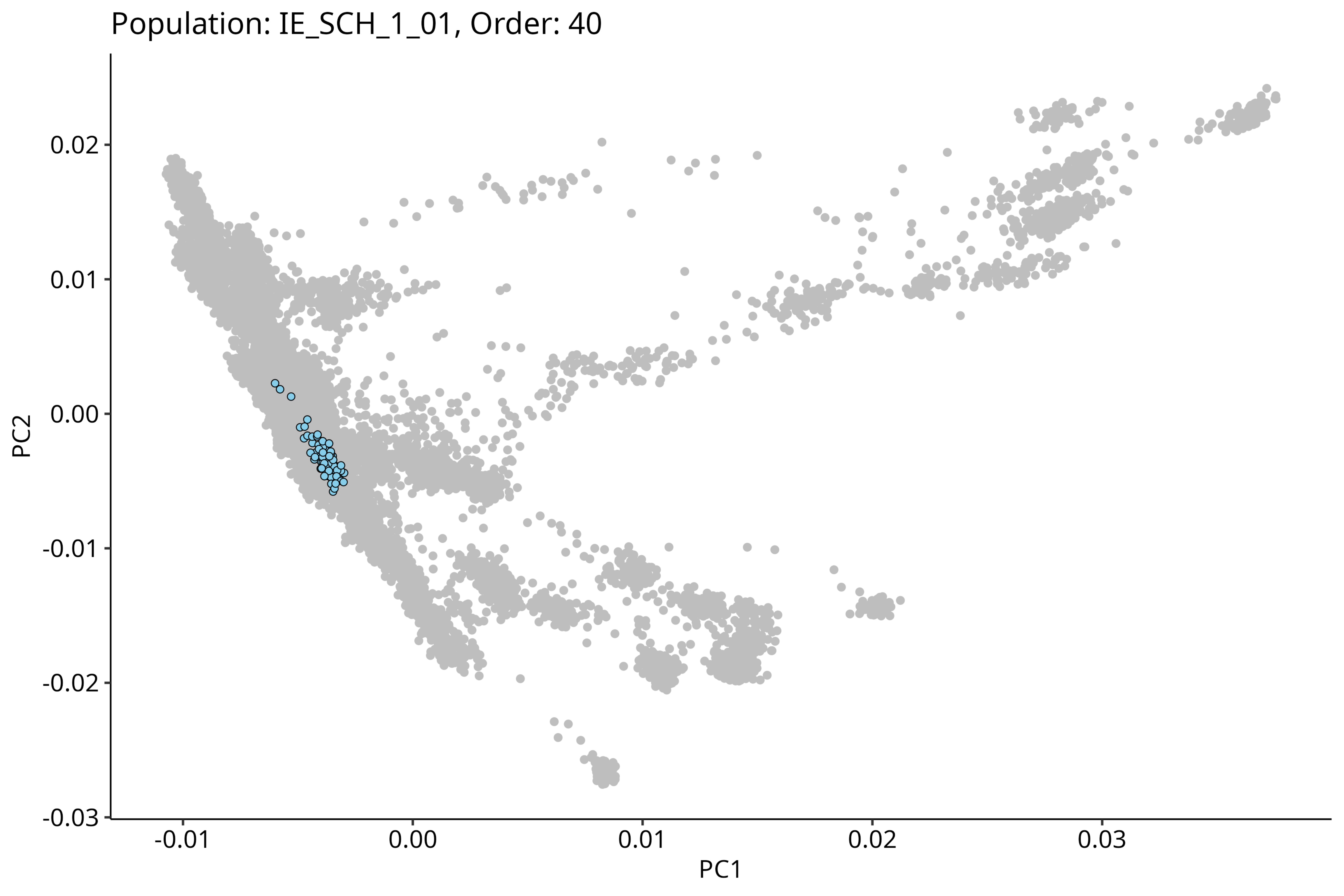
Population: IE_SCH_1_01, Order: 40
PC2
0.02
0.01
0.00
-0.01
-0.02
-0.03
-0.01
0.00
0.01
0.02
0.03
PC1

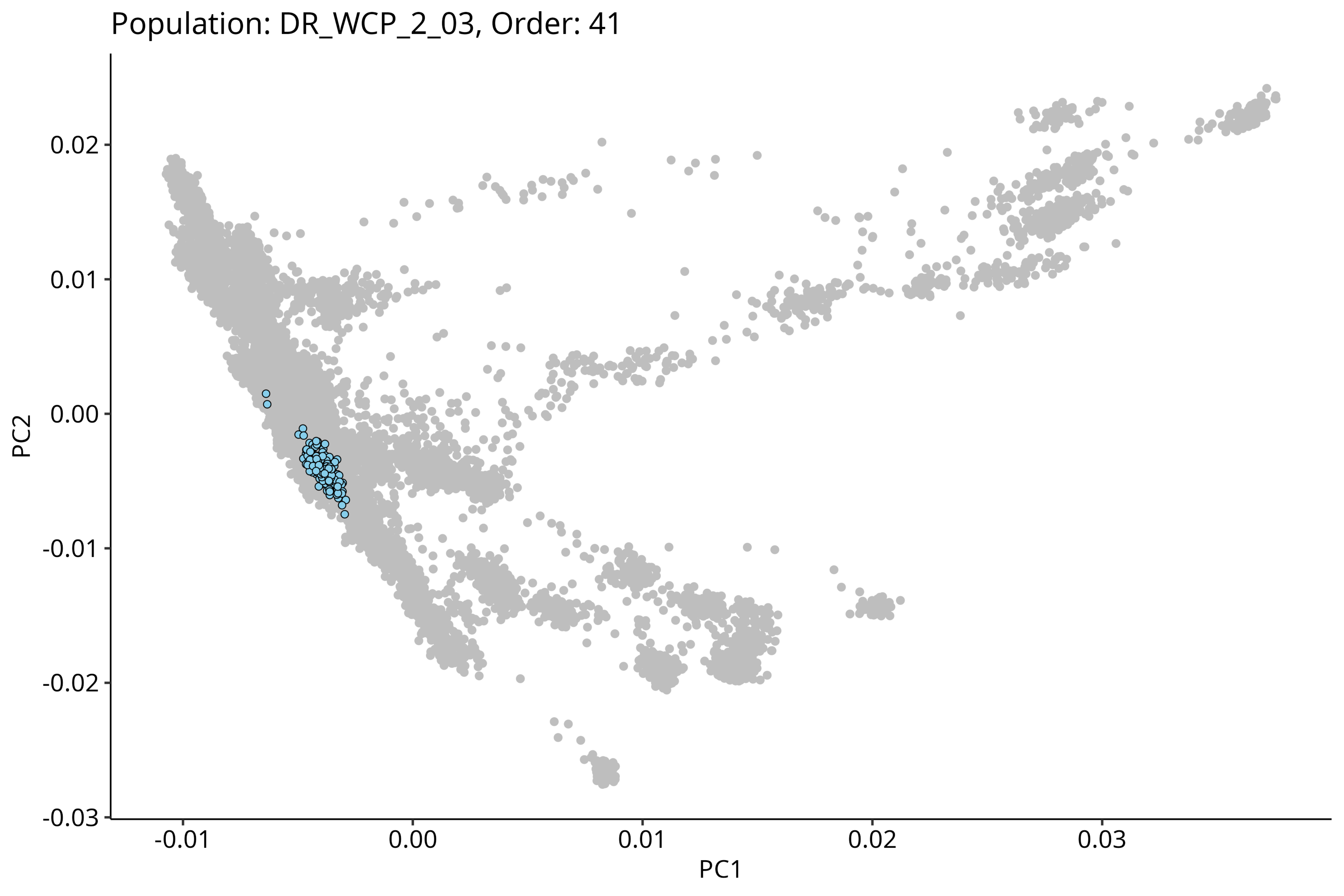
Population: DR_WCP_2_03, Order: 41
0.02
0.01
0.00
-0.01
-0.02
-0.03
PC2
-0.01
0.00
0.01
0.02
0.03
PC1

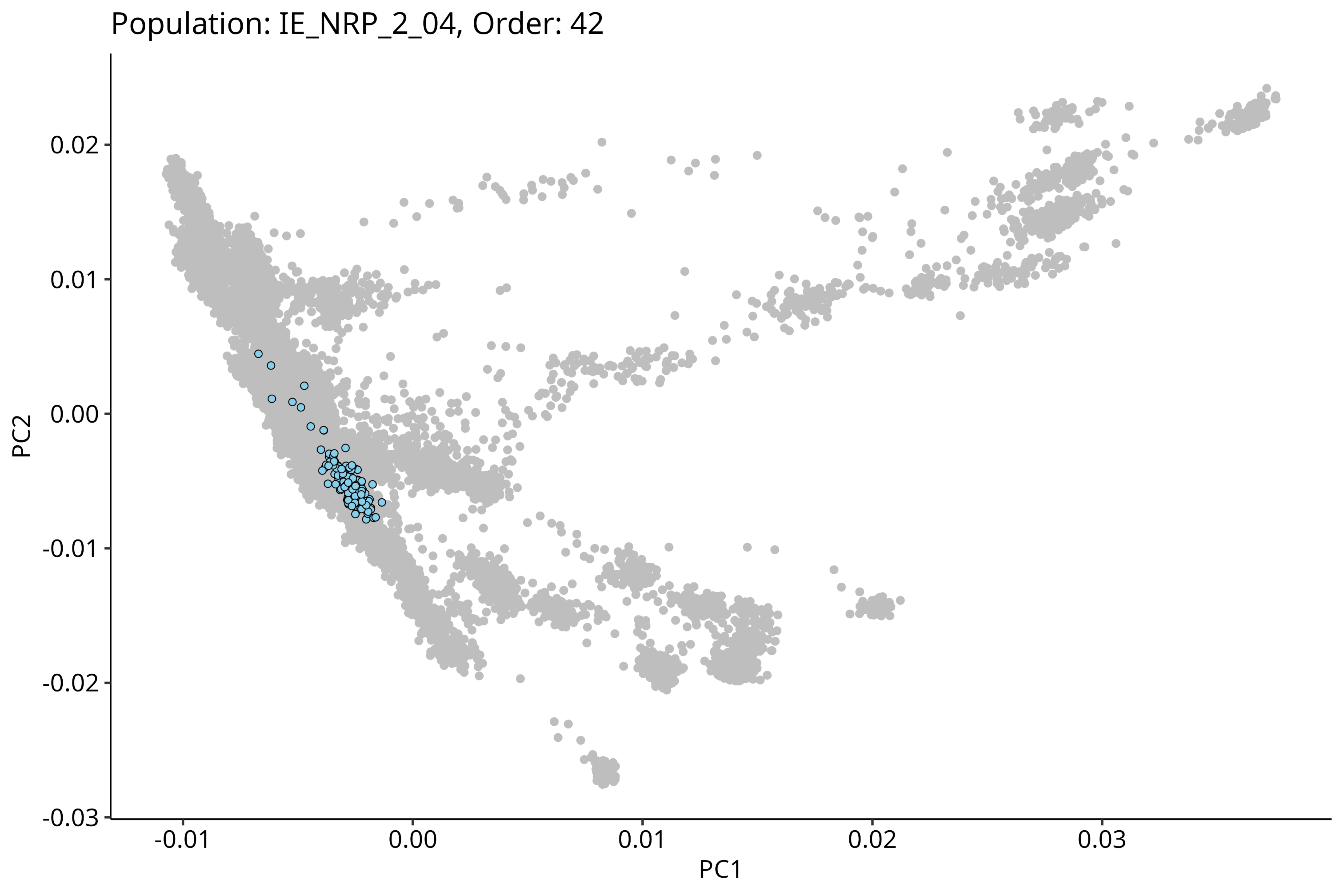
Population: IE_NRP_2_04, Order: 42
PC2
PC1
0.02
0.01
0.00
-0.01
-0.02
-0.03
-0.01
0.00
0.01
0.02
0.03

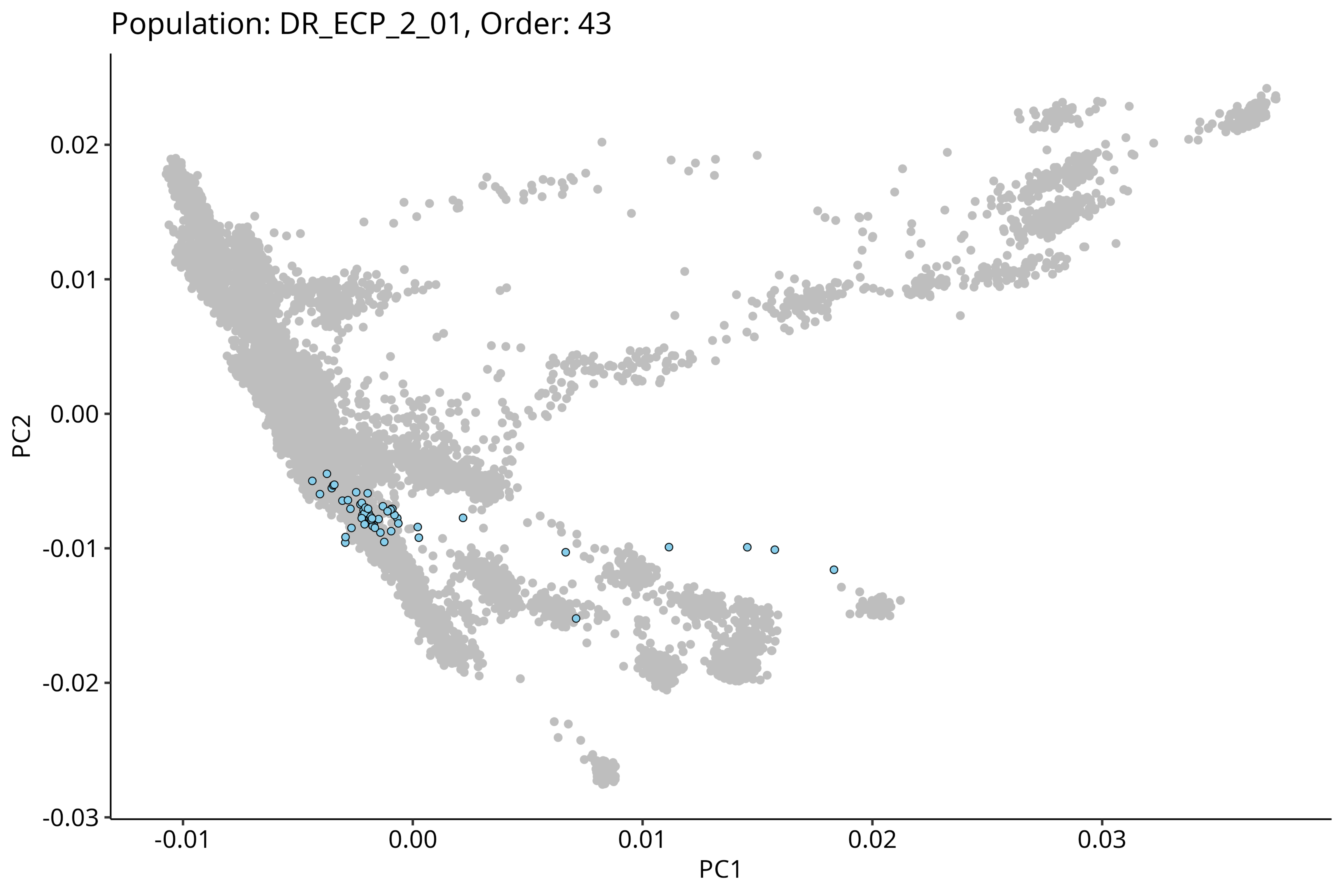
Population: DR_ECP_2_01, Order: 43
0.02
0.01
0.00
-0.01
-0.02
-0.03
PC2
-0.01
0.00
0.01
0.02
0.03
PC1

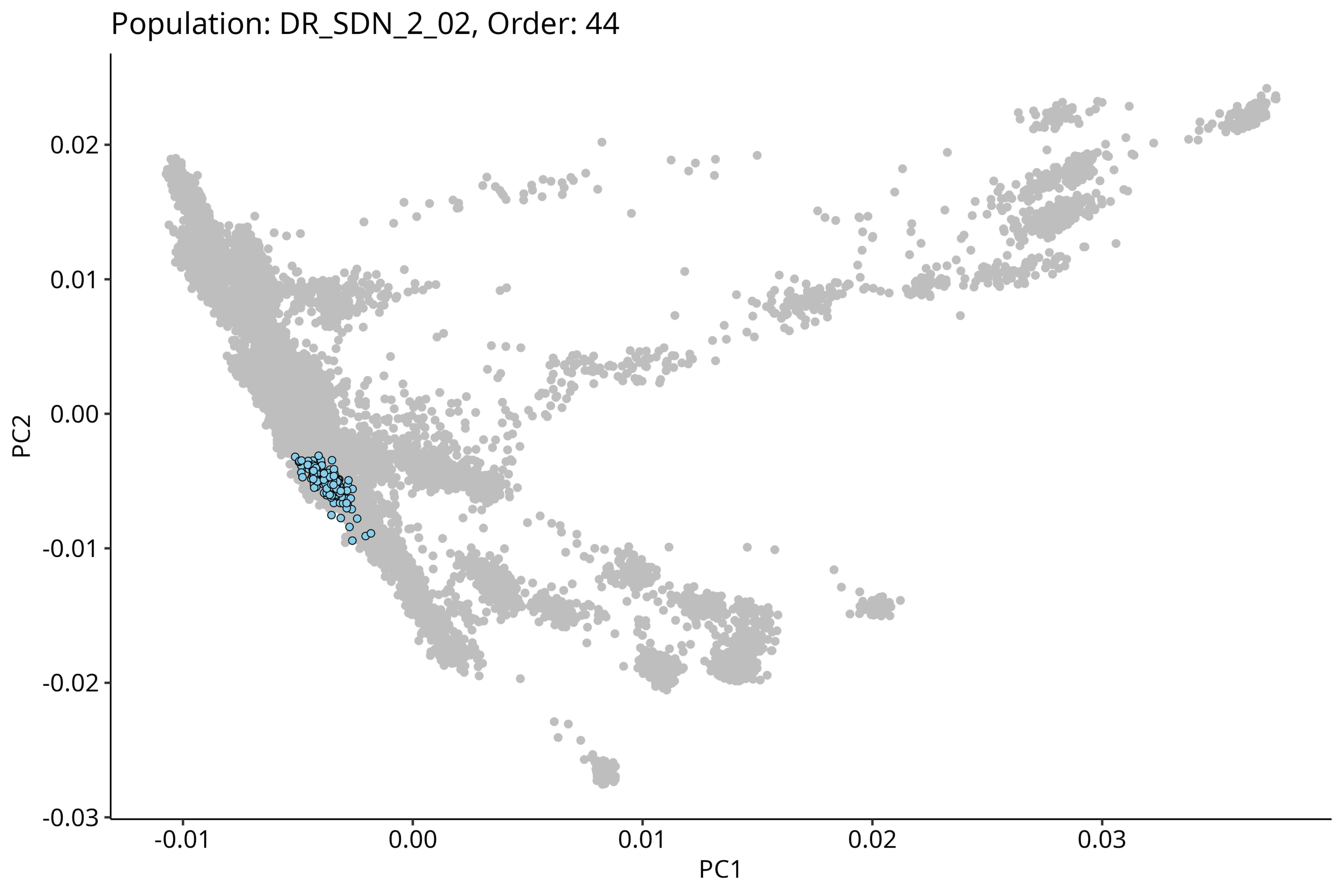
Population: DR_SDN_2_02, Order: 44
PC2
0.02
0.01
0.00
-0.01
-0.02
-0.03
PC1
-0.01
0.00
0.01
0.02
0.03

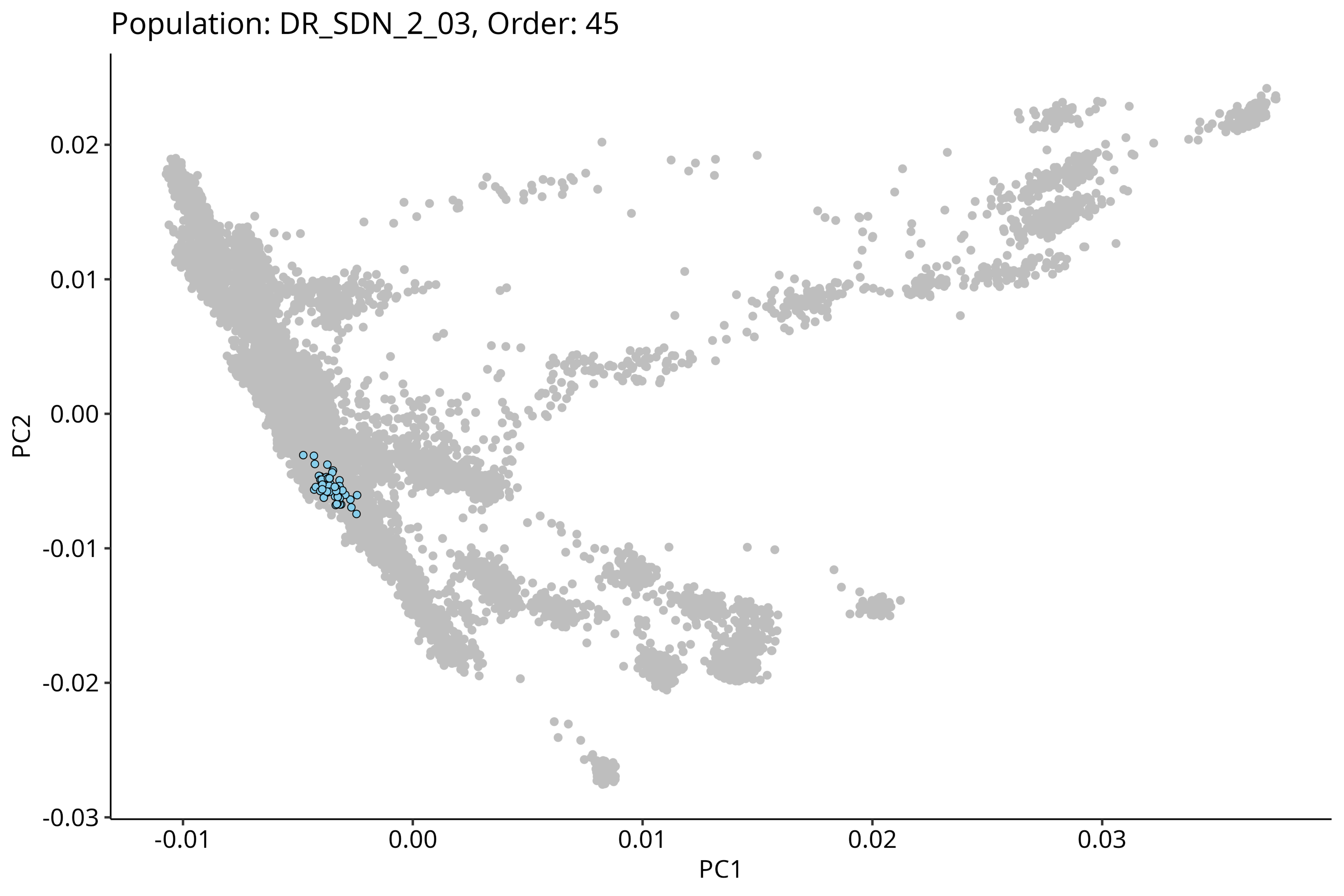
Population: DR_SDN_2_03, Order: 45
0.02
0.01
0.00
-0.01
-0.02
-0.03
PC2
-0.01
0.00
0.01
0.02
0.03
PC1

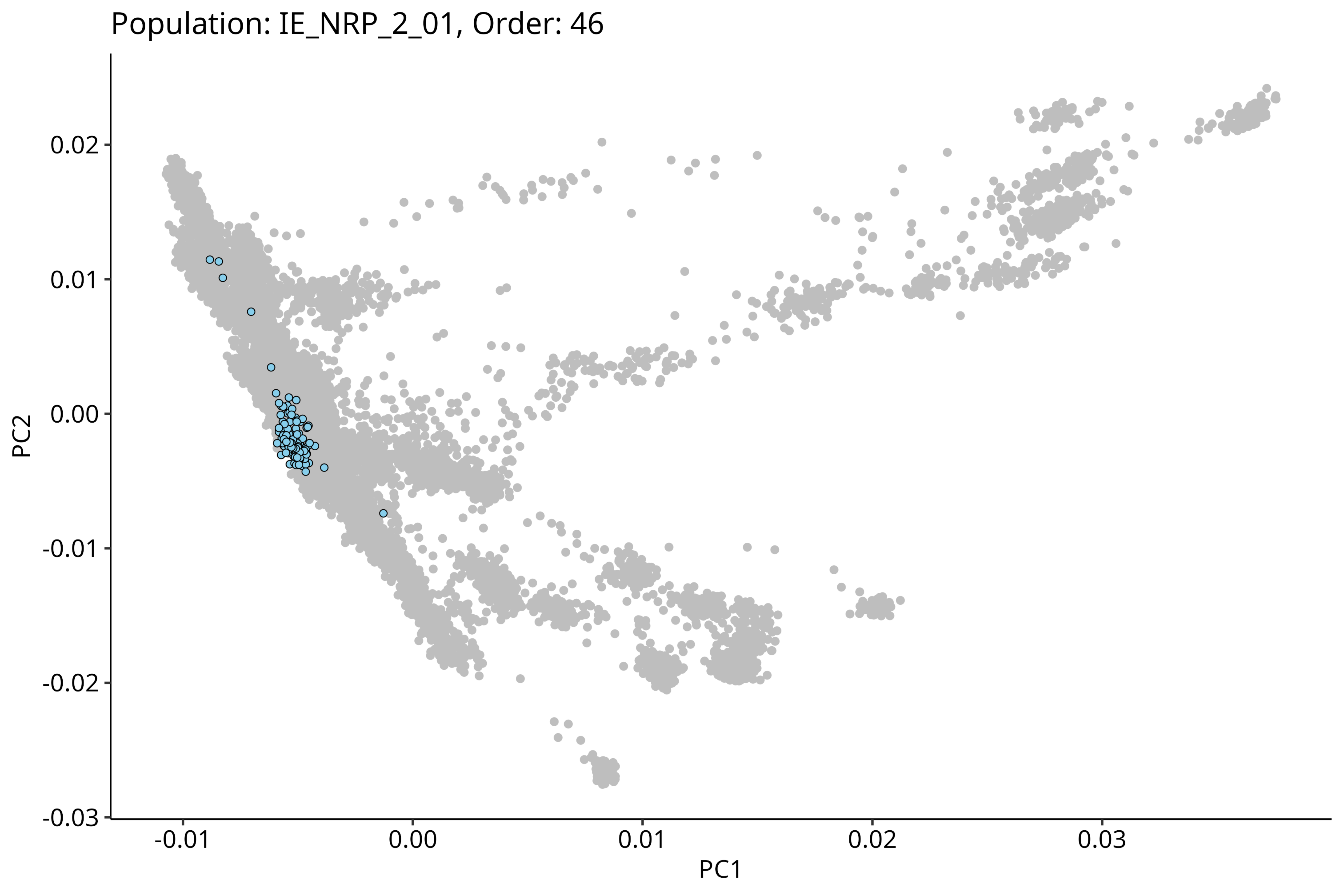
Population: IE_NRP_2_01, Order: 46
PC2
0.02
0.01
0.00
-0.01
-0.02
-0.03
PC1
-0.01
0.00
0.01
0.02
0.03

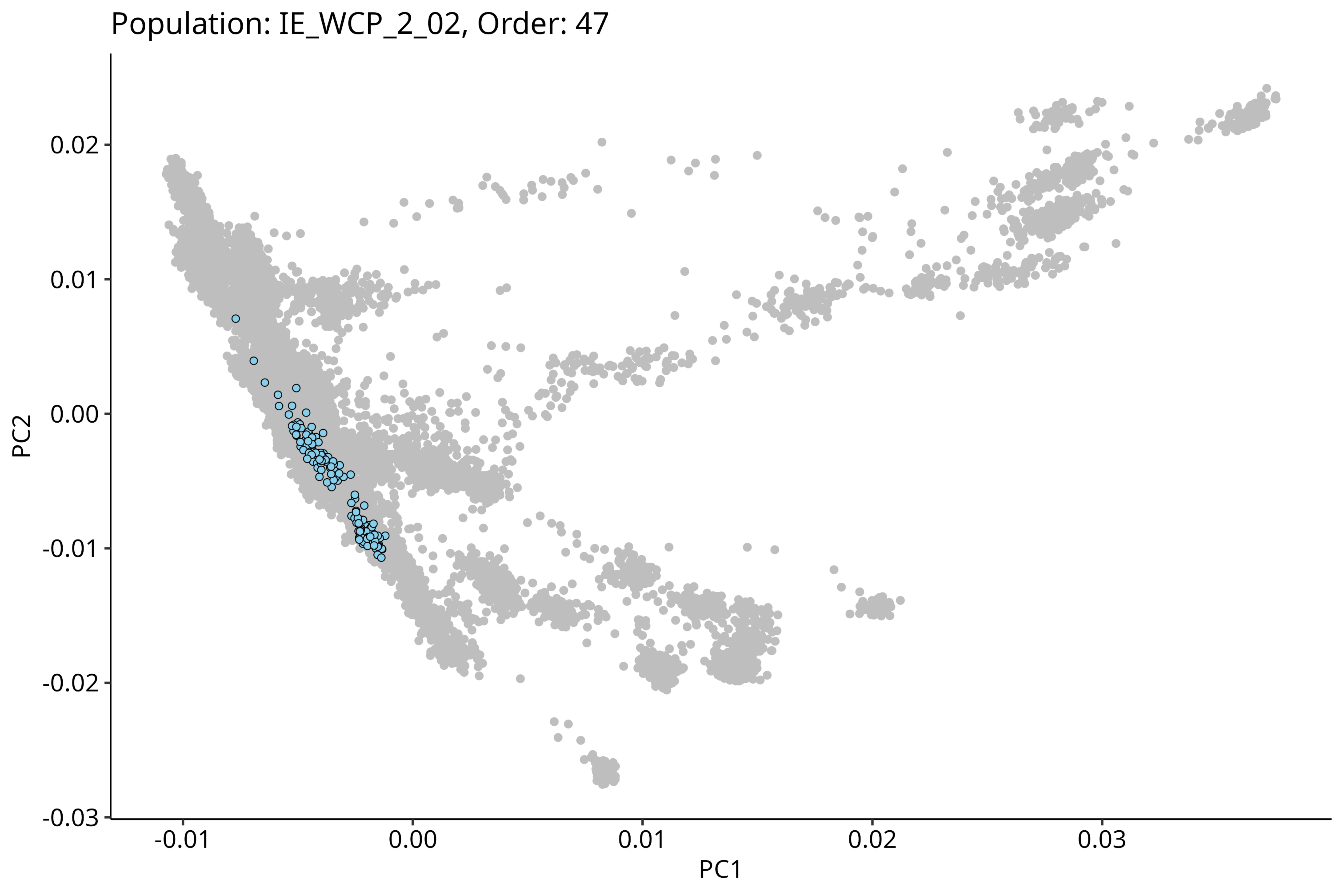

Population: IE_WCP_2_02, Order: 47
PC2
0.02
0.01
0.00
-0.01
-0.02
-0.03
PC1
-0.01
0.00
0.01
0.02
0.03

Population: IE_NRP_2_09, Order: 48

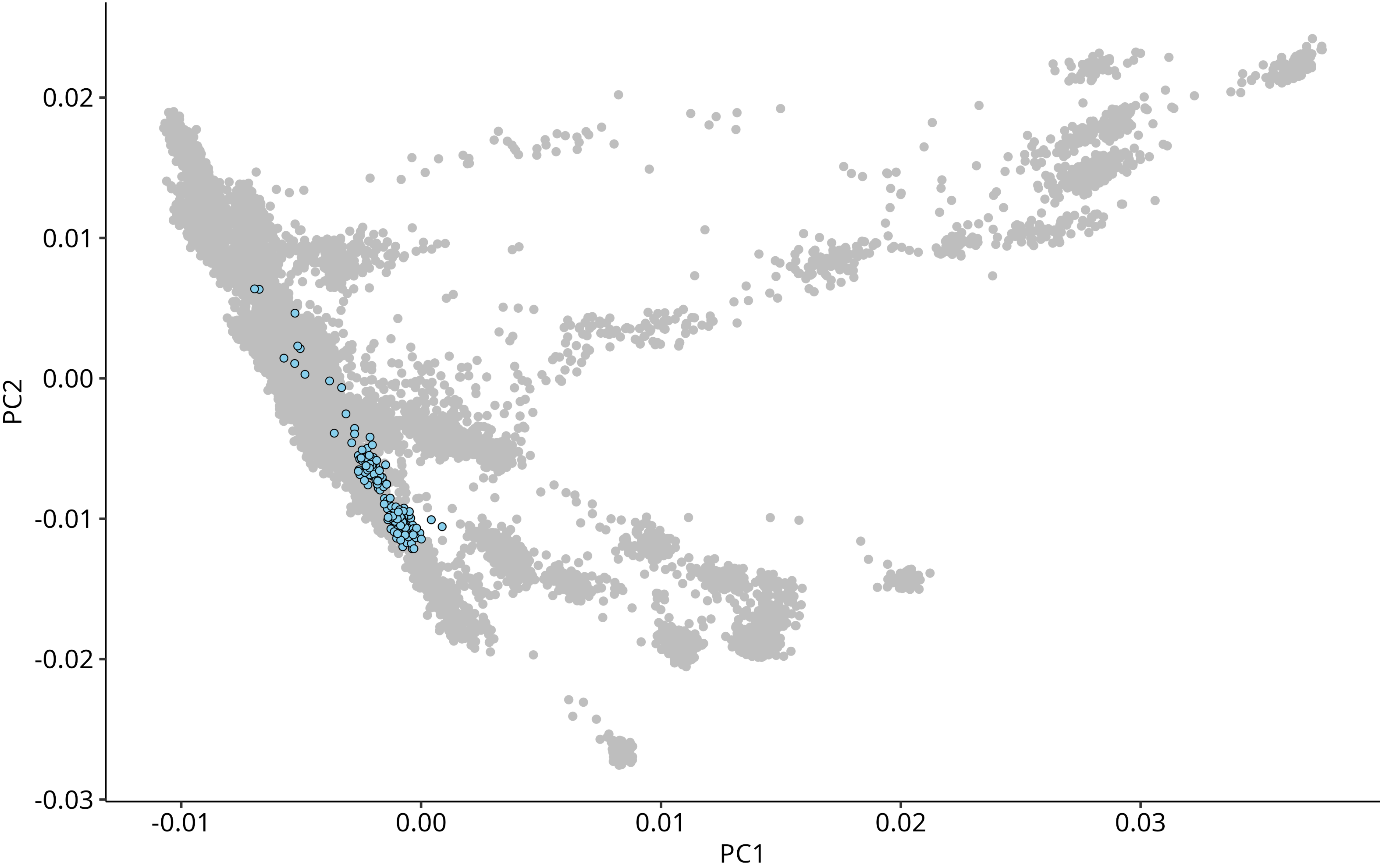

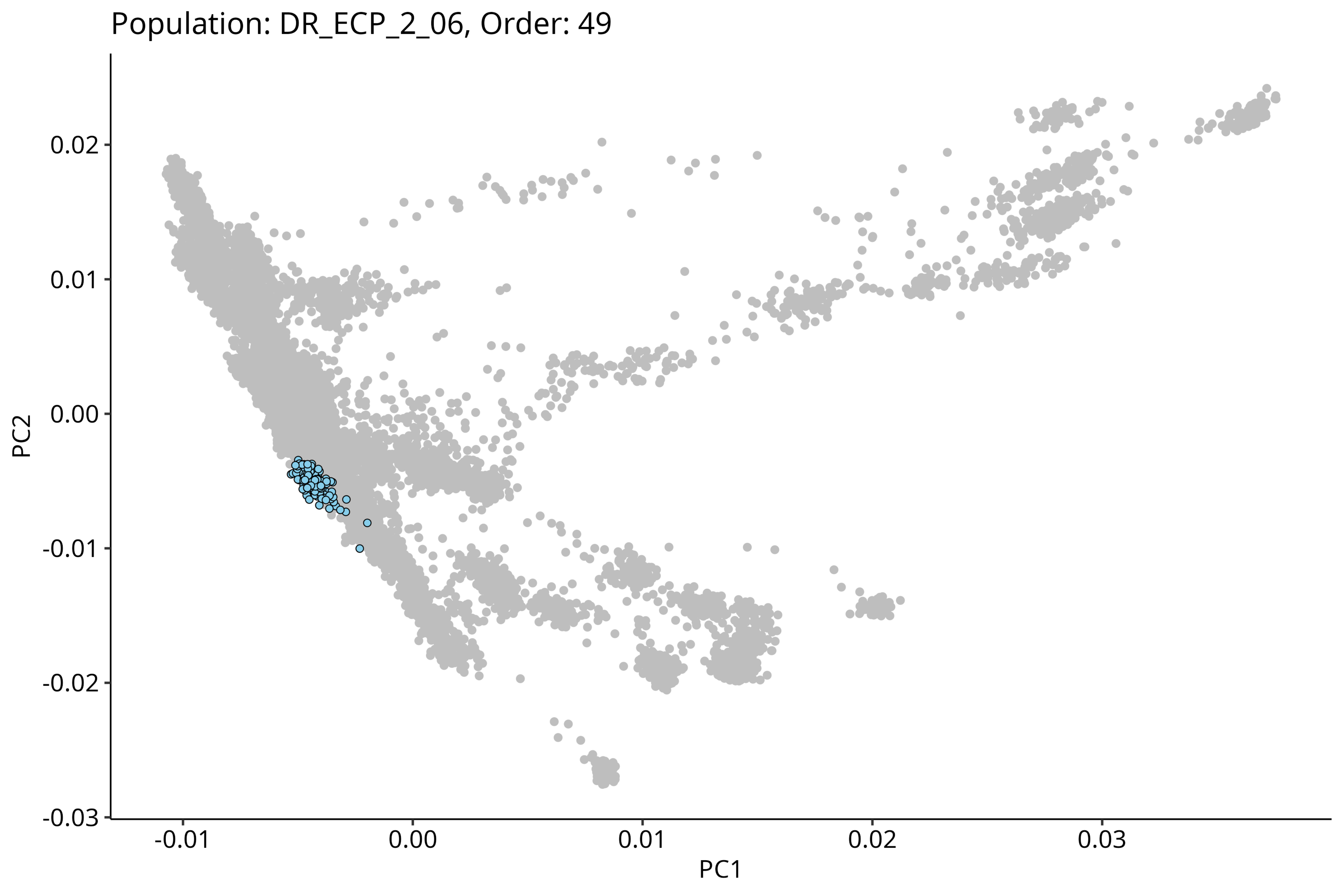
Population: DR_ECP_2_06, Order: 49
PC2
0.02
0.01
0.00
-0.01
-0.02
-0.03
PC1
-0.01
0.00
0.01
0.02
0.03

Population: DR_ECP_2_05, Order: 50

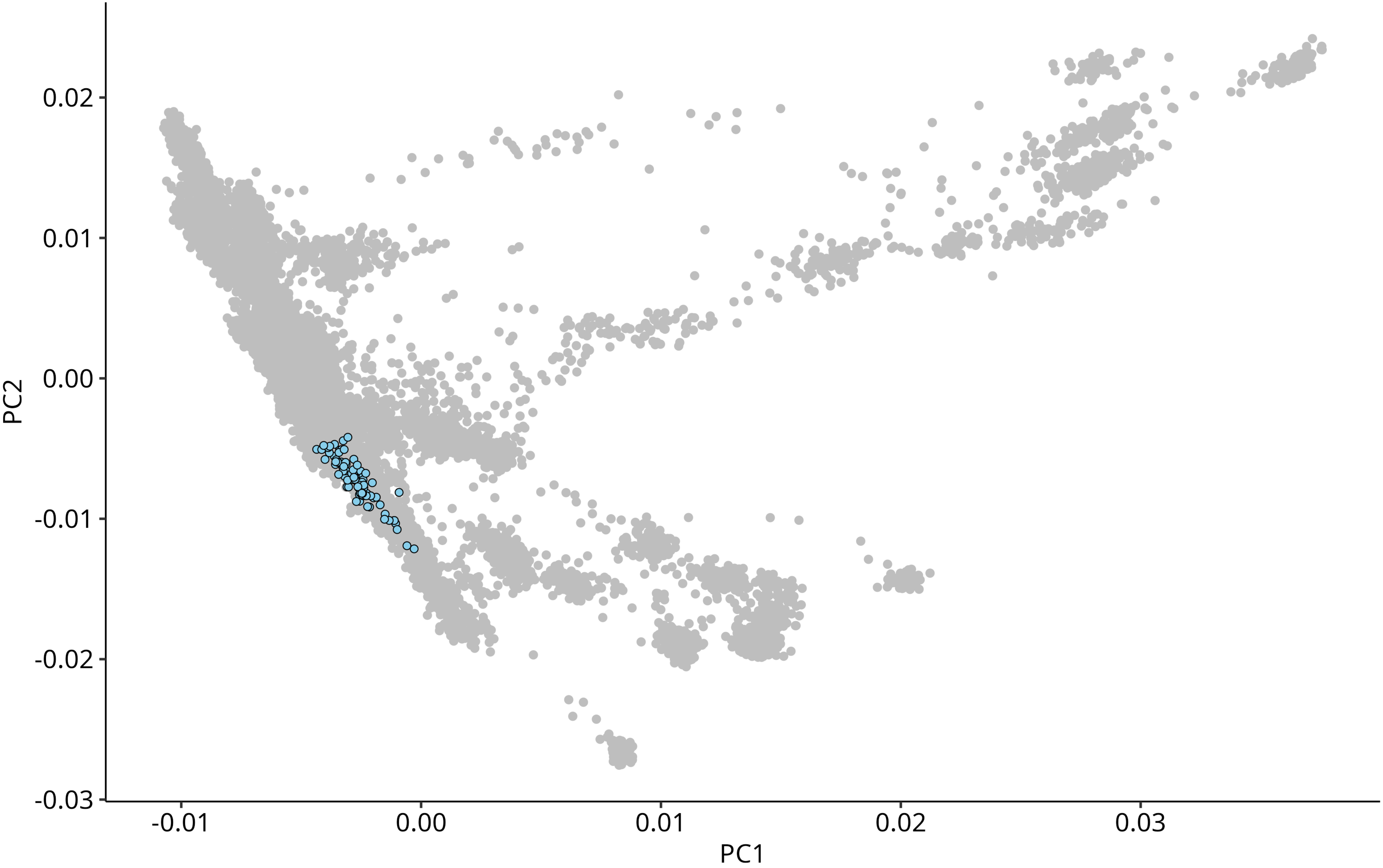

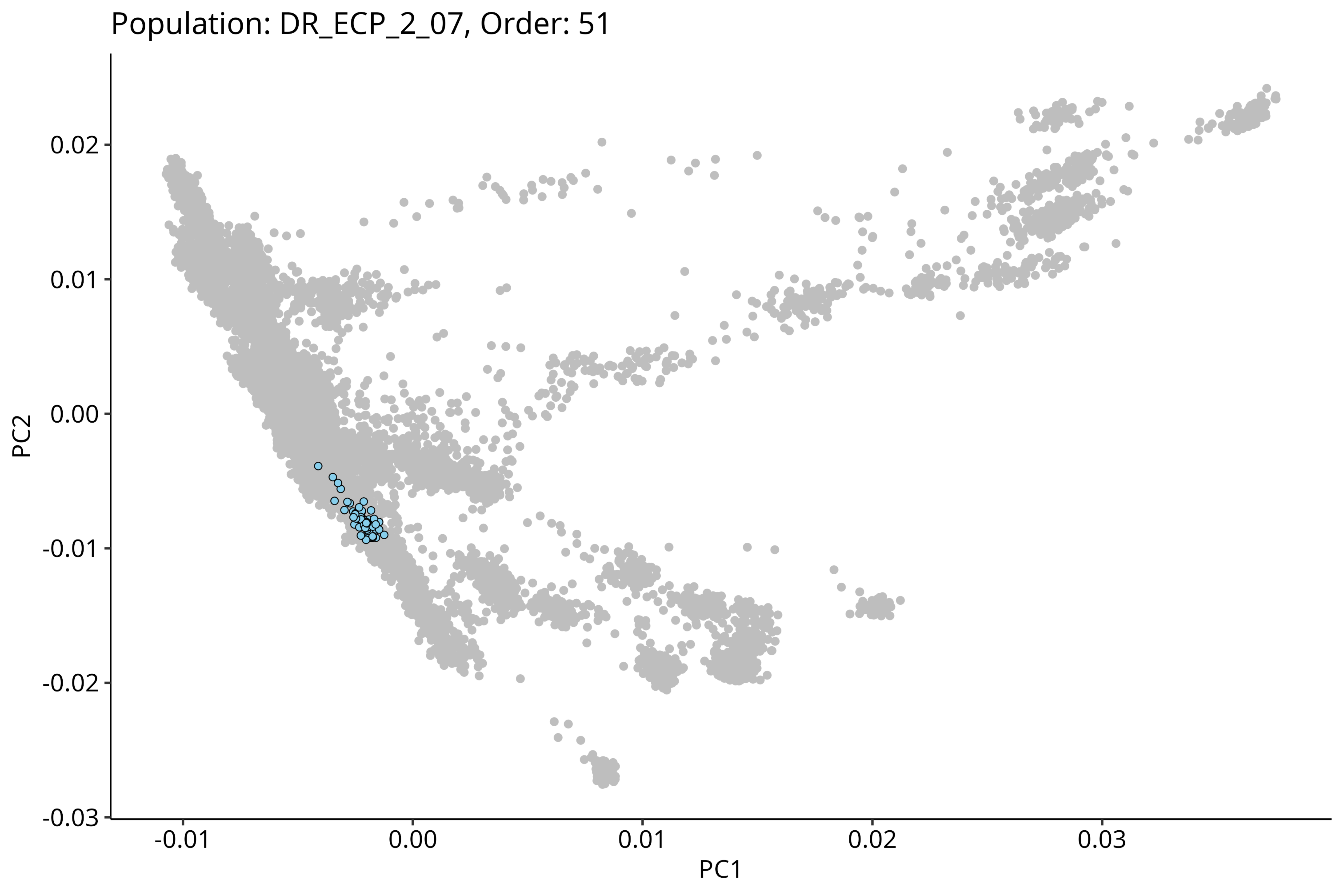
Population: DR_ECP_2_07, Order: 51
0.02
0.01
0.00
-0.01
-0.02
-0.03
PC2
-0.01
0.00
0.01
0.02
0.03
PC1

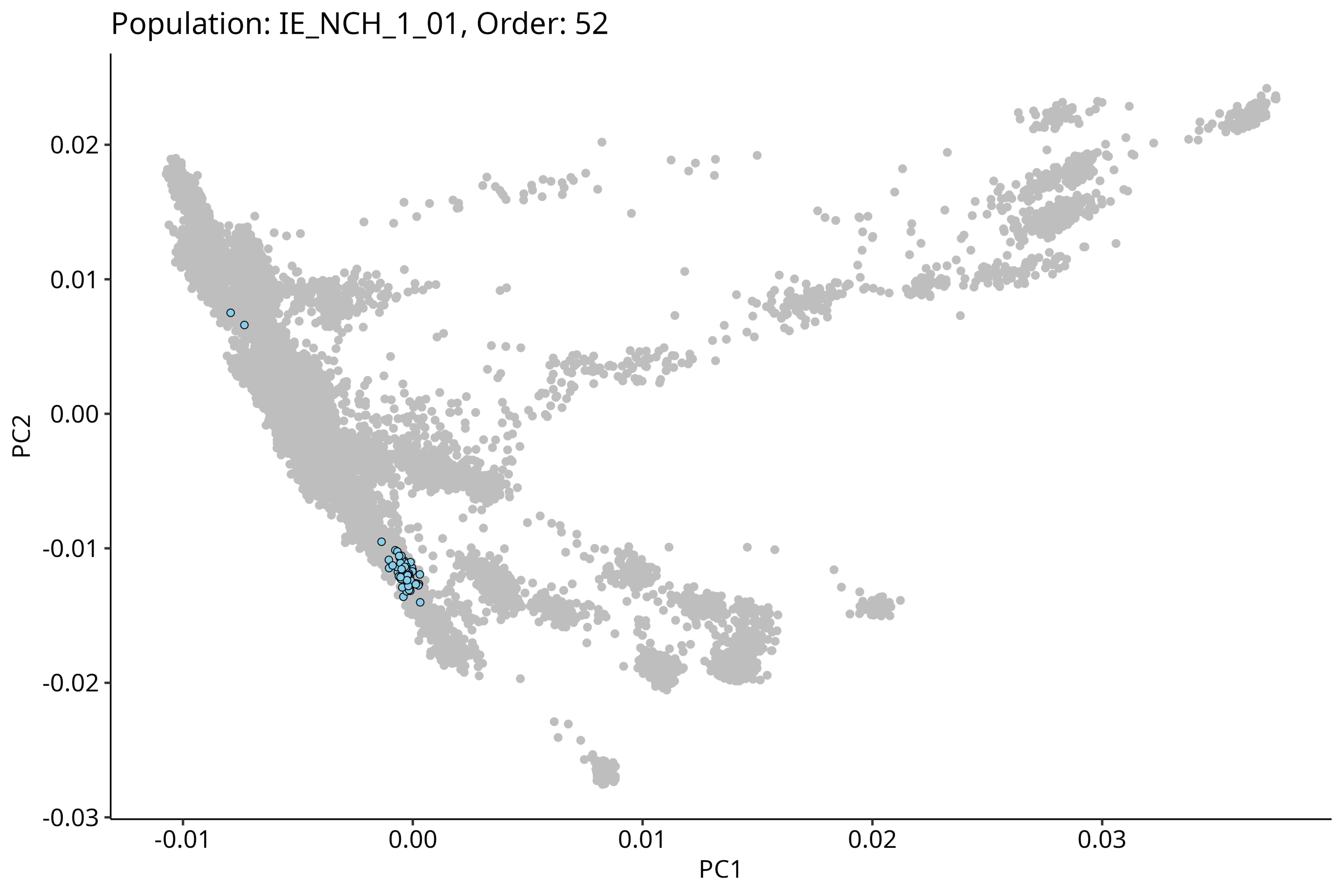
Population: IE_NCH_1_01, Order: 52
0.02
0.01
0.00
-0.01
-0.02
-0.03
PC2
-0.01
0.00
0.01
0.02
0.03
PC1

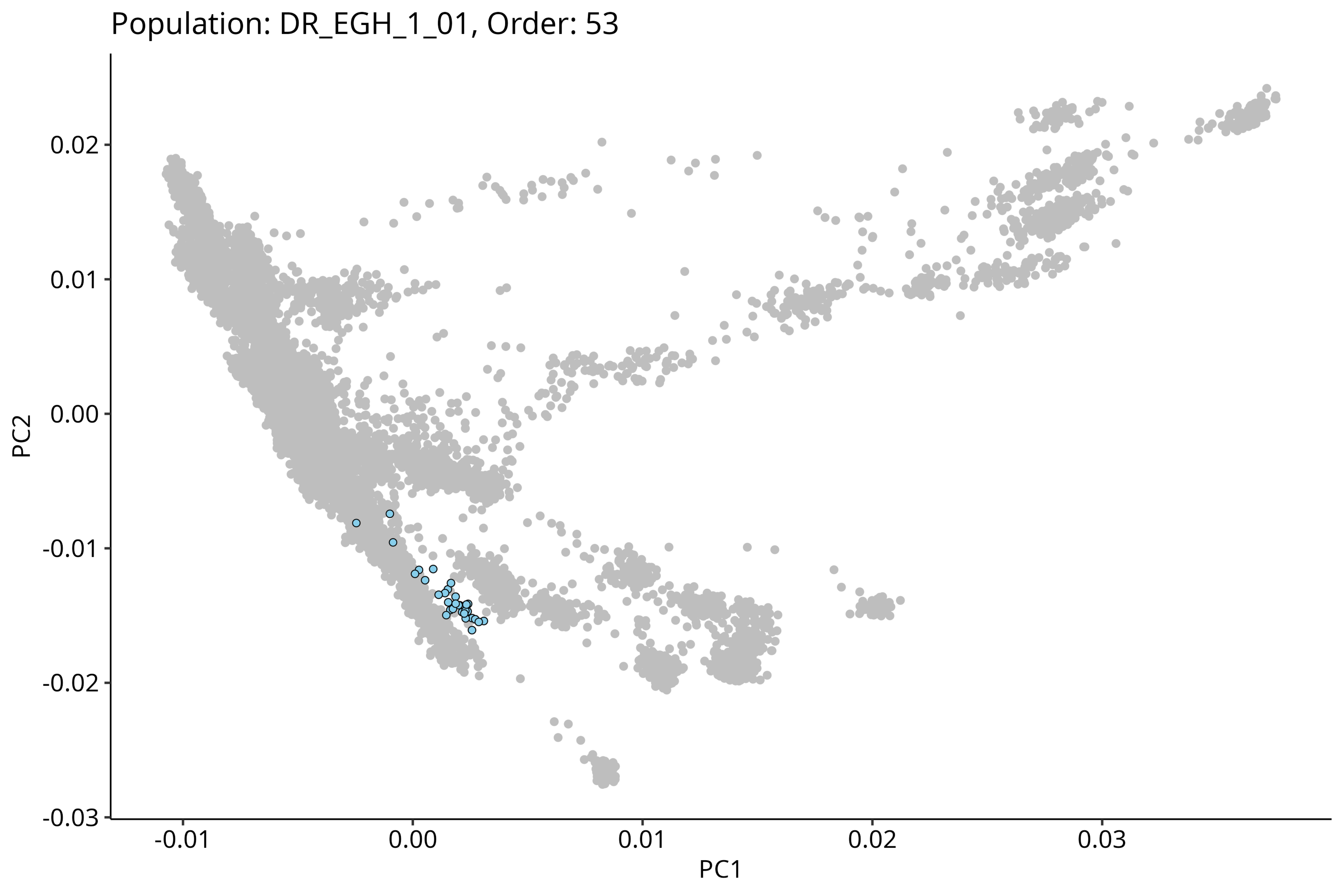
Population: DR_EGH_1_01, Order: 53
PC2
0.02
0.01
0.00
-0.01
-0.02
-0.03
-0.01
0.00
0.01
0.02
0.03
PC1

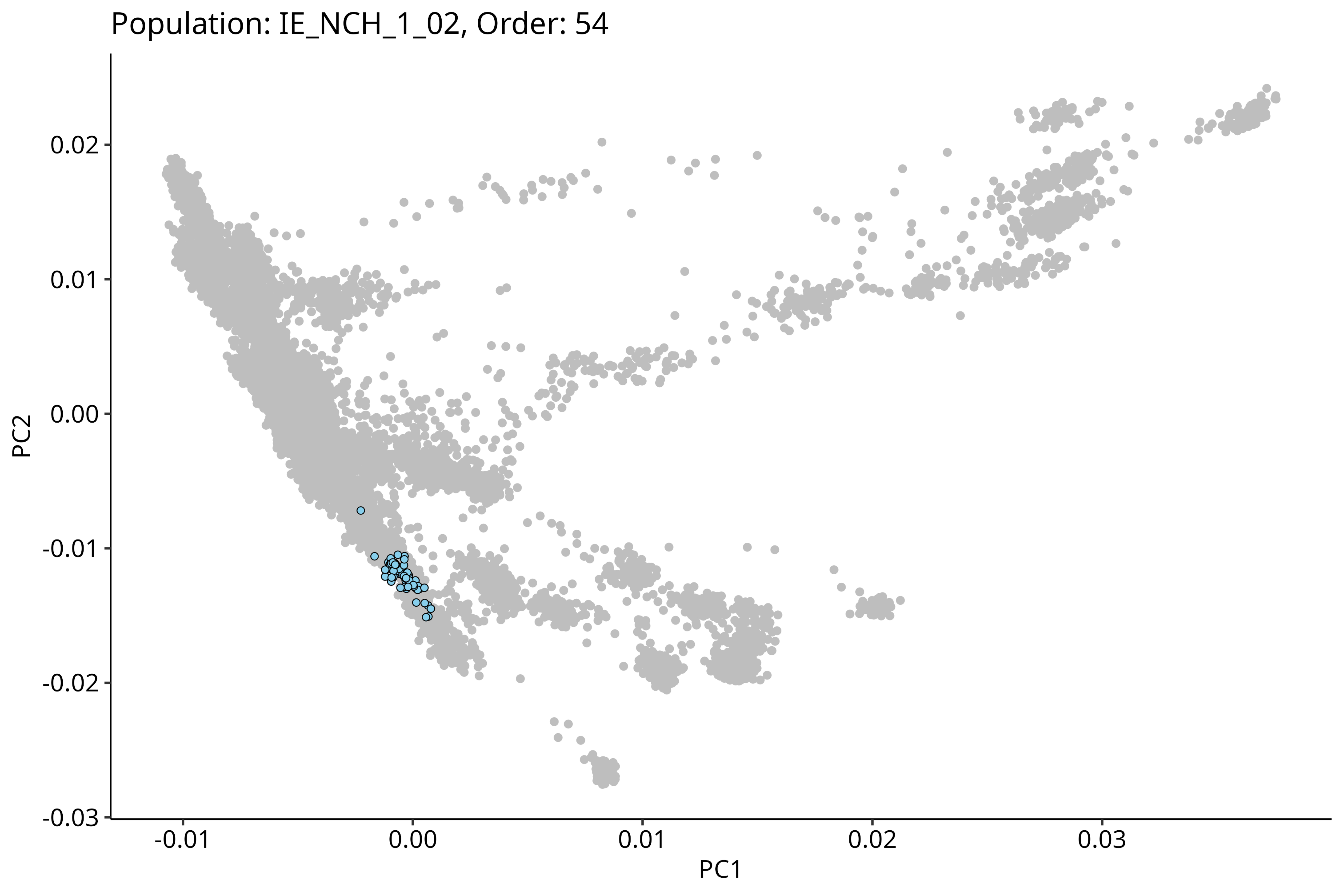

Population: IE_NCH_1_02, Order: 54
0.02
0.01
0.00
-0.01
-0.02
-0.03
PC2
-0.01
0.00
0.01
0.02
0.03
PC1

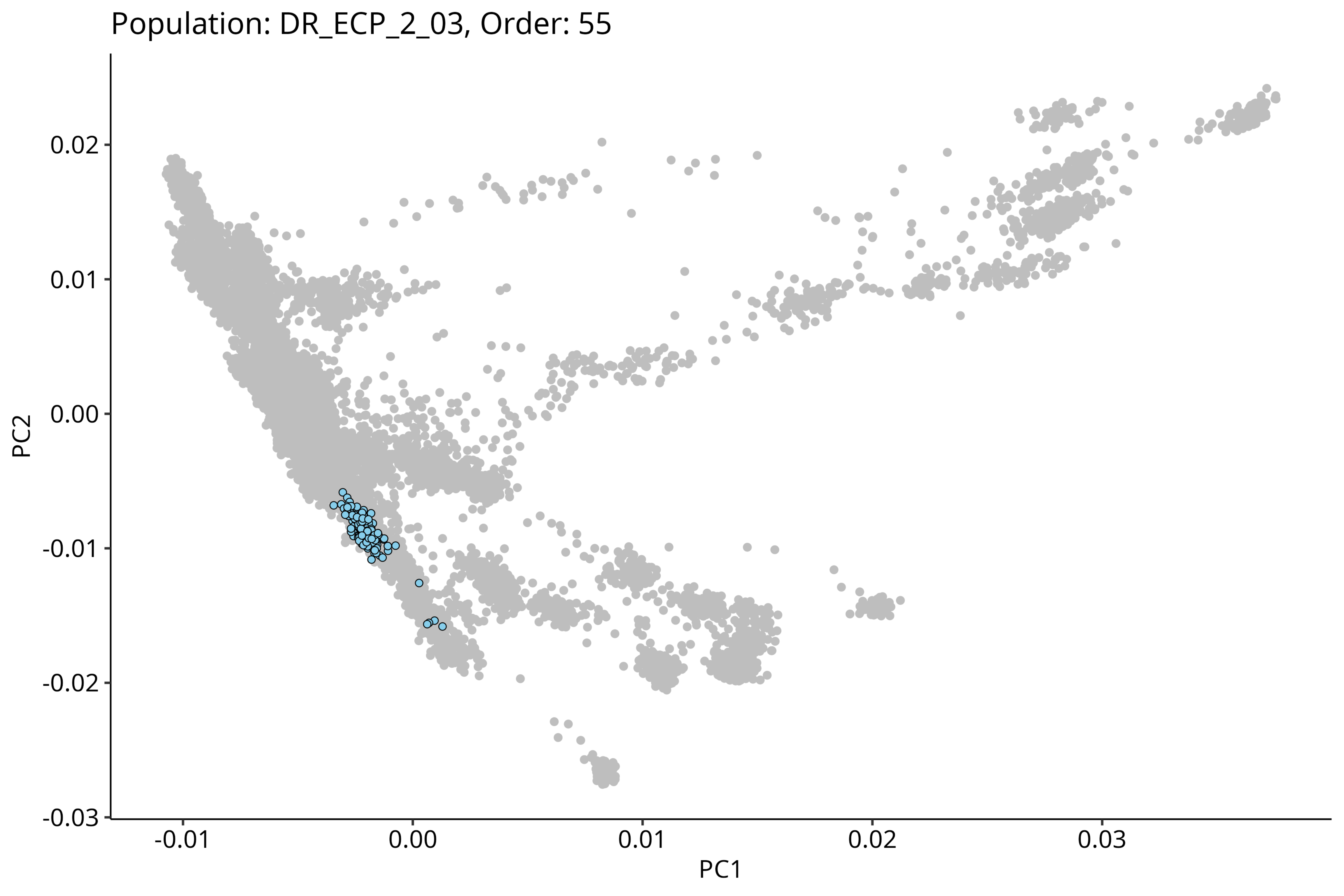

Population: DR_ECP_2_03, Order: 55
PC2
0.02
0.01
0.00
-0.01
-0.02
-0.03
PC1
-0.01
0.00
0.01
0.02
0.03

Population: DR_ECP_2_04, Order: 56

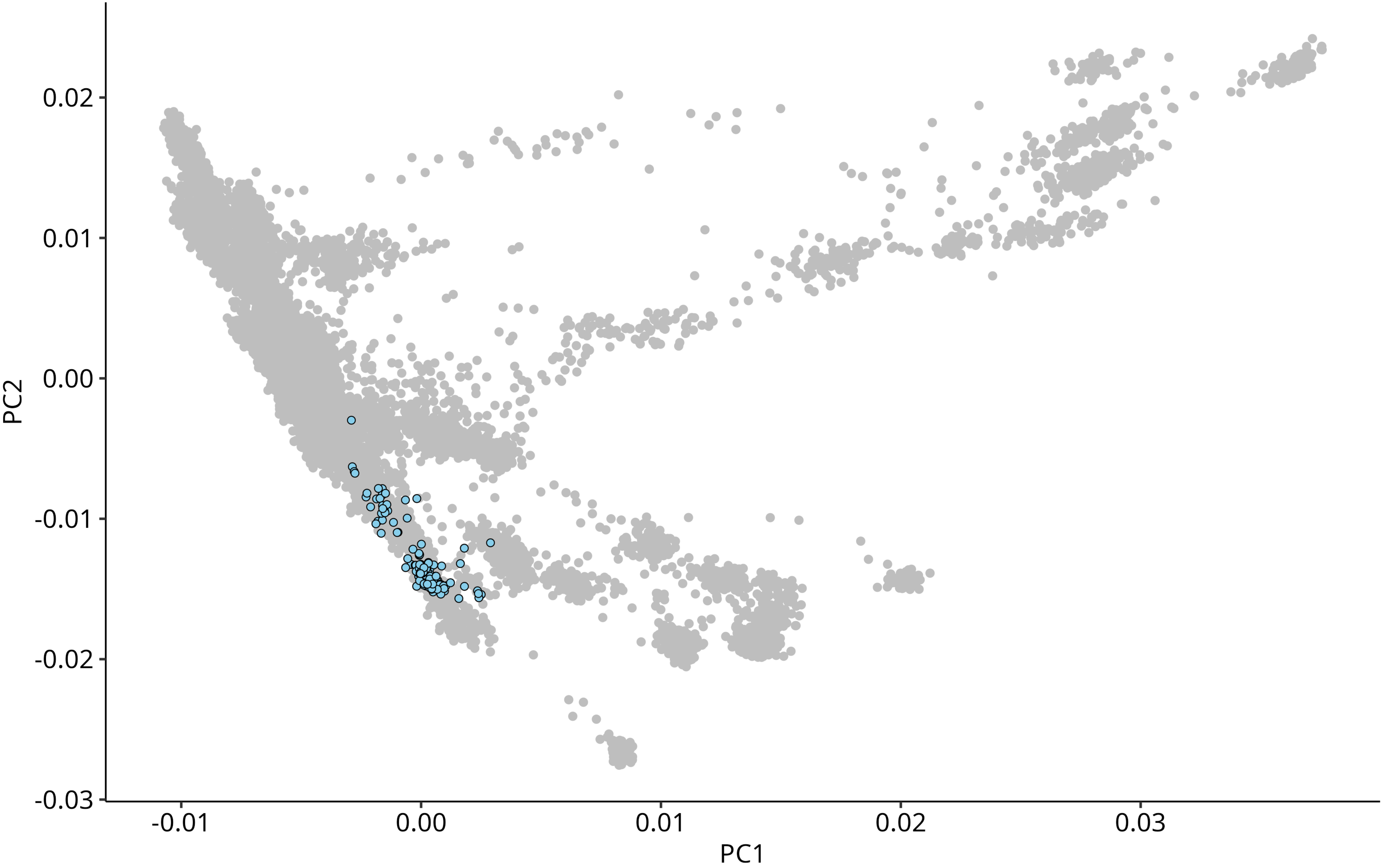

Population: DR_EGH_2_01, Order: 57

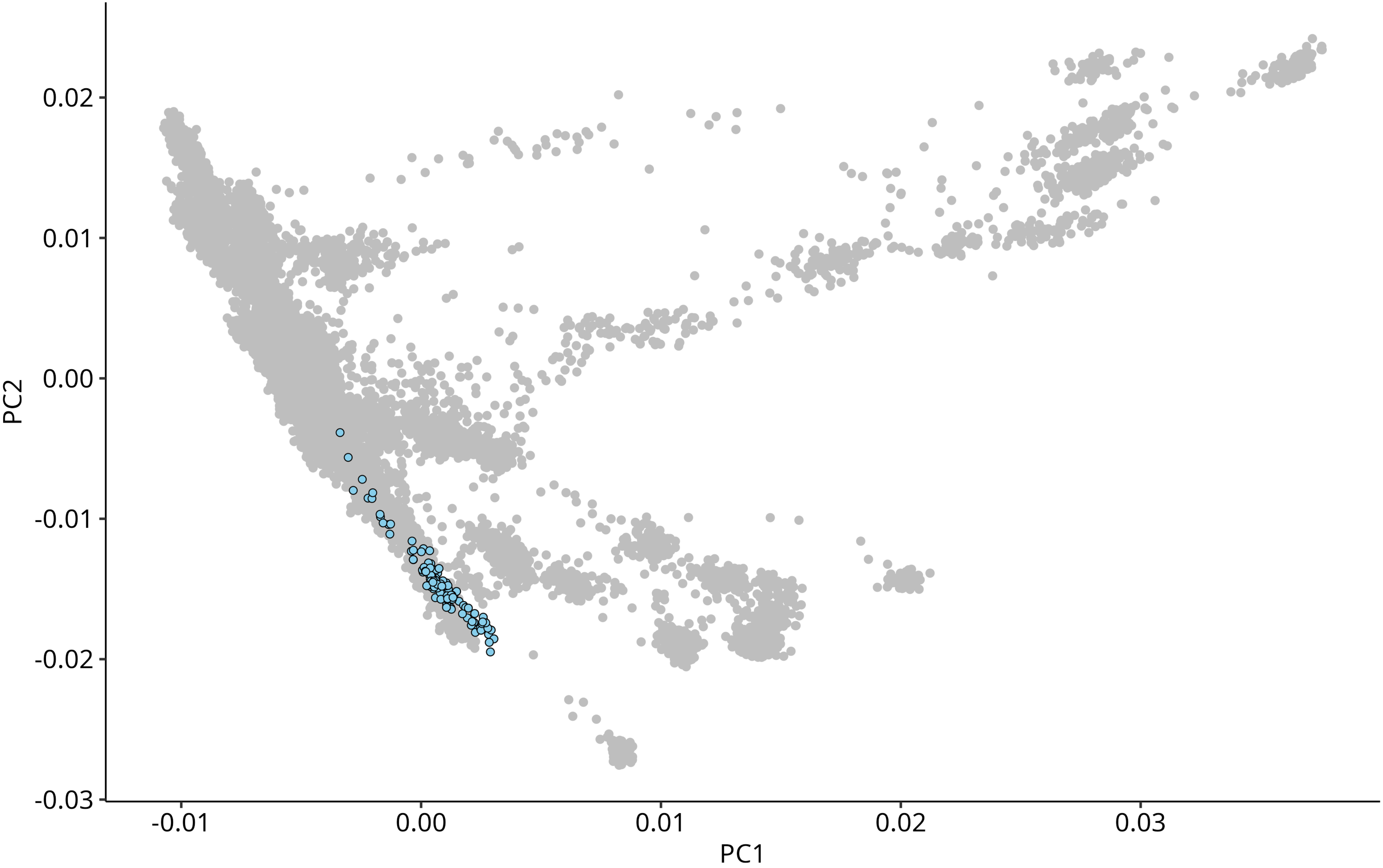

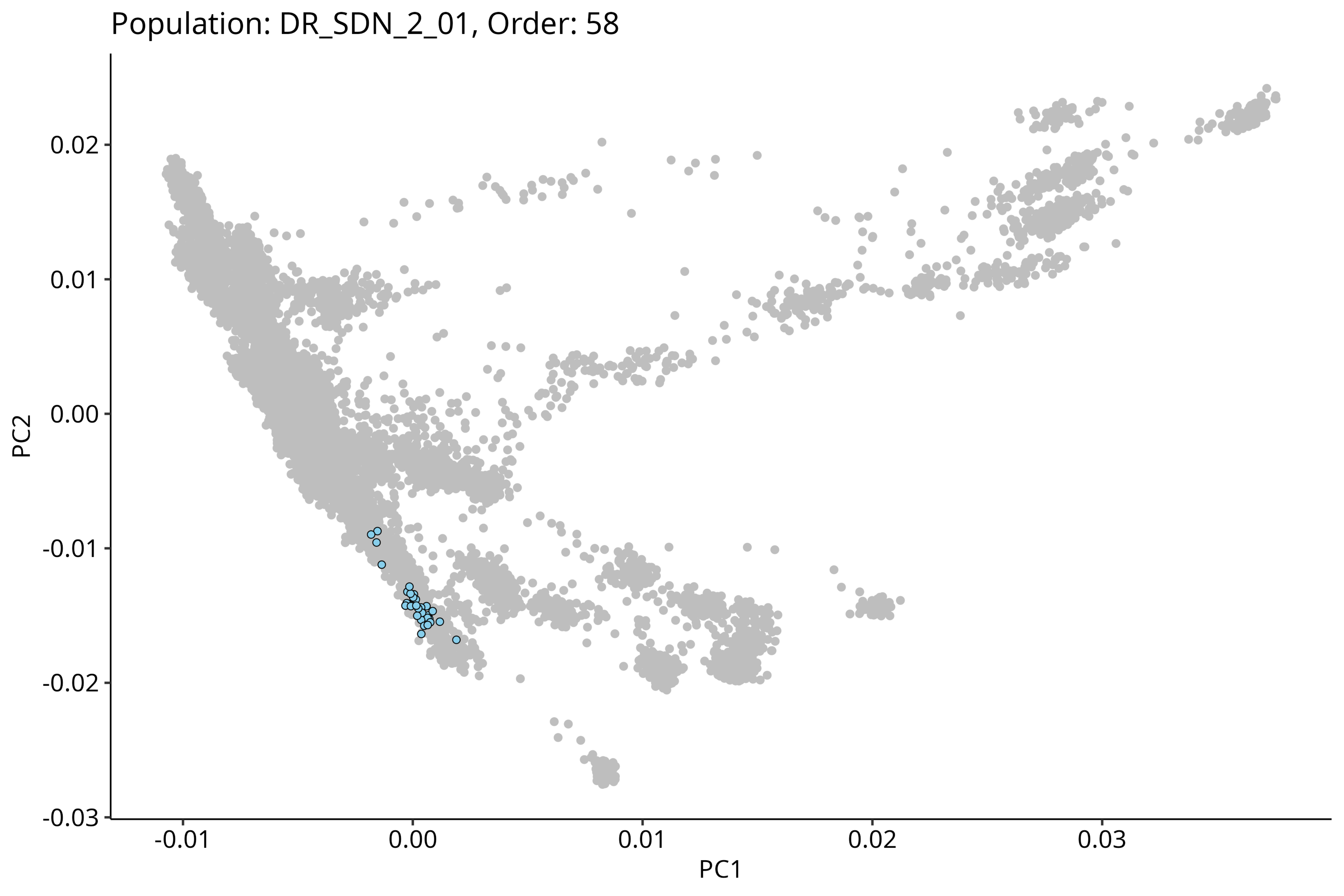
Population: DR_SDN_2_01, Order: 58
PC2
PC1
0.02
0.01
0.00
-0.01
-0.02
-0.03
-0.01
0.00
0.01
0.02
0.03

Population: DR_WGH_1_01, Order: 59

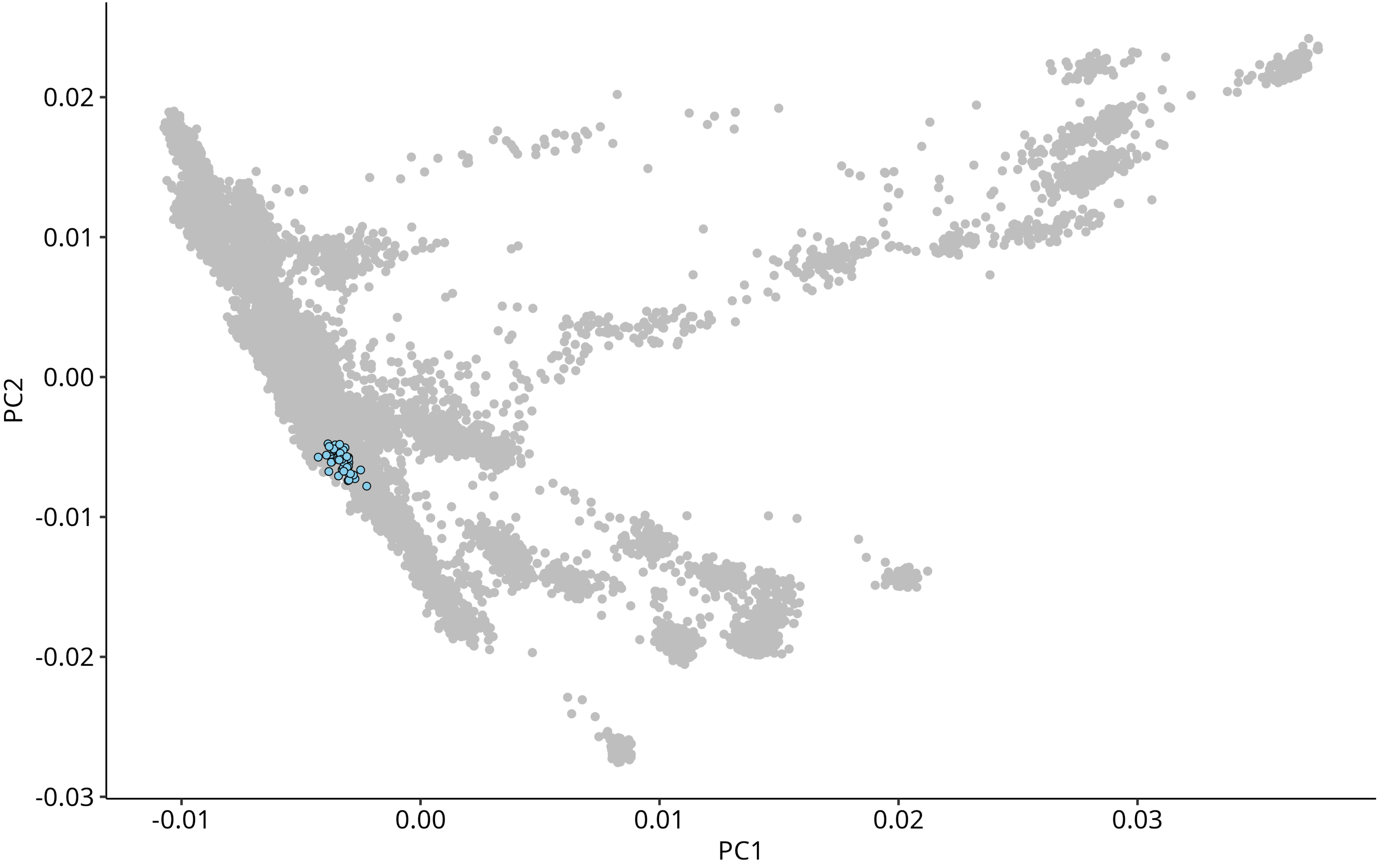

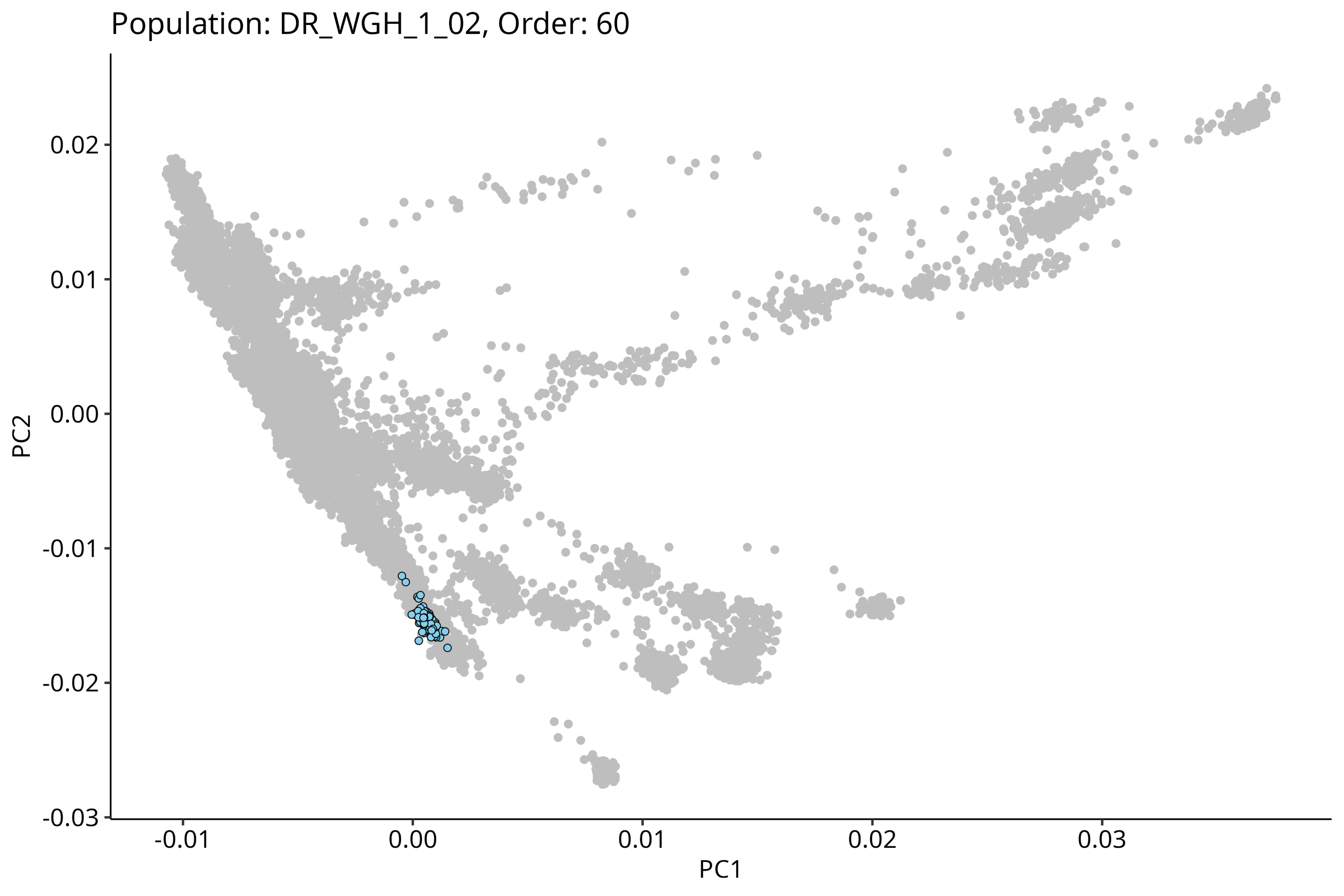
Population: DR_WGH_1_02, Order: 60
0.02
0.01
0.00
-0.01
-0.02
-0.03
PC2
-0.01
0.00
0.01
0.02
0.03
PC1

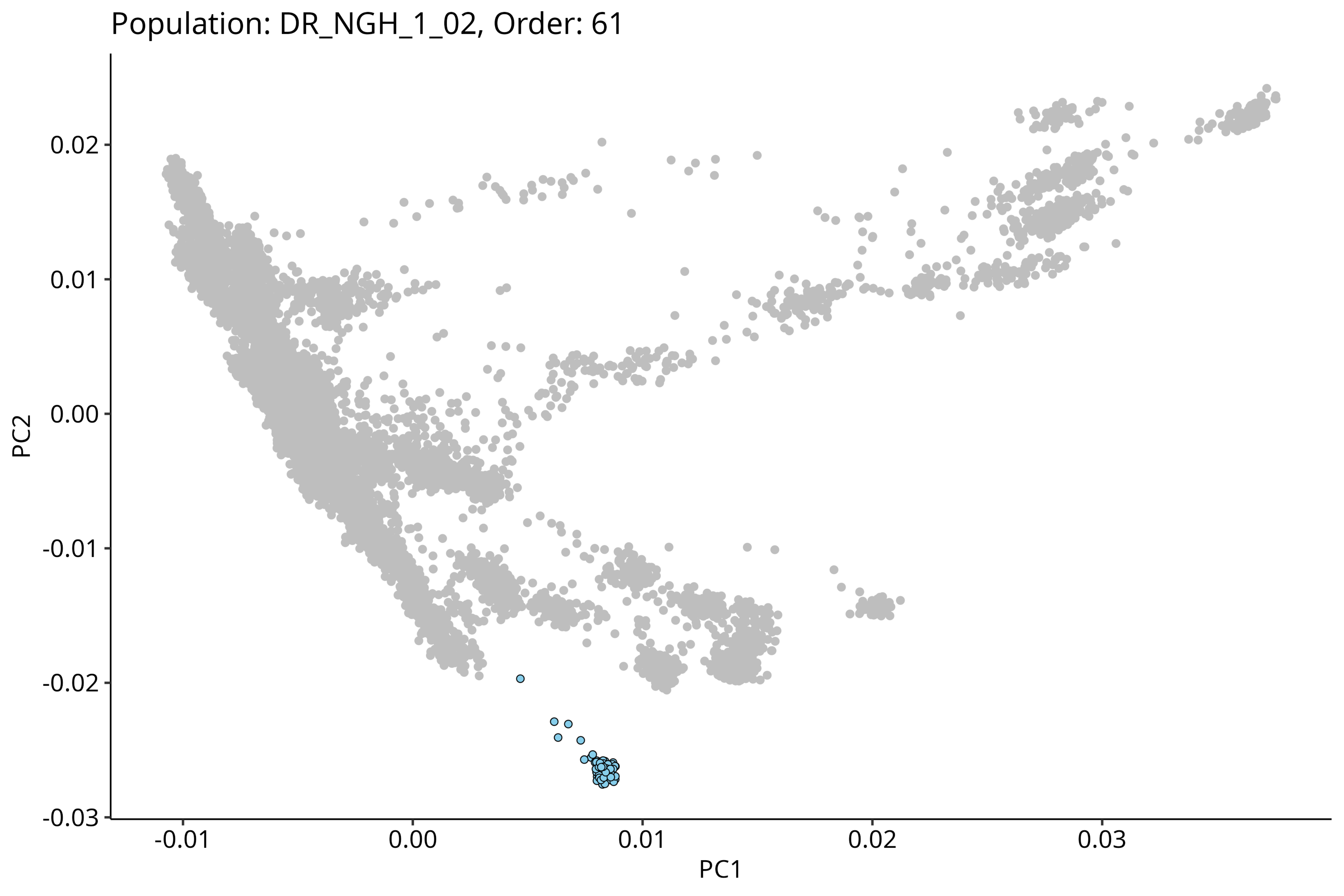
Population: DR_NGH_1_02, Order: 61
0.02
0.01
0.00
-0.01
-0.02
-0.03
PC2
-0.01
0.00
0.01
0.02
0.03
PC1

### Population: DR_NGH_1_01, Order: 62

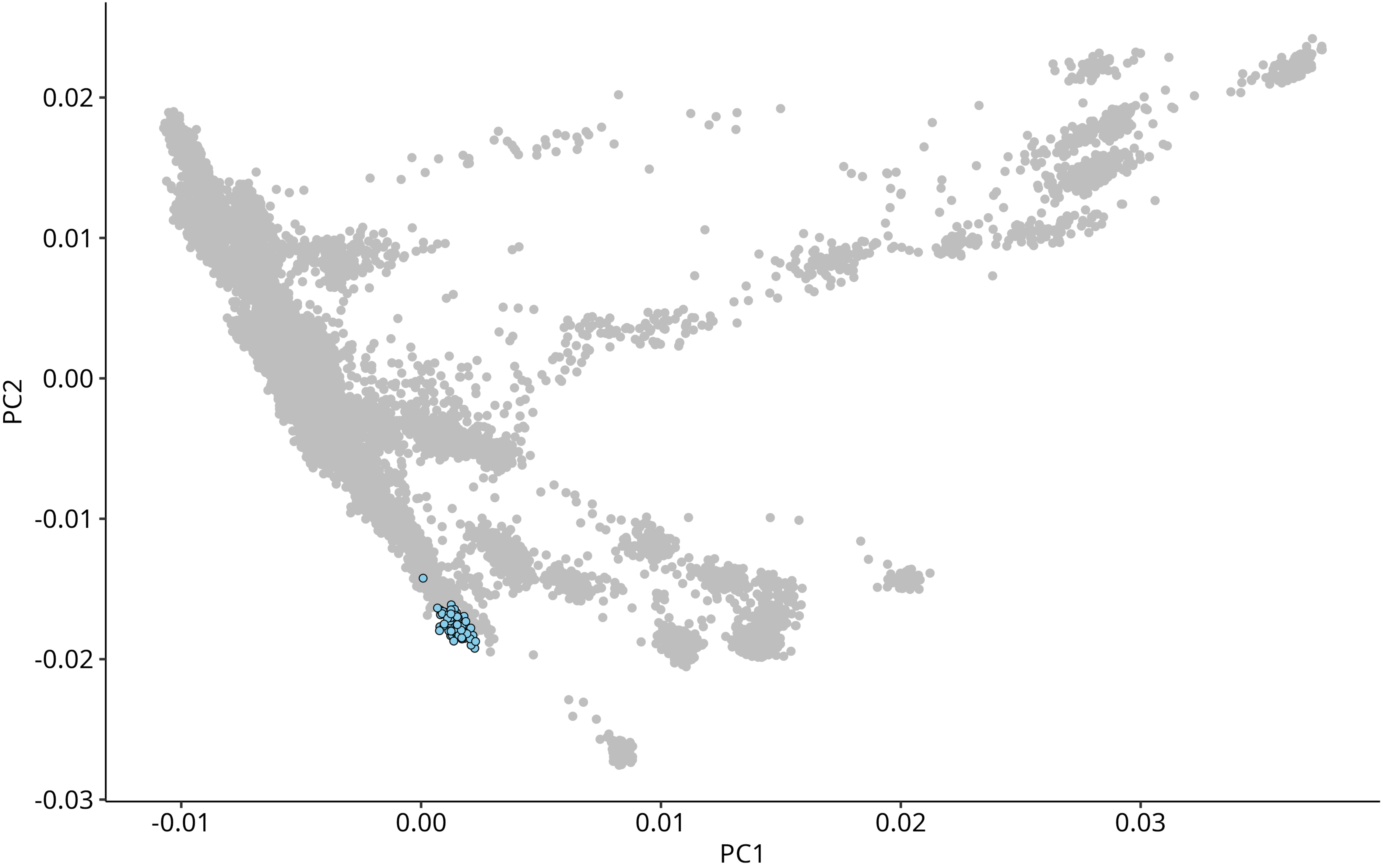

Population: AA_EPL_1_01, Order: 63

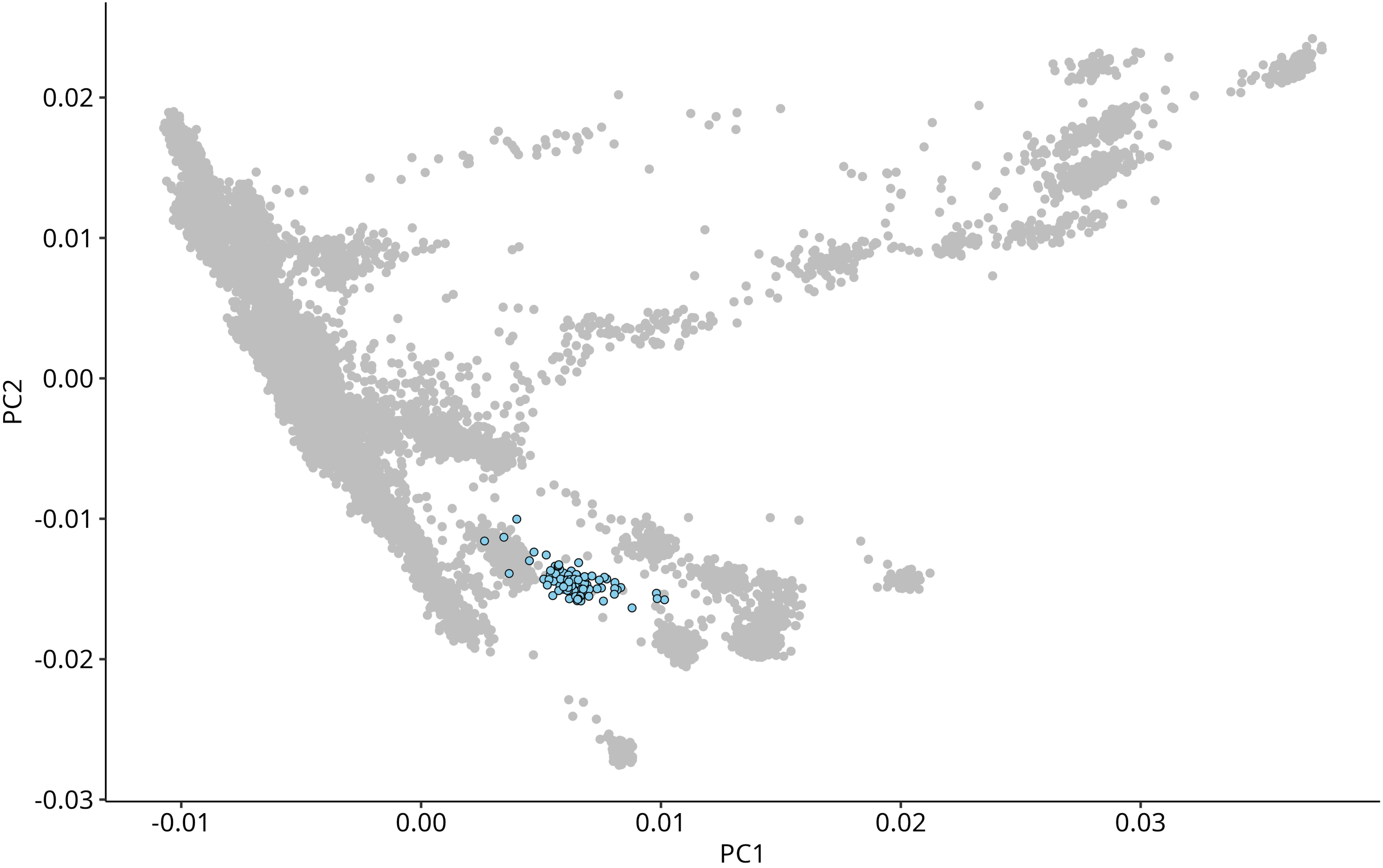

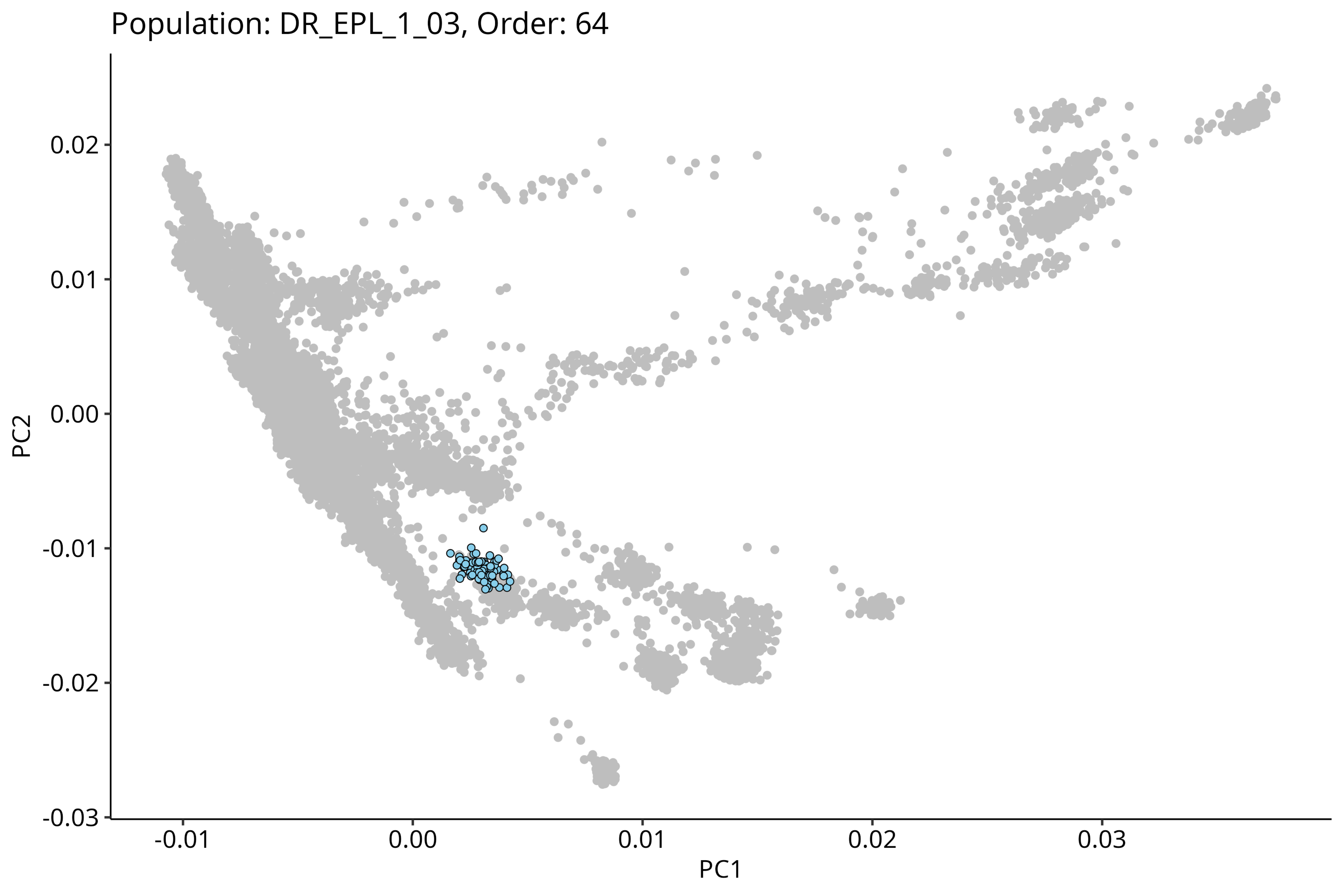

Population: DR_EPL_1_03, Order: 64
PC2
0.02
0.01
0.00
-0.01
-0.02
-0.03
PC1
-0.01
0.00
0.01
0.02
0.03

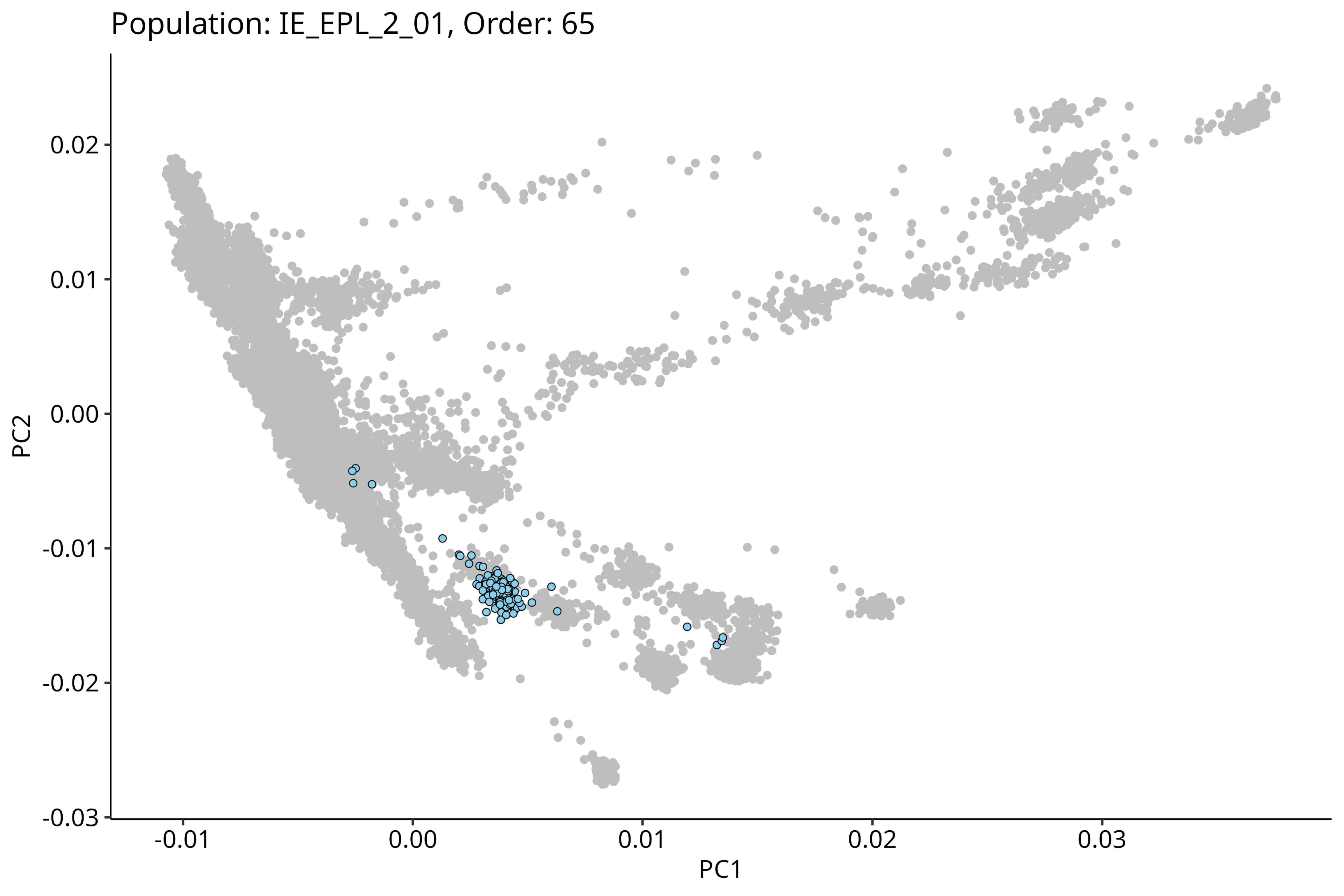
Population: IE_EPL_2_01, Order: 65
PC2
PC1
0.02
0.01
0.00
-0.01
-0.02
-0.03
-0.01
0.00
0.01
0.02
0.03

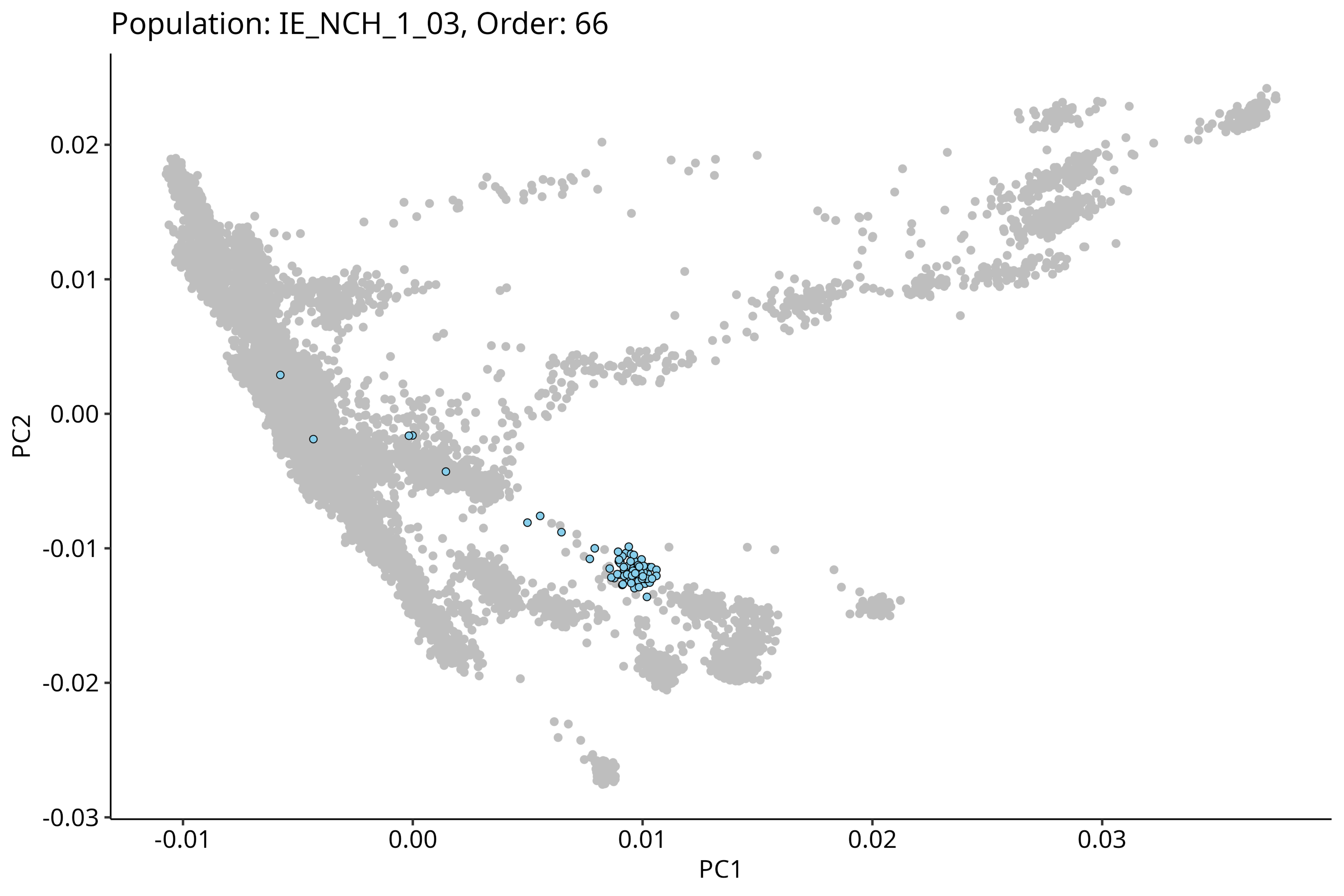

Population: IE_NCH_1_03, Order: 66
0.02
0.01
0.00
-0.01
-0.02
-0.03
PC2
-0.01
0.00
0.01
0.02
0.03
PC1

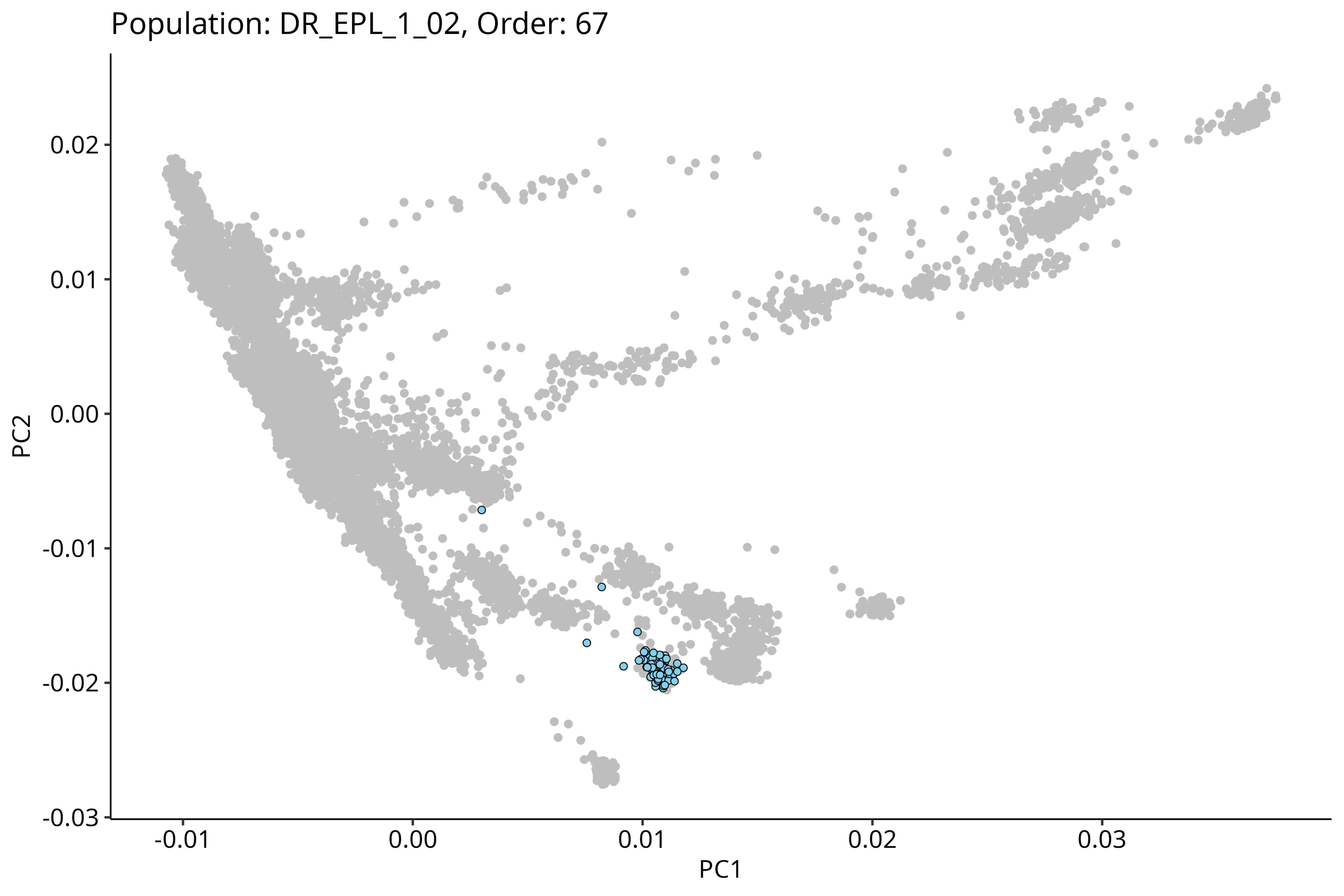

Population: DR_EPL_1_02, Order: 67
PC2
PC1
0.02
0.01
0.00
-0.01
-0.02
-0.03
-0.01
0.00
0.01
0.02
0.03

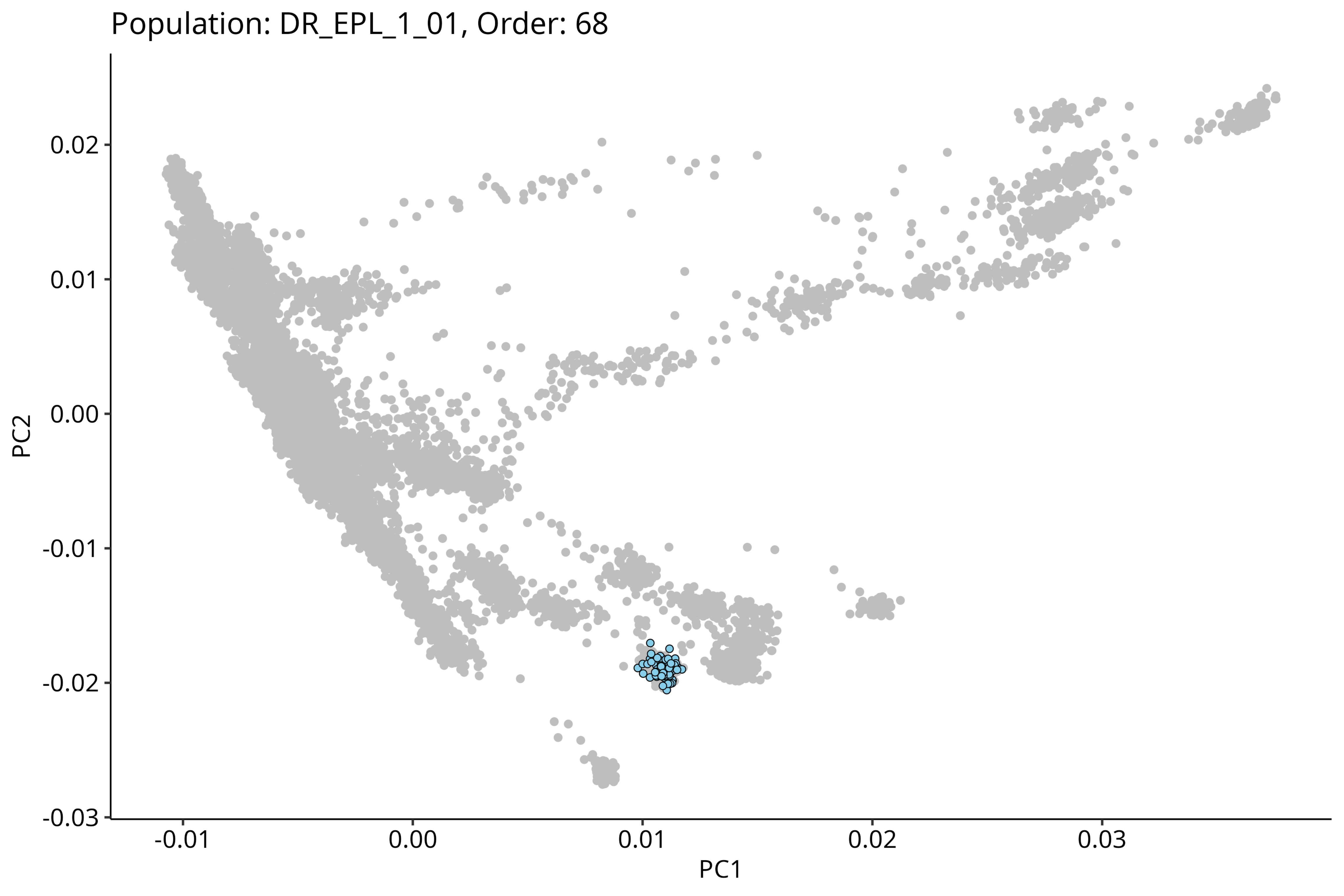
Population: DR_EPL_1_01, Order: 68
PC2
PC1
0.02
0.01
0.00
-0.01
-0.02
-0.03
-0.01
0.00
0.01
0.02
0.03

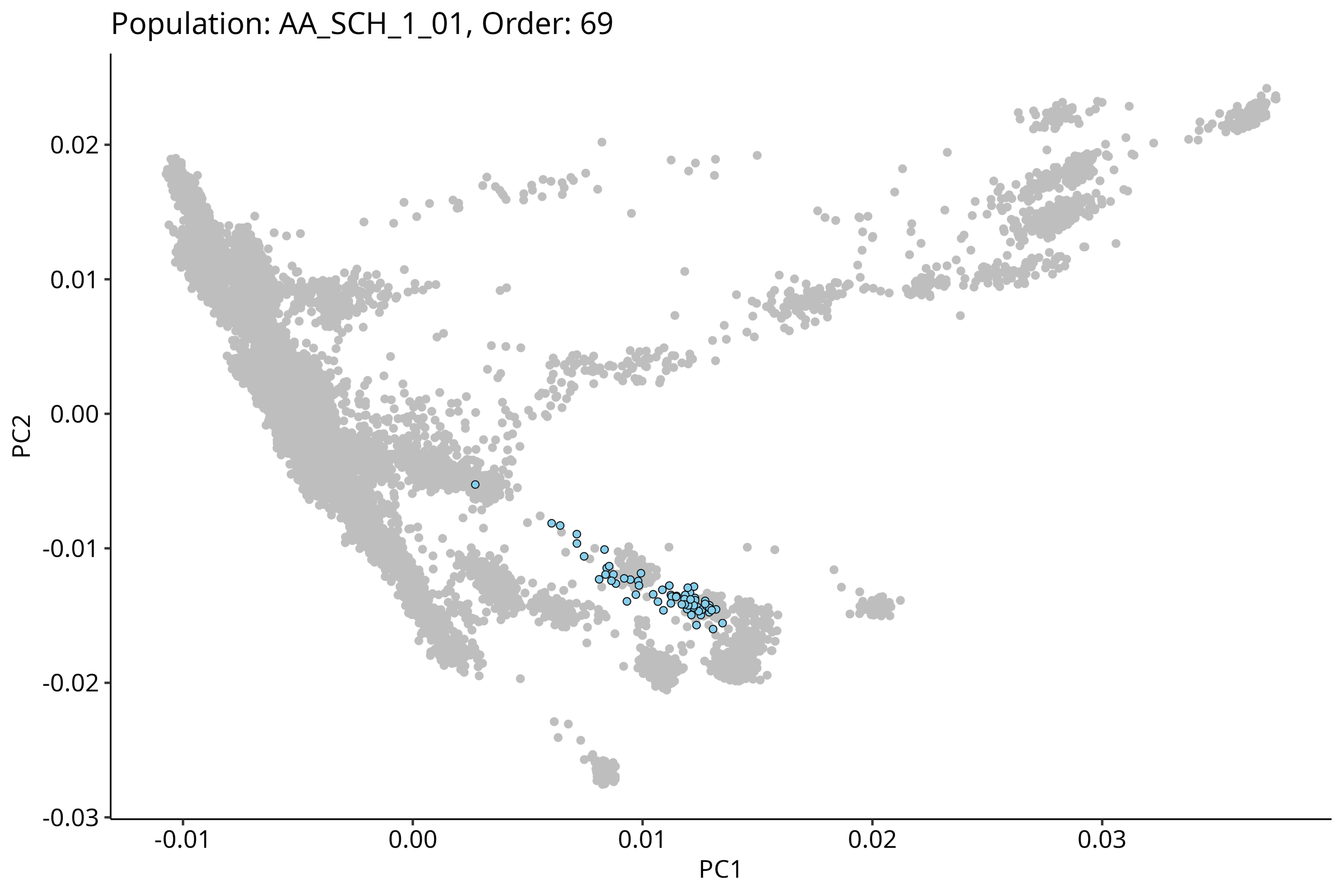
Population: AA_SCH_1_01, Order: 69
0.02
0.01
0.00
-0.01
-0.02
-0.03
PC2
-0.01
0.00
0.01
0.02
0.03
PC1

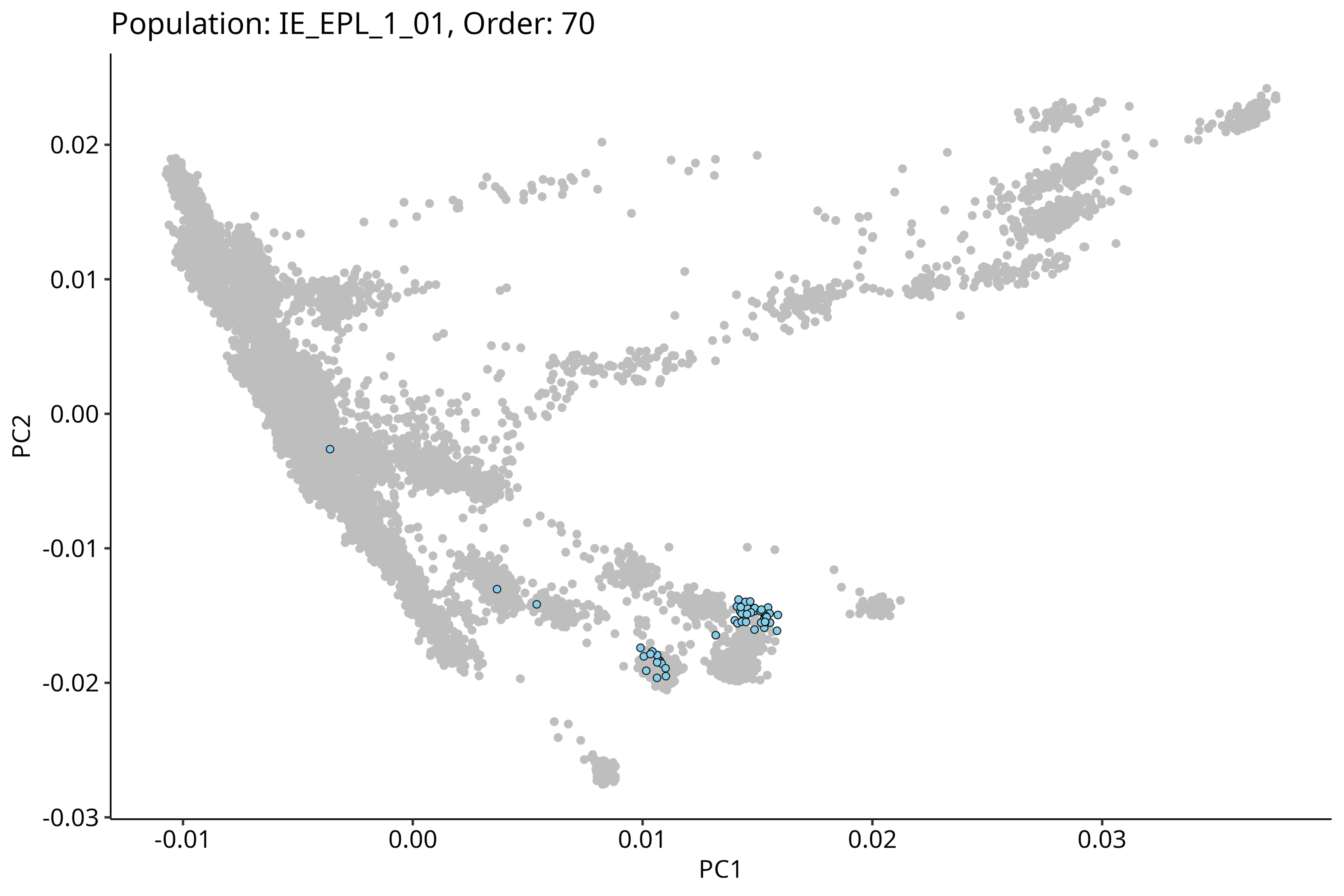

Population: IE_EPL_1_01, Order: 70
PC2
0.02
0.01
0.00
-0.01
-0.02
-0.03
-0.01
0.00
0.01
0.02
0.03
PC1

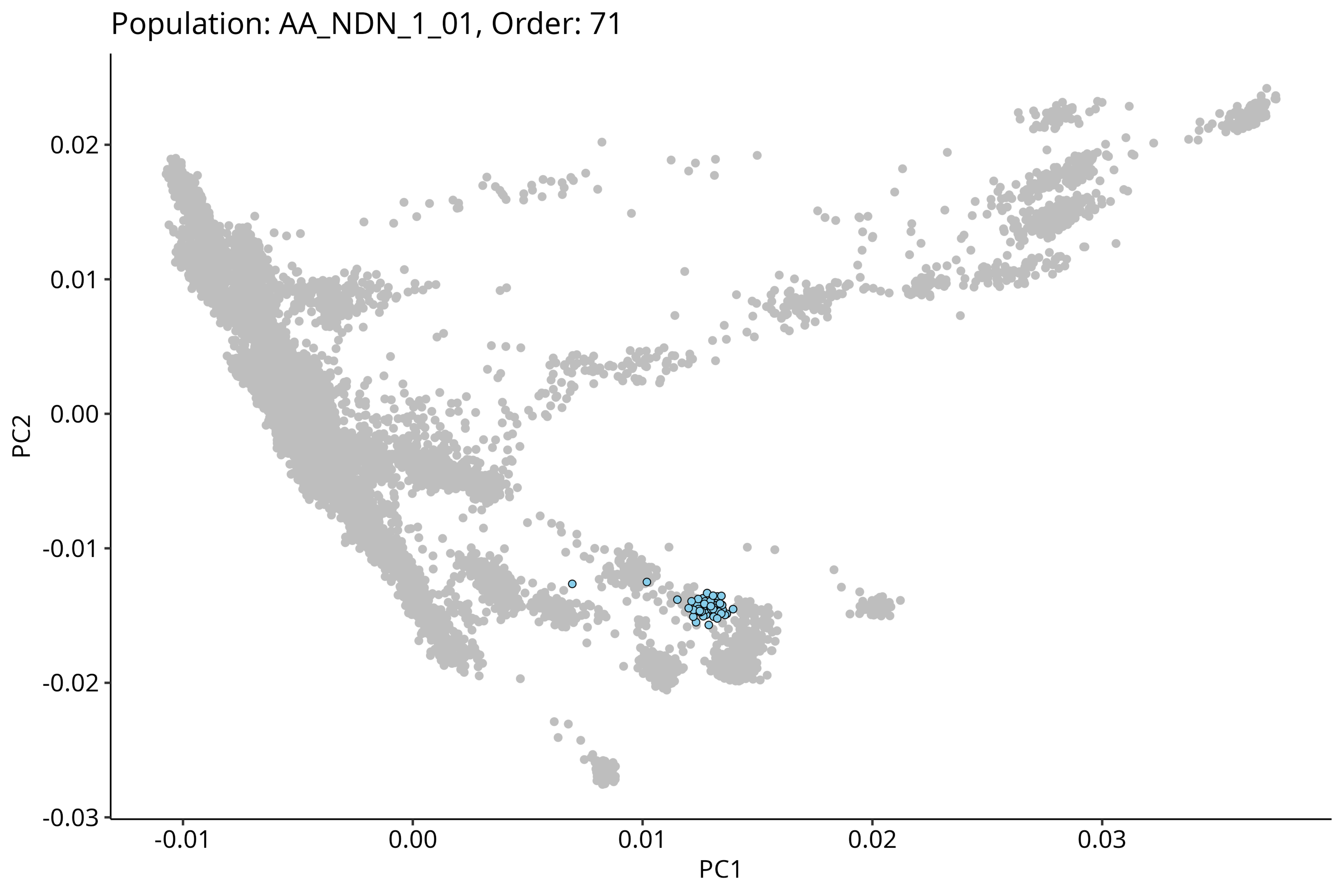
Population: AA_NDN_1_01, Order: 71
0.02
0.01
0.00
-0.01
-0.02
-0.03
PC2
-0.01
0.00
0.01
0.02
0.03
PC1

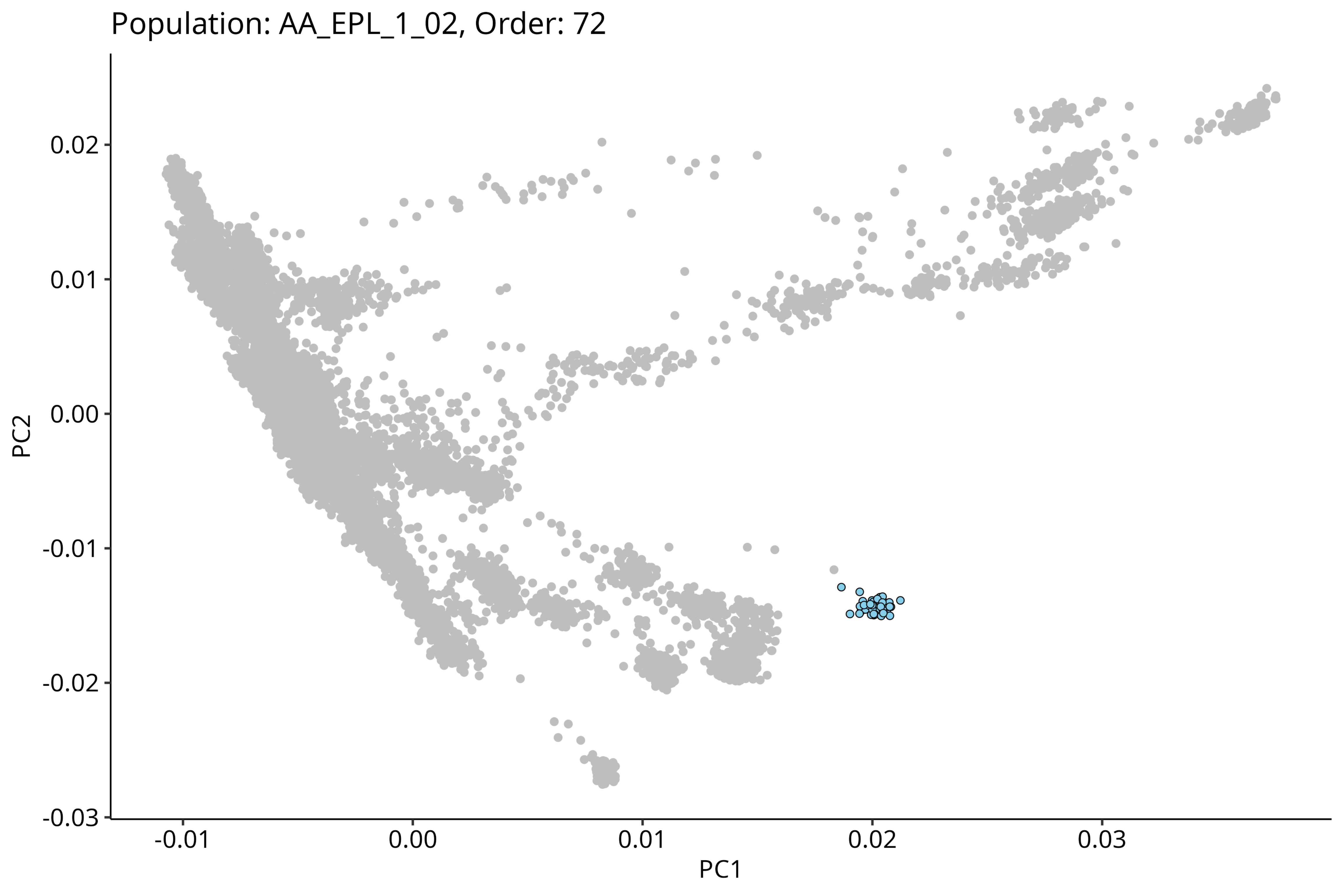

Population: AA_EPL_1_02, Order: 72
0.02
0.01
0.00
-0.01
-0.02
-0.03
PC2
-0.01
0.00
0.01
0.02
0.03
PC1

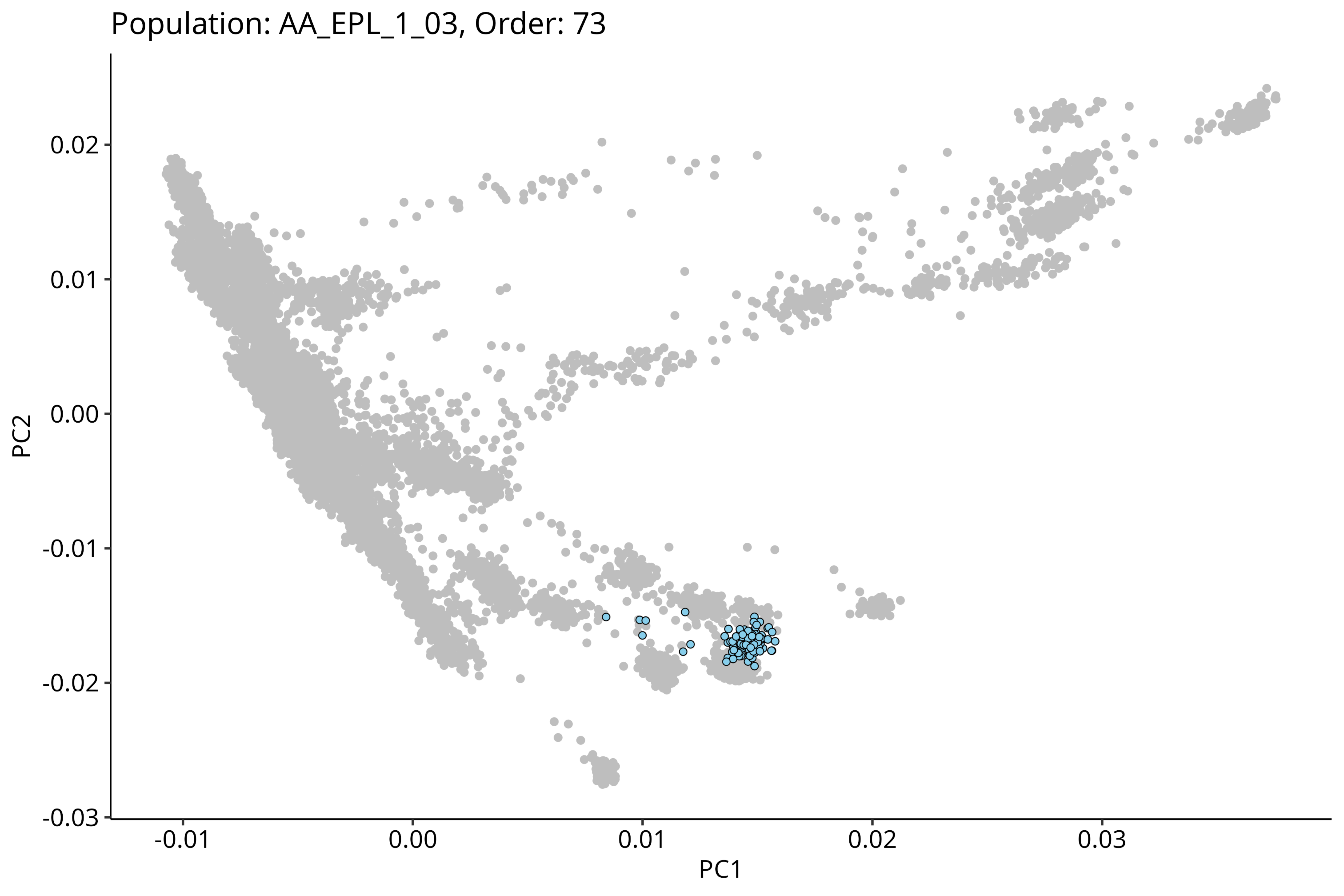

Population: AA_EPL_1_03, Order: 73
0.02
0.01
0.00
-0.01
-0.02
-0.03
PC2
-0.01
0.00
0.01
0.02
0.03
PC1

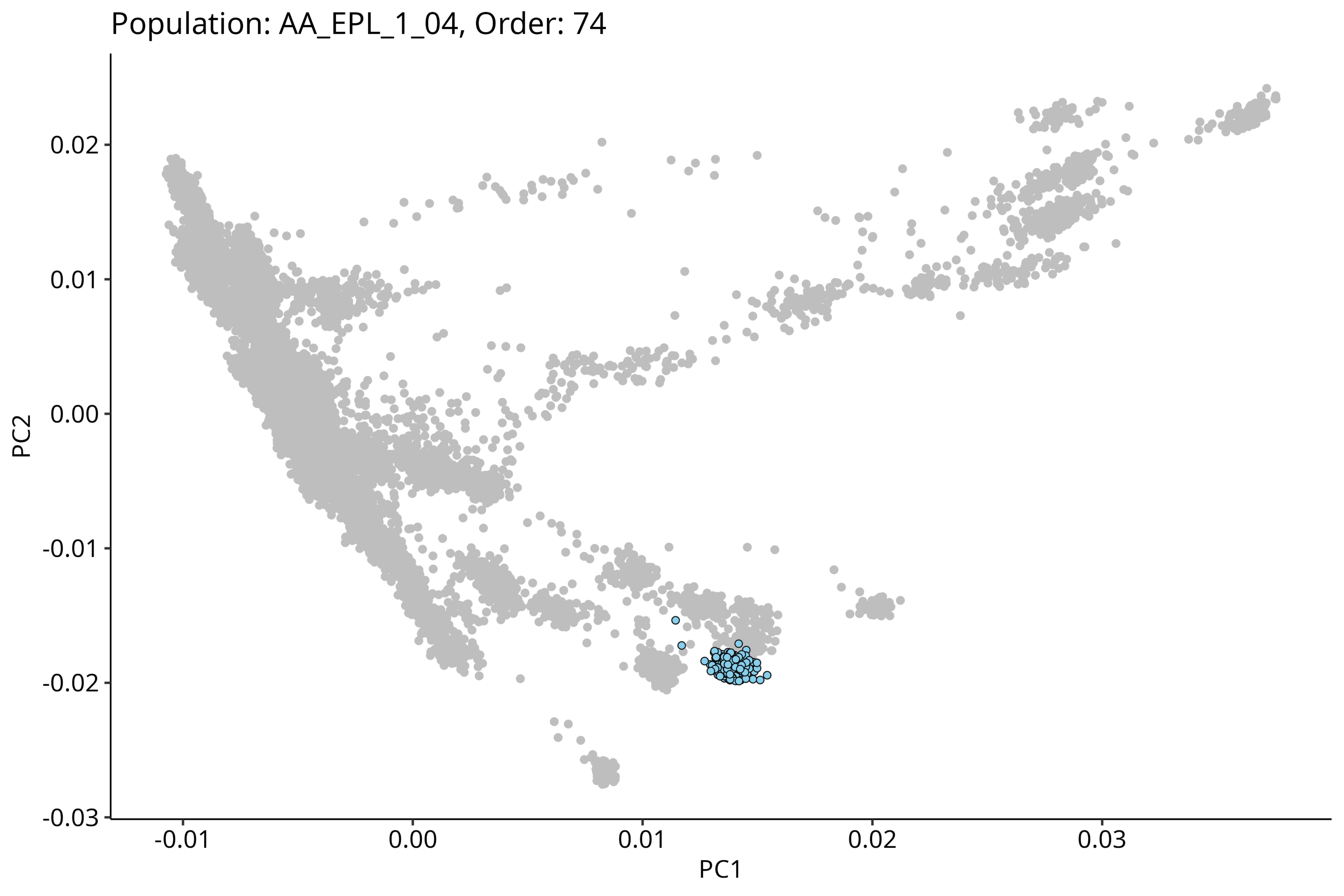
Population: AA_EPL_1_04, Order: 74
0.02
0.01
0.00
-0.01
-0.02
-0.03
PC2
-0.01
0.00
0.01
0.02
0.03
PC1

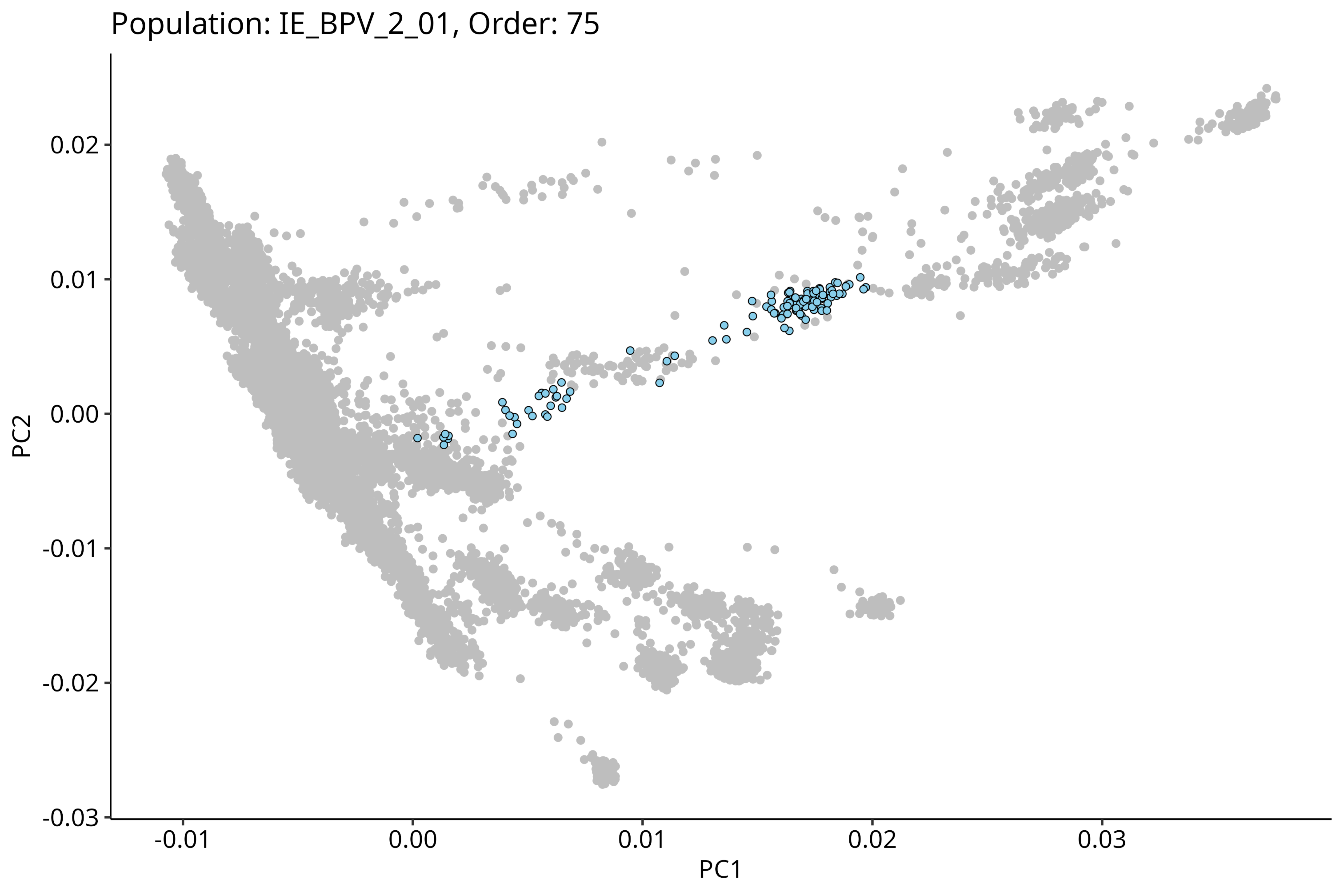
Population: IE_BPV_2_01, Order: 75
PC2
0.02
0.01
0.00
-0.01
-0.02
-0.03
PC1
-0.01
0.00
0.01
0.02
0.03

Population: IE_CHR_1_01, Order: 76

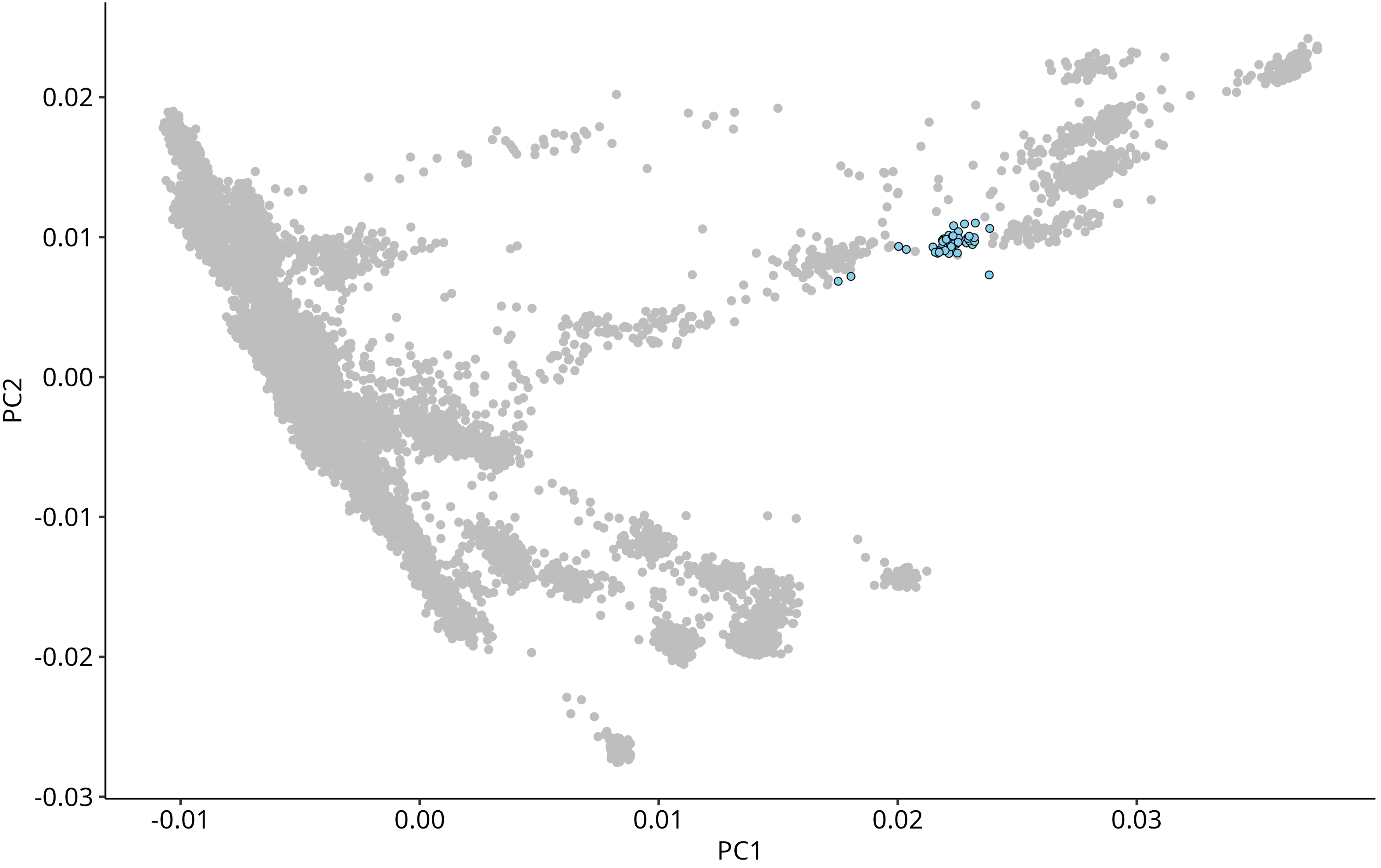

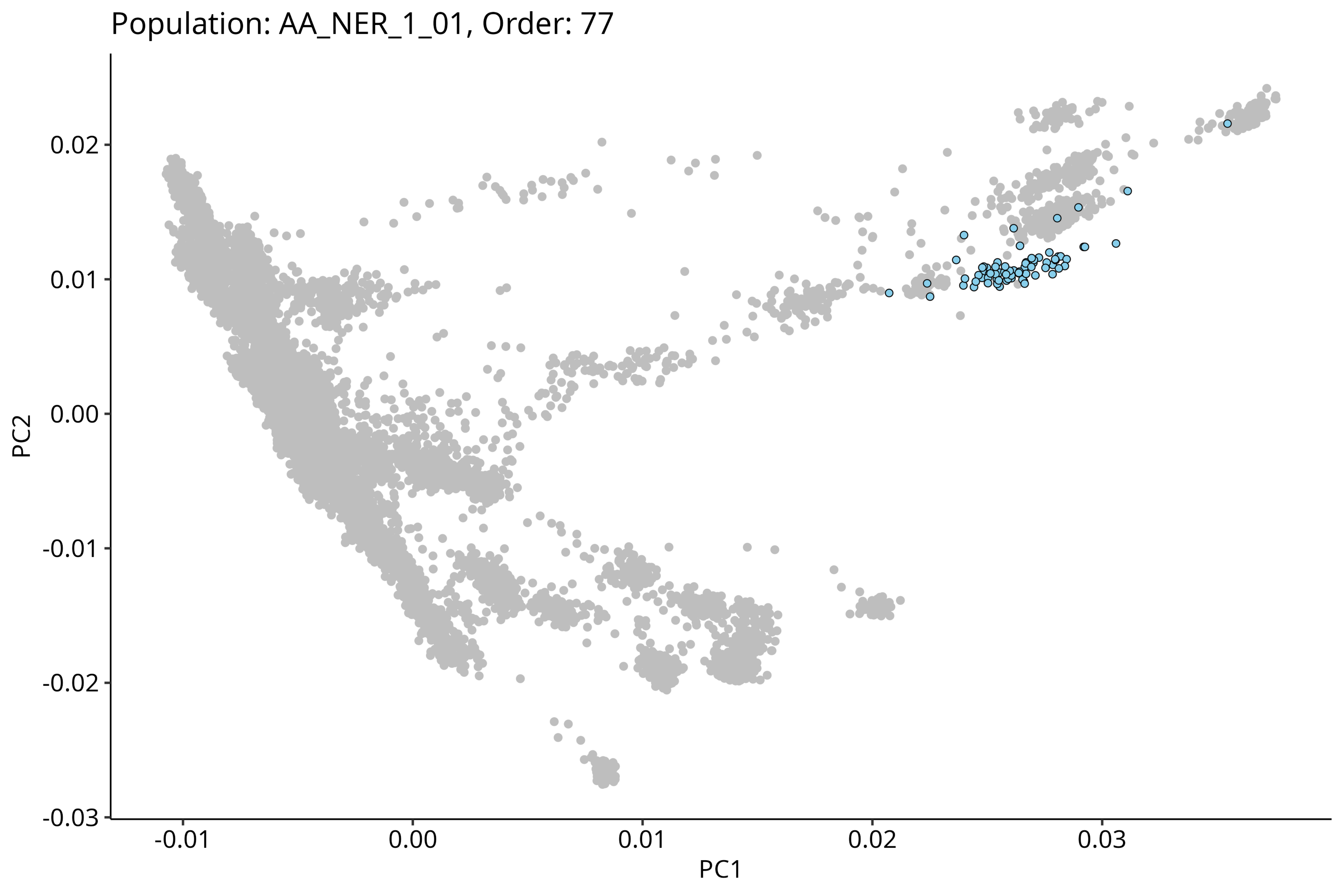

Population: AA_NER_1_01, Order: 77
0.02
0.01
0.00
-0.01
-0.02
-0.03
PC2
-0.01
0.00
0.01
0.02
0.03
PC1

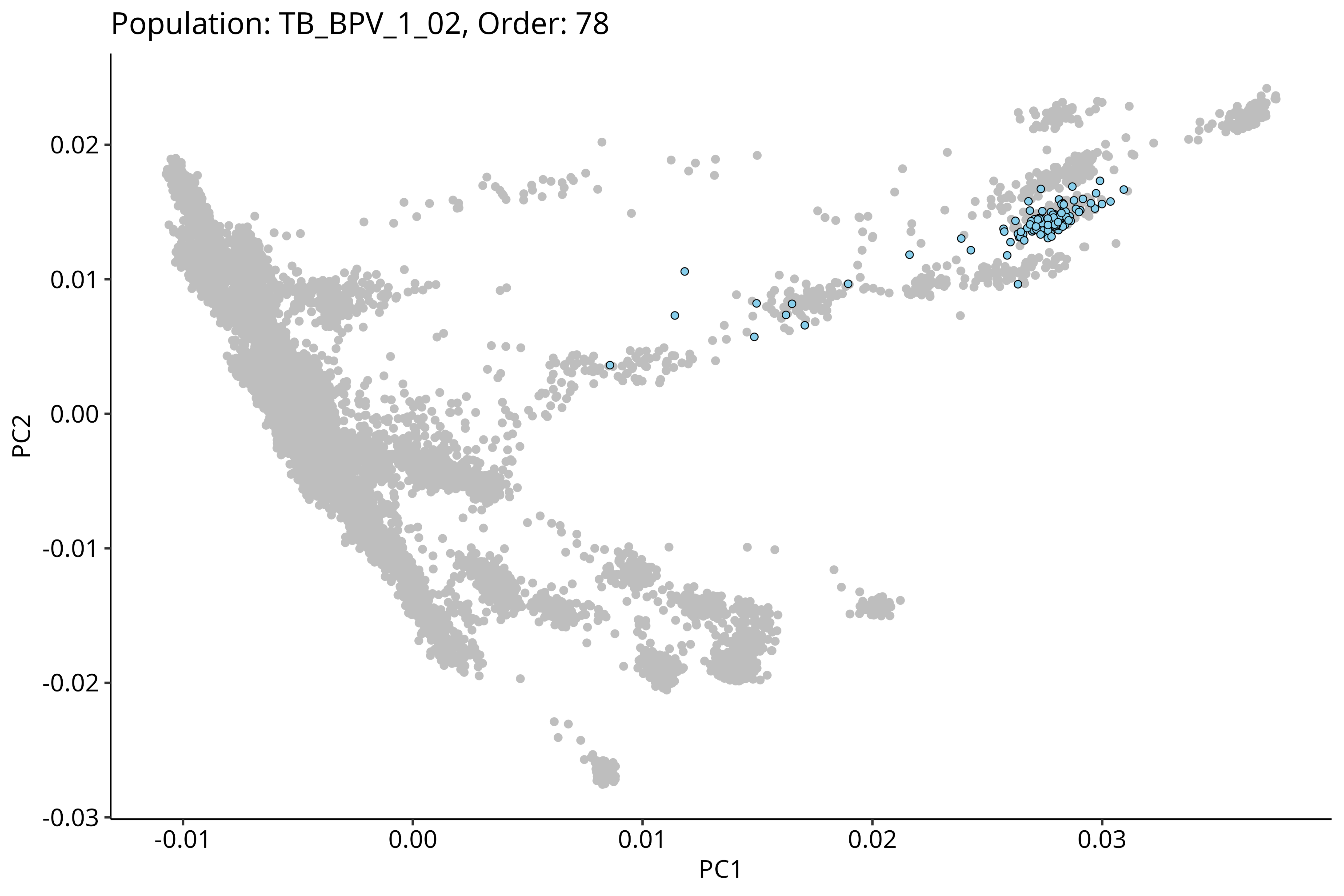
Population: TB_BPV_1_02, Order: 78
PC2
0.02
0.01
0.00
-0.01
-0.02
-0.03
-0.01
0.00
0.01
0.02
0.03
PC1

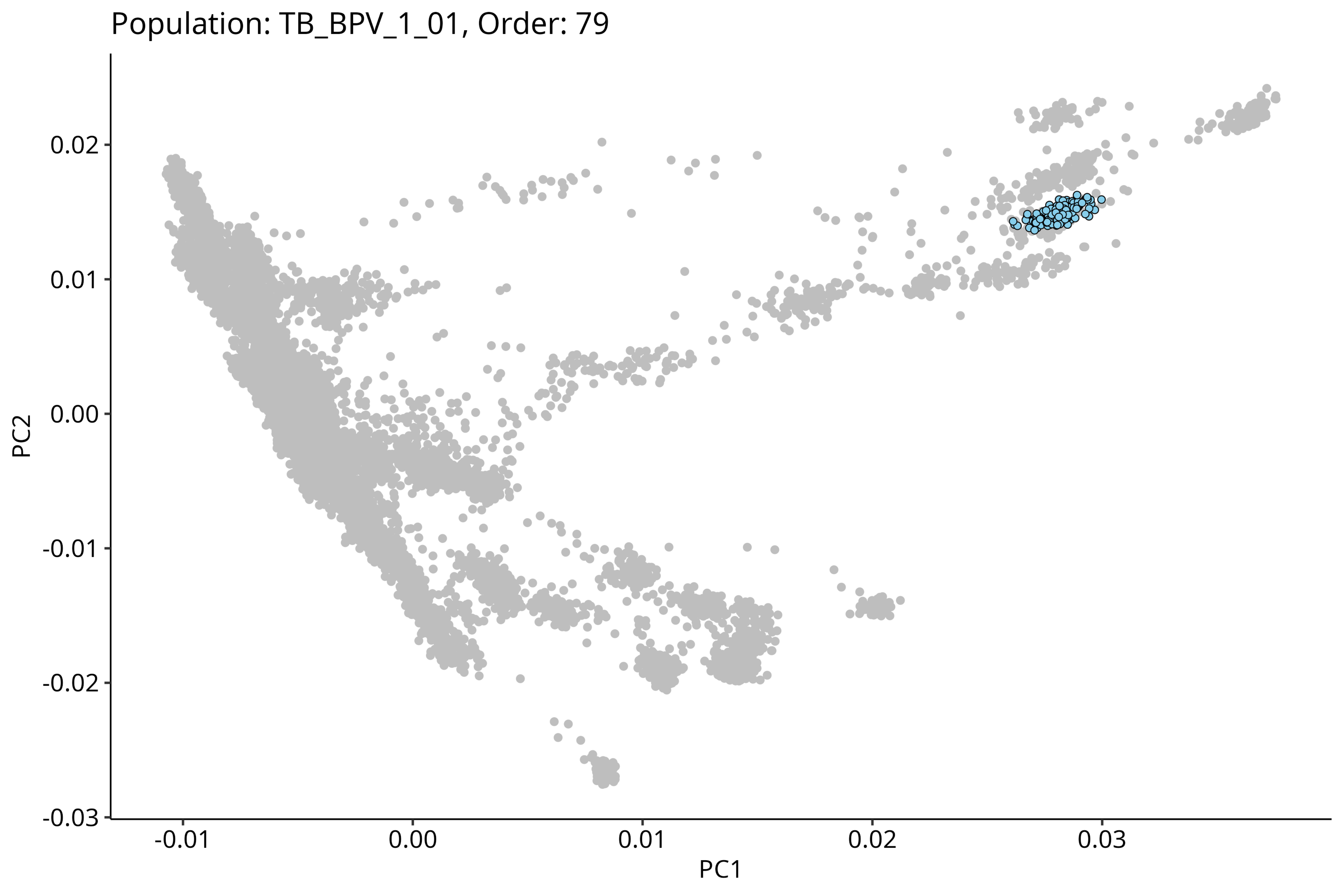
Population: TB_BPV_1_01, Order: 79
0.02
0.01
0.00
-0.01
-0.02
-0.03
PC2
-0.01
0.00
0.01
0.02
0.03
PC1

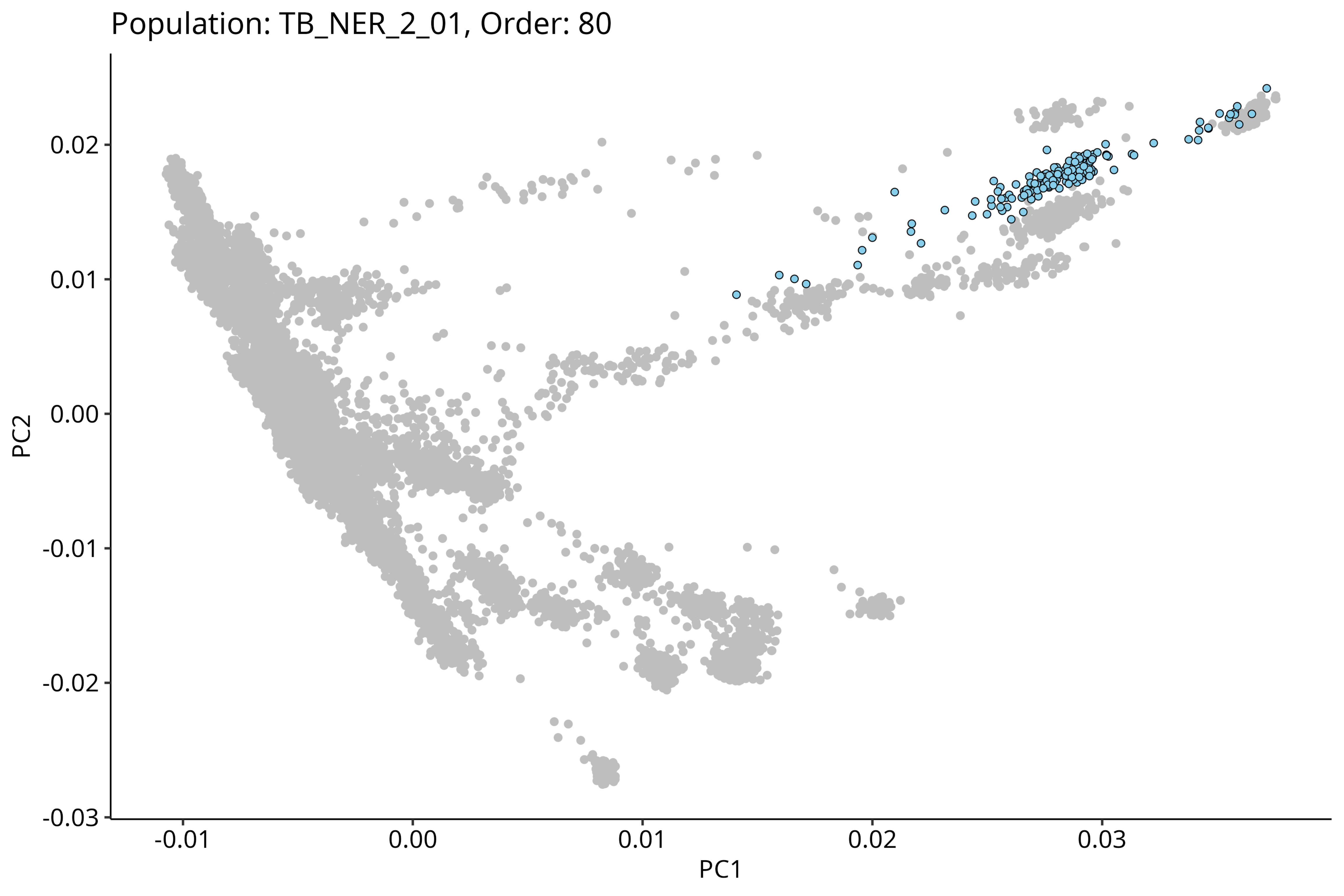

Population: TB_NER_2_01, Order: 80
PC2
PC1
0.02
0.01
0.00
-0.01
-0.02
-0.03
-0.01
0.00
0.01
0.02
0.03

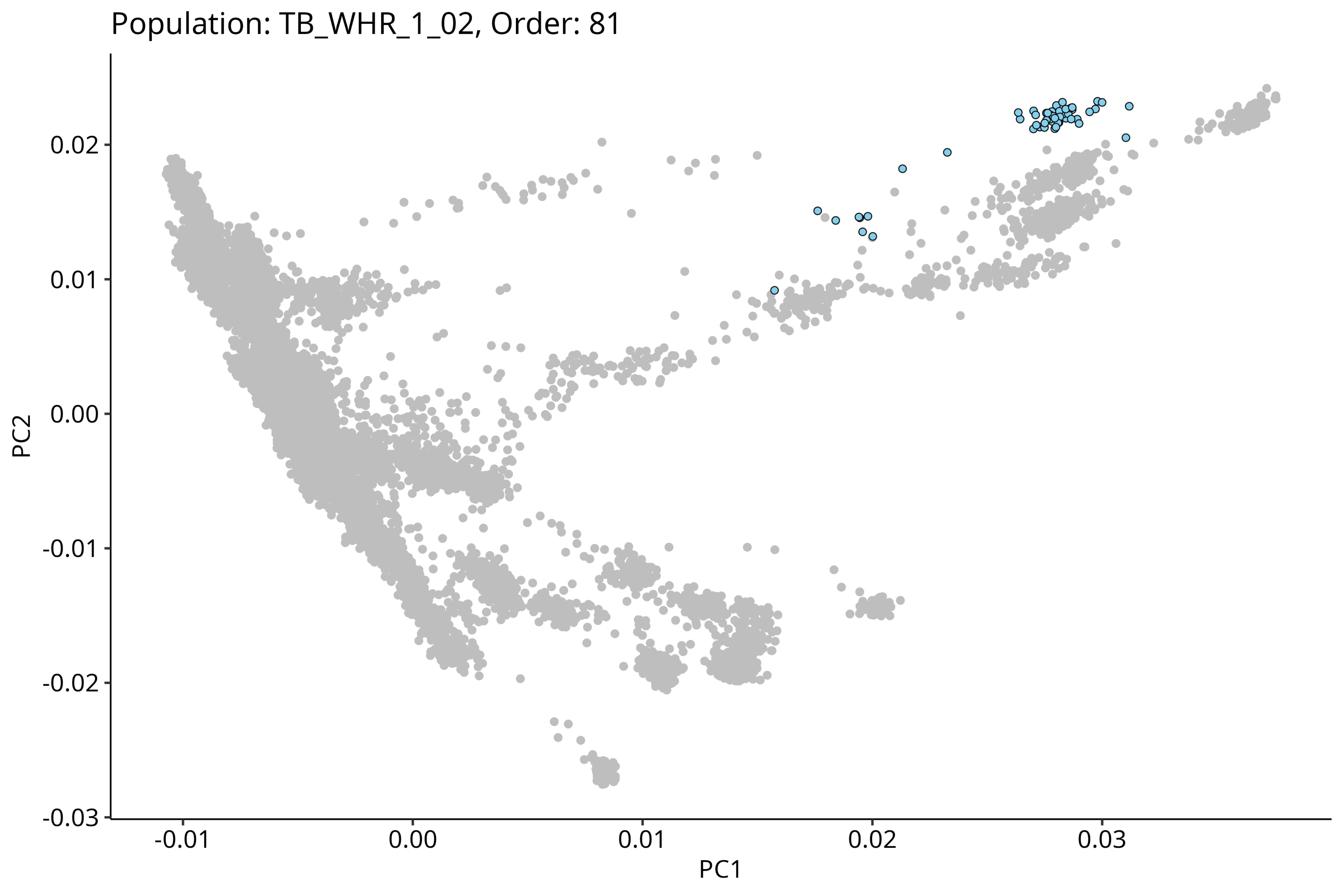

Population: TB_WHR_1_02, Order: 81
0.02
0.01
0.00
-0.01
-0.02
-0.03
PC2
-0.01
0.00
0.01
0.02
0.03
PC1

Population: TB_NER_1_01, Order: 82

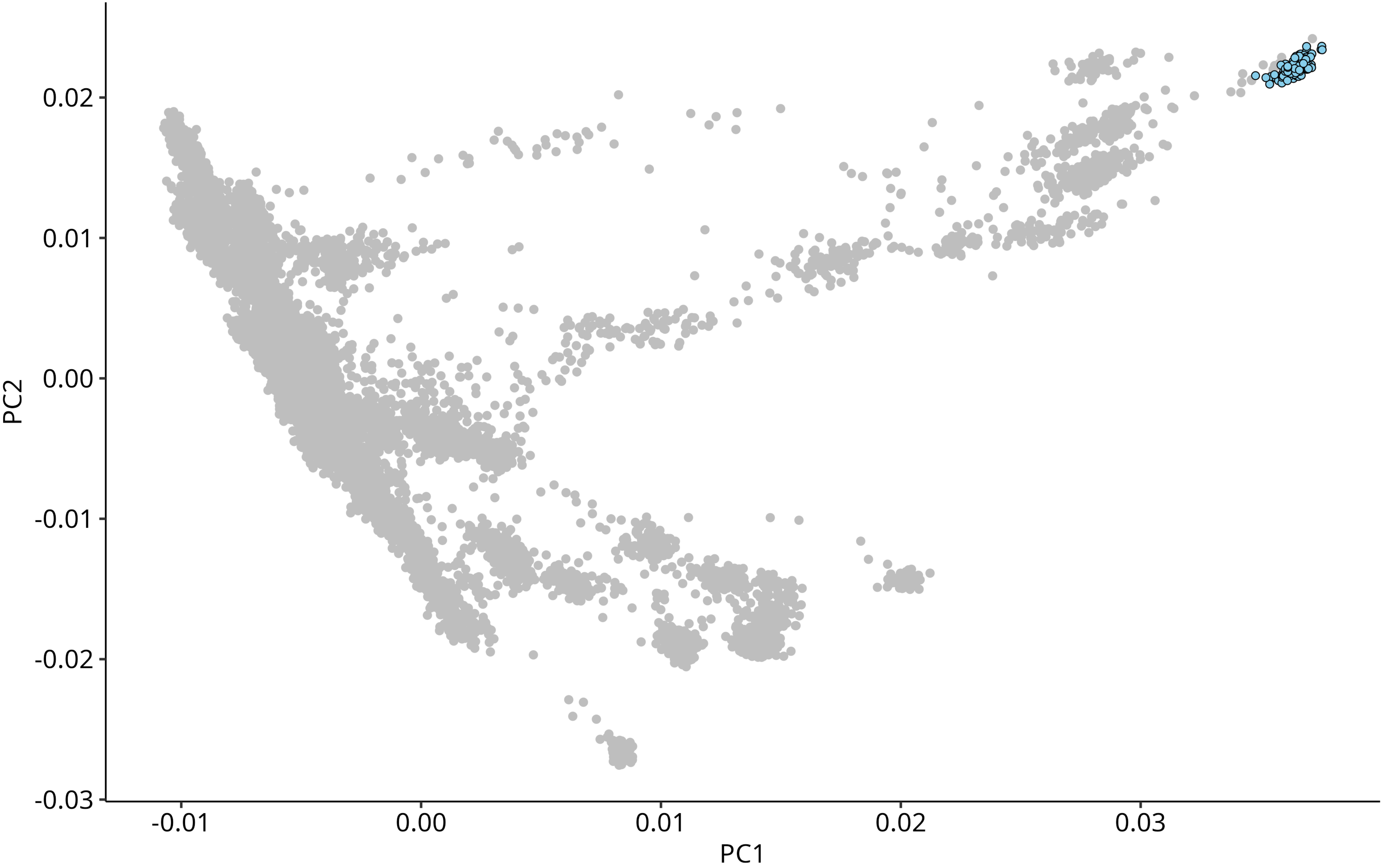
